# Temporal trends in the occurrence, mortality, and cost of brain disorders in Denmark

**DOI:** 10.1101/2024.09.18.24313799

**Authors:** Cecilia Hvitfeldt Fuglsang, Thomas Bøjer Rasmussen, Jan Håkon Rudolfsen, Jens Olsen, Niels Skipper, Sinna Pilgaard Ulrichsen, Henrik Toft Sørensen, Christian Fynbo Christiansen

**Author notes:** **Corresponding author:** Cecilia Hvitfeldt Fuglsang. **Declaration of interests:** The Department of Clinical Epidemiology, Aarhus University and Aarhus University Hospital, receives funding from private and public institutions in the form of research grants to (and administered by) Aarhus University. **Funding:** This work was funded by the Lundbeck Foundation (grant number: R433-2023-1140).

## Abstract

**Background:** Brain disorders, including neurological and mental disorders, are common and burdensome diseases.

**Aim:** To examine temporal trends in occurrence, mortality, and cost of brain disorders in Denmark for the period of 2015–2021.

**Methods:** We conducted a nationwide population-based cohort study using individual-level data recorded in the Danish health registries during 2011–2021. We computed the prevalence of any brain disorder and 25 individual brain disorders for each year from 2015 to 2021 and the incidence for the periods 2011–2015 and 2016–2021 by combining hospital diagnoses and filled prescriptions for relevant medications. We computed one-year hazard ratios (HRs) for mortality by comparing individuals with brain disorders to matched controls without. We also calculated attributable direct and indirect costs (i.e., lost productivity) of brain disorders.

**Results:** The prevalence of any brain disorder in Denmark was 33.2% in 2015, increasing to 35.2% in 2021. The three most prevalent conditions were depression (13.5% in 2021), sleep disorders (13.5% in 2021), and headache (7.9% in 2021). The incidence rate of any brain disorder was 1,792 and 1,634 per 100,000 person-years in 2011-2015 and 2016-2021, respectively. The one-year HR of mortality for any brain disorder was 5.5 (95% confidence internal [CI]: 5.4; 5.6) for 2011–2015 and 5.3 (95% CI: 5.2; 5.3) for 2016–2021. The total attributable direct costs for individuals with any brain disorder were €7.5 billion in both 2015 and 2021. In 2021, the total attributable indirect costs were highest for depression (∼€3.5 billion) and lowest for neuromuscular disorders (∼€53.5 million). Total indirect costs increased from €17.7 billion in 2015 to €23.2 billion in 2021. In 2021, the total indirect costs were highest for depression (∼€14 billion) and lowest for other neurodegenerative disorders (∼€60 million).

**Conclusion:** Brain disorders remain common, with a fivefold higher one-year mortality compared to persons without brain disorders. While total direct costs were similar in 2015 and 2021, total indirect costs increased over this period.

## Introduction

Disorders of the brain include neurologic diseases and mental health disorders. In 2021, The Global Burden of Disease study identified neurologic diseases as the primary cause of disability-adjusted life years (DALYs), with a contribution of 443 million DALYs that year alone.^1^ Between 1990 and 2019, mental disorders leapt in the DALY rankings from 13^th^ to 7^th^ place, with 125 million DALYs in 2019.^2^ The significant burden of these disorders is partly due to their high prevalence. According to data from the Global Burden of Disease study, 4 billion people (∼50%) worldwide were affected by a brain disorder in 2021.^3^ Such prevalence estimates may vary according to how brain disorders are identified. In a Danish study estimating the occurrence, mortality, and cost of brain disorders in 2011-2015, 20% of Danish residents alive on January 1, 2015, had been diagnosed in hospital with at least one brain disorder.^4^ The brain disorders in the Danish study were also associated with substantial costs. E.g., the direct costs of healthcare attributed to brain disorders were €5.2 billion in 2015.^4^ This was also evident in a study based on data from 30 European countries, where the total direct healthcare costs of brain disorders were estimated at €295 billion in 2010.^5^ To our knowledge, no other recent studies have examined the costs of brain disorders overall, nor the changes in these costs over time. In particular, the demographic shift towards older populations^6^ is likely to alter the prevalence and thus the costs of brain disorders. At the same time, both prevalence and costs may be affected by reduced mortality as the treatment of disorders such as stroke^7^ and multiple sclerosis has improved.^8^ Finally, increasing awareness of mental health disorders is likely contributing to increasing rates of diagnoses^9^ and costs. Thus, to ascertain the full burden of brain disorders, continuous updates are needed. Therefore, the aim of this study was to examine temporal trends in the occurrence, mortality, and cost of brain disorders in Denmark from 2015 to 2021.

## Methods

### Study design and setting

We conducted a nationwide population-based cohort study using individual-level data recorded in the Danish health registries during 2015–2021. In Denmark, all residents have access to the tax-funded healthcare system, and healthcare is free at the point of care.^10^ At immigration or birth, all persons are assigned a unique personal identifier, which is recorded in the Civil Registration System.^11^ This registry is continually updated with vital and immigration status. The personal identifier is used in all nationwide administrative and medical registries, thus enabling individual-level data linkage across these registries.^10^

### Study cohorts

The source population consisted of all individuals alive and residing in Denmark at some point between January 1, 1995, and December 31, 2021 (n=8,033,237). We examined 25 groups of brain disorders: alcohol abuse, anxiety disorders, bipolar disorder, brain tumors, cerebral palsy, dementia, depression, developmental and behavioral disorders, drug abuse, eating disorders, epilepsy, headache, infections of the central nervous system (CNS), intellectual disability, multiple sclerosis, neuromuscular disorders, other neurodegenerative disorders, Parkinson’s disease, personality disorders, polyneuropathy, schizophrenia spectrum disorders, sleep disorders, stress-related disorders, stroke, and traumatic brain injury (Appendix 1).^4^ For each of the 25 specific brain disorders and for each year during 2015–2021, we established a prevalent cohort comprising persons alive on 1 January of the given year who had a specific brain disorder recorded during the previous 20 years. Thus, for the 2015 prevalent cohort, we identified persons alive on January 1, 2015, who had a brain disorder diagnosis at some point during 1995–2014. We also established two incident cohorts for each brain disorder, consisting of persons with a first-time diagnosis recorded during 2011–2015 or 2016–2021. For the incident cohorts, we required no previous brain disorder diagnosis within 16 years before 2011 and 2016, respectively. Furthermore, we established a cohort of persons with any brain disorder based on all inclusion criteria for the 25 disorders. To avoid double counting, every person was counted only at the first diagnosis in any brain disorder cohort. The index date was the date of the brain disorder diagnosis for the incident cohorts and January 1 of the given year for the prevalent cohorts.

#### Matched comparison cohorts

For each individual brain disorder cohort and the cohort of any brain disorder, we created a matched comparison cohort. Each person with a brain disorder was matched exact with ten persons from the general population, by birth year and sex, using sampling with replacement. Matched persons were required to be living in Denmark and to not have been diagnosed with the brain disorder of interest prior to the index date of the person with the brain disorder. Controls were assigned the index date corresponding to that of their matched individual with a brain disorder.

#### Brain disorders

Brain disorders were identified by hospital diagnoses recorded in the Danish National Patient Registry (DNPR)^12^ and redeemed prescriptions recorded in the Danish National Prescription Registry (NPR)^13^ as proxies for brain disorder diagnoses treated outside hospitals. From the DNPR, we obtained information on all hospital contacts in Denmark from 1995 onwards. Diagnoses in the DNPR have been recorded with the World Health Organization’s International Classification of Diseases 10^th^ revision (ICD-10) codes since 1994.^12^ The NPR contains records of all redeemed prescriptions at community pharmacies since 1995.^13^ The definitions of the brain disorders in our primary analyses were adapted from those used in the sensitivity analysis of a previously published study on brain disorders in Denmark including redeemed prescriptions.^4^

#### Charlson comorbidity index and mortality

Information on comorbidities from the ten years preceding the index date was assessed using the Charlson Comorbidity index (CCI).^14^ We identified diagnoses in the DNPR as specified by the CCI (ICD-10 codes in Appendix 1) and calculated individual-level CCI score. Persons were then categorized according to low (CCI score = 0), moderate (CCI score = 1–2), or high (CCI score >2) comorbidity burden. Mortality was estimated using dates of death obtained from the civil registration system for persons who died during the study period.^11^

#### Costs

We estimated both direct and indirect costs of brain disorders. Direct costs included the costs of primary and hospital care. To estimate the costs of primary care, we obtained information on all primary care services provided by general practitioners (GPs), private practicing specialists, and dentists from the Danish Health Service Registry;^15^ information on nursing homes from the Nursing Home Registry;^16^ and information on sheltered accommodation, personal nursing and other personal care, home nurse visits, and hospital-based neurorehabilitation from Statistics Denmark.^17^

Costs of secondary care were estimated based on hospital inpatient admissions, outpatient clinic visits, and emergency department contacts according to the Diagnosis-Related Group (DRG) and Danish Ambulatory Grouping System (DAGS) tariffs. Since DRG/DAGS tariffs for psychiatric hospital contacts have been unavailable since 2019, values were imputed using data from previous years (2013–2018). A detailed description of this imputation is provided in Appendix 2. Medication costs of primary and secondary care were estimated using market prices for prescriptions filled at outpatient pharmacies recorded in the NPR,^13^ and in-hospital medication costs were included in the DRG/DAGS tariffs. Indirect costs were estimated as lost productivity using information on earned income from Statistics Denmark.

For the prevalent cohorts and their comparison cohorts, all cost information was obtained for the given year. For the incident cohorts, costs were obtained from the index date and one year thereafter. All costs and wages except medication costs were adjusted to 2023 levels according to Statistics Denmark’s consumer price index. Currency conversion from DKK to EUR was performed with a fixed currency exchange rate of EUR 1 = 7.45 DKK.

### Statistical analyses

#### Occurrence

The cohorts of persons with brain disorders were described according to age, sex, CCI conditions, and CCI scores. Occurrence was estimated as prevalence and incidence. Prevalence in a given year was assessed for persons alive on January 1 of that year and any diagnosis or redeemed prescription during the prior 20 years. The incidence rates per 100,000 person-years (PYs) were estimated for 2011–2015 and 2016–2021 according to first-time diagnoses or prescriptions.

#### Mortality

One-year mortality was estimated using the Kaplan Meier estimator. Using Cox regression, we computed crude and adjusted hazard ratios (HRs) of one-year mortality, comparing persons with brain disorders to persons in the comparison cohorts. HRs were adjusted for age, sex, and CCI score. Visual inspection of log (-log) survival plots did not indicate violation of the proportional hazards assumption.

#### Costs of illness

Using the human capital approach, we conducted a societal cost-of-illness analysis including individual-level direct and indirect costs. Both overall annual costs and the average annual cost per person were estimated for any brain disorder and for each brain disorder. For direct costs, we computed both actual direct costs and attributable direct costs; the latter were determined as the difference between the healthcare service costs for persons with brain disorders and the mean costs for the matched persons in the comparison cohorts. Since the costs of illness might be particularly high shortly after diagnosis, we computed the attributable direct costs per person in the first year after diagnosis for the incident cohorts. In terms of indirect costs, we estimated the loss of productivity for persons of working age (18–65 years) by subtracting the mean personal earned income of the matched comparison cohort from the personal income of persons with brain disorders before taxes. If a person with a brain disorder died in the year in which costs were obtained, the costs of the matched comparison cohort member were weighed to reflect the same length in days as that of the person with a brain disorder.

Finally, the attributable direct costs were computed by modeling the average annual cost per person for persons with a brain disorder using an ordinary least squares regression. All Danish residents were included, and the explanatory variables were age, sex, and the different brain disorders. Thus, the cost of each brain disorder was adjusted for comorbid brain disorders.

#### Sensitivity analyses

All analyses were repeated using three definitions of brain disorders. In the first sensitivity analysis, we included only brain disorders according to diagnoses in the DNPR (i.e., not identifying brain disorders by redeemed prescriptions). This definition was identical to that in the main analysis of a previously published study on brain disorders.^4^ In the second sensitivity analysis, we used the definition applied in the sensitivity analysis of the prior study.^4^ Thus, the first two sensitivity analyses enabled comparison with the previous study. In the third sensitivity analysis, we defined any brain disorders according to diagnoses in the DNPR; redeemed prescriptions in the NPR; and at least two contacts with neurologists, psychiatrists, or psychologists in private practice outside hospitals as recorded in the Danish Health Service Registry. This analysis was performed only for any brain disorder, because the diagnoses given by private practicing specialists outside hospitals are not recorded in any registry. Through this sensitivity analysis, we aimed to also include persons with brain disorder symptoms alone. Codes for the various definitions of brain disorders are available in Appendix 1.

Analyses were conducted using SAS V. 9.4, and visualizations were made in R version 4.3.3. The study was reported to the Danish Data Protection Agency by Aarhus University (record number 2016-051-00001, serial number 603). According to Danish law, no further approvals were required.

## Results

### Patient characteristics

For the 2015 prevalent cohort for any brain disorder, 44% were male, the median age was 51 years, and 73% had CCI = 0 (Appendix 3). For the 2021 prevalent cohort for any brain disorder, 44% were male, the median age was 52 years, and 72% had a CCI score = 0 (Figure 1 and Appendix 3). For the 2011–2015 incident cohort for any brain disorder, 48% were male, the median age was 44 years, and 80% had CCI = 0. For the 2016–2021 incident cohort for any brain disorder, 49% were male, the median age was 42 years, and 80% had CCI = 0 (Appendix 3). For both the prevalent and incident cohorts, the distribution of sex, age, and comorbidity burden varied by disease. For the 2016–2021 incident cohorts, the proportion of males was lowest for eating disorders and highest for drug abuse. The median age at diagnosis was lowest for cerebral palsy (13 years) and highest for dementia (81 years). The lowest proportion of persons with CCI = 0 was found for polyneuropathy (39%), whereas persons with eating disorders had the highest proportion (93%). Throughout 2015-2021, having concurrent anxiety disorders, depression, headache, sleep disorders, and stress-related disorders were common for patients with brain disorders (heat maps in Appendix 3). Patient characteristics by time period for both the prevalent and incident cohorts are provided in Appendix 3.

**Figure 1.**
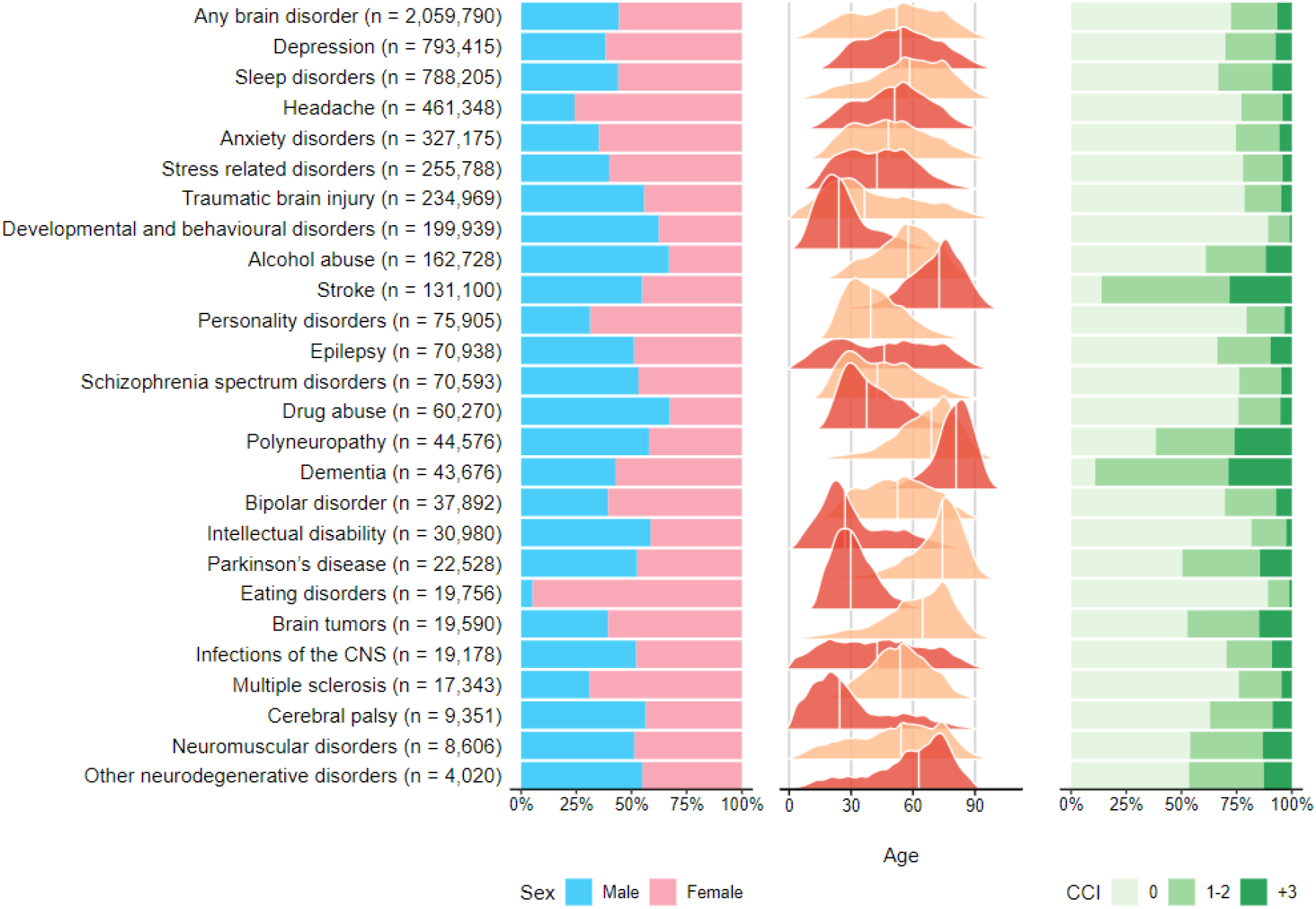
Patient characteristics of the 2021 prevalent cohort.

### Occurrence

The prevalence of any brain disorder in Denmark in 2015 was 33.2% (1,893,318 persons). This prevalence gradually increased to 35.2% (2,059,852 persons) in 2021 (Appendix 4). Throughout 2015—2021, the three most prevalent conditions were depression (13.5% in 2021), sleep disorders (13.5% in 2021), and headache (7.9% in 2021). The prevalence increased for most brain disorders, particularly for developmental and behavioral disorders (Figure 2). For cerebral palsy, dementia, CNS infections, and personality disorders, the prevalence remained stable, whereas the prevalence decreased for alcohol abuse, epilepsy, Parkinson’s disease, and traumatic brain injury.

**Figure 2.**
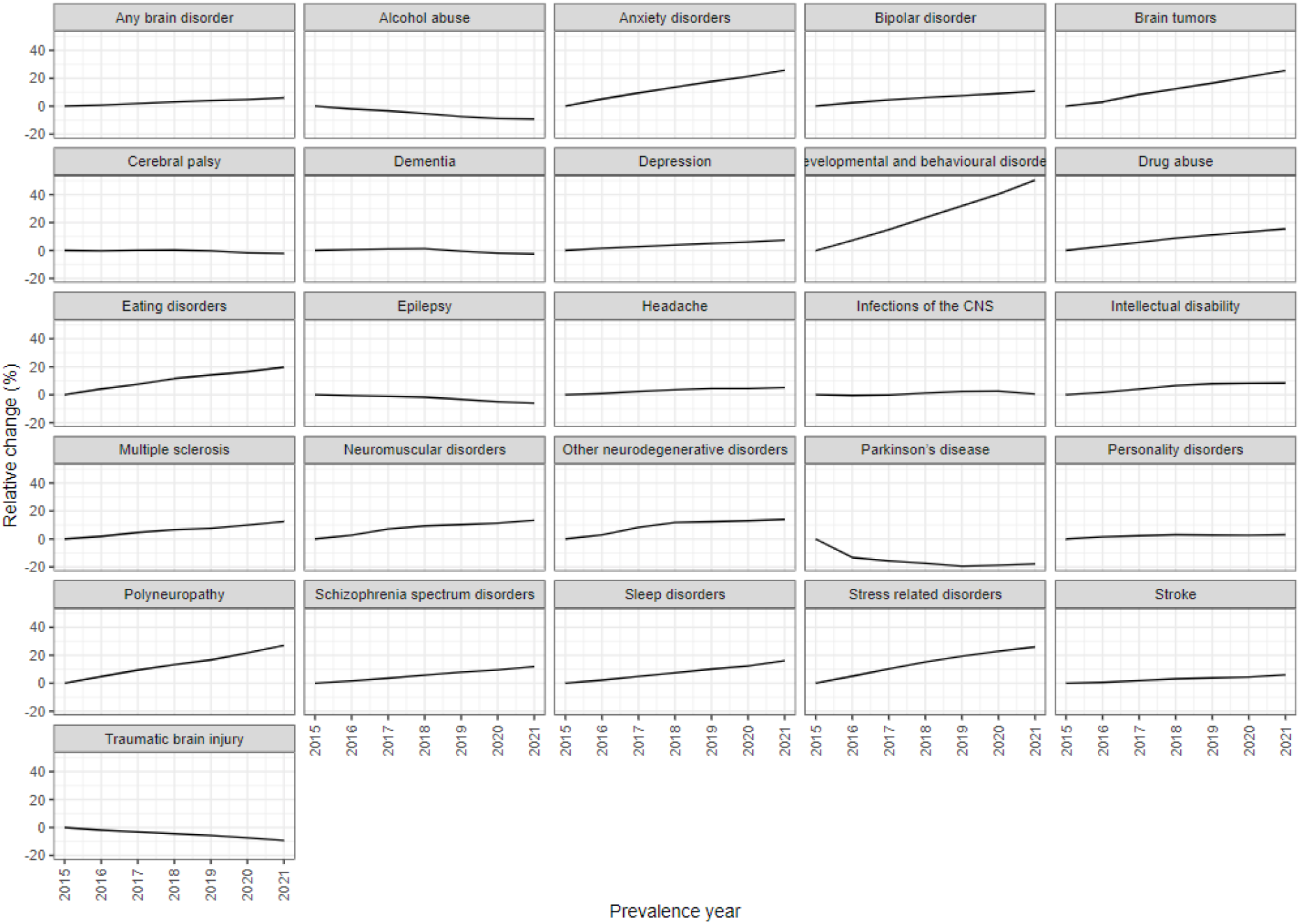
Relative changes in the prevalence of brain disorders in 2015–2021.

The incidence rate of any brain disorder decreased from 1,792 per 100,000 PYs in 2011–2015 to 1,634 per 100,000 PYs in 2016–2021 (Appendix 5). In both periods, the incidence rates were highest for sleep disorders (914 per 100,000 PYs in 2016–2021), depression (585 per 100,000 PYs in 2016–2021), and anxiety disorders (344 per 100,000 PYs in 2016–2021). The incidence rates for many conditions remained stable or decreased slightly between these time periods. However, the rates of developmental and behavioral disorders and sleep disorders increased between these periods.

### Mortality

In the incident cohorts for any brain disorder, 5% died within the first year after a diagnosis/redeemed prescription. The adjusted HR of one-year mortality after any incident brain disorder, with respect to the comparison cohort, was 5.5 (95% confidence interval [CI]: 5.4; 5.6) for 2011–2015 and 5.3 (95% CI: 5.2; 5.3) for 2016–2021 (Figure 3, Appendix 6). The HRs varied considerably by disorder (for 2016–2021, between 1.5 [95% CI: 1.4; 1.6] for polyneuropathy and 13.9 [95% CI: 12.3; 15.7] for other neurodegenerative disorders). The HRs for the individual disease groups were lower in 2016–2021 than in 2011–2015 for brain tumors, dementia, developmental and behavioral disorders, headache, neuromuscular disorders, stroke, and particularly multiple sclerosis. However, for depression, drug abuse, epilepsy, other neurodegenerative disorders, schizophrenia spectrum disorders, and sleep disorders, the HRs were higher in 2016–2021.

**Figure 3.**
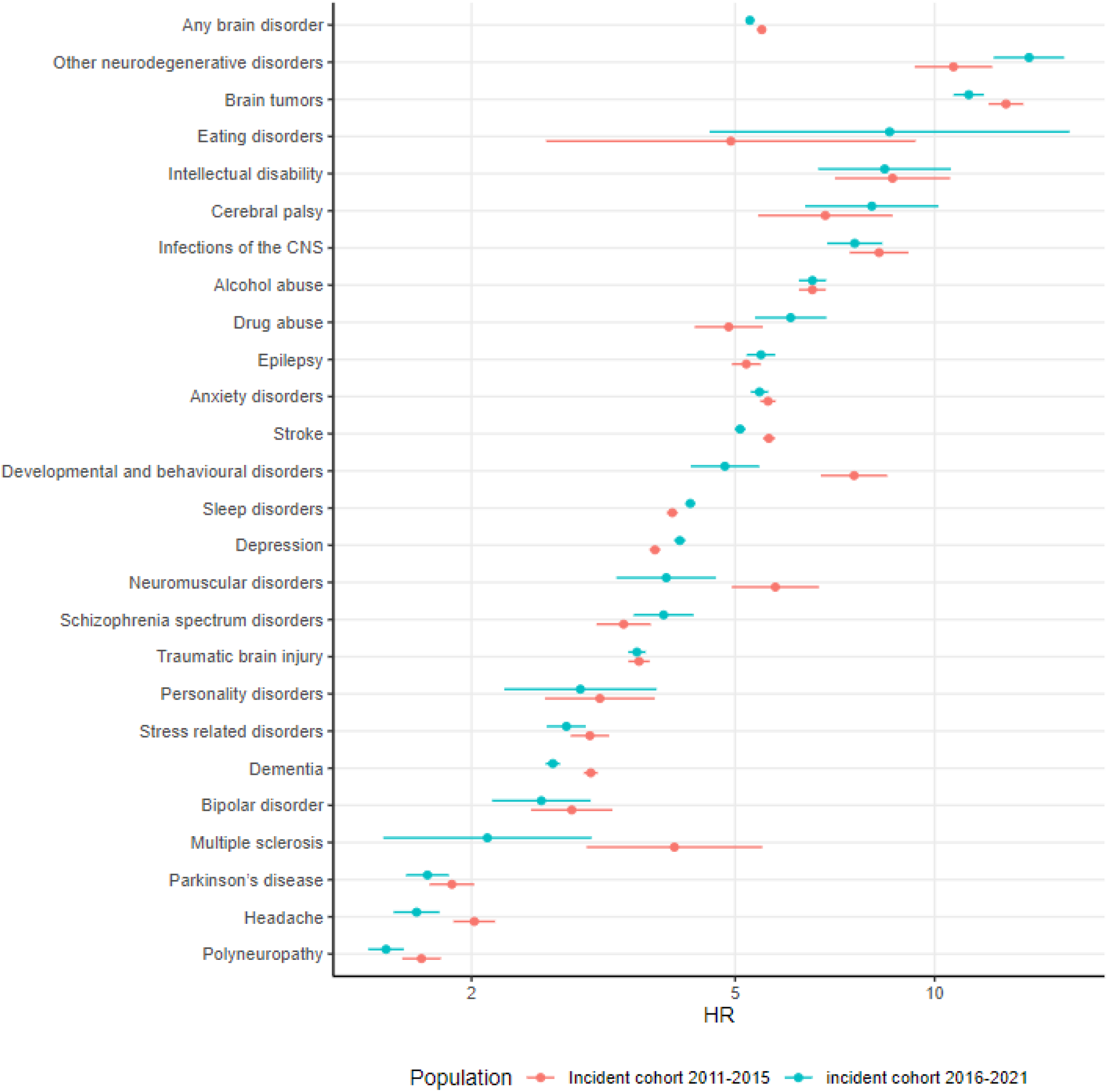
Adjusted one-year mortality hazard ratios (HRs) for the 2011–2015 and 2016–2021 incident cohorts by brain disorder.

### Costs

Overall, the total attributable direct costs for any prevalent brain disorder were estimated at €7.5 billion in both 2015 and 2021 (Figure 4, Appendix 7). The attributable direct costs per person for the prevalent cohorts decreased from €4,022 in 2015 to €3,680 in 2021, largely because of decreased costs of hospital care. In 2018, both total and per-person attributable direct costs decreased followed by an increase in 2019. Indirect costs due to lost productivity increased gradually, from €17.7 billion in 2015 to €23.2 billion in 2021 in total, and from €14,315 per person in 2015 to €17,587 per person in 2021 (Figure 4, Appendix 7). Both direct and indirect costs varied considerably among brain disorders (Figure 5). In 2021, the total attributable direct costs were highest for persons with depression (∼€3.5 billion) and lowest for neuromuscular disorders (∼€53.5 million), whereas the total attributable direct cost per person was highest for dementia (∼€33,600) and lowest for headache (∼€1,000) (Appendix 7). Total attributable indirect costs were also highest for depression (∼€14 billion in 2021) and lowest for other neurodegenerative disorders (∼€60 million), whereas the indirect costs per person were highest for dementia (∼€47,000) and lowest for headache (∼€5,000). Overall, the trends in direct costs varied by disease. However, for most conditions, the total direct costs per person were lower in 2021 than 2015 (except for cerebral palsy, CNS infections, Parkinson’s disease, and traumatic brain injury for which the costs were higher, and alcohol abuse and multiple sclerosis for which the costs were similar). The indirect costs per person were higher in 2021 than 2015 for all disease groups. The total actual direct costs and costs for incident cohorts are reported in Appendix 7.

**Figure 4.**
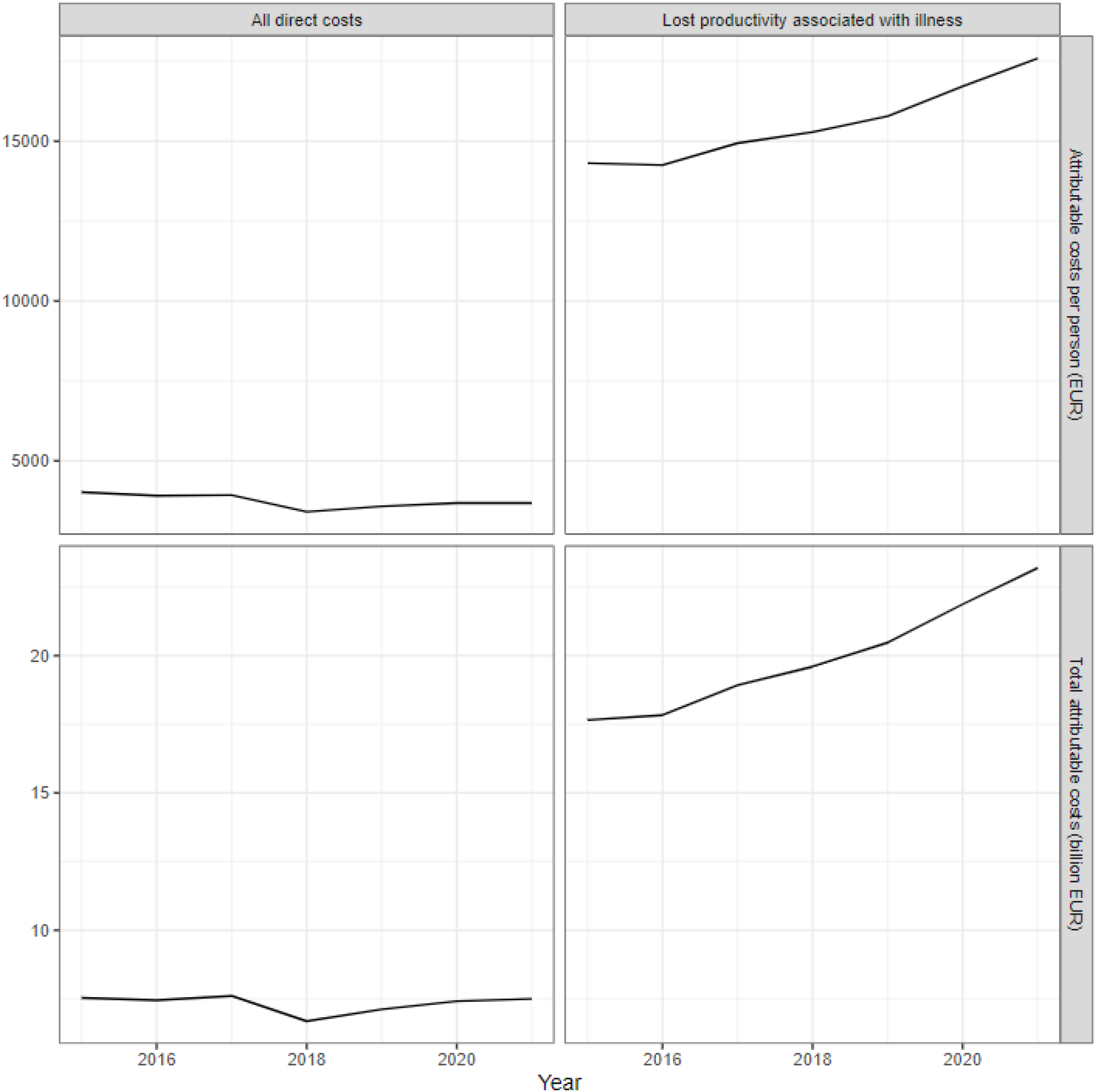
Attributable direct and indirect costs, per person and in total, for persons with brain disorders, 2015–2021.

**Figure 5.**
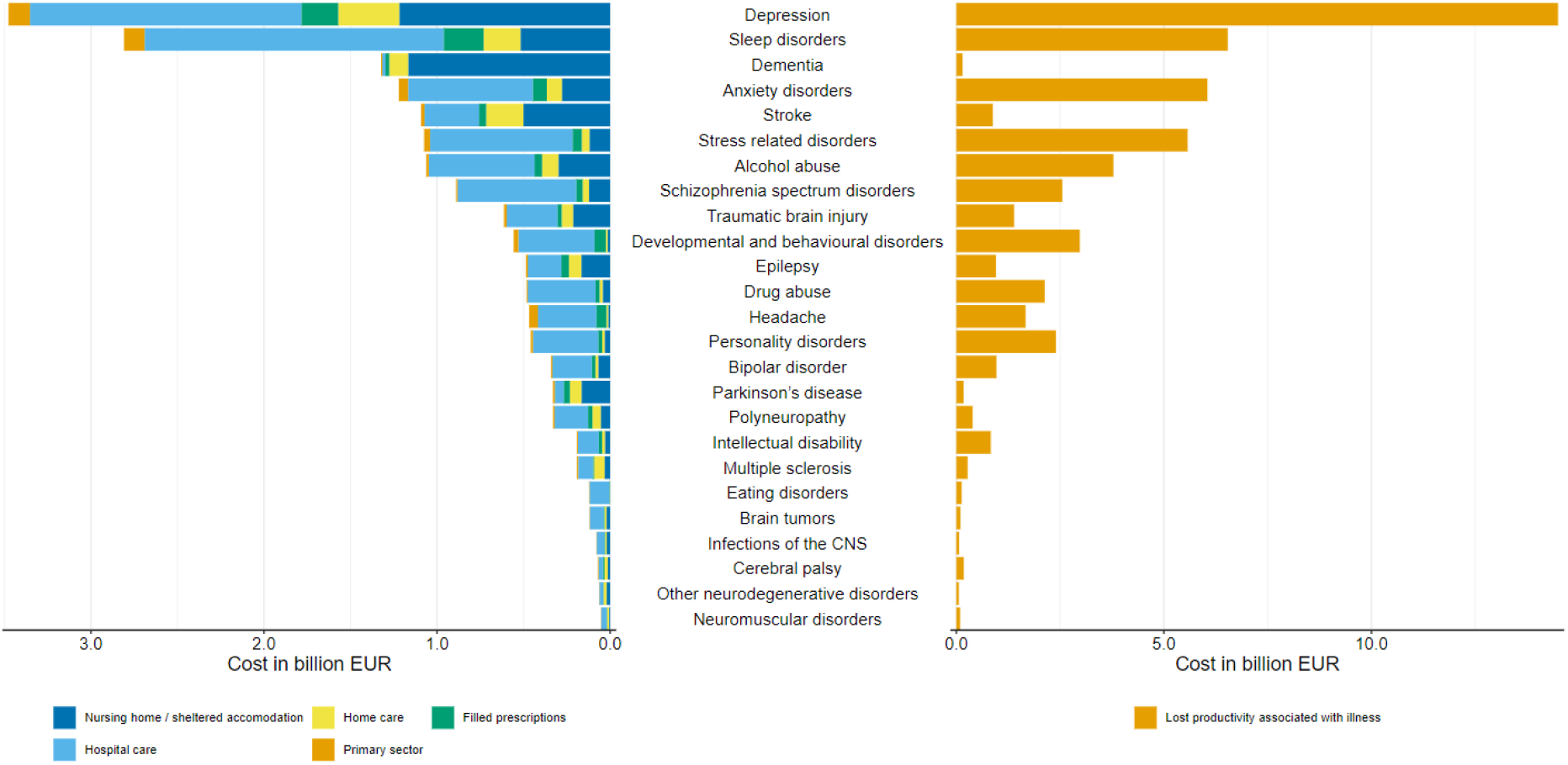
Total attributable direct costs (left) and indirect costs (right) for 2021 according to brain disorder. Disorders are sorted according to direct costs.

After accounting for the costs of comorbid brain disorders through an ordinary least squares regression, we observed the highest direct attributable costs per person for dementia throughout 2015–2021 (∼€21K in 2021) (Appendix 8).

### Sensitivity analyses

In the first sensitivity analysis, in which brain disorders were identified using hospital diagnostic codes, both the prevalence and incidence of any brain disorder were consistently lower than those in the main analysis in all periods (Appendix 9). The prevalence of any brain disorder was 21.4% in 2021, and the incidence for 2016–2021 was 1,331 per 100,000 PYs. The difference between this analysis and the main analysis was driven primarily by alcohol abuse, anxiety disorders, depression, developmental and behavioral disorders, headache, and sleep disorders. The adjusted HR of mortality for any brain disorder was similar to the one in the main analysis for 2011–2015 (HR: 5.5, 95% CI: 5.4; 5.6), but lower for 2016–2021 (HR 4.9, 95% CI: 4.8; 4.9). Both direct and indirect attributable costs for any brain disorder were lower in total but higher per person than observed in the main analysis for the prevalent cohorts. The temporal trends in attributable direct and indirect costs for any brain disorder were similar to those in the main analysis.

In the second sensitivity analysis, using the same definition as that in the sensitivity analyses of the previous Danish study,^4^ i.e., without the modifications, the prevalence was slightly lower and the incidence slightly higher than observed in the main analysis (Appendix 10). For any brain disorder, the HRs for mortality for the incident cohorts were similar to those in the main analysis. The temporal trends in occurrence, mortality, and costs of illness for any brain disorder were similar to those in the main analysis. In the third sensitivity analysis, to identify any brain disorder, we used diagnoses; redeemed prescriptions; and contacts with psychiatrists, psychologists, and neurologists outside hospitals. In that analysis, the prevalence of brain disorders was 41.4% in 2015 and 43.7% in 2021. Thus, as in the main analysis, the prevalence increased somewhat, whereas the incidence decreased from 2015 to 2021 (Appendix 11). Mortality estimates for incident cohorts and temporal trends in costs of illness were similar to those in the main analysis.

## Discussion

In 2021, one-third of the Danish population lived with a brain disorder. Comparison of 2011–2015 to 2016–2021 indicated an overall 9% decrease in incidence of brain disorders in Denmark. However, between 2015 and 2021, the prevalence increased by 6% (or two percentage points). The one-year mortality for persons with brain disorders was fivefold that of persons free of brain disorders and did not show considerable variation between 2011–2015 and 2016–2021. Where the total direct attributable costs for persons with brain disorders were €7.5 billion in both 2015 and 2021, the direct costs per person were lower in 2021 than in 2015, and the indirect costs due to lost productivity, both in total and per person, increased over this period of time.

There are only few population-based studies on the overall epidemiology of brain disorders. Our estimated prevalence of brain disorders in the main analysis was lower than that of the Global Burden of Disease study (∼50%).^3^ However, when we included both diagnoses, prescriptions, and contacts with neurologists, psychiatrists, and psychologists, the prevalence reached 44%. The differences between our estimates and those of the Global Burden of Disease study might be due to geographical variations in disease prevalence, case ascertainment, validity of diagnoses, variation in included diagnoses, and our lack of inclusion of brain disorder diagnoses made solely by GPs in patients without redeemed prescriptions of specific medications. In terms of trends, the global prevalence of neurological disorders was 37.1% in 2015 and 37.6% in 2021, whereas the prevalence of mental disorders was 13.2% in 2015 and 14.4% in 2021 according to the results of the Global Burden of Disease study.^18^ Likewise, we observed an increasing prevalence of almost all included mental health disorders, and a similar trend has been observed in countries such as the Netherlands^19^ and Sweden.^20^ For incidence of any brain disorder, we reported a 9% decrease between 2011—2015 and 2016—2021. This could potentially be partly explained by decreased diagnostic efforts of non-communicable diseases during the COVID-19 pandemic and/or the introduction of a new reporting system in the DNPR in 2019.^21^

To our knowledge, no study has examined the overall temporal trends in mortality for brain disorders. In our study, the one-year mortality did not show considerable variation for any brain disorder. However, the temporal trends varied by disease. For diseases such as stroke and multiple sclerosis, we observed a declining mortality similar to that observed in previous studies in Western countries.^7,22-24^

As far as we are aware, temporal trends in the costs of brain disorders have not previously been reported collectively. However, a recent Danish study has estimated the average costs of mental disorders in 2004–2017.^25^ For any mental disorder, the attributable healthcare costs were €1 billion, while the costs due to income losses amounted to €5 billion (in 2017 prices). As in our study, depression was a major contributor to the total attributable healthcare costs. However, the estimated direct and indirect costs were lower than those of our study.^25^ This difference is likely due to differences in identification of mental disorders and the prices given in values of different years.^25^ When compared with the Danish study estimating costs in 2015, both direct and indirect costs in 2015 were higher in our study, even when accounting for inflation. However, this difference reflects that our study identified a higher number of people with brain disorders. When we used only hospital diagnoses to identify brain disorders, costs were similar to those of the previous Danish study. In the European study from 2010, the average costs of brain disorders were estimated, and the direct healthcare costs per person were highest for brain tumors and, as observed in our study, lowest for headache.^5^ The indirect costs per person were highest for neuromuscular disorders and lowest for eating disorders.^5^ Variations may exist according to country, time period, and case ascertainment, thereby potentially explaining the differences with respect to our results. In terms of indirect costs, we observed an increase in both total and per-person costs, probably reflecting an increase in real wages during the period.^26^

Although our study relied on nationwide population-based individual-level data, some limitations must be considered, including the risk of misclassification of brain disorders. Severe or acute disorders, such as Parkinson’s disease and stroke, were likely to be diagnosed in hospital and therefore recorded in the DNPR. However, other disorders, such as headache, may in some cases be treated exclusively by GPs, and patients who did not redeem a prescription specifically for the disorder (such as triptans) would not have been classified as having a brain disorder in our study.

Consequently, both the incidence and prevalence could have been underestimated. Furthermore, diseases might have been misdiagnosed. However, certain diagnosis codes for brain disorders recorded in the DNPR have been validated, showing relatively high positive predictive values (PPV) for diagnoses such as dementia (PPV ∼86–98%) and multiple sclerosis (PPV 95%).^12^ The validity of these codes often depends on both the diagnostic criteria and access to diagnostic modalities, which may change over time, potentially influencing our results. Treatment options and medication prices may also change over time; thus, in the identification of brain disorders through prescriptions, more cases of brain disorders might have been found in more recent years. Furthermore, changes in incidence might reflect true changes in incidence or potentially healthcare policy changes that increase the extent to which patients are treated at GPs rather than at hospitals. In that case, the number of individuals with brain disorders may be underestimated, as no diagnosis would be recorded in the DNPR. In terms of mortality, vital status in the civil registration system is updated daily, and since registration in the civil registration system is required by law, the accuracy is believed to be high.^11^ Mortality HRs were adjusted for the diseases included in the CCI. However, residual or unmeasured confounding, in particular from other comorbidities, might have remained. In terms of costs of illness, we were able to include the costs of both lost income and primary and secondary healthcare services. However, we were unable to obtain information on municipally-supported rehabilitation, assistance supplies, and transportation costs associated with treatment, and we did not estimate the costs of care provided by relatives or the costs of lost quality of life.

Moreover, DRG/DAGS tariffs were unavailable for recent psychiatric hospital contacts; therefore, we had to rely on imputation. We also identified an unexpected drop in direct costs in 2018, which may be related to changes in the hospital tariff system in that year.^27^ Finally, the indirect costs were assumed to be reflected in the difference in income between the brain disorder populations and the comparison populations. These results should be considered in the context of two opposing effects. First, short-term sick leave does not influence the wages of workers in Denmark and was therefore not observable in the registries used in this study. E.g., individuals with headache have frequent, but usually short periods of sick leave. Therefore, the production loss due to short-term sick leave may have been underestimated, as it was not accounted for in the data. Second, the analysis was performed based on the assumption that the value of wages was equal to the value of production. This simplification has previously been found to overestimate production costs, as the forgone production due to illness can be replaced by production by colleagues or individuals who would otherwise be unemployed. Therefore, the time taken to restore production to pre-disease levels is shorter than the duration of the sick leave. These friction costs depend on sector characteristics, labor market mobility, and the availability of alternative workers, but are usually lower than those determined through the human capital approach applied in our study.^28^

## Conclusions

Brain disorders remain common, and the costs and mortality for persons with these disorders continue to be high. Our findings underscore the persistent societal burden of brain disorders in Denmark. Policies intended to facilitate and ration care will become increasingly important as the prevalence of brain disorders grows and survival in this patient population may improve.

## Data Availability

The individual-level data used in the present study are not publicly available due to data protection rules in Denmark. Researchers who fulfil the requirements set by the data providers (Statistics Denmark and the Danish Health Data Authority) can obtain similar data.

## Notes

### Competing Interest Statement

The authors have declared no competing interest.

### Author Declarations

The study was reported to the Danish Data Protection Agency by Aarhus University (record number 2016-051-00001, serial number 603). According to Danish law, no further ethical approvals are required.

## References

1. Global, regional, and national burden of disorders affecting the nervous system, 1990-2021: a systematic analysis for the Global Burden of Disease Study 2021. Lancet Neurol. 2024;23(4):344–344.

2. Global, regional, and national burden of 12 mental disorders in 204 countries and territories, 1990-2019: a systematic analysis for the Global Burden of Disease Study 2019. Lancet Psychiatry. 2022;9(2):137–137.

3. Lei J, Gillespie K. Projected Global Burden of Brain Disorders Through 2050 (P7-15.001). Neurology. 2024;17_supplement_1(102):3234.

4. Vestergaard SV, Rasmussen TB, Stallknecht S, et al. Occurrence, mortality and cost of brain disorders in Denmark: a population-based cohort study. BMJ Open. 2020;10(11):e037564.

5. Olesen J, Gustavsson A, Svensson M, Wittchen HU, Jönsson B. The economic cost of brain disorders in Europe. Eur J Neurol. 2012;19(1):155–155.

6. Vollset SE, Goren E, Yuan CW, et al. Fertility, mortality, migration, and population scenarios for 195 countries and territories from 2017 to 2100: a forecasting analysis for the Global Burden of Disease Study. Lancet. 2020;396(10258):1285–1285.

7. Skajaa N, Adelborg K, Horváth-Puhó E, et al. Nationwide Trends in Incidence and Mortality of Stroke Among Younger and Older Adults in Denmark. Neurology. 2021;96(13):e1711–e1723.

8. European Brain Council. Rethinking MS in Denmark - A policy brief. https://www.braincouncil.eu/wp-content/uploads/2020/06/RETHINKING_MS_Denmark_EBC_pp_29112019.pdf. Published 2020. Accessed September 9, 2024.

9. Foulkes L, Andrews JL. Are mental health awareness efforts contributing to the rise in reported mental health problems? A call to test the prevalence inflation hypothesis. New Ideas in Psychology. 2023;69:101010.

10. Schmidt M, Schmidt SAJ, Adelborg K, et al. The Danish health care system and epidemiological research: from health care contacts to database records. Clin Epidemiol. 2019;11:563–591.

11. Schmidt M, Pedersen L, Sørensen HT. The Danish Civil Registration System as a tool in epidemiology. Eur J Epidemiol. 2014;29(8):541–541.

12. Schmidt M, Schmidt SA, Sandegaard JL, Ehrenstein V, Pedersen L, Sørensen HT. The Danish National Patient Registry: a review of content, data quality, and research potential. Clin Epidemiol. 2015;7:449–490.

13. Pottegård A, Schmidt SAJ, Wallach-Kildemoes H, Sørensen HT, Hallas J, Schmidt M. Data Resource Profile: The Danish National Prescription Registry. Int J Epidemiol. 2017;46(3):798–798f.

14. Charlson ME, Pompei P, Ales KL, MacKenzie CR. A new method of classifying prognostic comorbidity in longitudinal studies: development and validation. J Chronic Dis. 1987;40(5):373–373.

15. Andersen JS, Olivarius Nde F, Krasnik A. The Danish National Health Service Register. Scand J Public Health. 2011;39(7 Suppl):34–37.

16. The Danish Health Data Authority. Plejehjemsadresser og plejehjemsbeboere. https://sundhedsdatastyrelsen.dk/-/media/sds/filer/find-tal-og-analyser/almen-praksis-og-kommuner/plejehjem/plejehjemsadresser_plejehjemsbeboere.pdf?la=da. Published 2020. Updated July 2020. Accessed 14 August, 2024.

17. Statistics Denmark. Documentation of statistics. https://www.dst.dk/en/Statistik/dokumentation/documentationofstatistics. Accessed 14 August, 2024.

18. Institute for Health Metrics and Evaluation. GBD Results. Institute for Health Metrics and Evaluation. https://vizhub.healthdata.org/gbd-results/. Published 2024. Accessed August 22, 2024.

19. Ten Have M, Tuithof M, van Dorsselaer S, Schouten F, Luik AI, de Graaf R. Prevalence and trends of common mental disorders from 2007-2009 to 2019-2022: results from the Netherlands Mental Health Survey and Incidence Studies (NEMESIS), including comparison of prevalence rates before vs. during the COVID-19 pandemic. World Psychiatry. 2023;22(2):275–275.

20. Forslund T, Kosidou K, Wicks S, Dalman C. Trends in psychiatric diagnoses, medications and psychological therapies in a large Swedish region: a population-based study. BMC Psychiatry. 2020;20(1):328.

21. Sundhedsdatastyrelsen. Datakvalitetsrapport om LPR 2019 - overgangen fra LPR2 til LPR3. Sundhedsdatastyrelsen. https://sundhedsdatastyrelsen.dk/-/media/sds/filer/registre-og-services/nationale-sundhedsregistre/sygdomme-laegemidler-og-behandlinger/landspatientregisteret/lpr-i-fremtiden/datakvalitetsrapport-om-lpr-2019-_-overgangen-fra-lpr2-til-lpr3.pdf?la=da. Published 2020. Updated October 2 2020. Accessed September 5, 2024.

22. Joundi RA, Smith EE, Yu AYX, Rashid M, Fang J, Kapral MK. Temporal Trends in Case Fatality, Discharge Destination, and Admission to Long-term Care After Acute Stroke. Neurology. 2021;96(16):e2037–e2047.

23. Rotstein DL, Chen H, Wilton AS, et al. Temporal trends in multiple sclerosis prevalence and incidence in a large population. Neurology. 2018;90(16):e1435–e1441.

24. Leadbetter R, MacAskill M, Myall DJ, Taylor BV, Joshi P, Mason DF. Multiple sclerosis mortality in New Zealand: a nationwide prospective study. J Neurol Neurosurg Psychiatry. 2023;94(7):511–511.

25. Christensen MK, McGrath JJ, Momen NC, et al. The cost of mental disorders in Denmark: a register-based study. Npj Ment Health Res. 2022;1(1):1.

26. Statistics Denmark. Statistikbanken Standardberegnet lønindex. https://www.statistikbanken.dk/20326. Published 2024. Accessed September 3, 2024.

27. Dansk Cardiologisk Selskab. Diagnoseog procedurekodning og DRG. https://www.cardio.dk/drg-2023. Published 2023. Updated May 12 2023. Accessed September 6, 2024.

28. Drummond M, Schulper MJ, Claxton K, Stoddart GL, Torrance G. Methods for the economic evaluation of health care programmes. 3 edn ed: Oxford University Press; 2005.

