## Appendix 1 for "Temporal trends in the occurrence, mortality, and cost of brain disorders in Denmark"

| *Brain disorder* | *Code type* | *Included codes* | *Excluded subcodes* |
|---|---|---|---|
| Alcohol abuse | ICD-10 | F10 E24.4 G31.2 G62.1 G72.1 I42.6 K29.2 K70 K85.2 K86.0 Q86.0 Z50.2 Z71.4 | F10.0 |
| Alcohol abuse | ATC | V03AA N07BB | |
| Alcohol abuse | Indication | | |
| Anxiety disorders | ICD-10 | F40 F41 F42 | |
| Anxiety disorders | ATC | N06A | |
| Anxiety disorders | Indication | 371 830 163 | |
| Bipolar disorder | ICD-10 | F30 F31 | |
| Bipolar disorder | ATC | N05AN01 | |
| Bipolar disorder | Indication | | |
| Brain tumors | ICD-10 | C70 C71 C72 D32 D33 D42 D43 | |
| Brain tumors | ATC | | |
| Brain tumors | Indication | | |
| Cerebral palsy | ICD-10 | G80 | |
| Cerebral palsy | ATC | | |
| Cerebral palsy | Indication | | |
| Dementia | ICD-10 | B22.0A F00 F01 F02 F03 F10.73 F11.73 F12.73 F13.73 F14.73 F15.73 F16.73 F18.73 F19.73 G30 G31.0B G31.1 G31.8 G31.9 | |
| Dementia | ATC | N06D | |
| Dementia | Indication | | |
| Depression | ICD-10 | F32 F33 | |
| Depression | ATC | N06A | |
| Depression | Indication | 168 270 | |
| Developmental and behavioural disorders | ICD-10 | F84 F9 | F99 |
| Developmental and behavioural disorders | ATC | N06BA02 N06BA04 N06BA09 NA06BA12 C02AC02 | |
| Developmental and behavioural disorders | Indication | 375 376 391 | |
| Drug abuse | ICD-10 | F1 Z72.2 Z86.44 | F10 F11.0 F13.0 F17 F18.0 |
| Drug abuse | ATC | N07BC01 N07BC51 | |
| Drug abuse | Indication | | |
| Eating disorders | ICD-10 | F50.0 F50.1 F50.2 F50.3 | |
| Eating disorders | ATC | | |
| Eating disorders | Indication | | |
| Epilepsy | ICD-10 | G40 | |
| Epilepsy | ATC | | |
| Epilepsy | Indication | | |
| Headache | ICD-10 | G43 G44 | |
| Headache | ATC | N02C | |
| Headache | Indication | | |
| Infections of the CNS | ICD-10 | G0 A17 A32.1 A39.0 A52.1 A52.2 A52.3 A83 A84 A85 A87 A89 B00.3 B00.4 B01.0 B01.1 B02.0 B02.1 B58.2 B45.1 B37.5 | G08 G09 |
| Infections of the CNS | ATC | | |
| Infections of the CNS | Indication | | |

| Brain disorder | Code type | Included codes | Excluded subcodes |
|---|---|---|---|
| Intellectual disability | ICD-10 | F7 Q90 Q99.2 | |
| Intellectual disability | ATC | | |
| Intellectual disability | Indication | | |
| Multiple sclerosis | ICD-10 | G35 | |
| Multiple sclerosis | ATC | | |
| Multiple sclerosis | Indication | | |
| Neuromuscular disorders | ICD-10 | G70 G71 G72 G73 | |
| Neuromuscular disorders | ATC | | |
| Neuromuscular disorders | Indication | | |
| Other neurodegenerative disorders | ICD-10 | G10 G11 G12 G13 G14 | G13.0 G13.9 |
| Other neurodegenerative disorders | ATC | | |
| Other neurodegenerative disorders | Indication | | |
| Parkinson's disease | ICD-10 | G20 G23 | |
| Parkinson's disease | ATC | N04BA N04BB N04BD N04BX | |
| Parkinson's disease | Indication | | |
| Personality disorders | ICD-10 | F60 | |
| Personality disorders | ATC | | |
| Personality disorders | Indication | | |
| Polyneuropathy | ICD-10 | G60 G61 G62 G63 G64 G13.0 | |
| Polyneuropathy | ATC | | |
| Polyneuropathy | Indication | | |
| Schizophrenia spectrum disorders | ICD-10 | F2 | |
| Schizophrenia spectrum disorders | ATC | | |
| Schizophrenia spectrum disorders | Indication | | |
| Sleep disorders | ICD-10 | F51 G47.0 G47.1 G47.3 G47.4 | |
| Sleep disorders | ATC | N05CF01 N05CH01 N06BA07 N06BA14 N07XX04 N07XX11 | |
| Sleep disorders | Indication | | |
| Stress related disorders | ICD-10 | F43 | |
| Stress related disorders | ATC | | |
| Stress related disorders | Indication | | |
| Stroke | ICD-10 | I60 I61 I63 I64 | I60.8 |
| Stroke | ATC | | |
| Stroke | Indication | | |
| Traumatic brain injury | ICD-10 | F07.2 S02.0 S02.1 S02.7 S02.9 S06 | |
| Traumatic brain injury | ATC | | |
| Traumatic brain injury | Indication | | |

| | Code type | Included codes | Excluded subcodes |
|---|---|---|---|
| Myocardial infarction | ICD-8 | 410 | |
| Myocardial infarction | ICD-10 | I21 I22 I23 | |
| Congestive heart failure | ICD-8 | 427.09 427.10 427.11 427.19 428.99 782.49 | |
| Congestive heart failure | ICD-10 | I50 I11.0 I13.0 I13.2 | |
| Peripheral vascular disease | ICD-8 | 440 441 442 443 444 445 | |
| Peripheral vascular disease | ICD-10 | I70 I71 I72 I73 I74 I77 | |
| Cerebrovascular disease | ICD-8 | 430 431 432 433 434 435 436 437 438 | |
| Cerebrovascular disease | ICD-10 | I6 G45 G46 | |
| Dementia | ICD-8 | 290.09 290.1 293.09 | |
| Dementia | ICD-10 | F00 F01 F02 F03 F05.1 G30 | |
| Chronic pulmonary disease | ICD-8 | 490 491 492 493 515 516 517 518 | |
| Chronic pulmonary disease | ICD-10 | J40 J41 J42 J43 J44 J45 J46 J47 J60 J61 J62 J63 J64 J65 J66 J67 J68.4 J70.1 J70.3 J84.1 J92.0 J96.1 J98.2 J98.3 | |
| Connective tissue disease | ICD-8 | 712 716 734 446 135.99 | |
| Connective tissue disease | ICD-10 | M05 M06 M08 M09 M30 M31 M32 M33 M34 M35 M36 D86 | |
| Ulcer disease | ICD-8 | 530.91 530.98 531 532 533 534 | |
| Ulcer disease | ICD-10 | K22.1 K25 K26 K27 K28 | |
| Mild liver disease | ICD-8 | 571 573.01 573.04 | |
| Mild liver disease | ICD-10 | B18 K70.0 K70.1 K70.2 K70.3 K70.9 K71 K73 K74 K76.0 | |
| Diabetes without end-organ damage | ICD-8 | 249.00 249.06 249.07 249.09 250.00 250.06 250.07 250.09 | |
| Diabetes without end-organ damage | ICD-10 | E10.0 E10.1 E10.9 E11.0 E11.1 E11.9 | |
| Hemiplegia | ICD-8 | 344 | |
| Hemiplegia | ICD-10 | G81 G82 | |
| Moderate to severe renal disease | ICD-8 | 403 404 580 581 582 583 584 590.09 593.19 753.1 792 | |
| Moderate to severe renal disease | ICD-10 | I12 I13 N00 N01 N02 N03 N04 N05 N07 N11 N14 N17 N18 N19 Q61 | |
| Diabetes with end-organ damage | ICD-8 | 249.01 249.02 249.03 249.04 249.05 249.08 250.01 250.02 250.03 250.04 250.05 250.08 | |
| Diabetes with end-organ damage | ICD-10 | E10.2 E10.3 E10.4 E10.5 E10.6 E10.7 E10.8 E11.2 E11.3 E11.4 E11.5 E11.6 E11.7 E11.8 | |
| Solid cancer tumour | ICD-8 | 14 15 16 17 18 190 191 192 193 194 | |
| Solid cancer tumour | ICD-10 | C0 C1 C2 C3 C4 C5 C6 C70 C71 C72 C73 C74 C75 | |
| Leukaemia | ICD-8 | 204 205 206 207 | |
| Leukaemia | ICD-10 | C91 C92 C93 C94 C95 | |
| Lymphoma | ICD-8 | 200 201 202 203 275.59 | |
| Lymphoma | ICD-10 | C81 C82 C83 C84 C85 C88 C90 C96 | |
| Moderate to severe liver disease | ICD-8 | 070.00 070.02 070.04 070.06 070.08 573.00 456.0 | |
| Moderate to severe liver disease | ICD-10 | B15.0 B16.0 B16.2 B19.0 K70.4 K72 K76.6 I85 | |
| Metastatic solid tumour | ICD-8 | 195 196 197 198 199 | |
| Metastatic solid tumour | ICD-10 | C76 C77 C78 C79 C80 | |
| AIDS | ICD-8 | 079.83 | |
| AIDS | ICD-10 | B21 B22 B23 B24 | |

| ***Brain disorder*** | ***Code type*** | ***Included codes*** | ***Excluded subcodes*** |
|---|---|---|---|
| Alcohol abuse | ICD-10 | F10 E24.4 G31.2 G62.1 G72.1 I42.6 K29.2 K70 K85.2 K86.0 Q86.0 | F10.0 |
| Anxiety disorders | ICD-10 | F40 F41 F42 | |
| Bipolar disorder | ICD-10 | F30 F31 | |
| Brain tumors | ICD-10 | C70 C71 C72 D32 D33 D42 D43 | |
| Cerebral palsy | ICD-10 | G80 | |
| Dementia | ICD-10 | F00 F01 F02 F03 G30 G31.0B G31.1 G31.8 G31.9 | |
| Depression | ICD-10 | F32 F33 | |
| Developmental and behavioural disorders | ICD-10 | F84 F9 | F99 |
| Drug abuse | ICD-10 | F1 | F10 F17 |
| Eating disorders | ICD-10 | F50.0 F50.1 F50.2 F50.3 | |
| Epilepsy | ICD-10 | G40 | |
| Headache | ICD-10 | G43 G44 | |
| Infections of the CNS | ICD-10 | G0 A17 A32.1 A32.7 A39.0 A52.1 A52.2 A52.3 A69.2 A83 A84 A85 A87 A89 B00.3 B00.4 B01.0 B01.1 B02.0 B02.1 B58.2 B45.1 B37.5 | G08 G09 |
| Intellectual disability | ICD-10 | F7 Q90 Q99.2 | |
| Multiple sclerosis | ICD-10 | G35 | |
| Neuromuscular disorders | ICD-10 | G70 G71 G72 G73 | |
| Other neurodegenerative disorders | ICD-10 | G10 G11 G12.2G G13 G14 | G13.0 G13.9 |
| Parkinson's disease | ICD-10 | G20 G23 | |
| Personality disorders | ICD-10 | F60 | |
| Polyneuropathy | ICD-10 | G60 G61 G62 G63 G64 G13.0 | |
| Schizophrenia spectrum disorders | ICD-10 | F2 | |
| Sleep disorders | ICD-10 | F51 G47.0 G47.1 G47.3 G47.4 | |
| Stress related disorders | ICD-10 | F43 | |
| Stroke | ICD-10 | I60 I61 I63 I64 | I60.8 |
| Traumatic brain injury | ICD-10 | S02.0 S02.1 S02.7 S02.9 S06 | |

| Brain disorder | Code type | Included codes | Excluded subcodes |
|---|---|---|---|
| Alcohol abuse | ICD-10 | F10 E24.4 G31.2 G62.1 G72.1 I42.6 K29.2 K70 K85.2 K86.0 Q86.0 | F10.0 |
| Alcohol abuse | ATC | V03AA N07BB | |
| Alcohol abuse | Indication | | |
| Anxiety disorders | ICD-10 | F40 F41 F42 | |
| Anxiety disorders | ATC | N06A | |
| Anxiety disorders | Indication | 371 830 163 | |
| Bipolar disorder | ICD-10 | F30 F31 | |
| Bipolar disorder | ATC | N05AN01 | |
| Bipolar disorder | Indication | | |
| Brain tumors | ICD-10 | C70 C71 C72 D32 D33 D42 D43 | |
| Brain tumors | ATC | | |
| Brain tumors | Indication | | |
| Cerebral palsy | ICD-10 | G80 | |
| Cerebral palsy | ATC | | |
| Cerebral palsy | Indication | | |
| Dementia | ICD-10 | F00 F01 F02 F03 G30 G31.0B G31.1 G31.8 G31.9 | |
| Dementia | ATC | N06D | |
| Dementia | Indication | 330 331 329 838 | |
| Depression | ICD-10 | F32 F33 | |
| Depression | ATC | N06A | |
| Depression | Indication | 168 270 | |
| Developmental and behavioural disorders | ICD-10 | F84 F9 | F99 |
| Developmental and behavioural disorders | ATC | | |
| Developmental and behavioural disorders | Indication | | |
| Drug abuse | ICD-10 | F1 | F10 F17 |
| Drug abuse | ATC | N07BC01 N07BC51 | |
| Drug abuse | Indication | | |
| Eating disorders | ICD-10 | F50.0 F50.1 F50.2 F50.3 | |
| Eating disorders | ATC | | |
| Eating disorders | Indication | | |
| Epilepsy | ICD-10 | G40 | |
| Epilepsy | ATC | | |
| Epilepsy | Indication | | |
| Headache | ICD-10 | G43 G44 | |
| Headache | ATC | N02C | |
| Headache | Indication | 56 269 153 | |
| Infections of the CNS | ICD-10 | G0 A17 A32.1 A32.7 A39.0 A52.1 A52.2 A52.3 A69.2 A83 A84 A85 A87 A89 B00.3 B00.4 B01.0 B01.1 B02.0 B02.1 B58.2 B45.1 B37.5 | G08 G09 |
| Infections of the CNS | ATC | | |
| Infections of the CNS | Indication | | |
| Intellectual disability | ICD-10 | F7 Q90 Q99.2 | |
| Intellectual disability | ATC | | |

| *Brain disorder* | *Code type* | *Included codes* | *Excluded subcodes* |
|---|---|---|---|
| Intellectual disability | Indication | | |
| Multiple sclerosis | ICD-10 | G35 | |
| Multiple sclerosis | ATC | | |
| Multiple sclerosis | Indication | | |
| Neuromuscular disorders | ICD-10 | G70 G71 G72 G73 | |
| Neuromuscular disorders | ATC | | |
| Neuromuscular disorders | Indication | | |
| Other neurodegenerative disorders | ICD-10 | G10 G11 G12.2G G13 G14 | G13.0 G13.9 |
| Other neurodegenerative disorders | ATC | | |
| Other neurodegenerative disorders | Indication | | |
| Parkinson’s disease | ICD-10 | G20 G23 | |
| Parkinson’s disease | ATC | N04BA N04BB N04BD N04BX | |
| Parkinson’s disease | Indication | | |
| Personality disorders | ICD-10 | F60 | |
| Personality disorders | ATC | | |
| Personality disorders | Indication | | |
| Polyneuropathy | ICD-10 | G60 G61 G62 G63 G64 G13.0 | |
| Polyneuropathy | ATC | | |
| Polyneuropathy | Indication | | |
| Schizophrenia spectrum disorders | ICD-10 | F2 | |
| Schizophrenia spectrum disorders | ATC | | |
| Schizophrenia spectrum disorders | Indication | | |
| Sleep disorders | ICD-10 | F51 G47.0 G47.1 G47.3 G47.4 | |
| Sleep disorders | ATC | N05CF01 N05CH01 | |
| Sleep disorders | Indication | 372 165 166 373 170 | |
| Stress related disorders | ICD-10 | F43 | |
| Stress related disorders | ATC | | |
| Stress related disorders | Indication | | |
| Stroke | ICD-10 | I60 I61 I63 I64 | I60.8 |
| Stroke | ATC | | |
| Stroke | Indication | | |
| Traumatic brain injury | ICD-10 | S02.0 S02.1 S02.7 S02.9 S06 | |
| Traumatic brain injury | ATC | | |
| Traumatic brain injury | Indication | | |

| *Brain disorder* | *Code type* | *Included codes* | *Excluded subcodes* |
|---|---|---|---|
| Any brain disorder | ICD-10 | A17 A32.1 A39.0 A52.1 A52.2 A52.3 A83 A84 A85 A87 A89 B00.3 B00.4 B01.0 B01.1 B02.0 B02.1 B22.0A B37.5 B58.2 B45.1 C70 C71 C72 D32 D33 D42 D43 E24.4 F G I42.6 I60 I61 I63 I64 K29.2 K70 K85.2 K86.0 Q86.0 Q90 Q99.2 S02.0 S02.1 S02.7 S02.9 S06 Z50.2 Z71.4 Z72.2 Z86.44 | I60.8 |
| Any brain disorder | ATC | C02AC02 N02C N04BA N04BB N04BD N04BX N05AN01 N05CF01 N05CH01 N06A N06BA02 N06BA04 N06BA07 N06BA09 N06BA12 N06BA14 N06D N07BB N07BC01 N07BC51 N07XX04 N07XX11 V03AA | |
| Any brain disorder | SSSY | 18 24 26 35 63 | |
