## Appendix 2 for "Temporal trends in the occurrence, mortality, and cost of brain disorders in Denmark"

Appendix 2 – Imputation of missing DRG/DAGs data

We did not have data on DRG/DAGS costs of psychiatric hospital contacts in the period 2019-2022. Instead we imputed these costs using mean imputation using earlier DRG/DAGS cost data. Psychiatric hospital contacts in 2019-2022 were identified using psychiatric department codes. For each contact, we classified the contact as outpatient or inpatient based on length of stay (≥/<12 hours), and according to 2 digit ICD-10 diagnosis (i.e. F30). The DRG/DAGS cost was then imputed to be the mean cost of inpatient/outpatient psychiatric contacts with the same 2-digit ICD-10 diagnosis in the period 2013-2018.
