## Appendix 3 for "Temporal trends in the occurrence, mortality, and cost of brain disorders in Denmark"

| Disease group | Persons, N | Men, N (%)% | Age, median (Q1-Q3) | CCI: 0, N (%) | CCI: 1-2, N (%) | CCI: +3, N (%) |
|---|---|---|---|---|---|---|
| **Any brain disorder** | 507,789 | 245,053 (48.3) | 43.7 (23.0;64.7) | 404,606 (79.7) | 77,796 (15.3) | 25,387 (5.0) |
| **Alcohol abuse** | 45,094 | 30,684 (68.0) | 52.8 (39.0;65.1) | 32,911 (73.0) | 9,060 (20.1) | 3,123 (6.9) |
| **Anxiety disorders** | 117,585 | 44,496 (37.8) | 43.3 (27.1;62.1) | 84,467 (71.8) | 22,349 (19.0) | 10,769 (9.2) |
| **Bipolar disorder** | 11,985 | 4,933 (41.2) | 41.0 (29.2;54.2) | 9,582 (79.9) | 1,933 (16.1) | 470 (3.9) |
| **Brain tumors** | 9,739 | 4,352 (44.7) | 63.9 (50.6;72.8) | 6,034 (62.0) | 2,491 (25.6) | 1,214 (12.5) |
| **Cerebral palsy** | 2,333 | 1,301 (55.8) | 12.8 (2.5;52.7) | 1,443 (61.9) | 650 (27.9) | 240 (10.3) |
| **Dementia** | 45,905 | 19,117 (41.6) | 81.4 (74.7;86.7) | 21,248 (46.3) | 17,319 (37.7) | 7,338 (16.0) |
| **Depression** | 244,331 | 103,920 (42.5) | 48.3 (31.5;68.4) | 170,572 (69.8) | 52,295 (21.4) | 21,464 (8.8) |
| **Developmental and behavioural disorders** | 63,072 | 38,007 (60.3) | 16.4 (9.6;28.2) | 56,073 (88.9) | 6,233 (9.9) | 766 (1.2) |
| **Drug abuse** | 19,069 | 12,655 (66.4) | 26.9 (20.7;42.1) | 15,921 (83.5) | 2,520 (13.2) | 628 (3.3) |
| **Eating disorders** | 6,105 | 328 (5.4) | 19.8 (16.1;25.0) | 5,719 (93.7) | 359 (5.9) | 27 (0.4) |
| **Epilepsy** | 22,978 | 12,161 (52.9) | 52.3 (20.8;70.0) | 12,673 (55.2) | 6,964 (30.3) | 3,341 (14.5) |
| **Headache** | 105,727 | 28,525 (27.0) | 38.3 (25.3;50.0) | 89,760 (84.9) | 13,382 (12.7) | 2,585 (2.4) |
| **Infections of the CNS** | 6,757 | 3,590 (53.1) | 42.7 (22.2;65.0) | 4,641 (68.7) | 1,433 (21.2) | 683 (10.1) |
| **Intellectual disability** | 8,467 | 4,880 (57.6) | 17.3 (9.8;34.8) | 7,137 (84.3) | 1,158 (13.7) | 172 (2.0) |
| **Multiple sclerosis** | 4,124 | 1,342 (32.5) | 41.6 (32.2;51.8) | 3,488 (84.6) | 553 (13.4) | 83 (2.0) |
| **Neuromuscular disorders** | 3,373 | 1,757 (52.1) | 52.4 (33.0;67.3) | 2,087 (61.9) | 912 (27.0) | 374 (11.1) |
| **Other neurodegenerative disorders** | 2,199 | 1,197 (54.4) | 64.7 (51.6;72.6) | 1,388 (63.1) | 606 (27.6) | 205 (9.3) |
| **Parkinson's disease** | 11,429 | 6,438 (56.3) | 72.1 (63.2;79.0) | 6,284 (55.0) | 3,766 (33.0) | 1,379 (12.1) |
| **Personality disorders** | 21,886 | 6,882 (31.4) | 27.3 (21.3;38.7) | 19,225 (87.8) | 2,349 (10.7) | 312 (1.4) |
| **Polyneuropathy** | 19,514 | 11,692 (59.9) | 65.5 (54.1;74.6) | 9,060 (46.4) | 6,864 (35.2) | 3,590 (18.4) |
| **Schizophrenia spectrum disorders** | 19,387 | 10,265 (52.9) | 29.8 (21.1;49.0) | 15,858 (81.8) | 2,787 (14.4) | 742 (3.8) |
| **Sleep disorders** | 243,342 | 116,967 (48.1) | 51.7 (35.7;67.3) | 168,055 (69.1) | 52,260 (21.5) | 23,027 (9.5) |
| **Stress related disorders** | 82,275 | 34,722 (42.2) | 34.7 (20.6;48.6) | 68,102 (82.8) | 11,606 (14.1) | 2,567 (3.1) |
| **Stroke** | 67,899 | 35,520 (52.3) | 71.8 (61.1;81.3) | 35,646 (52.5) | 22,412 (33.0) | 9,841 (14.5) |
| **Traumatic brain injury** | 65,732 | 35,822 (54.5) | 34.6 (15.9;63.1) | 50,341 (76.6) | 11,360 (17.3) | 4,031 (6.1) |

| *Disease group* | *Persons, N* | *Men, N (%)%* | *Age, median (Q1-Q3)* | *CCI: 0, N (%)* | *CCI: 1-2, N (%)* | *CCI: +3, N (%)* |
|---|---|---|---|---|---|---|
| **Any brain disorder** | 571,268 | 279,475 (48.9) | 42.3 (22.2;65.7) | 456,000 (79.8) | 88,138 (15.4) | 27,130 (4.7) |
| **Alcohol abuse** | 48,772 | 29,018 (59.5) | 52.9 (39.4;65.1) | 35,382 (72.5) | 10,300 (21.1) | 3,090 (6.3) |
| **Anxiety disorders** | 120,219 | 44,171 (36.7) | 37.6 (24.1;57.9) | 92,987 (77.3) | 19,804 (16.5) | 7,428 (6.2) |
| **Bipolar disorder** | 11,764 | 4,758 (40.4) | 38.2 (26.7;53.0) | 9,683 (82.3) | 1,735 (14.7) | 346 (2.9) |
| **Brain tumors** | 13,152 | 5,704 (43.4) | 65.1 (52.0;74.5) | 8,187 (62.2) | 3,516 (26.7) | 1,449 (11.0) |
| **Cerebral palsy** | 2,143 | 1,201 (56.0) | 12.8 (2.1;55.7) | 1,423 (66.4) | 514 (24.0) | 206 (9.6) |
| **Dementia** | 52,791 | 23,349 (44.2) | 80.6 (74.7;85.9) | 25,214 (47.8) | 19,624 (37.2) | 7,953 (15.1) |
| **Depression** | 204,398 | 90,134 (44.1) | 48.2 (28.6;71.4) | 141,819 (69.4) | 44,327 (21.7) | 18,252 (8.9) |
| **Developmental and behavioural disorders** | 91,100 | 51,961 (57.0) | 17.1 (10.6;29.0) | 82,259 (90.3) | 8,179 (9.0) | 662 (0.7) |
| **Drug abuse** | 20,848 | 14,200 (68.1) | 27.1 (21.2;39.5) | 17,681 (84.8) | 2,564 (12.3) | 603 (2.9) |
| **Eating disorders** | 6,916 | 427 (6.2) | 18.8 (15.2;24.3) | 6,427 (92.9) | 469 (6.8) | 20 (0.3) |
| **Epilepsy** | 22,387 | 12,074 (53.9) | 54.9 (19.6;72.5) | 12,340 (55.1) | 6,816 (30.4) | 3,231 (14.4) |
| **Headache** | 118,820 | 33,825 (28.5) | 35.9 (24.3;48.9) | 102,304 (86.1) | 14,218 (12.0) | 2,298 (1.9) |
| **Infections of the CNS** | 7,672 | 4,003 (52.2) | 46.1 (25.4;67.0) | 5,270 (68.7) | 1,645 (21.4) | 757 (9.9) |
| **Intellectual disability** | 7,689 | 4,564 (59.4) | 17.1 (8.7;31.7) | 6,572 (85.5) | 958 (12.5) | 159 (2.1) |
| **Multiple sclerosis** | 4,745 | 1,608 (33.9) | 41.3 (30.9;52.3) | 4,067 (85.7) | 571 (12.0) | 107 (2.3) |
| **Neuromuscular disorders** | 3,324 | 1,695 (51.0) | 54.9 (34.6;70.2) | 2,099 (63.1) | 887 (26.7) | 338 (10.2) |
| **Other neurodegenerative disorders** | 2,683 | 1,446 (53.9) | 67.0 (55.3;74.1) | 1,681 (62.7) | 787 (29.3) | 215 (8.0) |
| **Parkinson's disease** | 13,996 | 8,155 (58.3) | 73.5 (65.6;79.4) | 7,605 (54.3) | 4,713 (33.7) | 1,678 (12.0) |
| **Personality disorders** | 19,657 | 5,284 (26.9) | 26.6 (21.3;36.7) | 17,469 (88.9) | 1,973 (10.0) | 215 (1.1) |
| **Polyneuropathy** | 24,544 | 14,753 (60.1) | 67.7 (56.2;75.8) | 9,652 (39.3) | 9,049 (36.9) | 5,843 (23.8) |
| **Schizophrenia spectrum disorders** | 23,479 | 11,937 (50.8) | 27.0 (20.3;44.6) | 19,841 (84.5) | 2,954 (12.6) | 684 (2.9) |
| **Sleep disorders** | 319,314 | 151,647 (47.5) | 50.2 (30.5;67.1) | 225,758 (70.7) | 67,543 (21.2) | 26,013 (8.1) |
| **Stress related disorders** | 95,899 | 38,924 (40.6) | 32.2 (19.0;48.7) | 80,587 (84.0) | 12,702 (13.2) | 2,610 (2.7) |
| **Stroke** | 84,569 | 46,023 (54.4) | 72.9 (62.5;81.1) | 44,879 (53.1) | 27,405 (32.4) | 12,285 (14.5) |
| **Traumatic brain injury** | 78,568 | 41,483 (52.8) | 40.5 (17.6;70.2) | 57,276 (72.9) | 15,274 (19.4) | 6,018 (7.7) |

| *Disease group* | *Persons, N* | *Men, N (%)%* | *Age, median (Q1-Q3)* | *CCI: 0, N (%)* | *CCI: 1-2, N (%)* | *CCI: +3, N (%)* |
|---|---|---|---|---|---|---|
| **Any brain disorder** | 1,893,227 | 831,353 (43.9) | 50.8 (34.6;65.9) | 1,385,392 (73.2) | 388,765 (20.5) | 119,070 (6.3) |
| **Alcohol abuse** | 174,478 | 119,964 (68.8) | 55.6 (45.8;64.9) | 110,034 (63.1) | 45,397 (26.0) | 19,047 (10.9) |
| **Anxiety disorders** | 253,661 | 89,076 (35.1) | 47.7 (34.7;61.6) | 186,595 (73.6) | 51,474 (20.3) | 15,592 (6.1) |
| **Bipolar disorder** | 33,312 | 13,174 (39.5) | 53.8 (41.1;66.0) | 22,861 (68.6) | 7,910 (23.7) | 2,541 (7.6) |
| **Brain tumors** | 15,207 | 6,212 (40.8) | 62.6 (48.6;71.8) | 8,018 (52.7) | 4,842 (31.8) | 2,347 (15.4) |
| **Cerebral palsy** | 9,307 | 5,220 (56.1) | 22.1 (13.6;36.9) | 5,711 (61.4) | 2,809 (30.2) | 787 (8.5) |
| **Dementia** | 43,644 | 17,491 (40.1) | 81.3 (73.4;87.2) | 3,671 (8.4) | 27,150 (62.2) | 12,823 (29.4) |
| **Depression** | 719,777 | 270,110 (37.5) | 52.0 (39.0;66.2) | 504,256 (70.1) | 161,997 (22.5) | 53,524 (7.4) |
| **Developmental and behavioural disorders** | 129,633 | 84,622 (65.3) | 21.0 (15.1;29.0) | 115,835 (89.4) | 12,682 (9.8) | 1,116 (0.9) |
| **Drug abuse** | 50,848 | 33,336 (65.6) | 38.7 (28.6;50.7) | 37,006 (72.8) | 10,690 (21.0) | 3,152 (6.2) |
| **Eating disorders** | 16,080 | 760 (4.7) | 29.5 (23.0;36.9) | 14,238 (88.5) | 1,657 (10.3) | 185 (1.2) |
| **Epilepsy** | 73,576 | 37,737 (51.3) | 43.6 (24.7;62.1) | 49,052 (66.7) | 17,797 (24.2) | 6,727 (9.1) |
| **Headache** | 427,362 | 100,837 (23.6) | 50.8 (38.7;62.5) | 331,833 (77.6) | 77,997 (18.3) | 17,532 (4.1) |
| **Infections of the CNS** | 18,582 | 9,854 (53.0) | 39.2 (20.9;56.9) | 13,506 (72.7) | 3,647 (19.6) | 1,429 (7.7) |
| **Intellectual disability** | 27,861 | 16,256 (58.3) | 25.3 (17.0;45.0) | 22,449 (80.6) | 4,614 (16.6) | 798 (2.9) |
| **Multiple sclerosis** | 15,023 | 4,692 (31.2) | 52.2 (42.5;62.2) | 11,401 (75.9) | 2,959 (19.7) | 663 (4.4) |
| **Neuromuscular disorders** | 7,390 | 3,809 (51.5) | 51.2 (33.4;66.5) | 4,177 (56.5) | 2,279 (30.8) | 934 (12.6) |
| **Other neurodegenerative disorders** | 3,435 | 1,924 (56.0) | 59.6 (43.3;70.4) | 1,878 (54.7) | 1,142 (33.2) | 415 (12.1) |
| **Parkinson's disease** | 26,701 | 13,170 (49.3) | 71.1 (60.2;79.1) | 14,526 (54.4) | 8,762 (32.8) | 3,413 (12.8) |
| **Personality disorders** | 71,762 | 24,690 (34.4) | 39.9 (30.6;51.0) | 56,496 (78.7) | 12,594 (17.5) | 2,672 (3.7) |
| **Polyneuropathy** | 34,186 | 19,786 (57.9) | 66.5 (54.9;75.6) | 14,600 (42.7) | 12,045 (35.2) | 7,541 (22.1) |
| **Schizophrenia spectrum disorders** | 61,455 | 32,714 (53.2) | 45.1 (32.1;57.7) | 46,016 (74.9) | 12,155 (19.8) | 3,284 (5.3) |
| **Sleep disorders** | 661,517 | 282,011 (42.6) | 58.9 (46.1;70.6) | 439,276 (66.4) | 163,008 (24.6) | 59,233 (9.0) |
| **Stress related disorders** | 197,765 | 78,197 (39.5) | 42.3 (29.3;54.1) | 153,644 (77.7) | 35,710 (18.1) | 8,411 (4.3) |
| **Stroke** | 120,402 | 64,266 (53.4) | 70.9 (61.1;79.9) | 15,635 (13.0) | 71,213 (59.1) | 33,554 (27.9) |
| **Traumatic brain injury** | 252,065 | 144,206 (57.2) | 35.1 (22.2;53.4) | 203,183 (80.6) | 38,652 (15.3) | 10,230 (4.1) |

| *Disease group* | *Persons, N* | *Men, N (%)%* | *Age, median (Q1-Q3)* | *CCI: 0, N (%)* | *CCI: 1-2, N (%)* | *CCI: +3, N (%)* |
|---|---|---|---|---|---|---|
| **Any brain disorder** | 1,925,983 | 847,205 (44.0) | 51.0 (34.5;66.1) | 1,403,506 (72.9) | 398,747 (20.7) | 123,730 (6.4) |
| **Alcohol abuse** | 172,888 | 118,778 (68.7) | 56.0 (46.0;65.4) | 107,697 (62.3) | 45,627 (26.4) | 19,564 (11.3) |
| **Anxiety disorders** | 269,249 | 94,814 (35.2) | 47.8 (34.6;61.7) | 198,067 (73.6) | 54,590 (20.3) | 16,592 (6.2) |
| **Bipolar disorder** | 34,495 | 13,619 (39.5) | 53.4 (40.6;65.8) | 23,704 (68.7) | 8,131 (23.6) | 2,660 (7.7) |
| **Brain tumors** | 15,828 | 6,423 (40.6) | 62.9 (49.1;72.3) | 8,318 (52.6) | 5,076 (32.1) | 2,434 (15.4) |
| **Cerebral palsy** | 9,378 | 5,265 (56.1) | 22.6 (13.9;37.8) | 5,727 (61.1) | 2,834 (30.2) | 817 (8.7) |
| **Dementia** | 44,330 | 17,987 (40.6) | 81.2 (73.5;87.2) | 3,718 (8.4) | 27,212 (61.4) | 13,400 (30.2) |
| **Depression** | 737,885 | 277,862 (37.7) | 52.3 (39.3;66.3) | 515,354 (69.8) | 166,867 (22.6) | 55,664 (7.5) |
| **Developmental and behavioural disorders** | 140,250 | 90,797 (64.7) | 21.4 (15.4;29.5) | 125,211 (89.3) | 13,781 (9.8) | 1,258 (0.9) |
| **Drug abuse** | 52,839 | 34,835 (65.9) | 38.4 (28.5;50.5) | 38,678 (73.2) | 10,951 (20.7) | 3,210 (6.1) |
| **Eating disorders** | 16,915 | 802 (4.7) | 29.5 (23.0;37.1) | 14,997 (88.7) | 1,724 (10.2) | 194 (1.1) |
| **Epilepsy** | 73,759 | 37,821 (51.3) | 44.0 (25.0;62.7) | 48,838 (66.2) | 17,960 (24.3) | 6,961 (9.4) |
| **Headache** | 435,360 | 103,120 (23.7) | 50.8 (38.5;62.5) | 336,755 (77.4) | 80,541 (18.5) | 18,064 (4.1) |
| **Infections of the CNS** | 18,670 | 9,916 (53.1) | 40.0 (21.4;57.8) | 13,372 (71.6) | 3,749 (20.1) | 1,549 (8.3) |
| **Intellectual disability** | 28,590 | 16,703 (58.4) | 25.4 (17.4;44.8) | 23,050 (80.6) | 4,705 (16.5) | 835 (2.9) |
| **Multiple sclerosis** | 15,450 | 4,825 (31.2) | 52.4 (42.6;62.5) | 11,702 (75.7) | 3,024 (19.6) | 724 (4.7) |
| **Neuromuscular disorders** | 7,670 | 3,948 (51.5) | 51.6 (33.8;66.9) | 4,257 (55.5) | 2,412 (31.4) | 1,001 (13.1) |
| **Other neurodegenerative disorders** | 3,571 | 1,995 (55.9) | 60.3 (44.2;70.9) | 1,935 (54.2) | 1,192 (33.4) | 444 (12.4) |
| **Parkinson’s disease** | 23,394 | 11,962 (51.1) | 72.3 (62.1;79.7) | 12,218 (52.2) | 7,968 (34.1) | 3,208 (13.7) |
| **Personality disorders** | 73,544 | 24,890 (33.8) | 39.7 (30.3;50.9) | 57,876 (78.7) | 12,912 (17.6) | 2,756 (3.7) |
| **Polyneuropathy** | 36,181 | 20,960 (57.9) | 66.9 (55.3;75.8) | 15,242 (42.1) | 12,743 (35.2) | 8,196 (22.7) |
| **Schizophrenia spectrum disorders** | 63,105 | 33,668 (53.4) | 44.7 (31.5;57.5) | 47,243 (74.9) | 12,437 (19.7) | 3,425 (5.4) |
| **Sleep disorders** | 683,154 | 293,064 (42.9) | 58.8 (45.8;70.8) | 452,244 (66.2) | 168,848 (24.7) | 62,062 (9.1) |
| **Stress related disorders** | 209,658 | 83,213 (39.7) | 42.4 (29.2;54.2) | 162,453 (77.5) | 38,160 (18.2) | 9,045 (4.3) |
| **Stroke** | 122,356 | 65,602 (53.6) | 71.2 (61.3;79.9) | 16,197 (13.2) | 71,758 (58.6) | 34,401 (28.1) |
| **Traumatic brain injury** | 249,807 | 142,370 (57.0) | 35.4 (22.5;54.1) | 200,195 (80.1) | 38,986 (15.6) | 10,626 (4.3) |

| Disease group | Persons, N | Men, N (%)% | Age, median (Q1-Q3) | CCI: 0, N (%) | CCI: 1-2, N (%) | CCI: +3, N (%) |
|---|---|---|---|---|---|---|
| **Any brain disorder** | 1,955,729 | 861,953 (44.1) | 51.2 (34.4;66.3) | 1,418,602 (72.5) | 408,706 (20.9) | 128,421 (6.6) |
| **Alcohol abuse** | 170,917 | 117,406 (68.7) | 56.5 (46.3;65.9) | 105,260 (61.6) | 45,697 (26.7) | 19,960 (11.7) |
| **Anxiety disorders** | 281,611 | 99,429 (35.3) | 47.8 (34.5;61.8) | 206,886 (73.5) | 57,317 (20.4) | 17,408 (6.2) |
| **Bipolar disorder** | 35,264 | 13,939 (39.5) | 53.1 (40.4;65.6) | 24,150 (68.5) | 8,378 (23.8) | 2,736 (7.8) |
| **Brain tumors** | 16,700 | 6,730 (40.3) | 63.3 (49.7;72.9) | 8,659 (51.9) | 5,379 (32.2) | 2,662 (15.9) |
| **Cerebral palsy** | 9,441 | 5,294 (56.1) | 22.8 (14.1;38.8) | 5,739 (60.8) | 2,837 (30.0) | 865 (9.2) |
| **Dementia** | 44,754 | 18,341 (41.0) | 81.1 (73.7;87.1) | 3,659 (8.2) | 27,408 (61.2) | 13,687 (30.6) |
| **Depression** | 749,360 | 283,098 (37.8) | 52.6 (39.6;66.5) | 521,572 (69.6) | 170,612 (22.8) | 57,176 (7.6) |
| **Developmental and behavioural disorders** | 150,893 | 96,980 (64.3) | 21.9 (15.8;30.1) | 134,471 (89.1) | 15,064 (10.0) | 1,358 (0.9) |
| **Drug abuse** | 54,506 | 36,128 (66.3) | 38.2 (28.5;50.5) | 40,014 (73.4) | 11,255 (20.6) | 3,237 (5.9) |
| **Eating disorders** | 17,512 | 851 (4.9) | 29.6 (23.2;37.4) | 15,513 (88.6) | 1,788 (10.2) | 211 (1.2) |
| **Epilepsy** | 73,679 | 37,793 (51.3) | 44.5 (25.2;63.3) | 48,563 (65.9) | 17,893 (24.3) | 7,223 (9.8) |
| **Headache** | 443,391 | 105,506 (23.8) | 50.9 (38.4;62.5) | 341,542 (77.0) | 83,056 (18.7) | 18,793 (4.2) |
| **Infections of the CNS** | 18,792 | 9,935 (52.9) | 40.7 (22.1;58.8) | 13,323 (70.9) | 3,838 (20.4) | 1,631 (8.7) |
| **Intellectual disability** | 29,377 | 17,153 (58.4) | 25.7 (17.8;44.5) | 23,629 (80.4) | 4,881 (16.6) | 867 (3.0) |
| **Multiple sclerosis** | 15,937 | 4,975 (31.2) | 52.6 (42.7;62.7) | 12,033 (75.5) | 3,143 (19.7) | 761 (4.8) |
| **Neuromuscular disorders** | 8,026 | 4,117 (51.3) | 51.9 (33.9;67.4) | 4,370 (54.4) | 2,585 (32.2) | 1,071 (13.3) |
| **Other neurodegenerative disorders** | 3,772 | 2,102 (55.7) | 61.4 (44.9;71.7) | 2,011 (53.3) | 1,259 (33.4) | 502 (13.3) |
| **Parkinson’s disease** | 22,803 | 11,732 (51.4) | 72.8 (63.0;79.9) | 11,741 (51.5) | 7,840 (34.4) | 3,222 (14.1) |
| **Personality disorders** | 74,392 | 24,887 (33.5) | 39.6 (30.3;50.7) | 58,485 (78.6) | 13,102 (17.6) | 2,805 (3.8) |
| **Polyneuropathy** | 37,911 | 21,891 (57.7) | 67.4 (55.7;76.2) | 15,753 (41.6) | 13,369 (35.3) | 8,789 (23.2) |
| **Schizophrenia spectrum disorders** | 64,554 | 34,474 (53.4) | 44.3 (31.1;57.4) | 48,249 (74.7) | 12,784 (19.8) | 3,521 (5.5) |
| **Sleep disorders** | 703,403 | 303,482 (43.1) | 58.7 (45.5;71.0) | 464,102 (66.0) | 174,756 (24.8) | 64,545 (9.2) |
| **Stress related disorders** | 221,000 | 87,924 (39.8) | 42.4 (29.1;54.4) | 170,769 (77.3) | 40,490 (18.3) | 9,741 (4.4) |
| **Stroke** | 124,371 | 67,040 (53.9) | 71.5 (61.4;79.9) | 16,713 (13.4) | 72,293 (58.1) | 35,365 (28.4) |
| **Traumatic brain injury** | 247,409 | 140,606 (56.8) | 35.6 (22.7;54.8) | 196,884 (79.6) | 39,457 (15.9) | 11,068 (4.5) |

| *Disease group* | *Persons, N* | *Men, N (%)%* | *Age, median (Q1-Q3)* | *CCI: 0, N (%)* | *CCI: 1-2, N (%)* | *CCI: +3, N (%)* |
|---|---|---|---|---|---|---|
| **Any brain disorder** | 1,986,097 | 876,941 (44.2) | 51.5 (34.3;66.5) | 1,435,479 (72.3) | 417,776 (21.0) | 132,842 (6.7) |
| **Alcohol abuse** | 168,137 | 115,437 (68.7) | 56.8 (46.5;66.3) | 102,496 (61.0) | 45,493 (27.1) | 20,148 (12.0) |
| **Anxiety disorders** | 293,381 | 103,806 (35.4) | 47.8 (34.4;61.8) | 215,700 (73.5) | 59,686 (20.3) | 17,995 (6.1) |
| **Bipolar disorder** | 36,012 | 14,192 (39.4) | 53.0 (40.1;65.5) | 24,666 (68.5) | 8,580 (23.8) | 2,766 (7.7) |
| **Brain tumors** | 17,417 | 6,947 (39.9) | 63.8 (50.3;73.4) | 9,006 (51.7) | 5,621 (32.3) | 2,790 (16.0) |
| **Cerebral palsy** | 9,499 | 5,339 (56.2) | 23.2 (14.4;39.2) | 5,757 (60.6) | 2,869 (30.2) | 873 (9.2) |
| **Dementia** | 45,055 | 18,740 (41.6) | 81.0 (73.8;86.9) | 3,780 (8.4) | 27,484 (61.0) | 13,791 (30.6) |
| **Depression** | 761,324 | 288,413 (37.9) | 52.9 (39.8;66.7) | 528,384 (69.4) | 174,165 (22.9) | 58,775 (7.7) |
| **Developmental and behavioural disorders** | 162,917 | 103,956 (63.8) | 22.4 (16.1;30.7) | 144,981 (89.0) | 16,430 (10.1) | 1,506 (0.9) |
| **Drug abuse** | 56,295 | 37,482 (66.6) | 38.0 (28.6;50.4) | 41,556 (73.8) | 11,483 (20.4) | 3,256 (5.8) |
| **Eating disorders** | 18,261 | 913 (5.0) | 29.6 (23.2;37.4) | 16,149 (88.4) | 1,894 (10.4) | 218 (1.2) |
| **Epilepsy** | 73,595 | 37,746 (51.3) | 45.0 (25.5;63.7) | 48,318 (65.7) | 17,953 (24.4) | 7,324 (10.0) |
| **Headache** | 450,575 | 107,701 (23.9) | 51.0 (38.2;62.5) | 345,962 (76.8) | 85,230 (18.9) | 19,383 (4.3) |
| **Infections of the CNS** | 19,136 | 10,071 (52.6) | 41.2 (22.6;59.1) | 13,521 (70.7) | 3,961 (20.7) | 1,654 (8.6) |
| **Intellectual disability** | 30,214 | 17,639 (58.4) | 25.9 (18.3;44.2) | 24,320 (80.5) | 5,021 (16.6) | 873 (2.9) |
| **Multiple sclerosis** | 16,308 | 5,065 (31.1) | 52.9 (43.0;62.9) | 12,300 (75.4) | 3,207 (19.7) | 801 (4.9) |
| **Neuromuscular disorders** | 8,225 | 4,215 (51.2) | 52.5 (34.2;67.6) | 4,476 (54.4) | 2,662 (32.4) | 1,087 (13.2) |
| **Other neurodegenerative disorders** | 3,910 | 2,170 (55.5) | 61.9 (45.3;71.9) | 2,072 (53.0) | 1,312 (33.6) | 526 (13.5) |
| **Parkinson's disease** | 22,474 | 11,666 (51.9) | 73.4 (63.8;80.2) | 11,285 (50.2) | 7,903 (35.2) | 3,286 (14.6) |
| **Personality disorders** | 75,280 | 24,722 (32.8) | 39.6 (30.4;50.7) | 59,107 (78.5) | 13,426 (17.8) | 2,747 (3.6) |
| **Polyneuropathy** | 39,453 | 22,760 (57.7) | 67.7 (56.1;76.5) | 16,171 (41.0) | 13,961 (35.4) | 9,321 (23.6) |
| **Schizophrenia spectrum disorders** | 66,221 | 35,357 (53.4) | 43.9 (30.7;57.2) | 49,523 (74.8) | 13,133 (19.8) | 3,565 (5.4) |
| **Sleep disorders** | 724,193 | 314,109 (43.4) | 58.7 (45.3;71.3) | 476,670 (65.8) | 180,454 (24.9) | 67,069 (9.3) |
| **Stress related disorders** | 231,981 | 92,500 (39.9) | 42.4 (29.0;54.6) | 178,989 (77.2) | 42,682 (18.4) | 10,310 (4.4) |
| **Stroke** | 126,470 | 68,484 (54.2) | 71.8 (61.5;79.9) | 16,883 (13.3) | 73,293 (58.0) | 36,294 (28.7) |
| **Traumatic brain injury** | 245,065 | 138,541 (56.5) | 35.8 (22.9;55.4) | 193,894 (79.1) | 39,691 (16.2) | 11,480 (4.7) |

| *Disease group* | *Persons, N* | *Men, N (%)%* | *Age, median (Q1-Q3)* | *CCI: 0, N (%)* | *CCI: 1-2, N (%)* | *CCI: +3, N (%)* |
|---|---|---|---|---|---|---|
| **Any brain disorder** | 2,012,559 | 890,383 (44.2) | 51.7 (34.1;66.6) | 1,455,311 (72.3) | 423,417 (21.0) | 133,831 (6.6) |
| **Alcohol abuse** | 165,080 | 113,103 (68.5) | 57.2 (46.7;66.7) | 100,223 (60.7) | 45,057 (27.3) | 19,800 (12.0) |
| **Anxiety disorders** | 304,999 | 108,127 (35.5) | 47.9 (34.3;61.9) | 225,089 (73.8) | 61,663 (20.2) | 18,247 (6.0) |
| **Bipolar disorder** | 36,614 | 14,424 (39.4) | 52.9 (39.9;65.4) | 25,235 (68.9) | 8,642 (23.6) | 2,737 (7.5) |
| **Brain tumors** | 18,107 | 7,177 (39.6) | 64.1 (50.7;73.9) | 9,422 (52.0) | 5,807 (32.1) | 2,878 (15.9) |
| **Cerebral palsy** | 9,485 | 5,348 (56.4) | 23.8 (14.8;40.0) | 5,858 (61.8) | 2,785 (29.4) | 842 (8.9) |
| **Dementia** | 44,369 | 18,643 (42.0) | 81.0 (74.0;86.8) | 3,944 (8.9) | 27,036 (60.9) | 13,389 (30.2) |
| **Depression** | 772,153 | 293,537 (38.0) | 53.3 (40.1;66.8) | 536,555 (69.5) | 176,591 (22.9) | 59,007 (7.6) |
| **Developmental and behavioural disorders** | 174,775 | 110,839 (63.4) | 22.9 (16.4;31.4) | 155,700 (89.1) | 17,405 (10.0) | 1,670 (1.0) |
| **Drug abuse** | 57,743 | 38,686 (67.0) | 37.8 (28.7;50.3) | 43,037 (74.5) | 11,459 (19.8) | 3,247 (5.6) |
| **Eating disorders** | 18,751 | 944 (5.0) | 29.7 (23.4;37.5) | 16,624 (88.7) | 1,897 (10.1) | 230 (1.2) |
| **Epilepsy** | 72,675 | 37,199 (51.2) | 45.4 (25.8;64.0) | 47,787 (65.8) | 17,775 (24.5) | 7,113 (9.8) |
| **Headache** | 455,832 | 109,469 (24.0) | 51.1 (38.0;62.6) | 350,116 (76.8) | 86,296 (18.9) | 19,420 (4.3) |
| **Infections of the CNS** | 19,426 | 10,204 (52.5) | 41.5 (23.1;59.3) | 13,761 (70.8) | 3,971 (20.4) | 1,694 (8.7) |
| **Intellectual disability** | 30,696 | 17,941 (58.4) | 26.3 (18.7;44.0) | 24,834 (80.9) | 4,988 (16.2) | 874 (2.8) |
| **Multiple sclerosis** | 16,514 | 5,096 (30.9) | 53.2 (43.3;63.2) | 12,484 (75.6) | 3,217 (19.5) | 813 (4.9) |
| **Neuromuscular disorders** | 8,330 | 4,285 (51.4) | 53.0 (34.6;67.9) | 4,530 (54.4) | 2,708 (32.5) | 1,092 (13.1) |
| **Other neurodegenerative disorders** | 3,943 | 2,177 (55.2) | 62.3 (45.9;72.4) | 2,107 (53.4) | 1,314 (33.3) | 522 (13.2) |
| **Parkinson's disease** | 21,955 | 11,438 (52.1) | 73.8 (64.3;80.5) | 10,923 (49.8) | 7,797 (35.5) | 3,235 (14.7) |
| **Personality disorders** | 75,407 | 24,388 (32.3) | 39.7 (30.6;50.7) | 59,372 (78.7) | 13,341 (17.7) | 2,694 (3.6) |
| **Polyneuropathy** | 40,756 | 23,454 (57.5) | 68.2 (56.6;76.8) | 16,641 (40.8) | 14,414 (35.4) | 9,701 (23.8) |
| **Schizophrenia spectrum disorders** | 67,736 | 36,162 (53.4) | 43.6 (30.5;57.0) | 51,006 (75.3) | 13,199 (19.5) | 3,531 (5.2) |
| **Sleep disorders** | 744,403 | 324,760 (43.6) | 58.5 (45.0;71.4) | 491,388 (66.0) | 184,995 (24.9) | 68,020 (9.1) |
| **Stress related disorders** | 241,254 | 96,318 (39.9) | 42.5 (29.1;54.8) | 186,678 (77.4) | 44,016 (18.2) | 10,560 (4.4) |
| **Stroke** | 127,765 | 69,374 (54.3) | 72.1 (61.7;80.0) | 17,280 (13.5) | 73,939 (57.9) | 36,546 (28.6) |
| **Traumatic brain injury** | 242,569 | 136,531 (56.3) | 36.0 (23.1;56.0) | 191,553 (79.0) | 39,485 (16.3) | 11,531 (4.8) |

| *Disease group* | *Persons, N* | *Men, N (%)%* | *Age, median (Q1-Q3)* | *CCI: 0, N (%)* | *CCI: 1-2, N (%)* | *CCI: +3, N (%)* |
|---|---|---|---|---|---|---|
| **Any brain disorder** | 2,036,578 | 902,486 (44.3) | 51.8 (34.1;66.8) | 1,473,761 (72.4) | 428,143 (21.0) | 134,674 (6.6) |
| **Alcohol abuse** | 163,409 | 110,763 (67.8) | 57.4 (46.8;66.9) | 99,352 (60.8) | 44,585 (27.3) | 19,472 (11.9) |
| **Anxiety disorders** | 315,922 | 112,063 (35.5) | 47.9 (34.2;61.9) | 234,527 (74.2) | 63,111 (20.0) | 18,284 (5.8) |
| **Bipolar disorder** | 37,332 | 14,749 (39.5) | 52.7 (39.6;65.2) | 25,863 (69.3) | 8,765 (23.5) | 2,704 (7.2) |
| **Brain tumors** | 18,925 | 7,487 (39.6) | 64.4 (51.2;74.4) | 9,884 (52.2) | 6,142 (32.5) | 2,899 (15.3) |
| **Cerebral palsy** | 9,401 | 5,294 (56.3) | 24.0 (14.9;40.5) | 5,848 (62.2) | 2,724 (29.0) | 829 (8.8) |
| **Dementia** | 43,971 | 18,679 (42.5) | 81.0 (74.2;86.7) | 4,330 (9.8) | 26,721 (60.8) | 12,920 (29.4) |
| **Depression** | 783,414 | 298,913 (38.2) | 53.6 (40.3;67.0) | 545,787 (69.7) | 178,646 (22.8) | 58,981 (7.5) |
| **Developmental and behavioural disorders** | 186,974 | 117,894 (63.1) | 23.4 (16.8;32.1) | 166,664 (89.1) | 18,511 (9.9) | 1,799 (1.0) |
| **Drug abuse** | 59,165 | 39,805 (67.3) | 37.6 (28.9;50.1) | 44,474 (75.2) | 11,504 (19.4) | 3,187 (5.4) |
| **Eating disorders** | 19,236 | 983 (5.1) | 29.7 (23.5;37.6) | 17,089 (88.8) | 1,907 (9.9) | 240 (1.2) |
| **Epilepsy** | 71,785 | 36,739 (51.2) | 45.7 (25.8;64.5) | 47,253 (65.8) | 17,574 (24.5) | 6,958 (9.7) |
| **Headache** | 458,823 | 110,569 (24.1) | 51.2 (37.8;62.6) | 352,774 (76.9) | 86,648 (18.9) | 19,401 (4.2) |
| **Infections of the CNS** | 19,598 | 10,286 (52.5) | 42.0 (23.7;59.9) | 13,859 (70.7) | 4,018 (20.5) | 1,721 (8.8) |
| **Intellectual disability** | 30,981 | 18,122 (58.5) | 26.7 (19.2;43.8) | 25,170 (81.2) | 4,969 (16.0) | 842 (2.7) |
| **Multiple sclerosis** | 16,959 | 5,265 (31.0) | 53.4 (43.4;63.4) | 12,868 (75.9) | 3,264 (19.2) | 827 (4.9) |
| **Neuromuscular disorders** | 8,454 | 4,350 (51.5) | 53.7 (35.1;68.5) | 4,592 (54.3) | 2,750 (32.5) | 1,112 (13.2) |
| **Other neurodegenerative disorders** | 3,991 | 2,201 (55.1) | 62.4 (46.4;72.9) | 2,107 (52.8) | 1,372 (34.4) | 512 (12.8) |
| **Parkinson's disease** | 22,293 | 11,603 (52.0) | 74.0 (64.4;80.5) | 11,118 (49.9) | 7,876 (35.3) | 3,299 (14.8) |
| **Personality disorders** | 75,678 | 24,089 (31.8) | 39.6 (30.7;50.5) | 59,896 (79.1) | 13,174 (17.4) | 2,608 (3.4) |
| **Polyneuropathy** | 42,756 | 24,640 (57.6) | 68.6 (57.0;77.2) | 16,913 (39.6) | 15,171 (35.5) | 10,672 (25.0) |
| **Schizophrenia spectrum disorders** | 69,161 | 36,887 (53.3) | 43.1 (30.3;56.8) | 52,424 (75.8) | 13,311 (19.2) | 3,426 (5.0) |
| **Sleep disorders** | 763,760 | 334,649 (43.8) | 58.5 (44.7;71.5) | 505,836 (66.2) | 189,120 (24.8) | 68,804 (9.0) |
| **Stress related disorders** | 249,655 | 99,728 (39.9) | 42.5 (29.1;55.1) | 193,863 (77.7) | 45,074 (18.1) | 10,718 (4.3) |
| **Stroke** | 129,226 | 70,494 (54.6) | 72.4 (61.9;80.0) | 17,664 (13.7) | 74,763 (57.9) | 36,799 (28.5) |
| **Traumatic brain injury** | 239,933 | 134,397 (56.0) | 36.2 (23.3;56.7) | 189,139 (78.8) | 39,334 (16.4) | 11,460 (4.8) |

| *Disease group* | *Persons, N* | *Men, N (%)%* | *Age, median (Q1-Q3)* | *CCI: 0, N (%)* | *CCI: 1-2, N (%)* | *CCI: +3, N (%)* |
|---|---|---|---|---|---|---|
| **Any brain disorder** | 2,059,790 | 913,088 (44.3) | 52.0 (34.1;67.0) | 1,491,867 (72.4) | 432,712 (21.0) | 135,211 (6.6) |
| **Alcohol abuse** | 162,728 | 108,788 (66.9) | 57.6 (46.8;67.3) | 99,365 (61.1) | 44,199 (27.2) | 19,164 (11.8) |
| **Anxiety disorders** | 327,175 | 115,786 (35.4) | 48.0 (34.2;62.1) | 244,408 (74.7) | 64,428 (19.7) | 18,339 (5.6) |
| **Bipolar disorder** | 37,892 | 14,935 (39.4) | 52.5 (39.3;64.9) | 26,390 (69.6) | 8,826 (23.3) | 2,676 (7.1) |
| **Brain tumors** | 19,590 | 7,717 (39.4) | 64.7 (51.5;74.9) | 10,308 (52.6) | 6,383 (32.6) | 2,899 (14.8) |
| **Cerebral palsy** | 9,351 | 5,277 (56.4) | 24.4 (15.2;40.9) | 5,887 (63.0) | 2,649 (28.3) | 815 (8.7) |
| **Dementia** | 43,676 | 18,690 (42.8) | 80.8 (74.4;86.5) | 4,768 (10.9) | 26,370 (60.4) | 12,538 (28.7) |
| **Depression** | 793,415 | 303,226 (38.2) | 54.0 (40.5;67.2) | 554,720 (69.9) | 180,126 (22.7) | 58,569 (7.4) |
| **Developmental and behavioural disorders** | 199,939 | 124,966 (62.5) | 24.0 (17.2;32.8) | 178,504 (89.3) | 19,541 (9.8) | 1,894 (0.9) |
| **Drug abuse** | 60,270 | 40,585 (67.3) | 37.6 (29.2;49.9) | 45,668 (75.8) | 11,480 (19.0) | 3,122 (5.2) |
| **Eating disorders** | 19,756 | 1,030 (5.2) | 29.8 (23.6;37.4) | 17,607 (89.1) | 1,929 (9.8) | 220 (1.1) |
| **Epilepsy** | 70,938 | 36,289 (51.2) | 46.0 (26.1;64.8) | 46,863 (66.1) | 17,289 (24.4) | 6,786 (9.6) |
| **Headache** | 461,348 | 111,720 (24.2) | 51.1 (37.5;62.7) | 355,452 (77.0) | 86,555 (18.8) | 19,341 (4.2) |
| **Infections of the CNS** | 19,178 | 10,009 (52.2) | 42.6 (24.1;60.9) | 13,505 (70.4) | 3,949 (20.6) | 1,724 (9.0) |
| **Intellectual disability** | 30,980 | 18,149 (58.6) | 27.1 (19.6;43.5) | 25,316 (81.7) | 4,877 (15.7) | 787 (2.5) |
| **Multiple sclerosis** | 17,343 | 5,373 (31.0) | 53.8 (43.6;63.7) | 13,184 (76.0) | 3,356 (19.4) | 803 (4.6) |
| **Neuromuscular disorders** | 8,606 | 4,409 (51.2) | 54.2 (35.5;69.0) | 4,643 (54.0) | 2,834 (32.9) | 1,129 (13.1) |
| **Other neurodegenerative disorders** | 4,020 | 2,205 (54.9) | 62.8 (46.8;73.2) | 2,142 (53.3) | 1,370 (34.1) | 508 (12.6) |
| **Parkinson's disease** | 22,528 | 11,803 (52.4) | 74.3 (64.5;80.6) | 11,325 (50.3) | 7,963 (35.3) | 3,240 (14.4) |
| **Personality disorders** | 75,905 | 23,686 (31.2) | 39.5 (30.9;50.4) | 60,350 (79.5) | 13,041 (17.2) | 2,514 (3.3) |
| **Polyneuropathy** | 44,576 | 25,804 (57.9) | 68.9 (57.5;77.4) | 17,121 (38.4) | 15,874 (35.6) | 11,581 (26.0) |
| **Schizophrenia spectrum disorders** | 70,593 | 37,517 (53.1) | 42.7 (30.2;56.7) | 53,783 (76.2) | 13,449 (19.1) | 3,361 (4.8) |
| **Sleep disorders** | 788,205 | 346,249 (43.9) | 58.3 (44.2;71.6) | 524,381 (66.5) | 194,377 (24.7) | 69,447 (8.8) |
| **Stress related disorders** | 255,788 | 101,929 (39.8) | 42.6 (29.2;55.5) | 199,245 (77.9) | 45,835 (17.9) | 10,708 (4.2) |
| **Stroke** | 131,100 | 71,736 (54.7) | 72.6 (62.2;80.2) | 17,988 (13.7) | 76,189 (58.1) | 36,923 (28.2) |
| **Traumatic brain injury** | 234,969 | 130,891 (55.7) | 36.4 (23.5;57.4) | 184,708 (78.6) | 38,911 (16.6) | 11,350 (4.8) |

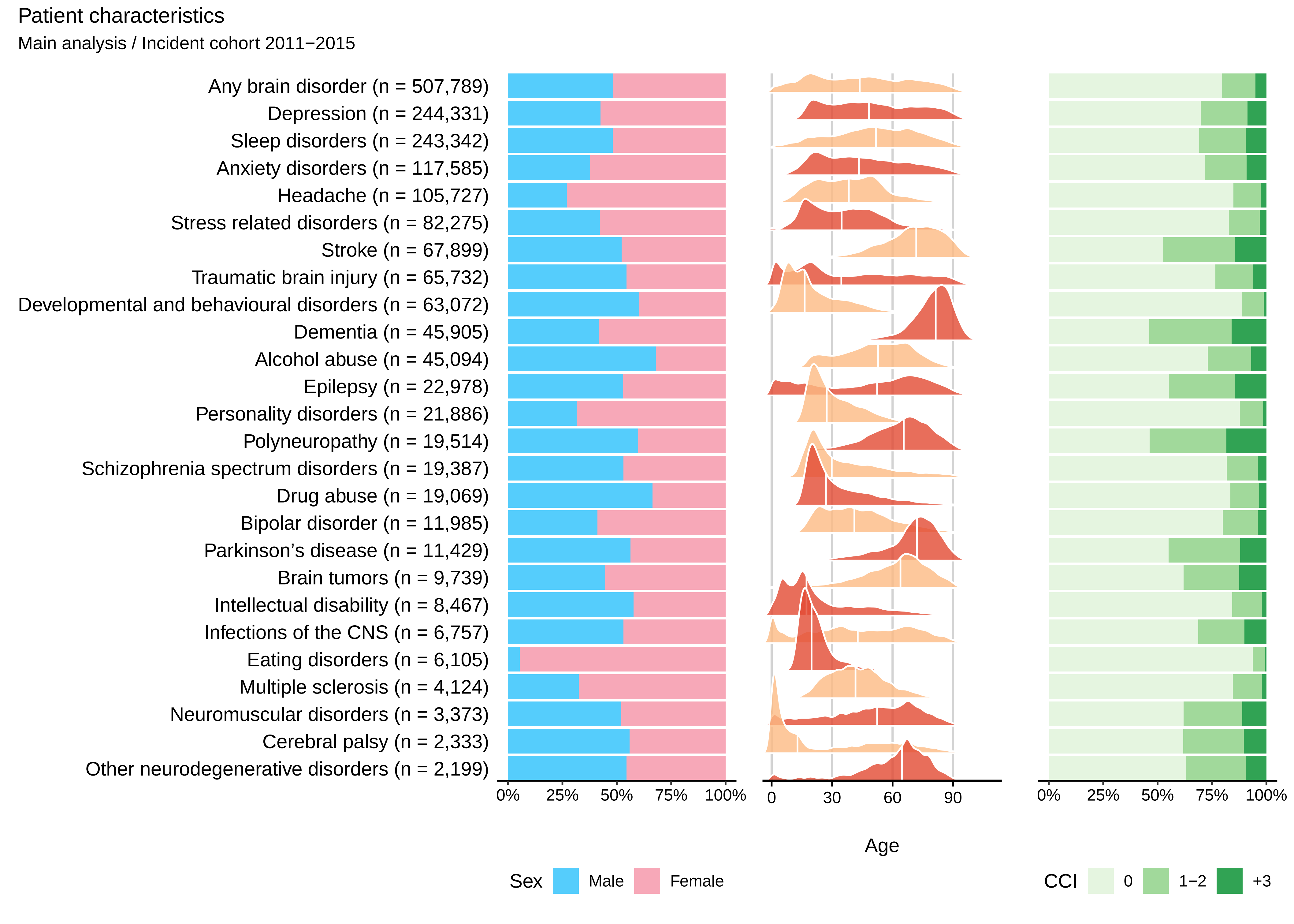
Patient characteristics
Main analysis / Incident cohort 2011−2015
Any brain disorder (n = 507,789)
Depression (n = 244,331)
Sleep disorders (n = 243,342)
Anxiety disorders (n = 117,585)
Headache (n = 105,727)
Stress related disorders (n = 82,275)
Stroke (n = 67,899)
Traumatic brain injury (n = 65,732)
Developmental and behavioural disorders (n = 63,072)
Dementia (n = 45,905)
Alcohol abuse (n = 45,094)
Epilepsy (n = 22,978)
Personality disorders (n = 21,886)
Polyneuropathy (n = 19,514)
Schizophrenia spectrum disorders (n = 19,387)
Drug abuse (n = 19,069)
Bipolar disorder (n = 11,985)
Parkinson’s disease (n = 11,429)
Brain tumors (n = 9,739)
Intellectual disability (n = 8,467)
Infections of the CNS (n = 6,757)
Eating disorders (n = 6,105)
Multiple sclerosis (n = 4,124)
Neuromuscular disorders (n = 3,373)
Cerebral palsy (n = 2,333)
Other neurodegenerative disorders (n = 2,199)
0% 25% 50% 75% 100%
0 30 60 90
Age
0% 25% 50% 75% 100%
Sex Male Female
CCI 0 1−2 +3

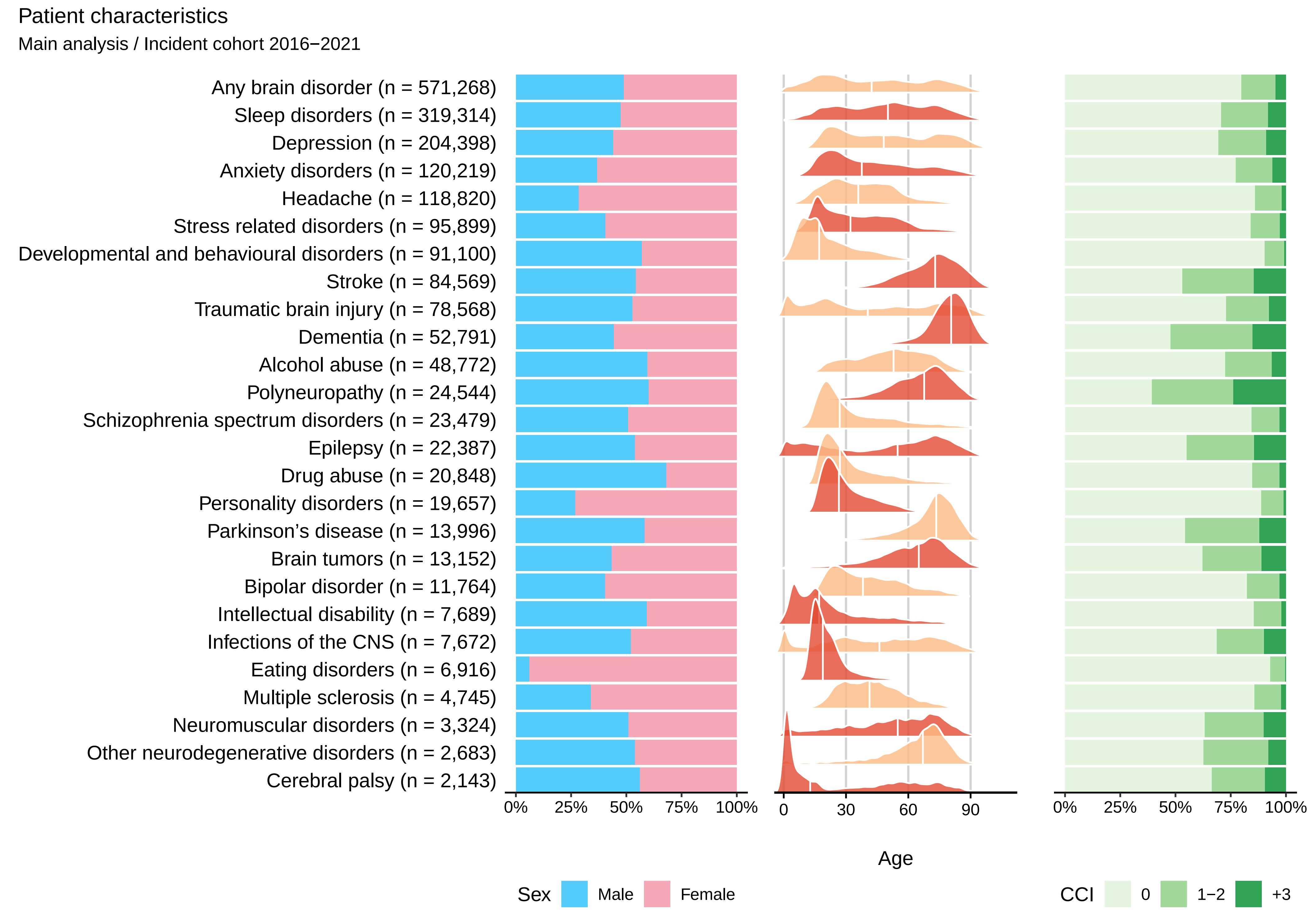

Patient characteristics
Main analysis / Incident cohort 2016−2021
Any brain disorder (n = 571,268)
Sleep disorders (n = 319,314)
Depression (n = 204,398)
Anxiety disorders (n = 120,219)
Headache (n = 118,820)
Stress related disorders (n = 95,899)
Developmental and behavioural disorders (n = 91,100)
Stroke (n = 84,569)
Traumatic brain injury (n = 78,568)
Dementia (n = 52,791)
Alcohol abuse (n = 48,772)
Polyneuropathy (n = 24,544)
Schizophrenia spectrum disorders (n = 23,479)
Epilepsy (n = 22,387)
Drug abuse (n = 20,848)
Personality disorders (n = 19,657)
Parkinson's disease (n = 13,996)
Brain tumors (n = 13,152)
Bipolar disorder (n = 11,764)
Intellectual disability (n = 7,689)
Infections of the CNS (n = 7,672)
Eating disorders (n = 6,916)
Multiple sclerosis (n = 4,745)
Neuromuscular disorders (n = 3,324)
Other neurodegenerative disorders (n = 2,683)
Cerebral palsy (n = 2,143)
0% 25% 50% 75% 100%
0 30 60 90
Age
0% 25% 50% 75% 100%
Sex Male Female
CCI 0 1−2 +3

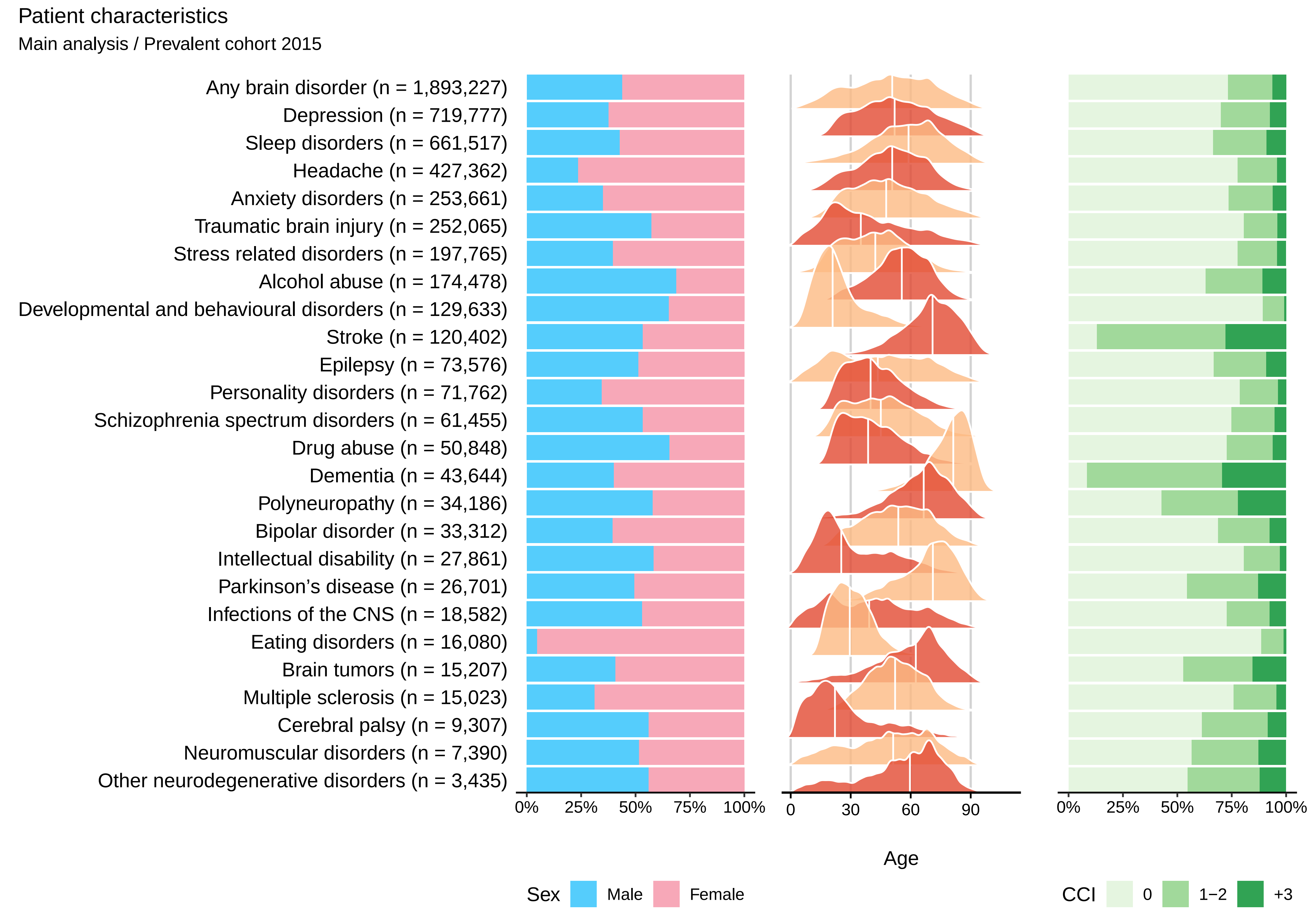

Patient characteristics
Main analysis / Prevalent cohort 2015
Any brain disorder (n = 1,893,227)
Depression (n = 719,777)
Sleep disorders (n = 661,517)
Headache (n = 427,362)
Anxiety disorders (n = 253,661)
Traumatic brain injury (n = 252,065)
Stress related disorders (n = 197,765)
Alcohol abuse (n = 174,478)
Developmental and behavioural disorders (n = 129,633)
Stroke (n = 120,402)
Epilepsy (n = 73,576)
Personality disorders (n = 71,762)
Schizophrenia spectrum disorders (n = 61,455)
Drug abuse (n = 50,848)
Dementia (n = 43,644)
Polyneuropathy (n = 34,186)
Bipolar disorder (n = 33,312)
Intellectual disability (n = 27,861)
Parkinson's disease (n = 26,701)
Infections of the CNS (n = 18,582)
Eating disorders (n = 16,080)
Brain tumors (n = 15,207)
Multiple sclerosis (n = 15,023)
Cerebral palsy (n = 9,307)
Neuromuscular disorders (n = 7,390)
Other neurodegenerative disorders (n = 3,435)
0% 25% 50% 75% 100%
0 30 60 90
Age
0% 25% 50% 75% 100%
Sex Male Female
CCI 0 1−2 +3

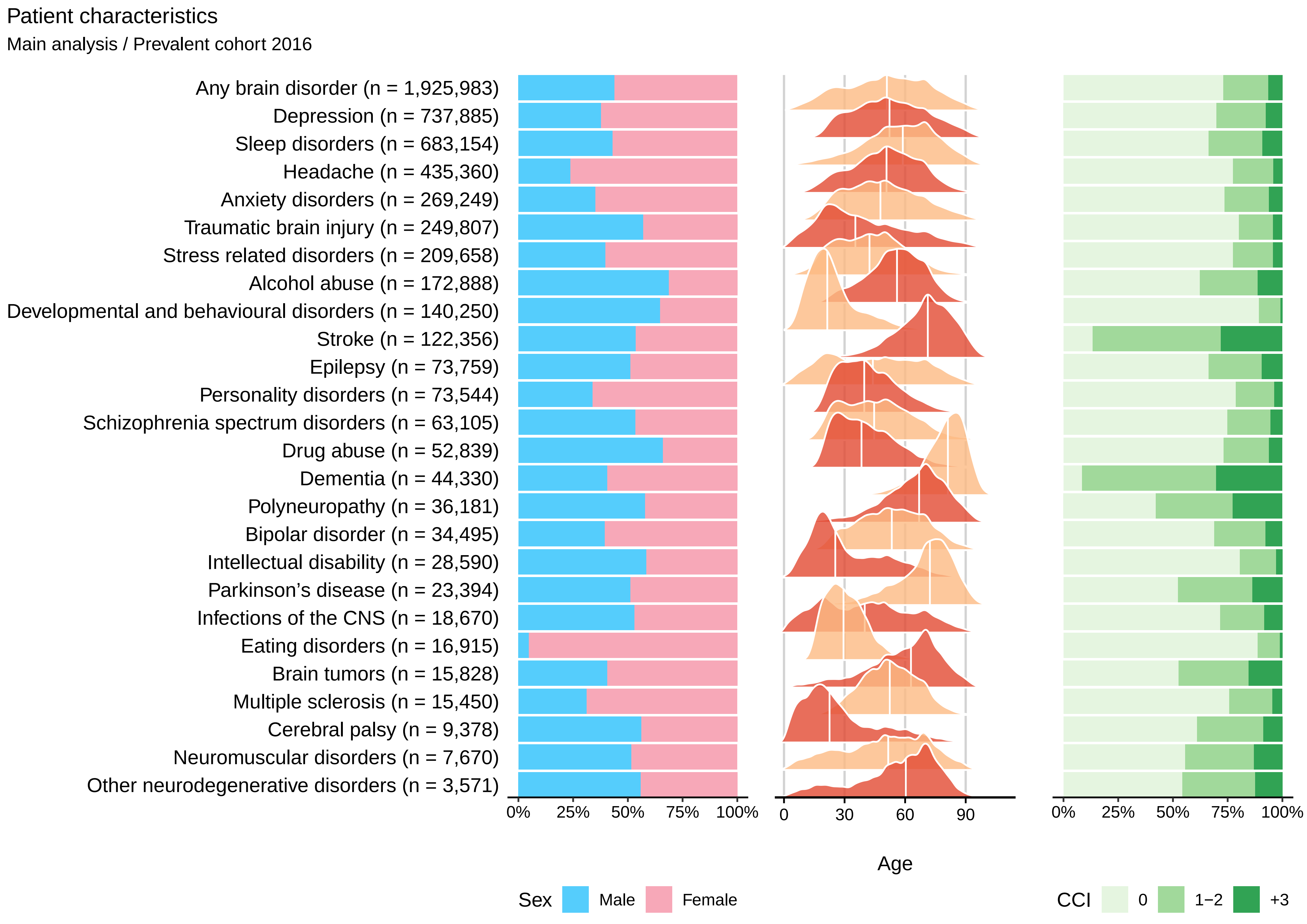

Patient characteristics
Main analysis / Prevalent cohort 2016
Any brain disorder (n = 1,925,983)
Depression (n = 737,885)
Sleep disorders (n = 683,154)
Headache (n = 435,360)
Anxiety disorders (n = 269,249)
Traumatic brain injury (n = 249,807)
Stress related disorders (n = 209,658)
Alcohol abuse (n = 172,888)
Developmental and behavioural disorders (n = 140,250)
Stroke (n = 122,356)
Epilepsy (n = 73,759)
Personality disorders (n = 73,544)
Schizophrenia spectrum disorders (n = 63,105)
Drug abuse (n = 52,839)
Dementia (n = 44,330)
Polyneuropathy (n = 36,181)
Bipolar disorder (n = 34,495)
Intellectual disability (n = 28,590)
Parkinson's disease (n = 23,394)
Infections of the CNS (n = 18,670)
Eating disorders (n = 16,915)
Brain tumors (n = 15,828)
Multiple sclerosis (n = 15,450)
Cerebral palsy (n = 9,378)
Neuromuscular disorders (n = 7,670)
Other neurodegenerative disorders (n = 3,571)
0% 25% 50% 75% 100%
0 30 60 90
Age
0% 25% 50% 75% 100%
Sex Male Female
CCI 0 1−2 +3

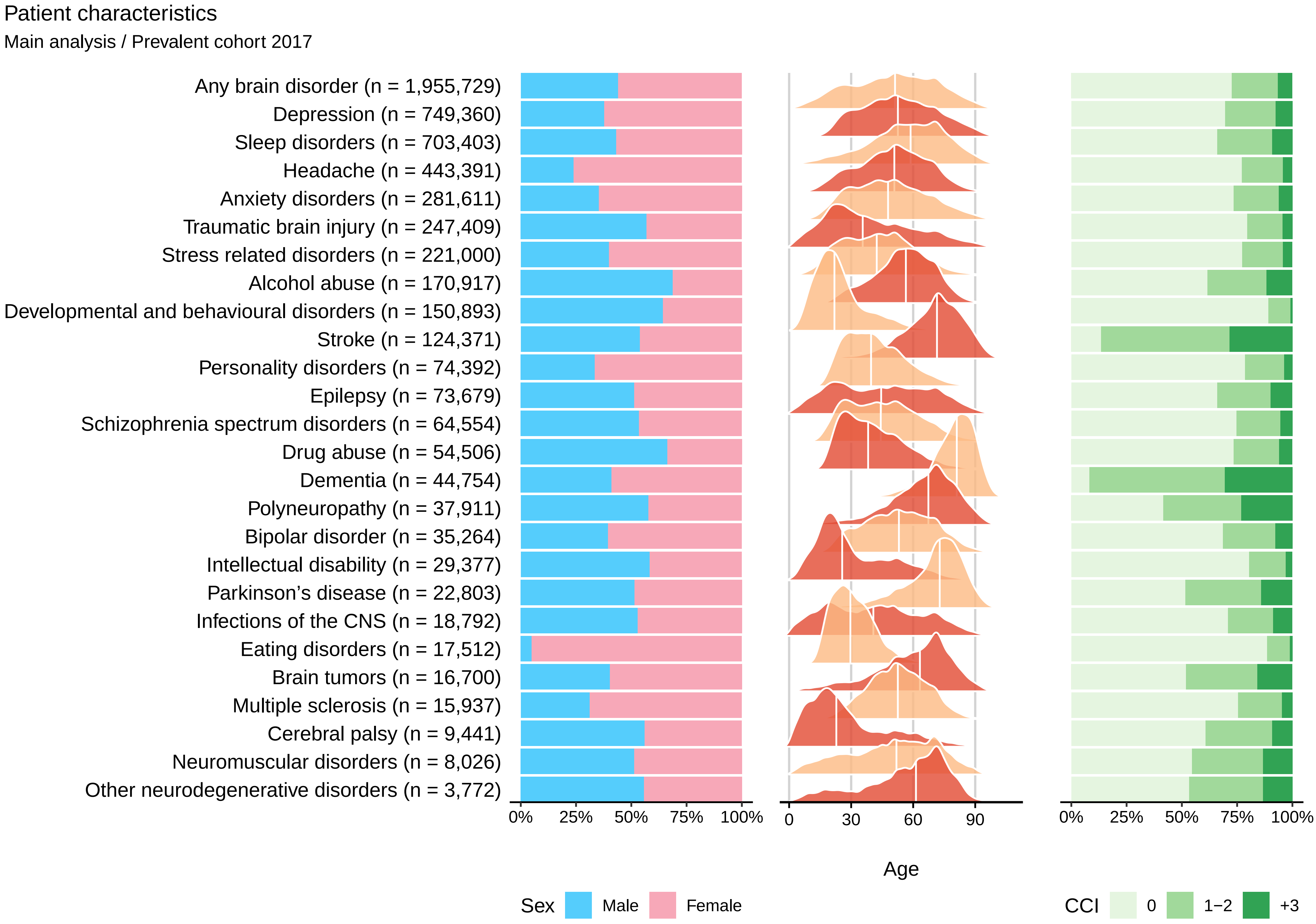
Patient characteristics
Main analysis / Prevalent cohort 2017
Any brain disorder (n = 1,955,729)
Depression (n = 749,360)
Sleep disorders (n = 703,403)
Headache (n = 443,391)
Anxiety disorders (n = 281,611)
Traumatic brain injury (n = 247,409)
Stress related disorders (n = 221,000)
Alcohol abuse (n = 170,917)
Developmental and behavioural disorders (n = 150,893)
Stroke (n = 124,371)
Personality disorders (n = 74,392)
Epilepsy (n = 73,679)
Schizophrenia spectrum disorders (n = 64,554)
Drug abuse (n = 54,506)
Dementia (n = 44,754)
Polyneuropathy (n = 37,911)
Bipolar disorder (n = 35,264)
Intellectual disability (n = 29,377)
Parkinson’s disease (n = 22,803)
Infections of the CNS (n = 18,792)
Eating disorders (n = 17,512)
Brain tumors (n = 16,700)
Multiple sclerosis (n = 15,937)
Cerebral palsy (n = 9,441)
Neuromuscular disorders (n = 8,026)
Other neurodegenerative disorders (n = 3,772)
0% 25% 50% 75% 100%
0 30 60 90
Age
0% 25% 50% 75% 100%
Sex Male Female
CCI 0 1−2 +3

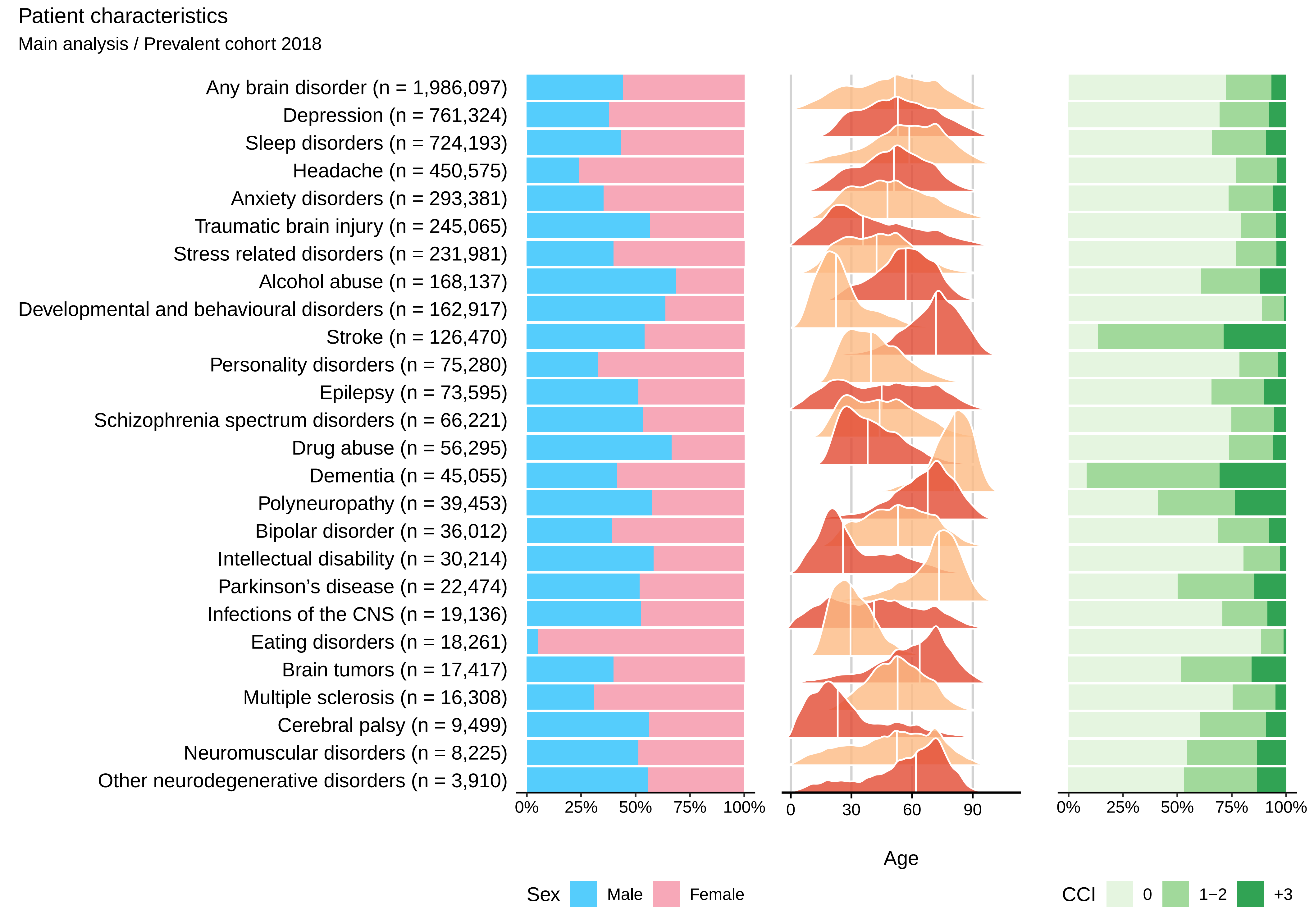
Patient characteristics
Main analysis / Prevalent cohort 2018
Any brain disorder (n = 1,986,097)
Depression (n = 761,324)
Sleep disorders (n = 724,193)
Headache (n = 450,575)
Anxiety disorders (n = 293,381)
Traumatic brain injury (n = 245,065)
Stress related disorders (n = 231,981)
Alcohol abuse (n = 168,137)
Developmental and behavioural disorders (n = 162,917)
Stroke (n = 126,470)
Personality disorders (n = 75,280)
Epilepsy (n = 73,595)
Schizophrenia spectrum disorders (n = 66,221)
Drug abuse (n = 56,295)
Dementia (n = 45,055)
Polyneuropathy (n = 39,453)
Bipolar disorder (n = 36,012)
Intellectual disability (n = 30,214)
Parkinson's disease (n = 22,474)
Infections of the CNS (n = 19,136)
Eating disorders (n = 18,261)
Brain tumors (n = 17,417)
Multiple sclerosis (n = 16,308)
Cerebral palsy (n = 9,499)
Neuromuscular disorders (n = 8,225)
Other neurodegenerative disorders (n = 3,910)
0%
25%
50%
75%
100%
0
30
60
90
Age
0%
25%
50%
75%
100%
Sex
Male
Female
CCI
0
1−2
+3

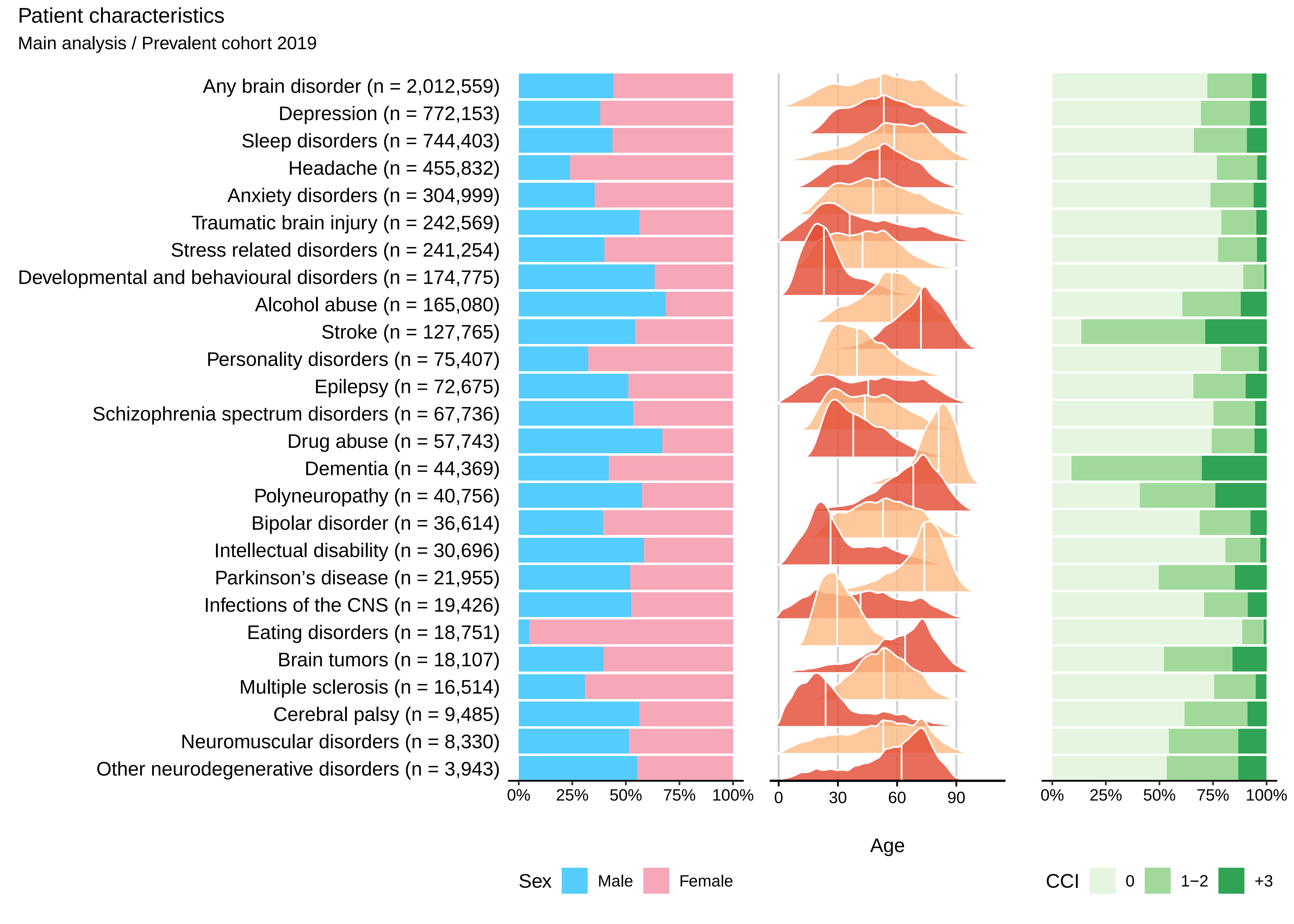
Patient characteristics
Main analysis / Prevalent cohort 2019
Any brain disorder (n = 2,012,559)
Depression (n = 772,153)
Sleep disorders (n = 744,403)
Headache (n = 455,832)
Anxiety disorders (n = 304,999)
Traumatic brain injury (n = 242,569)
Stress related disorders (n = 241,254)
Developmental and behavioural disorders (n = 174,775)
Alcohol abuse (n = 165,080)
Stroke (n = 127,765)
Personality disorders (n = 75,407)
Epilepsy (n = 72,675)
Schizophrenia spectrum disorders (n = 67,736)
Drug abuse (n = 57,743)
Dementia (n = 44,369)
Polyneuropathy (n = 40,756)
Bipolar disorder (n = 36,614)
Intellectual disability (n = 30,696)
Parkinson's disease (n = 21,955)
Infections of the CNS (n = 19,426)
Eating disorders (n = 18,751)
Brain tumors (n = 18,107)
Multiple sclerosis (n = 16,514)
Cerebral palsy (n = 9,485)
Neuromuscular disorders (n = 8,330)
Other neurodegenerative disorders (n = 3,943)
0% 25% 50% 75% 100%
0 30 60 90
Age
0% 25% 50% 75% 100%
Sex Male Female
CCI 0 1−2 +3

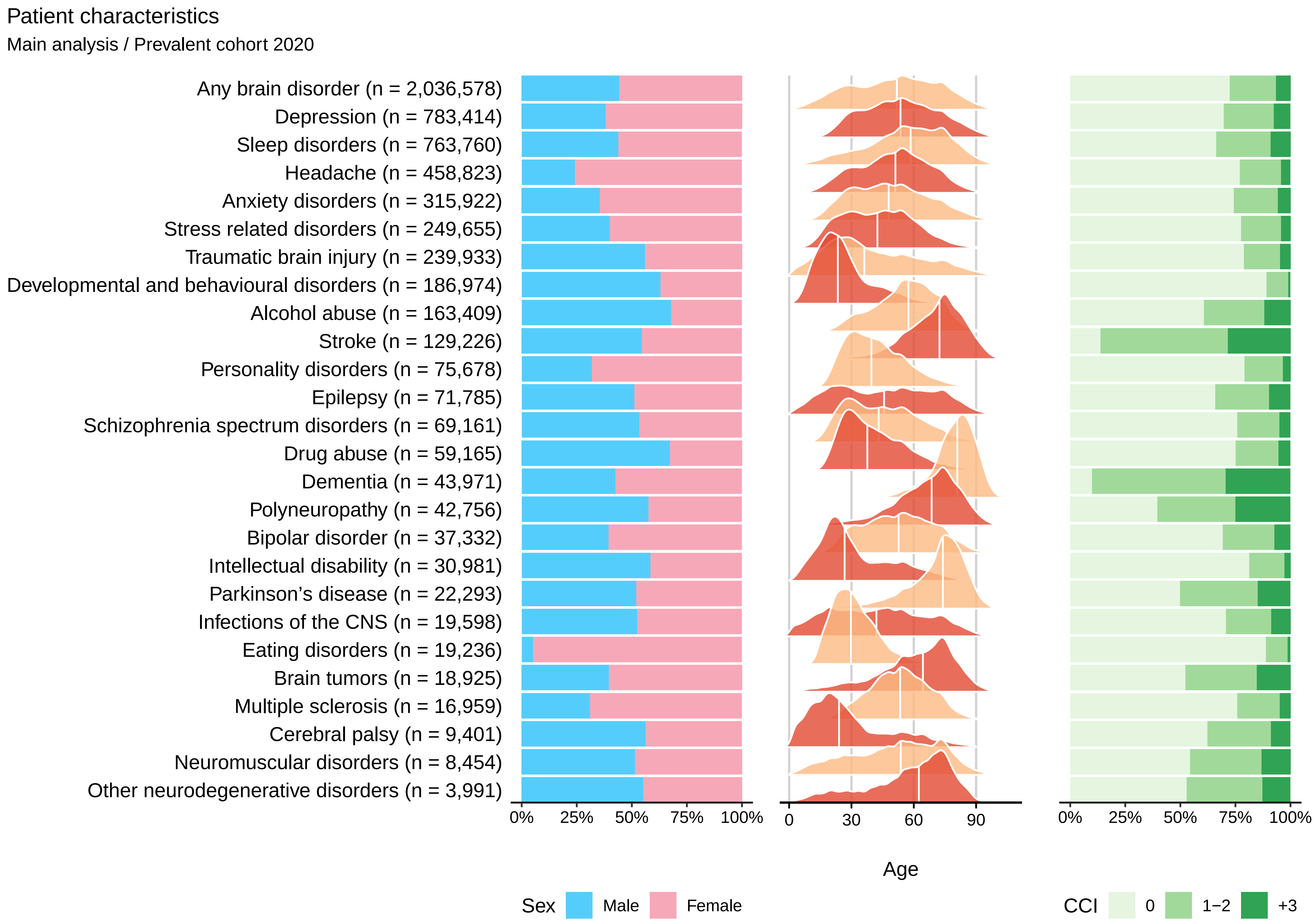

Patient characteristics
Main analysis / Prevalent cohort 2020
Any brain disorder (n = 2,036,578)
Depression (n = 783,414)
Sleep disorders (n = 763,760)
Headache (n = 458,823)
Anxiety disorders (n = 315,922)
Stress related disorders (n = 249,655)
Traumatic brain injury (n = 239,933)
Developmental and behavioural disorders (n = 186,974)
Alcohol abuse (n = 163,409)
Stroke (n = 129,226)
Personality disorders (n = 75,678)
Epilepsy (n = 71,785)
Schizophrenia spectrum disorders (n = 69,161)
Drug abuse (n = 59,165)
Dementia (n = 43,971)
Polyneuropathy (n = 42,756)
Bipolar disorder (n = 37,332)
Intellectual disability (n = 30,981)
Parkinson’s disease (n = 22,293)
Infections of the CNS (n = 19,598)
Eating disorders (n = 19,236)
Brain tumors (n = 18,925)
Multiple sclerosis (n = 16,959)
Cerebral palsy (n = 9,401)
Neuromuscular disorders (n = 8,454)
Other neurodegenerative disorders (n = 3,991)
0% 25% 50% 75% 100%
0 30 60 90
Age
0% 25% 50% 75% 100%
Sex Male Female
CCI 0 1−2 +3

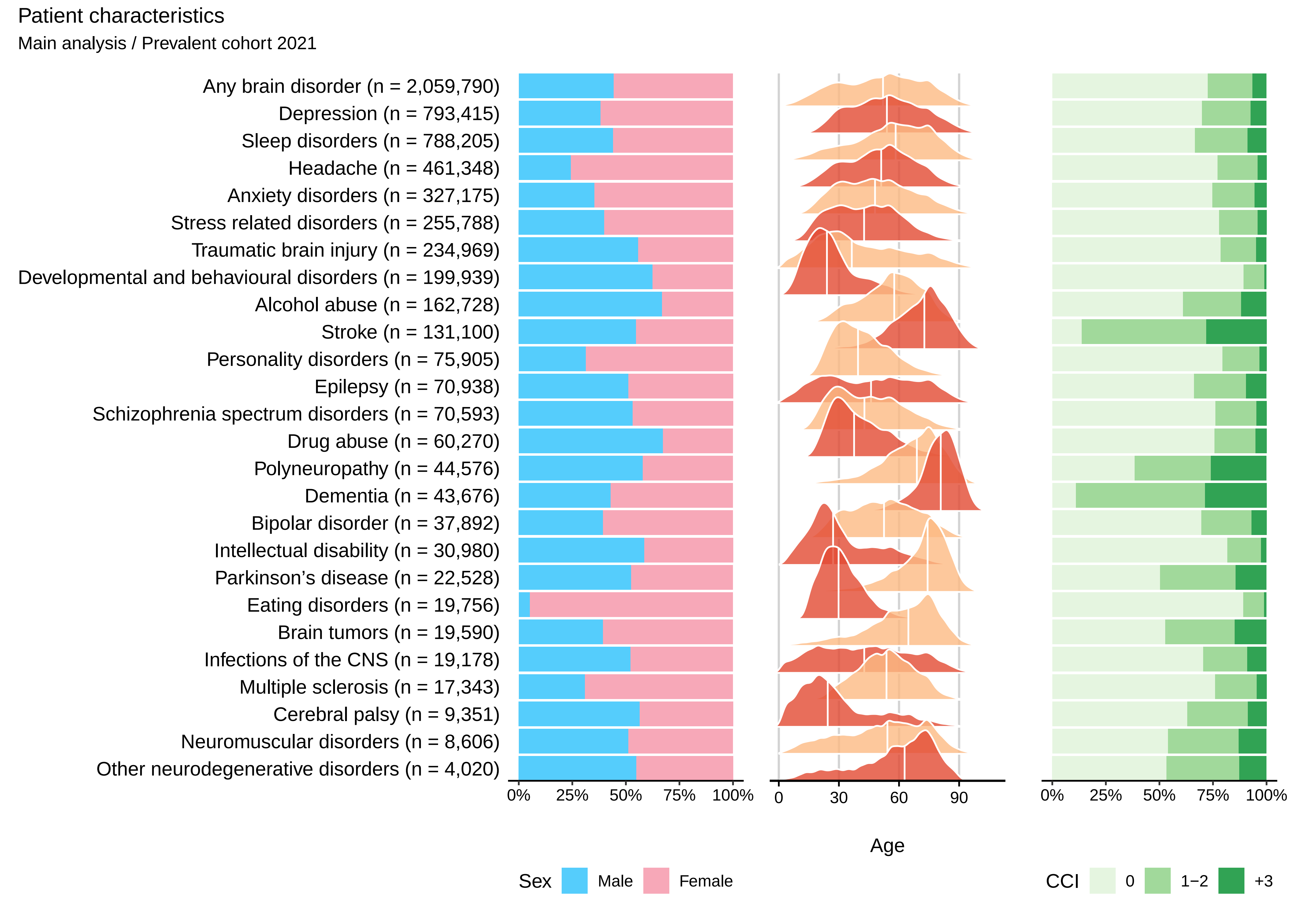

Patient characteristics
Main analysis / Prevalent cohort 2021
Any brain disorder (n = 2,059,790)
Depression (n = 793,415)
Sleep disorders (n = 788,205)
Headache (n = 461,348)
Anxiety disorders (n = 327,175)
Stress related disorders (n = 255,788)
Traumatic brain injury (n = 234,969)
Developmental and behavioural disorders (n = 199,939)
Alcohol abuse (n = 162,728)
Stroke (n = 131,100)
Personality disorders (n = 75,905)
Epilepsy (n = 70,938)
Schizophrenia spectrum disorders (n = 70,593)
Drug abuse (n = 60,270)
Polyneuropathy (n = 44,576)
Dementia (n = 43,676)
Bipolar disorder (n = 37,892)
Intellectual disability (n = 30,980)
Parkinson's disease (n = 22,528)
Eating disorders (n = 19,756)
Brain tumors (n = 19,590)
Infections of the CNS (n = 19,178)
Multiple sclerosis (n = 17,343)
Cerebral palsy (n = 9,351)
Neuromuscular disorders (n = 8,606)
Other neurodegenerative disorders (n = 4,020)
0% 25% 50% 75% 100%
0 30 60 90
Age
0% 25% 50% 75% 100%
Sex Male Female
CCI 0 1−2 +3

### Concurrent diseases

#### Main analysis − Incident cohort 2011−2015

| | Alcohol abuse | Anxiety disorders | Bipolar disorder | Brain tumors | Cerebral palsy | Dementia | Depression | Developmental and behavioural disorders | Drug abuse | Eating disorders | Epilepsy | Headache | Infections of the CNS | Intellectual disability | Multiple sclerosis | Neuromuscular disorders | Other neurodegenerative disorders | Parkinson's disease | Personality disorders | Polyneuropathy | Schizophrenia spectrum disorders | Sleep disorders | Stress related disorders | Stroke | Traumatic brain injury |
|---|---|---|---|---|---|---|---|---|---|---|---|---|---|---|---|---|---|---|---|---|---|---|---|---|---|
| Alcohol abuse | | 9.1 | 18.8 | 5.2 | 4.2 | 8.3 | 6.3 | 5.8 | 23.7 | 2.3 | 11.1 | 2.9 | 6.6 | 5.6 | 3.7 | 6.3 | 6.7 | 5.2 | 11.9 | 15.4 | 13.5 | 6 | 9.6 | 9.2 | 8.1 |
| Anxiety disorders | 13.1 | | 33.3 | 6.1 | 5.4 | 9.5 | 11.5 | 13.9 | 22.7 | 17.9 | 8.6 | 7.8 | 5.7 | 11.1 | 7.1 | 6.8 | 7.6 | 9.4 | 36.6 | 8.4 | 24.7 | 9.6 | 18 | 6.6 | 6.4 |
| Bipolar disorder | 2.3 | 2.2 | | 1 | 0.7 | 1.8 | 0.7 | 1.8 | 5 | 1.4 | 1.6 | 0.7 | 1.1 | 1.5 | 0.8 | 1.2 | 1 | 2.1 | 5.2 | 1.6 | 5.3 | 1.3 | 2.4 | 1.3 | 1 |
| Brain tumors | 0.3 | 0.7 | 0.3 | | 2 | 0.9 | 0.6 | 0.1 | 0.2 | 0.2 | 7 | 0.4 | 3 | 0.5 | 0.8 | 1.2 | 1 | 0.9 | 0.1 | 0.8 | 0.3 | 0.8 | 0.3 | 1.2 | 0.5 |
| Cerebral palsy | 0.1 | 0.2 | 0.2 | 0.2 | | 0.1 | 0.1 | 0.5 | 0.2 | 0.3 | 1.7 | 0.1 | 0.6 | 4.6 | 0.3 | 1 | 1.6 | 0.3 | 0.2 | 0.3 | 0.3 | 0.3 | 0.3 | 0.2 | 0.2 |
| Dementia | 2.1 | 2.7 | 2.3 | 1.9 | 2.6 | | 3.9 | 0.2 | 1.1 | 0.1 | 5.8 | 0.2 | 2.3 | 2 | 0.8 | 1.2 | 7 | 9.3 | 0.5 | 2 | 3.2 | 1.6 | 0.8 | 5.3 | 2.9 |
| Depression | 37.3 | 55.5 | 79.4 | 20.3 | 16 | 40.3 | | 26.2 | 46.3 | 32 | 26.5 | 21.9 | 18.3 | 18.4 | 22.2 | 21.3 | 26 | 34.8 | 67.5 | 29.1 | 48.8 | 28.3 | 45.5 | 23.8 | 19.3 |
| Developmental and behavioural disorders | 4.8 | 8 | 12.2 | 1 | 6.7 | 0.4 | 3.7 | | 22.9 | 9.6 | 4.7 | 3 | 2.2 | 40.8 | 2.4 | 2.4 | 1.2 | 0.8 | 19.9 | 0.9 | 17.2 | 7.3 | 10.1 | 0.5 | 3.6 |
| Drug abuse | 7.7 | 4.1 | 10.7 | 0.8 | 1.1 | 1.1 | 1.9 | 6 | | 2.8 | 3 | 1.4 | 2 | 4.3 | 1.2 | 1.5 | 1 | 1 | 13.5 | 1.8 | 17.8 | 1.9 | 5.6 | 1.2 | 2.1 |
| Eating disorders | 0.6 | 1.4 | 2.1 | 0.2 | 0.1 | 0 | 0.7 | 1.2 | 1.5 | | 0.4 | 0.7 | 0.5 | 0.7 | 0.5 | 0.4 | 0 | 0.1 | 4.3 | 0.2 | 1.7 | 0.6 | 1.3 | 0.1 | 0.4 |
| Epilepsy | 2.6 | 2.5 | 3 | 8.7 | 21.5 | 3.8 | 2.1 | 2.7 | 3.3 | 1.7 | | 1.7 | 5.2 | 15.1 | 2.3 | 3.6 | 4.2 | 3.3 | 2.9 | 3 | 3.9 | 2.3 | 2.5 | 3.3 | 3.1 |
| Headache | 7.3 | 13 | 14 | 12.4 | 4.2 | 6.4 | 11.2 | 5.7 | 9 | 7 | 8.8 | | 10.2 | 3.5 | 14 | 11.1 | 8.7 | 10.3 | 12.2 | 11.1 | 8.3 | 11.8 | 11.8 | 8.7 | 7.3 |
| Infections of the CNS | 0.4 | 0.5 | 0.4 | 1.1 | 2.6 | 0.5 | 0.5 | 0.5 | 0.6 | 0.4 | 2.1 | 0.6 | | 0.9 | 1.6 | 1.2 | 1 | 0.5 | 0.4 | 0.9 | 0.6 | 0.5 | 0.5 | 0.8 | 0.5 |
| Intellectual disability | 0.8 | 1.2 | 1.5 | 0.4 | 12.4 | 1 | 0.7 | 4 | 2.5 | 1 | 3.4 | 0.4 | 0.7 | | 0.4 | 1.2 | 1 | 0.6 | 2.1 | 0.2 | 4.1 | 1.1 | 1.6 | 0.4 | 0.8 |
| Multiple sclerosis | 0.3 | 0.4 | 0.3 | 0.6 | 0.7 | 0.3 | 0.4 | 0.1 | 0.2 | 0.1 | 0.8 | 0.3 | 0.5 | 0.1 | | 0.5 | 1.7 | 0.4 | 0.1 | 0.5 | 0.1 | 0.7 | 0.2 | 0.4 | 0.2 |
| Neuromuscular disorders | 0.2 | 0.2 | 0.2 | 0.4 | 0.9 | 0.2 | 0.2 | 0.1 | 0.2 | 0.1 | 0.3 | 0.1 | 0.4 | 0.6 | 0.6 | | 4 | 0.3 | 0.1 | 1.3 | 0.1 | 0.2 | 0.2 | 0.3 | 0.2 |
| Other neurodegenerative disorders | 0.1 | 0.2 | 0.1 | 0.1 | 1.5 | 0.4 | 0.2 | 0.1 | 0.1 | 0 | 0.2 | 0.1 | 0.2 | 0.3 | 0.8 | 2.4 | | 0.6 | 0.1 | 0.6 | 0.1 | 0.2 | 0.1 | 0.1 | 0.1 |
| Parkinson's disease | 0.6 | 1 | 0.7 | 1.4 | 1.7 | 6.7 | 1.2 | 0.2 | 0.5 | 0 | 1.7 | 0.5 | 0.9 | 0.2 | 0.7 | 1.7 | 4.5 | | 0.2 | 2.7 | 0.9 | 1.2 | 0.5 | 1.9 | 1 |
| Personality disorders | 4.6 | 6.4 | 16.5 | 1.2 | 1 | 1 | 2.1 | 6.4 | 14.6 | 10.8 | 2.3 | 2.8 | 1.8 | 5.2 | 2.8 | 2 | 1.4 | 1.6 | | 2 | 13.9 | 2.8 | 6.6 | 1.2 | 2 |
| Polyneuropathy | 2.1 | 1.1 | 0.8 | 1.5 | 1.6 | 2.7 | 1.3 | 0.2 | 0.9 | 0.2 | 1.9 | 0.6 | 2 | 0.3 | 3.1 | 6.8 | 7.9 | 3.2 | 0.5 | | 0.8 | 1.2 | 0.7 | 2.3 | 1.1 |
| Schizophrenia spectrum disorders | 5.2 | 4.2 | 14 | 1.6 | 1.6 | 3 | 2.1 | 3.3 | 16.8 | 3.4 | 3.1 | 1.3 | 1.9 | 8.8 | 1.5 | 1.5 | 2.1 | 2.1 | 9.2 | 1.7 | | 2.4 | 5.1 | 1.6 | 1.7 |
| Sleep disorders | 25.1 | 29.3 | 41.3 | 22.5 | 14.4 | 29.8 | 25.6 | 14.2 | 27.3 | 10.5 | 21.8 | 15.9 | 17.6 | 13.2 | 15.9 | 20.8 | 24.1 | 27.7 | 25.8 | 30 | 24.5 | | 23.3 | 24.6 | 15.3 |
| Stress related disorders | 12.4 | 15.1 | 31.2 | 4 | 5.2 | 3.7 | 8.7 | 13.9 | 30 | 16.6 | 6 | 6.8 | 5.2 | 14.2 | 5.6 | 4.7 | 4.7 | 4.4 | 40.5 | 5.8 | 30.1 | 7.8 | | 3.7 | 5.8 |
| Stroke | 5.2 | 4.6 | 2.8 | 9.6 | 14.2 | 16.8 | 6.5 | 0.6 | 2.1 | 0.4 | 21.2 | 2 | 8.5 | 1.9 | 3.3 | 5.3 | 7.1 | 10.1 | 1 | 8.7 | 2.7 | 4.6 | 2 | | 6.9 |
| Traumatic brain injury | 9.8 | 7 | 9.6 | 5 | 7.8 | 7.2 | 6.2 | 8.3 | 14.6 | 7.6 | 11.4 | 7.3 | 8.2 | 7.9 | 5.5 | 6.1 | 6.4 | 5.1 | 11.2 | 6.4 | 10.8 | 5.7 | 8.8 | 7.5 | |
| Myocardial infarction | 1.7 | 1.9 | 1 | 2.2 | 1.1 | 4.9 | 2.2 | 0.2 | 0.7 | 0 | 2.5 | 0.5 | 1.6 | 0.3 | 0.6 | 2.8 | 2.5 | 3.4 | 0.4 | 3.8 | 0.8 | 2.4 | 0.9 | 4.8 | 1.7 |
| Congestive heart failure | 2.6 | 2.3 | 1.3 | 2.3 | 1.4 | 8 | 2.9 | 0.2 | 1 | 0.1 | 3.5 | 0.5 | 2.2 | 0.6 | 0.4 | 3.8 | 2.9 | 4.2 | 0.3 | 5.7 | 1.4 | 3 | 0.9 | 6.7 | 2.5 |
| Peripheral vascular disease | 2.3 | 1.9 | 0.9 | 2.3 | 1.7 | 5.5 | 2.4 | 0.2 | 0.9 | 0.4 | 3.2 | 0.7 | 2.2 | 0.3 | 0.8 | 3 | 3 | 3.7 | 0.4 | 5.7 | 0.9 | 2.3 | 0.8 | 5.4 | 1.7 |
| Cerebrovascular disease | 5.4 | 5.8 | 3.4 | 9.9 | 15.1 | 20.4 | 7.8 | 0.7 | 2.5 | 0.3 | 22.4 | 2.4 | 7.8 | 2 | 3.8 | 6.7 | 9.9 | 13.6 | 1.2 | 10.6 | 3 | 5.6 | 2.5 | 11.7 | 6.6 |
| Chronic pulmonary disease | 7.3 | 7.6 | 6.1 | 6.3 | 8.1 | 10.8 | 6.8 | 6.2 | 5.9 | 3 | 8.3 | 4.6 | 7.1 | 6.4 | 3.7 | 10.1 | 9.2 | 8 | 4.8 | 11 | 5.8 | 7.4 | 5.7 | 9.6 | 6.5 |
| Connective tissue disease | 1.4 | 2.2 | 1.7 | 2.7 | 2.1 | 4.1 | 2.4 | 1.1 | 1.1 | 0.9 | 2.4 | 1.9 | 3.4 | 1.5 | 2 | 9 | 2.5 | 3.5 | 1.4 | 5.8 | 1.4 | 2.4 | 1.7 | 3.7 | 1.8 |
| Ulcer disease | 2.5 | 1.6 | 1.4 | 1.6 | 1.4 | 4.3 | 1.7 | 0.3 | 1.4 | 0.2 | 2.3 | 0.7 | 2 | 0.6 | 0.7 | 2 | 1.8 | 2.7 | 0.7 | 3.3 | 1.3 | 1.6 | 1 | 3.3 | 1.5 |
| Mild liver disease | 2.2 | 1.2 | 1.6 | 0.9 | 0.5 | 1.2 | 1 | 0.6 | 2.6 | 0.3 | 2 | 0.6 | 1.9 | 0.5 | 0.4 | 1.5 | 1 | 1 | 1.2 | 3.2 | 2 | 1 | 1.2 | 1.6 | 1.3 |
| Diabetes without end−organ damage | 5 | 4.4 | 3.7 | 5.3 | 3.1 | 9.9 | 4.8 | 0.9 | 2.7 | 0.9 | 6.3 | 1.9 | 5.4 | 2.2 | 2 | 5.7 | 5.2 | 8.1 | 1.9 | 16.3 | 3.1 | 5 | 3 | 9 | 3.6 |
| Hemiplegia | 0.2 | 0.3 | 0.2 | 0.5 | 14 | 0.3 | 0.3 | 0.2 | 0.2 | 0.1 | 1.3 | 0.2 | 1.1 | 1.8 | 1.2 | 1.2 | 2.7 | 0.6 | 0.1 | 0.5 | 0.2 | 0.4 | 0.3 | 0.3 | 0.3 |
| Moderate to severe renal disease | 1.5 | 1.5 | 0.9 | 1.7 | 1.5 | 4.2 | 1.9 | 0.4 | 1.1 | 0.4 | 2.8 | 0.6 | 2.7 | 0.9 | 0.5 | 4.4 | 2 | 2.8 | 0.6 | 5.1 | 1 | 2.2 | 1 | 4 | 1.6 |
| Diabetes with end−organ damage | 2.4 | 2.1 | 1.6 | 2.5 | 2 | 5.5 | 2.6 | 0.3 | 1.3 | 0.3 | 3.7 | 0.8 | 2.9 | 0.9 | 0.9 | 2.9 | 2.8 | 4.6 | 0.9 | 11.4 | 1.5 | 2.5 | 1.3 | 5.2 | 1.9 |
| Solid cancer tumour | 5.6 | 9.6 | 3.2 | 14.8 | 3.6 | 12.7 | 8.1 | 1.2 | 1.9 | 0.4 | 10.8 | 3.6 | 7.3 | 1.2 | 2.3 | 7.4 | 7.7 | 11 | 1 | 12.4 | 2.3 | 10.4 | 2.8 | 11.4 | 4.4 |
| Leukaemia | 0.1 | 0.2 | 0.2 | 0.5 | 0.1 | 0.3 | 0.3 | 0.1 | 0.1 | 0 | 0.4 | 0.1 | 0.9 | 0 | 0 | 0.7 | 0.4 | 0.3 | 0 | 0.7 | 0.1 | 0.4 | 0.2 | 0.4 | 0.2 |
| Lymphoma | 0.3 | 0.5 | 0.3 | 1.4 | 0.2 | 0.7 | 0.5 | 0.1 | 0.2 | 0 | 0.6 | 0.2 | 1.2 | 0.1 | 0.2 | 1.2 | 0.7 | 0.6 | 0.1 | 1.7 | 0.2 | 0.8 | 0.3 | 0.8 | 0.3 |
| Moderate to severe liver disease | 1.2 | 0.3 | 0.3 | 0.2 | 0.2 | 0.5 | 0.2 | 0.1 | 0.3 | 0 | 0.7 | 0.1 | 0.8 | 0.1 | 0.1 | 0.8 | 0.5 | 0.3 | 0.2 | 1 | 0.3 | 0.3 | 0.2 | 0.6 | 0.4 |
| Metastatic solid tumour | 0.5 | 2.6 | 0.3 | 4.6 | 0.6 | 0.7 | 1.5 | 0.5 | 0.2 | 0 | 2.1 | 0.6 | 1.2 | 0.1 | 0.3 | 0.8 | 0.5 | 0.8 | 0.1 | 1.5 | 0.2 | 2.2 | 0.3 | 1.3 | 0.4 |
| AIDS | 0.2 | 0.1 | 0.2 | 0.1 | 0 | 0.1 | 0.1 | 0.1 | 0.3 | 0 | 0.1 | 0.1 | 0.7 | 0.1 | 0 | 0.1 | 0 | 0.1 | 0.2 | 0.3 | 0.3 | 0.1 | 0.2 | 0.1 | 0.1 |

Percent: 60, 40, 20, 0

### Concurrent diseases

#### Main analysis − Incident cohort 2016−2021

| | Alcohol abuse | Anxiety disorders | Bipolar disorder | Brain tumors | Cerebral palsy | Dementia | Depression | Developmental and behavioural disorders | Drug abuse | Eating disorders | Epilepsy | Headache | Infections of the CNS | Intellectual disability | Multiple sclerosis | Neuromuscular disorders | Other neurodegenerative disorders | Parkinson's disease | Personality disorders | Polyneuropathy | Schizophrenia spectrum disorders | Sleep disorders | Stress related disorders | Stroke | Traumatic brain injury |
|---|---|---|---|---|---|---|---|---|---|---|---|---|---|---|---|---|---|---|---|---|---|---|---|---|---|
| Alcohol abuse | | 6.5 | 14.1 | 4.1 | 3 | 7.3 | 5.1 | 3.9 | 19.3 | 1.5 | 8.6 | 2.4 | 6.5 | 4.7 | 2.8 | 4.9 | 5.6 | 4.9 | 8.7 | 11.8 | 9.5 | 5.1 | 6.8 | 7.9 | 6.9 |
| Anxiety disorders | 15.7 | | 32.7 | 7.3 | 5.2 | 9.4 | 12.3 | 15.3 | 21.9 | 14.8 | 7.9 | 9.3 | 7.4 | 11.4 | 8.4 | 8 | 6.8 | 9.3 | 38.5 | 9.3 | 26.7 | 11.6 | 18.4 | 7.1 | 7.3 |
| Bipolar disorder | 2.2 | 1.6 | | 1 | 0.8 | 1.6 | 0.7 | 1.8 | 4.8 | 1.3 | 1.2 | 0.9 | 1.3 | 1.2 | 0.9 | 0.8 | 1.2 | 1.9 | 4.9 | 1.4 | 4.7 | 1.5 | 2.1 | 1.1 | 1.1 |
| Brain tumors | 0.5 | 0.6 | 0.3 | | 1.6 | 1.1 | 0.8 | 0.1 | 0.2 | 0.1 | 8 | 0.4 | 2.4 | 0.5 | 0.8 | 0.8 | 1.3 | 1.3 | 0.2 | 1 | 0.3 | 0.9 | 0.4 | 1.3 | 0.7 |
| Cerebral palsy | 0.1 | 0.2 | 0.2 | 0.2 | | 0.1 | 0.2 | 0.4 | 0.2 | 0.2 | 1.7 | 0.1 | 0.4 | 3.9 | 0.5 | 1 | 1.9 | 0.3 | 0.1 | 0.2 | 0.3 | 0.3 | 0.2 | 0.2 | 0.2 |
| Dementia | 1.7 | 1.6 | 1.4 | 1.7 | 1.5 | | 4.5 | 0.2 | 0.7 | 0.1 | 5.2 | 0.2 | 1.9 | 1.3 | 0.7 | 1.1 | 4.9 | 9.7 | 0.2 | 1.9 | 1.7 | 1.8 | 0.6 | 4.6 | 3.5 |
| Depression | 39.2 | 42.5 | 71.7 | 21.8 | 14.6 | 36.5 | | 23.7 | 40.3 | 22.5 | 23.8 | 21.1 | 19.7 | 16 | 20.6 | 20.7 | 25.5 | 31.5 | 61.3 | 29 | 42.8 | 28.2 | 38 | 23.3 | 19.9 |
| Developmental and behavioural disorders | 7.1 | 12.5 | 15.6 | 1.3 | 5.6 | 0.4 | 6.5 | | 26.7 | 12.4 | 5.4 | 4.8 | 3.3 | 46 | 3.3 | 2.9 | 1.8 | 0.9 | 25.7 | 1.5 | 24 | 10.1 | 15.4 | 0.8 | 4.4 |
| Drug abuse | 7.9 | 3.9 | 10.2 | 0.8 | 1 | 0.9 | 2 | 4.5 | | 2.1 | 2.3 | 1.5 | 2.1 | 5.3 | 1.3 | 1.2 | 0.6 | 0.9 | 12.4 | 1.6 | 15.4 | 2.3 | 5.1 | 1.1 | 2.1 |
| Eating disorders | 0.8 | 1.5 | 2.4 | 0.2 | 0.1 | 0 | 0.8 | 1.3 | 1.3 | | 0.3 | 0.8 | 0.5 | 0.5 | 0.5 | 0.6 | 0.1 | 0.2 | 4.7 | 0.2 | 1.8 | 0.8 | 1.3 | 0.1 | 0.5 |
| Epilepsy | 2.2 | 2 | 2.1 | 6.9 | 16.5 | 3.2 | 2 | 1.9 | 2.9 | 1.4 | | 1.4 | 4.7 | 12.3 | 1.7 | 2.3 | 2.6 | 2.8 | 1.9 | 2.6 | 3.1 | 2 | 2.1 | 2.8 | 3 |
| Headache | 12.4 | 12.9 | 14.2 | 12.8 | 4.1 | 6.8 | 10.9 | 6.6 | 9.3 | 8.1 | 8.3 | | 12.1 | 4 | 13.6 | 10.6 | 10.3 | 9.5 | 13 | 10.7 | 9.1 | 12 | 12.2 | 8.7 | 8.3 |
| Infections of the CNS | 0.5 | 0.4 | 0.6 | 0.9 | 2.5 | 0.6 | 0.5 | 0.4 | 0.6 | 0.4 | 2.2 | 0.6 | | 0.9 | 2.8 | 0.7 | 0.7 | 0.6 | 0.5 | 0.8 | 0.5 | 0.5 | 0.5 | 0.7 | 0.5 |
| Intellectual disability | 0.9 | 1.3 | 1.2 | 0.3 | 7.6 | 0.7 | 0.8 | 2.8 | 2.7 | 0.5 | 3 | 0.5 | 0.6 | | 0.5 | 0.8 | 0.9 | 0.4 | 1.7 | 0.4 | 3.7 | 1 | 1.5 | 0.4 | 0.8 |
| Multiple sclerosis | 0.4 | 0.3 | 0.2 | 0.5 | 0.6 | 0.3 | 0.4 | 0.1 | 0.2 | 0.1 | 0.8 | 0.3 | 0.7 | 0.1 | | 0.6 | 0.8 | 0.5 | 0.2 | 0.4 | 0.2 | 0.6 | 0.3 | 0.4 | 0.3 |
| Neuromuscular disorders | 0.3 | 0.2 | 0.2 | 0.3 | 1.3 | 0.3 | 0.2 | 0.1 | 0.2 | 0.1 | 0.4 | 0.2 | 0.3 | 0.4 | 0.3 | | 3.1 | 0.4 | 0.2 | 1 | 0.2 | 0.3 | 0.2 | 0.3 | 0.2 |
| Other neurodegenerative disorders | 0.1 | 0.2 | 0.1 | 0.1 | 1.2 | 0.3 | 0.2 | 0 | 0 | 0 | 0.1 | 0.1 | 0.1 | 0.2 | 0.6 | 2.2 | | 0.8 | 0 | 0.5 | 0.1 | 0.2 | 0.1 | 0.2 | 0.2 |
| Parkinson's disease | 0.9 | 0.6 | 0.4 | 1 | 1.2 | 7.9 | 1.2 | 0.1 | 0.4 | 0 | 1.3 | 0.3 | 0.8 | 0.2 | 0.3 | 1.4 | 4.4 | | 0.2 | 1.9 | 0.5 | 1.1 | 0.4 | 1.5 | 1.2 |
| Personality disorders | 4.6 | 5.1 | 12.9 | 1.2 | 0.5 | 0.6 | 1.9 | 4.7 | 11.2 | 6.7 | 1.9 | 2.9 | 1.9 | 4.4 | 2.9 | 1.6 | 0.9 | 1 | | 1.9 | 11.5 | 2.8 | 4.8 | 1.1 | 2 |
| Polyneuropathy | 2.6 | 1 | 0.8 | 1.9 | 1.2 | 3.2 | 1.5 | 0.2 | 0.7 | 0.2 | 2.1 | 0.6 | 2.2 | 0.4 | 2 | 7.6 | 9 | 3.7 | 0.4 | | 0.7 | 1.3 | 0.8 | 2.9 | 1.5 |
| Schizophrenia spectrum disorders | 4.5 | 4.3 | 13.1 | 1.3 | 1.5 | 2.2 | 2.5 | 3.4 | 14.6 | 3.3 | 2.8 | 1.6 | 2 | 9.3 | 1.9 | 1.2 | 1 | 1.6 | 8.2 | 1.6 | | 2.8 | 4.7 | 1.4 | 1.9 |
| Sleep disorders | 30.7 | 30.3 | 43.9 | 24.6 | 15.2 | 30.7 | 29 | 18.7 | 30.4 | 13.7 | 23.6 | 17 | 19.9 | 17.5 | 18.3 | 23.1 | 26.4 | 28.9 | 32.4 | 32.3 | 29.1 | | 25.1 | 25.3 | 18.6 |
| Stress related disorders | 13.9 | 17.4 | 31.2 | 4.8 | 4.2 | 3.9 | 11.1 | 16 | 30.8 | 15.4 | 6 | 8.5 | 6.2 | 17.2 | 7.5 | 5.1 | 4.1 | 4.5 | 42.2 | 6.5 | 31.9 | 10.1 | | 4.1 | 6.6 |
| Stroke | 4.4 | 3.2 | 2.2 | 8.7 | 14.6 | 14.7 | 6.7 | 0.5 | 1.8 | 0.3 | 20.9 | 1.7 | 7.5 | 1.7 | 3.1 | 5.7 | 7 | 9.8 | 0.7 | 9 | 2 | 4.6 | 1.9 | | 8.1 |
| Traumatic brain injury | 8.3 | 6 | 8 | 4.8 | 7.8 | 7 | 5.7 | 6.6 | 12.4 | 6.2 | 10.7 | 7 | 7.5 | 6.7 | 5.6 | 4.6 | 5.4 | 5.3 | 9.1 | 5.9 | 8.7 | 5.7 | 7.3 | 7.3 | |
| Myocardial infarction | 1.5 | 1.4 | 0.8 | 2.1 | 1.1 | 3.8 | 1.9 | 0.2 | 0.8 | 0 | 2.3 | 0.4 | 1.4 | 0.2 | 0.6 | 2.5 | 2.4 | 2.9 | 0.3 | 4 | 0.7 | 1.9 | 0.8 | 4.2 | 1.7 |
| Congestive heart failure | 2 | 1.6 | 0.8 | 2.7 | 1.5 | 6.3 | 2.7 | 0.2 | 0.9 | 0.1 | 3.2 | 0.4 | 2.2 | 0.6 | 0.4 | 3.4 | 2.9 | 3.5 | 0.3 | 6.6 | 0.9 | 2.6 | 0.8 | 5.8 | 3 |
| Peripheral vascular disease | 2 | 1.2 | 0.7 | 2.1 | 1.5 | 4.6 | 2.1 | 0.2 | 0.7 | 0.3 | 2.6 | 0.6 | 2 | 0.3 | 0.8 | 2 | 2.3 | 3.2 | 0.4 | 5.8 | 0.8 | 1.8 | 0.7 | 4.6 | 1.9 |
| Cerebrovascular disease | 4.8 | 4 | 2.8 | 8.9 | 14.6 | 18.2 | 7.9 | 0.6 | 2 | 0.3 | 22.6 | 2.2 | 7.5 | 1.8 | 3.7 | 6.8 | 8.2 | 12.8 | 0.8 | 10.6 | 2.3 | 5.5 | 2.3 | 10.1 | 7.8 |
| Chronic pulmonary disease | 7.5 | 7.3 | 5.5 | 7.3 | 7 | 10.5 | 7 | 5.5 | 5.8 | 3.6 | 8 | 4.9 | 7.5 | 6.1 | 2.8 | 9.5 | 10.6 | 7.8 | 4.8 | 11.6 | 5.4 | 7.4 | 5.4 | 9.4 | 6.8 |
| Connective tissue disease | 2.9 | 2 | 1.7 | 3.3 | 1.8 | 4.2 | 2.5 | 1.1 | 1.1 | 1.3 | 2.6 | 1.8 | 4 | 1.4 | 1.9 | 8.4 | 3.9 | 4 | 1.3 | 6 | 1.3 | 2.5 | 1.7 | 4 | 2.2 |
| Ulcer disease | 1.8 | 0.9 | 0.7 | 1.2 | 1.3 | 2.7 | 1.1 | 0.2 | 0.9 | 0.2 | 1.6 | 0.5 | 1.4 | 0.4 | 0.6 | 1.1 | 1.7 | 1.5 | 0.4 | 2.6 | 0.7 | 1 | 0.6 | 2.1 | 1.2 |
| Mild liver disease | 1.7 | 0.9 | 1.2 | 0.8 | 0.7 | 1 | 0.9 | 0.4 | 2 | 0.2 | 1.4 | 0.5 | 1.8 | 0.4 | 0.4 | 1.2 | 0.7 | 0.8 | 0.8 | 2.5 | 1.4 | 0.9 | 1 | 1.3 | 1.2 |
| Diabetes without end−organ damage | 4 | 2.9 | 2.8 | 4.7 | 2.5 | 8.6 | 4.2 | 0.9 | 2.1 | 1 | 5.4 | 1.5 | 5 | 2 | 1.6 | 5.4 | 4 | 6.6 | 1.7 | 23.5 | 2.5 | 3.8 | 2.5 | 8 | 3.8 |
| Hemiplegia | 0.2 | 0.2 | 0.1 | 0.4 | 10.5 | 0.2 | 0.3 | 0.1 | 0.2 | 0 | 1.3 | 0.1 | 0.7 | 1.4 | 1.2 | 0.7 | 2.5 | 0.5 | 0.1 | 0.4 | 0.2 | 0.3 | 0.2 | 0.3 | 0.3 |
| Moderate to severe renal disease | 1.7 | 1.4 | 1 | 2.4 | 1.8 | 4.8 | 2.4 | 0.4 | 1.1 | 0.3 | 3.4 | 0.7 | 3.1 | 1.1 | 0.5 | 3.5 | 2 | 3.4 | 0.5 | 7.7 | 1 | 2.4 | 1 | 4.7 | 2.4 |
| Diabetes with end−organ damage | 2.1 | 1.3 | 1.4 | 2.4 | 1.2 | 4.6 | 2.3 | 0.3 | 1 | 0.2 | 3.4 | 0.7 | 3 | 1.1 | 0.8 | 3.3 | 2.3 | 3.5 | 0.7 | 20.1 | 1.2 | 2.1 | 1.2 | 5.1 | 2.1 |
| Solid cancer tumour | 6.2 | 7.3 | 3.2 | 14.7 | 4.8 | 15.1 | 9.1 | 0.8 | 1.8 | 0.4 | 12.5 | 3 | 7.5 | 1.1 | 2.6 | 8.4 | 9.1 | 13.4 | 1 | 14 | 2.1 | 9.7 | 2.9 | 13.9 | 6.4 |
| Leukaemia | 0.2 | 0.2 | 0.1 | 0.5 | 0.1 | 0.4 | 0.3 | 0.1 | 0.1 | 0 | 0.4 | 0.1 | 0.8 | 0.1 | 0 | 0.3 | 0.3 | 0.5 | 0 | 0.8 | 0.1 | 0.4 | 0.1 | 0.5 | 0.3 |
| Lymphoma | 0.4 | 0.4 | 0.2 | 1.5 | 0.2 | 1 | 0.6 | 0.1 | 0.2 | 0 | 0.9 | 0.2 | 1.4 | 0.1 | 0.2 | 0.9 | 0.8 | 0.9 | 0.1 | 2.3 | 0.2 | 0.8 | 0.2 | 0.9 | 0.5 |
| Moderate to severe liver disease | 1 | 0.2 | 0.2 | 0.2 | 0.2 | 0.4 | 0.2 | 0.1 | 0.4 | 0 | 0.5 | 0.1 | 0.7 | 0.1 | 0.2 | 0.4 | 0.1 | 0.4 | 0.1 | 0.9 | 0.2 | 0.3 | 0.2 | 0.4 | 0.3 |
| Metastatic solid tumour | 0.5 | 1.4 | 0.2 | 2.6 | 0.7 | 0.7 | 1.2 | 0.1 | 0.2 | 0 | 2 | 0.3 | 1 | 0.1 | 0.1 | 0.9 | 0.7 | 0.6 | 0.1 | 1.4 | 0.2 | 1.4 | 0.2 | 1.2 | 0.5 |
| AIDS | 0.1 | 0.1 | 0.2 | 0.1 | 0.1 | 0 | 0.1 | 0.1 | 0.3 | 0 | 0.1 | 0.1 | 0.6 | 0 | 0 | 0.2 | 0 | 0.1 | 0.2 | 0.2 | 0.3 | 0.1 | 0.2 | 0.1 | 0.1 |

Percent: 60, 40, 20, 0

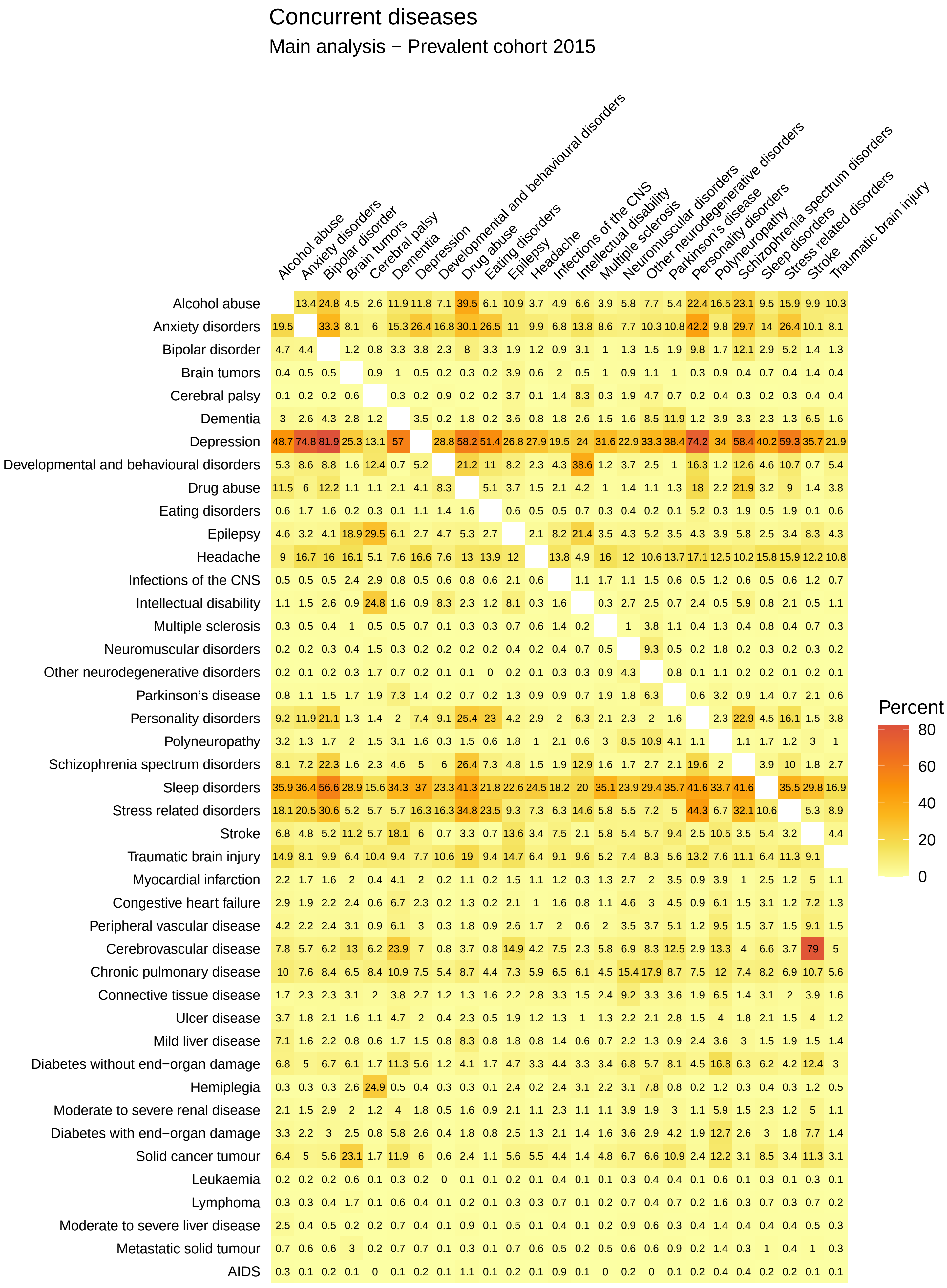
Concurrent diseases
Main analysis − Prevalent cohort 2015
Alcohol abuse
Anxiety disorders
Bipolar disorder
Brain tumors
Cerebral palsy
Dementia
Depression
Developmental and behavioural disorders
Drug abuse
Eating disorders
Epilepsy
Headache
Infections of the CNS
Intellectual disability
Multiple sclerosis
Neuromuscular disorders
Other neurodegenerative disorders
Parkinson's disease
Personality disorders
Polyneuropathy
Schizophrenia spectrum disorders
Sleep disorders
Stress related disorders
Stroke
Traumatic brain injury
Myocardial infarction
Congestive heart failure
Peripheral vascular disease
Cerebrovascular disease
Chronic pulmonary disease
Connective tissue disease
Ulcer disease
Mild liver disease
Diabetes without end−organ damage
Hemiplegia
Moderate to severe renal disease
Diabetes with end−organ damage
Solid cancer tumour
Leukaemia
Lymphoma
Moderate to severe liver disease
Metastatic solid tumour
AIDS
Percent
80
60
40
20
0

### Concurrent diseases

#### Main analysis − Prevalent cohort 2016

| | Alcohol abuse | Anxiety disorders | Bipolar disorder | Brain tumors | Cerebral palsy | Dementia | Depression | Developmental and behavioural disorders | Drug abuse | Eating disorders | Epilepsy | Headache | Infections of the CNS | Intellectual disability | Multiple sclerosis | Neuromuscular disorders | Other neurodegenerative disorders | Parkinson's disease | Personality disorders | Polyneuropathy | Schizophrenia spectrum disorders | Sleep disorders | Stress related disorders | Stroke | Traumatic brain injury |
|---|---|---|---|---|---|---|---|---|---|---|---|---|---|---|---|---|---|---|---|---|---|---|---|---|---|
| Alcohol abuse | | 13 | 24.4 | 4.3 | 2.6 | 11.9 | 11.6 | 7.1 | 38.4 | 5.9 | 10.7 | 3.6 | 4.9 | 6.7 | 3.8 | 5.5 | 7.7 | 5.4 | 21.5 | 15.8 | 22.7 | 9.2 | 15.3 | 9.8 | 10.1 |
| Anxiety disorders | 20.3 | | 34.5 | 8.5 | 6.2 | 15 | 27.1 | 17.8 | 31 | 27.6 | 11.5 | 10.5 | 7.2 | 14.6 | 8.8 | 8.2 | 10.2 | 11.8 | 43.7 | 10.2 | 30.8 | 14.6 | 27.3 | 10.3 | 8.6 |
| Bipolar disorder | 4.9 | 4.4 | | 1.2 | 0.8 | 3.2 | 3.8 | 2.4 | 8.1 | 3.5 | 1.9 | 1.3 | 0.9 | 3 | 1 | 1.2 | 1.5 | 2.1 | 9.9 | 1.7 | 11.9 | 2.9 | 5.2 | 1.4 | 1.4 |
| Brain tumors | 0.4 | 0.5 | 0.6 | | 0.9 | 1 | 0.5 | 0.2 | 0.3 | 0.2 | 4 | 0.6 | 2.1 | 0.5 | 1 | 0.9 | 1.2 | 1.1 | 0.3 | 0.9 | 0.4 | 0.7 | 0.4 | 1.4 | 0.4 |
| Cerebral palsy | 0.1 | 0.2 | 0.2 | 0.6 | | 0.3 | 0.2 | 0.8 | 0.2 | 0.2 | 3.8 | 0.1 | 1.5 | 7.9 | 0.3 | 1.8 | 4.6 | 0.7 | 0.2 | 0.4 | 0.3 | 0.2 | 0.3 | 0.5 | 0.4 |
| Dementia | 3.1 | 2.5 | 4.2 | 2.7 | 1.4 | | 3.4 | 0.2 | 1.7 | 0.2 | 3.7 | 0.8 | 1.9 | 2.7 | 1.4 | 1.7 | 7.8 | 13.7 | 1.1 | 3.9 | 3.2 | 2.3 | 1.2 | 6.4 | 1.7 |
| Depression | 49.6 | 74.3 | 82.2 | 25.5 | 13.5 | 55.9 | | 29.4 | 58.2 | 51.6 | 27.3 | 28.5 | 20 | 24.4 | 31.7 | 23.4 | 32.8 | 40.9 | 74.9 | 34.4 | 59 | 40.7 | 59.6 | 35.6 | 22.3 |
| Developmental and behavioural disorders | 5.7 | 9.3 | 9.9 | 1.7 | 12.7 | 0.8 | 5.6 | | 22.8 | 11.8 | 8.5 | 2.5 | 4.5 | 39.9 | 1.3 | 3.9 | 2.5 | 1.2 | 17.8 | 1.2 | 13.7 | 5.1 | 11.6 | 0.8 | 5.8 |
| Drug abuse | 11.7 | 6.1 | 12.4 | 1.1 | 1.1 | 2.1 | 4.2 | 8.6 | | 5.2 | 3.8 | 1.6 | 2.1 | 4.6 | 1 | 1.4 | 1.1 | 1.3 | 18.1 | 2.1 | 22.3 | 3.2 | 9 | 1.4 | 3.9 |
| Eating disorders | 0.6 | 1.7 | 1.7 | 0.2 | 0.4 | 0.1 | 1.2 | 1.4 | 1.7 | | 0.6 | 0.5 | 0.5 | 0.7 | 0.3 | 0.4 | 0.2 | 0.1 | 5.3 | 0.3 | 2 | 0.6 | 1.9 | 0.1 | 0.6 |
| Epilepsy | 4.6 | 3.1 | 4.1 | 18.5 | 29.7 | 6.2 | 2.7 | 4.5 | 5.2 | 2.6 | | 2 | 8.3 | 21.1 | 3.5 | 4.2 | 5.1 | 3.7 | 4.2 | 3.7 | 5.7 | 2.5 | 3.4 | 8.2 | 4.3 |
| Headache | 9 | 16.9 | 16.2 | 16 | 5.5 | 7.5 | 16.8 | 7.9 | 13 | 14 | 12.1 | | 14.1 | 5 | 16 | 12.2 | 10.3 | 13.4 | 17.4 | 12.5 | 10.4 | 15.9 | 16.1 | 12.3 | 11.2 |
| Infections of the CNS | 0.5 | 0.5 | 0.5 | 2.5 | 3 | 0.8 | 0.5 | 0.6 | 0.7 | 0.6 | 2.1 | 0.6 | | 1.1 | 1.7 | 1.1 | 1.4 | 0.6 | 0.5 | 1.2 | 0.6 | 0.5 | 0.6 | 1.2 | 0.7 |
| Intellectual disability | 1.1 | 1.5 | 2.5 | 0.9 | 24.1 | 1.7 | 0.9 | 8.1 | 2.5 | 1.3 | 8.2 | 0.3 | 1.6 | | 0.3 | 2.6 | 2.3 | 0.7 | 2.5 | 0.5 | 5.9 | 0.9 | 2.1 | 0.5 | 1.1 |
| Multiple sclerosis | 0.3 | 0.5 | 0.5 | 1 | 0.5 | 0.5 | 0.7 | 0.1 | 0.3 | 0.3 | 0.7 | 0.6 | 1.4 | 0.2 | | 1 | 3.6 | 1.1 | 0.4 | 1.3 | 0.4 | 0.8 | 0.4 | 0.7 | 0.3 |
| Neuromuscular disorders | 0.2 | 0.2 | 0.3 | 0.4 | 1.5 | 0.3 | 0.2 | 0.2 | 0.2 | 0.2 | 0.4 | 0.2 | 0.4 | 0.7 | 0.5 | | 8.9 | 0.5 | 0.2 | 1.9 | 0.2 | 0.3 | 0.2 | 0.4 | 0.2 |
| Other neurodegenerative disorders | 0.2 | 0.1 | 0.2 | 0.3 | 1.7 | 0.6 | 0.2 | 0.1 | 0.1 | 0 | 0.2 | 0.1 | 0.3 | 0.3 | 0.8 | 4.1 | | 0.9 | 0.1 | 1.1 | 0.1 | 0.2 | 0.1 | 0.2 | 0.1 |
| Parkinson's disease | 0.7 | 1 | 1.4 | 1.6 | 1.8 | 7.2 | 1.3 | 0.2 | 0.6 | 0.1 | 1.2 | 0.7 | 0.8 | 0.6 | 1.6 | 1.6 | 6 | | 0.5 | 2.9 | 0.8 | 1.3 | 0.6 | 1.9 | 0.6 |
| Personality disorders | 9.2 | 11.9 | 21.2 | 1.3 | 1.4 | 1.9 | 7.5 | 9.3 | 25.2 | 23 | 4.2 | 2.9 | 2 | 6.5 | 2 | 2.3 | 2 | 1.7 | | 2.2 | 22.5 | 4.6 | 15.9 | 1.5 | 3.8 |
| Polyneuropathy | 3.3 | 1.4 | 1.7 | 2.1 | 1.5 | 3.2 | 1.7 | 0.3 | 1.4 | 0.5 | 1.8 | 1 | 2.3 | 0.6 | 3 | 8.8 | 11.3 | 4.6 | 1.1 | | 1.2 | 1.8 | 1.2 | 3.1 | 1.1 |
| Schizophrenia spectrum disorders | 8.3 | 7.2 | 21.8 | 1.5 | 2.3 | 4.5 | 5 | 6.2 | 26.6 | 7.4 | 4.9 | 1.5 | 1.9 | 13.1 | 1.6 | 1.7 | 2.5 | 2.3 | 19.3 | 2 | | 3.9 | 9.9 | 1.8 | 2.8 |
| Sleep disorders | 36.5 | 37.2 | 57.1 | 29 | 17 | 34.6 | 37.7 | 25.1 | 41.3 | 22.8 | 23.5 | 24.9 | 18.9 | 21.3 | 36.3 | 24.8 | 30.1 | 37.2 | 42.3 | 34.3 | 42.1 | | 36.2 | 30.2 | 17.5 |
| Stress related disorders | 18.6 | 21.2 | 31.6 | 5.3 | 6.3 | 5.8 | 16.9 | 17.3 | 35.7 | 24.1 | 9.6 | 7.8 | 6.6 | 15.5 | 5.9 | 5.9 | 7.3 | 5.4 | 45.4 | 6.9 | 33 | 11.1 | | 5.4 | 9.2 |
| Stroke | 7 | 4.7 | 5 | 11 | 6.2 | 17.7 | 5.9 | 0.7 | 3.3 | 0.7 | 13.6 | 3.4 | 7.6 | 2.1 | 5.8 | 5.6 | 5.9 | 9.9 | 2.5 | 10.5 | 3.5 | 5.4 | 3.2 | | 4.5 |
| Traumatic brain injury | 14.6 | 7.9 | 9.8 | 6.5 | 10.4 | 9.4 | 7.6 | 10.3 | 18.4 | 9.1 | 14.5 | 6.4 | 9 | 9.5 | 5.1 | 7.2 | 8.3 | 5.9 | 12.9 | 7.5 | 10.9 | 6.4 | 11 | 9.2 | |
| Myocardial infarction | 2.3 | 1.6 | 1.6 | 1.9 | 0.4 | 4 | 2 | 0.2 | 1.1 | 0.2 | 1.6 | 1.1 | 1.2 | 0.3 | 1.3 | 2.6 | 2.1 | 3.4 | 0.9 | 3.9 | 1 | 2.5 | 1.2 | 4.8 | 1.1 |
| Congestive heart failure | 3 | 1.9 | 2.1 | 2.4 | 0.7 | 6.7 | 2.3 | 0.2 | 1.3 | 0.2 | 2.2 | 1 | 1.6 | 0.7 | 1.1 | 4.7 | 2.8 | 4.6 | 0.9 | 6.1 | 1.5 | 3.1 | 1.1 | 7.2 | 1.3 |
| Peripheral vascular disease | 4.3 | 2.2 | 2.3 | 3.2 | 0.8 | 6.2 | 2.9 | 0.3 | 1.7 | 0.8 | 2.6 | 1.7 | 2.1 | 0.5 | 2.1 | 3.6 | 4.2 | 5.2 | 1.2 | 9.5 | 1.4 | 3.6 | 1.5 | 9.1 | 1.5 |
| Cerebrovascular disease | 7.9 | 5.5 | 6.2 | 12.6 | 6.9 | 23.5 | 6.9 | 0.8 | 3.7 | 0.8 | 14.9 | 4.2 | 7.7 | 2.3 | 5.8 | 6.9 | 8.3 | 13.3 | 2.9 | 13.2 | 4 | 6.6 | 3.7 | 78.4 | 5.1 |
| Chronic pulmonary disease | 10.3 | 7.7 | 8.5 | 6.7 | 8.5 | 11.2 | 7.6 | 5.4 | 8.8 | 4.4 | 7.5 | 6 | 6.7 | 6.2 | 4.6 | 16.1 | 18.4 | 9.2 | 7.6 | 12.2 | 7.5 | 8.4 | 7.1 | 11 | 5.7 |
| Connective tissue disease | 1.7 | 2.4 | 2.3 | 3.1 | 1.9 | 3.9 | 2.8 | 1.2 | 1.3 | 1.7 | 2.3 | 2.9 | 3.4 | 1.6 | 2.5 | 9.6 | 3.1 | 3.8 | 2 | 6.7 | 1.4 | 3.2 | 2.1 | 4 | 1.6 |
| Ulcer disease | 3.6 | 1.7 | 2 | 1.5 | 1.2 | 4.4 | 1.9 | 0.4 | 2.1 | 0.5 | 1.8 | 1.2 | 1.4 | 0.9 | 1.2 | 2.2 | 1.9 | 3 | 1.5 | 3.8 | 1.7 | 1.9 | 1.4 | 3.7 | 1.2 |
| Mild liver disease | 7.2 | 1.6 | 2.1 | 0.8 | 0.6 | 1.7 | 1.6 | 0.8 | 7.9 | 0.8 | 1.9 | 0.9 | 1.4 | 0.6 | 0.8 | 2.2 | 1.2 | 1 | 2.4 | 3.6 | 3 | 1.5 | 1.9 | 1.5 | 1.4 |
| Diabetes without end−organ damage | 6.9 | 5 | 6.8 | 6.1 | 2 | 11.5 | 5.7 | 1.3 | 4.1 | 1.6 | 4.9 | 3.4 | 4.5 | 3.3 | 3.4 | 7 | 5.7 | 8.6 | 4.5 | 17.5 | 6.4 | 6.3 | 4.3 | 12.6 | 3.1 |
| Hemiplegia | 0.3 | 0.3 | 0.3 | 2.5 | 24.6 | 0.5 | 0.4 | 0.3 | 0.3 | 0.1 | 2.4 | 0.2 | 2.4 | 3 | 2.2 | 3.1 | 7.4 | 0.9 | 0.2 | 1.2 | 0.3 | 0.4 | 0.3 | 1.2 | 0.5 |
| Moderate to severe renal disease | 2.3 | 1.5 | 3 | 2.1 | 1.3 | 4.5 | 1.9 | 0.5 | 1.6 | 0.8 | 2.2 | 1.2 | 2.5 | 1.2 | 1.3 | 4.2 | 2.3 | 3.4 | 1.2 | 6.1 | 1.6 | 2.4 | 1.3 | 5.2 | 1.2 |
| Diabetes with end−organ damage | 3.3 | 2.1 | 2.9 | 2.4 | 0.9 | 5.7 | 2.6 | 0.4 | 1.8 | 0.8 | 2.5 | 1.3 | 2.1 | 1.3 | 1.6 | 3.5 | 2.9 | 4.3 | 1.9 | 12.7 | 2.6 | 2.9 | 1.8 | 7.6 | 1.4 |
| Solid cancer tumour | 6.8 | 5.1 | 5.7 | 23.4 | 1.8 | 12.6 | 6.2 | 0.6 | 2.4 | 1.1 | 5.9 | 5.6 | 4.9 | 1.5 | 5 | 7.1 | 6.6 | 11.7 | 2.5 | 12.9 | 3.2 | 8.7 | 3.5 | 11.8 | 3.2 |
| Leukaemia | 0.2 | 0.2 | 0.2 | 0.6 | 0.1 | 0.4 | 0.2 | 0 | 0.1 | 0.1 | 0.2 | 0.2 | 0.4 | 0.1 | 0.1 | 0.3 | 0.5 | 0.4 | 0.1 | 0.7 | 0.1 | 0.3 | 0.1 | 0.4 | 0.1 |
| Lymphoma | 0.3 | 0.3 | 0.4 | 1.9 | 0.1 | 0.7 | 0.4 | 0.1 | 0.2 | 0.1 | 0.4 | 0.3 | 0.7 | 0.1 | 0.2 | 0.8 | 0.6 | 0.8 | 0.2 | 1.7 | 0.3 | 0.7 | 0.3 | 0.7 | 0.2 |
| Moderate to severe liver disease | 2.6 | 0.4 | 0.4 | 0.2 | 0.3 | 0.7 | 0.4 | 0.1 | 0.8 | 0.1 | 0.5 | 0.1 | 0.4 | 0.1 | 0.2 | 0.9 | 0.6 | 0.2 | 0.4 | 1.4 | 0.4 | 0.4 | 0.4 | 0.5 | 0.3 |
| Metastatic solid tumour | 0.8 | 0.6 | 0.5 | 2.9 | 0.2 | 0.7 | 0.7 | 0.1 | 0.2 | 0.1 | 0.7 | 0.6 | 0.6 | 0.2 | 0.5 | 0.6 | 0.4 | 0.9 | 0.2 | 1.6 | 0.3 | 1 | 0.4 | 1 | 0.3 |
| AIDS | 0.3 | 0.1 | 0.2 | 0.1 | 0 | 0.1 | 0.2 | 0.1 | 1 | 0.1 | 0.2 | 0.1 | 0.9 | 0.1 | 0 | 0.1 | 0 | 0.1 | 0.2 | 0.4 | 0.4 | 0.2 | 0.2 | 0.1 | 0.1 |

Percent: 0, 20, 40, 60, 80

### Concurrent diseases

#### Main analysis − Prevalent cohort 2017

| | Alcohol abuse | Anxiety disorders | Bipolar disorder | Brain tumors | Cerebral palsy | Dementia | Depression | Developmental and behavioural disorders | Drug abuse | Eating disorders | Epilepsy | Headache | Infections of the CNS | Intellectual disability | Multiple sclerosis | Neuromuscular disorders | Other neurodegenerative disorders | Parkinson's disease | Personality disorders | Polyneuropathy | Schizophrenia spectrum disorders | Sleep disorders | Stress related disorders | Stroke | Traumatic brain injury |
|---|---|---|---|---|---|---|---|---|---|---|---|---|---|---|---|---|---|---|---|---|---|---|---|---|---|
| Alcohol abuse | | 12.7 | 23.9 | 4.1 | 2.6 | 11.8 | 11.4 | 7 | 37.2 | 5.8 | 10.5 | 3.5 | 5 | 6.8 | 3.7 | 5.5 | 7.2 | 5.3 | 20.8 | 15.3 | 22.2 | 9 | 14.8 | 9.8 | 9.9 |
| Anxiety disorders | 20.9 | | 35.5 | 8.6 | 6.6 | 14.6 | 27.7 | 18.7 | 31.7 | 28.4 | 11.8 | 10.9 | 7.4 | 15.2 | 9 | 8.5 | 10.3 | 12 | 44.9 | 10.3 | 31.7 | 15.2 | 28 | 10.3 | 8.9 |
| Bipolar disorder | 4.9 | 4.4 | | 1.2 | 0.9 | 3.1 | 3.9 | 2.5 | 8.2 | 3.5 | 1.9 | 1.3 | 1 | 2.9 | 1.1 | 1.3 | 1.4 | 2.1 | 10 | 1.7 | 11.7 | 2.9 | 5.2 | 1.4 | 1.4 |
| Brain tumors | 0.4 | 0.5 | 0.6 | | 1 | 1 | 0.6 | 0.2 | 0.3 | 0.2 | 4.1 | 0.6 | 2.1 | 0.5 | 1 | 0.8 | 1.2 | 1.1 | 0.3 | 1 | 0.4 | 0.7 | 0.4 | 1.5 | 0.4 |
| Cerebral palsy | 0.1 | 0.2 | 0.2 | 0.6 | | 0.3 | 0.2 | 0.8 | 0.2 | 0.2 | 3.8 | 0.1 | 1.5 | 7.6 | 0.3 | 1.8 | 4.6 | 0.8 | 0.2 | 0.4 | 0.3 | 0.2 | 0.3 | 0.5 | 0.4 |
| Dementia | 3.1 | 2.3 | 4 | 2.8 | 1.4 | | 3.3 | 0.2 | 1.7 | 0.2 | 3.8 | 0.8 | 1.9 | 2.5 | 1.3 | 1.8 | 7.6 | 14.1 | 1.1 | 3.9 | 2.9 | 2.2 | 1.2 | 6.2 | 1.7 |
| Depression | 50.1 | 73.7 | 82.4 | 26 | 13.7 | 54.4 | | 29.8 | 58 | 51.6 | 27.5 | 28.9 | 20.4 | 24.5 | 31.8 | 23.3 | 32.5 | 41.3 | 75.5 | 34.3 | 59.3 | 40.9 | 59.5 | 35.3 | 22.6 |
| Developmental and behavioural disorders | 6.2 | 10 | 10.7 | 1.8 | 13 | 0.8 | 6 | | 24.2 | 12.6 | 8.9 | 2.8 | 4.7 | 41 | 1.5 | 4.1 | 2.6 | 1.1 | 19.1 | 1.3 | 14.9 | 5.8 | 12.5 | 0.8 | 6.2 |
| Drug abuse | 11.9 | 6.1 | 12.6 | 1.1 | 1.1 | 2 | 4.2 | 8.8 | | 5.1 | 3.8 | 1.6 | 2.1 | 4.9 | 1 | 1.4 | 1.1 | 1.3 | 18.2 | 2.1 | 22.6 | 3.2 | 9 | 1.4 | 3.9 |
| Eating disorders | 0.6 | 1.8 | 1.7 | 0.2 | 0.3 | 0.1 | 1.2 | 1.5 | 1.6 | | 0.6 | 0.6 | 0.6 | 0.8 | 0.3 | 0.4 | 0.2 | 0.1 | 5.4 | 0.3 | 2 | 0.6 | 2 | 0.1 | 0.6 |
| Epilepsy | 4.5 | 3.1 | 4 | 18 | 29.6 | 6.2 | 2.7 | 4.4 | 5.1 | 2.5 | | 2 | 8.5 | 20.8 | 3.5 | 4.1 | 4.9 | 3.8 | 4.1 | 3.7 | 5.6 | 2.5 | 3.3 | 8.2 | 4.2 |
| Headache | 9.1 | 17.2 | 16.3 | 16.1 | 5.7 | 7.6 | 17.1 | 8.2 | 12.9 | 14.3 | 12.2 | | 14.5 | 5.2 | 16.2 | 12.5 | 10.7 | 13.4 | 17.7 | 12.7 | 10.7 | 16 | 16.4 | 12.3 | 11.5 |
| Infections of the CNS | 0.5 | 0.5 | 0.5 | 2.4 | 2.9 | 0.8 | 0.5 | 0.6 | 0.7 | 0.6 | 2.2 | 0.6 | | 1.1 | 1.7 | 1.1 | 1.3 | 0.7 | 0.5 | 1.2 | 0.5 | 0.5 | 0.6 | 1.2 | 0.7 |
| Intellectual disability | 1.2 | 1.6 | 2.4 | 0.9 | 23.8 | 1.6 | 1 | 8 | 2.7 | 1.3 | 8.3 | 0.3 | 1.6 | | 0.3 | 2.6 | 2.2 | 0.7 | 2.6 | 0.5 | 5.9 | 0.9 | 2.2 | 0.5 | 1.1 |
| Multiple sclerosis | 0.3 | 0.5 | 0.5 | 0.9 | 0.6 | 0.5 | 0.7 | 0.2 | 0.3 | 0.3 | 0.8 | 0.6 | 1.4 | 0.2 | | 1 | 3.5 | 1 | 0.4 | 1.3 | 0.4 | 0.8 | 0.4 | 0.7 | 0.3 |
| Neuromuscular disorders | 0.3 | 0.2 | 0.3 | 0.4 | 1.5 | 0.3 | 0.2 | 0.2 | 0.2 | 0.2 | 0.5 | 0.2 | 0.5 | 0.7 | 0.5 | | 8.8 | 0.5 | 0.2 | 1.9 | 0.2 | 0.3 | 0.2 | 0.4 | 0.2 |
| Other neurodegenerative disorders | 0.2 | 0.1 | 0.2 | 0.3 | 1.8 | 0.6 | 0.2 | 0.1 | 0.1 | 0 | 0.2 | 0.1 | 0.3 | 0.3 | 0.8 | 4.1 | | 1 | 0.1 | 1.1 | 0.1 | 0.2 | 0.1 | 0.2 | 0.1 |
| Parkinson's disease | 0.7 | 1 | 1.3 | 1.5 | 1.8 | 7.2 | 1.3 | 0.2 | 0.6 | 0.2 | 1.2 | 0.7 | 0.8 | 0.6 | 1.4 | 1.6 | 5.9 | | 0.5 | 2.9 | 0.8 | 1.2 | 0.6 | 1.8 | 0.6 |
| Personality disorders | 9.1 | 11.9 | 21.2 | 1.4 | 1.3 | 1.8 | 7.5 | 9.4 | 24.9 | 22.8 | 4.1 | 3 | 2 | 6.5 | 2 | 2.3 | 1.9 | 1.6 | | 2.2 | 22 | 4.6 | 15.6 | 1.5 | 3.8 |
| Polyneuropathy | 3.4 | 1.4 | 1.8 | 2.2 | 1.5 | 3.3 | 1.7 | 0.3 | 1.4 | 0.6 | 1.9 | 1.1 | 2.4 | 0.6 | 3 | 9.1 | 11.5 | 4.8 | 1.1 | | 1.2 | 1.9 | 1.2 | 3.2 | 1.1 |
| Schizophrenia spectrum disorders | 8.4 | 7.3 | 21.4 | 1.6 | 2.3 | 4.2 | 5.1 | 6.4 | 26.8 | 7.3 | 4.9 | 1.6 | 1.8 | 13 | 1.6 | 1.6 | 2.2 | 2.3 | 19.1 | 2 | | 3.9 | 9.9 | 1.8 | 2.8 |
| Sleep disorders | 36.8 | 37.8 | 57.6 | 29.6 | 17.8 | 34.7 | 38.4 | 26.8 | 41.5 | 24.1 | 24.1 | 25.4 | 19.6 | 22.4 | 37.3 | 25.7 | 31.2 | 38 | 43.1 | 34.6 | 42.6 | | 36.9 | 30.4 | 18.1 |
| Stress related disorders | 19.1 | 22 | 32.5 | 5.6 | 6.5 | 5.9 | 17.6 | 18.3 | 36.5 | 24.7 | 9.8 | 8.2 | 6.9 | 16.5 | 6.2 | 6.2 | 7.2 | 5.7 | 46.3 | 7 | 34 | 11.6 | | 5.6 | 9.6 |
| Stroke | 7.1 | 4.6 | 5 | 11 | 6.5 | 17.3 | 5.9 | 0.7 | 3.3 | 0.7 | 13.8 | 3.5 | 7.7 | 2.1 | 5.7 | 5.7 | 5.9 | 9.9 | 2.5 | 10.6 | 3.5 | 5.4 | 3.1 | | 4.7 |
| Traumatic brain injury | 14.4 | 7.8 | 9.7 | 6.5 | 10.2 | 9.4 | 7.5 | 10.1 | 17.9 | 8.9 | 14.3 | 6.4 | 8.7 | 9.5 | 5.1 | 7.2 | 8 | 6.1 | 12.6 | 7.2 | 10.8 | 6.4 | 10.7 | 9.3 | |
| Myocardial infarction | 2.3 | 1.6 | 1.6 | 2 | 0.4 | 3.9 | 2 | 0.2 | 1.1 | 0.2 | 1.6 | 1.1 | 1.2 | 0.3 | 1.2 | 2.6 | 2 | 3.2 | 0.9 | 4 | 1.1 | 2.4 | 1.3 | 4.7 | 1.1 |
| Congestive heart failure | 3.1 | 1.9 | 2 | 2.6 | 0.7 | 6.5 | 2.3 | 0.2 | 1.3 | 0.3 | 2.2 | 1 | 1.7 | 0.8 | 1.2 | 4.8 | 3.1 | 4.4 | 0.9 | 6.2 | 1.6 | 3.1 | 1.2 | 7.2 | 1.4 |
| Peripheral vascular disease | 4.4 | 2.2 | 2.2 | 3.3 | 0.9 | 6.2 | 2.9 | 0.3 | 1.7 | 0.8 | 2.7 | 1.7 | 2.2 | 0.6 | 2.1 | 3.7 | 4.6 | 5.3 | 1.2 | 9.7 | 1.4 | 3.6 | 1.5 | 9.1 | 1.6 |
| Cerebrovascular disease | 8 | 5.4 | 6.1 | 12.7 | 7.4 | 23.1 | 6.8 | 0.8 | 3.7 | 0.9 | 15.1 | 4.2 | 7.8 | 2.3 | 5.8 | 7.1 | 8.2 | 13.2 | 2.8 | 13.3 | 3.8 | 6.5 | 3.7 | 77.9 | 5.2 |
| Chronic pulmonary disease | 10.6 | 7.8 | 8.6 | 7 | 8.7 | 11.2 | 7.8 | 5.4 | 8.8 | 4.3 | 7.6 | 6.2 | 7 | 6.2 | 4.7 | 16.7 | 19.7 | 9.3 | 7.7 | 12.5 | 7.6 | 8.6 | 7.1 | 11.1 | 5.9 |
| Connective tissue disease | 1.8 | 2.4 | 2.4 | 3.4 | 1.8 | 4 | 2.8 | 1.2 | 1.3 | 1.7 | 2.3 | 2.9 | 3.7 | 1.7 | 2.5 | 9.9 | 3.2 | 3.9 | 2 | 6.9 | 1.5 | 3.2 | 2.1 | 4.1 | 1.7 |
| Ulcer disease | 3.5 | 1.6 | 1.9 | 1.4 | 1.2 | 4.2 | 1.8 | 0.4 | 2.1 | 0.4 | 1.7 | 1.1 | 1.4 | 0.9 | 1.2 | 2.2 | 1.8 | 3 | 1.4 | 3.6 | 1.7 | 1.8 | 1.4 | 3.6 | 1.2 |
| Mild liver disease | 7.3 | 1.6 | 2.1 | 0.9 | 0.6 | 1.7 | 1.6 | 0.8 | 7.7 | 0.8 | 1.8 | 0.9 | 1.5 | 0.6 | 0.7 | 2.1 | 1.3 | 0.9 | 2.4 | 3.4 | 3 | 1.5 | 1.9 | 1.6 | 1.4 |
| Diabetes without end−organ damage | 7 | 5 | 6.8 | 6.4 | 2 | 11.7 | 5.8 | 1.3 | 4.2 | 1.6 | 5 | 3.4 | 4.7 | 3.4 | 3.4 | 7.3 | 6.1 | 8.8 | 4.6 | 17.8 | 6.5 | 6.4 | 4.4 | 12.7 | 3.2 |
| Hemiplegia | 0.4 | 0.3 | 0.3 | 2.6 | 24.7 | 0.5 | 0.4 | 0.3 | 0.3 | 0.1 | 2.4 | 0.2 | 2.4 | 2.9 | 2.4 | 3.1 | 7.6 | 0.9 | 0.2 | 1.2 | 0.3 | 0.4 | 0.3 | 1.2 | 0.5 |
| Moderate to severe renal disease | 2.4 | 1.6 | 3.2 | 2.3 | 1.3 | 4.7 | 2 | 0.5 | 1.7 | 0.9 | 2.3 | 1.2 | 2.7 | 1.3 | 1.2 | 4.4 | 2.4 | 3.7 | 1.2 | 6.4 | 1.6 | 2.5 | 1.3 | 5.4 | 1.3 |
| Diabetes with end−organ damage | 3.2 | 2.1 | 2.7 | 2.4 | 0.9 | 5.6 | 2.6 | 0.4 | 1.7 | 0.7 | 2.5 | 1.3 | 2.1 | 1.3 | 1.6 | 3.4 | 2.9 | 4.4 | 1.8 | 12.8 | 2.5 | 2.9 | 1.8 | 7.5 | 1.4 |
| Solid cancer tumour | 7.2 | 5.3 | 5.9 | 23.7 | 2.1 | 13.3 | 6.4 | 0.7 | 2.5 | 1.2 | 6.2 | 5.8 | 5.1 | 1.5 | 5.1 | 7.4 | 7.2 | 12.4 | 2.5 | 13.4 | 3.2 | 8.9 | 3.6 | 12.3 | 3.5 |
| Leukaemia | 0.2 | 0.2 | 0.2 | 0.6 | 0.1 | 0.4 | 0.2 | 0 | 0.1 | 0 | 0.2 | 0.2 | 0.4 | 0.1 | 0.1 | 0.4 | 0.5 | 0.3 | 0.1 | 0.7 | 0.1 | 0.3 | 0.2 | 0.4 | 0.1 |
| Lymphoma | 0.4 | 0.4 | 0.4 | 2 | 0.1 | 0.7 | 0.4 | 0.1 | 0.2 | 0.1 | 0.4 | 0.3 | 0.8 | 0.1 | 0.3 | 0.7 | 0.6 | 0.8 | 0.2 | 1.7 | 0.3 | 0.7 | 0.3 | 0.8 | 0.3 |
| Moderate to severe liver disease | 2.6 | 0.4 | 0.4 | 0.3 | 0.3 | 0.7 | 0.4 | 0.1 | 0.8 | 0.2 | 0.5 | 0.1 | 0.4 | 0.1 | 0.2 | 0.9 | 0.6 | 0.2 | 0.4 | 1.4 | 0.4 | 0.4 | 0.4 | 0.5 | 0.3 |
| Metastatic solid tumour | 0.8 | 0.6 | 0.5 | 3 | 0.2 | 0.7 | 0.7 | 0.1 | 0.3 | 0.1 | 0.7 | 0.6 | 0.6 | 0.2 | 0.5 | 0.6 | 0.7 | 1 | 0.2 | 1.5 | 0.3 | 1 | 0.4 | 1 | 0.3 |
| AIDS | 0.3 | 0.1 | 0.2 | 0.1 | 0.1 | 0.1 | 0.2 | 0.1 | 1 | 0.1 | 0.2 | 0.1 | 1 | 0.1 | 0 | 0.1 | 0 | 0.1 | 0.2 | 0.4 | 0.4 | 0.2 | 0.2 | 0.1 | 0.1 |

Percent: 0, 20, 40, 60, 80

### Concurrent diseases

#### Main analysis − Prevalent cohort 2018

| | Alcohol abuse | Anxiety disorders | Bipolar disorder | Brain tumors | Cerebral palsy | Dementia | Depression | Developmental and behavioural disorders | Drug abuse | Eating disorders | Epilepsy | Headache | Infections of the CNS | Intellectual disability | Multiple sclerosis | Neuromuscular disorders | Other neurodegenerative disorders | Parkinson's disease | Personality disorders | Polyneuropathy | Schizophrenia spectrum disorders | Sleep disorders | Stress related disorders | Stroke | Traumatic brain injury |
|---|---|---|---|---|---|---|---|---|---|---|---|---|---|---|---|---|---|---|---|---|---|---|---|---|---|
| Alcohol abuse | | 12.3 | 23.4 | 4 | 2.5 | 11.6 | 11.2 | 6.9 | 36.2 | 5.6 | 10.2 | 3.4 | 4.9 | 6.7 | 3.7 | 5.3 | 7.2 | 5.2 | 20 | 14.8 | 21.4 | 8.7 | 14.2 | 9.6 | 9.7 |
| Anxiety disorders | 21.5 | | 36.3 | 9.1 | 6.8 | 14.1 | 28.1 | 19.4 | 32.2 | 29.1 | 12 | 11.3 | 7.7 | 15.6 | 9.3 | 8.7 | 10.5 | 12.1 | 46 | 10.5 | 32.7 | 15.6 | 28.7 | 10.3 | 9.2 |
| Bipolar disorder | 5 | 4.5 | | 1.2 | 0.9 | 3 | 3.9 | 2.6 | 8.2 | 3.6 | 1.9 | 1.3 | 1.1 | 2.8 | 1.1 | 1.3 | 1.5 | 2.1 | 10 | 1.7 | 11.5 | 2.9 | 5.2 | 1.4 | 1.4 |
| Brain tumors | 0.4 | 0.5 | 0.6 | | 1.1 | 1.1 | 0.6 | 0.2 | 0.3 | 0.2 | 4.2 | 0.6 | 2.1 | 0.5 | 1 | 0.8 | 1.1 | 1.2 | 0.3 | 1 | 0.4 | 0.7 | 0.4 | 1.5 | 0.5 |
| Cerebral palsy | 0.1 | 0.2 | 0.2 | 0.6 | | 0.3 | 0.2 | 0.8 | 0.2 | 0.2 | 3.8 | 0.1 | 1.5 | 7.5 | 0.4 | 1.8 | 4.4 | 0.8 | 0.2 | 0.4 | 0.3 | 0.2 | 0.3 | 0.5 | 0.4 |
| Dementia | 3.1 | 2.2 | 3.8 | 2.8 | 1.3 | | 3.1 | 0.2 | 1.6 | 0.1 | 3.7 | 0.8 | 1.8 | 2.4 | 1.3 | 1.8 | 7.4 | 15.2 | 1 | 3.8 | 2.8 | 2.2 | 1.2 | 6.1 | 1.7 |
| Depression | 50.8 | 73 | 82.5 | 26.5 | 13.8 | 53 | | 30.1 | 57.8 | 51.4 | 27.6 | 29.2 | 20.7 | 24.8 | 31.8 | 23.8 | 33.2 | 41.7 | 75.9 | 34.5 | 59.5 | 41.1 | 59.4 | 35.1 | 22.9 |
| Developmental and behavioural disorders | 6.7 | 10.8 | 11.8 | 1.8 | 13.1 | 0.8 | 6.4 | | 25.6 | 13.6 | 9.2 | 3.1 | 4.9 | 42.3 | 1.6 | 4.3 | 2.8 | 1.3 | 20.4 | 1.4 | 16.1 | 6.4 | 13.5 | 0.9 | 6.5 |
| Drug abuse | 12.1 | 6.2 | 12.7 | 1.1 | 1.1 | 2 | 4.3 | 8.9 | | 5.2 | 3.8 | 1.6 | 2.2 | 5.1 | 1 | 1.5 | 1 | 1.3 | 18.2 | 2 | 22.8 | 3.2 | 9.1 | 1.4 | 4 |
| Eating disorders | 0.6 | 1.8 | 1.8 | 0.2 | 0.4 | 0.1 | 1.2 | 1.5 | 1.7 | | 0.6 | 0.6 | 0.6 | 0.8 | 0.3 | 0.4 | 0.2 | 0.1 | 5.4 | 0.3 | 2 | 0.6 | 2 | 0.1 | 0.7 |
| Epilepsy | 4.5 | 3 | 3.8 | 17.6 | 29.5 | 6.1 | 2.7 | 4.2 | 5 | 2.5 | | 2 | 8.4 | 20.4 | 3.4 | 4 | 4.7 | 3.7 | 4 | 3.6 | 5.5 | 2.5 | 3.2 | 8.1 | 4.2 |
| Headache | 9.1 | 17.4 | 16.5 | 16.4 | 5.9 | 7.6 | 17.3 | 8.4 | 13 | 14.5 | 12.3 | | 15 | 5.4 | 16.3 | 12.4 | 11.1 | 13.4 | 18.1 | 12.7 | 11 | 16.1 | 16.6 | 12.4 | 11.8 |
| Infections of the CNS | 0.6 | 0.5 | 0.6 | 2.4 | 3 | 0.8 | 0.5 | 0.6 | 0.7 | 0.6 | 2.2 | 0.6 | | 1 | 1.7 | 1 | 1.3 | 0.6 | 0.5 | 1.1 | 0.5 | 0.5 | 0.6 | 1.2 | 0.7 |
| Intellectual disability | 1.2 | 1.6 | 2.4 | 0.8 | 23.7 | 1.6 | 1 | 7.8 | 2.8 | 1.3 | 8.4 | 0.4 | 1.6 | | 0.3 | 2.6 | 2.2 | 0.7 | 2.6 | 0.4 | 5.9 | 1 | 2.2 | 0.5 | 1.1 |
| Multiple sclerosis | 0.4 | 0.5 | 0.5 | 0.9 | 0.6 | 0.5 | 0.7 | 0.2 | 0.3 | 0.3 | 0.8 | 0.6 | 1.5 | 0.2 | | 1 | 3.4 | 0.9 | 0.4 | 1.2 | 0.4 | 0.9 | 0.4 | 0.7 | 0.3 |
| Neuromuscular disorders | 0.3 | 0.2 | 0.3 | 0.4 | 1.6 | 0.3 | 0.3 | 0.2 | 0.2 | 0.2 | 0.4 | 0.2 | 0.4 | 0.7 | 0.5 | | 8.7 | 0.6 | 0.2 | 1.9 | 0.2 | 0.3 | 0.2 | 0.4 | 0.2 |
| Other neurodegenerative disorders | 0.2 | 0.1 | 0.2 | 0.3 | 1.8 | 0.6 | 0.2 | 0.1 | 0.1 | 0 | 0.2 | 0.1 | 0.3 | 0.3 | 0.8 | 4.1 | | 1.1 | 0.1 | 1.2 | 0.1 | 0.2 | 0.1 | 0.2 | 0.1 |
| Parkinson's disease | 0.7 | 0.9 | 1.3 | 1.5 | 1.8 | 7.6 | 1.2 | 0.2 | 0.5 | 0.2 | 1.1 | 0.7 | 0.7 | 0.5 | 1.3 | 1.6 | 6 | | 0.5 | 2.9 | 0.8 | 1.2 | 0.6 | 1.8 | 0.6 |
| Personality disorders | 9 | 11.8 | 21 | 1.4 | 1.3 | 1.7 | 7.5 | 9.4 | 24.4 | 22.4 | 4.1 | 3 | 2.1 | 6.4 | 2 | 2.3 | 1.9 | 1.6 | | 2.1 | 21.5 | 4.5 | 15.3 | 1.5 | 3.8 |
| Polyneuropathy | 3.5 | 1.4 | 1.9 | 2.2 | 1.6 | 3.4 | 1.8 | 0.3 | 1.4 | 0.6 | 1.9 | 1.1 | 2.4 | 0.6 | 3 | 9.2 | 11.8 | 5 | 1.1 | | 1.1 | 1.9 | 1.2 | 3.3 | 1.2 |
| Schizophrenia spectrum disorders | 8.4 | 7.4 | 21.1 | 1.6 | 2.3 | 4.1 | 5.2 | 6.5 | 26.9 | 7.4 | 4.9 | 1.6 | 1.9 | 13 | 1.6 | 1.7 | 2.2 | 2.4 | 18.9 | 1.9 | | 3.9 | 10 | 1.8 | 2.9 |
| Sleep disorders | 37.3 | 38.5 | 58 | 30.4 | 18.7 | 34.8 | 39.1 | 28.3 | 41.8 | 25.2 | 24.8 | 25.9 | 20.3 | 23.5 | 38.1 | 26.2 | 31 | 39.1 | 43.7 | 34.9 | 43.1 | | 37.5 | 30.7 | 18.7 |
| Stress related disorders | 19.6 | 22.7 | 33.2 | 5.9 | 7 | 6 | 18.1 | 19.2 | 37.4 | 25.1 | 10.1 | 8.5 | 7.1 | 17 | 6.4 | 6.6 | 7.2 | 5.9 | 47 | 7.2 | 35 | 12 | | 5.7 | 9.9 |
| Stroke | 7.2 | 4.4 | 4.9 | 10.8 | 6.7 | 17.1 | 5.8 | 0.7 | 3.2 | 0.7 | 13.9 | 3.5 | 7.7 | 2.1 | 5.5 | 5.6 | 6 | 10 | 2.5 | 10.6 | 3.4 | 5.4 | 3.1 | | 4.8 |
| Traumatic brain injury | 14.1 | 7.7 | 9.6 | 6.5 | 9.9 | 9.4 | 7.4 | 9.8 | 17.4 | 8.8 | 14 | 6.4 | 8.8 | 9.2 | 4.9 | 7 | 8.1 | 6.4 | 12.3 | 7.2 | 10.6 | 6.3 | 10.5 | 9.3 | |
| Myocardial infarction | 2.3 | 1.6 | 1.6 | 2 | 0.4 | 3.8 | 1.9 | 0.2 | 1.1 | 0.2 | 1.6 | 1.1 | 1.2 | 0.3 | 1.2 | 2.5 | 2 | 3.1 | 0.9 | 4 | 1.1 | 2.4 | 1.3 | 4.5 | 1.1 |
| Congestive heart failure | 3.1 | 1.8 | 2 | 2.6 | 0.7 | 6.3 | 2.2 | 0.2 | 1.3 | 0.3 | 2.2 | 1 | 1.6 | 0.7 | 1.2 | 4.6 | 2.9 | 4.4 | 0.9 | 6.2 | 1.5 | 3 | 1.1 | 7.1 | 1.4 |
| Peripheral vascular disease | 4.6 | 2.2 | 2.2 | 3.3 | 1 | 6.1 | 3 | 0.4 | 1.6 | 0.9 | 2.8 | 1.7 | 2.1 | 0.6 | 2 | 3.7 | 4.1 | 5.4 | 1.2 | 9.9 | 1.4 | 3.6 | 1.5 | 9.1 | 1.7 |
| Cerebrovascular disease | 8.1 | 5.2 | 6 | 12.5 | 7.6 | 22.7 | 6.8 | 0.8 | 3.6 | 0.9 | 15.1 | 4.2 | 8 | 2.3 | 5.6 | 6.8 | 8 | 13.3 | 2.7 | 13.3 | 3.7 | 6.4 | 3.6 | 77.8 | 5.3 |
| Chronic pulmonary disease | 11 | 7.9 | 8.7 | 7.3 | 8.8 | 10.9 | 8 | 5.4 | 8.8 | 4.3 | 7.7 | 6.4 | 7.2 | 6.3 | 5 | 16.9 | 20 | 9.4 | 7.8 | 12.8 | 7.7 | 8.7 | 7.3 | 11.2 | 6.1 |
| Connective tissue disease | 1.9 | 2.4 | 2.4 | 3.4 | 1.8 | 3.9 | 2.8 | 1.3 | 1.3 | 1.8 | 2.4 | 3 | 3.7 | 1.6 | 2.5 | 10 | 3.4 | 4.1 | 2 | 7 | 1.5 | 3.3 | 2.2 | 4.2 | 1.8 |
| Ulcer disease | 3.4 | 1.5 | 1.9 | 1.4 | 1.3 | 3.9 | 1.7 | 0.4 | 2 | 0.4 | 1.7 | 1.1 | 1.4 | 0.9 | 1.1 | 2.1 | 1.9 | 2.7 | 1.4 | 3.5 | 1.6 | 1.8 | 1.3 | 3.4 | 1.1 |
| Mild liver disease | 7.4 | 1.6 | 2.1 | 1 | 0.5 | 1.6 | 1.6 | 0.8 | 7.4 | 0.8 | 1.8 | 0.9 | 1.4 | 0.6 | 0.7 | 2 | 1.4 | 0.9 | 2.3 | 3.5 | 3 | 1.5 | 1.9 | 1.6 | 1.4 |
| Diabetes without end−organ damage | 7.1 | 5 | 6.9 | 6.4 | 2 | 11.9 | 5.9 | 1.4 | 4.1 | 1.7 | 5.1 | 3.4 | 4.6 | 3.4 | 3.4 | 7.2 | 6.4 | 9 | 4.5 | 18.1 | 6.5 | 6.5 | 4.4 | 12.7 | 3.2 |
| Hemiplegia | 0.4 | 0.3 | 0.3 | 2.5 | 24.5 | 0.5 | 0.4 | 0.3 | 0.3 | 0.1 | 2.4 | 0.2 | 2.5 | 2.8 | 2.4 | 3 | 7.9 | 1 | 0.2 | 1.2 | 0.3 | 0.4 | 0.3 | 1.3 | 0.5 |
| Moderate to severe renal disease | 2.6 | 1.6 | 3.1 | 2.5 | 1.4 | 4.9 | 2.1 | 0.6 | 1.7 | 0.9 | 2.4 | 1.3 | 2.7 | 1.3 | 1.3 | 4.1 | 2.3 | 3.9 | 1.2 | 6.7 | 1.6 | 2.6 | 1.4 | 5.6 | 1.4 |
| Diabetes with end−organ damage | 3.3 | 2.1 | 2.7 | 2.5 | 0.9 | 5.6 | 2.6 | 0.5 | 1.8 | 0.8 | 2.5 | 1.3 | 2 | 1.3 | 1.5 | 3.4 | 3 | 4.4 | 1.8 | 13.1 | 2.5 | 2.9 | 1.8 | 7.4 | 1.5 |
| Solid cancer tumour | 7.5 | 5.4 | 6 | 23.1 | 2.2 | 13.6 | 6.6 | 0.7 | 2.5 | 1.3 | 6.4 | 5.8 | 5.2 | 1.5 | 5.4 | 7.6 | 7.2 | 13.2 | 2.6 | 13.8 | 3.2 | 9 | 3.6 | 12.7 | 3.7 |
| Leukaemia | 0.2 | 0.2 | 0.2 | 0.6 | 0.1 | 0.4 | 0.2 | 0.1 | 0.1 | 0 | 0.2 | 0.2 | 0.4 | 0.1 | 0.1 | 0.3 | 0.4 | 0.3 | 0.1 | 0.7 | 0.1 | 0.3 | 0.2 | 0.4 | 0.1 |
| Lymphoma | 0.4 | 0.4 | 0.4 | 2 | 0.1 | 0.8 | 0.5 | 0.1 | 0.2 | 0.1 | 0.4 | 0.4 | 0.8 | 0.1 | 0.3 | 0.7 | 0.7 | 0.9 | 0.2 | 1.8 | 0.3 | 0.7 | 0.3 | 0.8 | 0.3 |
| Moderate to severe liver disease | 2.7 | 0.4 | 0.4 | 0.3 | 0.3 | 0.6 | 0.4 | 0.1 | 0.8 | 0.1 | 0.5 | 0.1 | 0.4 | 0.1 | 0.2 | 0.8 | 0.6 | 0.2 | 0.4 | 1.4 | 0.5 | 0.4 | 0.4 | 0.5 | 0.3 |
| Metastatic solid tumour | 0.8 | 0.6 | 0.5 | 3 | 0.2 | 0.8 | 0.6 | 0.1 | 0.2 | 0.1 | 0.7 | 0.5 | 0.6 | 0.2 | 0.5 | 0.8 | 0.6 | 0.9 | 0.2 | 1.5 | 0.3 | 0.9 | 0.4 | 1 | 0.3 |
| AIDS | 0.3 | 0.1 | 0.2 | 0.1 | 0.1 | 0.1 | 0.2 | 0.1 | 0.9 | 0.1 | 0.2 | 0.1 | 1 | 0.1 | 0 | 0.2 | 0 | 0.1 | 0.2 | 0.4 | 0.4 | 0.2 | 0.2 | 0.1 | 0.1 |

Percent: 0, 20, 40, 60, 80

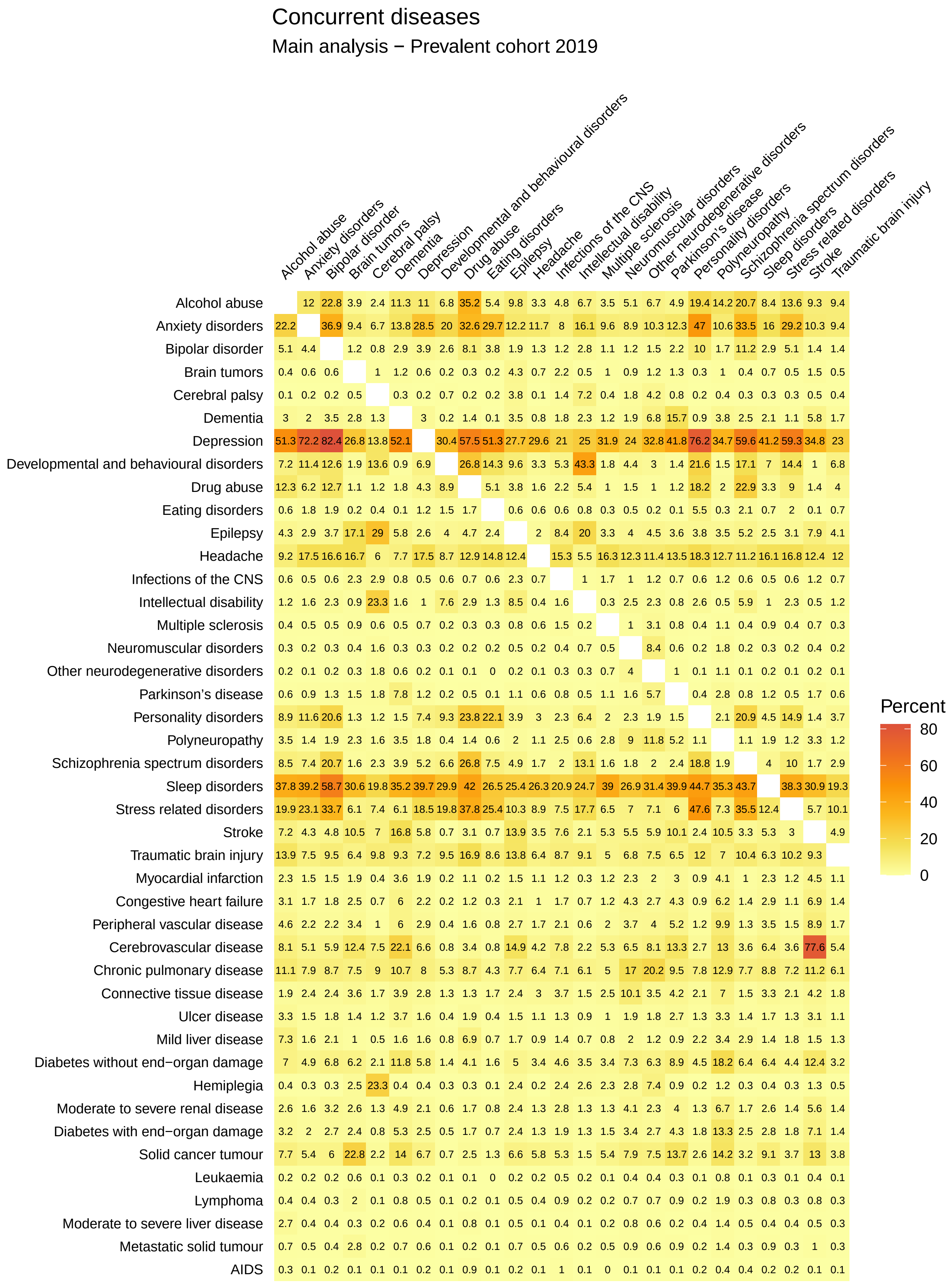
Concurrent diseases
Main analysis − Prevalent cohort 2019
Alcohol abuse
Anxiety disorders
Bipolar disorder
Brain tumors
Cerebral palsy
Dementia
Depression
Developmental and behavioural disorders
Drug abuse
Eating disorders
Epilepsy
Headache
Infections of the CNS
Intellectual disability
Multiple sclerosis
Neuromuscular disorders
Other neurodegenerative disorders
Parkinson's disease
Personality disorders
Polyneuropathy
Schizophrenia spectrum disorders
Sleep disorders
Stress related disorders
Stroke
Traumatic brain injury
Myocardial infarction
Congestive heart failure
Peripheral vascular disease
Cerebrovascular disease
Chronic pulmonary disease
Connective tissue disease
Ulcer disease
Mild liver disease
Diabetes without end−organ damage
Hemiplegia
Moderate to severe renal disease
Diabetes with end−organ damage
Solid cancer tumour
Leukaemia
Lymphoma
Moderate to severe liver disease
Metastatic solid tumour
AIDS
Percent
80
60
40
20
0

#### Concurrent diseases

##### Main analysis − Prevalent cohort 2020

| | Alcohol abuse | Anxiety disorders | Bipolar disorder | Brain tumors | Cerebral palsy | Dementia | Depression | Developmental and behavioural disorders | Drug abuse | Eating disorders | Epilepsy | Headache | Infections of the CNS | Intellectual disability | Multiple sclerosis | Neuromuscular disorders | Other neurodegenerative disorders | Parkinson's disease | Personality disorders | Polyneuropathy | Schizophrenia spectrum disorders | Sleep disorders | Stress related disorders | Stroke | Traumatic brain injury |
|---|---|---|---|---|---|---|---|---|---|---|---|---|---|---|---|---|---|---|---|---|---|---|---|---|---|
| Alcohol abuse | | 11.7 | 22.2 | 3.8 | 2.3 | 11 | 10.8 | 6.8 | 34.3 | 5.3 | 9.5 | 3.4 | 4.8 | 6.7 | 3.6 | 5.1 | 6.1 | 5.2 | 18.8 | 13.6 | 20 | 8.2 | 13.2 | 9.1 | 9.2 |
| Anxiety disorders | 22.7 | | 37.4 | 9.3 | 6.8 | 13.3 | 28.9 | 20.8 | 33.1 | 30.3 | 12.4 | 12.1 | 8.3 | 16.7 | 9.6 | 9.1 | 10.1 | 12.5 | 48.1 | 10.6 | 34.4 | 16.5 | 29.8 | 10.2 | 9.7 |
| Bipolar disorder | 5.1 | 4.4 | | 1.2 | 0.9 | 2.8 | 3.9 | 2.7 | 8.1 | 3.9 | 1.8 | 1.4 | 1.2 | 2.7 | 1.1 | 1.2 | 1.5 | 2.2 | 10 | 1.6 | 10.9 | 2.9 | 5.1 | 1.4 | 1.4 |
| Brain tumors | 0.4 | 0.6 | 0.6 | | 1 | 1.2 | 0.6 | 0.2 | 0.3 | 0.2 | 4.3 | 0.7 | 2.2 | 0.5 | 1.1 | 0.9 | 1.2 | 1.3 | 0.3 | 1.1 | 0.4 | 0.8 | 0.5 | 1.5 | 0.5 |
| Cerebral palsy | 0.1 | 0.2 | 0.2 | 0.5 | | 0.3 | 0.2 | 0.7 | 0.2 | 0.2 | 3.8 | 0.1 | 1.4 | 6.9 | 0.3 | 1.8 | 4.3 | 0.7 | 0.1 | 0.4 | 0.3 | 0.3 | 0.3 | 0.5 | 0.4 |
| Dementia | 3 | 1.9 | 3.3 | 2.7 | 1.2 | | 2.9 | 0.2 | 1.3 | 0.1 | 3.4 | 0.7 | 1.8 | 2.2 | 1.2 | 1.7 | 6.2 | 16.2 | 0.9 | 3.7 | 2.4 | 2 | 1.1 | 5.6 | 1.7 |
| Depression | 51.9 | 71.6 | 82.4 | 26.8 | 13.9 | 51.3 | | 31.1 | 57.5 | 50.8 | 27.8 | 29.9 | 21.3 | 25.4 | 31.8 | 24.4 | 32.8 | 41.8 | 76.6 | 34.5 | 59.8 | 41.4 | 59.3 | 34.5 | 23.1 |
| Developmental and behavioural disorders | 7.8 | 12.3 | 13.7 | 2 | 13.9 | 0.9 | 7.4 | | 28.3 | 15.2 | 9.9 | 3.6 | 5.4 | 44.3 | 2 | 4.7 | 3.2 | 1.5 | 22.9 | 1.6 | 18.4 | 7.7 | 15.4 | 1 | 7.2 |
| Drug abuse | 12.4 | 6.2 | 12.8 | 1 | 1.2 | 1.8 | 4.3 | 8.9 | | 5.1 | 3.8 | 1.7 | 2.1 | 5.7 | 1.1 | 1.4 | 0.9 | 1.2 | 18.3 | 1.9 | 23 | 3.3 | 9.1 | 1.4 | 4.1 |
| Eating disorders | 0.6 | 1.8 | 2 | 0.2 | 0.4 | 0.1 | 1.2 | 1.6 | 1.7 | | 0.6 | 0.6 | 0.6 | 0.8 | 0.3 | 0.5 | 0.2 | 0.1 | 5.5 | 0.3 | 2.1 | 0.7 | 2 | 0.1 | 0.7 |
| Epilepsy | 4.2 | 2.8 | 3.6 | 16.4 | 28.7 | 5.6 | 2.5 | 3.8 | 4.6 | 2.3 | | 1.9 | 8.2 | 19.6 | 3.2 | 4 | 4.4 | 3.6 | 3.6 | 3.4 | 5.1 | 2.5 | 3 | 7.7 | 4.1 |
| Headache | 9.6 | 17.5 | 16.7 | 16.5 | 5.9 | 7.7 | 17.5 | 8.9 | 12.8 | 14.9 | 12.3 | | 15.7 | 5.7 | 16.2 | 12.4 | 11.4 | 13.2 | 18.5 | 12.6 | 11.2 | 16 | 16.9 | 12.3 | 12.2 |
| Infections of the CNS | 0.6 | 0.5 | 0.6 | 2.2 | 2.9 | 0.8 | 0.5 | 0.6 | 0.7 | 0.6 | 2.2 | 0.7 | | 1 | 1.8 | 1 | 1.2 | 0.7 | 0.6 | 1.2 | 0.6 | 0.6 | 0.6 | 1.1 | 0.7 |
| Intellectual disability | 1.3 | 1.6 | 2.2 | 0.8 | 22.8 | 1.6 | 1 | 7.3 | 3 | 1.3 | 8.5 | 0.4 | 1.6 | | 0.3 | 2.4 | 2.2 | 0.7 | 2.6 | 0.5 | 5.9 | 1.1 | 2.3 | 0.5 | 1.2 |
| Multiple sclerosis | 0.4 | 0.5 | 0.5 | 1 | 0.6 | 0.5 | 0.7 | 0.2 | 0.3 | 0.3 | 0.8 | 0.6 | 1.5 | 0.2 | | 1 | 2.9 | 0.9 | 0.4 | 1.1 | 0.4 | 0.9 | 0.4 | 0.7 | 0.3 |
| Neuromuscular disorders | 0.3 | 0.2 | 0.3 | 0.4 | 1.6 | 0.3 | 0.3 | 0.2 | 0.2 | 0.2 | 0.5 | 0.2 | 0.4 | 0.6 | 0.5 | | 8.3 | 0.6 | 0.2 | 1.8 | 0.2 | 0.3 | 0.2 | 0.4 | 0.2 |
| Other neurodegenerative disorders | 0.1 | 0.1 | 0.2 | 0.2 | 1.8 | 0.6 | 0.2 | 0.1 | 0.1 | 0 | 0.2 | 0.1 | 0.3 | 0.3 | 0.7 | 3.9 | | 1.1 | 0.1 | 1.1 | 0.1 | 0.2 | 0.1 | 0.2 | 0.1 |
| Parkinson's disease | 0.7 | 0.9 | 1.3 | 1.5 | 1.8 | 8.2 | 1.2 | 0.2 | 0.5 | 0.1 | 1.1 | 0.6 | 0.8 | 0.5 | 1.1 | 1.6 | 5.9 | | 0.4 | 2.8 | 0.7 | 1.2 | 0.6 | 1.7 | 0.6 |
| Personality disorders | 8.7 | 11.5 | 20.2 | 1.3 | 1.2 | 1.5 | 7.4 | 9.3 | 23.4 | 21.7 | 3.8 | 3.1 | 2.3 | 6.4 | 2 | 2.1 | 1.8 | 1.5 | | 2 | 20.4 | 4.5 | 14.6 | 1.4 | 3.7 |
| Polyneuropathy | 3.6 | 1.4 | 1.8 | 2.4 | 1.7 | 3.6 | 1.9 | 0.4 | 1.4 | 0.6 | 2 | 1.2 | 2.6 | 0.6 | 2.7 | 9.1 | 11.7 | 5.4 | 1.1 | | 1.1 | 2 | 1.3 | 3.5 | 1.2 |
| Schizophrenia spectrum disorders | 8.5 | 7.5 | 20.2 | 1.5 | 2.3 | 3.7 | 5.3 | 6.8 | 26.9 | 7.6 | 4.9 | 1.7 | 2 | 13.3 | 1.6 | 1.8 | 1.9 | 2.3 | 18.6 | 1.8 | | 4 | 10 | 1.7 | 2.9 |
| Sleep disorders | 38.3 | 39.8 | 59 | 31.2 | 21 | 35.6 | 40.4 | 31.6 | 42.8 | 27.3 | 26.1 | 26.7 | 21.7 | 26.2 | 39.5 | 27.6 | 32.2 | 40.9 | 45.6 | 35.6 | 44.4 | | 39 | 31.3 | 19.9 |
| Stress related disorders | 20.2 | 23.5 | 34.1 | 6.3 | 7.7 | 6 | 18.9 | 20.6 | 38.2 | 25.9 | 10.5 | 9.2 | 7.6 | 18.3 | 6.6 | 7.2 | 6.9 | 6.2 | 48.2 | 7.4 | 36.3 | 12.8 | | 5.8 | 10.3 |
| Stroke | 7.2 | 4.2 | 4.7 | 10.3 | 7.1 | 16.4 | 5.7 | 0.7 | 3 | 0.7 | 13.8 | 3.5 | 7.6 | 2 | 5.2 | 5.7 | 5.7 | 10 | 2.3 | 10.5 | 3.2 | 5.3 | 3 | | 5 |
| Traumatic brain injury | 13.5 | 7.3 | 9.3 | 6.4 | 9.6 | 9.3 | 7.1 | 9.2 | 16.5 | 8.4 | 13.6 | 6.4 | 8.7 | 9 | 4.9 | 6.7 | 7.4 | 6.6 | 11.8 | 7 | 10.2 | 6.2 | 9.9 | 9.3 | |
| Myocardial infarction | 2.3 | 1.5 | 1.5 | 1.9 | 0.5 | 3.5 | 1.8 | 0.2 | 1.1 | 0.2 | 1.5 | 1.1 | 1.1 | 0.3 | 1.2 | 2.3 | 2.1 | 3.1 | 0.9 | 4.1 | 1 | 2.2 | 1.2 | 4.3 | 1.1 |
| Congestive heart failure | 3.1 | 1.7 | 1.8 | 2.5 | 0.7 | 5.7 | 2.2 | 0.2 | 1.2 | 0.2 | 2.1 | 1 | 1.7 | 0.7 | 1.1 | 4.5 | 2.6 | 4.3 | 0.8 | 6.4 | 1.3 | 2.8 | 1.1 | 6.8 | 1.4 |
| Peripheral vascular disease | 4.6 | 2.1 | 2.1 | 3.2 | 1 | 5.8 | 2.9 | 0.4 | 1.5 | 0.8 | 2.7 | 1.7 | 2.2 | 0.6 | 1.9 | 3.5 | 3.8 | 5.1 | 1.1 | 10.1 | 1.3 | 3.4 | 1.5 | 8.7 | 1.7 |
| Cerebrovascular disease | 7.9 | 4.8 | 5.7 | 12 | 7.5 | 21.6 | 6.5 | 0.8 | 3.3 | 0.8 | 14.7 | 4.1 | 7.6 | 2.2 | 5.1 | 6.5 | 7.7 | 13.1 | 2.6 | 13 | 3.5 | 6.3 | 3.4 | 77.2 | 5.4 |
| Chronic pulmonary disease | 11 | 7.8 | 8.6 | 7.5 | 9 | 10.5 | 8 | 5.1 | 8.5 | 4.2 | 7.7 | 6.5 | 7 | 6.1 | 5 | 17 | 20.1 | 9.3 | 7.6 | 12.9 | 7.7 | 8.7 | 7.1 | 11.1 | 6.1 |
| Connective tissue disease | 2 | 2.5 | 2.4 | 3.7 | 1.7 | 4 | 2.9 | 1.2 | 1.3 | 1.8 | 2.4 | 3 | 3.8 | 1.5 | 2.4 | 9.9 | 3.6 | 4.2 | 2 | 7.1 | 1.4 | 3.3 | 2.2 | 4.3 | 1.9 |
| Ulcer disease | 3.1 | 1.4 | 1.6 | 1.3 | 1.1 | 3.4 | 1.6 | 0.4 | 1.7 | 0.5 | 1.5 | 1 | 1.2 | 0.8 | 1 | 1.8 | 2 | 2.5 | 1.2 | 3.1 | 1.3 | 1.6 | 1.2 | 2.9 | 1 |
| Mild liver disease | 7.2 | 1.6 | 2 | 1.1 | 0.6 | 1.5 | 1.6 | 0.8 | 6.6 | 0.7 | 1.7 | 0.9 | 1.4 | 0.7 | 0.9 | 2 | 1.1 | 1 | 2.1 | 3.3 | 2.8 | 1.4 | 1.7 | 1.5 | 1.3 |
| Diabetes without end−organ damage | 6.8 | 4.7 | 6.7 | 6 | 2.2 | 11.6 | 5.7 | 1.4 | 3.9 | 1.6 | 4.9 | 3.3 | 4.5 | 3.4 | 3.4 | 7.2 | 6.1 | 8.8 | 4.4 | 19.9 | 6.2 | 6.2 | 4.3 | 12 | 3.2 |
| Hemiplegia | 0.4 | 0.3 | 0.3 | 2.4 | 22.5 | 0.4 | 0.4 | 0.3 | 0.3 | 0.1 | 2.4 | 0.2 | 2.4 | 2.5 | 2.3 | 2.7 | 7.2 | 0.9 | 0.2 | 1.1 | 0.3 | 0.4 | 0.3 | 1.3 | 0.5 |
| Moderate to severe renal disease | 2.7 | 1.6 | 3.2 | 2.6 | 1.4 | 4.9 | 2.1 | 0.6 | 1.7 | 0.8 | 2.5 | 1.3 | 2.8 | 1.3 | 1.4 | 4 | 2.5 | 4.1 | 1.2 | 7.2 | 1.6 | 2.6 | 1.4 | 5.7 | 1.4 |
| Diabetes with end−organ damage | 3.1 | 2 | 2.6 | 2.4 | 0.9 | 5.1 | 2.5 | 0.5 | 1.6 | 0.8 | 2.4 | 1.3 | 2 | 1.3 | 1.5 | 3.4 | 2.7 | 4.3 | 1.8 | 15.8 | 2.4 | 2.8 | 1.8 | 6.9 | 1.5 |
| Solid cancer tumour | 7.8 | 5.4 | 6 | 22.7 | 2.3 | 13.9 | 6.8 | 0.7 | 2.5 | 1.3 | 6.7 | 5.9 | 5.6 | 1.5 | 5.5 | 8.3 | 7.9 | 13.8 | 2.6 | 14.4 | 3.1 | 9.1 | 3.7 | 13.2 | 3.9 |
| Leukaemia | 0.2 | 0.2 | 0.2 | 0.5 | 0.1 | 0.4 | 0.2 | 0 | 0.1 | 0 | 0.2 | 0.2 | 0.5 | 0.2 | 0.1 | 0.4 | 0.4 | 0.4 | 0.1 | 0.8 | 0.1 | 0.3 | 0.1 | 0.4 | 0.1 |
| Lymphoma | 0.4 | 0.4 | 0.3 | 2.1 | 0.1 | 0.8 | 0.5 | 0.1 | 0.2 | 0.1 | 0.5 | 0.4 | 0.9 | 0.1 | 0.3 | 0.7 | 0.9 | 0.9 | 0.2 | 1.9 | 0.3 | 0.8 | 0.3 | 0.9 | 0.3 |
| Moderate to severe liver disease | 2.7 | 0.3 | 0.4 | 0.3 | 0.2 | 0.6 | 0.4 | 0.1 | 0.8 | 0.1 | 0.5 | 0.1 | 0.4 | 0.1 | 0.2 | 0.8 | 0.6 | 0.3 | 0.3 | 1.3 | 0.4 | 0.4 | 0.4 | 0.5 | 0.3 |
| Metastatic solid tumour | 0.7 | 0.5 | 0.4 | 2.5 | 0.2 | 0.6 | 0.6 | 0.1 | 0.2 | 0.1 | 0.7 | 0.5 | 0.5 | 0.1 | 0.4 | 0.8 | 0.6 | 0.8 | 0.2 | 1.4 | 0.2 | 0.8 | 0.3 | 0.9 | 0.3 |
| AIDS | 0.3 | 0.1 | 0.2 | 0.1 | 0.1 | 0.1 | 0.2 | 0.1 | 0.8 | 0.1 | 0.2 | 0.1 | 1 | 0.1 | 0 | 0.2 | 0.1 | 0.1 | 0.2 | 0.4 | 0.3 | 0.2 | 0.2 | 0.2 | 0.1 |

Percent: 80, 60, 40, 20, 0

### Concurrent diseases

#### Main analysis − Prevalent cohort 2021

| | Alcohol abuse | Anxiety disorders | Bipolar disorder | Brain tumors | Cerebral palsy | Dementia | Depression | Developmental and behavioural disorders | Drug abuse | Eating disorders | Epilepsy | Headache | Infections of the CNS | Intellectual disability | Multiple sclerosis | Neuromuscular disorders | Other neurodegenerative disorders | Parkinson's disease | Personality disorders | Polyneuropathy | Schizophrenia spectrum disorders | Sleep disorders | Stress related disorders | Stroke | Traumatic brain injury |
|---|---|---|---|---|---|---|---|---|---|---|---|---|---|---|---|---|---|---|---|---|---|---|---|---|---|
| Alcohol abuse | | 11.5 | 21.8 | 3.8 | 2.3 | 10.6 | 10.7 | 6.8 | 33.7 | 5.3 | 9.2 | 3.6 | 4.9 | 6.7 | 3.8 | 5.2 | 6 | 5.5 | 18.4 | 13.1 | 19.5 | 8.1 | 12.9 | 8.8 | 9 |
| Anxiety disorders | 23.1 | | 37.7 | 9.4 | 7 | 13.2 | 29.2 | 21.6 | 33.4 | 30.7 | 12.6 | 12.5 | 8.5 | 17.4 | 9.8 | 9.3 | 10.1 | 12.5 | 49.1 | 10.8 | 35.1 | 16.9 | 30.3 | 10.1 | 10 |
| Bipolar disorder | 5.1 | 4.4 | | 1.1 | 1 | 2.8 | 3.9 | 2.8 | 8.2 | 3.9 | 1.8 | 1.4 | 1.3 | 2.6 | 1 | 1.2 | 1.3 | 2.1 | 9.9 | 1.6 | 10.7 | 2.9 | 5.1 | 1.3 | 1.5 |
| Brain tumors | 0.5 | 0.6 | 0.6 | | 0.9 | 1.2 | 0.7 | 0.2 | 0.3 | 0.2 | 4.4 | 0.7 | 2.2 | 0.5 | 1.1 | 0.9 | 1.2 | 1.3 | 0.3 | 1.1 | 0.4 | 0.8 | 0.5 | 1.5 | 0.5 |
| Cerebral palsy | 0.1 | 0.2 | 0.2 | 0.4 | | 0.2 | 0.2 | 0.6 | 0.2 | 0.2 | 3.7 | 0.1 | 1.4 | 6.5 | 0.3 | 1.8 | 4.1 | 0.7 | 0.1 | 0.4 | 0.3 | 0.3 | 0.3 | 0.5 | 0.4 |
| Dementia | 2.8 | 1.8 | 3.2 | 2.7 | 1.1 | | 2.8 | 0.2 | 1.2 | 0.1 | 3.3 | 0.7 | 1.8 | 2.1 | 1.3 | 1.8 | 6.2 | 16.5 | 0.8 | 3.6 | 2.2 | 2 | 1 | 5.4 | 1.7 |
| Depression | 52.3 | 70.9 | 82.3 | 26.9 | 14.1 | 50.4 | | 31.6 | 57.5 | 50.2 | 27.8 | 30.1 | 21.6 | 25.9 | 31.7 | 24.2 | 32.9 | 41.5 | 76.9 | 34.4 | 59.8 | 41.6 | 59.3 | 34.1 | 23.4 |
| Developmental and behavioural disorders | 8.3 | 13.2 | 14.9 | 2.1 | 13.8 | 0.9 | 8 | | 29.7 | 16.2 | 10.2 | 4 | 5.6 | 45.3 | 2.2 | 4.8 | 3.2 | 1.6 | 24.4 | 1.7 | 19.7 | 8.6 | 16.5 | 1.1 | 7.6 |
| Drug abuse | 12.5 | 6.2 | 13 | 1 | 1.2 | 1.7 | 4.4 | 9 | | 5 | 3.8 | 1.7 | 2.1 | 6 | 1.1 | 1.5 | 0.9 | 1.2 | 18.2 | 1.8 | 23 | 3.4 | 9.1 | 1.4 | 4.1 |
| Eating disorders | 0.6 | 1.9 | 2.1 | 0.2 | 0.4 | 0.1 | 1.3 | 1.6 | 1.6 | | 0.6 | 0.6 | 0.6 | 0.8 | 0.3 | 0.5 | 0.2 | 0.1 | 5.5 | 0.3 | 2.1 | 0.7 | 2 | 0.1 | 0.7 |
| Epilepsy | 4 | 2.7 | 3.4 | 16 | 28.4 | 5.3 | 2.5 | 3.6 | 4.5 | 2.2 | | 1.9 | 8.1 | 19.2 | 3.2 | 3.9 | 4.3 | 3.6 | 3.5 | 3.3 | 5 | 2.4 | 2.9 | 7.4 | 4 |
| Headache | 10.1 | 17.6 | 16.8 | 16.3 | 5.9 | 7.6 | 17.5 | 9.1 | 12.8 | 15 | 12.4 | | 16 | 5.9 | 16.1 | 12.5 | 11.7 | 13.2 | 18.8 | 12.4 | 11.4 | 15.9 | 17 | 12.2 | 12.5 |
| Infections of the CNS | 0.6 | 0.5 | 0.6 | 2.1 | 2.8 | 0.8 | 0.5 | 0.5 | 0.7 | 0.6 | 2.2 | 0.7 | | 0.9 | 1.9 | 1 | 1.1 | 0.7 | 0.6 | 1.2 | 0.6 | 0.6 | 0.6 | 1.1 | 0.7 |
| Intellectual disability | 1.3 | 1.6 | 2.2 | 0.8 | 21.6 | 1.5 | 1 | 7 | 3.1 | 1.3 | 8.4 | 0.4 | 1.5 | | 0.3 | 2.3 | 2.1 | 0.6 | 2.6 | 0.5 | 5.9 | 1.1 | 2.3 | 0.5 | 1.2 |
| Multiple sclerosis | 0.4 | 0.5 | 0.5 | 1 | 0.6 | 0.5 | 0.7 | 0.2 | 0.3 | 0.2 | 0.8 | 0.6 | 1.7 | 0.2 | | 1 | 2.7 | 0.9 | 0.4 | 1.1 | 0.4 | 0.9 | 0.4 | 0.7 | 0.4 |
| Neuromuscular disorders | 0.3 | 0.2 | 0.3 | 0.4 | 1.7 | 0.4 | 0.3 | 0.2 | 0.2 | 0.2 | 0.5 | 0.2 | 0.4 | 0.6 | 0.5 | | 8.1 | 0.6 | 0.2 | 1.7 | 0.2 | 0.3 | 0.2 | 0.4 | 0.2 |
| Other neurodegenerative disorders | 0.1 | 0.1 | 0.1 | 0.3 | 1.8 | 0.6 | 0.2 | 0.1 | 0.1 | 0 | 0.2 | 0.1 | 0.2 | 0.3 | 0.6 | 3.8 | | 1.1 | 0.1 | 1 | 0.1 | 0.2 | 0.1 | 0.2 | 0.1 |
| Parkinson's disease | 0.8 | 0.9 | 1.3 | 1.5 | 1.7 | 8.5 | 1.2 | 0.2 | 0.5 | 0.2 | 1.1 | 0.6 | 0.8 | 0.5 | 1.2 | 1.5 | 6.1 | | 0.4 | 2.7 | 0.7 | 1.2 | 0.5 | 1.7 | 0.6 |
| Personality disorders | 8.6 | 11.4 | 19.9 | 1.2 | 1.1 | 1.4 | 7.4 | 9.3 | 23 | 21.2 | 3.7 | 3.1 | 2.2 | 6.5 | 2 | 2 | 1.8 | 1.4 | | 2 | 19.9 | 4.5 | 14.4 | 1.3 | 3.7 |
| Polyneuropathy | 3.6 | 1.5 | 1.9 | 2.5 | 1.7 | 3.6 | 1.9 | 0.4 | 1.3 | 0.6 | 2.1 | 1.2 | 2.7 | 0.7 | 2.7 | 9.1 | 11.6 | 5.3 | 1.2 | | 1.2 | 2 | 1.3 | 3.6 | 1.3 |
| Schizophrenia spectrum disorders | 8.4 | 7.6 | 19.9 | 1.5 | 2.3 | 3.5 | 5.3 | 6.9 | 26.9 | 7.6 | 5 | 1.7 | 2.2 | 13.5 | 1.6 | 1.7 | 1.9 | 2.1 | 18.5 | 1.9 | | 4.1 | 10.1 | 1.7 | 3 |
| Sleep disorders | 39.1 | 40.8 | 60.1 | 31.6 | 22.3 | 36.1 | 41.3 | 33.9 | 44 | 28.5 | 27.1 | 27.2 | 22.9 | 27.9 | 40.2 | 28.2 | 32.9 | 41.9 | 46.9 | 36 | 45.6 | | 40.2 | 31.7 | 20.7 |
| Stress related disorders | 20.3 | 23.7 | 34.3 | 6.3 | 7.8 | 5.8 | 19.1 | 21.1 | 38.6 | 25.9 | 10.6 | 9.4 | 7.8 | 18.9 | 6.6 | 7.2 | 6.8 | 6.2 | 48.5 | 7.5 | 36.7 | 13 | | 5.8 | 10.5 |
| Stroke | 7.1 | 4.1 | 4.6 | 10.1 | 7.2 | 16.1 | 5.6 | 0.7 | 2.9 | 0.7 | 13.7 | 3.5 | 7.6 | 1.9 | 5.2 | 5.6 | 6 | 10.1 | 2.3 | 10.5 | 3.2 | 5.3 | 3 | | 5.1 |
| Traumatic brain injury | 13 | 7.2 | 9 | 6.4 | 9.3 | 9.2 | 6.9 | 8.9 | 15.9 | 8.2 | 13.4 | 6.4 | 8.5 | 8.8 | 4.8 | 6.5 | 7.5 | 6.6 | 11.4 | 6.8 | 9.9 | 6.2 | 9.7 | 9.2 | |
| Myocardial infarction | 2.2 | 1.4 | 1.5 | 1.9 | 0.5 | 3.3 | 1.8 | 0.2 | 1 | 0.2 | 1.4 | 1 | 1.2 | 0.3 | 1.2 | 2.3 | 2 | 2.9 | 0.8 | 4.1 | 1 | 2.2 | 1.2 | 4.2 | 1.1 |
| Congestive heart failure | 3 | 1.6 | 1.7 | 2.4 | 0.8 | 5.5 | 2.1 | 0.2 | 1.2 | 0.2 | 2.1 | 0.9 | 1.7 | 0.7 | 1.1 | 4.5 | 2.8 | 4.2 | 0.8 | 6.4 | 1.3 | 2.8 | 1.1 | 6.6 | 1.4 |
| Peripheral vascular disease | 4.5 | 2 | 2.1 | 3.2 | 1 | 5.8 | 2.8 | 0.4 | 1.5 | 0.8 | 2.7 | 1.7 | 2.2 | 0.6 | 1.9 | 3.4 | 4 | 5 | 1.1 | 10.1 | 1.3 | 3.4 | 1.5 | 8.5 | 1.7 |
| Cerebrovascular disease | 7.7 | 4.6 | 5.5 | 11.7 | 7.5 | 20.8 | 6.3 | 0.8 | 3.1 | 0.7 | 14.5 | 4.1 | 7.7 | 2 | 5.1 | 6.3 | 7.9 | 13.1 | 2.4 | 12.8 | 3.4 | 6.2 | 3.3 | 77 | 5.4 |
| Chronic pulmonary disease | 10.8 | 7.6 | 8.5 | 7.4 | 9 | 10.3 | 7.8 | 5 | 8.2 | 4.1 | 7.5 | 6.4 | 7.1 | 5.9 | 4.9 | 17.2 | 19.6 | 8.9 | 7.6 | 12.8 | 7.5 | 8.6 | 6.9 | 10.9 | 6 |
| Connective tissue disease | 2.2 | 2.4 | 2.4 | 3.7 | 1.6 | 4 | 2.8 | 1.2 | 1.3 | 1.7 | 2.4 | 3 | 3.8 | 1.4 | 2.4 | 9.9 | 3.5 | 4.2 | 2 | 7.1 | 1.4 | 3.3 | 2.1 | 4.2 | 1.9 |
| Ulcer disease | 3 | 1.3 | 1.5 | 1.3 | 1.1 | 3.2 | 1.5 | 0.3 | 1.6 | 0.5 | 1.4 | 1 | 1.2 | 0.7 | 1 | 1.6 | 1.8 | 2.3 | 1.1 | 2.9 | 1.2 | 1.5 | 1.1 | 2.8 | 1 |
| Mild liver disease | 7.1 | 1.5 | 2 | 1.1 | 0.6 | 1.4 | 1.5 | 0.8 | 6.3 | 0.7 | 1.6 | 0.9 | 1.4 | 0.7 | 0.9 | 1.9 | 1.2 | 1 | 2 | 3.1 | 2.8 | 1.4 | 1.7 | 1.5 | 1.3 |
| Diabetes without end−organ damage | 6.5 | 4.6 | 6.5 | 5.7 | 2.1 | 11.3 | 5.5 | 1.4 | 3.8 | 1.6 | 4.9 | 3.2 | 4.6 | 3.4 | 3.3 | 7.1 | 5.6 | 8.5 | 4.3 | 21.6 | 6.1 | 6 | 4.2 | 11.7 | 3.2 |
| Hemiplegia | 0.3 | 0.3 | 0.3 | 2.3 | 21.6 | 0.4 | 0.4 | 0.2 | 0.3 | 0.1 | 2.3 | 0.2 | 2.3 | 2.3 | 2.1 | 2.5 | 7 | 0.9 | 0.2 | 1 | 0.3 | 0.4 | 0.3 | 1.3 | 0.5 |
| Moderate to severe renal disease | 2.7 | 1.6 | 3.2 | 2.5 | 1.4 | 5 | 2.1 | 0.6 | 1.7 | 0.7 | 2.5 | 1.3 | 2.9 | 1.3 | 1.4 | 4 | 2.4 | 4 | 1.2 | 7.7 | 1.7 | 2.6 | 1.4 | 5.6 | 1.5 |
| Diabetes with end−organ damage | 3 | 1.9 | 2.6 | 2.2 | 0.9 | 4.7 | 2.5 | 0.5 | 1.6 | 0.7 | 2.3 | 1.2 | 2 | 1.3 | 1.4 | 3.5 | 2.8 | 4 | 1.8 | 18.3 | 2.3 | 2.7 | 1.8 | 6.7 | 1.5 |
| Solid cancer tumour | 7.9 | 5.4 | 6.1 | 22.4 | 2.3 | 14.1 | 6.9 | 0.8 | 2.4 | 1.3 | 6.8 | 5.9 | 5.8 | 1.4 | 5.6 | 8.9 | 8.3 | 14.1 | 2.6 | 14.7 | 3.1 | 9.1 | 3.7 | 13.5 | 4.1 |
| Leukaemia | 0.2 | 0.2 | 0.2 | 0.5 | 0.1 | 0.4 | 0.2 | 0.1 | 0.1 | 0 | 0.3 | 0.2 | 0.5 | 0.2 | 0.2 | 0.4 | 0.4 | 0.4 | 0.1 | 0.8 | 0.1 | 0.3 | 0.1 | 0.4 | 0.2 |
| Lymphoma | 0.4 | 0.4 | 0.3 | 2.1 | 0.2 | 0.9 | 0.5 | 0.1 | 0.2 | 0.1 | 0.5 | 0.4 | 1 | 0.1 | 0.3 | 0.8 | 0.9 | 1 | 0.2 | 2 | 0.3 | 0.8 | 0.3 | 0.9 | 0.3 |
| Moderate to severe liver disease | 2.6 | 0.3 | 0.4 | 0.3 | 0.2 | 0.6 | 0.4 | 0.1 | 0.7 | 0.1 | 0.5 | 0.1 | 0.4 | 0.1 | 0.2 | 0.7 | 0.6 | 0.2 | 0.3 | 1.3 | 0.4 | 0.3 | 0.4 | 0.5 | 0.3 |
| Metastatic solid tumour | 0.7 | 0.4 | 0.4 | 2.5 | 0.2 | 0.6 | 0.6 | 0.1 | 0.2 | 0.1 | 0.7 | 0.5 | 0.6 | 0.2 | 0.4 | 0.8 | 0.4 | 0.9 | 0.2 | 1.4 | 0.2 | 0.8 | 0.3 | 1 | 0.3 |
| AIDS | 0.3 | 0.1 | 0.2 | 0.1 | 0.1 | 0.1 | 0.2 | 0.1 | 0.8 | 0.1 | 0.2 | 0.1 | 1 | 0.1 | 0 | 0.2 | 0 | 0.1 | 0.2 | 0.4 | 0.3 | 0.2 | 0.2 | 0.2 | 0.1 |

Percent: 0, 20, 40, 60, 80
