## Appendix 4 for "Temporal trends in the occurrence, mortality, and cost of brain disorders in Denmark"

| | *Prevalence, N (%)* | *Prevalence per 100,000 persons (95% CI)* |
|---|---|---|
| Any brain disorder | 1,893,318 (33.2) | 33,184 (33,137-33,231) |
| Alcohol abuse | 174,496 (3.1) | 3,058 (3,044-3,073) |
| Anxiety disorders | 253,686 (4.4) | 4,446 (4,429-4,464) |
| Bipolar disorder | 33,314 (0.6) | 584 (578-590) |
| Brain tumors | 15,212 (0.3) | 267 (262-271) |
| Cerebral palsy | 9,308 (0.2) | 163 (160-166) |
| Dementia | 43,663 (0.8) | 765 (758-772) |
| Depression | 719,788 (12.6) | 12,616 (12,586-12,645) |
| Developmental and behavioural disorders | 129,663 (2.3) | 2,273 (2,260-2,285) |
| Drug abuse | 50,859 (0.9) | 891 (884-899) |
| Eating disorders | 16,081 (0.3) | 282 (278-286) |
| Epilepsy | 73,582 (1.3) | 1,290 (1,280-1,299) |
| Headache | 427,370 (7.5) | 7,490 (7,468-7,513) |
| Infections of the CNS | 18,586 (0.3) | 326 (321-330) |
| Intellectual disability | 27,868 (0.5) | 488 (483-494) |
| Multiple sclerosis | 15,023 (0.3) | 263 (259-268) |
| Neuromuscular disorders | 7,391 (0.1) | 130 (127-133) |
| Other neurodegenerative disorders | 3,435 (0.1) | 60 (58-62) |
| Parkinson’s disease | 26,701 (0.5) | 468 (462-474) |
| Personality disorders | 71,766 (1.3) | 1,258 (1,249-1,267) |
| Polyneuropathy | 34,187 (0.6) | 599 (593-606) |
| Schizophrenia spectrum disorders | 61,462 (1.1) | 1,077 (1,069-1,086) |
| Sleep disorders | 661,519 (11.6) | 11,594 (11,566-11,622) |
| Stress related disorders | 197,786 (3.5) | 3,467 (3,451-3,482) |
| Stroke | 120,435 (2.1) | 2,111 (2,099-2,123) |
| Traumatic brain injury | 252,108 (4.4) | 4,419 (4,401-4,436) |

| | *Prevalence, N (%)* | *Prevalence per 100,000 persons (95% CI)* |
|---|---|---|
| Any brain disorder | 1,926,077 (33.4) | 33,424 (33,377-33,472) |
| Alcohol abuse | 172,908 (3.0) | 3,001 (2,986-3,015) |
| Anxiety disorders | 269,266 (4.7) | 4,673 (4,655-4,690) |
| Bipolar disorder | 34,504 (0.6) | 599 (592-605) |
| Brain tumors | 15,828 (0.3) | 275 (270-279) |
| Cerebral palsy | 9,378 (0.2) | 163 (159-166) |
| Dementia | 44,364 (0.8) | 770 (763-777) |
| Depression | 737,896 (12.8) | 12,805 (12,776-12,834) |
| Developmental and behavioural disorders | 140,283 (2.4) | 2,434 (2,422-2,447) |
| Drug abuse | 52,863 (0.9) | 917 (910-925) |
| Eating disorders | 16,915 (0.3) | 294 (289-298) |
| Epilepsy | 73,771 (1.3) | 1,280 (1,271-1,289) |
| Headache | 435,367 (7.6) | 7,555 (7,533-7,578) |
| Infections of the CNS | 18,671 (0.3) | 324 (319-329) |
| Intellectual disability | 28,593 (0.5) | 496 (490-502) |
| Multiple sclerosis | 15,450 (0.3) | 268 (264-272) |
| Neuromuscular disorders | 7,670 (0.1) | 133 (130-136) |
| Other neurodegenerative disorders | 3,571 (0.1) | 62 (60-64) |
| Parkinson's disease | 23,394 (0.4) | 406 (401-411) |
| Personality disorders | 73,566 (1.3) | 1,277 (1,267-1,286) |
| Polyneuropathy | 36,182 (0.6) | 628 (621-634) |
| Schizophrenia spectrum disorders | 63,115 (1.1) | 1,095 (1,087-1,104) |
| Sleep disorders | 683,166 (11.9) | 11,855 (11,827-11,884) |
| Stress related disorders | 209,698 (3.6) | 3,639 (3,623-3,655) |
| Stroke | 122,391 (2.1) | 2,124 (2,112-2,136) |
| Traumatic brain injury | 249,852 (4.3) | 4,336 (4,319-4,353) |

| | *Prevalence, N (%)* | *Prevalence per 100,000 persons (95% CI)* |
|---|---|---|
| Any brain disorder | 1,955,822 (33.8) | 33,825 (33,778-33,872) |
| Alcohol abuse | 170,934 (3.0) | 2,956 (2,942-2,970) |
| Anxiety disorders | 281,621 (4.9) | 4,871 (4,853-4,889) |
| Bipolar disorder | 35,266 (0.6) | 610 (604-616) |
| Brain tumors | 16,702 (0.3) | 289 (285-293) |
| Cerebral palsy | 9,442 (0.2) | 163 (160-167) |
| Dementia | 44,757 (0.8) | 774 (767-781) |
| Depression | 749,368 (13.0) | 12,960 (12,931-12,989) |
| Developmental and behavioural disorders | 150,901 (2.6) | 2,610 (2,597-2,623) |
| Drug abuse | 54,521 (0.9) | 943 (935-951) |
| Eating disorders | 17,512 (0.3) | 303 (298-307) |
| Epilepsy | 73,701 (1.3) | 1,275 (1,265-1,284) |
| Headache | 443,400 (7.7) | 7,668 (7,646-7,691) |
| Infections of the CNS | 18,798 (0.3) | 325 (320-330) |
| Intellectual disability | 29,379 (0.5) | 508 (502-514) |
| Multiple sclerosis | 15,940 (0.3) | 276 (271-280) |
| Neuromuscular disorders | 8,027 (0.1) | 139 (136-142) |
| Other neurodegenerative disorders | 3,772 (0.1) | 65 (63-67) |
| Parkinson's disease | 22,803 (0.4) | 394 (389-400) |
| Personality disorders | 74,393 (1.3) | 1,287 (1,277-1,296) |
| Polyneuropathy | 37,920 (0.7) | 656 (649-662) |
| Schizophrenia spectrum disorders | 64,558 (1.1) | 1,117 (1,108-1,125) |
| Sleep disorders | 703,408 (12.2) | 12,165 (12,137-12,194) |
| Stress related disorders | 221,022 (3.8) | 3,822 (3,807-3,838) |
| Stroke | 124,390 (2.2) | 2,151 (2,139-2,163) |
| Traumatic brain injury | 247,456 (4.3) | 4,280 (4,263-4,297) |

| | *Prevalence, N (%)* | *Prevalence per 100,000 persons (95% CI)* |
|---|---|---|
| Any brain disorder | 1,986,200 (34.2) | 34,191 (34,143-34,238) |
| Alcohol abuse | 168,151 (2.9) | 2,895 (2,881-2,908) |
| Anxiety disorders | 293,398 (5.1) | 5,051 (5,032-5,069) |
| Bipolar disorder | 36,015 (0.6) | 620 (614-626) |
| Brain tumors | 17,420 (0.3) | 300 (295-304) |
| Cerebral palsy | 9,501 (0.2) | 164 (160-167) |
| Dementia | 45,059 (0.8) | 776 (769-783) |
| Depression | 761,338 (13.1) | 13,106 (13,076-13,135) |
| Developmental and behavioural disorders | 162,935 (2.8) | 2,805 (2,791-2,818) |
| Drug abuse | 56,304 (1.0) | 969 (961-977) |
| Eating disorders | 18,261 (0.3) | 314 (310-319) |
| Epilepsy | 73,601 (1.3) | 1,267 (1,258-1,276) |
| Headache | 450,583 (7.8) | 7,756 (7,734-7,779) |
| Infections of the CNS | 19,142 (0.3) | 330 (325-334) |
| Intellectual disability | 30,217 (0.5) | 520 (514-526) |
| Multiple sclerosis | 16,308 (0.3) | 281 (276-285) |
| Neuromuscular disorders | 8,225 (0.1) | 142 (139-145) |
| Other neurodegenerative disorders | 3,911 (0.1) | 67 (65-69) |
| Parkinson's disease | 22,474 (0.4) | 387 (382-392) |
| Personality disorders | 75,283 (1.3) | 1,296 (1,287-1,305) |
| Polyneuropathy | 39,453 (0.7) | 679 (672-686) |
| Schizophrenia spectrum disorders | 66,225 (1.1) | 1,140 (1,131-1,149) |
| Sleep disorders | 724,196 (12.5) | 12,466 (12,438-12,495) |
| Stress related disorders | 232,012 (4.0) | 3,994 (3,978-4,010) |
| Stroke | 126,495 (2.2) | 2,177 (2,166-2,190) |
| Traumatic brain injury | 245,120 (4.2) | 4,220 (4,203-4,236) |

| | *Prevalence, N (%)* | *Prevalence per 100,000 persons (95% CI)* |
|---|---|---|
| Any brain disorder | 2,012,655 (34.5) | 34,527 (34,479-34,575) |
| Alcohol abuse | 165,090 (2.8) | 2,832 (2,818-2,846) |
| Anxiety disorders | 305,008 (5.2) | 5,232 (5,214-5,251) |
| Bipolar disorder | 36,617 (0.6) | 628 (622-635) |
| Brain tumors | 18,110 (0.3) | 311 (306-315) |
| Cerebral palsy | 9,485 (0.2) | 163 (159-166) |
| Dementia | 44,370 (0.8) | 761 (754-768) |
| Depression | 772,157 (13.2) | 13,246 (13,217-13,276) |
| Developmental and behavioural disorders | 174,775 (3.0) | 2,998 (2,984-3,012) |
| Drug abuse | 57,758 (1.0) | 991 (983-999) |
| Eating disorders | 18,751 (0.3) | 322 (317-326) |
| Epilepsy | 72,679 (1.2) | 1,247 (1,238-1,256) |
| Headache | 455,840 (7.8) | 7,820 (7,797-7,843) |
| Infections of the CNS | 19,429 (0.3) | 333 (329-338) |
| Intellectual disability | 30,697 (0.5) | 527 (521-533) |
| Multiple sclerosis | 16,515 (0.3) | 283 (279-288) |
| Neuromuscular disorders | 8,330 (0.1) | 143 (140-146) |
| Other neurodegenerative disorders | 3,943 (0.1) | 68 (66-70) |
| Parkinson’s disease | 21,955 (0.4) | 377 (372-382) |
| Personality disorders | 75,410 (1.3) | 1,294 (1,284-1,303) |
| Polyneuropathy | 40,756 (0.7) | 699 (692-706) |
| Schizophrenia spectrum disorders | 67,745 (1.2) | 1,162 (1,153-1,171) |
| Sleep disorders | 744,407 (12.8) | 12,770 (12,741-12,799) |
| Stress related disorders | 241,284 (4.1) | 4,139 (4,123-4,156) |
| Stroke | 127,804 (2.2) | 2,192 (2,180-2,205) |
| Traumatic brain injury | 242,622 (4.2) | 4,162 (4,146-4,179) |

| | *Prevalence, N (%)* | *Prevalence per 100,000 persons (95% CI)* |
|---|---|---|
| Any brain disorder | 2,036,638 (34.7) | 34,746 (34,698-34,793) |
| Alcohol abuse | 163,416 (2.8) | 2,788 (2,774-2,801) |
| Anxiety disorders | 315,928 (5.4) | 5,390 (5,371-5,409) |
| Bipolar disorder | 37,333 (0.6) | 637 (630-643) |
| Brain tumors | 18,927 (0.3) | 323 (318-328) |
| Cerebral palsy | 9,403 (0.2) | 160 (157-164) |
| Dementia | 43,973 (0.8) | 750 (743-757) |
| Depression | 783,417 (13.4) | 13,365 (13,336-13,395) |
| Developmental and behavioural disorders | 186,974 (3.2) | 3,190 (3,175-3,204) |
| Drug abuse | 59,175 (1.0) | 1,010 (1,001-1,018) |
| Eating disorders | 19,236 (0.3) | 328 (324-333) |
| Epilepsy | 71,789 (1.2) | 1,225 (1,216-1,234) |
| Headache | 458,826 (7.8) | 7,828 (7,805-7,850) |
| Infections of the CNS | 19,599 (0.3) | 334 (330-339) |
| Intellectual disability | 30,982 (0.5) | 529 (523-534) |
| Multiple sclerosis | 16,960 (0.3) | 289 (285-294) |
| Neuromuscular disorders | 8,454 (0.1) | 144 (141-147) |
| Other neurodegenerative disorders | 3,991 (0.1) | 68 (66-70) |
| Parkinson's disease | 22,293 (0.4) | 380 (375-385) |
| Personality disorders | 75,678 (1.3) | 1,291 (1,282-1,300) |
| Polyneuropathy | 42,758 (0.7) | 729 (723-736) |
| Schizophrenia spectrum disorders | 69,162 (1.2) | 1,180 (1,171-1,189) |
| Sleep disorders | 763,765 (13.0) | 13,030 (13,001-13,059) |
| Stress related disorders | 249,672 (4.3) | 4,259 (4,243-4,276) |
| Stroke | 129,247 (2.2) | 2,205 (2,193-2,217) |
| Traumatic brain injury | 239,984 (4.1) | 4,094 (4,078-4,111) |

| | *Prevalence, N (%)* | *Prevalence per 100,000 persons (95% CI)* |
|---|---|---|
| Any brain disorder | 2,059,852 (35.2) | 35,171 (35,123-35,219) |
| Alcohol abuse | 162,735 (2.8) | 2,779 (2,765-2,792) |
| Anxiety disorders | 327,178 (5.6) | 5,586 (5,567-5,606) |
| Bipolar disorder | 37,892 (0.6) | 647 (641-654) |
| Brain tumors | 19,592 (0.3) | 335 (330-339) |
| Cerebral palsy | 9,351 (0.2) | 160 (156-163) |
| Dementia | 43,678 (0.7) | 746 (739-753) |
| Depression | 793,419 (13.5) | 13,547 (13,518-13,577) |
| Developmental and behavioural disorders | 199,940 (3.4) | 3,414 (3,399-3,429) |
| Drug abuse | 60,279 (1.0) | 1,029 (1,021-1,037) |
| Eating disorders | 19,756 (0.3) | 337 (333-342) |
| Epilepsy | 70,944 (1.2) | 1,211 (1,202-1,220) |
| Headache | 461,353 (7.9) | 7,877 (7,855-7,900) |
| Infections of the CNS | 19,179 (0.3) | 327 (323-332) |
| Intellectual disability | 30,980 (0.5) | 529 (523-535) |
| Multiple sclerosis | 17,343 (0.3) | 296 (292-301) |
| Neuromuscular disorders | 8,606 (0.1) | 147 (144-150) |
| Other neurodegenerative disorders | 4,020 (0.1) | 69 (67-71) |
| Parkinson’s disease | 22,528 (0.4) | 385 (380-390) |
| Personality disorders | 75,906 (1.3) | 1,296 (1,287-1,305) |
| Polyneuropathy | 44,577 (0.8) | 761 (754-768) |
| Schizophrenia spectrum disorders | 70,597 (1.2) | 1,205 (1,197-1,214) |
| Sleep disorders | 788,209 (13.5) | 13,458 (13,429-13,488) |
| Stress related disorders | 255,796 (4.4) | 4,368 (4,351-4,385) |
| Stroke | 131,130 (2.2) | 2,239 (2,227-2,251) |
| Traumatic brain injury | 235,012 (4.0) | 4,013 (3,997-4,029) |

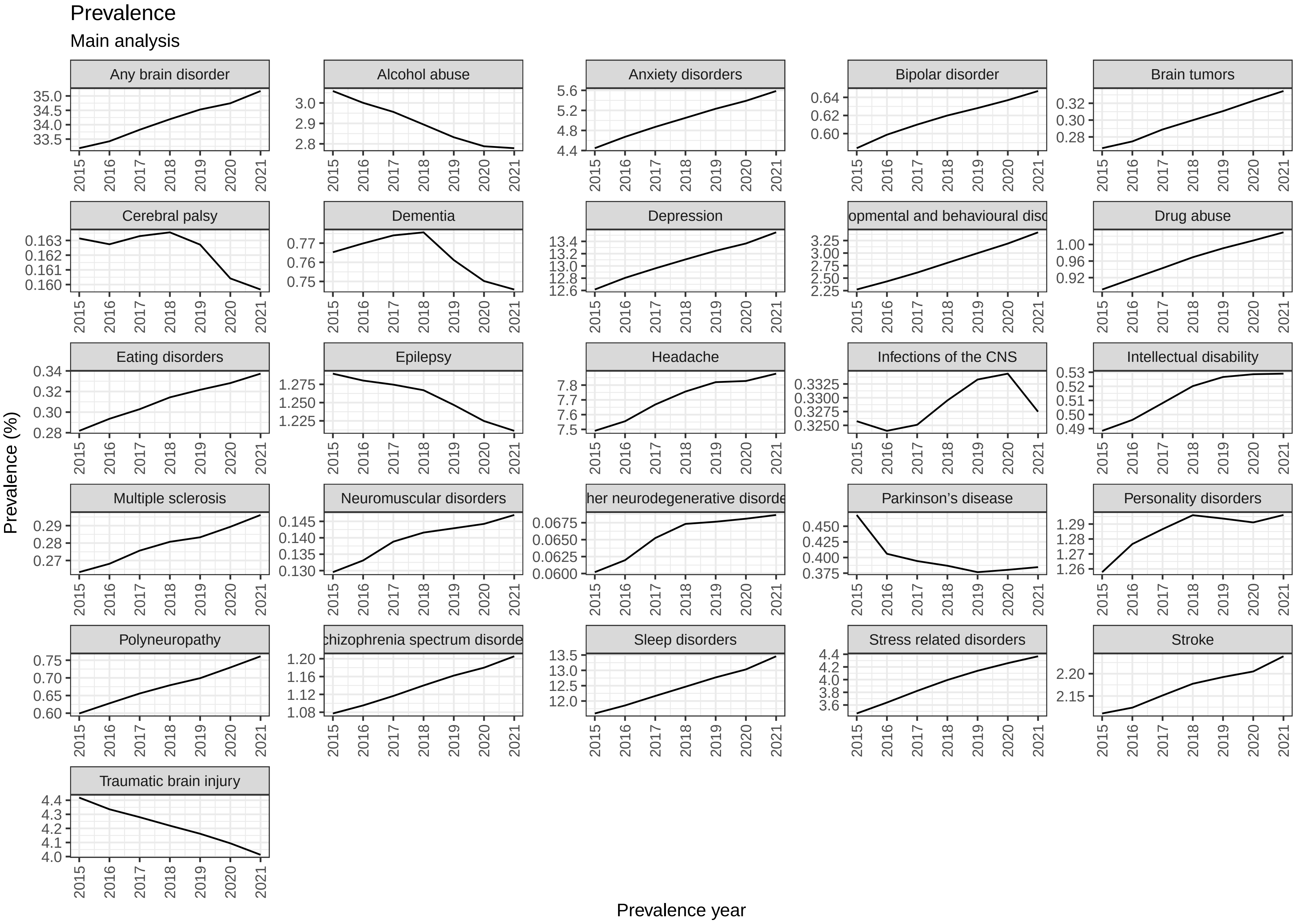
Prevalence
Main analysis
Any brain disorder
Alcohol abuse
Anxiety disorders
Bipolar disorder
Brain tumors
Cerebral palsy
Dementia
Depression
opmental and behavioural disc
Drug abuse
Eating disorders
Epilepsy
Headache
Infections of the CNS
Intellectual disability
Multiple sclerosis
Neuromuscular disorders
her neurodegenerative disorde
Parkinson's disease
Personality disorders
Polyneuropathy
chizophrenia spectrum disorde
Sleep disorders
Stress related disorders
Stroke
Traumatic brain injury
Prevalence (%)
Prevalence year

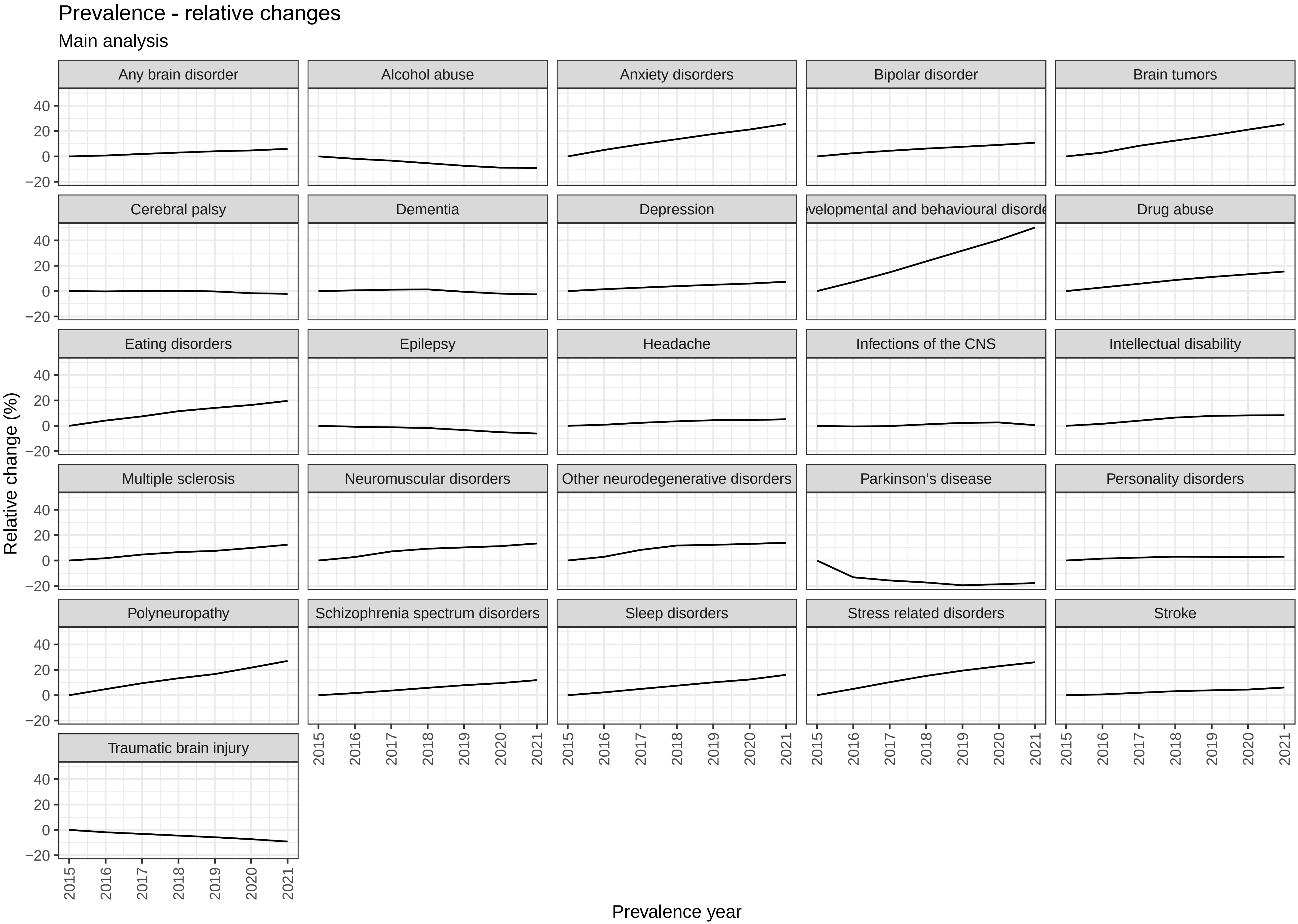

Prevalence - relative changes
Main analysis
Any brain disorder
Alcohol abuse
Anxiety disorders
Bipolar disorder
Brain tumors
Cerebral palsy
Dementia
Depression
evelopmental and behavioural disorde
Drug abuse
Eating disorders
Epilepsy
Headache
Infections of the CNS
Intellectual disability
Multiple sclerosis
Neuromuscular disorders
Other neurodegenerative disorders
Parkinson's disease
Personality disorders
Polyneuropathy
Schizophrenia spectrum disorders
Sleep disorders
Stress related disorders
Stroke
Traumatic brain injury
40
20
0
−20
2015
2016
2017
2018
2019
2020
2021
Relative change (%)
Prevalence year
