## Appendix 5 for "Temporal trends in the occurrence, mortality, and cost of brain disorders in Denmark"

| | *Incidence, N* | *Incidence rate per 100,000 person-years (95% CI)* |
|---|---|---|
| Any brain disorder | 507,789 | 1,792 (1,787-1,797) |
| Alcohol abuse | 45,094 | 159 (158-161) |
| Anxiety disorders | 117,585 | 415 (413-417) |
| Bipolar disorder | 11,985 | 42 (42-43) |
| Brain tumors | 9,739 | 34 (34-35) |
| Cerebral palsy | 2,333 | 8 (8-9) |
| Dementia | 45,905 | 162 (160-163) |
| Depression | 244,331 | 862 (859-865) |
| Developmental and behavioural disorders | 63,072 | 223 (221-224) |
| Drug abuse | 19,069 | 67 (66-68) |
| Eating disorders | 6,105 | 22 (21-22) |
| Epilepsy | 22,978 | 81 (80-82) |
| Headache | 105,727 | 373 (371-375) |
| Infections of the CNS | 6,757 | 24 (23-24) |
| Intellectual disability | 8,467 | 30 (29-31) |
| Multiple sclerosis | 4,124 | 15 (14-15) |
| Neuromuscular disorders | 3,373 | 12 (12-12) |
| Other neurodegenerative disorders | 2,199 | 8 (7-8) |
| Parkinson’s disease | 11,429 | 40 (40-41) |
| Personality disorders | 21,886 | 77 (76-78) |
| Polyneuropathy | 19,514 | 69 (68-70) |
| Schizophrenia spectrum disorders | 19,387 | 68 (67-69) |
| Sleep disorders | 243,342 | 859 (855-862) |
| Stress related disorders | 82,275 | 290 (288-292) |
| Stroke | 67,899 | 240 (238-241) |
| Traumatic brain injury | 65,732 | 232 (230-234) |

| | *Incidence, N* | *Incidence rate per 100,000 person-years (95% CI)* |
|---|---|---|
| Any brain disorder | 571,268 | 1,634 (1,630-1,639) |
| Alcohol abuse | 48,772 | 140 (138-141) |
| Anxiety disorders | 120,219 | 344 (342-346) |
| Bipolar disorder | 11,764 | 34 (33-34) |
| Brain tumors | 13,152 | 38 (37-38) |
| Cerebral palsy | 2,143 | 6 (6-6) |
| Dementia | 52,791 | 151 (150-152) |
| Depression | 204,398 | 585 (582-587) |
| Developmental and behavioural disorders | 91,100 | 261 (259-262) |
| Drug abuse | 20,848 | 60 (59-60) |
| Eating disorders | 6,916 | 20 (19-20) |
| Epilepsy | 22,387 | 64 (63-65) |
| Headache | 118,820 | 340 (338-342) |
| Infections of the CNS | 7,672 | 22 (21-22) |
| Intellectual disability | 7,689 | 22 (22-22) |
| Multiple sclerosis | 4,745 | 14 (13-14) |
| Neuromuscular disorders | 3,324 | 10 (9-10) |
| Other neurodegenerative disorders | 2,683 | 8 (7-8) |
| Parkinson's disease | 13,996 | 40 (39-41) |
| Personality disorders | 19,657 | 56 (55-57) |
| Polyneuropathy | 24,544 | 70 (69-71) |
| Schizophrenia spectrum disorders | 23,479 | 67 (66-68) |
| Sleep disorders | 319,314 | 914 (910-917) |
| Stress related disorders | 95,899 | 274 (273-276) |
| Stroke | 84,569 | 242 (240-244) |
| Traumatic brain injury | 78,568 | 225 (223-226) |

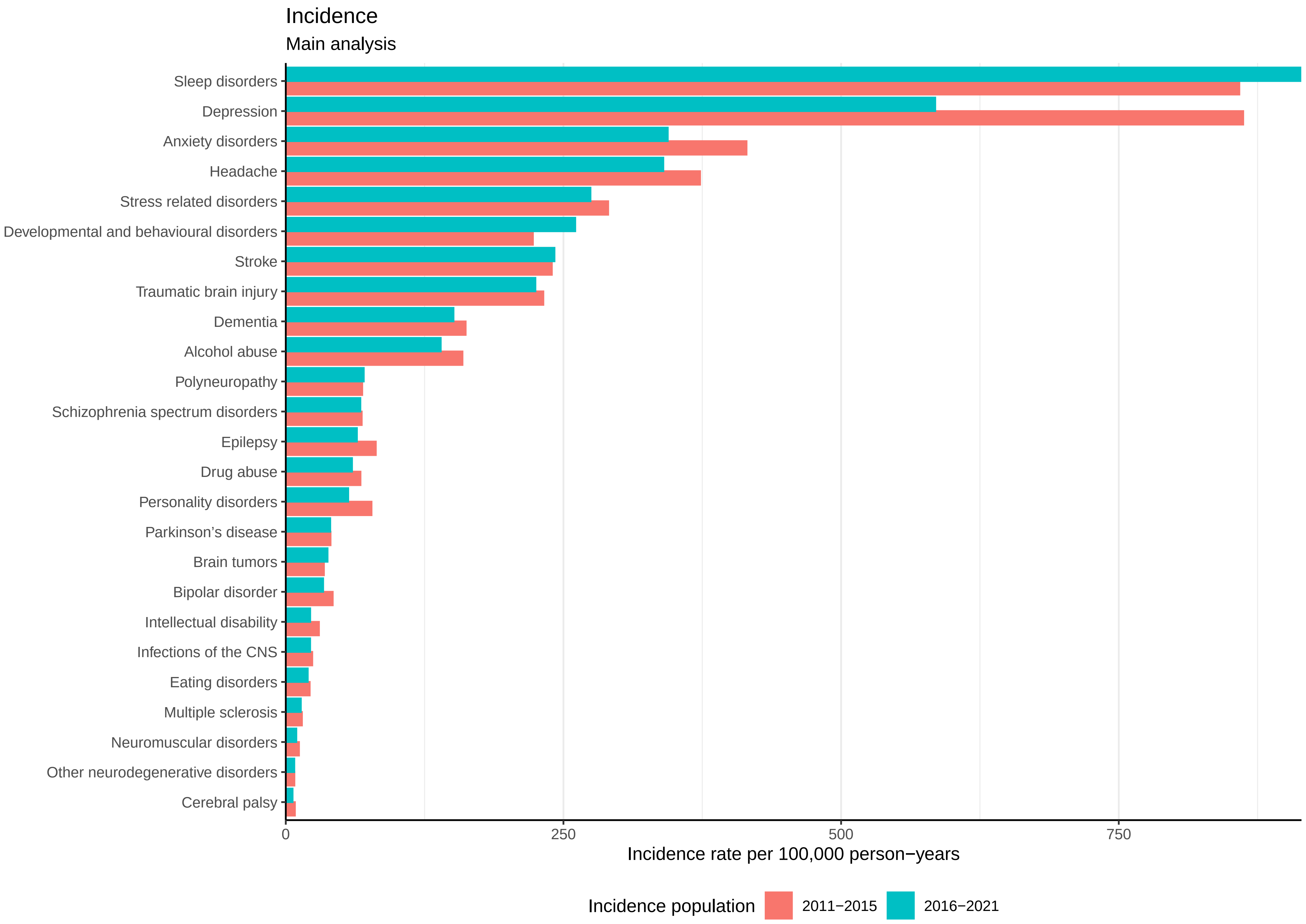
Incidence
Main analysis
Sleep disorders
Depression
Anxiety disorders
Headache
Stress related disorders
Developmental and behavioural disorders
Stroke
Traumatic brain injury
Dementia
Alcohol abuse
Polyneuropathy
Schizophrenia spectrum disorders
Epilepsy
Drug abuse
Personality disorders
Parkinson's disease
Brain tumors
Bipolar disorder
Intellectual disability
Infections of the CNS
Eating disorders
Multiple sclerosis
Neuromuscular disorders
Other neurodegenerative disorders
Cerebral palsy
0
250
500
750
Incidence rate per 100,000 person−years
Incidence population
2011−2015
2016−2021
