## Appendix 6 for "Temporal trends in the occurrence, mortality, and cost of brain disorders in Denmark"

| *Brain disorder* | *Patient group* | | *Comparison group* | | |
|---|---|---|---|---|---|
| | *Persons, N* | *Deaths, N(%)* | *Persons, N* | *Deaths, N(%)* | *Adjusted HR (95% CI)* |
| Any brain disorder | 505,611 | 28,013 (5.5) | 5,047,497 | 49,696 (1.0) | 5.5 (5.4-5.6) |
| Alcohol abuse | 44,942 | 3,109 (6.9) | 448,984 | 4,355 (1.0) | 6.5 (6.2-6.9) |
| Anxiety disorders | 117,257 | 9,584 (8.2) | 1,169,145 | 13,980 (1.2) | 5.6 (5.5-5.8) |
| Bipolar disorder | 11,937 | 261 (2.2) | 119,046 | 773 (0.6) | 2.8 (2.5-3.3) |
| Brain tumors | 9,721 | 2,293 (23.6) | 97,200 | 2,077 (2.1) | 12.8 (12.1-13.6) |
| Cerebral palsy | 2,331 | 144 (6.2) | 23,235 | 172 (0.7) | 6.8 (5.4-8.6) |
| Dementia | 45,887 | 8,608 (18.8) | 458,791 | 30,835 (6.7) | 3.0 (3.0-3.1) |
| Depression | 243,549 | 16,978 (7.0) | 2,429,771 | 38,862 (1.6) | 3.8 (3.7-3.9) |
| Developmental and behavioural disorders | 62,876 | 596 (0.9) | 627,022 | 619 (0.1) | 7.6 (6.7-8.5) |
| Drug abuse | 18,968 | 459 (2.4) | 189,034 | 782 (0.4) | 4.9 (4.3-5.5) |
| Eating disorders | 6,087 | 14 (0.2) | 60,492 | 32 (0.1) | 4.9 (2.6-9.4) |
| Epilepsy | 22,907 | 2,776 (12.1) | 229,083 | 4,167 (1.8) | 5.2 (4.9-5.5) |
| Headache | 105,365 | 938 (0.9) | 1,050,242 | 3,616 (0.3) | 2.0 (1.9-2.2) |
| Infections of the CNS | 6,733 | 709 (10.5) | 67,224 | 858 (1.3) | 8.2 (7.4-9.1) |
| Intellectual disability | 8,450 | 181 (2.1) | 84,228 | 211 (0.3) | 8.6 (7.1-10.5) |
| Multiple sclerosis | 4,110 | 59 (1.4) | 40,986 | 144 (0.4) | 4.0 (3.0-5.5) |
| Neuromuscular disorders | 3,368 | 293 (8.7) | 33,627 | 447 (1.3) | 5.7 (4.9-6.7) |
| Other neurodegenerative disorders | 2,197 | 418 (19.0) | 21,946 | 442 (2.0) | 10.7 (9.3-12.2) |
| Parkinson's disease | 11,421 | 773 (6.8) | 114,143 | 4,008 (3.5) | 1.9 (1.7-2.0) |
| Personality disorders | 21,832 | 153 (0.7) | 216,677 | 369 (0.2) | 3.1 (2.6-3.8) |
| Polyneuropathy | 19,498 | 1,087 (5.6) | 194,792 | 4,868 (2.5) | 1.7 (1.6-1.8) |
| Schizophrenia spectrum disorders | 19,269 | 593 (3.1) | 192,355 | 1,686 (0.9) | 3.4 (3.1-3.7) |
| Sleep disorders | 242,726 | 16,699 (6.9) | 2,423,741 | 35,150 (1.5) | 4.0 (3.9-4.1) |
| Stress related disorders | 81,999 | 1,243 (1.5) | 817,602 | 3,371 (0.4) | 3.0 (2.8-3.2) |
| Stroke | 67,848 | 14,052 (20.7) | 678,201 | 28,724 (4.2) | 5.6 (5.5-5.7) |
| Traumatic brain injury | 65,532 | 3,851 (5.9) | 654,214 | 10,932 (1.7) | 3.6 (3.4-3.7) |

| *Brain disorder* | *Patient group* | | *Comparison group* | | |
|---|---|---|---|---|---|
| | *Persons, N* | *Deaths, N(%)* | *Persons, N* | *Deaths, N(%)* | *Adjusted HR (95% CI)* |
| Any brain disorder | 567,664 | 29,186 (5.1) | 5,669,442 | 56,268 (1.0) | 5.3 (5.2-5.3) |
| Alcohol abuse | 48,536 | 2,971 (6.1) | 485,215 | 4,299 (0.9) | 6.5 (6.2-6.9) |
| Anxiety disorders | 119,617 | 6,858 (5.7) | 1,192,680 | 11,549 (1.0) | 5.4 (5.3-5.6) |
| Bipolar disorder | 11,688 | 170 (1.5) | 116,643 | 592 (0.5) | 2.5 (2.1-3.0) |
| Brain tumors | 13,134 | 2,783 (21.2) | 131,183 | 2,910 (2.2) | 11.3 (10.7-11.9) |
| Cerebral palsy | 2,133 | 142 (6.7) | 21,330 | 166 (0.8) | 8.0 (6.4-10.1) |
| Dementia | 52,762 | 8,052 (15.3) | 527,650 | 32,683 (6.2) | 2.7 (2.6-2.7) |
| Depression | 203,290 | 15,436 (7.6) | 2,029,831 | 34,990 (1.7) | 4.1 (4.0-4.2) |
| Developmental and behavioural disorders | 90,722 | 432 (0.5) | 903,961 | 775 (0.1) | 4.8 (4.3-5.4) |
| Drug abuse | 20,722 | 449 (2.2) | 206,126 | 631 (0.3) | 6.1 (5.4-6.9) |
| Eating disorders | 6,884 | 18 (0.3) | 68,550 | 26 (0.0) | 8.6 (4.6-16.0) |
| Epilepsy | 22,295 | 2,772 (12.4) | 223,007 | 4,260 (1.9) | 5.5 (5.2-5.7) |
| Headache | 118,174 | 722 (0.6) | 1,177,908 | 3,751 (0.3) | 1.7 (1.5-1.8) |
| Infections of the CNS | 7,651 | 758 (9.9) | 76,303 | 1,030 (1.3) | 7.6 (6.9-8.3) |
| Intellectual disability | 7,663 | 135 (1.8) | 76,393 | 162 (0.2) | 8.4 (6.7-10.6) |
| Multiple sclerosis | 4,722 | 36 (0.8) | 47,101 | 165 (0.4) | 2.1 (1.5-3.0) |
| Neuromuscular disorders | 3,318 | 188 (5.7) | 33,099 | 444 (1.3) | 3.9 (3.3-4.7) |
| Other neurodegenerative disorders | 2,680 | 570 (21.3) | 26,779 | 484 (1.8) | 13.9 (12.3-15.7) |
| Parkinson’s disease | 13,982 | 836 (6.0) | 139,783 | 4,834 (3.5) | 1.7 (1.6-1.8) |
| Personality disorders | 19,604 | 75 (0.4) | 194,153 | 216 (0.1) | 2.9 (2.2-3.8) |
| Polyneuropathy | 24,518 | 1,277 (5.2) | 244,947 | 6,133 (2.5) | 1.5 (1.4-1.6) |
| Schizophrenia spectrum disorders | 23,308 | 509 (2.2) | 232,484 | 1,251 (0.5) | 3.9 (3.5-4.3) |
| Sleep disorders | 318,207 | 20,113 (6.3) | 3,175,163 | 42,825 (1.3) | 4.3 (4.2-4.4) |
| Stress related disorders | 95,445 | 1,156 (1.2) | 951,691 | 3,506 (0.4) | 2.8 (2.6-3.0) |
| Stroke | 84,507 | 15,736 (18.6) | 844,596 | 35,403 (4.2) | 5.1 (5.0-5.2) |
| Traumatic brain injury | 78,275 | 5,723 (7.3) | 781,137 | 16,834 (2.2) | 3.6 (3.4-3.7) |

### Mortality
Main analysis / Incident cohort 2011-2015

| Brain disorder | Patient group -Deaths (%) | Comparison group- Deaths (%) | Hazard Ratio | Adjusted hazard ratio (95% CI) |
|---|---|---|---|---|
| Any brain disorder | 28,013 (5.5) | 49,696 (1.0) | | 5.5 (5.4-5.6) |
| Brain tumors | 2,293 (23.6) | 2,077 (2.1) | | 12.8 (12.1-13.6) |
| Other neurodegenerative disorders | 418 (19.0) | 442 (2.0) | | 10.7 (9.3-12.2) |
| Intellectual disability | 181 (2.1) | 211 (0.3) | | 8.6 (7.1-10.5) |
| Infections of the CNS | 709 (10.5) | 858 (1.3) | | 8.2 (7.4-9.1) |
| Developmental and behavioural disorders | 596 (0.9) | 619 (0.1) | | 7.6 (6.7-8.5) |
| Cerebral palsy | 144 (6.2) | 172 (0.7) | | 6.8 (5.4-8.6) |
| Alcohol abuse | 3,109 (6.9) | 4,355 (1.0) | | 6.5 (6.2-6.9) |
| Neuromuscular disorders | 293 (8.7) | 447 (1.3) | | 5.7 (4.9-6.7) |
| Stroke | 14,052 (20.7) | 28,724 (4.2) | | 5.6 (5.5-5.7) |
| Anxiety disorders | 9,584 (8.2) | 13,980 (1.2) | | 5.6 (5.5-5.8) |
| Epilepsy | 2,776 (12.1) | 4,167 (1.8) | | 5.2 (4.9-5.5) |
| Eating disorders | 14 (0.2) | 32 (0.1) | | 4.9 (2.6-9.4) |
| Drug abuse | 459 (2.4) | 782 (0.4) | | 4.9 (4.3-5.5) |
| Multiple sclerosis | 59 (1.4) | 144 (0.4) | | 4.0 (3.0-5.5) |
| Sleep disorders | 16,699 (6.9) | 35,150 (1.5) | | 4.0 (3.9-4.1) |
| Depression | 16,978 (7.0) | 38,862 (1.6) | | 3.8 (3.7-3.9) |
| Traumatic brain injury | 3,851 (5.9) | 10,932 (1.7) | | 3.6 (3.4-3.7) |
| Schizophrenia spectrum disorders | 593 (3.1) | 1,686 (0.9) | | 3.4 (3.1-3.7) |
| Personality disorders | 153 (0.7) | 369 (0.2) | | 3.1 (2.6-3.8) |
| Dementia | 8,608 (18.8) | 30,835 (6.7) | | 3.0 (3.0-3.1) |
| Stress related disorders | 1,243 (1.5) | 3,371 (0.4) | | 3.0 (2.8-3.2) |
| Bipolar disorder | 261 (2.2) | 773 (0.6) | | 2.8 (2.5-3.3) |
| Headache | 938 (0.9) | 3,616 (0.3) | | 2.0 (1.9-2.2) |
| Parkinson’s disease | 773 (6.8) | 4,008 (3.5) | | 1.9 (1.7-2.0) |
| Polyneuropathy | 1,087 (5.6) | 4,868 (2.5) | | 1.7 (1.6-1.8) |

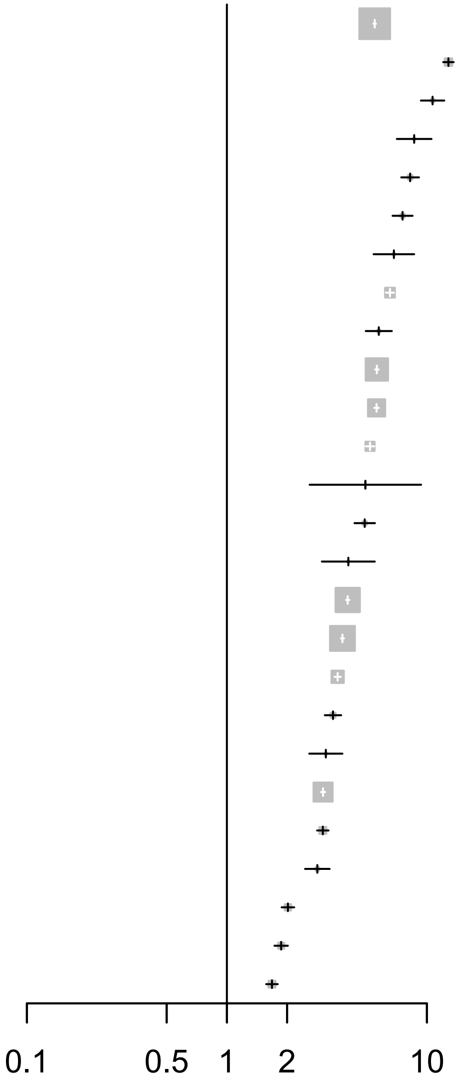

### Mortality

Main analysis / Incident cohort 2016-2021

| Brain disorder | Patient group -Deaths (%) | Comparison group- Deaths (%) | Adjusted hazard ratio (95% CI) |
|---|---|---|---|
| Any brain disorder | 29,186 (5.1) | 56,268 (1.0) | 5.3 (5.2-5.3) |
| Other neurodegenerative disorders | 570 (21.3) | 484 (1.8) | 13.9 (12.3-15.7) |
| Brain tumors | 2,783 (21.2) | 2,910 (2.2) | 11.3 (10.7-11.9) |
| Eating disorders | 18 (0.3) | 26 (0.0) | 8.6 (4.6-16.0) |
| Intellectual disability | 135 (1.8) | 162 (0.2) | 8.4 (6.7-10.6) |
| Cerebral palsy | 142 (6.7) | 166 (0.8) | 8.0 (6.4-10.1) |
| Infections of the CNS | 758 (9.9) | 1,030 (1.3) | 7.6 (6.9-8.3) |
| Alcohol abuse | 2,971 (6.1) | 4,299 (0.9) | 6.5 (6.2-6.9) |
| Drug abuse | 449 (2.2) | 631 (0.3) | 6.1 (5.4-6.9) |
| Epilepsy | 2,772 (12.4) | 4,260 (1.9) | 5.5 (5.2-5.7) |
| Anxiety disorders | 6,858 (5.7) | 11,549 (1.0) | 5.4 (5.3-5.6) |
| Stroke | 15,736 (18.6) | 35,403 (4.2) | 5.1 (5.0-5.2) |
| Developmental and behavioural disorders | 432 (0.5) | 775 (0.1) | 4.8 (4.3-5.4) |
| Sleep disorders | 20,113 (6.3) | 42,825 (1.3) | 4.3 (4.2-4.4) |
| Depression | 15,436 (7.6) | 34,990 (1.7) | 4.1 (4.0-4.2) |
| Neuromuscular disorders | 188 (5.7) | 444 (1.3) | 3.9 (3.3-4.7) |
| Schizophrenia spectrum disorders | 509 (2.2) | 1,251 (0.5) | 3.9 (3.5-4.3) |
| Traumatic brain injury | 5,723 (7.3) | 16,834 (2.2) | 3.6 (3.4-3.7) |
| Personality disorders | 75 (0.4) | 216 (0.1) | 2.9 (2.2-3.8) |
| Stress related disorders | 1,156 (1.2) | 3,506 (0.4) | 2.8 (2.6-3.0) |
| Dementia | 8,052 (15.3) | 32,683 (6.2) | 2.7 (2.6-2.7) |
| Bipolar disorder | 170 (1.5) | 592 (0.5) | 2.5 (2.1-3.0) |
| Multiple sclerosis | 36 (0.8) | 165 (0.4) | 2.1 (1.5-3.0) |
| Parkinson’s disease | 836 (6.0) | 4,834 (3.5) | 1.7 (1.6-1.8) |
| Headache | 722 (0.6) | 3,751 (0.3) | 1.7 (1.5-1.8) |
| Polyneuropathy | 1,277 (5.2) | 6,133 (2.5) | 1.5 (1.4-1.6) |

Hazard Ratio

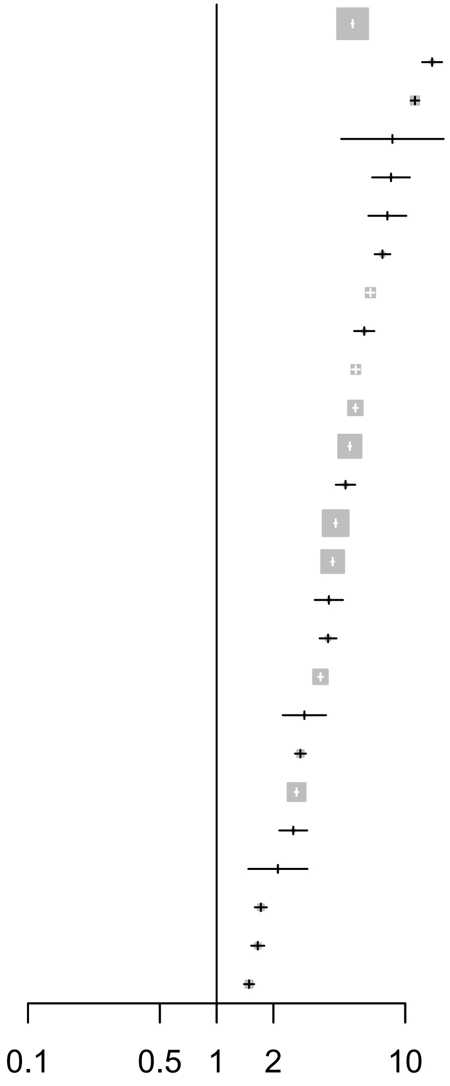

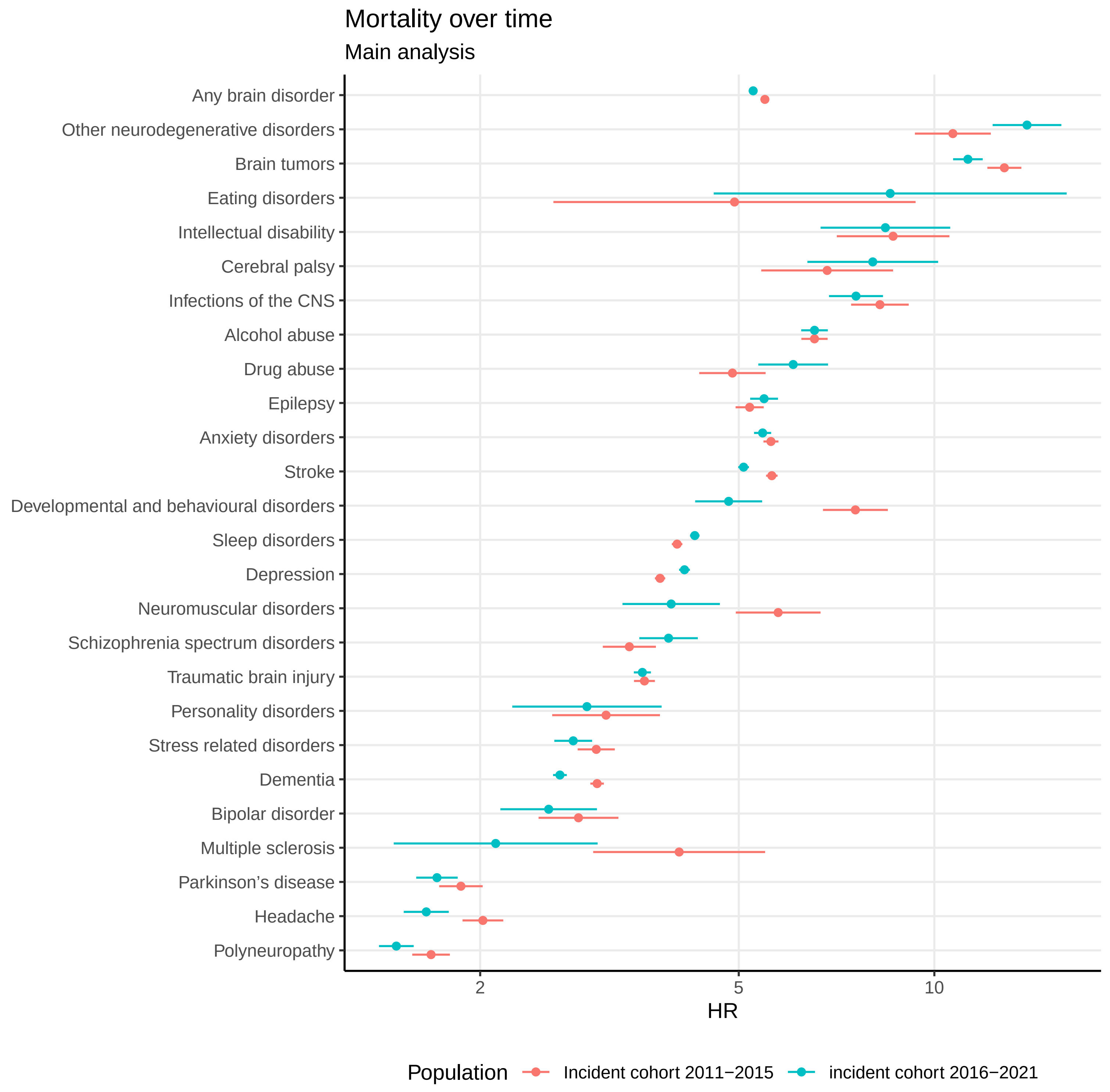
Mortality over time
Main analysis
Any brain disorder
Other neurodegenerative disorders
Brain tumors
Eating disorders
Intellectual disability
Cerebral palsy
Infections of the CNS
Alcohol abuse
Drug abuse
Epilepsy
Anxiety disorders
Stroke
Developmental and behavioural disorders
Sleep disorders
Depression
Neuromuscular disorders
Schizophrenia spectrum disorders
Traumatic brain injury
Personality disorders
Stress related disorders
Dementia
Bipolar disorder
Multiple sclerosis
Parkinson’s disease
Headache
Polyneuropathy
2
5
10
HR
Population
Incident cohort 2011−2015
incident cohort 2016−2021
