## Appendix 7 for "Temporal trends in the occurrence, mortality, and cost of brain disorders in Denmark"

***Main analysis / Incident cohort 2011-2015***

| *Brain disorder* | *Cost component* | *Actual costs (EUR)* | *Actual costs per person* | *Attributable costs (EUR)* | *Attributable costs per person* |
|---|---|---|---|---|---|
| Any brain disorder | Primary sector | 227,555,854 | 467 | 100,080,050 | 205 |
| | Hospital care | 4,770,420,309 | 9,794 | 3,959,386,556 | 8,129 |
| | Filled prescriptions | 179,427,338 | 368 | 99,280,589 | 204 |
| | Home care | 260,408,791 | 535 | 183,621,191 | 377 |
| | Nursing home / sheltered accomodation | 515,266,552 | 1,058 | 401,162,312 | 824 |
| | Lost productivity associated with illness | . | . | 2,779,679,495 | 9,795 |
| | Total cost (excl. lost productivity) | 5,953,078,844 | 12,222 | 4,743,530,698 | 9,739 |
| Alcohol abuse | Primary sector | 20,127,619 | 469 | 7,000,119 | 163 |
| | Hospital care | 668,578,170 | 15,594 | 569,677,811 | 13,287 |
| | Filled prescriptions | 24,342,314 | 568 | 12,869,818 | 300 |
| | Home care | 30,004,738 | 700 | 22,306,723 | 520 |
| | Nursing home / sheltered accomodation | 49,741,408 | 1,160 | 33,827,559 | 789 |
| | Lost productivity associated with illness | . | . | 762,934,471 | 25,045 |
| | Total cost (excl. lost productivity) | 792,794,250 | 18,491 | 645,682,030 | 15,059 |
| Anxiety disorders | Primary sector | 66,788,573 | 604 | 33,047,422 | 299 |
| | Hospital care | 1,151,652,888 | 10,419 | 911,210,152 | 8,244 |
| | Filled prescriptions | 75,114,298 | 680 | 48,973,145 | 443 |
| | Home care | 78,192,508 | 707 | 49,133,393 | 445 |
| | Nursing home / sheltered accomodation | 235,130,585 | 2,127 | 162,622,310 | 1,471 |
| | Lost productivity associated with illness | . | . | 1,562,894,213 | 19,940 |
| | Total cost (excl. lost productivity) | 1,606,878,852 | 14,538 | 1,204,986,421 | 10,902 |
| Bipolar disorder | Primary sector | 6,837,296 | 579 | 3,298,962 | 279 |
| | Hospital care | 308,206,933 | 26,106 | 283,900,568 | 24,047 |
| | Filled prescriptions | 12,139,655 | 1,028 | 9,489,822 | 804 |
| | Home care | 4,664,385 | 395 | 2,840,276 | 241 |
| | Nursing home / sheltered accomodation | 13,436,982 | 1,138 | 9,659,967 | 818 |
| | Lost productivity associated with illness | . | . | 283,038,554 | 28,506 |
| | Total cost (excl. lost productivity) | 345,285,250 | 29,246 | 309,189,595 | 26,189 |
| Brain tumors | Primary sector | 4,761,849 | 574 | 1,579,218 | 190 |
| | Hospital care | 412,782,835 | 49,756 | 387,192,567 | 46,671 |
| | Filled prescriptions | 5,507,517 | 664 | 2,391,849 | 288 |
| | Home care | 11,769,533 | 1,419 | 8,085,964 | 975 |
| | Nursing home / sheltered accomodation | 11,569,206 | 1,395 | 2,782,933 | 335 |

***Main analysis / Incident cohort 2011-2015***

| Brain disorder | Cost component | Actual costs (EUR) | Actual costs per person | Attributable costs (EUR) | Attributable costs per person |
|---|---|---|---|---|---|
| | Lost productivity associated with illness | . | . | 21,102,148 | 5,700 |
| | Total cost (excl. lost productivity) | 446,390,939 | 53,807 | 402,032,531 | 48,460 |
| Cerebral palsy | Primary sector | 1,456,451 | 652 | 861,951 | 386 |
| | Hospital care | 39,147,738 | 17,536 | 35,730,055 | 16,006 |
| | Filled prescriptions | 1,595,967 | 715 | 1,231,165 | 552 |
| | Home care | 5,813,039 | 2,604 | 5,487,033 | 2,458 |
| | Nursing home / sheltered accomodation | 8,193,681 | 3,670 | 7,511,856 | 3,365 |
| | Lost productivity associated with illness | . | . | 22,858,092 | 36,573 |
| | Total cost (excl. lost productivity) | 56,206,876 | 25,178 | 50,822,061 | 22,766 |
| Dementia | Primary sector | 25,705,785 | 635 | 4,567,214 | 113 |
| | Hospital care | 539,446,726 | 13,335 | 341,018,488 | 8,430 |
| | Filled prescriptions | 46,605,754 | 1,152 | 23,216,390 | 574 |
| | Home care | 214,584,171 | 5,305 | 143,578,706 | 3,549 |
| | Nursing home / sheltered accomodation | 745,671,823 | 18,433 | 599,057,356 | 14,809 |
| | Lost productivity associated with illness | . | . | 93,206,727 | 35,346 |
| | Total cost (excl. lost productivity) | 1,572,014,259 | 38,860 | 1,111,438,155 | 27,475 |
| Depression | Primary sector | 147,640,690 | 634 | 74,693,347 | 321 |
| | Hospital care | 2,044,238,520 | 8,774 | 1,522,315,097 | 6,534 |
| | Filled prescriptions | 136,162,274 | 584 | 80,245,217 | 344 |
| | Home care | 237,725,249 | 1,020 | 161,067,222 | 691 |
| | Nursing home / sheltered accomodation | 731,794,154 | 3,141 | 571,100,050 | 2,451 |
| | Lost productivity associated with illness | . | . | 2,964,198,806 | 18,844 |
| | Total cost (excl. lost productivity) | 3,297,560,886 | 14,154 | 2,409,420,933 | 10,342 |
| Developmental and behavioural disorders | Primary sector | 25,463,549 | 407 | 14,521,833 | 232 |
| | Hospital care | 588,260,407 | 9,403 | 523,086,546 | 8,361 |
| | Filled prescriptions | 45,256,709 | 723 | 39,951,000 | 639 |
| | Home care | 3,299,134 | 53 | 2,235,918 | 36 |
| | Nursing home / sheltered accomodation | 6,841,371 | 109 | 5,008,830 | 80 |
| | Lost productivity associated with illness | . | . | 588,601,731 | 22,180 |
| | Total cost (excl. lost productivity) | 669,121,169 | 10,696 | 584,804,127 | 9,348 |
| Drug abuse | Primary sector | 7,215,732 | 385 | 3,113,579 | 166 |
| | Hospital care | 420,316,215 | 22,454 | 392,737,383 | 20,981 |
| | Filled prescriptions | 13,185,955 | 704 | 10,349,214 | 553 |

*Main analysis / Incident cohort 2011-2015*

| *Brain disorder* | *Cost component* | *Actual costs (EUR)* | *Actual costs per person* | *Attributable costs (EUR)* | *Attributable costs per person* |
|---|---|---|---|---|---|
| | Home care | 8,046,416 | 430 | 6,306,102 | 337 |
| | Nursing home / sheltered accomodation | 10,599,399 | 566 | 6,547,935 | 350 |
| | Lost productivity associated with illness | . | . | 423,394,446 | 27,504 |
| | Total cost (excl. lost productivity) | 459,363,716 | 24,540 | 419,054,214 | 22,387 |
| Eating disorders | Primary sector | 2,105,606 | 346 | 643,700 | 106 |
| | Hospital care | 220,014,012 | 36,140 | 210,934,156 | 34,649 |
| | Filled prescriptions | 1,384,863 | 227 | 630,977 | 104 |
| | Home care | 338,539 | 56 | 247,205 | 41 |
| | Nursing home / sheltered accomodation | 447,829 | 74 | 374,603 | 62 |
| | Lost productivity associated with illness | . | . | 26,789,067 | 7,268 |
| | Total cost (excl. lost productivity) | 224,290,850 | 36,843 | 212,830,640 | 34,960 |
| Epilepsy | Primary sector | 11,405,817 | 540 | 4,595,806 | 218 |
| | Hospital care | 448,947,402 | 21,253 | 395,207,916 | 18,709 |
| | Filled prescriptions | 17,250,052 | 817 | 11,319,395 | 536 |
| | Home care | 39,717,638 | 1,880 | 31,910,613 | 1,511 |
| | Nursing home / sheltered accomodation | 99,465,592 | 4,709 | 79,519,465 | 3,764 |
| | Lost productivity associated with illness | . | . | 188,581,749 | 21,759 |
| | Total cost (excl. lost productivity) | 616,786,502 | 29,198 | 522,553,195 | 24,737 |
| Headache | Primary sector | 47,860,098 | 456 | 18,234,818 | 174 |
| | Hospital care | 425,340,165 | 4,053 | 232,724,089 | 2,217 |
| | Filled prescriptions | 36,059,780 | 344 | 16,392,602 | 156 |
| | Home care | 9,730,769 | 93 | 971,074 | 9 |
| | Nursing home / sheltered accomodation | 12,262,165 | 117 | -2,851,121 | -27 |
| | Lost productivity associated with illness | . | . | 324,186,786 | 3,779 |
| | Total cost (excl. lost productivity) | 531,252,977 | 5,062 | 265,471,463 | 2,529 |
| Infections of the CNS | Primary sector | 2,905,752 | 470 | 992,094 | 160 |
| | Hospital care | 222,855,724 | 36,026 | 209,479,876 | 33,864 |
| | Filled prescriptions | 3,075,685 | 497 | 1,614,429 | 261 |
| | Home care | 7,520,277 | 1,216 | 6,211,741 | 1,004 |
| | Nursing home / sheltered accomodation | 9,567,627 | 1,547 | 6,154,355 | 995 |
| | Lost productivity associated with illness | . | . | 16,445,087 | 4,959 |
| | Total cost (excl. lost productivity) | 245,925,064 | 39,756 | 224,452,496 | 36,284 |
| Intellectual disability | Primary sector | 3,255,411 | 390 | 1,532,396 | 184 |
| | Hospital care | 129,888,412 | 15,573 | 118,713,461 | 14,234 |
| | Filled prescriptions | 5,653,343 | 678 | 4,596,678 | 551 |
| | Home care | 4,159,806 | 499 | 3,660,270 | 439 |
| | Nursing home / sheltered accomodation | 7,661,621 | 919 | 6,859,432 | 822 |
| | Lost productivity associated with illness | . | . | 119,663,543 | 34,356 |
| | Total cost (excl. lost productivity) | 150,618,592 | 18,059 | 135,362,238 | 16,230 |

***Main analysis / Incident cohort 2011-2015***

| *Brain disorder* | *Cost component* | *Actual costs (EUR)* | *Actual costs per person* | *Attributable costs (EUR)* | *Attributable costs per person* |
|---|---|---|---|---|---|
| Multiple sclerosis | Primary sector | 2,434,817 | 597 | 1,201,017 | 294 |
| | Hospital care | 76,976,497 | 18,866 | 68,832,811 | 16,870 |
| | Filled prescriptions | 1,569,360 | 385 | 703,711 | 172 |
| | Home care | 2,821,710 | 692 | 2,495,673 | 612 |
| | Nursing home / sheltered accomodation | 1,972,382 | 483 | 1,510,507 | 370 |
| | Lost productivity associated with illness | . | . | 31,875,001 | 8,655 |
| | Total cost (excl. lost productivity) | 85,774,765 | 21,022 | 74,743,719 | 18,319 |
| Neuromuscular disorders | Primary sector | 1,760,759 | 557 | 721,167 | 228 |
| | Hospital care | 73,254,387 | 23,194 | 65,296,441 | 20,674 |
| | Filled prescriptions | 2,097,662 | 664 | 1,198,621 | 380 |
| | Home care | 2,978,477 | 943 | 2,146,196 | 680 |
| | Nursing home / sheltered accomodation | 2,672,693 | 846 | 652,864 | 207 |
| | Lost productivity associated with illness | . | . | 15,526,631 | 8,996 |
| | Total cost (excl. lost productivity) | 82,763,979 | 26,205 | 70,015,290 | 22,168 |
| Other neurodegenerative disorders | Primary sector | 1,527,685 | 778 | 777,241 | 396 |
| | Hospital care | 43,893,296 | 22,344 | 37,640,543 | 19,161 |
| | Filled prescriptions | 2,766,029 | 1,408 | 2,046,494 | 1,042 |
| | Home care | 8,733,058 | 4,446 | 7,978,552 | 4,062 |
| | Nursing home / sheltered accomodation | 7,954,013 | 4,049 | 6,139,863 | 3,126 |
| | Lost productivity associated with illness | . | . | 14,740,786 | 18,850 |
| | Total cost (excl. lost productivity) | 64,874,080 | 33,025 | 54,582,694 | 27,786 |
| Parkinson’s disease | Primary sector | 10,071,539 | 916 | 5,099,499 | 464 |
| | Hospital care | 105,739,654 | 9,618 | 60,073,635 | 5,464 |
| | Filled prescriptions | 13,560,034 | 1,233 | 8,394,289 | 764 |
| | Home care | 29,255,932 | 2,661 | 21,302,649 | 1,938 |
| | Nursing home / sheltered accomodation | 57,170,923 | 5,200 | 35,278,719 | 3,209 |
| | Lost productivity associated with illness | . | . | 61,545,896 | 21,989 |
| | Total cost (excl. lost productivity) | 215,798,082 | 19,628 | 130,148,790 | 11,838 |
| Personality disorders | Primary sector | 9,545,569 | 438 | 3,904,986 | 179 |
| | Hospital care | 446,637,130 | 20,510 | 413,666,858 | 18,996 |
| | Filled prescriptions | 11,972,493 | 550 | 8,791,843 | 404 |
| | Home care | 2,264,370 | 104 | 1,279,220 | 59 |
| | Nursing home / sheltered accomodation | 6,441,433 | 296 | 5,170,930 | 237 |
| | Lost productivity associated with illness | . | . | 495,764,001 | 25,530 |
| | Total cost (excl. lost productivity) | 476,860,997 | 21,898 | 432,813,836 | 19,875 |
| Polyneuropathy | Primary sector | 12,769,046 | 676 | 5,208,213 | 276 |
| | Hospital care | 317,261,896 | 16,796 | 250,719,443 | 13,273 |
| | Filled prescriptions | 18,651,987 | 987 | 10,993,641 | 582 |
| | Home care | 25,758,093 | 1,364 | 15,230,758 | 806 |

***Main analysis / Incident cohort 2011-2015***

| *Brain disorder* | *Cost component* | *Actual costs (EUR)* | *Actual costs per person* | *Attributable costs (EUR)* | *Attributable costs per person* |
|---|---|---|---|---|---|
| | Nursing home / sheltered accomodation | 22,875,427 | 1,211 | -3,837,710 | -203 |
| | Lost productivity associated with illness | . | . | 165,406,236 | 20,421 |
| | Total cost (excl. lost productivity) | 397,316,449 | 21,035 | 278,314,345 | 14,734 |
| Schizophrenia spectrum disorders | Primary sector | 7,166,569 | 378 | 2,361,166 | 125 |
| | Hospital care | 856,720,642 | 45,175 | 824,370,820 | 43,469 |
| | Filled prescriptions | 14,817,946 | 781 | 11,363,320 | 599 |
| | Home care | 11,054,388 | 583 | 7,324,063 | 386 |
| | Nursing home / sheltered accomodation | 39,060,090 | 2,060 | 28,979,372 | 1,528 |
| | Lost productivity associated with illness | . | . | 438,924,509 | 30,703 |
| | Total cost (excl. lost productivity) | 928,819,634 | 48,977 | 874,398,741 | 46,107 |
| Sleep disorders | Primary sector | 128,311,646 | 553 | 55,689,687 | 240 |
| | Hospital care | 2,214,649,835 | 9,549 | 1,684,335,338 | 7,263 |
| | Filled prescriptions | 150,202,935 | 648 | 91,700,250 | 395 |
| | Home care | 138,844,344 | 599 | 69,956,942 | 302 |
| | Nursing home / sheltered accomodation | 310,735,400 | 1,340 | 134,941,747 | 582 |
| | Lost productivity associated with illness | . | . | 1,181,197,040 | 8,153 |
| | Total cost (excl. lost productivity) | 2,942,744,159 | 12,689 | 2,036,623,964 | 8,782 |
| Stress related disorders | Primary sector | 40,181,998 | 494 | 19,320,599 | 237 |
| | Hospital care | 1,268,153,485 | 15,580 | 1,133,697,776 | 13,928 |
| | Filled prescriptions | 36,442,053 | 448 | 22,372,353 | 275 |
| | Home care | 19,644,954 | 241 | 11,939,881 | 147 |
| | Nursing home / sheltered accomodation | 44,277,134 | 544 | 28,989,774 | 356 |
| | Lost productivity associated with illness | . | . | 1,390,144,263 | 23,151 |
| | Total cost (excl. lost productivity) | 1,408,699,623 | 17,306 | 1,216,320,383 | 14,943 |
| Stroke | Primary sector | 37,626,711 | 664 | 13,182,929 | 233 |
| | Hospital care | 1,975,042,560 | 34,838 | 1,759,528,558 | 31,037 |
| | Filled prescriptions | 47,119,081 | 831 | 21,300,452 | 376 |
| | Home care | 145,036,426 | 2,558 | 98,926,906 | 1,745 |
| | Nursing home / sheltered accomodation | 307,135,751 | 5,418 | 176,341,087 | 3,111 |
| | Lost productivity associated with illness | . | . | 317,767,914 | 17,826 |
| | Total cost (excl. lost productivity) | 2,511,960,528 | 44,309 | 2,069,279,933 | 36,500 |
| Traumatic brain injury | Primary sector | 25,597,391 | 408 | 7,870,033 | 125 |
| | Hospital care | 752,293,178 | 11,981 | 623,438,991 | 9,929 |
| | Filled prescriptions | 21,990,657 | 350 | 7,788,645 | 124 |
| | Home care | 52,730,725 | 840 | 31,967,348 | 509 |
| | Nursing home / sheltered accomodation | 129,712,559 | 2,066 | 69,816,711 | 1,112 |
| | Lost productivity associated with illness | . | . | 252,141,703 | 8,441 |
| | Total cost (excl. lost productivity) | 982,324,510 | 15,644 | 740,881,727 | 11,799 |

***Main analysis / Incident cohort 2016-2021***

| *Brain disorder* | *Cost component* | *Actual costs (EUR)* | *Actual costs per person* | *Attributable costs (EUR)* | *Attributable costs per person* |
|---|---|---|---|---|---|
| Any brain disorder | Primary sector | 255,738,267 | 466 | 116,148,266 | 212 |
| | Hospital care | 4,670,782,367 | 8,506 | 3,843,965,581 | 7,000 |
| | Filled prescriptions | 199,055,918 | 362 | 104,374,577 | 190 |
| | Home care | 242,843,604 | 442 | 173,548,050 | 316 |
| | Nursing home / sheltered accomodation | 565,055,852 | 1,029 | 439,748,748 | 801 |
| | Lost productivity associated with illness | . | . | 3,138,405,046 | 10,148 |
| | Total cost (excl. lost productivity) | 5,933,476,008 | 10,805 | 4,677,785,222 | 8,519 |
| Alcohol abuse | Primary sector | 23,093,013 | 495 | 8,567,731 | 184 |
| | Hospital care | 563,948,956 | 12,090 | 464,901,830 | 9,967 |
| | Filled prescriptions | 26,841,605 | 575 | 14,370,515 | 308 |
| | Home care | 29,729,025 | 637 | 23,105,601 | 495 |
| | Nursing home / sheltered accomodation | 47,331,490 | 1,015 | 32,011,071 | 686 |
| | Lost productivity associated with illness | . | . | 911,372,384 | 27,125 |
| | Total cost (excl. lost productivity) | 690,944,089 | 14,813 | 542,956,748 | 11,640 |
| Anxiety disorders | Primary sector | 71,340,584 | 620 | 38,510,114 | 335 |
| | Hospital care | 913,278,634 | 7,942 | 707,046,096 | 6,149 |
| | Filled prescriptions | 53,157,093 | 462 | 29,094,645 | 253 |
| | Home care | 46,325,681 | 403 | 26,823,012 | 233 |
| | Nursing home / sheltered accomodation | 150,613,504 | 1,310 | 93,291,455 | 811 |
| | Lost productivity associated with illness | . | . | 1,650,365,166 | 20,144 |
| | Total cost (excl. lost productivity) | 1,234,715,496 | 10,737 | 894,765,321 | 7,781 |
| Bipolar disorder | Primary sector | 6,295,582 | 541 | 2,970,414 | 255 |
| | Hospital care | 229,967,740 | 19,776 | 208,969,504 | 17,971 |
| | Filled prescriptions | 7,602,213 | 654 | 5,276,260 | 454 |
| | Home care | 2,797,918 | 241 | 1,729,276 | 149 |
| | Nursing home / sheltered accomodation | 9,806,875 | 843 | 7,333,683 | 631 |
| | Lost productivity associated with illness | . | . | 259,189,392 | 26,149 |
| | Total cost (excl. lost productivity) | 256,470,328 | 22,056 | 226,279,137 | 19,459 |
| Brain tumors | Primary sector | 6,619,549 | 581 | 2,173,058 | 191 |
| | Hospital care | 472,356,911 | 41,443 | 440,373,452 | 38,637 |
| | Filled prescriptions | 7,621,185 | 669 | 3,290,005 | 289 |
| | Home care | 13,442,208 | 1,179 | 9,340,532 | 819 |
| | Nursing home / sheltered accomodation | 17,586,335 | 1,543 | 6,015,639 | 528 |

***Main analysis / Incident cohort 2016-2021***

| *Brain disorder* | *Cost component* | *Actual costs (EUR)* | *Actual costs per person* | *Attributable costs (EUR)* | *Attributable costs per person* |
|---|---|---|---|---|---|
| | Lost productivity associated with illness | . | . | 29,576,411 | 5,949 |
| | Total cost (excl. lost productivity) | 517,626,188 | 45,414 | 461,192,687 | 40,463 |
| Cerebral palsy | Primary sector | 1,297,043 | 636 | 744,065 | 365 |
| | Hospital care | 26,925,179 | 13,199 | 23,815,879 | 11,675 |
| | Filled prescriptions | 1,487,241 | 729 | 1,140,045 | 559 |
| | Home care | 3,320,685 | 1,628 | 3,113,529 | 1,526 |
| | Nursing home / sheltered accomodation | 6,258,268 | 3,068 | 5,484,914 | 2,689 |
| | Lost productivity associated with illness | . | . | 19,899,678 | 36,851 |
| | Total cost (excl. lost productivity) | 39,288,417 | 19,260 | 34,298,432 | 16,813 |
| Dementia | Primary sector | 32,288,482 | 673 | 6,418,108 | 134 |
| | Hospital care | 450,190,598 | 9,377 | 234,775,209 | 4,890 |
| | Filled prescriptions | 49,956,886 | 1,041 | 20,342,142 | 424 |
| | Home care | 207,839,827 | 4,329 | 146,145,107 | 3,044 |
| | Nursing home / sheltered accomodation | 747,831,571 | 15,576 | 606,045,976 | 12,623 |
| | Lost productivity associated with illness | . | . | 103,942,279 | 42,201 |
| | Total cost (excl. lost productivity) | 1,488,107,363 | 30,995 | 1,013,726,543 | 21,114 |
| Depression | Primary sector | 131,420,124 | 678 | 71,085,844 | 367 |
| | Hospital care | 1,543,980,263 | 7,965 | 1,141,431,726 | 5,888 |
| | Filled prescriptions | 104,438,802 | 539 | 55,821,334 | 288 |
| | Home care | 180,651,580 | 932 | 125,139,262 | 646 |
| | Nursing home / sheltered accomodation | 688,763,255 | 3,553 | 543,331,593 | 2,803 |
| | Lost productivity associated with illness | . | . | 2,632,516,867 | 20,697 |
| | Total cost (excl. lost productivity) | 2,649,254,024 | 13,666 | 1,936,809,759 | 9,991 |
| Developmental and behavioural disorders | Primary sector | 43,330,306 | 478 | 27,808,976 | 307 |
| | Hospital care | 569,885,114 | 6,288 | 487,294,102 | 5,377 |
| | Filled prescriptions | 47,847,887 | 528 | 40,299,798 | 445 |
| | Home care | 3,504,767 | 39 | 2,194,967 | 24 |
| | Nursing home / sheltered accomodation | 9,566,997 | 106 | 7,413,556 | 82 |
| | Lost productivity associated with illness | . | . | 812,829,536 | 19,875 |
| | Total cost (excl. lost productivity) | 674,135,070 | 7,438 | 565,011,399 | 6,234 |
| Drug abuse | Primary sector | 7,354,068 | 359 | 3,055,771 | 149 |
| | Hospital care | 355,854,617 | 17,362 | 330,765,493 | 16,138 |
| | Filled prescriptions | 9,098,649 | 444 | 6,411,152 | 313 |

***Main analysis / Incident cohort 2016-2021***

| *Brain disorder* | *Cost component* | *Actual costs (EUR)* | *Actual costs per person* | *Attributable costs (EUR)* | *Attributable costs per person* |
|---|---|---|---|---|---|
| | Home care | 4,854,010 | 237 | 3,694,627 | 180 |
| | Nursing home / sheltered accomodation | 7,495,050 | 366 | 4,812,176 | 235 |
| | Lost productivity associated with illness | . | . | 493,472,840 | 28,697 |
| | Total cost (excl. lost productivity) | 384,656,395 | 18,768 | 348,739,219 | 17,015 |
| Eating disorders | Primary sector | 2,093,952 | 304 | 546,355 | 79 |
| | Hospital care | 212,916,135 | 30,893 | 204,146,053 | 29,620 |
| | Filled prescriptions | 1,136,640 | 165 | 397,679 | 58 |
| | Home care | 156,002 | 23 | 107,184 | 16 |
| | Nursing home / sheltered accomodation | 66,796 | 10 | 34,280 | 5 |
| | Lost productivity associated with illness | . | . | 26,564,779 | 6,980 |
| | Total cost (excl. lost productivity) | 216,369,525 | 31,394 | 205,231,551 | 29,778 |
| Epilepsy | Primary sector | 10,760,119 | 523 | 4,067,519 | 198 |
| | Hospital care | 387,367,955 | 18,839 | 339,566,835 | 16,514 |
| | Filled prescriptions | 17,019,350 | 828 | 11,053,768 | 538 |
| | Home care | 35,132,939 | 1,709 | 28,703,812 | 1,396 |
| | Nursing home / sheltered accomodation | 99,953,751 | 4,861 | 80,808,758 | 3,930 |
| | Lost productivity associated with illness | . | . | 167,555,032 | 22,175 |
| | Total cost (excl. lost productivity) | 550,234,115 | 26,760 | 464,200,692 | 22,576 |
| Headache | Primary sector | 52,594,617 | 446 | 20,815,708 | 176 |
| | Hospital care | 390,769,082 | 3,310 | 197,144,327 | 1,670 |
| | Filled prescriptions | 33,691,506 | 285 | 13,571,750 | 115 |
| | Home care | 7,864,997 | 67 | 203,977 | 2 |
| | Nursing home / sheltered accomodation | 12,665,253 | 107 | -3,460,548 | -29 |
| | Lost productivity associated with illness | . | . | 366,106,090 | 3,807 |
| | Total cost (excl. lost productivity) | 497,585,455 | 4,215 | 228,275,215 | 1,934 |
| Infections of the CNS | Primary sector | 3,358,238 | 475 | 1,157,975 | 164 |
| | Hospital care | 238,992,713 | 33,826 | 224,054,679 | 31,712 |
| | Filled prescriptions | 3,638,487 | 515 | 1,872,121 | 265 |
| | Home care | 6,017,033 | 852 | 4,560,057 | 645 |
| | Nursing home / sheltered accomodation | 10,889,900 | 1,541 | 6,779,617 | 960 |
| | Lost productivity associated with illness | . | . | 23,498,353 | 6,007 |
| | Total cost (excl. lost productivity) | 262,896,371 | 37,209 | 238,424,450 | 33,745 |
| Intellectual disability | Primary sector | 2,671,418 | 352 | 1,188,631 | 157 |
| | Hospital care | 98,695,951 | 13,016 | 90,588,554 | 11,947 |
| | Filled prescriptions | 3,999,727 | 527 | 3,151,028 | 416 |
| | Home care | 2,656,661 | 350 | 2,411,214 | 318 |
| | Nursing home / sheltered accomodation | 6,166,896 | 813 | 5,640,297 | 744 |
| | Lost productivity associated with illness | . | . | 116,149,905 | 36,896 |
| | Total cost (excl. lost productivity) | 114,190,653 | 15,060 | 102,979,725 | 13,581 |

***Main analysis / Incident cohort 2016-2021***

| *Brain disorder* | *Cost component* | *Actual costs (EUR)* | *Actual costs per person* | *Attributable costs (EUR)* | *Attributable costs per person* |
|---|---|---|---|---|---|
| Multiple sclerosis | Primary sector | 2,785,855 | 591 | 1,403,061 | 298 |
| | Hospital care | 58,963,034 | 12,508 | 50,423,721 | 10,697 |
| | Filled prescriptions | 1,423,027 | 302 | 460,620 | 98 |
| | Home care | 2,918,357 | 619 | 2,656,066 | 563 |
| | Nursing home / sheltered accomodation | 1,607,313 | 341 | 1,056,916 | 224 |
| | Lost productivity associated with illness | . | . | 33,746,491 | 7,946 |
| | Total cost (excl. lost productivity) | 67,697,585 | 14,361 | 56,000,384 | 11,880 |
| Neuromuscular disorders | Primary sector | 1,828,515 | 572 | 746,511 | 234 |
| | Hospital care | 52,102,956 | 16,308 | 44,164,341 | 13,823 |
| | Filled prescriptions | 2,230,134 | 698 | 1,249,926 | 391 |
| | Home care | 3,195,135 | 1,000 | 2,330,983 | 730 |
| | Nursing home / sheltered accomodation | 2,751,462 | 861 | 645,599 | 202 |
| | Lost productivity associated with illness | . | . | 22,109,413 | 12,967 |
| | Total cost (excl. lost productivity) | 62,108,203 | 19,439 | 49,137,361 | 15,380 |
| Other neurodegenerative disorders | Primary sector | 1,946,854 | 821 | 1,011,142 | 426 |
| | Hospital care | 45,022,278 | 18,986 | 37,957,719 | 16,007 |
| | Filled prescriptions | 2,522,819 | 1,064 | 1,571,194 | 663 |
| | Home care | 7,611,334 | 3,210 | 6,734,948 | 2,840 |
| | Nursing home / sheltered accomodation | 6,554,861 | 2,764 | 4,563,429 | 1,924 |
| | Lost productivity associated with illness | . | . | 20,955,974 | 24,283 |
| | Total cost (excl. lost productivity) | 63,658,146 | 26,845 | 51,838,432 | 21,861 |
| Parkinson’s disease | Primary sector | 14,046,320 | 1,038 | 7,782,205 | 575 |
| | Hospital care | 102,236,708 | 7,552 | 49,899,522 | 3,686 |
| | Filled prescriptions | 13,453,068 | 994 | 6,695,571 | 495 |
| | Home care | 30,582,205 | 2,259 | 22,652,601 | 1,673 |
| | Nursing home / sheltered accomodation | 68,027,214 | 5,025 | 43,244,877 | 3,194 |
| | Lost productivity associated with illness | . | . | 63,096,707 | 21,757 |
| | Total cost (excl. lost productivity) | 228,345,516 | 16,867 | 130,274,776 | 9,623 |
| Personality disorders | Primary sector | 7,981,971 | 408 | 2,946,594 | 150 |
| | Hospital care | 316,134,945 | 16,142 | 288,528,402 | 14,732 |
| | Filled prescriptions | 6,360,619 | 325 | 3,756,177 | 192 |
| | Home care | 1,257,176 | 64 | 718,691 | 37 |
| | Nursing home / sheltered accomodation | 3,192,780 | 163 | 2,320,431 | 118 |
| | Lost productivity associated with illness | . | . | 448,508,457 | 25,617 |
| | Total cost (excl. lost productivity) | 334,927,491 | 17,101 | 298,270,295 | 15,230 |
| Polyneuropathy | Primary sector | 15,838,848 | 664 | 6,078,483 | 255 |
| | Hospital care | 339,716,381 | 14,250 | 262,106,401 | 10,994 |
| | Filled prescriptions | 27,308,714 | 1,145 | 17,039,296 | 715 |
| | Home care | 29,054,869 | 1,219 | 18,245,521 | 765 |

***Main analysis / Incident cohort 2016-2021***

| *Brain disorder* | *Cost component* | *Actual costs (EUR)* | *Actual costs per person* | *Attributable costs (EUR)* | *Attributable costs per person* |
|---|---|---|---|---|---|
| | Nursing home / sheltered accomodation | 27,230,000 | 1,142 | -5,079,028 | -213 |
| | Lost productivity associated with illness | . | . | 218,189,377 | 23,721 |
| | Total cost (excl. lost productivity) | 439,148,812 | 18,420 | 298,390,673 | 12,516 |
| Schizophrenia spectrum disorders | Primary sector | 7,896,094 | 342 | 2,327,551 | 101 |
| | Hospital care | 738,618,574 | 32,004 | 706,155,352 | 30,597 |
| | Filled prescriptions | 9,096,656 | 394 | 5,373,958 | 233 |
| | Home care | 8,568,347 | 371 | 6,064,354 | 263 |
| | Nursing home / sheltered accomodation | 31,937,880 | 1,384 | 25,013,863 | 1,084 |
| | Lost productivity associated with illness | . | . | 550,196,711 | 31,512 |
| | Total cost (excl. lost productivity) | 796,117,552 | 34,495 | 744,935,077 | 32,277 |
| Sleep disorders | Primary sector | 167,533,081 | 549 | 75,931,173 | 249 |
| | Hospital care | 2,469,425,093 | 8,090 | 1,872,714,176 | 6,135 |
| | Filled prescriptions | 178,042,094 | 583 | 105,361,618 | 345 |
| | Home care | 147,845,246 | 484 | 78,975,905 | 259 |
| | Nursing home / sheltered accomodation | 449,425,132 | 1,472 | 248,006,961 | 812 |
| | Lost productivity associated with illness | . | . | 1,991,703,568 | 10,343 |
| | Total cost (excl. lost productivity) | 3,412,270,645 | 11,178 | 2,380,989,833 | 7,800 |
| Stress related disorders | Primary sector | 44,795,231 | 472 | 21,608,861 | 228 |
| | Hospital care | 1,130,404,785 | 11,902 | 995,266,722 | 10,479 |
| | Filled prescriptions | 32,112,617 | 338 | 16,672,577 | 176 |
| | Home care | 17,577,057 | 185 | 10,949,961 | 115 |
| | Nursing home / sheltered accomodation | 42,952,550 | 452 | 27,049,157 | 285 |
| | Lost productivity associated with illness | . | . | 1,609,069,934 | 24,067 |
| | Total cost (excl. lost productivity) | 1,267,842,241 | 13,349 | 1,071,547,278 | 11,283 |
| Stroke | Primary sector | 47,943,785 | 664 | 16,075,868 | 223 |
| | Hospital care | 2,278,022,053 | 31,559 | 2,024,131,193 | 28,041 |
| | Filled prescriptions | 61,869,551 | 857 | 27,452,044 | 380 |
| | Home care | 146,375,480 | 2,028 | 99,145,951 | 1,374 |
| | Nursing home / sheltered accomodation | 351,525,262 | 4,870 | 197,597,071 | 2,737 |
| | Lost productivity associated with illness | . | . | 403,734,447 | 19,594 |
| | Total cost (excl. lost productivity) | 2,885,736,132 | 39,978 | 2,364,402,127 | 32,755 |
| Traumatic brain injury | Primary sector | 32,651,874 | 440 | 10,359,923 | 139 |
| | Hospital care | 820,521,769 | 11,045 | 669,339,670 | 9,010 |
| | Filled prescriptions | 30,200,678 | 407 | 11,308,836 | 152 |
| | Home care | 66,502,942 | 895 | 40,685,718 | 548 |
| | Nursing home / sheltered accomodation | 202,594,689 | 2,727 | 118,566,608 | 1,596 |
| | Lost productivity associated with illness | . | . | 232,642,491 | 6,926 |
| | Total cost (excl. lost productivity) | 1,152,471,952 | 15,513 | 850,260,756 | 11,445 |

***Main analysis / Prevalent cohort 2015***

| *Brain disorder* | *Cost component* | *Actual costs (EUR)* | *Actual costs per person* | *Attributable costs (EUR)* | *Attributable costs per person* |
|---|---|---|---|---|---|
| Any brain disorder | Primary sector | 782,233,596 | 418 | 256,104,947 | 137 |
| | Hospital care | 7,099,761,558 | 3,791 | 3,777,738,007 | 2,017 |
| | Filled prescriptions | 838,086,861 | 447 | 493,508,775 | 264 |
| | Home care | 939,375,158 | 502 | 663,566,657 | 354 |
| | Nursing home / sheltered accomodation | 2,773,176,455 | 1,481 | 2,342,360,496 | 1,251 |
| | Lost productivity associated with illness | . | . | 17,652,945,147 | 14,315 |
| | Total cost (excl. lost productivity) | 12,432,633,627 | 6,638 | 7,533,278,882 | 4,022 |
| Alcohol abuse | Primary sector | 69,152,392 | 403 | 14,305,493 | 83 |
| | Hospital care | 1,102,390,934 | 6,429 | 694,920,697 | 4,052 |
| | Filled prescriptions | 106,982,549 | 624 | 58,916,138 | 344 |
| | Home care | 111,998,581 | 653 | 84,240,655 | 491 |
| | Nursing home / sheltered accomodation | 338,123,359 | 1,972 | 285,213,844 | 1,663 |
| | Lost productivity associated with illness | . | . | 3,813,452,150 | 30,649 |
| | Total cost (excl. lost productivity) | 1,728,647,815 | 10,081 | 1,137,596,827 | 6,634 |
| Anxiety disorders | Primary sector | 124,496,228 | 497 | 43,284,339 | 173 |
| | Hospital care | 1,240,139,597 | 4,946 | 684,064,568 | 2,728 |
| | Filled prescriptions | 154,135,312 | 615 | 91,839,863 | 366 |
| | Home care | 141,569,632 | 565 | 83,798,790 | 334 |
| | Nursing home / sheltered accomodation | 486,443,733 | 1,940 | 334,648,410 | 1,335 |
| | Lost productivity associated with illness | . | . | 3,983,488,779 | 20,787 |
| | Total cost (excl. lost productivity) | 2,146,784,502 | 8,562 | 1,237,635,970 | 4,936 |
| Bipolar disorder | Primary sector | 18,585,822 | 565 | 6,993,264 | 213 |
| | Hospital care | 329,268,692 | 10,011 | 245,918,198 | 7,477 |
| | Filled prescriptions | 33,650,572 | 1,023 | 23,792,557 | 723 |
| | Home care | 24,016,729 | 730 | 15,472,671 | 470 |
| | Nursing home / sheltered accomodation | 96,305,790 | 2,928 | 74,885,106 | 2,277 |
| | Lost productivity associated with illness | . | . | 729,450,785 | 31,202 |
| | Total cost (excl. lost productivity) | 501,827,606 | 15,258 | 367,061,796 | 11,161 |
| Brain tumors | Primary sector | 7,757,673 | 526 | 2,015,312 | 137 |
| | Hospital care | 118,501,141 | 8,032 | 73,289,553 | 4,967 |
| | Filled prescriptions | 9,030,374 | 612 | 3,547,087 | 240 |
| | Home care | 17,335,029 | 1,175 | 11,053,545 | 749 |
| | Nursing home / sheltered accomodation | 32,913,778 | 2,231 | 15,406,080 | 1,044 |

***Main analysis / Prevalent cohort 2015***

| *Brain disorder* | *Cost component* | *Actual costs (EUR)* | *Actual costs per person* | *Attributable costs (EUR)* | *Attributable costs per person* |
|---|---|---|---|---|---|
| | Lost productivity associated with illness | . | . | 72,075,182 | 9,732 |
| | Total cost (excl. lost productivity) | 185,537,995 | 12,575 | 105,311,577 | 7,138 |
| Cerebral palsy | Primary sector | 6,384,551 | 691 | 4,462,664 | 483 |
| | Hospital care | 46,844,503 | 5,073 | 33,705,985 | 3,650 |
| | Filled prescriptions | 6,712,756 | 727 | 5,480,807 | 594 |
| | Home care | 14,876,674 | 1,611 | 14,458,053 | 1,566 |
| | Nursing home / sheltered accomodation | 11,975,865 | 1,297 | 11,007,736 | 1,192 |
| | Lost productivity associated with illness | . | . | 144,842,628 | 27,796 |
| | Total cost (excl. lost productivity) | 86,794,348 | 9,400 | 69,115,244 | 7,485 |
| Dementia | Primary sector | 22,936,620 | 580 | 2,313,374 | 58 |
| | Hospital care | 236,596,125 | 5,980 | 52,184,095 | 1,319 |
| | Filled prescriptions | 42,301,833 | 1,069 | 19,566,966 | 495 |
| | Home care | 168,599,833 | 4,261 | 101,171,731 | 2,557 |
| | Nursing home / sheltered accomodation | 1,352,378,117 | 34,181 | 1,197,688,179 | 30,271 |
| | Lost productivity associated with illness | . | . | 156,360,245 | 39,705 |
| | Total cost (excl. lost productivity) | 1,822,812,529 | 46,071 | 1,372,924,346 | 34,700 |
| Depression | Primary sector | 348,762,742 | 491 | 117,107,284 | 165 |
| | Hospital care | 3,286,427,278 | 4,631 | 1,699,879,350 | 2,395 |
| | Filled prescriptions | 417,178,794 | 588 | 242,696,001 | 342 |
| | Home care | 514,945,916 | 726 | 330,381,642 | 466 |
| | Nursing home / sheltered accomodation | 1,778,225,201 | 2,506 | 1,377,807,415 | 1,941 |
| | Lost productivity associated with illness | . | . | 11,028,639,617 | 21,697 |
| | Total cost (excl. lost productivity) | 6,345,539,931 | 8,941 | 3,767,871,692 | 5,309 |
| Developmental and behavioural disorders | Primary sector | 37,900,656 | 293 | 15,237,851 | 118 |
| | Hospital care | 489,461,063 | 3,783 | 350,695,238 | 2,711 |
| | Filled prescriptions | 78,747,385 | 609 | 67,672,893 | 523 |
| | Home care | 7,293,190 | 56 | 5,002,791 | 39 |
| | Nursing home / sheltered accomodation | 13,908,547 | 108 | 10,662,644 | 82 |
| | Lost productivity associated with illness | . | . | 1,421,129,724 | 17,729 |
| | Total cost (excl. lost productivity) | 627,310,842 | 4,849 | 449,271,418 | 3,472 |
| Drug abuse | Primary sector | 18,352,695 | 365 | 5,713,030 | 114 |
| | Hospital care | 476,206,166 | 9,463 | 393,634,431 | 7,822 |
| | Filled prescriptions | 40,222,713 | 799 | 31,281,061 | 622 |

***Main analysis / Prevalent cohort 2015***

| *Brain disorder* | *Cost component* | *Actual costs (EUR)* | *Actual costs per person* | *Attributable costs (EUR)* | *Attributable costs per person* |
|---|---|---|---|---|---|
| | Home care | 21,944,826 | 436 | 17,577,183 | 349 |
| | Nursing home / sheltered accomodation | 54,686,016 | 1,087 | 44,795,751 | 890 |
| | Lost productivity associated with illness | . | . | 1,586,073,664 | 34,874 |
| | Total cost (excl. lost productivity) | 611,412,416 | 12,150 | 493,001,457 | 9,797 |
| Eating disorders | Primary sector | 6,849,665 | 427 | 2,036,461 | 127 |
| | Hospital care | 141,375,424 | 8,813 | 112,451,833 | 7,010 |
| | Filled prescriptions | 5,134,371 | 320 | 2,670,754 | 166 |
| | Home care | 1,075,080 | 67 | 719,407 | 45 |
| | Nursing home / sheltered accomodation | 1,647,012 | 103 | 1,396,218 | 87 |
| | Lost productivity associated with illness | . | . | 98,949,074 | 6,813 |
| | Total cost (excl. lost productivity) | 156,081,552 | 9,730 | 119,274,674 | 7,436 |
| Epilepsy | Primary sector | 31,967,438 | 441 | 10,405,193 | 144 |
| | Hospital care | 414,148,572 | 5,718 | 254,853,362 | 3,518 |
| | Filled prescriptions | 57,623,808 | 796 | 40,260,686 | 556 |
| | Home care | 75,564,134 | 1,043 | 60,279,350 | 832 |
| | Nursing home / sheltered accomodation | 207,973,683 | 2,871 | 166,402,529 | 2,297 |
| | Lost productivity associated with illness | . | . | 885,587,468 | 19,216 |
| | Total cost (excl. lost productivity) | 787,277,635 | 10,869 | 532,201,119 | 7,347 |
| Headache | Primary sector | 189,223,258 | 445 | 45,861,403 | 108 |
| | Hospital care | 1,345,379,939 | 3,162 | 370,898,346 | 872 |
| | Filled prescriptions | 179,112,545 | 421 | 67,345,488 | 158 |
| | Home care | 89,600,334 | 211 | 10,008,546 | 24 |
| | Nursing home / sheltered accomodation | 187,967,137 | 442 | 9,313,024 | 22 |
| | Lost productivity associated with illness | . | . | 1,124,067,021 | 3,502 |
| | Total cost (excl. lost productivity) | 1,991,283,213 | 4,680 | 503,426,806 | 1,183 |
| Infections of the CNS | Primary sector | 6,731,734 | 366 | 1,634,997 | 89 |
| | Hospital care | 76,462,138 | 4,152 | 39,394,843 | 2,139 |
| | Filled prescriptions | 7,218,660 | 392 | 3,190,978 | 173 |
| | Home care | 11,174,841 | 607 | 8,142,837 | 442 |
| | Nursing home / sheltered accomodation | 25,257,409 | 1,372 | 17,330,789 | 941 |
| | Lost productivity associated with illness | . | . | 54,337,333 | 4,672 |
| | Total cost (excl. lost productivity) | 126,844,781 | 6,888 | 69,694,445 | 3,785 |
| Intellectual disability | Primary sector | 11,344,833 | 410 | 5,223,757 | 189 |
| | Hospital care | 161,192,376 | 5,827 | 119,542,064 | 4,321 |
| | Filled prescriptions | 21,750,057 | 786 | 17,520,703 | 633 |
| | Home care | 14,661,044 | 530 | 13,247,410 | 479 |
| | Nursing home / sheltered accomodation | 31,524,079 | 1,140 | 28,467,112 | 1,029 |
| | Lost productivity associated with illness | . | . | 610,616,245 | 34,095 |
| | Total cost (excl. lost productivity) | 240,472,390 | 8,693 | 184,001,046 | 6,651 |

***Main analysis / Prevalent cohort 2015***

| *Brain disorder* | *Cost component* | *Actual costs (EUR)* | *Actual costs per person* | *Attributable costs (EUR)* | *Attributable costs per person* |
|---|---|---|---|---|---|
| Multiple sclerosis | Primary sector | 12,096,775 | 813 | 6,926,721 | 466 |
| | Hospital care | 106,970,567 | 7,192 | 70,569,113 | 4,744 |
| | Filled prescriptions | 9,912,316 | 666 | 5,638,864 | 379 |
| | Home care | 54,459,504 | 3,661 | 52,280,198 | 3,515 |
| | Nursing home / sheltered accomodation | 34,795,208 | 2,339 | 30,102,266 | 2,024 |
| | Lost productivity associated with illness | . | . | 221,303,492 | 18,942 |
| | Total cost (excl. lost productivity) | 218,234,370 | 14,672 | 165,517,162 | 11,128 |
| Neuromuscular disorders | Primary sector | 4,258,791 | 585 | 1,897,586 | 261 |
| | Hospital care | 52,907,446 | 7,264 | 35,109,961 | 4,820 |
| | Filled prescriptions | 4,561,800 | 626 | 2,506,888 | 344 |
| | Home care | 9,557,772 | 1,312 | 7,658,069 | 1,051 |
| | Nursing home / sheltered accomodation | 9,793,012 | 1,344 | 4,965,945 | 682 |
| | Lost productivity associated with illness | . | . | 69,867,348 | 15,592 |
| | Total cost (excl. lost productivity) | 81,078,821 | 11,131 | 52,138,449 | 7,158 |
| Other neurodegenerative disorders | Primary sector | 2,523,386 | 765 | 1,359,225 | 412 |
| | Hospital care | 30,525,378 | 9,258 | 21,422,505 | 6,497 |
| | Filled prescriptions | 2,990,532 | 907 | 1,930,293 | 585 |
| | Home care | 14,609,999 | 4,431 | 13,619,810 | 4,131 |
| | Nursing home / sheltered accomodation | 21,142,083 | 6,412 | 18,538,151 | 5,622 |
| | Lost productivity associated with illness | . | . | 46,959,213 | 27,623 |
| | Total cost (excl. lost productivity) | 71,791,379 | 21,774 | 56,869,984 | 17,248 |
| Parkinson's disease | Primary sector | 20,132,021 | 782 | 8,561,214 | 332 |
| | Hospital care | 165,957,062 | 6,445 | 66,046,612 | 2,565 |
| | Filled prescriptions | 44,418,817 | 1,725 | 32,701,642 | 1,270 |
| | Home care | 74,396,146 | 2,889 | 55,503,099 | 2,155 |
| | Nursing home / sheltered accomodation | 202,730,802 | 7,873 | 145,044,412 | 5,633 |
| | Lost productivity associated with illness | . | . | 75,843,670 | 9,194 |
| | Total cost (excl. lost productivity) | 507,634,848 | 19,714 | 307,856,979 | 11,956 |
| Personality disorders | Primary sector | 33,070,646 | 463 | 11,882,689 | 166 |
| | Hospital care | 545,135,869 | 7,636 | 413,207,997 | 5,788 |
| | Filled prescriptions | 48,098,687 | 674 | 34,031,908 | 477 |
| | Home care | 19,128,288 | 268 | 13,895,600 | 195 |
| | Nursing home / sheltered accomodation | 51,390,311 | 720 | 41,701,088 | 584 |
| | Lost productivity associated with illness | . | . | 1,949,975,843 | 29,862 |
| | Total cost (excl. lost productivity) | 696,823,800 | 9,761 | 514,719,282 | 7,210 |
| Polyneuropathy | Primary sector | 20,445,880 | 613 | 6,749,603 | 202 |
| | Hospital care | 282,123,746 | 8,458 | 167,345,797 | 5,017 |
| | Filled prescriptions | 31,450,051 | 943 | 17,727,836 | 531 |
| | Home care | 56,400,109 | 1,691 | 37,262,313 | 1,117 |

***Main analysis / Prevalent cohort 2015***

| *Brain disorder* | *Cost component* | *Actual costs (EUR)* | *Actual costs per person* | *Attributable costs (EUR)* | *Attributable costs per person* |
|---|---|---|---|---|---|
| | Nursing home / sheltered accomodation | 97,916,044 | 2,936 | 41,703,174 | 1,250 |
| | Lost productivity associated with illness | . | . | 273,079,524 | 19,053 |
| | Total cost (excl. lost productivity) | 488,335,829 | 14,641 | 270,788,723 | 8,118 |
| Schizophrenia spectrum disorders | Primary sector | 24,982,333 | 411 | 7,208,242 | 119 |
| | Hospital care | 788,327,352 | 12,982 | 670,695,575 | 11,045 |
| | Filled prescriptions | 60,694,950 | 1,000 | 47,201,690 | 777 |
| | Home care | 42,530,160 | 700 | 33,193,932 | 547 |
| | Nursing home / sheltered accomodation | 157,124,464 | 2,588 | 134,942,078 | 2,222 |
| | Lost productivity associated with illness | . | . | 1,966,202,375 | 39,224 |
| | Total cost (excl. lost productivity) | 1,073,659,259 | 17,681 | 893,241,517 | 14,710 |
| Sleep disorders | Primary sector | 325,323,646 | 499 | 95,759,319 | 147 |
| | Hospital care | 3,222,667,468 | 4,942 | 1,552,669,974 | 2,381 |
| | Filled prescriptions | 405,025,667 | 621 | 210,128,724 | 322 |
| | Home care | 412,183,807 | 632 | 153,880,516 | 236 |
| | Nursing home / sheltered accomodation | 1,157,999,375 | 1,776 | 452,089,412 | 693 |
| | Lost productivity associated with illness | . | . | 4,397,228,549 | 11,534 |
| | Total cost (excl. lost productivity) | 5,523,199,963 | 8,470 | 2,464,527,945 | 3,780 |
| Stress related disorders | Primary sector | 84,634,253 | 430 | 27,813,728 | 141 |
| | Hospital care | 1,100,397,240 | 5,597 | 738,883,516 | 3,758 |
| | Filled prescriptions | 95,197,887 | 484 | 55,378,666 | 282 |
| | Home care | 54,515,050 | 277 | 35,194,494 | 179 |
| | Nursing home / sheltered accomodation | 145,113,941 | 738 | 102,012,357 | 519 |
| | Lost productivity associated with illness | . | . | 3,688,849,281 | 22,495 |
| | Total cost (excl. lost productivity) | 1,479,858,371 | 7,527 | 959,282,761 | 4,879 |
| Stroke | Primary sector | 70,179,238 | 606 | 18,874,404 | 163 |
| | Hospital care | 780,767,736 | 6,741 | 344,034,592 | 2,970 |
| | Filled prescriptions | 92,869,339 | 802 | 40,003,485 | 345 |
| | Home care | 275,192,054 | 2,376 | 187,523,630 | 1,619 |
| | Nursing home / sheltered accomodation | 791,410,704 | 6,833 | 524,033,647 | 4,525 |
| | Lost productivity associated with illness | . | . | 696,269,766 | 19,388 |
| | Total cost (excl. lost productivity) | 2,010,419,071 | 17,358 | 1,114,469,758 | 9,622 |
| Traumatic brain injury | Primary sector | 79,924,535 | 320 | 14,640,429 | 59 |
| | Hospital care | 792,316,607 | 3,168 | 327,142,860 | 1,308 |
| | Filled prescriptions | 76,441,164 | 306 | 26,708,633 | 107 |
| | Home care | 97,804,491 | 391 | 54,308,036 | 217 |
| | Nursing home / sheltered accomodation | 305,487,478 | 1,221 | 186,807,740 | 747 |
| | Lost productivity associated with illness | . | . | 1,446,426,218 | 8,318 |
| | Total cost (excl. lost productivity) | 1,351,974,275 | 5,406 | 609,607,699 | 2,437 |

***Main analysis / Prevalent cohort 2016***

| *Brain disorder* | *Cost component* | *Actual costs (EUR)* | *Actual costs per person* | *Attributable costs (EUR)* | *Attributable costs per person* |
|---|---|---|---|---|---|
| Any brain disorder | Primary sector | 775,078,978 | 407 | 254,579,331 | 134 |
| | Hospital care | 7,095,402,862 | 3,724 | 3,769,292,725 | 1,978 |
| | Filled prescriptions | 828,998,075 | 435 | 480,454,663 | 252 |
| | Home care | 827,074,234 | 434 | 590,870,343 | 310 |
| | Nursing home / sheltered accomodation | 2,789,238,868 | 1,464 | 2,355,478,235 | 1,236 |
| | Lost productivity associated with illness | . | . | 17,835,189,478 | 14,256 |
| | Total cost (excl. lost productivity) | 12,315,793,016 | 6,463 | 7,450,675,296 | 3,910 |
| Alcohol abuse | Primary sector | 66,920,344 | 394 | 13,429,550 | 79 |
| | Hospital care | 1,079,011,857 | 6,357 | 675,659,288 | 3,981 |
| | Filled prescriptions | 103,336,765 | 609 | 55,396,025 | 326 |
| | Home care | 93,673,738 | 552 | 69,024,724 | 407 |
| | Nursing home / sheltered accomodation | 347,994,930 | 2,050 | 292,848,903 | 1,725 |
| | Lost productivity associated with illness | . | . | 3,724,440,332 | 30,722 |
| | Total cost (excl. lost productivity) | 1,690,937,634 | 9,962 | 1,106,358,489 | 6,518 |
| Anxiety disorders | Primary sector | 127,937,774 | 481 | 44,578,578 | 168 |
| | Hospital care | 1,291,194,843 | 4,852 | 716,339,984 | 2,692 |
| | Filled prescriptions | 151,571,234 | 570 | 86,699,021 | 326 |
| | Home care | 124,362,135 | 467 | 71,937,504 | 270 |
| | Nursing home / sheltered accomodation | 482,468,302 | 1,813 | 326,811,646 | 1,228 |
| | Lost productivity associated with illness | . | . | 4,350,821,420 | 21,413 |
| | Total cost (excl. lost productivity) | 2,177,534,287 | 8,182 | 1,246,366,732 | 4,683 |
| Bipolar disorder | Primary sector | 18,433,399 | 541 | 6,846,899 | 201 |
| | Hospital care | 330,731,416 | 9,715 | 247,024,317 | 7,257 |
| | Filled prescriptions | 30,700,129 | 902 | 20,793,070 | 611 |
| | Home care | 19,828,186 | 582 | 12,606,167 | 370 |
| | Nursing home / sheltered accomodation | 92,465,633 | 2,716 | 71,433,862 | 2,098 |
| | Lost productivity associated with illness | . | . | 775,928,619 | 31,854 |
| | Total cost (excl. lost productivity) | 492,158,762 | 14,458 | 358,704,316 | 10,537 |
| Brain tumors | Primary sector | 7,838,703 | 510 | 1,964,492 | 128 |
| | Hospital care | 123,575,004 | 8,035 | 77,681,337 | 5,051 |
| | Filled prescriptions | 9,121,248 | 593 | 3,440,778 | 224 |
| | Home care | 15,233,128 | 991 | 9,619,581 | 626 |
| | Nursing home / sheltered accomodation | 34,615,194 | 2,251 | 16,503,098 | 1,073 |

***Main analysis / Prevalent cohort 2016***

| *Brain disorder* | *Cost component* | *Actual costs (EUR)* | *Actual costs per person* | *Attributable costs (EUR)* | *Attributable costs per person* |
|---|---|---|---|---|---|
| | Lost productivity associated with illness | . | . | 77,657,485 | 10,223 |
| | Total cost (excl. lost productivity) | 190,383,277 | 12,379 | 109,209,286 | 7,101 |
| Cerebral palsy | Primary sector | 6,447,493 | 693 | 4,521,471 | 486 |
| | Hospital care | 48,917,348 | 5,259 | 35,360,003 | 3,802 |
| | Filled prescriptions | 6,458,876 | 694 | 5,206,697 | 560 |
| | Home care | 14,288,704 | 1,536 | 13,910,973 | 1,496 |
| | Nursing home / sheltered accomodation | 12,929,498 | 1,390 | 11,706,030 | 1,259 |
| | Lost productivity associated with illness | . | . | 163,241,223 | 30,882 |
| | Total cost (excl. lost productivity) | 89,041,918 | 9,573 | 70,705,173 | 7,602 |
| Dementia | Primary sector | 21,925,386 | 546 | 1,453,576 | 36 |
| | Hospital care | 236,759,288 | 5,894 | 54,647,074 | 1,361 |
| | Filled prescriptions | 42,231,682 | 1,051 | 19,019,600 | 474 |
| | Home care | 149,612,943 | 3,725 | 90,255,891 | 2,247 |
| | Nursing home / sheltered accomodation | 1,367,608,307 | 34,049 | 1,213,820,294 | 30,220 |
| | Lost productivity associated with illness | . | . | 155,381,378 | 40,296 |
| | Total cost (excl. lost productivity) | 1,818,137,606 | 45,265 | 1,379,196,435 | 34,337 |
| Depression | Primary sector | 347,127,752 | 477 | 116,420,170 | 160 |
| | Hospital care | 3,294,563,972 | 4,527 | 1,691,747,756 | 2,325 |
| | Filled prescriptions | 407,981,042 | 561 | 230,879,876 | 317 |
| | Home care | 453,753,407 | 624 | 293,907,436 | 404 |
| | Nursing home / sheltered accomodation | 1,768,114,592 | 2,430 | 1,362,865,845 | 1,873 |
| | Lost productivity associated with illness | . | . | 11,102,969,647 | 21,364 |
| | Total cost (excl. lost productivity) | 6,271,540,763 | 8,618 | 3,695,821,083 | 5,079 |
| Developmental and behavioural disorders | Primary sector | 40,648,764 | 290 | 16,526,034 | 118 |
| | Hospital care | 511,695,378 | 3,656 | 362,296,384 | 2,589 |
| | Filled prescriptions | 78,860,414 | 563 | 67,247,495 | 480 |
| | Home care | 6,708,480 | 48 | 4,496,188 | 32 |
| | Nursing home / sheltered accomodation | 14,316,887 | 102 | 10,664,737 | 76 |
| | Lost productivity associated with illness | . | . | 1,572,232,135 | 17,701 |
| | Total cost (excl. lost productivity) | 652,229,923 | 4,660 | 461,230,838 | 3,295 |
| Drug abuse | Primary sector | 18,260,748 | 349 | 5,572,418 | 107 |
| | Hospital care | 474,409,572 | 9,073 | 391,788,484 | 7,493 |
| | Filled prescriptions | 38,393,737 | 734 | 29,437,928 | 563 |

***Main analysis / Prevalent cohort 2016***

| *Brain disorder* | *Cost component* | *Actual costs (EUR)* | *Actual costs per person* | *Attributable costs (EUR)* | *Attributable costs per person* |
|---|---|---|---|---|---|
| | Home care | 17,994,755 | 344 | 13,974,550 | 267 |
| | Nursing home / sheltered accomodation | 55,031,130 | 1,053 | 45,099,191 | 863 |
| | Lost productivity associated with illness | . | . | 1,646,624,247 | 34,839 |
| | Total cost (excl. lost productivity) | 604,089,941 | 11,554 | 485,872,571 | 9,293 |
| Eating disorders | Primary sector | 6,886,547 | 408 | 2,026,193 | 120 |
| | Hospital care | 136,007,704 | 8,057 | 106,530,948 | 6,311 |
| | Filled prescriptions | 4,941,165 | 293 | 2,539,152 | 150 |
| | Home care | 917,025 | 54 | 568,817 | 34 |
| | Nursing home / sheltered accomodation | 1,747,417 | 104 | 1,442,101 | 85 |
| | Lost productivity associated with illness | . | . | 113,849,152 | 7,414 |
| | Total cost (excl. lost productivity) | 150,499,857 | 8,915 | 113,107,211 | 6,700 |
| Epilepsy | Primary sector | 31,378,102 | 432 | 10,242,760 | 141 |
| | Hospital care | 413,962,098 | 5,701 | 256,682,926 | 3,535 |
| | Filled prescriptions | 55,668,245 | 767 | 38,380,197 | 529 |
| | Home care | 68,787,102 | 947 | 55,115,789 | 759 |
| | Nursing home / sheltered accomodation | 214,445,994 | 2,953 | 172,049,062 | 2,369 |
| | Lost productivity associated with illness | . | . | 896,049,841 | 19,557 |
| | Total cost (excl. lost productivity) | 784,241,540 | 10,800 | 532,470,733 | 7,333 |
| Headache | Primary sector | 187,045,704 | 431 | 45,312,413 | 105 |
| | Hospital care | 1,347,837,379 | 3,109 | 370,761,366 | 855 |
| | Filled prescriptions | 176,679,921 | 408 | 66,218,246 | 153 |
| | Home care | 74,992,130 | 173 | 4,915,301 | 11 |
| | Nursing home / sheltered accomodation | 189,993,502 | 438 | 6,522,446 | 15 |
| | Lost productivity associated with illness | . | . | 1,160,150,765 | 3,546 |
| | Total cost (excl. lost productivity) | 1,976,548,636 | 4,560 | 493,729,772 | 1,139 |
| Infections of the CNS | Primary sector | 6,639,943 | 359 | 1,581,275 | 85 |
| | Hospital care | 77,250,976 | 4,176 | 39,878,593 | 2,155 |
| | Filled prescriptions | 7,080,681 | 383 | 3,045,460 | 165 |
| | Home care | 10,725,262 | 580 | 7,935,896 | 429 |
| | Nursing home / sheltered accomodation | 26,643,117 | 1,440 | 18,018,020 | 974 |
| | Lost productivity associated with illness | . | . | 56,885,202 | 4,888 |
| | Total cost (excl. lost productivity) | 128,339,980 | 6,937 | 70,459,244 | 3,808 |
| Intellectual disability | Primary sector | 11,503,428 | 405 | 5,353,272 | 189 |
| | Hospital care | 157,326,502 | 5,544 | 114,888,212 | 4,049 |
| | Filled prescriptions | 20,932,786 | 738 | 16,813,851 | 593 |
| | Home care | 13,732,896 | 484 | 12,252,446 | 432 |
| | Nursing home / sheltered accomodation | 31,811,639 | 1,121 | 28,660,394 | 1,010 |
| | Lost productivity associated with illness | . | . | 641,887,298 | 34,232 |
| | Total cost (excl. lost productivity) | 235,307,250 | 8,292 | 177,968,176 | 6,271 |

***Main analysis / Prevalent cohort 2016***

| *Brain disorder* | *Cost component* | *Actual costs (EUR)* | *Actual costs per person* | *Attributable costs (EUR)* | *Attributable costs per person* |
|---|---|---|---|---|---|
| Multiple sclerosis | Primary sector | 12,140,477 | 793 | 6,945,813 | 454 |
| | Hospital care | 123,288,244 | 8,051 | 86,960,148 | 5,678 |
| | Filled prescriptions | 9,703,236 | 634 | 5,395,340 | 352 |
| | Home care | 51,717,917 | 3,377 | 49,854,559 | 3,255 |
| | Nursing home / sheltered accomodation | 34,616,874 | 2,260 | 29,615,792 | 1,934 |
| | Lost productivity associated with illness | . | . | 228,405,698 | 19,125 |
| | Total cost (excl. lost productivity) | 231,466,748 | 15,115 | 178,771,653 | 11,674 |
| Neuromuscular disorders | Primary sector | 4,322,743 | 572 | 1,900,437 | 252 |
| | Hospital care | 54,329,416 | 7,192 | 35,574,802 | 4,709 |
| | Filled prescriptions | 4,806,337 | 636 | 2,665,396 | 353 |
| | Home care | 8,885,283 | 1,176 | 7,169,700 | 949 |
| | Nursing home / sheltered accomodation | 10,623,765 | 1,406 | 5,396,726 | 714 |
| | Lost productivity associated with illness | . | . | 72,243,890 | 15,620 |
| | Total cost (excl. lost productivity) | 82,967,543 | 10,983 | 52,707,061 | 6,977 |
| Other neurodegenerative disorders | Primary sector | 2,580,063 | 751 | 1,362,848 | 397 |
| | Hospital care | 31,219,702 | 9,090 | 21,424,662 | 6,238 |
| | Filled prescriptions | 3,601,488 | 1,049 | 2,432,036 | 708 |
| | Home care | 14,613,362 | 4,255 | 13,698,977 | 3,989 |
| | Nursing home / sheltered accomodation | 21,198,167 | 6,172 | 18,300,423 | 5,328 |
| | Lost productivity associated with illness | . | . | 47,044,007 | 27,099 |
| | Total cost (excl. lost productivity) | 73,212,783 | 21,317 | 57,218,946 | 16,660 |
| Parkinson’s disease | Primary sector | 18,374,181 | 818 | 8,363,156 | 372 |
| | Hospital care | 151,120,273 | 6,729 | 63,644,818 | 2,834 |
| | Filled prescriptions | 41,696,593 | 1,857 | 31,269,769 | 1,392 |
| | Home care | 63,087,197 | 2,809 | 47,610,320 | 2,120 |
| | Nursing home / sheltered accomodation | 201,166,970 | 8,958 | 151,647,996 | 6,753 |
| | Lost productivity associated with illness | . | . | 104,794,210 | 16,194 |
| | Total cost (excl. lost productivity) | 475,445,215 | 21,171 | 302,536,058 | 13,471 |
| Personality disorders | Primary sector | 32,965,469 | 451 | 12,025,497 | 164 |
| | Hospital care | 531,867,399 | 7,272 | 399,535,862 | 5,463 |
| | Filled prescriptions | 44,111,804 | 603 | 30,345,429 | 415 |
| | Home care | 15,915,204 | 218 | 11,521,150 | 158 |
| | Nursing home / sheltered accomodation | 49,968,684 | 683 | 40,079,928 | 548 |
| | Lost productivity associated with illness | . | . | 2,018,584,084 | 30,120 |
| | Total cost (excl. lost productivity) | 674,828,560 | 9,227 | 493,507,865 | 6,748 |
| Polyneuropathy | Primary sector | 21,305,752 | 604 | 7,107,206 | 202 |
| | Hospital care | 294,419,525 | 8,351 | 174,257,419 | 4,943 |
| | Filled prescriptions | 33,021,912 | 937 | 18,523,191 | 525 |
| | Home care | 47,173,402 | 1,338 | 29,515,622 | 837 |

*Main analysis / Prevalent cohort 2016*

| *Brain disorder* | *Cost component* | *Actual costs (EUR)* | *Actual costs per person* | *Attributable costs (EUR)* | *Attributable costs per person* |
|---|---|---|---|---|---|
| | Nursing home / sheltered accomodation | 104,181,244 | 2,955 | 43,579,446 | 1,236 |
| | Lost productivity associated with illness | . | . | 311,632,453 | 21,012 |
| | Total cost (excl. lost productivity) | 500,101,835 | 14,185 | 272,982,884 | 7,743 |
| Schizophrenia spectrum disorders | Primary sector | 24,486,063 | 393 | 6,911,036 | 111 |
| | Hospital care | 774,136,108 | 12,421 | 656,470,179 | 10,533 |
| | Filled prescriptions | 55,506,240 | 891 | 42,089,281 | 675 |
| | Home care | 33,831,782 | 543 | 25,947,589 | 416 |
| | Nursing home / sheltered accomodation | 157,207,224 | 2,522 | 135,071,806 | 2,167 |
| | Lost productivity associated with illness | . | . | 2,053,506,848 | 39,851 |
| | Total cost (excl. lost productivity) | 1,045,167,418 | 16,770 | 866,489,890 | 13,903 |
| Sleep disorders | Primary sector | 326,951,278 | 485 | 97,076,014 | 144 |
| | Hospital care | 3,286,796,045 | 4,879 | 1,597,890,580 | 2,372 |
| | Filled prescriptions | 406,966,367 | 604 | 209,102,900 | 310 |
| | Home care | 357,865,400 | 531 | 130,969,131 | 194 |
| | Nursing home / sheltered accomodation | 1,175,133,259 | 1,744 | 463,078,239 | 687 |
| | Lost productivity associated with illness | . | . | 4,640,018,455 | 11,798 |
| | Total cost (excl. lost productivity) | 5,553,712,349 | 8,244 | 2,498,116,865 | 3,708 |
| Stress related disorders | Primary sector | 87,251,568 | 419 | 29,094,573 | 140 |
| | Hospital care | 1,147,583,890 | 5,507 | 773,279,684 | 3,710 |
| | Filled prescriptions | 93,812,681 | 450 | 52,738,700 | 253 |
| | Home care | 48,830,675 | 234 | 30,747,254 | 148 |
| | Nursing home / sheltered accomodation | 148,257,989 | 711 | 103,606,223 | 497 |
| | Lost productivity associated with illness | . | . | 4,025,738,334 | 23,167 |
| | Total cost (excl. lost productivity) | 1,525,736,802 | 7,321 | 989,466,435 | 4,748 |
| Stroke | Primary sector | 69,225,933 | 587 | 18,092,076 | 153 |
| | Hospital care | 763,872,390 | 6,475 | 323,755,061 | 2,744 |
| | Filled prescriptions | 94,213,532 | 799 | 40,293,713 | 342 |
| | Home care | 247,961,522 | 2,102 | 171,166,909 | 1,451 |
| | Nursing home / sheltered accomodation | 784,623,526 | 6,651 | 514,132,531 | 4,358 |
| | Lost productivity associated with illness | . | . | 718,074,660 | 19,876 |
| | Total cost (excl. lost productivity) | 1,959,896,904 | 16,614 | 1,067,440,289 | 9,048 |
| Traumatic brain injury | Primary sector | 77,467,966 | 313 | 14,178,421 | 57 |
| | Hospital care | 787,141,712 | 3,177 | 333,342,346 | 1,345 |
| | Filled prescriptions | 74,313,283 | 300 | 25,571,994 | 103 |
| | Home care | 87,224,361 | 352 | 48,076,034 | 194 |
| | Nursing home / sheltered accomodation | 314,363,826 | 1,269 | 192,687,255 | 778 |
| | Lost productivity associated with illness | . | . | 995,812,735 | 5,807 |
| | Total cost (excl. lost productivity) | 1,340,511,147 | 5,411 | 613,856,049 | 2,478 |

***Main analysis / Prevalent cohort 2017***

| *Brain disorder* | *Cost component* | *Actual costs (EUR)* | *Actual costs per person* | *Attributable costs (EUR)* | *Attributable costs per person* |
|---|---|---|---|---|---|
| Any brain disorder | Primary sector | 796,362,868 | 412 | 260,611,772 | 135 |
| | Hospital care | 7,580,813,548 | 3,918 | 3,910,508,011 | 2,021 |
| | Filled prescriptions | 810,281,097 | 419 | 458,154,598 | 237 |
| | Home care | 866,285,607 | 448 | 628,307,720 | 325 |
| | Nursing home / sheltered accomodation | 2,788,158,994 | 1,441 | 2,351,214,344 | 1,215 |
| | Lost productivity associated with illness | . | . | 18,923,869,010 | 14,941 |
| | Total cost (excl. lost productivity) | 12,841,902,114 | 6,637 | 7,608,796,444 | 3,933 |
| Alcohol abuse | Primary sector | 67,535,068 | 403 | 13,564,011 | 81 |
| | Hospital care | 1,108,477,504 | 6,607 | 674,212,106 | 4,018 |
| | Filled prescriptions | 96,886,032 | 577 | 49,976,657 | 298 |
| | Home care | 105,835,773 | 631 | 79,140,243 | 472 |
| | Nursing home / sheltered accomodation | 356,707,018 | 2,126 | 298,948,077 | 1,782 |
| | Lost productivity associated with illness | . | . | 3,728,706,019 | 31,582 |
| | Total cost (excl. lost productivity) | 1,735,441,394 | 10,344 | 1,115,841,093 | 6,651 |
| Anxiety disorders | Primary sector | 134,489,230 | 483 | 46,427,332 | 167 |
| | Hospital care | 1,375,672,724 | 4,940 | 725,644,814 | 2,606 |
| | Filled prescriptions | 146,294,262 | 525 | 80,337,872 | 289 |
| | Home care | 128,372,886 | 461 | 73,252,709 | 263 |
| | Nursing home / sheltered accomodation | 467,892,002 | 1,680 | 312,501,012 | 1,122 |
| | Lost productivity associated with illness | . | . | 4,589,368,703 | 21,618 |
| | Total cost (excl. lost productivity) | 2,252,721,103 | 8,090 | 1,238,163,738 | 4,447 |
| Bipolar disorder | Primary sector | 18,890,350 | 543 | 6,906,423 | 198 |
| | Hospital care | 328,624,301 | 9,441 | 236,345,028 | 6,790 |
| | Filled prescriptions | 27,733,757 | 797 | 17,861,160 | 513 |
| | Home care | 21,190,769 | 609 | 13,741,044 | 395 |
| | Nursing home / sheltered accomodation | 90,491,150 | 2,600 | 69,854,880 | 2,007 |
| | Lost productivity associated with illness | . | . | 806,080,425 | 32,107 |
| | Total cost (excl. lost productivity) | 486,930,327 | 13,988 | 344,708,535 | 9,903 |
| Brain tumors | Primary sector | 8,482,543 | 523 | 2,125,486 | 131 |
| | Hospital care | 134,793,436 | 8,310 | 82,432,870 | 5,082 |
| | Filled prescriptions | 9,133,586 | 563 | 3,171,288 | 196 |
| | Home care | 17,086,753 | 1,053 | 10,811,761 | 667 |
| | Nursing home / sheltered accomodation | 39,047,697 | 2,407 | 19,858,136 | 1,224 |

***Main analysis / Prevalent cohort 2017***

| Brain disorder | Cost component | Actual costs (EUR) | Actual costs per person | Attributable costs (EUR) | Attributable costs per person |
|---|---|---|---|---|---|
| | Lost productivity associated with illness | . | . | 79,219,752 | 10,010 |
| | Total cost (excl. lost productivity) | 208,544,015 | 12,857 | 118,399,541 | 7,299 |
| Cerebral palsy | Primary sector | 6,380,318 | 681 | 4,425,981 | 472 |
| | Hospital care | 50,135,694 | 5,352 | 35,932,578 | 3,836 |
| | Filled prescriptions | 6,589,978 | 704 | 5,368,550 | 573 |
| | Home care | 15,351,078 | 1,639 | 14,864,710 | 1,587 |
| | Nursing home / sheltered accomodation | 13,175,468 | 1,407 | 12,147,222 | 1,297 |
| | Lost productivity associated with illness | . | . | 153,689,104 | 28,867 |
| | Total cost (excl. lost productivity) | 91,632,535 | 9,782 | 72,739,041 | 7,765 |
| Dementia | Primary sector | 22,813,495 | 562 | 1,619,464 | 40 |
| | Hospital care | 237,811,331 | 5,861 | 41,768,126 | 1,029 |
| | Filled prescriptions | 40,005,236 | 986 | 16,763,782 | 413 |
| | Home care | 157,754,743 | 3,888 | 99,178,265 | 2,444 |
| | Nursing home / sheltered accomodation | 1,360,274,786 | 33,524 | 1,210,124,235 | 29,824 |
| | Lost productivity associated with illness | . | . | 154,238,600 | 40,653 |
| | Total cost (excl. lost productivity) | 1,818,659,591 | 44,821 | 1,369,453,872 | 33,751 |
| Depression | Primary sector | 355,866,901 | 481 | 118,008,592 | 160 |
| | Hospital care | 3,464,183,431 | 4,687 | 1,696,705,571 | 2,296 |
| | Filled prescriptions | 391,785,662 | 530 | 213,566,617 | 289 |
| | Home care | 466,007,644 | 630 | 301,563,831 | 408 |
| | Nursing home / sheltered accomodation | 1,729,171,711 | 2,339 | 1,318,341,850 | 1,784 |
| | Lost productivity associated with illness | . | . | 12,021,351,463 | 22,843 |
| | Total cost (excl. lost productivity) | 6,407,015,349 | 8,668 | 3,648,186,462 | 4,936 |
| Developmental and behavioural disorders | Primary sector | 43,966,288 | 292 | 17,745,418 | 118 |
| | Hospital care | 559,344,109 | 3,714 | 387,166,657 | 2,571 |
| | Filled prescriptions | 78,399,984 | 521 | 65,959,879 | 438 |
| | Home care | 7,665,000 | 51 | 5,140,928 | 34 |
| | Nursing home / sheltered accomodation | 14,315,206 | 95 | 10,777,152 | 72 |
| | Lost productivity associated with illness | . | . | 1,775,278,346 | 18,181 |
| | Total cost (excl. lost productivity) | 703,690,588 | 4,673 | 486,790,034 | 3,232 |
| Drug abuse | Primary sector | 18,719,237 | 347 | 5,660,520 | 105 |
| | Hospital care | 507,084,655 | 9,399 | 415,099,302 | 7,694 |
| | Filled prescriptions | 35,872,811 | 665 | 26,993,695 | 500 |

***Main analysis / Prevalent cohort 2017***

| *Brain disorder* | *Cost component* | *Actual costs (EUR)* | *Actual costs per person* | *Attributable costs (EUR)* | *Attributable costs per person* |
|---|---|---|---|---|---|
| | Home care | 21,738,140 | 403 | 17,598,284 | 326 |
| | Nursing home / sheltered accomodation | 55,408,592 | 1,027 | 45,178,012 | 837 |
| | Lost productivity associated with illness | . | . | 1,728,301,336 | 35,407 |
| | Total cost (excl. lost productivity) | 638,823,436 | 11,841 | 510,529,814 | 9,463 |
| Eating disorders | Primary sector | 7,001,602 | 401 | 1,955,999 | 112 |
| | Hospital care | 131,813,062 | 7,542 | 98,816,164 | 5,654 |
| | Filled prescriptions | 4,919,128 | 281 | 2,418,640 | 138 |
| | Home care | 875,276 | 50 | 557,572 | 32 |
| | Nursing home / sheltered accomodation | 1,691,543 | 97 | 1,425,374 | 82 |
| | Lost productivity associated with illness | . | . | 97,170,927 | 6,054 |
| | Total cost (excl. lost productivity) | 146,300,611 | 8,371 | 105,173,750 | 6,018 |
| Epilepsy | Primary sector | 31,545,596 | 435 | 10,034,737 | 138 |
| | Hospital care | 419,602,293 | 5,786 | 248,792,897 | 3,431 |
| | Filled prescriptions | 54,419,801 | 750 | 37,361,947 | 515 |
| | Home care | 73,487,350 | 1,013 | 58,713,321 | 810 |
| | Nursing home / sheltered accomodation | 215,913,202 | 2,977 | 173,737,461 | 2,396 |
| | Lost productivity associated with illness | . | . | 911,043,405 | 20,044 |
| | Total cost (excl. lost productivity) | 794,968,242 | 10,963 | 528,640,363 | 7,290 |
| Headache | Primary sector | 192,595,032 | 436 | 46,648,692 | 106 |
| | Hospital care | 1,471,843,669 | 3,334 | 401,270,495 | 909 |
| | Filled prescriptions | 172,135,038 | 390 | 63,216,219 | 143 |
| | Home care | 81,745,187 | 185 | 7,143,950 | 16 |
| | Nursing home / sheltered accomodation | 190,677,639 | 432 | 9,956,015 | 23 |
| | Lost productivity associated with illness | . | . | 1,233,186,562 | 3,702 |
| | Total cost (excl. lost productivity) | 2,108,996,565 | 4,778 | 528,235,372 | 1,197 |
| Infections of the CNS | Primary sector | 6,810,948 | 366 | 1,633,063 | 88 |
| | Hospital care | 84,828,116 | 4,559 | 44,465,948 | 2,390 |
| | Filled prescriptions | 7,194,215 | 387 | 3,165,381 | 170 |
| | Home care | 11,773,681 | 633 | 8,657,391 | 465 |
| | Nursing home / sheltered accomodation | 26,731,759 | 1,437 | 18,219,628 | 979 |
| | Lost productivity associated with illness | . | . | 54,418,160 | 4,667 |
| | Total cost (excl. lost productivity) | 137,338,719 | 7,381 | 76,141,413 | 4,092 |
| Intellectual disability | Primary sector | 11,797,744 | 405 | 5,441,113 | 187 |
| | Hospital care | 167,175,413 | 5,733 | 120,608,445 | 4,136 |
| | Filled prescriptions | 20,315,116 | 697 | 16,195,144 | 555 |
| | Home care | 13,272,438 | 455 | 11,863,399 | 407 |
| | Nursing home / sheltered accomodation | 32,892,773 | 1,128 | 29,710,288 | 1,019 |
| | Lost productivity associated with illness | . | . | 670,904,623 | 34,092 |
| | Total cost (excl. lost productivity) | 245,453,485 | 8,417 | 183,818,389 | 6,303 |

***Main analysis / Prevalent cohort 2017***

| *Brain disorder* | *Cost component* | *Actual costs (EUR)* | *Actual costs per person* | *Attributable costs (EUR)* | *Attributable costs per person* |
|---|---|---|---|---|---|
| Multiple sclerosis | Primary sector | 12,397,569 | 785 | 6,976,906 | 442 |
| | Hospital care | 145,794,571 | 9,235 | 105,198,199 | 6,663 |
| | Filled prescriptions | 9,408,996 | 596 | 5,060,389 | 321 |
| | Home care | 56,412,110 | 3,573 | 54,434,891 | 3,448 |
| | Nursing home / sheltered accomodation | 35,485,226 | 2,248 | 30,759,956 | 1,948 |
| | Lost productivity associated with illness | . | . | 234,141,526 | 19,072 |
| | Total cost (excl. lost productivity) | 259,498,473 | 16,437 | 202,430,340 | 12,822 |
| Neuromuscular disorders | Primary sector | 4,605,422 | 584 | 2,048,190 | 260 |
| | Hospital care | 59,051,246 | 7,487 | 38,301,340 | 4,856 |
| | Filled prescriptions | 4,929,441 | 625 | 2,742,879 | 348 |
| | Home care | 9,907,232 | 1,256 | 8,093,928 | 1,026 |
| | Nursing home / sheltered accomodation | 11,381,630 | 1,443 | 6,135,090 | 778 |
| | Lost productivity associated with illness | . | . | 77,031,541 | 16,062 |
| | Total cost (excl. lost productivity) | 89,874,971 | 11,394 | 57,321,427 | 7,267 |
| Other neurodegenerative disorders | Primary sector | 2,850,043 | 784 | 1,549,206 | 426 |
| | Hospital care | 33,298,215 | 9,164 | 22,143,670 | 6,094 |
| | Filled prescriptions | 3,256,705 | 896 | 2,058,867 | 567 |
| | Home care | 17,076,485 | 4,699 | 15,974,267 | 4,396 |
| | Nursing home / sheltered accomodation | 22,586,816 | 6,216 | 19,813,868 | 5,453 |
| | Lost productivity associated with illness | . | . | 52,992,639 | 29,408 |
| | Total cost (excl. lost productivity) | 79,068,264 | 21,759 | 61,539,879 | 16,936 |
| Parkinson’s disease | Primary sector | 18,859,387 | 861 | 8,829,628 | 403 |
| | Hospital care | 153,143,749 | 6,990 | 60,944,879 | 2,782 |
| | Filled prescriptions | 41,191,150 | 1,880 | 31,048,319 | 1,417 |
| | Home care | 66,942,087 | 3,056 | 52,320,490 | 2,388 |
| | Nursing home / sheltered accomodation | 197,950,245 | 9,035 | 148,068,782 | 6,759 |
| | Lost productivity associated with illness | . | . | 111,510,209 | 18,789 |
| | Total cost (excl. lost productivity) | 478,086,618 | 21,822 | 301,212,099 | 13,749 |
| Personality disorders | Primary sector | 33,199,105 | 449 | 11,856,064 | 160 |
| | Hospital care | 557,835,160 | 7,538 | 413,350,993 | 5,585 |
| | Filled prescriptions | 40,204,021 | 543 | 26,749,465 | 361 |
| | Home care | 16,988,795 | 230 | 12,470,122 | 169 |
| | Nursing home / sheltered accomodation | 48,175,896 | 651 | 39,280,584 | 531 |
| | Lost productivity associated with illness | . | . | 2,063,778,940 | 30,352 |
| | Total cost (excl. lost productivity) | 696,402,977 | 9,410 | 503,707,228 | 6,806 |
| Polyneuropathy | Primary sector | 22,782,733 | 616 | 7,519,906 | 203 |
| | Hospital care | 319,633,253 | 8,645 | 183,387,945 | 4,960 |
| | Filled prescriptions | 33,717,069 | 912 | 18,625,262 | 504 |
| | Home care | 52,763,606 | 1,427 | 33,674,124 | 911 |

*Main analysis / Prevalent cohort 2017*

| *Brain disorder* | *Cost component* | *Actual costs (EUR)* | *Actual costs per person* | *Attributable costs (EUR)* | *Attributable costs per person* |
|---|---|---|---|---|---|
| | Nursing home / sheltered accomodation | 109,511,506 | 2,962 | 45,899,817 | 1,241 |
| | Lost productivity associated with illness | . | . | 310,548,868 | 20,369 |
| | Total cost (excl. lost productivity) | 538,408,166 | 14,562 | 289,107,054 | 7,819 |
| Schizophrenia spectrum disorders | Primary sector | 24,948,590 | 391 | 6,846,919 | 107 |
| | Hospital care | 802,997,585 | 12,590 | 674,025,107 | 10,568 |
| | Filled prescriptions | 50,592,377 | 793 | 37,341,084 | 585 |
| | Home care | 36,944,922 | 579 | 28,858,728 | 452 |
| | Nursing home / sheltered accomodation | 155,479,972 | 2,438 | 133,773,496 | 2,097 |
| | Lost productivity associated with illness | . | . | 2,119,751,613 | 40,082 |
| | Total cost (excl. lost productivity) | 1,070,963,446 | 16,791 | 880,845,334 | 13,810 |
| Sleep disorders | Primary sector | 340,834,160 | 491 | 101,384,853 | 146 |
| | Hospital care | 3,566,183,610 | 5,141 | 1,720,581,079 | 2,480 |
| | Filled prescriptions | 402,544,395 | 580 | 204,463,103 | 295 |
| | Home care | 386,919,024 | 558 | 155,230,659 | 224 |
| | Nursing home / sheltered accomodation | 1,176,547,535 | 1,696 | 466,757,364 | 673 |
| | Lost productivity associated with illness | . | . | 4,897,580,631 | 12,089 |
| | Total cost (excl. lost productivity) | 5,873,028,724 | 8,466 | 2,648,417,058 | 3,818 |
| Stress related disorders | Primary sector | 92,297,687 | 420 | 30,613,081 | 139 |
| | Hospital care | 1,241,798,002 | 5,653 | 816,311,387 | 3,716 |
| | Filled prescriptions | 92,217,305 | 420 | 49,972,991 | 227 |
| | Home care | 53,513,887 | 244 | 33,063,787 | 151 |
| | Nursing home / sheltered accomodation | 156,807,707 | 714 | 111,894,019 | 509 |
| | Lost productivity associated with illness | . | . | 4,293,710,407 | 23,470 |
| | Total cost (excl. lost productivity) | 1,636,634,587 | 7,450 | 1,041,855,265 | 4,743 |
| Stroke | Primary sector | 71,374,240 | 596 | 18,334,243 | 153 |
| | Hospital care | 819,609,080 | 6,841 | 343,818,168 | 2,870 |
| | Filled prescriptions | 93,220,609 | 778 | 39,239,944 | 328 |
| | Home care | 259,556,363 | 2,166 | 182,732,464 | 1,525 |
| | Nursing home / sheltered accomodation | 781,619,583 | 6,524 | 515,729,845 | 4,304 |
| | Lost productivity associated with illness | . | . | 753,195,480 | 20,696 |
| | Total cost (excl. lost productivity) | 2,025,379,873 | 16,905 | 1,099,854,664 | 9,180 |
| Traumatic brain injury | Primary sector | 78,165,428 | 319 | 14,460,672 | 59 |
| | Hospital care | 823,346,339 | 3,357 | 337,277,124 | 1,375 |
| | Filled prescriptions | 72,140,178 | 294 | 24,615,802 | 100 |
| | Home care | 92,526,719 | 377 | 52,809,051 | 215 |
| | Nursing home / sheltered accomodation | 321,595,892 | 1,311 | 197,631,180 | 806 |
| | Lost productivity associated with illness | . | . | 1,379,431,254 | 8,162 |
| | Total cost (excl. lost productivity) | 1,387,774,556 | 5,658 | 626,793,830 | 2,555 |

***Main analysis / Prevalent cohort 2018***

| *Brain disorder* | *Cost component* | *Actual costs (EUR)* | *Actual costs per person* | *Attributable costs (EUR)* | *Attributable costs per person* |
|---|---|---|---|---|---|
| Any brain disorder | Primary sector | 809,948,459 | 412 | 276,237,446 | 141 |
| | Hospital care | 5,729,858,909 | 2,918 | 3,077,083,259 | 1,567 |
| | Filled prescriptions | 819,026,282 | 417 | 460,694,753 | 235 |
| | Home care | 730,645,658 | 372 | 524,835,157 | 267 |
| | Nursing home / sheltered accomodation | 2,776,942,675 | 1,414 | 2,349,435,356 | 1,196 |
| | Lost productivity associated with illness | . | . | 19,599,778,312 | 15,286 |
| | Total cost (excl. lost productivity) | 10,866,421,983 | 5,533 | 6,688,285,970 | 3,406 |
| Alcohol abuse | Primary sector | 67,359,274 | 409 | 14,688,211 | 89 |
| | Hospital care | 917,469,950 | 5,565 | 609,660,519 | 3,698 |
| | Filled prescriptions | 94,678,071 | 574 | 48,285,321 | 293 |
| | Home care | 88,047,154 | 534 | 65,594,738 | 398 |
| | Nursing home / sheltered accomodation | 361,071,884 | 2,190 | 302,204,476 | 1,833 |
| | Lost productivity associated with illness | . | . | 3,633,379,954 | 31,827 |
| | Total cost (excl. lost productivity) | 1,528,626,332 | 9,272 | 1,040,433,265 | 6,311 |
| Anxiety disorders | Primary sector | 140,159,405 | 483 | 49,420,988 | 170 |
| | Hospital care | 1,114,410,162 | 3,840 | 634,301,715 | 2,185 |
| | Filled prescriptions | 148,719,262 | 512 | 80,168,446 | 276 |
| | Home care | 109,496,396 | 377 | 61,988,663 | 214 |
| | Nursing home / sheltered accomodation | 454,712,963 | 1,567 | 298,096,398 | 1,027 |
| | Lost productivity associated with illness | . | . | 4,874,250,866 | 22,062 |
| | Total cost (excl. lost productivity) | 1,967,498,188 | 6,779 | 1,123,976,211 | 3,873 |
| Bipolar disorder | Primary sector | 19,472,133 | 548 | 7,367,971 | 207 |
| | Hospital care | 275,167,705 | 7,739 | 207,292,025 | 5,830 |
| | Filled prescriptions | 27,895,276 | 785 | 17,874,002 | 503 |
| | Home care | 18,411,917 | 518 | 12,219,385 | 344 |
| | Nursing home / sheltered accomodation | 88,029,769 | 2,476 | 67,757,156 | 1,906 |
| | Lost productivity associated with illness | . | . | 839,090,542 | 32,610 |
| | Total cost (excl. lost productivity) | 428,976,801 | 12,066 | 312,510,539 | 8,790 |
| Brain tumors | Primary sector | 8,942,234 | 529 | 2,285,582 | 135 |
| | Hospital care | 97,371,986 | 5,757 | 57,402,040 | 3,394 |
| | Filled prescriptions | 9,709,236 | 574 | 3,395,762 | 201 |
| | Home care | 14,807,501 | 876 | 8,962,969 | 530 |
| | Nursing home / sheltered accomodation | 39,750,948 | 2,350 | 19,471,173 | 1,151 |

***Main analysis / Prevalent cohort 2018***

| *Brain disorder* | *Cost component* | *Actual costs (EUR)* | *Actual costs per person* | *Attributable costs (EUR)* | *Attributable costs per person* |
|---|---|---|---|---|---|
| | Lost productivity associated with illness | . | . | 82,665,785 | 10,189 |
| | Total cost (excl. lost productivity) | 170,581,907 | 10,086 | 91,517,527 | 5,411 |
| Cerebral palsy | Primary sector | 6,677,649 | 709 | 4,703,041 | 499 |
| | Hospital care | 35,189,192 | 3,734 | 24,874,128 | 2,639 |
| | Filled prescriptions | 6,862,500 | 728 | 5,568,080 | 591 |
| | Home care | 12,161,833 | 1,291 | 11,758,131 | 1,248 |
| | Nursing home / sheltered accomodation | 13,814,864 | 1,466 | 12,758,871 | 1,354 |
| | Lost productivity associated with illness | . | . | 159,912,641 | 29,353 |
| | Total cost (excl. lost productivity) | 74,706,038 | 7,927 | 59,662,250 | 6,331 |
| Dementia | Primary sector | 25,664,049 | 633 | 4,144,564 | 102 |
| | Hospital care | 203,575,020 | 5,019 | 52,575,678 | 1,296 |
| | Filled prescriptions | 40,221,804 | 992 | 16,589,885 | 409 |
| | Home care | 133,995,613 | 3,303 | 85,417,825 | 2,106 |
| | Nursing home / sheltered accomodation | 1,352,271,411 | 33,337 | 1,206,597,762 | 29,746 |
| | Lost productivity associated with illness | . | . | 155,967,562 | 42,222 |
| | Total cost (excl. lost productivity) | 1,755,727,898 | 43,283 | 1,365,325,714 | 33,659 |
| Depression | Primary sector | 362,095,051 | 482 | 122,689,671 | 163 |
| | Hospital care | 2,709,302,887 | 3,609 | 1,438,941,872 | 1,917 |
| | Filled prescriptions | 393,262,133 | 524 | 211,555,176 | 282 |
| | Home care | 391,278,361 | 521 | 253,455,253 | 338 |
| | Nursing home / sheltered accomodation | 1,691,396,715 | 2,253 | 1,284,687,845 | 1,712 |
| | Lost productivity associated with illness | . | . | 12,480,032,525 | 23,454 |
| | Total cost (excl. lost productivity) | 5,547,335,147 | 7,390 | 3,311,329,817 | 4,412 |
| Developmental and behavioural disorders | Primary sector | 48,487,192 | 298 | 20,166,771 | 124 |
| | Hospital care | 477,885,442 | 2,940 | 341,003,295 | 2,098 |
| | Filled prescriptions | 82,896,868 | 510 | 69,063,248 | 425 |
| | Home care | 6,677,962 | 41 | 4,422,974 | 27 |
| | Nursing home / sheltered accomodation | 14,760,725 | 91 | 11,128,301 | 68 |
| | Lost productivity associated with illness | . | . | 1,999,961,448 | 18,542 |
| | Total cost (excl. lost productivity) | 630,708,189 | 3,880 | 445,784,588 | 2,742 |
| Drug abuse | Primary sector | 19,487,009 | 350 | 6,204,593 | 111 |
| | Hospital care | 422,429,693 | 7,588 | 354,448,025 | 6,367 |
| | Filled prescriptions | 35,003,499 | 629 | 25,956,958 | 466 |

***Main analysis / Prevalent cohort 2018***

| *Brain disorder* | *Cost component* | *Actual costs (EUR)* | *Actual costs per person* | *Attributable costs (EUR)* | *Attributable costs per person* |
|---|---|---|---|---|---|
| | Home care | 17,448,182 | 313 | 13,777,046 | 247 |
| | Nursing home / sheltered accomodation | 54,392,994 | 977 | 44,799,864 | 805 |
| | Lost productivity associated with illness | . | . | 1,806,769,720 | 35,796 |
| | Total cost (excl. lost productivity) | 548,761,376 | 9,858 | 445,186,485 | 7,997 |
| Eating disorders | Primary sector | 7,318,119 | 402 | 2,115,937 | 116 |
| | Hospital care | 113,902,664 | 6,252 | 89,307,018 | 4,902 |
| | Filled prescriptions | 5,183,680 | 285 | 2,645,304 | 145 |
| | Home care | 820,604 | 45 | 565,318 | 31 |
| | Nursing home / sheltered accomodation | 1,392,430 | 76 | 1,154,061 | 63 |
| | Lost productivity associated with illness | . | . | 106,002,713 | 6,335 |
| | Total cost (excl. lost productivity) | 128,617,497 | 7,060 | 95,787,639 | 5,258 |
| Epilepsy | Primary sector | 32,386,003 | 448 | 10,959,689 | 152 |
| | Hospital care | 317,022,357 | 4,383 | 191,638,807 | 2,649 |
| | Filled prescriptions | 55,553,373 | 768 | 38,322,922 | 530 |
| | Home care | 63,348,912 | 876 | 51,213,592 | 708 |
| | Nursing home / sheltered accomodation | 216,343,387 | 2,991 | 173,603,608 | 2,400 |
| | Lost productivity associated with illness | . | . | 906,689,730 | 20,100 |
| | Total cost (excl. lost productivity) | 684,654,032 | 9,465 | 465,738,618 | 6,439 |
| Headache | Primary sector | 194,214,527 | 433 | 47,925,249 | 107 |
| | Hospital care | 1,061,561,970 | 2,367 | 286,990,758 | 640 |
| | Filled prescriptions | 171,581,687 | 383 | 61,289,015 | 137 |
| | Home care | 67,746,725 | 151 | 5,414,837 | 12 |
| | Nursing home / sheltered accomodation | 187,694,070 | 418 | 5,075,816 | 11 |
| | Lost productivity associated with illness | . | . | 1,350,577,115 | 3,990 |
| | Total cost (excl. lost productivity) | 1,682,798,979 | 3,752 | 406,695,675 | 907 |
| Infections of the CNS | Primary sector | 7,080,754 | 374 | 1,791,593 | 95 |
| | Hospital care | 61,721,550 | 3,259 | 31,075,396 | 1,641 |
| | Filled prescriptions | 7,466,738 | 394 | 3,297,254 | 174 |
| | Home care | 9,481,521 | 501 | 7,009,546 | 370 |
| | Nursing home / sheltered accomodation | 26,608,248 | 1,405 | 18,106,159 | 956 |
| | Lost productivity associated with illness | . | . | 59,808,683 | 5,014 |
| | Total cost (excl. lost productivity) | 112,358,812 | 5,932 | 61,279,948 | 3,236 |
| Intellectual disability | Primary sector | 12,428,024 | 414 | 5,962,039 | 199 |
| | Hospital care | 132,991,928 | 4,433 | 98,186,528 | 3,273 |
| | Filled prescriptions | 21,295,618 | 710 | 17,089,822 | 570 |
| | Home care | 10,418,040 | 347 | 9,128,057 | 304 |
| | Nursing home / sheltered accomodation | 31,905,742 | 1,064 | 28,747,465 | 958 |
| | Lost productivity associated with illness | . | . | 708,833,721 | 34,263 |
| | Total cost (excl. lost productivity) | 209,039,353 | 6,968 | 159,113,911 | 5,304 |

***Main analysis / Prevalent cohort 2018***

| *Brain disorder* | *Cost component* | *Actual costs (EUR)* | *Actual costs per person* | *Attributable costs (EUR)* | *Attributable costs per person* |
|---|---|---|---|---|---|
| Multiple sclerosis | Primary sector | 12,896,279 | 799 | 7,402,099 | 458 |
| | Hospital care | 52,727,508 | 3,265 | 23,438,271 | 1,451 |
| | Filled prescriptions | 9,744,677 | 603 | 5,355,018 | 332 |
| | Home care | 49,512,601 | 3,066 | 47,660,112 | 2,951 |
| | Nursing home / sheltered accomodation | 35,798,623 | 2,217 | 30,784,193 | 1,906 |
| | Lost productivity associated with illness | . | . | 240,059,611 | 19,229 |
| | Total cost (excl. lost productivity) | 160,679,687 | 9,949 | 114,639,693 | 7,099 |
| Neuromuscular disorders | Primary sector | 4,764,141 | 588 | 2,145,248 | 265 |
| | Hospital care | 43,334,226 | 5,352 | 27,996,686 | 3,458 |
| | Filled prescriptions | 5,106,274 | 631 | 2,838,708 | 351 |
| | Home care | 8,803,527 | 1,087 | 7,206,334 | 890 |
| | Nursing home / sheltered accomodation | 11,622,769 | 1,435 | 6,458,257 | 798 |
| | Lost productivity associated with illness | . | . | 81,524,090 | 16,577 |
| | Total cost (excl. lost productivity) | 73,630,937 | 9,094 | 46,645,233 | 5,761 |
| Other neurodegenerative disorders | Primary sector | 3,076,052 | 823 | 1,720,403 | 460 |
| | Hospital care | 27,760,825 | 7,427 | 19,376,301 | 5,184 |
| | Filled prescriptions | 3,798,895 | 1,016 | 2,516,353 | 673 |
| | Home care | 14,479,912 | 3,874 | 13,500,824 | 3,612 |
| | Nursing home / sheltered accomodation | 22,712,637 | 6,077 | 19,642,389 | 5,255 |
| | Lost productivity associated with illness | . | . | 53,278,013 | 29,290 |
| | Total cost (excl. lost productivity) | 71,828,321 | 19,217 | 56,756,270 | 15,185 |
| Parkinson’s disease | Primary sector | 19,799,119 | 921 | 9,868,113 | 459 |
| | Hospital care | 124,404,605 | 5,786 | 55,921,328 | 2,601 |
| | Filled prescriptions | 41,825,232 | 1,945 | 31,688,979 | 1,474 |
| | Home care | 57,324,176 | 2,666 | 44,926,161 | 2,089 |
| | Nursing home / sheltered accomodation | 200,394,402 | 9,320 | 151,485,413 | 7,045 |
| | Lost productivity associated with illness | . | . | 134,446,803 | 24,172 |
| | Total cost (excl. lost productivity) | 443,747,534 | 20,637 | 293,889,995 | 13,668 |
| Personality disorders | Primary sector | 33,642,105 | 449 | 12,300,491 | 164 |
| | Hospital care | 446,383,220 | 5,963 | 342,831,574 | 4,580 |
| | Filled prescriptions | 38,802,761 | 518 | 25,298,299 | 338 |
| | Home care | 14,255,503 | 190 | 10,385,506 | 139 |
| | Nursing home / sheltered accomodation | 45,041,300 | 602 | 36,097,453 | 482 |
| | Lost productivity associated with illness | . | . | 2,134,747,683 | 30,943 |
| | Total cost (excl. lost productivity) | 578,124,890 | 7,723 | 426,913,322 | 5,703 |
| Polyneuropathy | Primary sector | 24,272,920 | 631 | 8,381,601 | 218 |
| | Hospital care | 238,981,435 | 6,216 | 133,887,795 | 3,482 |
| | Filled prescriptions | 35,012,746 | 911 | 19,250,312 | 501 |
| | Home care | 45,768,699 | 1,190 | 28,498,507 | 741 |

***Main analysis / Prevalent cohort 2018***

| *Brain disorder* | *Cost component* | *Actual costs (EUR)* | *Actual costs per person* | *Attributable costs (EUR)* | *Attributable costs per person* |
|---|---|---|---|---|---|
| | Nursing home / sheltered accomodation | 112,979,243 | 2,939 | 46,948,006 | 1,221 |
| | Lost productivity associated with illness | . | . | 315,600,013 | 20,343 |
| | Total cost (excl. lost productivity) | 457,015,042 | 11,887 | 236,966,221 | 6,163 |
| Schizophrenia spectrum disorders | Primary sector | 25,769,877 | 394 | 7,499,287 | 115 |
| | Hospital care | 663,285,873 | 10,142 | 569,325,794 | 8,706 |
| | Filled prescriptions | 49,977,095 | 764 | 36,564,195 | 559 |
| | Home care | 30,499,810 | 466 | 23,506,461 | 359 |
| | Nursing home / sheltered accomodation | 151,166,569 | 2,312 | 129,725,847 | 1,984 |
| | Lost productivity associated with illness | . | . | 2,204,081,780 | 40,630 |
| | Total cost (excl. lost productivity) | 920,699,224 | 14,079 | 766,621,584 | 11,723 |
| Sleep disorders | Primary sector | 350,878,477 | 491 | 107,479,090 | 151 |
| | Hospital care | 2,708,952,007 | 3,794 | 1,332,660,538 | 1,866 |
| | Filled prescriptions | 410,696,854 | 575 | 208,254,599 | 292 |
| | Home care | 320,792,834 | 449 | 122,771,834 | 172 |
| | Nursing home / sheltered accomodation | 1,170,259,229 | 1,639 | 462,497,178 | 648 |
| | Lost productivity associated with illness | . | . | 5,186,787,731 | 12,427 |
| | Total cost (excl. lost productivity) | 4,961,579,401 | 6,949 | 2,233,663,239 | 3,128 |
| Stress related disorders | Primary sector | 95,942,081 | 416 | 32,044,399 | 139 |
| | Hospital care | 1,023,783,444 | 4,441 | 710,719,140 | 3,083 |
| | Filled prescriptions | 95,082,557 | 412 | 50,908,415 | 221 |
| | Home care | 49,220,692 | 214 | 31,393,034 | 136 |
| | Nursing home / sheltered accomodation | 163,095,592 | 707 | 117,159,755 | 508 |
| | Lost productivity associated with illness | . | . | 4,597,322,791 | 23,986 |
| | Total cost (excl. lost productivity) | 1,427,124,366 | 6,190 | 942,224,743 | 4,087 |
| Stroke | Primary sector | 73,857,640 | 607 | 19,969,406 | 164 |
| | Hospital care | 662,718,763 | 5,447 | 303,682,026 | 2,496 |
| | Filled prescriptions | 95,413,657 | 784 | 40,741,332 | 335 |
| | Home care | 218,681,150 | 1,797 | 151,926,262 | 1,249 |
| | Nursing home / sheltered accomodation | 780,648,306 | 6,416 | 518,611,131 | 4,263 |
| | Lost productivity associated with illness | . | . | 765,518,082 | 20,907 |
| | Total cost (excl. lost productivity) | 1,831,319,515 | 15,052 | 1,034,930,156 | 8,507 |
| Traumatic brain injury | Primary sector | 78,731,349 | 324 | 15,666,775 | 65 |
| | Hospital care | 635,726,538 | 2,618 | 278,753,315 | 1,148 |
| | Filled prescriptions | 72,236,324 | 297 | 24,436,606 | 101 |
| | Home care | 78,426,912 | 323 | 43,706,733 | 180 |
| | Nursing home / sheltered accomodation | 327,189,559 | 1,347 | 203,028,939 | 836 |
| | Lost productivity associated with illness | . | . | 1,364,217,949 | 8,212 |
| | Total cost (excl. lost productivity) | 1,192,310,682 | 4,910 | 565,592,368 | 2,329 |

***Main analysis / Prevalent cohort 2019***

| *Brain disorder* | *Cost component* | *Actual costs (EUR)* | *Actual costs per person* | *Attributable costs (EUR)* | *Attributable costs per person* |
|---|---|---|---|---|---|
| Any brain disorder | Primary sector | 818,972,026 | 411 | 278,469,164 | 140 |
| | Hospital care | 6,547,500,439 | 3,288 | 3,396,253,803 | 1,706 |
| | Filled prescriptions | 868,317,097 | 436 | 474,382,734 | 238 |
| | Home care | 843,088,115 | 423 | 613,392,030 | 308 |
| | Nursing home / sheltered accomodation | 2,788,557,881 | 1,401 | 2,356,640,542 | 1,184 |
| | Lost productivity associated with illness | . | . | 20,474,390,030 | 15,785 |
| | Total cost (excl. lost productivity) | 11,866,435,558 | 5,960 | 7,119,138,273 | 3,576 |
| Alcohol abuse | Primary sector | 65,605,838 | 405 | 13,974,860 | 86 |
| | Hospital care | 969,641,508 | 5,989 | 612,252,963 | 3,781 |
| | Filled prescriptions | 96,478,924 | 596 | 46,989,972 | 290 |
| | Home care | 102,005,553 | 630 | 75,710,397 | 468 |
| | Nursing home / sheltered accomodation | 359,292,119 | 2,219 | 299,566,316 | 1,850 |
| | Lost productivity associated with illness | . | . | 3,624,608,077 | 32,770 |
| | Total cost (excl. lost productivity) | 1,593,023,942 | 9,839 | 1,048,494,509 | 6,476 |
| Anxiety disorders | Primary sector | 144,833,790 | 480 | 50,504,770 | 167 |
| | Hospital care | 1,256,081,515 | 4,161 | 674,272,638 | 2,233 |
| | Filled prescriptions | 156,741,428 | 519 | 80,846,599 | 268 |
| | Home care | 130,126,989 | 431 | 75,606,096 | 250 |
| | Nursing home / sheltered accomodation | 450,602,049 | 1,493 | 290,365,100 | 962 |
| | Lost productivity associated with illness | . | . | 5,174,829,594 | 22,524 |
| | Total cost (excl. lost productivity) | 2,138,385,770 | 7,083 | 1,171,595,203 | 3,881 |
| Bipolar disorder | Primary sector | 19,648,784 | 543 | 7,461,110 | 206 |
| | Hospital care | 297,838,794 | 8,233 | 218,218,936 | 6,032 |
| | Filled prescriptions | 29,408,438 | 813 | 18,632,419 | 515 |
| | Home care | 21,099,932 | 583 | 13,953,358 | 386 |
| | Nursing home / sheltered accomodation | 87,389,323 | 2,416 | 67,693,019 | 1,871 |
| | Lost productivity associated with illness | . | . | 873,030,675 | 33,222 |
| | Total cost (excl. lost productivity) | 455,385,271 | 12,589 | 325,958,843 | 9,011 |
| Brain tumors | Primary sector | 9,368,759 | 532 | 2,410,496 | 137 |
| | Hospital care | 119,907,917 | 6,811 | 70,868,435 | 4,026 |
| | Filled prescriptions | 10,730,418 | 610 | 3,732,956 | 212 |
| | Home care | 15,827,764 | 899 | 9,069,325 | 515 |
| | Nursing home / sheltered accomodation | 41,350,699 | 2,349 | 19,940,161 | 1,133 |

***Main analysis / Prevalent cohort 2019***

| *Brain disorder* | *Cost component* | *Actual costs (EUR)* | *Actual costs per person* | *Attributable costs (EUR)* | *Attributable costs per person* |
|---|---|---|---|---|---|
| | Lost productivity associated with illness | . | . | 89,171,309 | 10,698 |
| | Total cost (excl. lost productivity) | 197,185,557 | 11,201 | 106,021,373 | 6,022 |
| Cerebral palsy | Primary sector | 6,792,338 | 723 | 4,788,060 | 509 |
| | Hospital care | 40,595,806 | 4,318 | 28,477,461 | 3,029 |
| | Filled prescriptions | 7,624,181 | 811 | 6,248,096 | 665 |
| | Home care | 14,400,185 | 1,532 | 13,876,546 | 1,476 |
| | Nursing home / sheltered accomodation | 14,135,402 | 1,504 | 12,750,345 | 1,356 |
| | Lost productivity associated with illness | . | . | 164,320,670 | 29,731 |
| | Total cost (excl. lost productivity) | 83,547,912 | 8,887 | 66,140,508 | 7,036 |
| Dementia | Primary sector | 25,855,788 | 643 | 4,729,877 | 118 |
| | Hospital care | 190,857,695 | 4,746 | 20,447,725 | 508 |
| | Filled prescriptions | 43,805,914 | 1,089 | 18,730,996 | 466 |
| | Home care | 142,928,697 | 3,554 | 88,782,082 | 2,208 |
| | Nursing home / sheltered accomodation | 1,348,344,292 | 33,530 | 1,206,468,423 | 30,002 |
| | Lost productivity associated with illness | . | . | 151,799,017 | 43,396 |
| | Total cost (excl. lost productivity) | 1,751,792,386 | 43,562 | 1,339,159,102 | 33,301 |
| Depression | Primary sector | 365,284,246 | 479 | 122,579,630 | 161 |
| | Hospital care | 3,008,313,001 | 3,949 | 1,476,642,753 | 1,938 |
| | Filled prescriptions | 413,035,071 | 542 | 214,701,163 | 282 |
| | Home care | 447,408,704 | 587 | 289,683,736 | 380 |
| | Nursing home / sheltered accomodation | 1,673,349,107 | 2,196 | 1,266,414,388 | 1,662 |
| | Lost productivity associated with illness | . | . | 12,923,618,497 | 24,041 |
| | Total cost (excl. lost productivity) | 5,907,390,130 | 7,754 | 3,370,021,670 | 4,424 |
| Developmental and behavioural disorders | Primary sector | 53,869,993 | 309 | 22,619,298 | 130 |
| | Hospital care | 527,813,049 | 3,026 | 359,360,943 | 2,060 |
| | Filled prescriptions | 82,830,893 | 475 | 66,714,697 | 383 |
| | Home care | 8,239,132 | 47 | 5,380,715 | 31 |
| | Nursing home / sheltered accomodation | 16,295,570 | 93 | 11,961,388 | 69 |
| | Lost productivity associated with illness | . | . | 2,259,636,668 | 19,098 |
| | Total cost (excl. lost productivity) | 689,048,638 | 3,951 | 466,037,041 | 2,672 |
| Drug abuse | Primary sector | 19,951,813 | 349 | 6,303,717 | 110 |
| | Hospital care | 470,320,218 | 8,231 | 389,248,639 | 6,812 |
| | Filled prescriptions | 34,235,751 | 599 | 24,490,673 | 429 |

***Main analysis / Prevalent cohort 2019***

| *Brain disorder* | *Cost component* | *Actual costs (EUR)* | *Actual costs per person* | *Attributable costs (EUR)* | *Attributable costs per person* |
|---|---|---|---|---|---|
| | Home care | 19,173,838 | 336 | 15,098,915 | 264 |
| | Nursing home / sheltered accomodation | 52,926,416 | 926 | 43,805,178 | 767 |
| | Lost productivity associated with illness | . | . | 1,905,558,237 | 36,669 |
| | Total cost (excl. lost productivity) | 596,608,035 | 10,441 | 478,947,122 | 8,382 |
| Eating disorders | Primary sector | 7,690,183 | 411 | 2,269,033 | 121 |
| | Hospital care | 126,872,432 | 6,780 | 96,893,385 | 5,178 |
| | Filled prescriptions | 5,459,941 | 292 | 2,742,406 | 147 |
| | Home care | 964,445 | 52 | 608,331 | 33 |
| | Nursing home / sheltered accomodation | 1,347,059 | 72 | 1,050,035 | 56 |
| | Lost productivity associated with illness | . | . | 112,998,470 | 6,567 |
| | Total cost (excl. lost productivity) | 142,334,060 | 7,606 | 103,563,191 | 5,534 |
| Epilepsy | Primary sector | 32,257,004 | 451 | 11,089,754 | 155 |
| | Hospital care | 341,737,537 | 4,783 | 199,541,779 | 2,793 |
| | Filled prescriptions | 61,114,254 | 855 | 42,844,579 | 600 |
| | Home care | 73,263,797 | 1,025 | 59,190,288 | 828 |
| | Nursing home / sheltered accomodation | 211,368,532 | 2,958 | 168,626,926 | 2,360 |
| | Lost productivity associated with illness | . | . | 925,348,708 | 20,796 |
| | Total cost (excl. lost productivity) | 719,741,124 | 10,074 | 481,293,327 | 6,737 |
| Headache | Primary sector | 196,527,262 | 433 | 48,574,421 | 107 |
| | Hospital care | 1,257,322,777 | 2,771 | 341,773,014 | 753 |
| | Filled prescriptions | 179,571,889 | 396 | 61,196,644 | 135 |
| | Home care | 79,471,446 | 175 | 6,654,909 | 15 |
| | Nursing home / sheltered accomodation | 190,657,054 | 420 | 6,414,668 | 14 |
| | Lost productivity associated with illness | . | . | 1,415,568,641 | 4,134 |
| | Total cost (excl. lost productivity) | 1,903,550,428 | 4,195 | 464,613,656 | 1,024 |
| Infections of the CNS | Primary sector | 7,271,211 | 378 | 1,874,606 | 97 |
| | Hospital care | 75,449,782 | 3,920 | 40,035,562 | 2,080 |
| | Filled prescriptions | 8,184,205 | 425 | 3,674,352 | 191 |
| | Home care | 10,432,207 | 542 | 7,372,751 | 383 |
| | Nursing home / sheltered accomodation | 26,274,477 | 1,365 | 17,083,742 | 888 |
| | Lost productivity associated with illness | . | . | 58,995,572 | 4,852 |
| | Total cost (excl. lost productivity) | 127,611,882 | 6,630 | 70,041,014 | 3,639 |
| Intellectual disability | Primary sector | 12,917,146 | 424 | 6,213,705 | 204 |
| | Hospital care | 141,934,339 | 4,659 | 102,072,395 | 3,350 |
| | Filled prescriptions | 23,061,708 | 757 | 18,548,753 | 609 |
| | Home care | 11,994,914 | 394 | 10,612,566 | 348 |
| | Nursing home / sheltered accomodation | 32,176,288 | 1,056 | 29,273,780 | 961 |
| | Lost productivity associated with illness | . | . | 752,564,799 | 35,011 |
| | Total cost (excl. lost productivity) | 222,084,396 | 7,290 | 166,721,199 | 5,473 |

***Main analysis / Prevalent cohort 2019***

| *Brain disorder* | *Cost component* | *Actual costs (EUR)* | *Actual costs per person* | *Attributable costs (EUR)* | *Attributable costs per person* |
|---|---|---|---|---|---|
| Multiple sclerosis | Primary sector | 13,095,724 | 800 | 7,508,402 | 459 |
| | Hospital care | 113,935,899 | 6,964 | 79,149,786 | 4,838 |
| | Filled prescriptions | 10,232,810 | 625 | 5,510,887 | 337 |
| | Home care | 57,231,639 | 3,498 | 55,152,961 | 3,371 |
| | Nursing home / sheltered accomodation | 35,586,891 | 2,175 | 30,413,135 | 1,859 |
| | Lost productivity associated with illness | . | . | 249,950,537 | 19,907 |
| | Total cost (excl. lost productivity) | 230,082,963 | 14,064 | 177,735,172 | 10,864 |
| Neuromuscular disorders | Primary sector | 4,922,963 | 599 | 2,246,813 | 274 |
| | Hospital care | 48,459,643 | 5,901 | 30,198,958 | 3,677 |
| | Filled prescriptions | 5,436,278 | 662 | 2,958,835 | 360 |
| | Home care | 10,730,744 | 1,307 | 9,011,039 | 1,097 |
| | Nursing home / sheltered accomodation | 11,941,626 | 1,454 | 6,471,262 | 788 |
| | Lost productivity associated with illness | . | . | 81,639,311 | 16,433 |
| | Total cost (excl. lost productivity) | 81,491,255 | 9,923 | 50,886,906 | 6,196 |
| Other neurodegenerative disorders | Primary sector | 3,060,624 | 811 | 1,697,403 | 450 |
| | Hospital care | 28,811,909 | 7,631 | 18,639,722 | 4,937 |
| | Filled prescriptions | 3,352,315 | 888 | 1,961,894 | 520 |
| | Home care | 16,372,646 | 4,336 | 15,290,597 | 4,050 |
| | Nursing home / sheltered accomodation | 22,971,840 | 6,084 | 19,828,037 | 5,251 |
| | Lost productivity associated with illness | . | . | 54,502,400 | 30,079 |
| | Total cost (excl. lost productivity) | 74,569,334 | 19,749 | 57,417,654 | 15,207 |
| Parkinson’s disease | Primary sector | 19,833,367 | 943 | 10,178,665 | 484 |
| | Hospital care | 129,851,474 | 6,174 | 52,882,614 | 2,515 |
| | Filled prescriptions | 44,013,533 | 2,093 | 33,360,366 | 1,586 |
| | Home care | 63,600,706 | 3,024 | 49,756,523 | 2,366 |
| | Nursing home / sheltered accomodation | 202,223,108 | 9,615 | 155,442,672 | 7,391 |
| | Lost productivity associated with illness | . | . | 147,961,613 | 28,124 |
| | Total cost (excl. lost productivity) | 459,522,188 | 21,850 | 301,620,841 | 14,342 |
| Personality disorders | Primary sector | 33,815,104 | 451 | 12,263,434 | 163 |
| | Hospital care | 482,110,489 | 6,424 | 360,346,802 | 4,801 |
| | Filled prescriptions | 38,555,775 | 514 | 24,295,948 | 324 |
| | Home care | 17,147,855 | 228 | 12,898,962 | 172 |
| | Nursing home / sheltered accomodation | 43,866,693 | 584 | 35,316,343 | 471 |
| | Lost productivity associated with illness | . | . | 2,199,066,934 | 31,703 |
| | Total cost (excl. lost productivity) | 615,495,917 | 8,201 | 445,121,488 | 5,931 |
| Polyneuropathy | Primary sector | 24,773,321 | 623 | 8,437,621 | 212 |
| | Hospital care | 293,964,675 | 7,397 | 167,936,770 | 4,226 |
| | Filled prescriptions | 38,239,584 | 962 | 20,410,306 | 514 |
| | Home care | 55,610,277 | 1,399 | 35,214,276 | 886 |

***Main analysis / Prevalent cohort 2019***

| *Brain disorder* | *Cost component* | *Actual costs (EUR)* | *Actual costs per person* | *Attributable costs (EUR)* | *Attributable costs per person* |
|---|---|---|---|---|---|
| | Nursing home / sheltered accomodation | 119,388,118 | 3,004 | 49,106,312 | 1,236 |
| | Lost productivity associated with illness | . | . | 332,983,353 | 21,246 |
| | Total cost (excl. lost productivity) | 531,975,975 | 13,386 | 281,105,285 | 7,074 |
| Schizophrenia spectrum disorders | Primary sector | 26,218,851 | 392 | 7,568,236 | 113 |
| | Hospital care | 748,156,431 | 11,181 | 637,426,346 | 9,526 |
| | Filled prescriptions | 52,071,729 | 778 | 37,696,218 | 563 |
| | Home care | 34,786,593 | 520 | 26,838,808 | 401 |
| | Nursing home / sheltered accomodation | 147,669,845 | 2,207 | 127,249,541 | 1,902 |
| | Lost productivity associated with illness | . | . | 2,301,619,956 | 41,307 |
| | Total cost (excl. lost productivity) | 1,008,903,449 | 15,078 | 836,779,148 | 12,505 |
| Sleep disorders | Primary sector | 357,210,380 | 486 | 109,254,875 | 149 |
| | Hospital care | 3,170,140,496 | 4,317 | 1,541,456,957 | 2,099 |
| | Filled prescriptions | 438,390,129 | 597 | 216,889,046 | 295 |
| | Home care | 377,378,974 | 514 | 150,337,642 | 205 |
| | Nursing home / sheltered accomodation | 1,184,174,077 | 1,613 | 473,759,055 | 645 |
| | Lost productivity associated with illness | . | . | 5,462,880,437 | 12,713 |
| | Total cost (excl. lost productivity) | 5,527,294,056 | 7,527 | 2,491,697,575 | 3,393 |
| Stress related disorders | Primary sector | 100,138,553 | 418 | 33,393,827 | 139 |
| | Hospital care | 1,128,853,386 | 4,707 | 748,223,074 | 3,120 |
| | Filled prescriptions | 100,442,361 | 419 | 52,046,561 | 217 |
| | Home care | 54,982,408 | 229 | 35,032,009 | 146 |
| | Nursing home / sheltered accomodation | 166,099,746 | 693 | 119,296,519 | 497 |
| | Lost productivity associated with illness | . | . | 4,860,367,742 | 24,384 |
| | Total cost (excl. lost productivity) | 1,550,516,455 | 6,466 | 987,991,990 | 4,120 |
| Stroke | Primary sector | 73,797,161 | 599 | 19,829,953 | 161 |
| | Hospital care | 716,324,131 | 5,814 | 296,039,250 | 2,403 |
| | Filled prescriptions | 102,014,507 | 828 | 42,164,349 | 342 |
| | Home care | 250,600,009 | 2,034 | 175,414,889 | 1,424 |
| | Nursing home / sheltered accomodation | 778,256,871 | 6,317 | 513,783,651 | 4,170 |
| | Lost productivity associated with illness | . | . | 805,957,219 | 21,988 |
| | Total cost (excl. lost productivity) | 1,920,992,679 | 15,593 | 1,047,232,092 | 8,500 |
| Traumatic brain injury | Primary sector | 78,986,049 | 329 | 15,857,551 | 66 |
| | Hospital care | 690,095,972 | 2,871 | 280,442,657 | 1,167 |
| | Filled prescriptions | 75,371,835 | 314 | 24,501,095 | 102 |
| | Home care | 92,339,719 | 384 | 53,177,869 | 221 |
| | Nursing home / sheltered accomodation | 329,991,013 | 1,373 | 206,677,016 | 860 |
| | Lost productivity associated with illness | . | . | 1,368,869,599 | 8,375 |
| | Total cost (excl. lost productivity) | 1,266,784,589 | 5,270 | 580,656,189 | 2,416 |

***Main analysis / Prevalent cohort 2020***

| *Brain disorder* | *Cost component* | *Actual costs (EUR)* | *Actual costs per person* | *Attributable costs (EUR)* | *Attributable costs per person* |
|---|---|---|---|---|---|
| Any brain disorder | Primary sector | 833,725,652 | 414 | 281,377,585 | 140 |
| | Hospital care | 6,932,063,260 | 3,439 | 3,583,750,840 | 1,778 |
| | Filled prescriptions | 902,722,178 | 448 | 493,667,336 | 245 |
| | Home care | 946,819,291 | 470 | 684,054,761 | 339 |
| | Nursing home / sheltered accomodation | 2,802,647,913 | 1,390 | 2,375,674,472 | 1,179 |
| | Lost productivity associated with illness | . | . | 21,870,933,132 | 16,714 |
| | Total cost (excl. lost productivity) | 12,417,978,294 | 6,161 | 7,418,524,993 | 3,681 |
| Alcohol abuse | Primary sector | 65,200,296 | 407 | 13,611,666 | 85 |
| | Hospital care | 993,535,295 | 6,195 | 619,156,395 | 3,861 |
| | Filled prescriptions | 96,993,726 | 605 | 46,384,227 | 289 |
| | Home care | 118,327,027 | 738 | 90,129,203 | 562 |
| | Nursing home / sheltered accomodation | 359,576,528 | 2,242 | 297,580,712 | 1,856 |
| | Lost productivity associated with illness | . | . | 3,747,690,581 | 34,605 |
| | Total cost (excl. lost productivity) | 1,633,632,871 | 10,187 | 1,066,862,204 | 6,652 |
| Anxiety disorders | Primary sector | 149,542,673 | 478 | 51,082,394 | 163 |
| | Hospital care | 1,342,240,492 | 4,288 | 707,220,435 | 2,260 |
| | Filled prescriptions | 166,616,089 | 532 | 86,029,993 | 275 |
| | Home care | 144,594,833 | 462 | 80,991,130 | 259 |
| | Nursing home / sheltered accomodation | 447,075,918 | 1,428 | 284,850,223 | 910 |
| | Lost productivity associated with illness | . | . | 5,666,172,279 | 23,820 |
| | Total cost (excl. lost productivity) | 2,250,070,005 | 7,189 | 1,210,174,174 | 3,866 |
| Bipolar disorder | Primary sector | 20,139,618 | 546 | 7,659,307 | 208 |
| | Hospital care | 317,935,662 | 8,621 | 233,647,117 | 6,335 |
| | Filled prescriptions | 31,648,049 | 858 | 20,557,658 | 557 |
| | Home care | 24,161,547 | 655 | 16,099,593 | 437 |
| | Nursing home / sheltered accomodation | 87,798,293 | 2,381 | 68,178,939 | 1,849 |
| | Lost productivity associated with illness | . | . | 927,408,394 | 34,467 |
| | Total cost (excl. lost productivity) | 481,683,170 | 13,060 | 346,142,614 | 9,385 |
| Brain tumors | Primary sector | 9,856,721 | 535 | 2,535,488 | 138 |
| | Hospital care | 129,386,568 | 7,023 | 75,710,305 | 4,110 |
| | Filled prescriptions | 11,664,009 | 633 | 4,113,380 | 223 |
| | Home care | 17,580,223 | 954 | 8,926,944 | 485 |
| | Nursing home / sheltered accomodation | 43,668,615 | 2,370 | 20,992,280 | 1,139 |

*Main analysis / Prevalent cohort 2020*

| Brain disorder | Cost component | Actual costs (EUR) | Actual costs per person | Attributable costs (EUR) | Attributable costs per person |
|---|---|---|---|---|---|
| | Lost productivity associated with illness | . | . | 96,465,473 | 11,200 |
| | Total cost (excl. lost productivity) | 212,156,136 | 11,516 | 112,278,397 | 6,095 |
| Cerebral palsy | Primary sector | 6,169,094 | 661 | 4,152,670 | 445 |
| | Hospital care | 38,091,301 | 4,079 | 24,964,832 | 2,674 |
| | Filled prescriptions | 8,170,309 | 875 | 6,746,739 | 723 |
| | Home care | 15,798,819 | 1,692 | 15,189,573 | 1,627 |
| | Nursing home / sheltered accomodation | 13,885,651 | 1,487 | 12,435,138 | 1,332 |
| | Lost productivity associated with illness | . | . | 167,982,853 | 30,459 |
| | Total cost (excl. lost productivity) | 82,115,174 | 8,794 | 63,488,951 | 6,799 |
| Dementia | Primary sector | 26,720,129 | 669 | 5,259,296 | 132 |
| | Hospital care | 202,344,537 | 5,064 | 24,225,157 | 606 |
| | Filled prescriptions | 48,102,487 | 1,204 | 22,688,937 | 568 |
| | Home care | 164,469,705 | 4,116 | 104,605,994 | 2,618 |
| | Nursing home / sheltered accomodation | 1,331,608,245 | 33,327 | 1,194,944,389 | 29,906 |
| | Lost productivity associated with illness | . | . | 151,449,693 | 44,981 |
| | Total cost (excl. lost productivity) | 1,773,245,103 | 44,380 | 1,351,723,772 | 33,830 |
| Depression | Primary sector | 369,809,915 | 478 | 121,109,199 | 157 |
| | Hospital care | 3,193,445,886 | 4,129 | 1,563,664,163 | 2,022 |
| | Filled prescriptions | 431,074,075 | 557 | 224,052,960 | 290 |
| | Home care | 500,417,810 | 647 | 321,272,965 | 415 |
| | Nursing home / sheltered accomodation | 1,662,594,901 | 2,150 | 1,252,901,858 | 1,620 |
| | Lost productivity associated with illness | . | . | 13,849,049,818 | 25,526 |
| | Total cost (excl. lost productivity) | 6,157,342,587 | 7,961 | 3,483,001,145 | 4,503 |
| Developmental and behavioural disorders | Primary sector | 58,894,227 | 316 | 24,308,537 | 130 |
| | Hospital care | 590,148,837 | 3,163 | 401,107,973 | 2,150 |
| | Filled prescriptions | 80,362,462 | 431 | 61,718,327 | 331 |
| | Home care | 8,672,923 | 46 | 5,396,746 | 29 |
| | Nursing home / sheltered accomodation | 17,448,331 | 94 | 12,754,426 | 68 |
| | Lost productivity associated with illness | . | . | 2,617,609,994 | 20,222 |
| | Total cost (excl. lost productivity) | 755,526,781 | 4,049 | 505,286,009 | 2,708 |
| Drug abuse | Primary sector | 20,390,670 | 348 | 6,372,879 | 109 |
| | Hospital care | 483,173,520 | 8,241 | 397,631,974 | 6,782 |
| | Filled prescriptions | 33,737,492 | 575 | 23,495,725 | 401 |

***Main analysis / Prevalent cohort 2020***

| *Brain disorder* | *Cost component* | *Actual costs (EUR)* | *Actual costs per person* | *Attributable costs (EUR)* | *Attributable costs per person* |
|---|---|---|---|---|---|
| | Home care | 21,720,004 | 370 | 17,542,911 | 299 |
| | Nursing home / sheltered accomodation | 51,005,917 | 870 | 41,628,474 | 710 |
| | Lost productivity associated with illness | . | . | 2,024,090,608 | 37,926 |
| | Total cost (excl. lost productivity) | 610,027,603 | 10,405 | 486,671,964 | 8,301 |
| Eating disorders | Primary sector | 8,190,029 | 427 | 2,528,989 | 132 |
| | Hospital care | 138,766,605 | 7,232 | 106,464,575 | 5,549 |
| | Filled prescriptions | 5,673,053 | 296 | 2,738,738 | 143 |
| | Home care | 1,157,475 | 60 | 778,924 | 41 |
| | Nursing home / sheltered accomodation | 1,635,669 | 85 | 1,391,409 | 73 |
| | Lost productivity associated with illness | . | . | 125,055,219 | 7,121 |
| | Total cost (excl. lost productivity) | 155,422,833 | 8,100 | 113,902,635 | 5,936 |
| Epilepsy | Primary sector | 31,662,738 | 448 | 10,394,892 | 147 |
| | Hospital care | 349,889,796 | 4,951 | 201,253,447 | 2,848 |
| | Filled prescriptions | 67,960,740 | 962 | 49,437,916 | 700 |
| | Home care | 77,948,168 | 1,103 | 62,528,479 | 885 |
| | Nursing home / sheltered accomodation | 210,401,036 | 2,977 | 168,511,479 | 2,385 |
| | Lost productivity associated with illness | . | . | 947,912,706 | 21,743 |
| | Total cost (excl. lost productivity) | 737,862,478 | 10,441 | 492,126,214 | 6,964 |
| Headache | Primary sector | 199,385,533 | 436 | 49,940,627 | 109 |
| | Hospital care | 1,311,798,094 | 2,871 | 347,526,093 | 761 |
| | Filled prescriptions | 183,573,513 | 402 | 60,749,164 | 133 |
| | Home care | 91,440,299 | 200 | 10,592,814 | 23 |
| | Nursing home / sheltered accomodation | 193,379,949 | 423 | 8,247,273 | 18 |
| | Lost productivity associated with illness | . | . | 1,528,438,223 | 4,440 |
| | Total cost (excl. lost productivity) | 1,979,577,388 | 4,333 | 477,055,971 | 1,044 |
| Infections of the CNS | Primary sector | 7,395,600 | 381 | 1,905,738 | 98 |
| | Hospital care | 80,430,121 | 4,144 | 42,793,626 | 2,205 |
| | Filled prescriptions | 8,416,242 | 434 | 3,661,332 | 189 |
| | Home care | 12,258,484 | 632 | 8,578,012 | 442 |
| | Nursing home / sheltered accomodation | 28,326,258 | 1,459 | 19,052,946 | 982 |
| | Lost productivity associated with illness | . | . | 65,291,798 | 5,315 |
| | Total cost (excl. lost productivity) | 136,826,706 | 7,049 | 75,991,655 | 3,915 |
| Intellectual disability | Primary sector | 12,642,144 | 411 | 5,776,780 | 188 |
| | Hospital care | 154,701,141 | 5,026 | 111,624,359 | 3,627 |
| | Filled prescriptions | 24,203,736 | 786 | 19,601,398 | 637 |
| | Home care | 12,148,694 | 395 | 10,534,688 | 342 |
| | Nursing home / sheltered accomodation | 31,979,890 | 1,039 | 28,820,066 | 936 |
| | Lost productivity associated with illness | . | . | 798,909,765 | 36,150 |
| | Total cost (excl. lost productivity) | 235,675,606 | 7,657 | 176,357,291 | 5,730 |

***Main analysis / Prevalent cohort 2020***

| *Brain disorder* | *Cost component* | *Actual costs (EUR)* | *Actual costs per person* | *Attributable costs (EUR)* | *Attributable costs per person* |
|---|---|---|---|---|---|
| Multiple sclerosis | Primary sector | 12,714,467 | 757 | 7,010,644 | 417 |
| | Hospital care | 122,232,986 | 7,275 | 84,537,895 | 5,031 |
| | Filled prescriptions | 10,499,177 | 625 | 5,541,921 | 330 |
| | Home care | 58,619,714 | 3,489 | 56,077,799 | 3,338 |
| | Nursing home / sheltered accomodation | 36,552,707 | 2,176 | 30,745,732 | 1,830 |
| | Lost productivity associated with illness | . | . | 265,403,104 | 20,657 |
| | Total cost (excl. lost productivity) | 240,619,052 | 14,321 | 183,913,990 | 10,946 |
| Neuromuscular disorders | Primary sector | 4,843,482 | 580 | 2,092,840 | 251 |
| | Hospital care | 52,611,576 | 6,304 | 32,881,188 | 3,940 |
| | Filled prescriptions | 5,573,527 | 668 | 2,968,406 | 356 |
| | Home care | 11,993,566 | 1,437 | 9,768,526 | 1,170 |
| | Nursing home / sheltered accomodation | 11,784,298 | 1,412 | 5,824,288 | 698 |
| | Lost productivity associated with illness | . | . | 88,920,253 | 17,777 |
| | Total cost (excl. lost productivity) | 86,806,449 | 10,401 | 53,535,248 | 6,415 |
| Other neurodegenerative disorders | Primary sector | 3,030,682 | 791 | 1,623,486 | 424 |
| | Hospital care | 28,817,314 | 7,519 | 17,948,170 | 4,683 |
| | Filled prescriptions | 3,304,791 | 862 | 1,862,590 | 486 |
| | Home care | 18,389,229 | 4,798 | 17,118,901 | 4,467 |
| | Nursing home / sheltered accomodation | 23,125,178 | 6,034 | 19,738,541 | 5,150 |
| | Lost productivity associated with illness | . | . | 58,812,535 | 32,244 |
| | Total cost (excl. lost productivity) | 76,667,194 | 20,004 | 58,291,690 | 15,209 |
| Parkinson’s disease | Primary sector | 20,107,956 | 944 | 10,206,657 | 479 |
| | Hospital care | 138,096,380 | 6,481 | 56,186,586 | 2,637 |
| | Filled prescriptions | 45,779,779 | 2,148 | 34,646,189 | 1,626 |
| | Home care | 74,830,280 | 3,512 | 58,722,305 | 2,756 |
| | Nursing home / sheltered accomodation | 209,435,430 | 9,829 | 162,312,498 | 7,617 |
| | Lost productivity associated with illness | . | . | 158,513,144 | 29,953 |
| | Total cost (excl. lost productivity) | 488,249,824 | 22,913 | 322,074,234 | 15,115 |
| Personality disorders | Primary sector | 33,975,590 | 451 | 12,173,212 | 162 |
| | Hospital care | 502,831,407 | 6,675 | 374,294,983 | 4,969 |
| | Filled prescriptions | 39,277,368 | 521 | 24,605,754 | 327 |
| | Home care | 18,438,288 | 245 | 13,750,915 | 183 |
| | Nursing home / sheltered accomodation | 41,239,372 | 547 | 33,278,370 | 442 |
| | Lost productivity associated with illness | . | . | 2,324,853,244 | 33,359 |
| | Total cost (excl. lost productivity) | 635,762,025 | 8,440 | 458,103,232 | 6,081 |
| Polyneuropathy | Primary sector | 26,044,024 | 625 | 8,589,205 | 206 |
| | Hospital care | 328,288,566 | 7,881 | 188,504,491 | 4,525 |
| | Filled prescriptions | 42,502,049 | 1,020 | 23,233,376 | 558 |
| | Home care | 66,337,024 | 1,593 | 41,833,360 | 1,004 |

***Main analysis / Prevalent cohort 2020***

| *Brain disorder* | *Cost component* | *Actual costs (EUR)* | *Actual costs per person* | *Attributable costs (EUR)* | *Attributable costs per person* |
|---|---|---|---|---|---|
| | Nursing home / sheltered accomodation | 125,629,582 | 3,016 | 54,339,261 | 1,305 |
| | Lost productivity associated with illness | . | . | 362,380,942 | 22,531 |
| | Total cost (excl. lost productivity) | 588,801,244 | 14,135 | 316,499,695 | 7,598 |
| Schizophrenia spectrum disorders | Primary sector | 26,730,454 | 391 | 7,668,319 | 112 |
| | Hospital care | 795,722,386 | 11,630 | 677,831,234 | 9,907 |
| | Filled prescriptions | 53,372,949 | 780 | 38,377,189 | 561 |
| | Home care | 40,455,340 | 591 | 31,752,344 | 464 |
| | Nursing home / sheltered accomodation | 144,019,153 | 2,105 | 124,516,989 | 1,820 |
| | Lost productivity associated with illness | . | . | 2,428,486,677 | 42,556 |
| | Total cost (excl. lost productivity) | 1,060,300,282 | 15,498 | 880,146,075 | 12,864 |
| Sleep disorders | Primary sector | 368,829,179 | 489 | 113,459,111 | 151 |
| | Hospital care | 3,410,875,783 | 4,525 | 1,672,468,381 | 2,219 |
| | Filled prescriptions | 459,311,905 | 609 | 227,228,443 | 301 |
| | Home care | 432,650,952 | 574 | 177,140,001 | 235 |
| | Nursing home / sheltered accomodation | 1,200,698,393 | 1,593 | 493,145,720 | 654 |
| | Lost productivity associated with illness | . | . | 5,998,477,351 | 13,603 |
| | Total cost (excl. lost productivity) | 5,872,366,212 | 7,791 | 2,683,441,657 | 3,560 |
| Stress related disorders | Primary sector | 103,501,010 | 417 | 33,998,023 | 137 |
| | Hospital care | 1,222,075,122 | 4,923 | 808,195,185 | 3,255 |
| | Filled prescriptions | 106,128,467 | 427 | 54,235,168 | 218 |
| | Home care | 61,720,983 | 249 | 38,561,005 | 155 |
| | Nursing home / sheltered accomodation | 165,957,961 | 668 | 118,001,282 | 475 |
| | Lost productivity associated with illness | . | . | 5,272,184,424 | 25,610 |
| | Total cost (excl. lost productivity) | 1,659,383,544 | 6,684 | 1,052,990,661 | 4,242 |
| Stroke | Primary sector | 74,799,977 | 600 | 19,115,405 | 153 |
| | Hospital care | 773,669,068 | 6,201 | 325,554,315 | 2,609 |
| | Filled prescriptions | 106,000,850 | 850 | 43,608,081 | 350 |
| | Home care | 276,332,892 | 2,215 | 191,156,016 | 1,532 |
| | Nursing home / sheltered accomodation | 777,477,161 | 6,231 | 515,427,657 | 4,131 |
| | Lost productivity associated with illness | . | . | 846,104,640 | 23,125 |
| | Total cost (excl. lost productivity) | 2,008,279,948 | 16,096 | 1,094,861,475 | 8,775 |
| Traumatic brain injury | Primary sector | 79,437,920 | 334 | 15,839,264 | 67 |
| | Hospital care | 720,494,064 | 3,030 | 293,315,074 | 1,234 |
| | Filled prescriptions | 77,855,323 | 327 | 25,254,545 | 106 |
| | Home care | 102,394,940 | 431 | 56,983,192 | 240 |
| | Nursing home / sheltered accomodation | 336,599,017 | 1,416 | 211,568,257 | 890 |
| | Lost productivity associated with illness | . | . | 1,369,830,185 | 8,537 |
| | Total cost (excl. lost productivity) | 1,316,781,263 | 5,538 | 602,960,332 | 2,536 |

***Main analysis / Prevalent cohort 2021***

| *Brain disorder* | *Cost component* | *Actual costs (EUR)* | *Actual costs per person* | *Attributable costs (EUR)* | *Attributable costs per person* |
|---|---|---|---|---|---|
| Any brain disorder | Primary sector | 865,449,326 | 425 | 292,607,841 | 144 |
| | Hospital care | 6,957,360,854 | 3,414 | 3,591,877,299 | 1,763 |
| | Filled prescriptions | 885,263,634 | 434 | 478,703,967 | 235 |
| | Home care | 1,068,554,217 | 524 | 782,066,718 | 384 |
| | Nursing home / sheltered accomodation | 2,778,761,929 | 1,364 | 2,354,587,950 | 1,155 |
| | Lost productivity associated with illness | . | . | 23,195,589,686 | 17,587 |
| | Total cost (excl. lost productivity) | 12,555,389,960 | 6,161 | 7,499,843,775 | 3,680 |
| Alcohol abuse | Primary sector | 67,300,900 | 422 | 14,458,107 | 91 |
| | Hospital care | 980,102,459 | 6,143 | 611,778,103 | 3,835 |
| | Filled prescriptions | 92,924,408 | 582 | 43,049,156 | 270 |
| | Home care | 127,719,661 | 801 | 95,668,121 | 600 |
| | Nursing home / sheltered accomodation | 359,310,183 | 2,252 | 297,553,937 | 1,865 |
| | Lost productivity associated with illness | . | . | 3,782,912,370 | 35,399 |
| | Total cost (excl. lost productivity) | 1,627,357,611 | 10,200 | 1,062,507,424 | 6,660 |
| Anxiety disorders | Primary sector | 158,358,090 | 489 | 53,574,497 | 165 |
| | Hospital care | 1,369,234,383 | 4,225 | 721,603,130 | 2,227 |
| | Filled prescriptions | 161,604,316 | 499 | 79,949,762 | 247 |
| | Home care | 160,334,044 | 495 | 88,359,547 | 273 |
| | Nursing home / sheltered accomodation | 439,743,447 | 1,357 | 277,490,154 | 856 |
| | Lost productivity associated with illness | . | . | 6,048,778,636 | 24,577 |
| | Total cost (excl. lost productivity) | 2,289,274,280 | 7,064 | 1,220,977,091 | 3,768 |
| Bipolar disorder | Primary sector | 20,641,768 | 551 | 7,702,439 | 206 |
| | Hospital care | 313,497,127 | 8,374 | 229,334,400 | 6,126 |
| | Filled prescriptions | 30,029,258 | 802 | 19,129,686 | 511 |
| | Home care | 26,559,466 | 709 | 17,831,202 | 476 |
| | Nursing home / sheltered accomodation | 86,021,259 | 2,298 | 67,327,780 | 1,798 |
| | Lost productivity associated with illness | . | . | 966,287,536 | 35,154 |
| | Total cost (excl. lost productivity) | 476,748,878 | 12,735 | 341,325,507 | 9,117 |
| Brain tumors | Primary sector | 10,321,932 | 541 | 2,535,444 | 133 |
| | Hospital care | 136,319,065 | 7,151 | 81,197,461 | 4,259 |
| | Filled prescriptions | 11,154,889 | 585 | 3,477,652 | 182 |
| | Home care | 21,286,988 | 1,117 | 11,860,170 | 622 |
| | Nursing home / sheltered accomodation | 43,788,764 | 2,297 | 20,342,030 | 1,067 |

*Main analysis / Prevalent cohort 2021*

| *Brain disorder* | *Cost component* | *Actual costs (EUR)* | *Actual costs per person* | *Attributable costs (EUR)* | *Attributable costs per person* |
|---|---|---|---|---|---|
| | Lost productivity associated with illness | . | . | 98,700,847 | 11,172 |
| | Total cost (excl. lost productivity) | 222,871,638 | 11,691 | 119,412,758 | 6,264 |
| Cerebral palsy | Primary sector | 6,526,579 | 703 | 4,433,803 | 478 |
| | Hospital care | 39,715,357 | 4,279 | 27,067,968 | 2,916 |
| | Filled prescriptions | 8,172,995 | 881 | 6,814,448 | 734 |
| | Home care | 20,839,416 | 2,245 | 20,237,373 | 2,180 |
| | Nursing home / sheltered accomodation | 14,122,589 | 1,522 | 12,869,341 | 1,387 |
| | Lost productivity associated with illness | . | . | 178,819,263 | 32,365 |
| | Total cost (excl. lost productivity) | 89,376,936 | 9,630 | 71,422,933 | 7,695 |
| Dementia | Primary sector | 28,178,314 | 715 | 6,728,510 | 171 |
| | Hospital care | 192,519,124 | 4,886 | 19,182,599 | 487 |
| | Filled prescriptions | 45,349,547 | 1,151 | 21,176,777 | 537 |
| | Home care | 173,724,521 | 4,409 | 110,604,105 | 2,807 |
| | Nursing home / sheltered accomodation | 1,291,677,913 | 32,781 | 1,165,450,860 | 29,577 |
| | Lost productivity associated with illness | . | . | 151,063,138 | 47,178 |
| | Total cost (excl. lost productivity) | 1,731,449,419 | 43,942 | 1,323,142,851 | 33,579 |
| Depression | Primary sector | 383,346,326 | 490 | 124,166,046 | 159 |
| | Hospital care | 3,202,289,382 | 4,089 | 1,569,365,821 | 2,004 |
| | Filled prescriptions | 418,259,695 | 534 | 211,712,958 | 270 |
| | Home care | 554,289,410 | 708 | 354,368,581 | 453 |
| | Nursing home / sheltered accomodation | 1,625,729,847 | 2,076 | 1,216,841,595 | 1,554 |
| | Lost productivity associated with illness | . | . | 14,494,797,018 | 26,533 |
| | Total cost (excl. lost productivity) | 6,183,914,660 | 7,897 | 3,476,455,002 | 4,440 |
| Developmental and behavioural disorders | Primary sector | 65,555,288 | 329 | 27,079,126 | 136 |
| | Hospital care | 645,356,618 | 3,235 | 439,697,147 | 2,204 |
| | Filled prescriptions | 85,769,986 | 430 | 65,894,680 | 330 |
| | Home care | 15,279,082 | 77 | 10,968,398 | 55 |
| | Nursing home / sheltered accomodation | 18,329,076 | 92 | 13,617,787 | 68 |
| | Lost productivity associated with illness | . | . | 2,974,061,152 | 21,032 |
| | Total cost (excl. lost productivity) | 830,290,051 | 4,162 | 557,257,137 | 2,793 |
| Drug abuse | Primary sector | 21,070,889 | 353 | 6,310,682 | 106 |
| | Hospital care | 479,970,816 | 8,038 | 392,641,067 | 6,575 |
| | Filled prescriptions | 33,393,380 | 559 | 23,123,277 | 387 |

***Main analysis / Prevalent cohort 2021***

| *Brain disorder* | *Cost component* | *Actual costs (EUR)* | *Actual costs per person* | *Attributable costs (EUR)* | *Attributable costs per person* |
|---|---|---|---|---|---|
| | Home care | 24,389,108 | 408 | 19,628,464 | 329 |
| | Nursing home / sheltered accomodation | 50,061,106 | 838 | 41,348,726 | 692 |
| | Lost productivity associated with illness | . | . | 2,126,992,387 | 39,064 |
| | Total cost (excl. lost productivity) | 608,885,299 | 10,197 | 483,052,216 | 8,089 |
| Eating disorders | Primary sector | 8,641,707 | 439 | 2,598,253 | 132 |
| | Hospital care | 146,421,706 | 7,434 | 113,859,541 | 5,781 |
| | Filled prescriptions | 5,778,497 | 293 | 2,767,124 | 140 |
| | Home care | 1,331,351 | 68 | 876,233 | 44 |
| | Nursing home / sheltered accomodation | 1,544,507 | 78 | 1,335,872 | 68 |
| | Lost productivity associated with illness | . | . | 128,547,028 | 7,148 |
| | Total cost (excl. lost productivity) | 163,717,768 | 8,313 | 121,437,022 | 6,166 |
| Epilepsy | Primary sector | 32,506,905 | 466 | 10,897,467 | 156 |
| | Hospital care | 341,166,561 | 4,889 | 193,687,437 | 2,775 |
| | Filled prescriptions | 61,579,885 | 882 | 43,449,062 | 623 |
| | Home care | 90,457,061 | 1,296 | 73,229,813 | 1,049 |
| | Nursing home / sheltered accomodation | 206,567,179 | 2,960 | 165,040,260 | 2,365 |
| | Lost productivity associated with illness | . | . | 955,388,750 | 22,365 |
| | Total cost (excl. lost productivity) | 732,277,592 | 10,493 | 486,304,040 | 6,968 |
| Headache | Primary sector | 205,374,145 | 447 | 50,684,166 | 110 |
| | Hospital care | 1,296,643,356 | 2,824 | 337,913,801 | 736 |
| | Filled prescriptions | 178,301,477 | 388 | 58,068,888 | 126 |
| | Home care | 102,319,046 | 223 | 11,295,389 | 25 |
| | Nursing home / sheltered accomodation | 193,365,245 | 421 | 9,886,883 | 22 |
| | Lost productivity associated with illness | . | . | 1,668,262,550 | 4,824 |
| | Total cost (excl. lost productivity) | 1,976,003,268 | 4,303 | 467,849,127 | 1,019 |
| Infections of the CNS | Primary sector | 7,497,166 | 395 | 1,912,670 | 101 |
| | Hospital care | 81,890,282 | 4,313 | 45,078,092 | 2,374 |
| | Filled prescriptions | 8,085,316 | 426 | 3,462,765 | 182 |
| | Home care | 13,335,824 | 702 | 9,531,439 | 502 |
| | Nursing home / sheltered accomodation | 28,634,403 | 1,508 | 19,348,414 | 1,019 |
| | Lost productivity associated with illness | . | . | 66,261,206 | 5,569 |
| | Total cost (excl. lost productivity) | 139,442,991 | 7,345 | 79,333,379 | 4,179 |
| Intellectual disability | Primary sector | 13,261,092 | 431 | 6,153,184 | 200 |
| | Hospital care | 163,662,708 | 5,321 | 120,867,101 | 3,930 |
| | Filled prescriptions | 23,887,092 | 777 | 19,313,685 | 628 |
| | Home care | 18,852,632 | 613 | 17,187,854 | 559 |
| | Nursing home / sheltered accomodation | 31,600,748 | 1,027 | 28,735,945 | 934 |
| | Lost productivity associated with illness | . | . | 829,514,511 | 37,029 |
| | Total cost (excl. lost productivity) | 251,264,272 | 8,170 | 192,257,769 | 6,251 |

***Main analysis / Prevalent cohort 2021***

| *Brain disorder* | *Cost component* | *Actual costs (EUR)* | *Actual costs per person* | *Attributable costs (EUR)* | *Attributable costs per person* |
|---|---|---|---|---|---|
| Multiple sclerosis | Primary sector | 13,533,717 | 787 | 7,489,392 | 436 |
| | Hospital care | 124,524,169 | 7,245 | 86,356,566 | 5,024 |
| | Filled prescriptions | 9,993,905 | 581 | 4,934,470 | 287 |
| | Home care | 65,155,136 | 3,791 | 62,238,975 | 3,621 |
| | Nursing home / sheltered accomodation | 37,086,868 | 2,158 | 31,155,102 | 1,813 |
| | Lost productivity associated with illness | . | . | 272,092,544 | 20,861 |
| | Total cost (excl. lost productivity) | 250,293,795 | 14,562 | 192,174,505 | 11,181 |
| Neuromuscular disorders | Primary sector | 5,031,654 | 593 | 2,149,818 | 254 |
| | Hospital care | 50,903,497 | 6,003 | 31,136,241 | 3,672 |
| | Filled prescriptions | 5,436,854 | 641 | 2,839,909 | 335 |
| | Home care | 14,358,581 | 1,693 | 12,077,950 | 1,424 |
| | Nursing home / sheltered accomodation | 11,264,841 | 1,329 | 5,313,674 | 627 |
| | Lost productivity associated with illness | . | . | 89,802,597 | 17,800 |
| | Total cost (excl. lost productivity) | 86,995,427 | 10,260 | 53,517,593 | 6,312 |
| Other neurodegenerative disorders | Primary sector | 3,170,840 | 823 | 1,721,498 | 447 |
| | Hospital care | 33,657,967 | 8,733 | 22,917,182 | 5,946 |
| | Filled prescriptions | 3,059,422 | 794 | 1,566,196 | 406 |
| | Home care | 19,085,341 | 4,952 | 17,603,690 | 4,568 |
| | Nursing home / sheltered accomodation | 23,222,011 | 6,025 | 19,728,940 | 5,119 |
| | Lost productivity associated with illness | . | . | 59,682,920 | 32,631 |
| | Total cost (excl. lost productivity) | 82,195,582 | 21,328 | 63,537,507 | 16,486 |
| Parkinson’s disease | Primary sector | 21,124,590 | 981 | 10,992,945 | 511 |
| | Hospital care | 133,885,055 | 6,220 | 52,435,222 | 2,436 |
| | Filled prescriptions | 45,474,761 | 2,113 | 34,629,277 | 1,609 |
| | Home care | 85,491,344 | 3,972 | 68,511,691 | 3,183 |
| | Nursing home / sheltered accomodation | 209,733,073 | 9,744 | 164,087,837 | 7,623 |
| | Lost productivity associated with illness | . | . | 174,516,672 | 32,841 |
| | Total cost (excl. lost productivity) | 495,708,822 | 23,030 | 330,656,972 | 15,362 |
| Personality disorders | Primary sector | 34,947,485 | 463 | 12,343,879 | 163 |
| | Hospital care | 506,465,838 | 6,704 | 378,135,983 | 5,005 |
| | Filled prescriptions | 36,991,862 | 490 | 22,503,788 | 298 |
| | Home care | 20,435,946 | 271 | 15,324,793 | 203 |
| | Nursing home / sheltered accomodation | 38,200,140 | 506 | 30,305,047 | 401 |
| | Lost productivity associated with illness | . | . | 2,399,317,450 | 34,284 |
| | Total cost (excl. lost productivity) | 637,041,269 | 8,433 | 458,613,490 | 6,071 |
| Polyneuropathy | Primary sector | 27,796,232 | 640 | 9,258,656 | 213 |
| | Hospital care | 336,539,484 | 7,745 | 193,299,601 | 4,449 |
| | Filled prescriptions | 45,063,592 | 1,037 | 25,465,907 | 586 |
| | Home care | 76,074,731 | 1,751 | 48,881,338 | 1,125 |

***Main analysis / Prevalent cohort 2021***

| *Brain disorder* | *Cost component* | *Actual costs (EUR)* | *Actual costs per person* | *Attributable costs (EUR)* | *Attributable costs per person* |
|---|---|---|---|---|---|
| | Nursing home / sheltered accomodation | 126,302,376 | 2,907 | 52,959,914 | 1,219 |
| | Lost productivity associated with illness | . | . | 393,226,038 | 24,042 |
| | Total cost (excl. lost productivity) | 611,776,415 | 14,079 | 329,865,416 | 7,592 |
| Schizophrenia spectrum disorders | Primary sector | 27,818,015 | 399 | 7,757,907 | 111 |
| | Hospital care | 806,291,724 | 11,550 | 687,190,047 | 9,844 |
| | Filled prescriptions | 51,384,380 | 736 | 36,588,948 | 524 |
| | Home care | 46,322,900 | 664 | 36,935,829 | 529 |
| | Nursing home / sheltered accomodation | 140,161,981 | 2,008 | 121,682,604 | 1,743 |
| | Lost productivity associated with illness | . | . | 2,552,794,600 | 43,721 |
| | Total cost (excl. lost productivity) | 1,071,979,001 | 15,356 | 890,155,335 | 12,752 |
| Sleep disorders | Primary sector | 387,686,990 | 498 | 119,075,274 | 153 |
| | Hospital care | 3,485,909,890 | 4,482 | 1,729,270,066 | 2,223 |
| | Filled prescriptions | 459,868,931 | 591 | 229,721,529 | 295 |
| | Home care | 497,728,808 | 640 | 214,187,061 | 275 |
| | Nursing home / sheltered accomodation | 1,213,969,644 | 1,561 | 517,012,424 | 665 |
| | Lost productivity associated with illness | . | . | 6,539,006,557 | 14,356 |
| | Total cost (excl. lost productivity) | 6,045,164,263 | 7,772 | 2,809,266,354 | 3,612 |
| Stress related disorders | Primary sector | 108,404,313 | 426 | 34,736,261 | 137 |
| | Hospital care | 1,245,735,577 | 4,898 | 824,071,456 | 3,240 |
| | Filled prescriptions | 104,753,617 | 412 | 52,611,879 | 207 |
| | Home care | 72,280,953 | 284 | 45,542,248 | 179 |
| | Nursing home / sheltered accomodation | 166,517,118 | 655 | 118,067,867 | 464 |
| | Lost productivity associated with illness | . | . | 5,571,484,154 | 26,429 |
| | Total cost (excl. lost productivity) | 1,697,691,578 | 6,675 | 1,075,029,712 | 4,227 |
| Stroke | Primary sector | 77,577,444 | 614 | 20,311,441 | 161 |
| | Hospital care | 763,050,453 | 6,036 | 315,333,542 | 2,494 |
| | Filled prescriptions | 102,704,139 | 812 | 40,725,047 | 322 |
| | Home care | 308,795,566 | 2,443 | 215,929,137 | 1,708 |
| | Nursing home / sheltered accomodation | 759,625,321 | 6,009 | 500,300,815 | 3,958 |
| | Lost productivity associated with illness | . | . | 878,570,058 | 24,152 |
| | Total cost (excl. lost productivity) | 2,011,752,924 | 15,914 | 1,092,599,982 | 8,643 |
| Traumatic brain injury | Primary sector | 80,534,550 | 346 | 16,217,542 | 70 |
| | Hospital care | 713,802,173 | 3,068 | 293,606,401 | 1,262 |
| | Filled prescriptions | 75,190,007 | 323 | 24,021,612 | 103 |
| | Home care | 116,464,191 | 501 | 66,574,483 | 286 |
| | Nursing home / sheltered accomodation | 337,430,820 | 1,450 | 213,029,152 | 916 |
| | Lost productivity associated with illness | . | . | 1,392,226,424 | 8,926 |
| | Total cost (excl. lost productivity) | 1,323,421,740 | 5,688 | 613,449,190 | 2,636 |

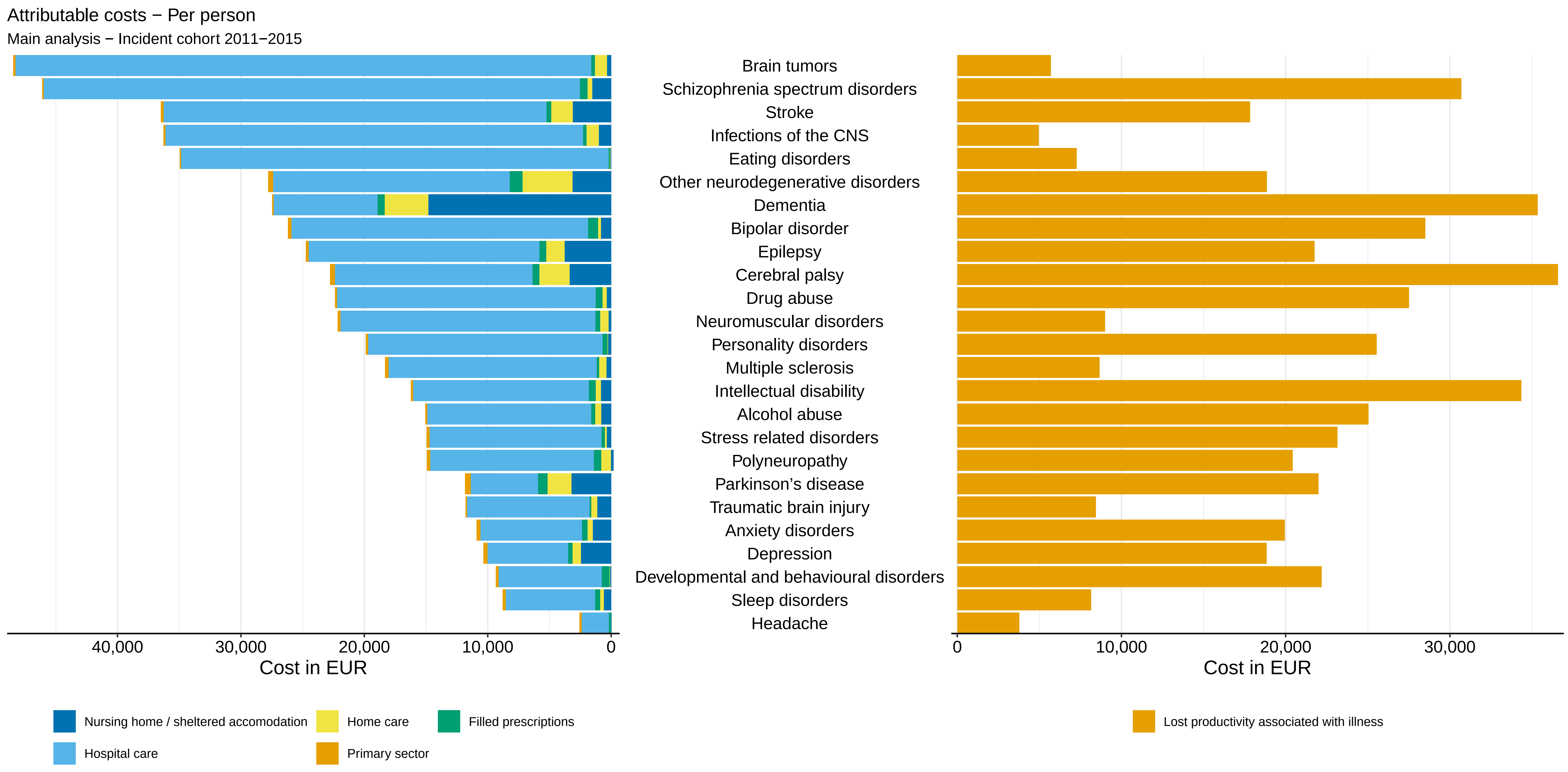
Attributable costs − Per person
Main analysis − Incident cohort 2011−2015
Brain tumors
Schizophrenia spectrum disorders
Stroke
Infections of the CNS
Eating disorders
Other neurodegenerative disorders
Dementia
Bipolar disorder
Epilepsy
Cerebral palsy
Drug abuse
Neuromuscular disorders
Personality disorders
Multiple sclerosis
Intellectual disability
Alcohol abuse
Stress related disorders
Polyneuropathy
Parkinson's disease
Traumatic brain injury
Anxiety disorders
Depression
Developmental and behavioural disorders
Sleep disorders
Headache
40,000
30,000
20,000
10,000
0
Cost in EUR
0
10,000
20,000
30,000
Cost in EUR
Nursing home / sheltered accomodation
Home care
Filled prescriptions
Hospital care
Primary sector
Lost productivity associated with illness

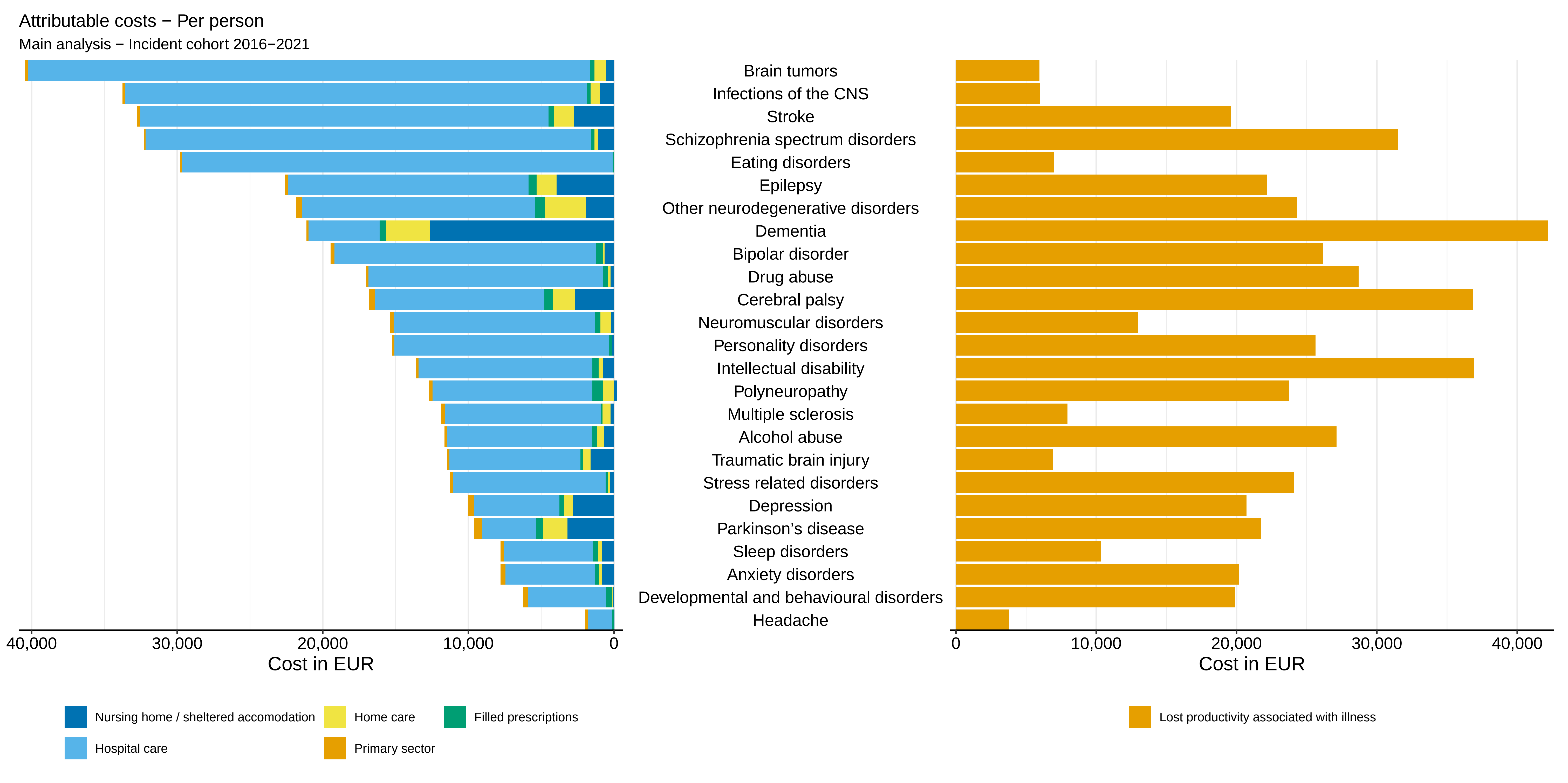
Attributable costs − Per person
Main analysis − Incident cohort 2016−2021
Brain tumors
Infections of the CNS
Stroke
Schizophrenia spectrum disorders
Eating disorders
Epilepsy
Other neurodegenerative disorders
Dementia
Bipolar disorder
Drug abuse
Cerebral palsy
Neuromuscular disorders
Personality disorders
Intellectual disability
Polyneuropathy
Multiple sclerosis
Alcohol abuse
Traumatic brain injury
Stress related disorders
Depression
Parkinson’s disease
Sleep disorders
Anxiety disorders
Developmental and behavioural disorders
Headache
40,000
30,000
20,000
10,000
0
Cost in EUR
0
10,000
20,000
30,000
40,000
Cost in EUR
Nursing home / sheltered accomodation
Home care
Filled prescriptions
Hospital care
Primary sector
Lost productivity associated with illness

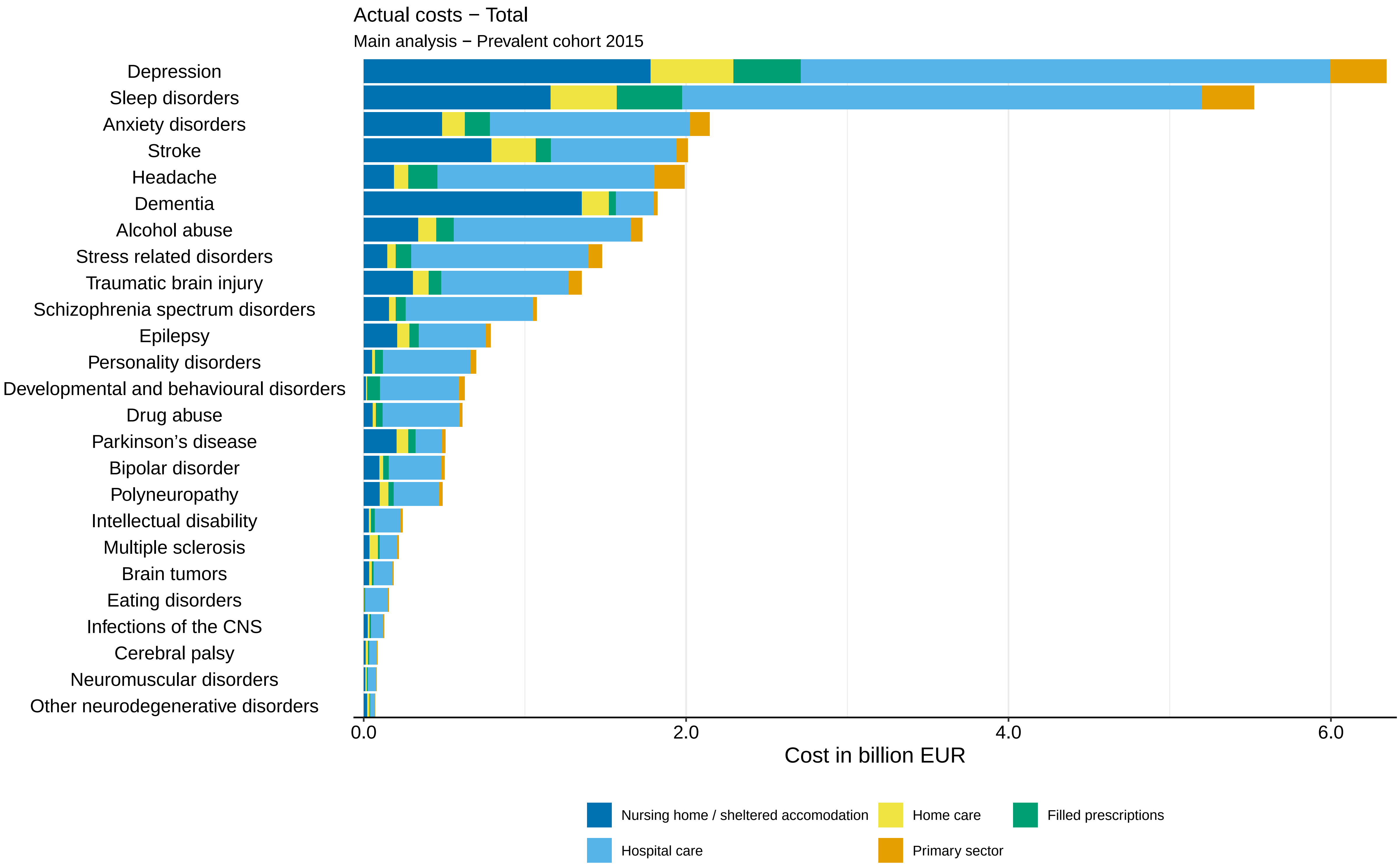

Actual costs − Total
Main analysis − Prevalent cohort 2015
Depression
Sleep disorders
Anxiety disorders
Stroke
Headache
Dementia
Alcohol abuse
Stress related disorders
Traumatic brain injury
Schizophrenia spectrum disorders
Epilepsy
Personality disorders
Developmental and behavioural disorders
Drug abuse
Parkinson’s disease
Bipolar disorder
Polyneuropathy
Intellectual disability
Multiple sclerosis
Brain tumors
Eating disorders
Infections of the CNS
Cerebral palsy
Neuromuscular disorders
Other neurodegenerative disorders
0.0
2.0
4.0
6.0
Cost in billion EUR
Nursing home / sheltered accomodation
Home care
Filled prescriptions
Hospital care
Primary sector

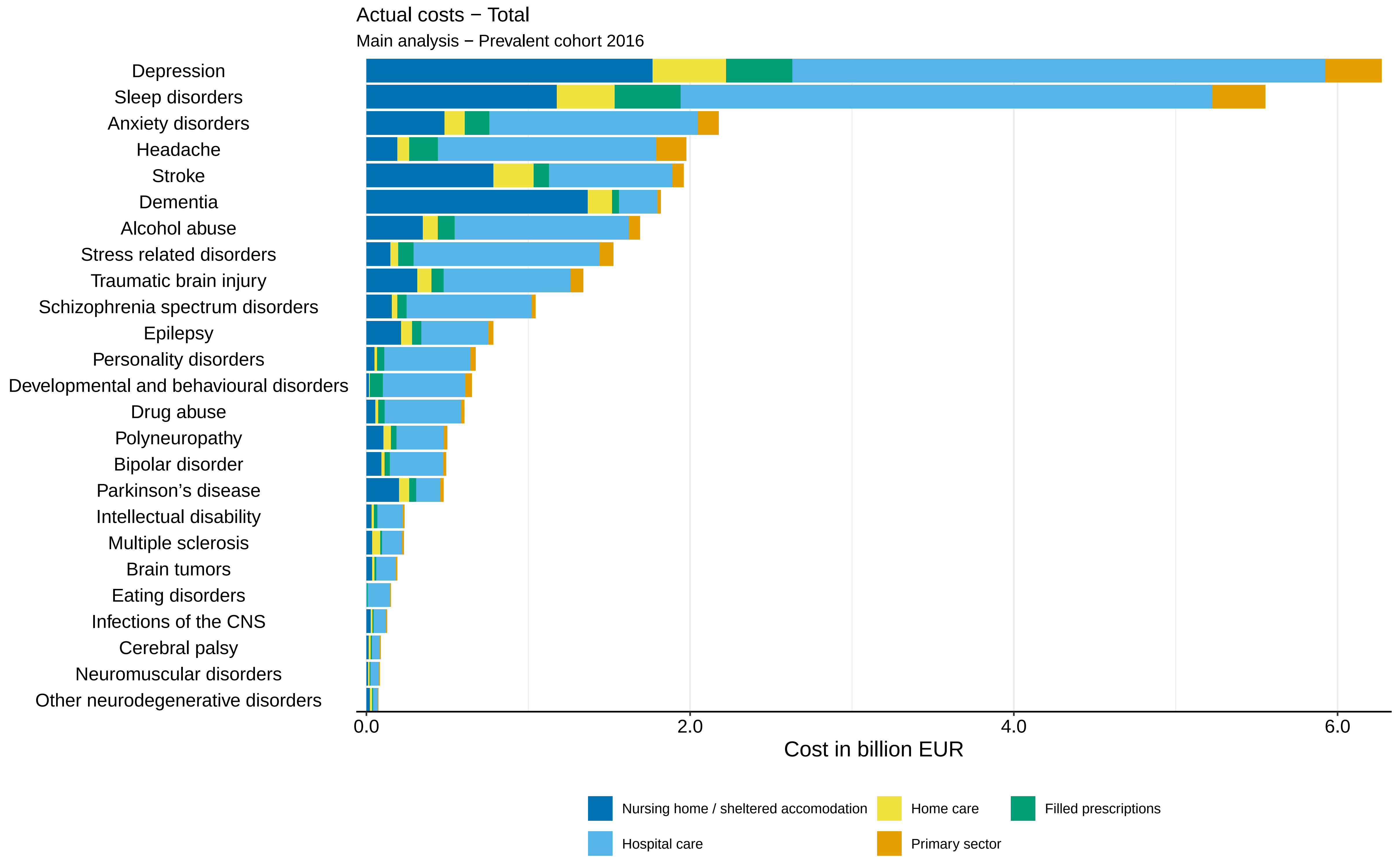
Actual costs − Total
Main analysis − Prevalent cohort 2016
Depression
Sleep disorders
Anxiety disorders
Headache
Stroke
Dementia
Alcohol abuse
Stress related disorders
Traumatic brain injury
Schizophrenia spectrum disorders
Epilepsy
Personality disorders
Developmental and behavioural disorders
Drug abuse
Polyneuropathy
Bipolar disorder
Parkinson’s disease
Intellectual disability
Multiple sclerosis
Brain tumors
Eating disorders
Infections of the CNS
Cerebral palsy
Neuromuscular disorders
Other neurodegenerative disorders
0.0
2.0
4.0
6.0
Cost in billion EUR
Nursing home / sheltered accomodation
Home care
Filled prescriptions
Hospital care
Primary sector

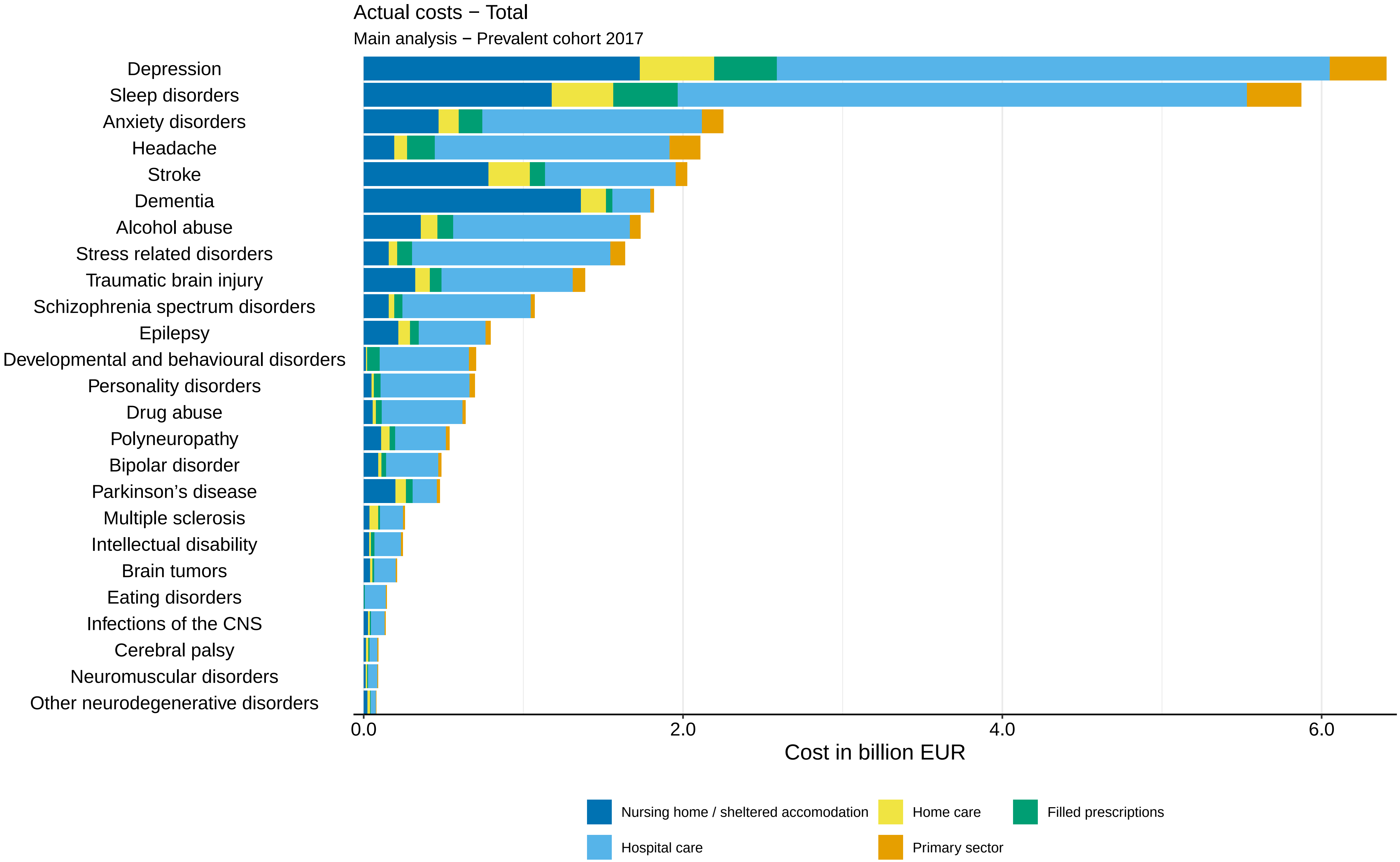

Actual costs − Total
Main analysis − Prevalent cohort 2017
Depression
Sleep disorders
Anxiety disorders
Headache
Stroke
Dementia
Alcohol abuse
Stress related disorders
Traumatic brain injury
Schizophrenia spectrum disorders
Epilepsy
Developmental and behavioural disorders
Personality disorders
Drug abuse
Polyneuropathy
Bipolar disorder
Parkinson’s disease
Multiple sclerosis
Intellectual disability
Brain tumors
Eating disorders
Infections of the CNS
Cerebral palsy
Neuromuscular disorders
Other neurodegenerative disorders
0.0
2.0
4.0
6.0
Cost in billion EUR
Nursing home / sheltered accomodation
Home care
Filled prescriptions
Hospital care
Primary sector

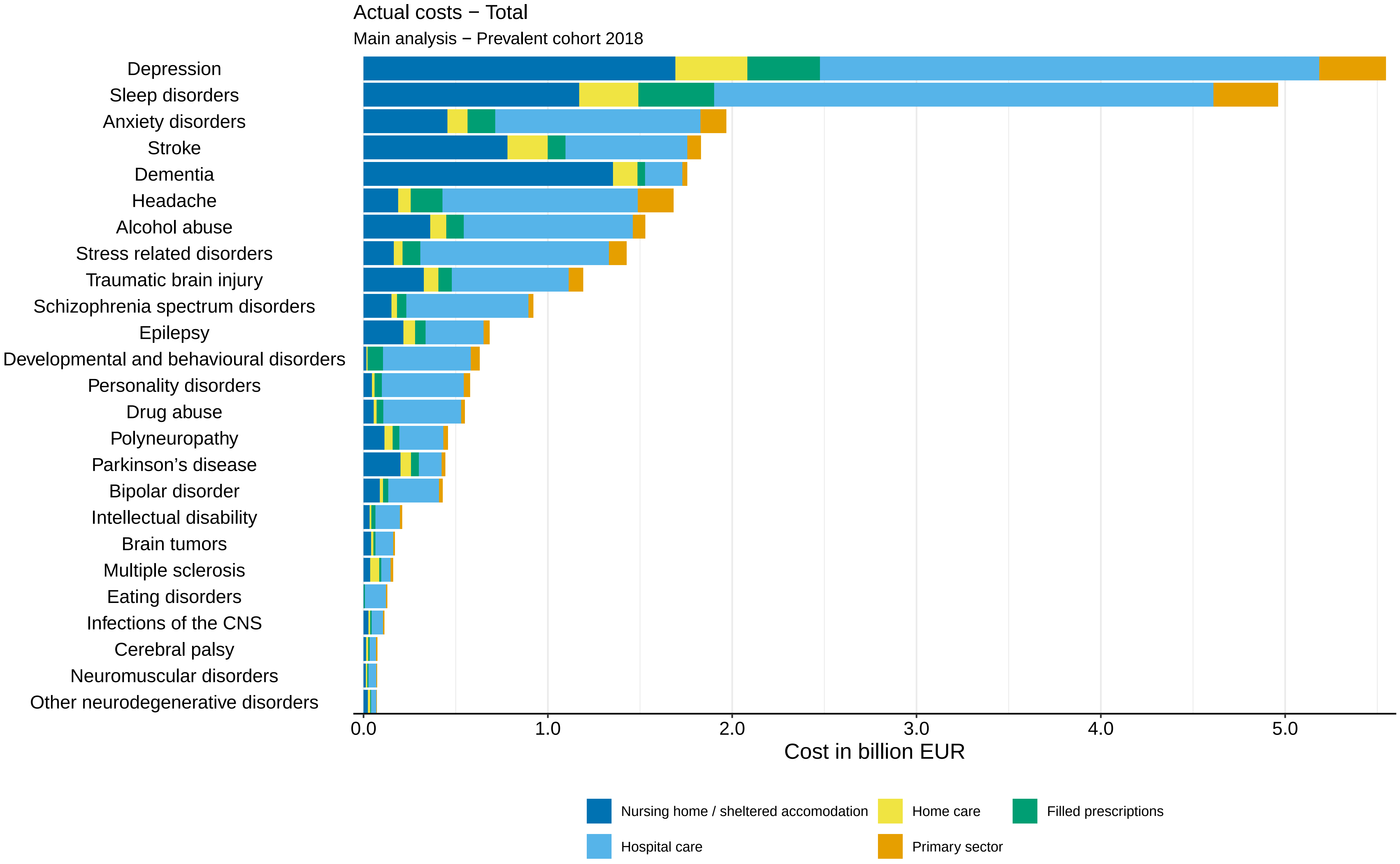

Actual costs − Total
Main analysis − Prevalent cohort 2018
Depression
Sleep disorders
Anxiety disorders
Stroke
Dementia
Headache
Alcohol abuse
Stress related disorders
Traumatic brain injury
Schizophrenia spectrum disorders
Epilepsy
Developmental and behavioural disorders
Personality disorders
Drug abuse
Polyneuropathy
Parkinson’s disease
Bipolar disorder
Intellectual disability
Brain tumors
Multiple sclerosis
Eating disorders
Infections of the CNS
Cerebral palsy
Neuromuscular disorders
Other neurodegenerative disorders
0.0
1.0
2.0
3.0
4.0
5.0
Cost in billion EUR
Nursing home / sheltered accomodation
Home care
Filled prescriptions
Hospital care
Primary sector

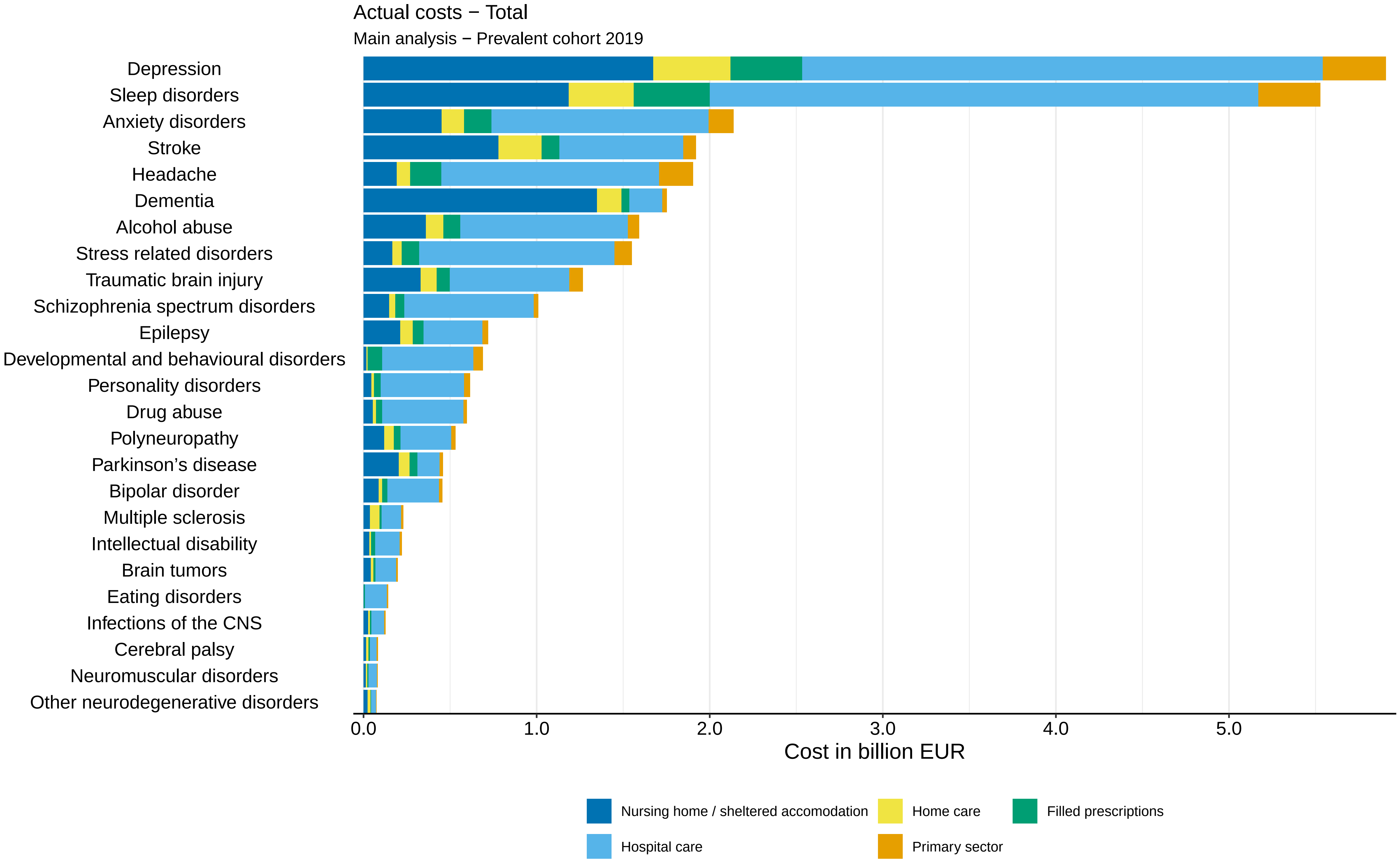

Actual costs − Total
Main analysis − Prevalent cohort 2019
Depression
Sleep disorders
Anxiety disorders
Stroke
Headache
Dementia
Alcohol abuse
Stress related disorders
Traumatic brain injury
Schizophrenia spectrum disorders
Epilepsy
Developmental and behavioural disorders
Personality disorders
Drug abuse
Polyneuropathy
Parkinson's disease
Bipolar disorder
Multiple sclerosis
Intellectual disability
Brain tumors
Eating disorders
Infections of the CNS
Cerebral palsy
Neuromuscular disorders
Other neurodegenerative disorders
0.0
1.0
2.0
3.0
4.0
5.0
Cost in billion EUR
Nursing home / sheltered accomodation
Home care
Filled prescriptions
Hospital care
Primary sector

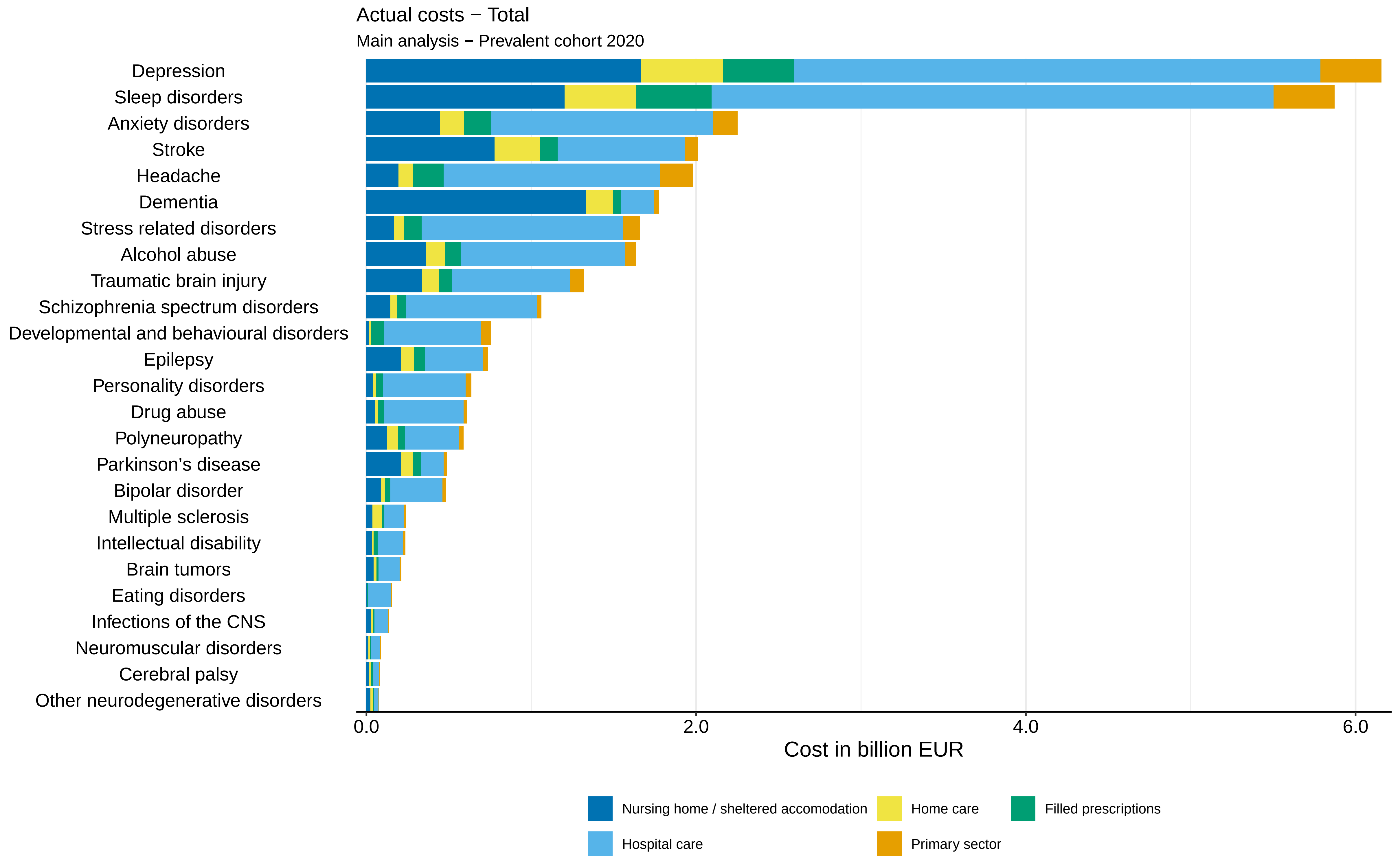

Actual costs − Total
Main analysis − Prevalent cohort 2020
Depression
Sleep disorders
Anxiety disorders
Stroke
Headache
Dementia
Stress related disorders
Alcohol abuse
Traumatic brain injury
Schizophrenia spectrum disorders
Developmental and behavioural disorders
Epilepsy
Personality disorders
Drug abuse
Polyneuropathy
Parkinson's disease
Bipolar disorder
Multiple sclerosis
Intellectual disability
Brain tumors
Eating disorders
Infections of the CNS
Neuromuscular disorders
Cerebral palsy
Other neurodegenerative disorders
0.0
2.0
4.0
6.0
Cost in billion EUR
Nursing home / sheltered accomodation
Home care
Filled prescriptions
Hospital care
Primary sector

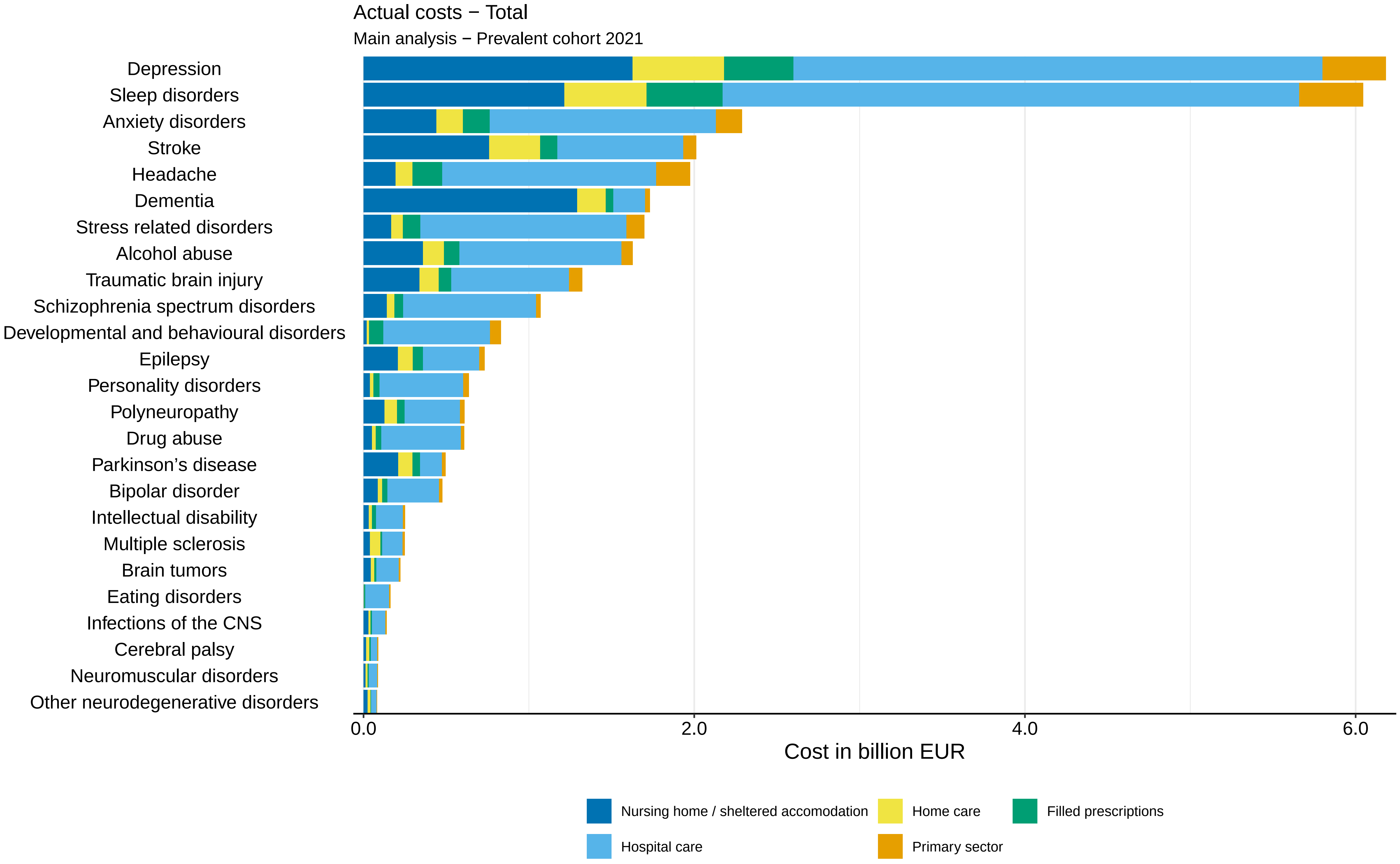
Actual costs − Total
Main analysis − Prevalent cohort 2021
Depression
Sleep disorders
Anxiety disorders
Stroke
Headache
Dementia
Stress related disorders
Alcohol abuse
Traumatic brain injury
Schizophrenia spectrum disorders
Developmental and behavioural disorders
Epilepsy
Personality disorders
Polyneuropathy
Drug abuse
Parkinson's disease
Bipolar disorder
Intellectual disability
Multiple sclerosis
Brain tumors
Eating disorders
Infections of the CNS
Cerebral palsy
Neuromuscular disorders
Other neurodegenerative disorders
0.0
2.0
4.0
6.0
Cost in billion EUR
Nursing home / sheltered accomodation
Home care
Filled prescriptions
Hospital care
Primary sector

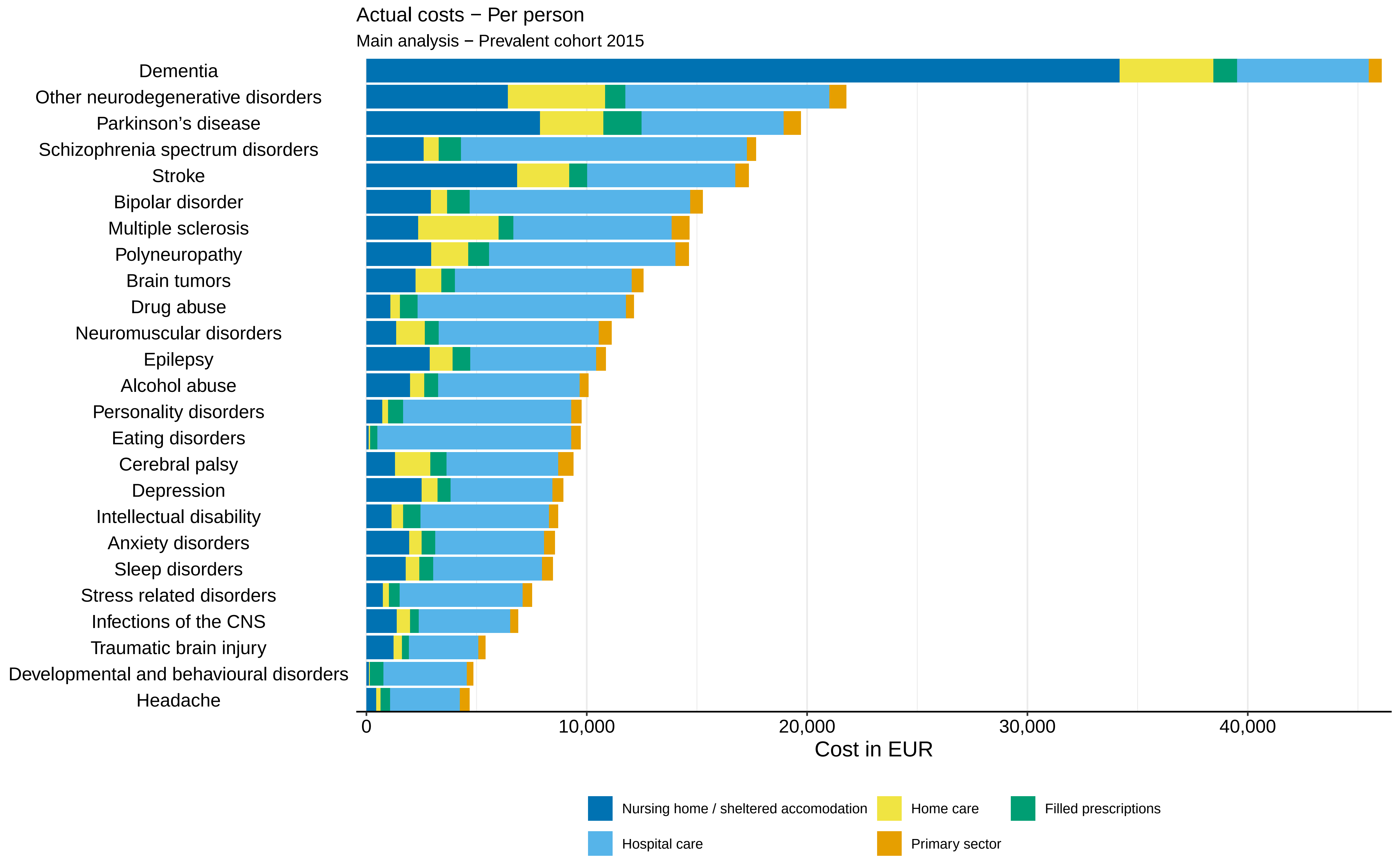

Actual costs − Per person
Main analysis − Prevalent cohort 2015
Dementia
Other neurodegenerative disorders
Parkinson's disease
Schizophrenia spectrum disorders
Stroke
Bipolar disorder
Multiple sclerosis
Polyneuropathy
Brain tumors
Drug abuse
Neuromuscular disorders
Epilepsy
Alcohol abuse
Personality disorders
Eating disorders
Cerebral palsy
Depression
Intellectual disability
Anxiety disorders
Sleep disorders
Stress related disorders
Infections of the CNS
Traumatic brain injury
Developmental and behavioural disorders
Headache
0
10,000
20,000
30,000
40,000
Cost in EUR
Nursing home / sheltered accomodation
Home care
Filled prescriptions
Hospital care
Primary sector

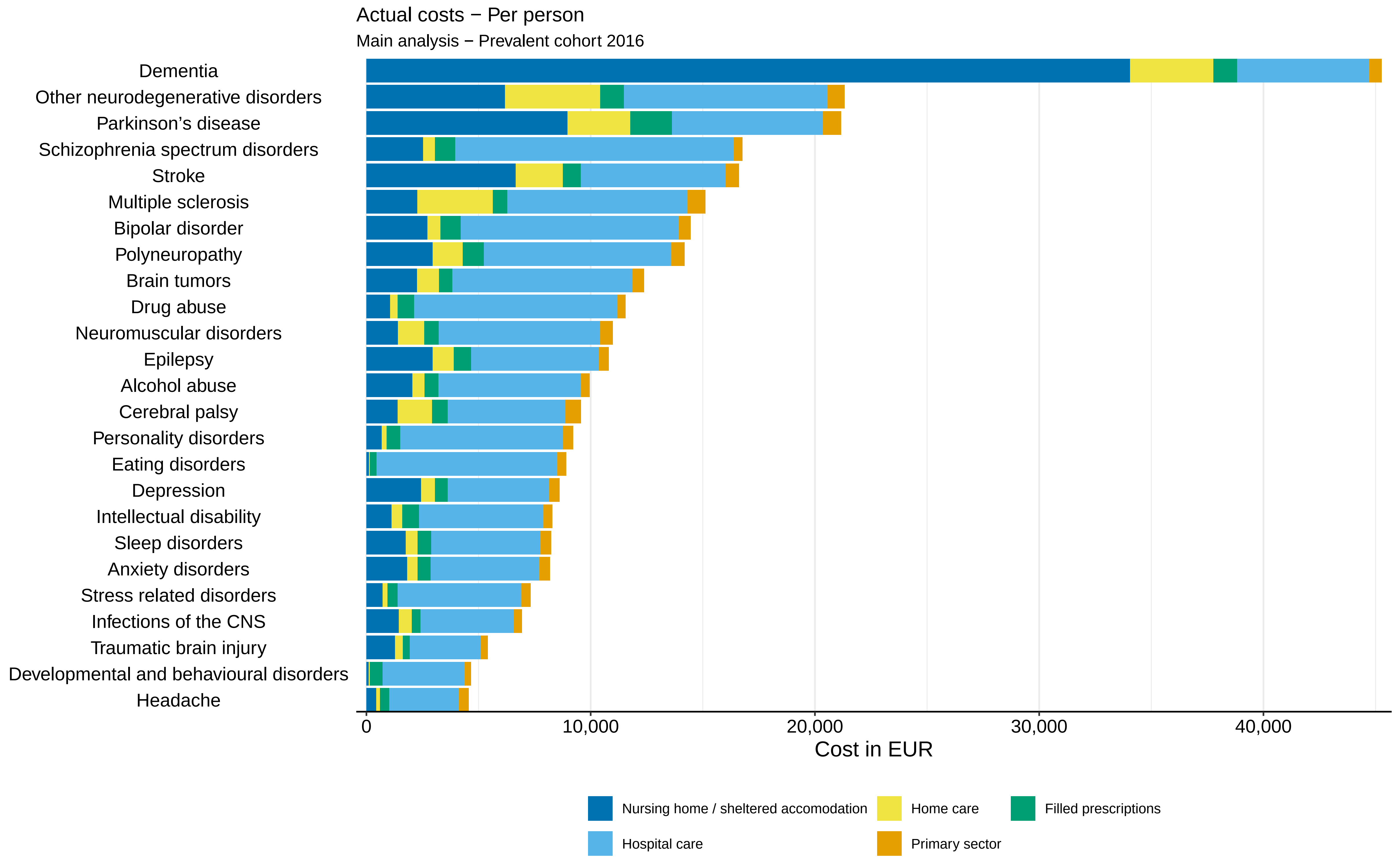
Actual costs − Per person
Main analysis − Prevalent cohort 2016
Dementia
Other neurodegenerative disorders
Parkinson’s disease
Schizophrenia spectrum disorders
Stroke
Multiple sclerosis
Bipolar disorder
Polyneuropathy
Brain tumors
Drug abuse
Neuromuscular disorders
Epilepsy
Alcohol abuse
Cerebral palsy
Personality disorders
Eating disorders
Depression
Intellectual disability
Sleep disorders
Anxiety disorders
Stress related disorders
Infections of the CNS
Traumatic brain injury
Developmental and behavioural disorders
Headache
0
10,000
20,000
30,000
40,000
Cost in EUR
Nursing home / sheltered accomodation
Home care
Filled prescriptions
Hospital care
Primary sector

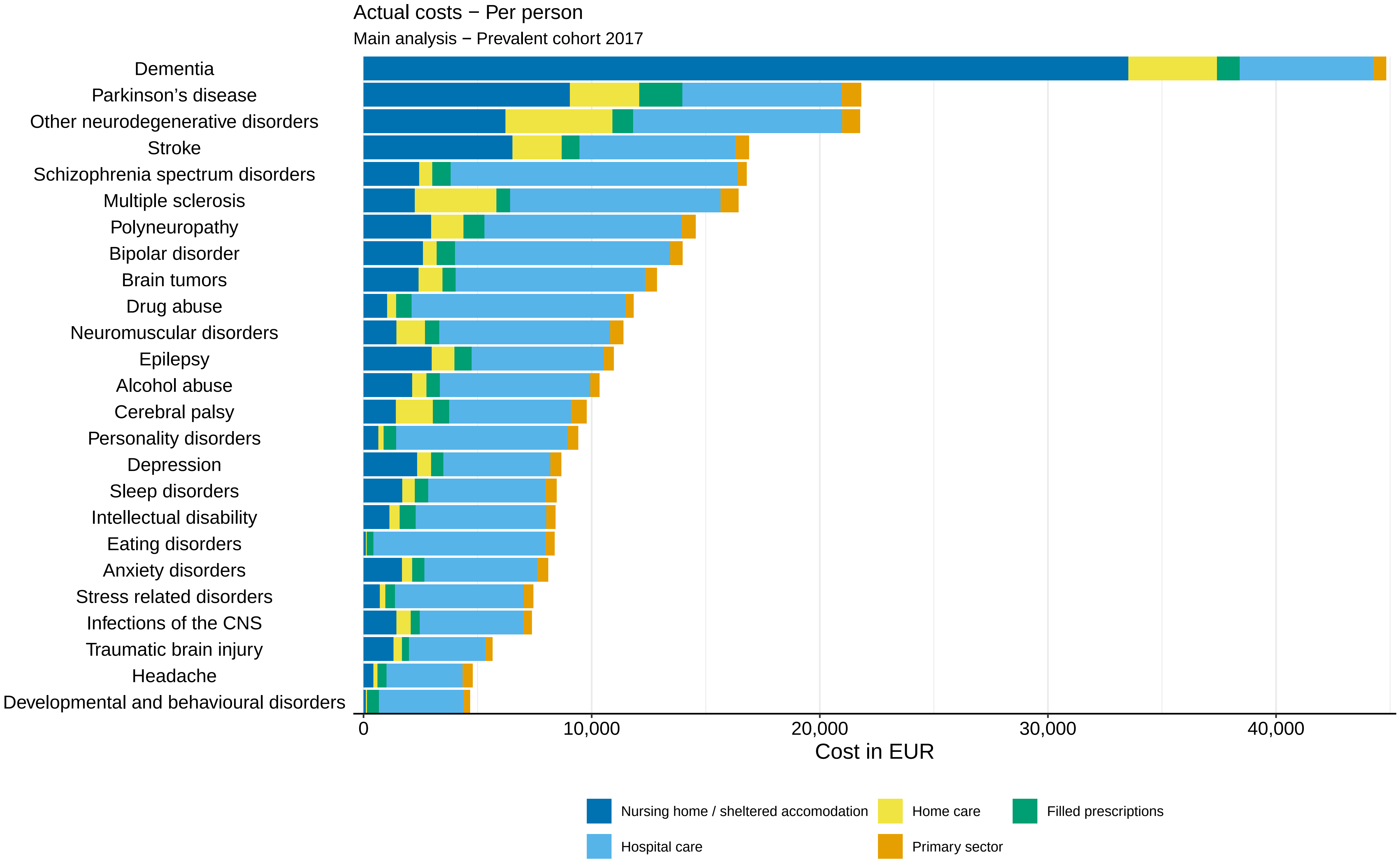

Actual costs − Per person
Main analysis − Prevalent cohort 2017
Dementia
Parkinson's disease
Other neurodegenerative disorders
Stroke
Schizophrenia spectrum disorders
Multiple sclerosis
Polyneuropathy
Bipolar disorder
Brain tumors
Drug abuse
Neuromuscular disorders
Epilepsy
Alcohol abuse
Cerebral palsy
Personality disorders
Depression
Sleep disorders
Intellectual disability
Eating disorders
Anxiety disorders
Stress related disorders
Infections of the CNS
Traumatic brain injury
Headache
Developmental and behavioural disorders
0
10,000
20,000
30,000
40,000
Cost in EUR
Nursing home / sheltered accomodation
Home care
Filled prescriptions
Hospital care
Primary sector

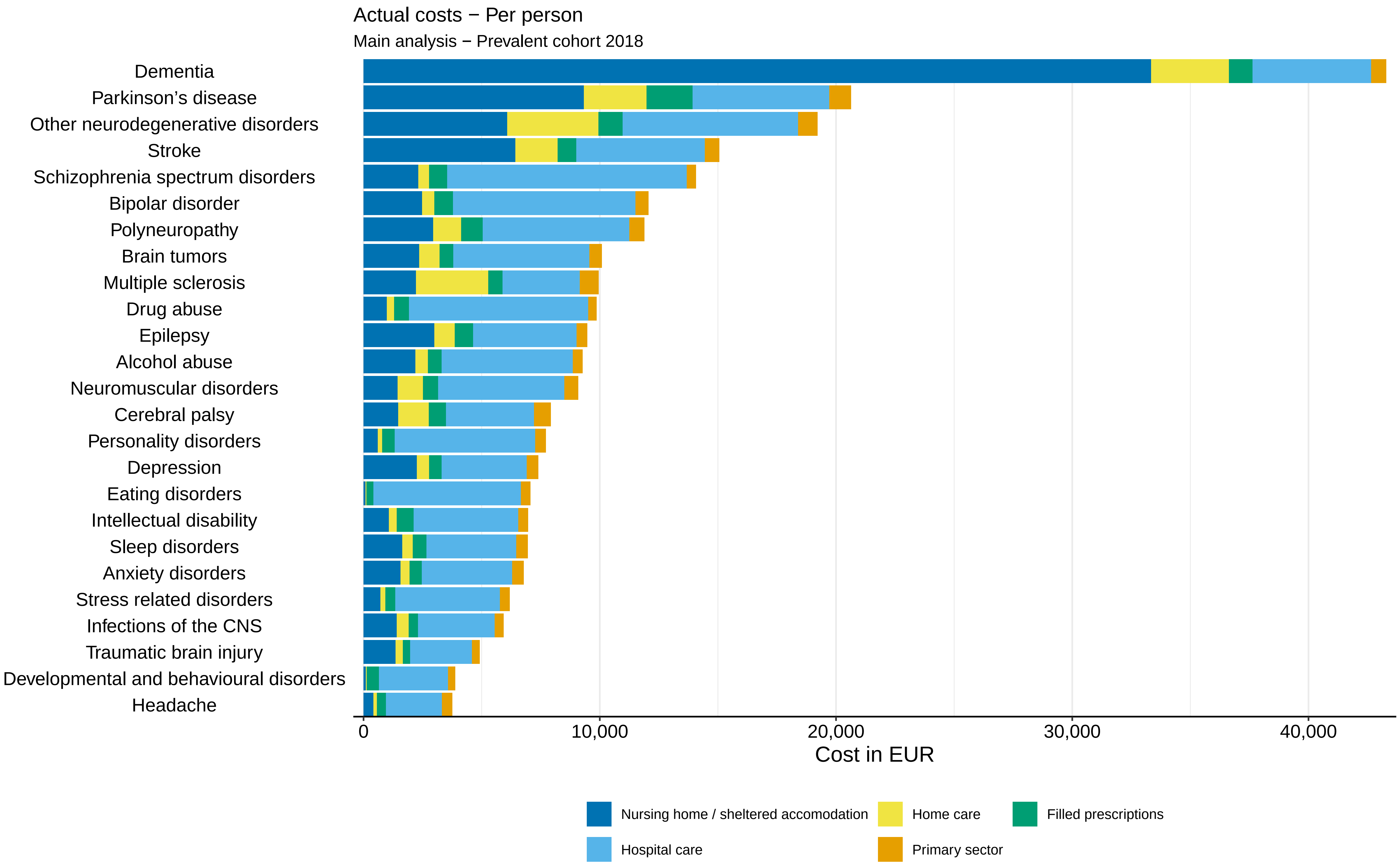

Actual costs − Per person
Main analysis − Prevalent cohort 2018
Dementia
Parkinson’s disease
Other neurodegenerative disorders
Stroke
Schizophrenia spectrum disorders
Bipolar disorder
Polyneuropathy
Brain tumors
Multiple sclerosis
Drug abuse
Epilepsy
Alcohol abuse
Neuromuscular disorders
Cerebral palsy
Personality disorders
Depression
Eating disorders
Intellectual disability
Sleep disorders
Anxiety disorders
Stress related disorders
Infections of the CNS
Traumatic brain injury
Developmental and behavioural disorders
Headache
0
10,000
20,000
30,000
40,000
Cost in EUR
Nursing home / sheltered accomodation
Home care
Filled prescriptions
Hospital care
Primary sector

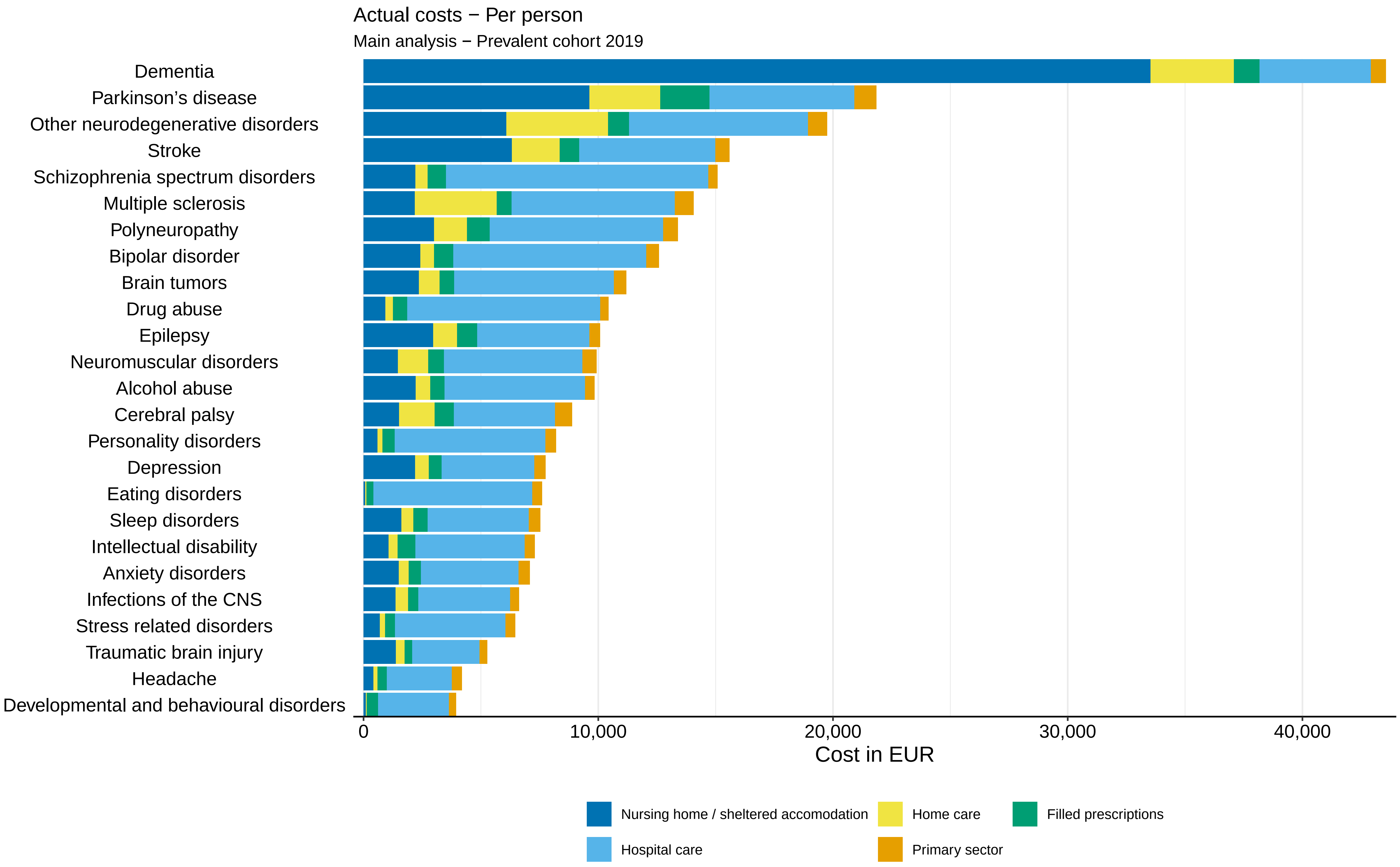

Actual costs − Per person
Main analysis − Prevalent cohort 2019
Dementia
Parkinson’s disease
Other neurodegenerative disorders
Stroke
Schizophrenia spectrum disorders
Multiple sclerosis
Polyneuropathy
Bipolar disorder
Brain tumors
Drug abuse
Epilepsy
Neuromuscular disorders
Alcohol abuse
Cerebral palsy
Personality disorders
Depression
Eating disorders
Sleep disorders
Intellectual disability
Anxiety disorders
Infections of the CNS
Stress related disorders
Traumatic brain injury
Headache
Developmental and behavioural disorders
0
10,000
20,000
30,000
40,000
Cost in EUR
Nursing home / sheltered accomodation
Home care
Filled prescriptions
Hospital care
Primary sector

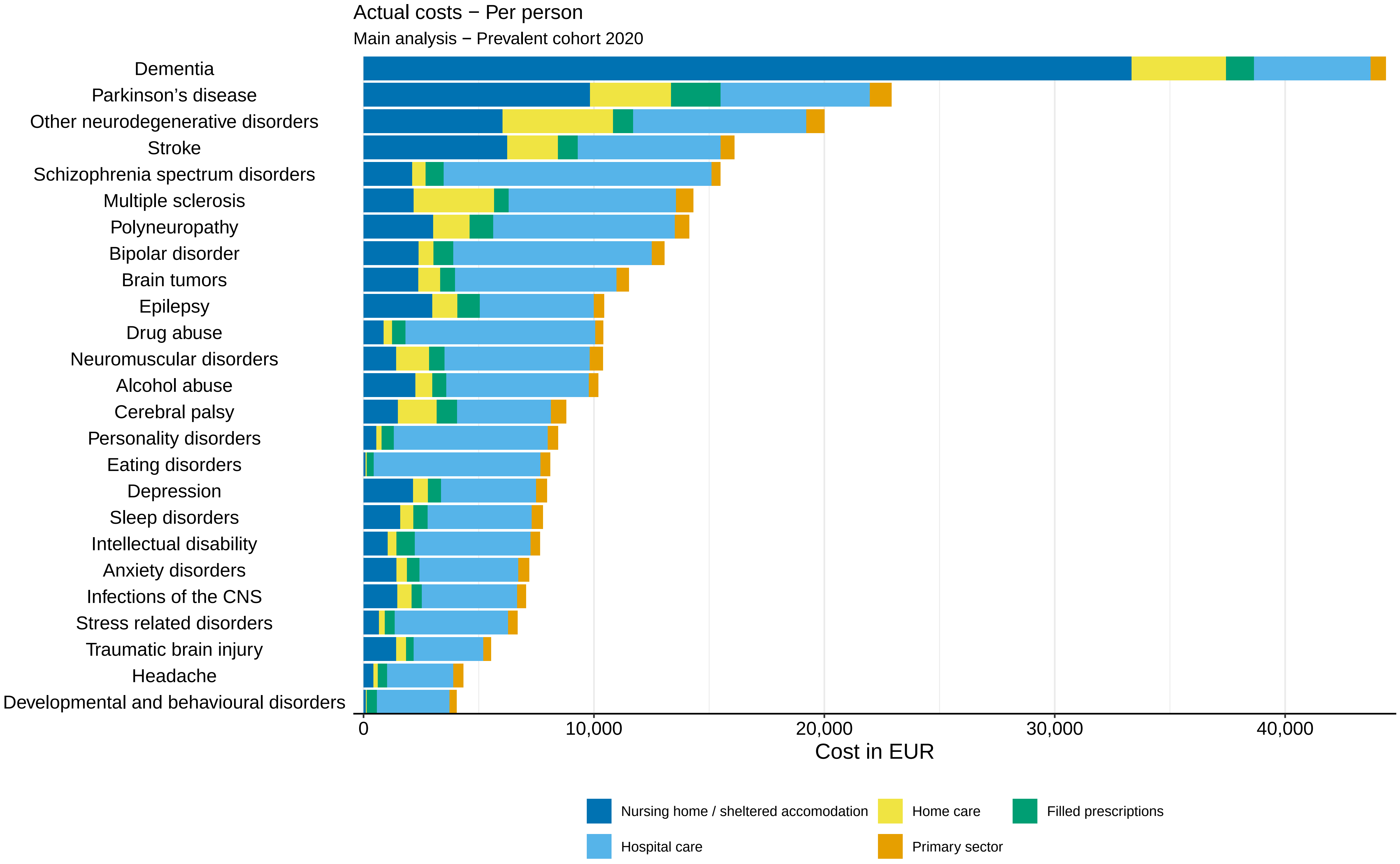

Actual costs − Per person
Main analysis − Prevalent cohort 2020
Dementia
Parkinson’s disease
Other neurodegenerative disorders
Stroke
Schizophrenia spectrum disorders
Multiple sclerosis
Polyneuropathy
Bipolar disorder
Brain tumors
Epilepsy
Drug abuse
Neuromuscular disorders
Alcohol abuse
Cerebral palsy
Personality disorders
Eating disorders
Depression
Sleep disorders
Intellectual disability
Anxiety disorders
Infections of the CNS
Stress related disorders
Traumatic brain injury
Headache
Developmental and behavioural disorders
0
10,000
20,000
30,000
40,000
Cost in EUR
Nursing home / sheltered accomodation
Home care
Filled prescriptions
Hospital care
Primary sector

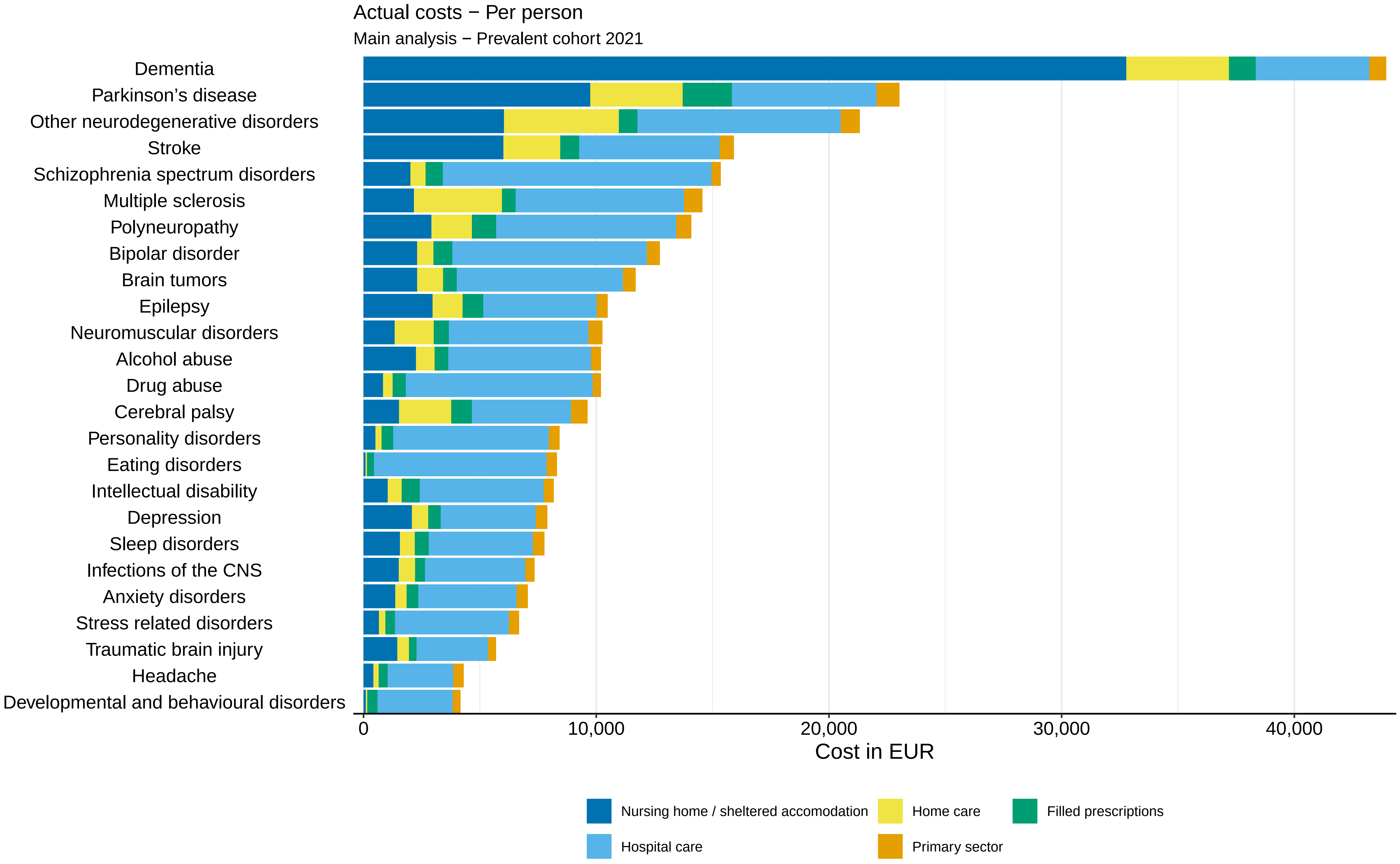

Actual costs − Per person
Main analysis − Prevalent cohort 2021
Dementia
Parkinson’s disease
Other neurodegenerative disorders
Stroke
Schizophrenia spectrum disorders
Multiple sclerosis
Polyneuropathy
Bipolar disorder
Brain tumors
Epilepsy
Neuromuscular disorders
Alcohol abuse
Drug abuse
Cerebral palsy
Personality disorders
Eating disorders
Intellectual disability
Depression
Sleep disorders
Infections of the CNS
Anxiety disorders
Stress related disorders
Traumatic brain injury
Headache
Developmental and behavioural disorders
0
10,000
20,000
30,000
40,000
Cost in EUR
Nursing home / sheltered accomodation
Home care
Filled prescriptions
Hospital care
Primary sector

Attributable costs − Total

Main analysis − Prevalent cohort 2015

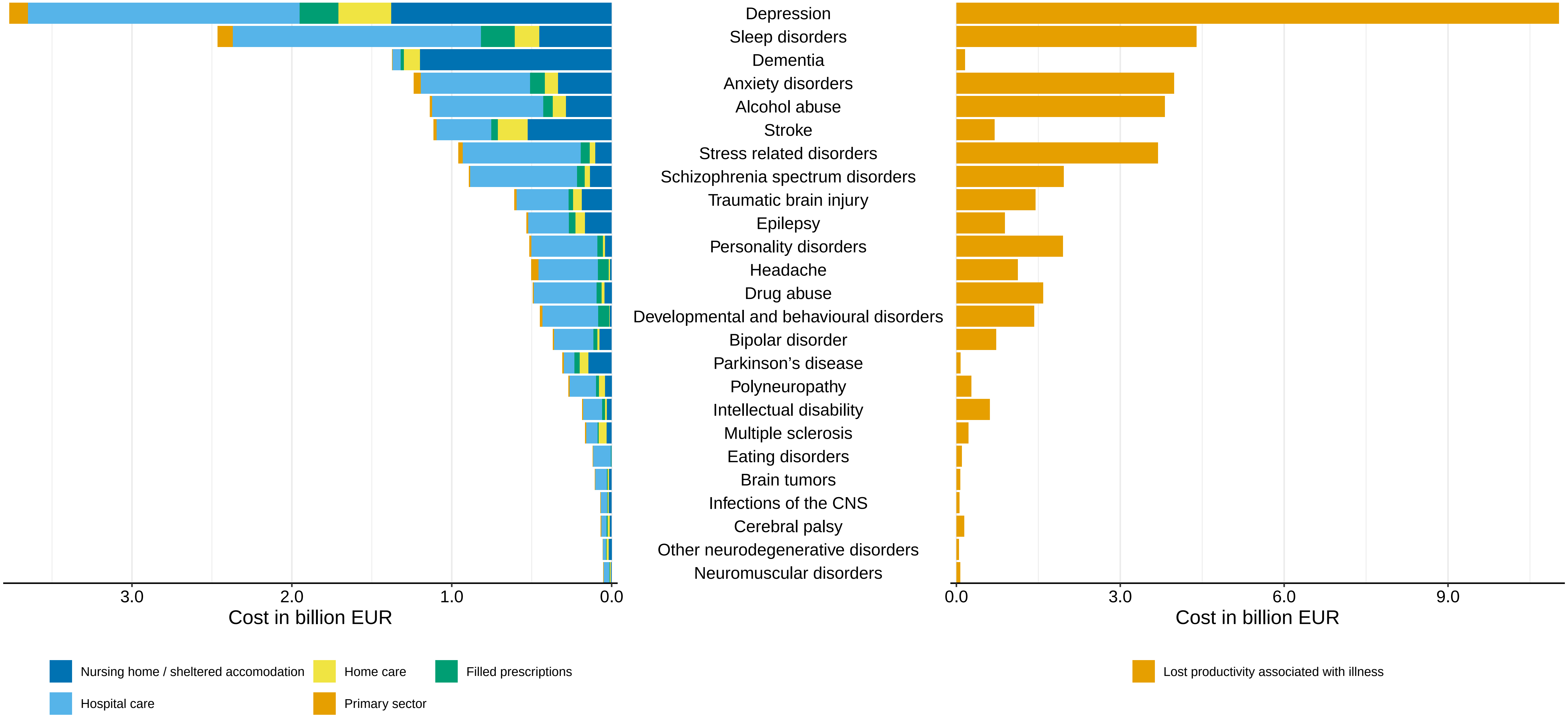

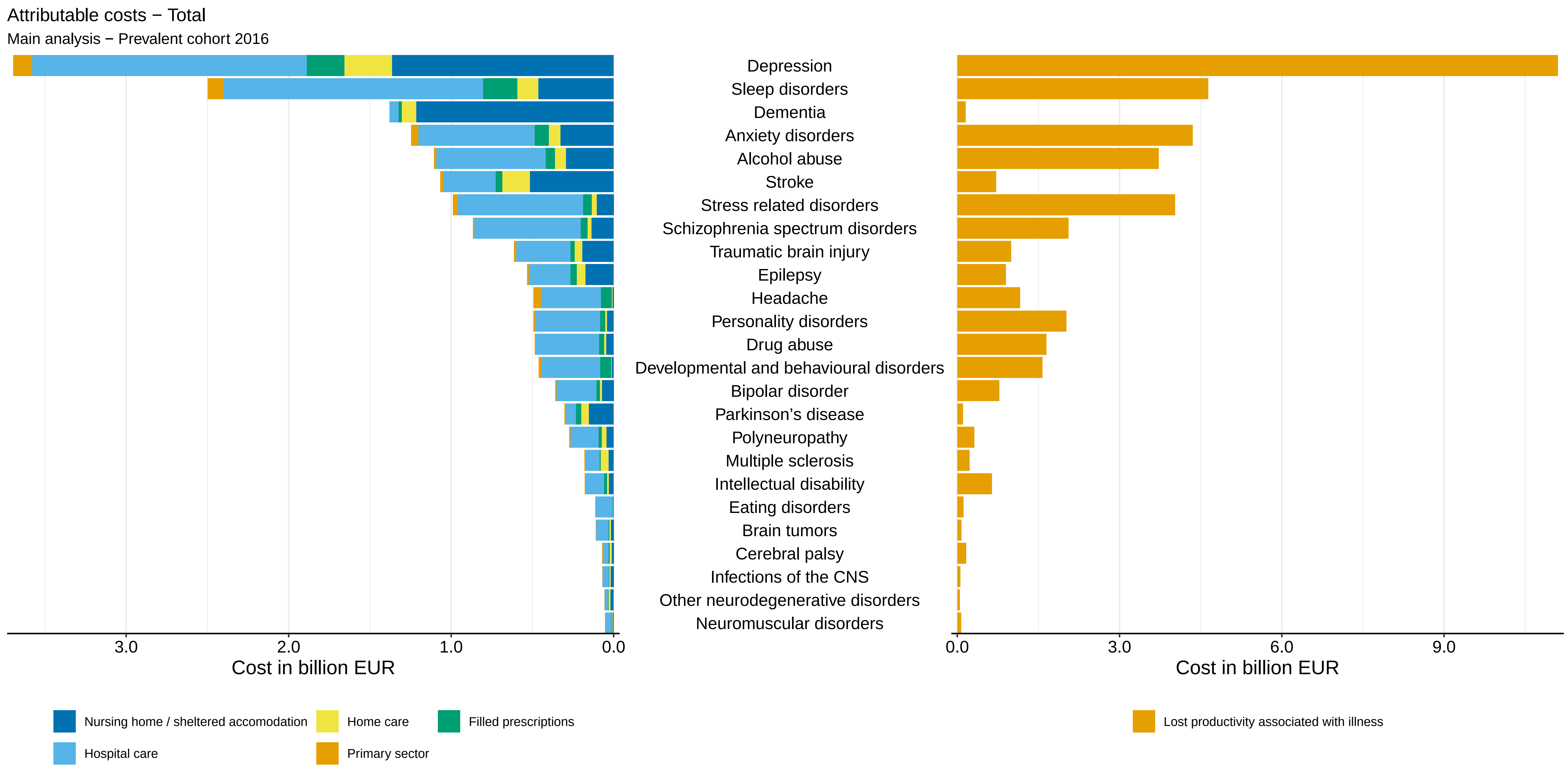

Attributable costs − Total
Main analysis − Prevalent cohort 2016
Depression
Sleep disorders
Dementia
Anxiety disorders
Alcohol abuse
Stroke
Stress related disorders
Schizophrenia spectrum disorders
Traumatic brain injury
Epilepsy
Headache
Personality disorders
Drug abuse
Developmental and behavioural disorders
Bipolar disorder
Parkinson’s disease
Polyneuropathy
Multiple sclerosis
Intellectual disability
Eating disorders
Brain tumors
Cerebral palsy
Infections of the CNS
Other neurodegenerative disorders
Neuromuscular disorders
3.0
2.0
1.0
0.0
Cost in billion EUR
0.0
3.0
6.0
9.0
Cost in billion EUR
Nursing home / sheltered accomodation
Home care
Filled prescriptions
Hospital care
Primary sector
Lost productivity associated with illness

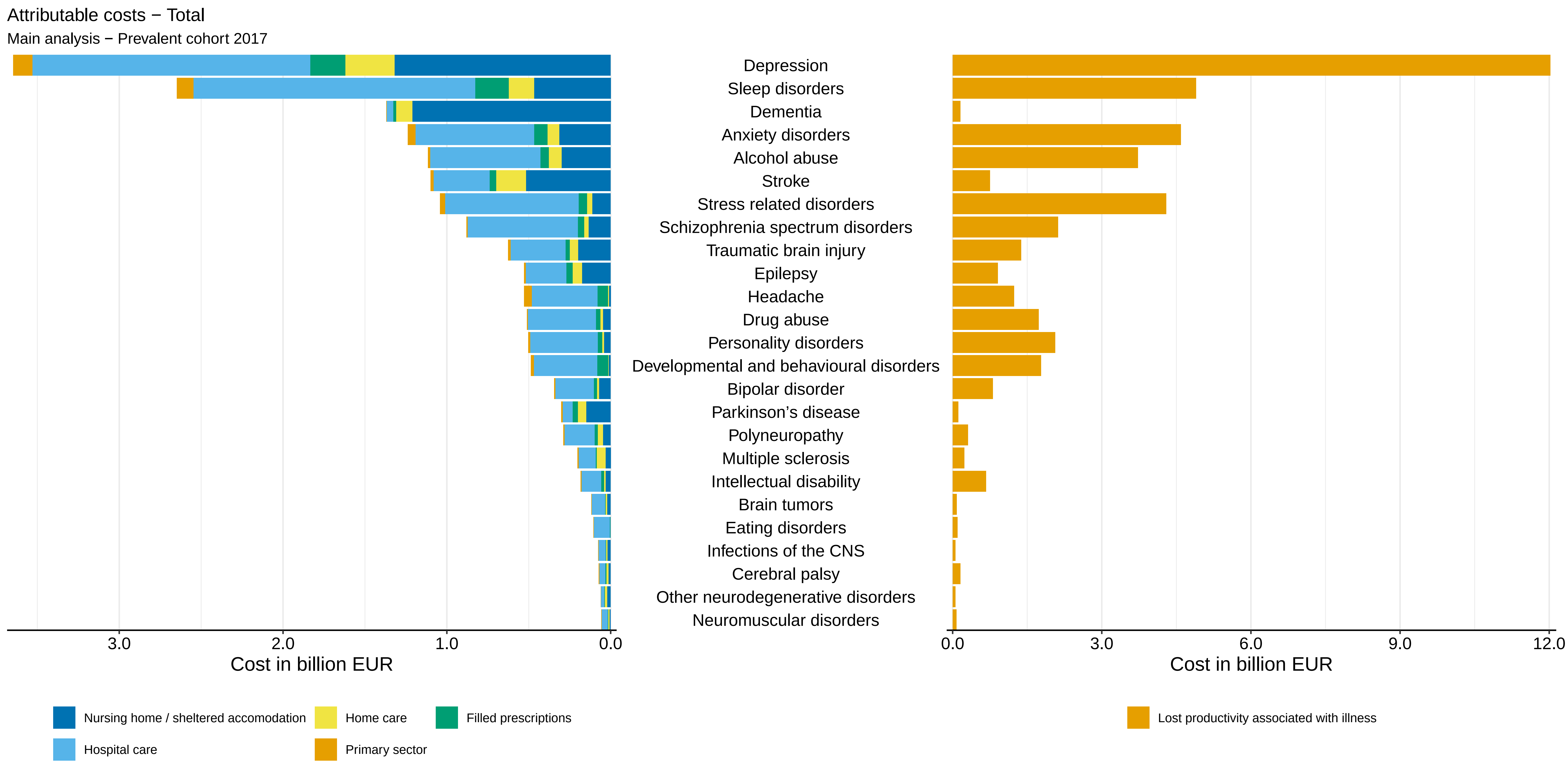
Attributable costs − Total
Main analysis − Prevalent cohort 2017
Depression
Sleep disorders
Dementia
Anxiety disorders
Alcohol abuse
Stroke
Stress related disorders
Schizophrenia spectrum disorders
Traumatic brain injury
Epilepsy
Headache
Drug abuse
Personality disorders
Developmental and behavioural disorders
Bipolar disorder
Parkinson’s disease
Polyneuropathy
Multiple sclerosis
Intellectual disability
Brain tumors
Eating disorders
Infections of the CNS
Cerebral palsy
Other neurodegenerative disorders
Neuromuscular disorders
3.0
2.0
1.0
0.0
Cost in billion EUR
0.0
3.0
6.0
9.0
12.0
Cost in billion EUR
Nursing home / sheltered accomodation
Home care
Filled prescriptions
Hospital care
Primary sector
Lost productivity associated with illness

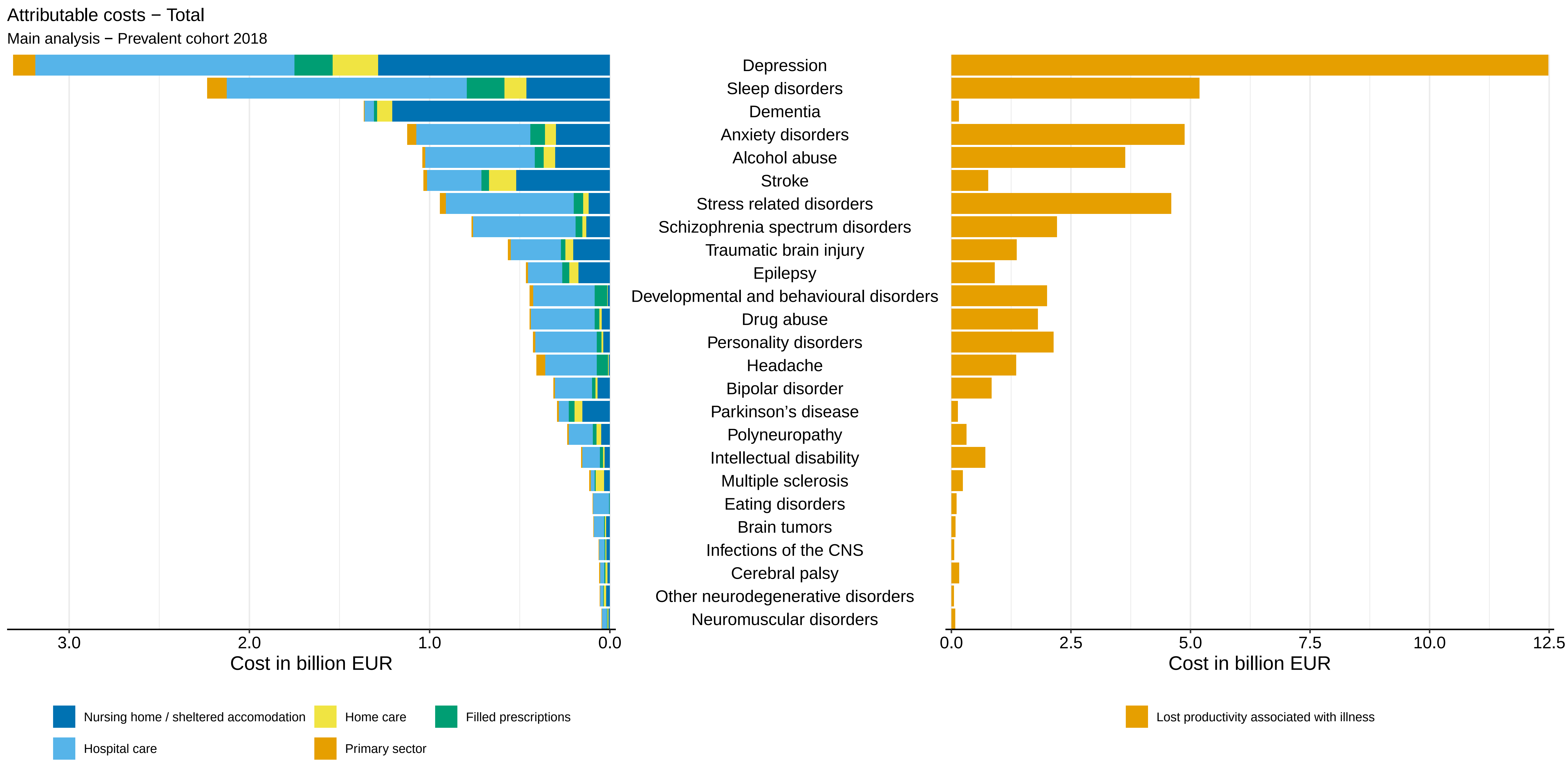

Attributable costs − Total
Main analysis − Prevalent cohort 2018
Depression
Sleep disorders
Dementia
Anxiety disorders
Alcohol abuse
Stroke
Stress related disorders
Schizophrenia spectrum disorders
Traumatic brain injury
Epilepsy
Developmental and behavioural disorders
Drug abuse
Personality disorders
Headache
Bipolar disorder
Parkinson’s disease
Polyneuropathy
Intellectual disability
Multiple sclerosis
Eating disorders
Brain tumors
Infections of the CNS
Cerebral palsy
Other neurodegenerative disorders
Neuromuscular disorders
3.0
2.0
1.0
0.0
Cost in billion EUR
0.0
2.5
5.0
7.5
10.0
12.5
Cost in billion EUR
Nursing home / sheltered accomodation
Home care
Filled prescriptions
Hospital care
Primary sector
Lost productivity associated with illness

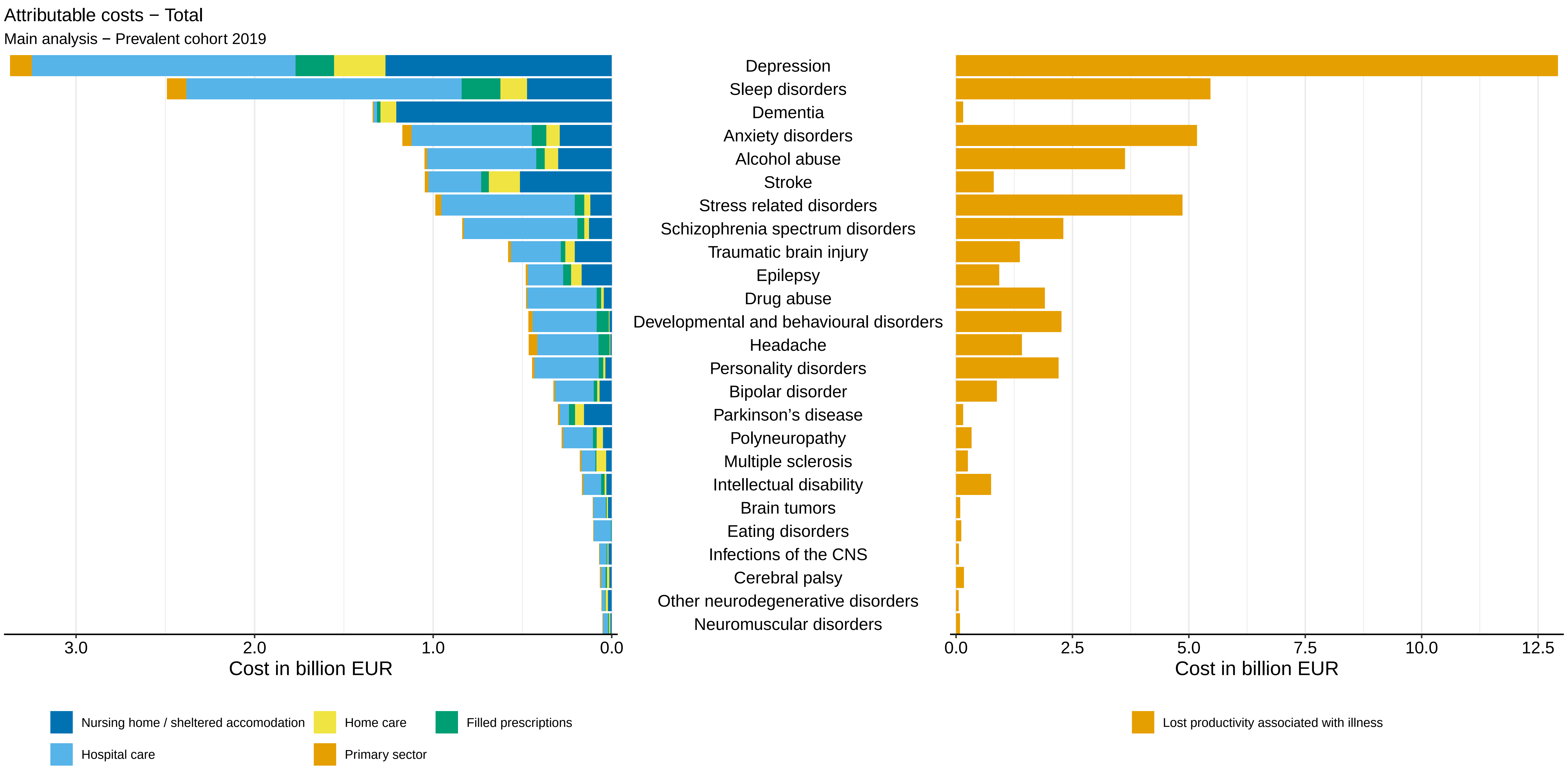

Attributable costs − Total
Main analysis − Prevalent cohort 2019
Depression
Sleep disorders
Dementia
Anxiety disorders
Alcohol abuse
Stroke
Stress related disorders
Schizophrenia spectrum disorders
Traumatic brain injury
Epilepsy
Drug abuse
Developmental and behavioural disorders
Headache
Personality disorders
Bipolar disorder
Parkinson’s disease
Polyneuropathy
Multiple sclerosis
Intellectual disability
Brain tumors
Eating disorders
Infections of the CNS
Cerebral palsy
Other neurodegenerative disorders
Neuromuscular disorders
3.0
2.0
1.0
0.0
Cost in billion EUR
0.0
2.5
5.0
7.5
10.0
12.5
Cost in billion EUR
Nursing home / sheltered accomodation
Home care
Filled prescriptions
Hospital care
Primary sector
Lost productivity associated with illness

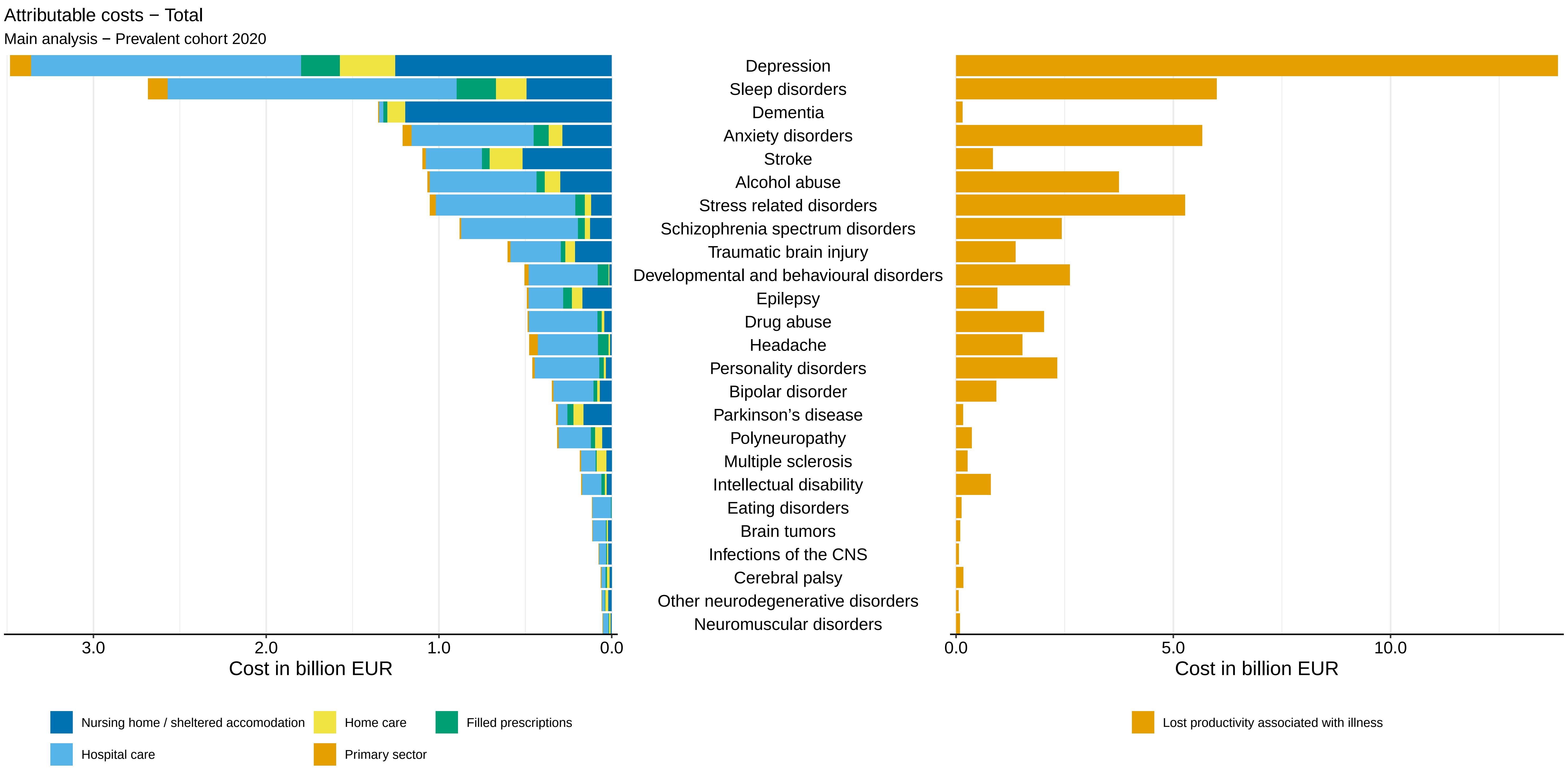
Attributable costs − Total
Main analysis − Prevalent cohort 2020
Depression
Sleep disorders
Dementia
Anxiety disorders
Stroke
Alcohol abuse
Stress related disorders
Schizophrenia spectrum disorders
Traumatic brain injury
Developmental and behavioural disorders
Epilepsy
Drug abuse
Headache
Personality disorders
Bipolar disorder
Parkinson’s disease
Polyneuropathy
Multiple sclerosis
Intellectual disability
Eating disorders
Brain tumors
Infections of the CNS
Cerebral palsy
Other neurodegenerative disorders
Neuromuscular disorders
3.0
2.0
1.0
0.0
Cost in billion EUR
0.0
5.0
10.0
Cost in billion EUR
Nursing home / sheltered accomodation
Home care
Filled prescriptions
Hospital care
Primary sector
Lost productivity associated with illness

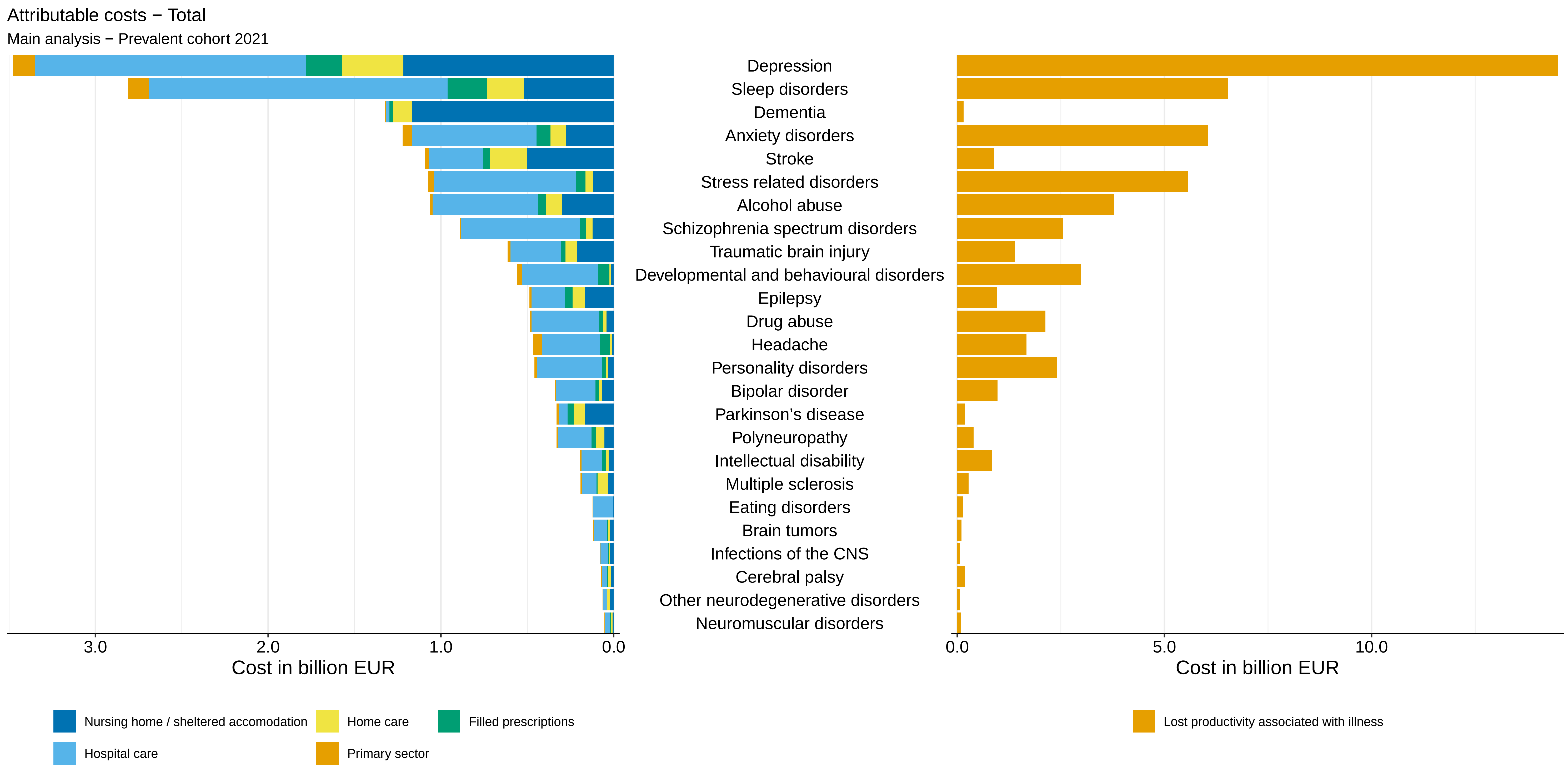

Attributable costs − Total
Main analysis − Prevalent cohort 2021
Depression
Sleep disorders
Dementia
Anxiety disorders
Stroke
Stress related disorders
Alcohol abuse
Schizophrenia spectrum disorders
Traumatic brain injury
Developmental and behavioural disorders
Epilepsy
Drug abuse
Headache
Personality disorders
Bipolar disorder
Parkinson’s disease
Polyneuropathy
Intellectual disability
Multiple sclerosis
Eating disorders
Brain tumors
Infections of the CNS
Cerebral palsy
Other neurodegenerative disorders
Neuromuscular disorders
3.0
2.0
1.0
0.0
Cost in billion EUR
0.0
5.0
10.0
Cost in billion EUR
Nursing home / sheltered accomodation
Home care
Filled prescriptions
Hospital care
Primary sector
Lost productivity associated with illness

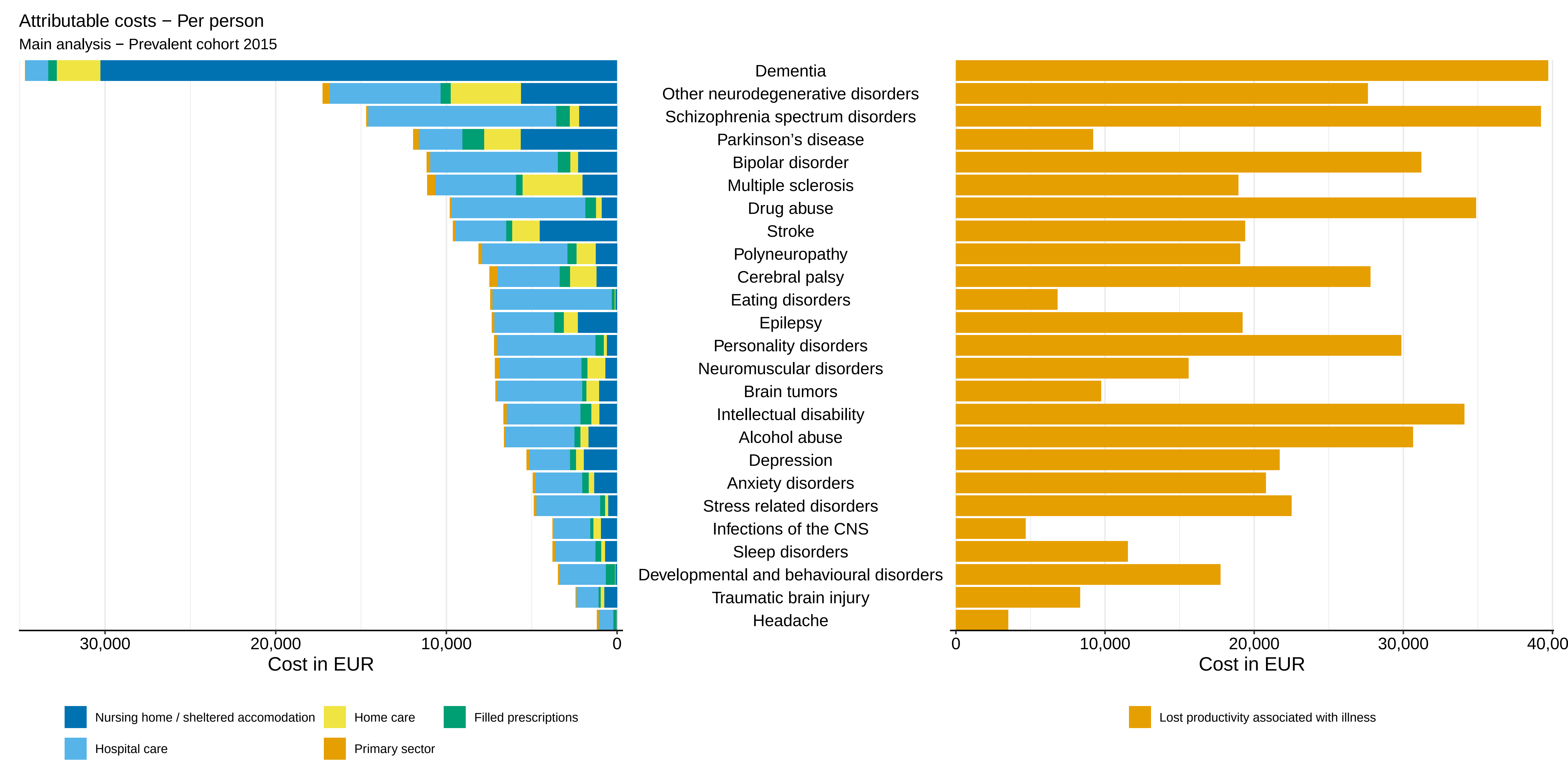

Attributable costs − Per person
Main analysis − Prevalent cohort 2015
Dementia
Other neurodegenerative disorders
Schizophrenia spectrum disorders
Parkinson’s disease
Bipolar disorder
Multiple sclerosis
Drug abuse
Stroke
Polyneuropathy
Cerebral palsy
Eating disorders
Epilepsy
Personality disorders
Neuromuscular disorders
Brain tumors
Intellectual disability
Alcohol abuse
Depression
Anxiety disorders
Stress related disorders
Infections of the CNS
Sleep disorders
Developmental and behavioural disorders
Traumatic brain injury
Headache
30,000
20,000
10,000
0
Cost in EUR
0
10,000
20,000
30,000
40,00
Cost in EUR
Nursing home / sheltered accomodation
Home care
Filled prescriptions
Hospital care
Primary sector
Lost productivity associated with illness

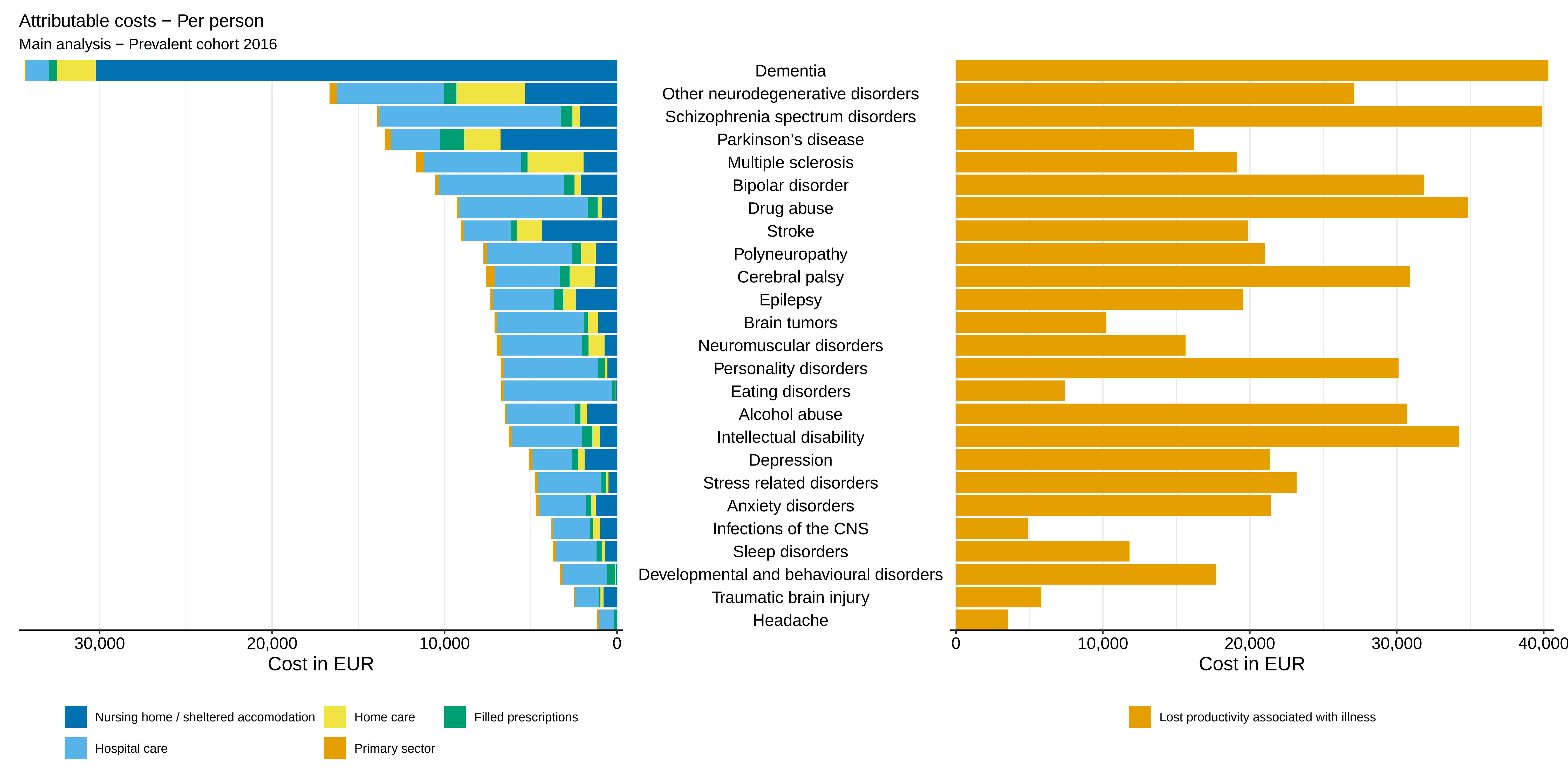
Attributable costs − Per person
Main analysis − Prevalent cohort 2016
Dementia
Other neurodegenerative disorders
Schizophrenia spectrum disorders
Parkinson’s disease
Multiple sclerosis
Bipolar disorder
Drug abuse
Stroke
Polyneuropathy
Cerebral palsy
Epilepsy
Brain tumors
Neuromuscular disorders
Personality disorders
Eating disorders
Alcohol abuse
Intellectual disability
Depression
Stress related disorders
Anxiety disorders
Infections of the CNS
Sleep disorders
Developmental and behavioural disorders
Traumatic brain injury
Headache
30,000
20,000
10,000
0
Cost in EUR
0
10,000
20,000
30,000
40,000
Cost in EUR
Nursing home / sheltered accomodation
Home care
Filled prescriptions
Hospital care
Primary sector
Lost productivity associated with illness

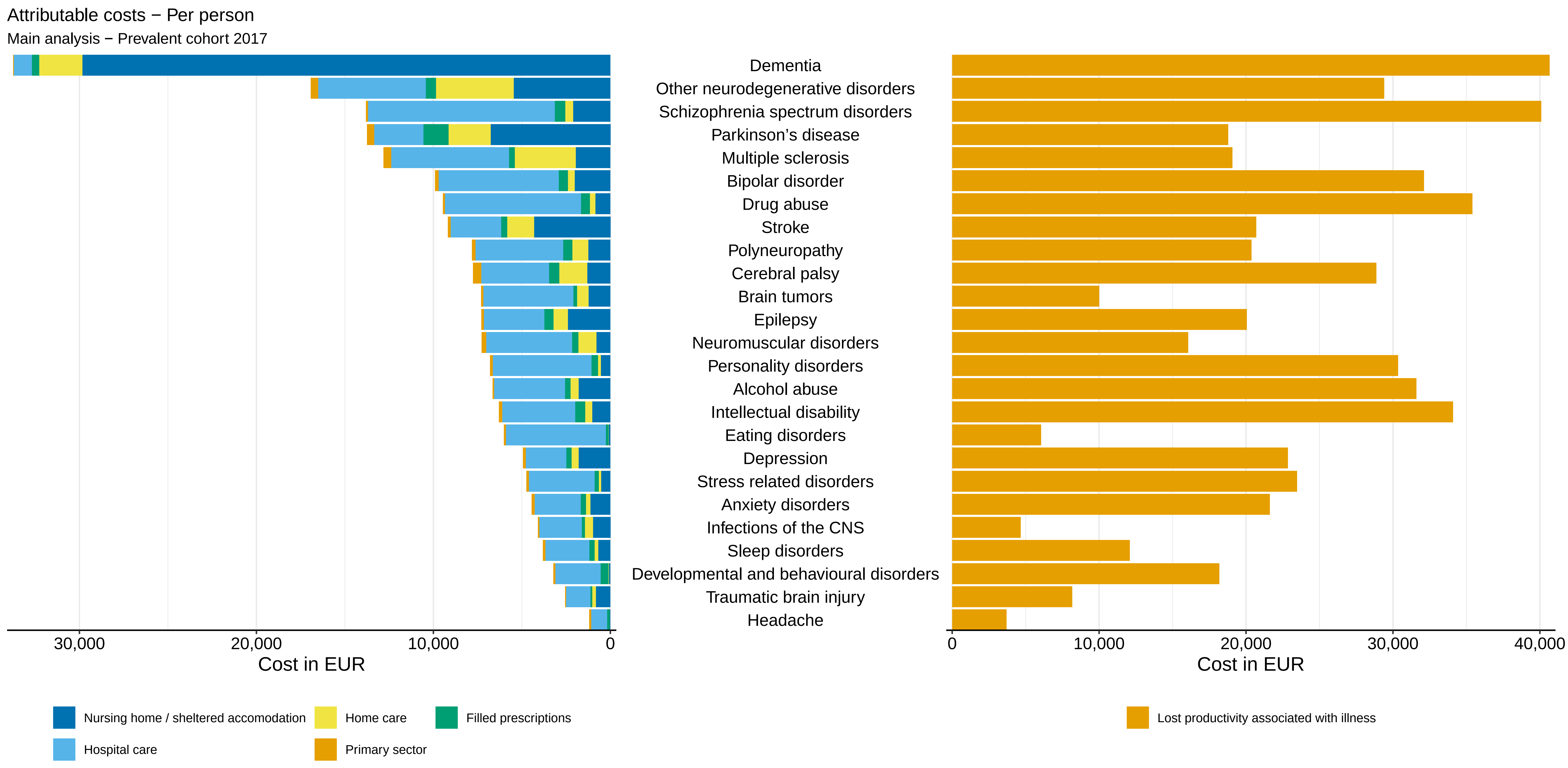

Attributable costs − Per person
Main analysis − Prevalent cohort 2017
Dementia
Other neurodegenerative disorders
Schizophrenia spectrum disorders
Parkinson's disease
Multiple sclerosis
Bipolar disorder
Drug abuse
Stroke
Polyneuropathy
Cerebral palsy
Brain tumors
Epilepsy
Neuromuscular disorders
Personality disorders
Alcohol abuse
Intellectual disability
Eating disorders
Depression
Stress related disorders
Anxiety disorders
Infections of the CNS
Sleep disorders
Developmental and behavioural disorders
Traumatic brain injury
Headache
30,000
20,000
10,000
0
Cost in EUR
0
10,000
20,000
30,000
40,000
Cost in EUR
Nursing home / sheltered accomodation
Home care
Filled prescriptions
Hospital care
Primary sector
Lost productivity associated with illness

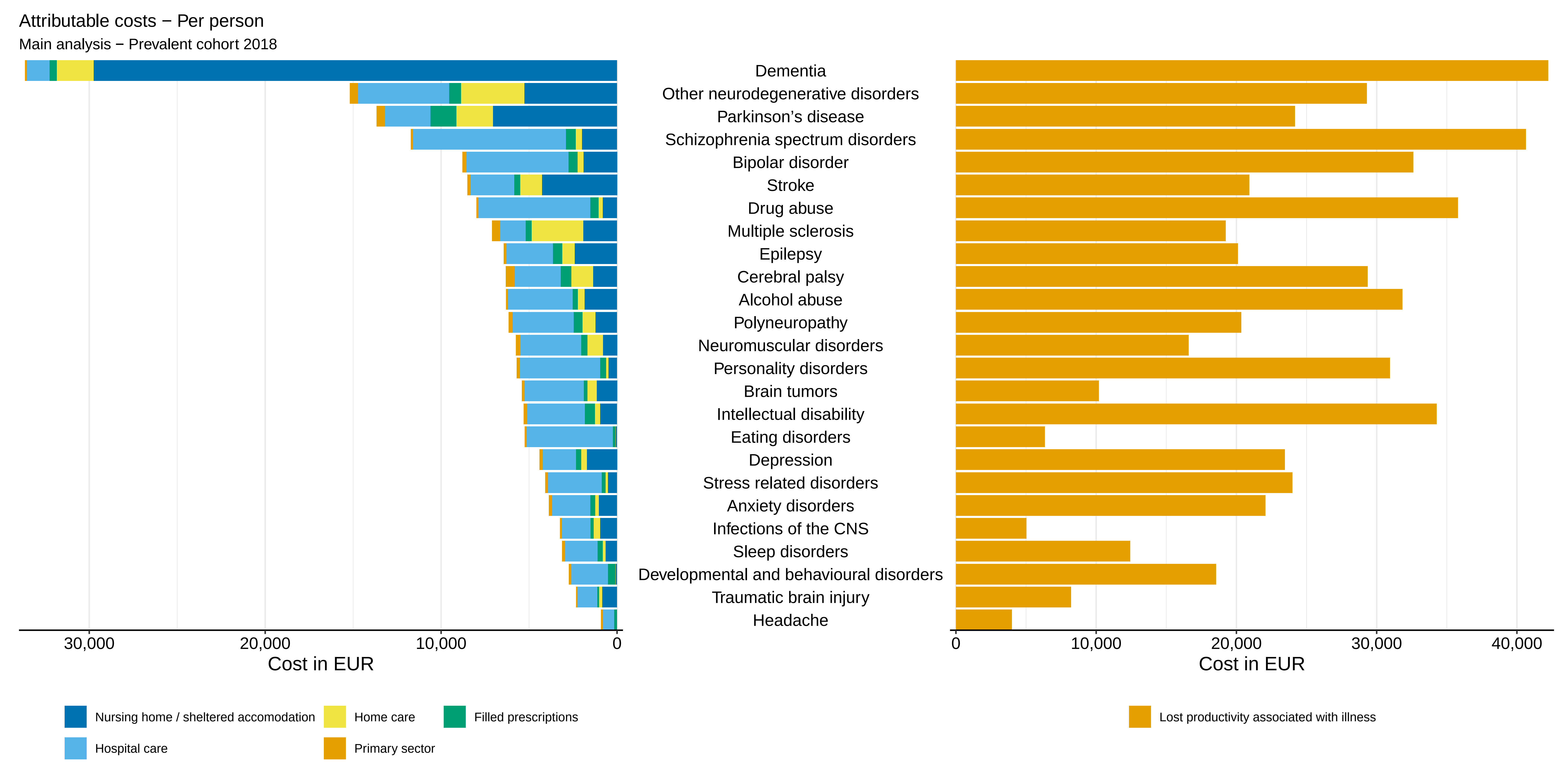

Attributable costs − Per person
Main analysis − Prevalent cohort 2018
Dementia
Other neurodegenerative disorders
Parkinson's disease
Schizophrenia spectrum disorders
Bipolar disorder
Stroke
Drug abuse
Multiple sclerosis
Epilepsy
Cerebral palsy
Alcohol abuse
Polyneuropathy
Neuromuscular disorders
Personality disorders
Brain tumors
Intellectual disability
Eating disorders
Depression
Stress related disorders
Anxiety disorders
Infections of the CNS
Sleep disorders
Developmental and behavioural disorders
Traumatic brain injury
Headache
30,000
20,000
10,000
0
Cost in EUR
0
10,000
20,000
30,000
40,000
Cost in EUR
Nursing home / sheltered accomodation
Home care
Filled prescriptions
Hospital care
Primary sector
Lost productivity associated with illness

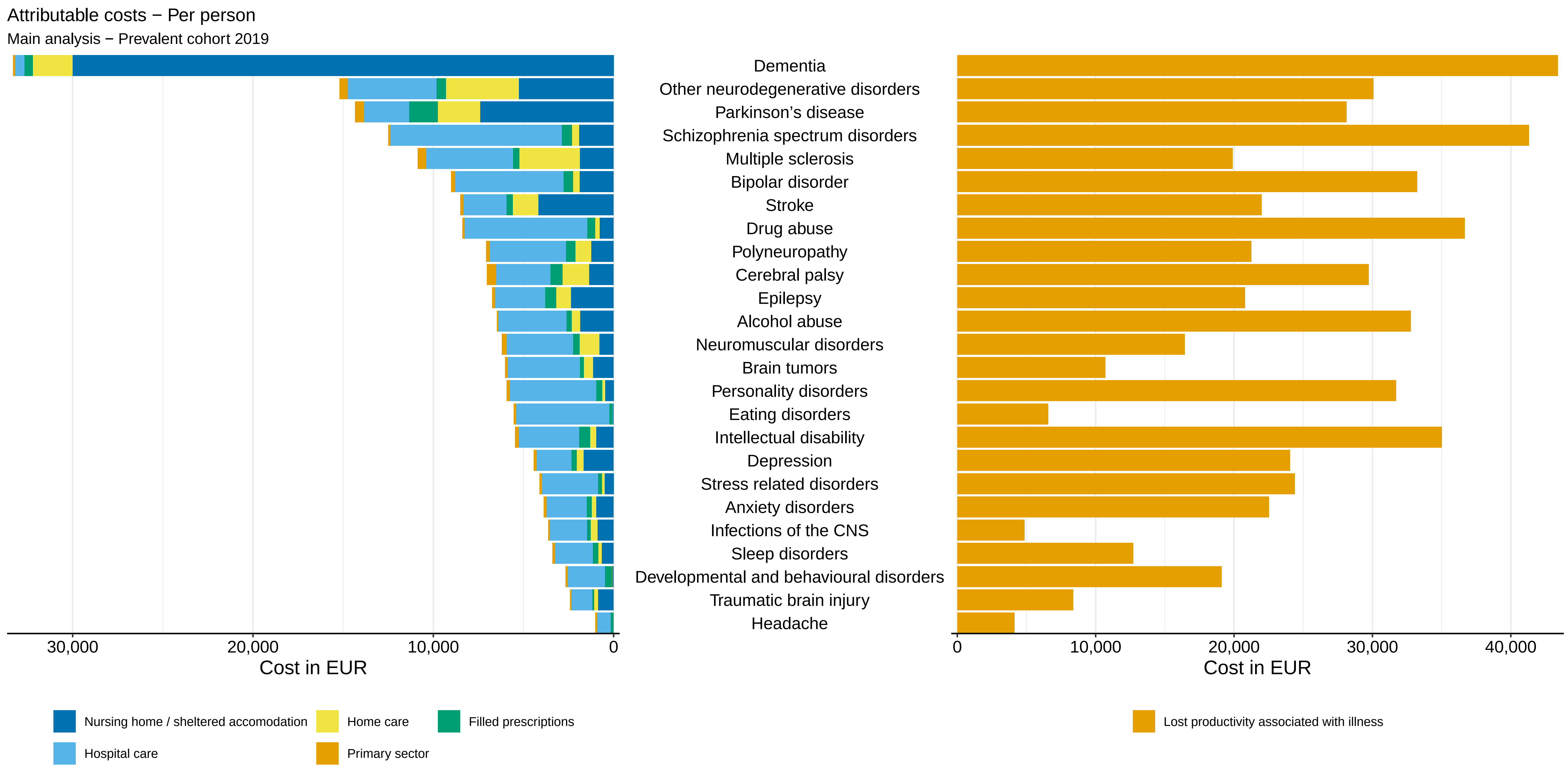

Attributable costs − Per person
Main analysis − Prevalent cohort 2019
Dementia
Other neurodegenerative disorders
Parkinson's disease
Schizophrenia spectrum disorders
Multiple sclerosis
Bipolar disorder
Stroke
Drug abuse
Polyneuropathy
Cerebral palsy
Epilepsy
Alcohol abuse
Neuromuscular disorders
Brain tumors
Personality disorders
Eating disorders
Intellectual disability
Depression
Stress related disorders
Anxiety disorders
Infections of the CNS
Sleep disorders
Developmental and behavioural disorders
Traumatic brain injury
Headache
30,000
20,000
10,000
0
Cost in EUR
0
10,000
20,000
30,000
40,000
Cost in EUR
Nursing home / sheltered accomodation
Home care
Filled prescriptions
Hospital care
Primary sector
Lost productivity associated with illness

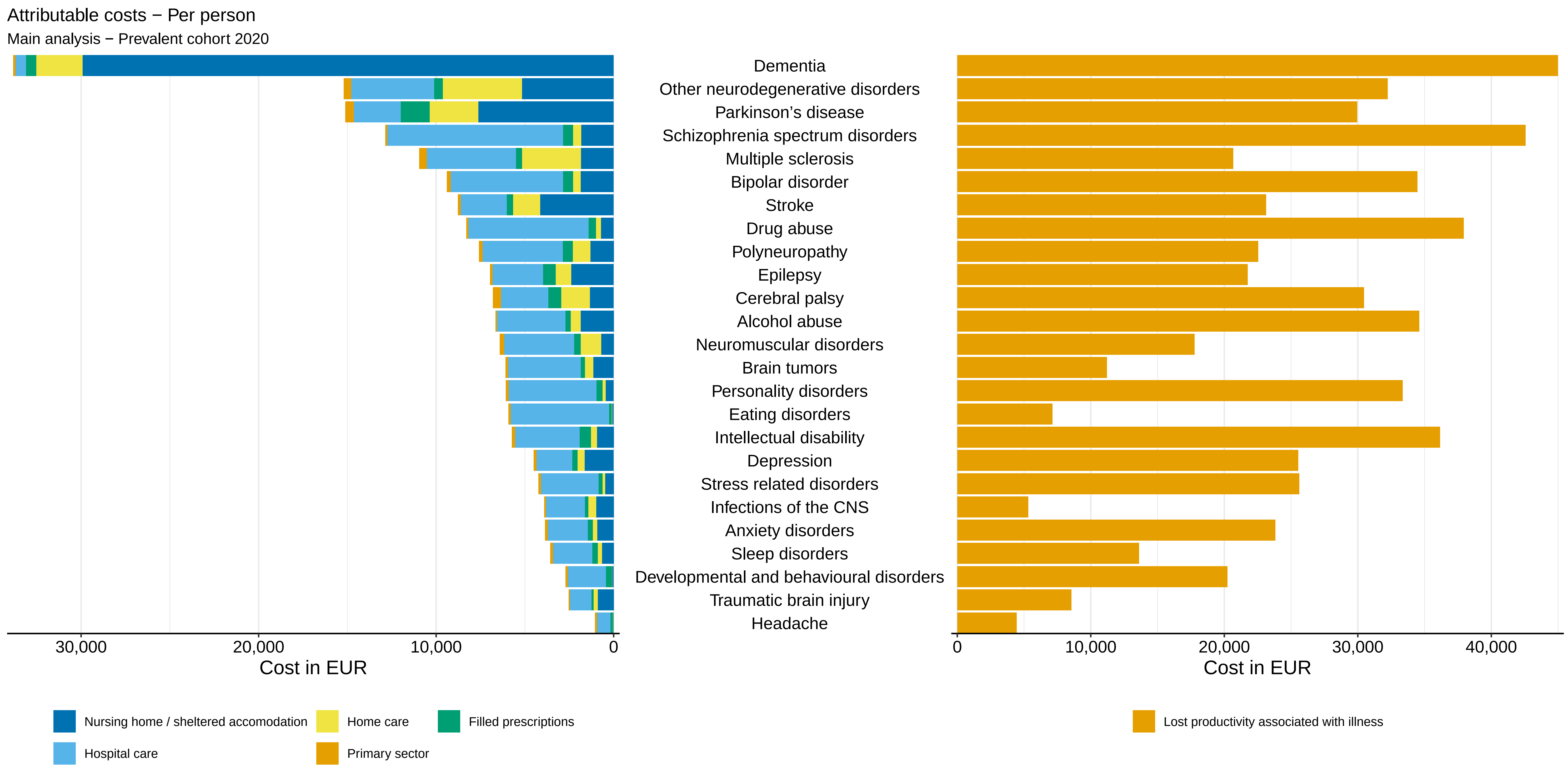

Attributable costs − Per person
Main analysis − Prevalent cohort 2020
Dementia
Other neurodegenerative disorders
Parkinson's disease
Schizophrenia spectrum disorders
Multiple sclerosis
Bipolar disorder
Stroke
Drug abuse
Polyneuropathy
Epilepsy
Cerebral palsy
Alcohol abuse
Neuromuscular disorders
Brain tumors
Personality disorders
Eating disorders
Intellectual disability
Depression
Stress related disorders
Infections of the CNS
Anxiety disorders
Sleep disorders
Developmental and behavioural disorders
Traumatic brain injury
Headache
30,000
20,000
10,000
0
Cost in EUR
0
10,000
20,000
30,000
40,000
Cost in EUR
Nursing home / sheltered accomodation
Home care
Filled prescriptions
Hospital care
Primary sector
Lost productivity associated with illness

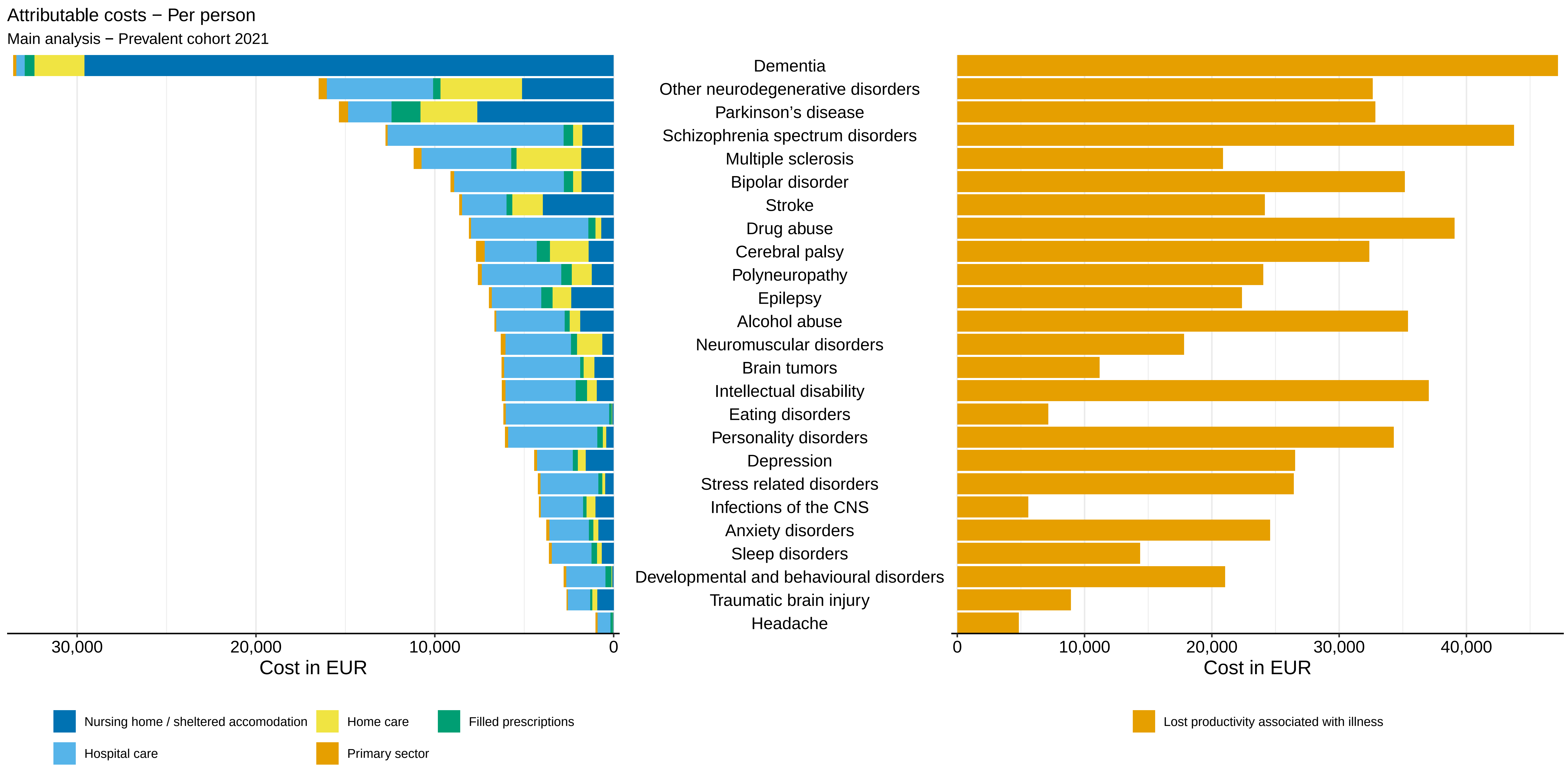
Attributable costs − Per person
Main analysis − Prevalent cohort 2021
Dementia
Other neurodegenerative disorders
Parkinson's disease
Schizophrenia spectrum disorders
Multiple sclerosis
Bipolar disorder
Stroke
Drug abuse
Cerebral palsy
Polyneuropathy
Epilepsy
Alcohol abuse
Neuromuscular disorders
Brain tumors
Intellectual disability
Eating disorders
Personality disorders
Depression
Stress related disorders
Infections of the CNS
Anxiety disorders
Sleep disorders
Developmental and behavioural disorders
Traumatic brain injury
Headache
30,000
20,000
10,000
0
Cost in EUR
0
10,000
20,000
30,000
40,000
Cost in EUR
Nursing home / sheltered accomodation
Home care
Filled prescriptions
Hospital care
Primary sector
Lost productivity associated with illness

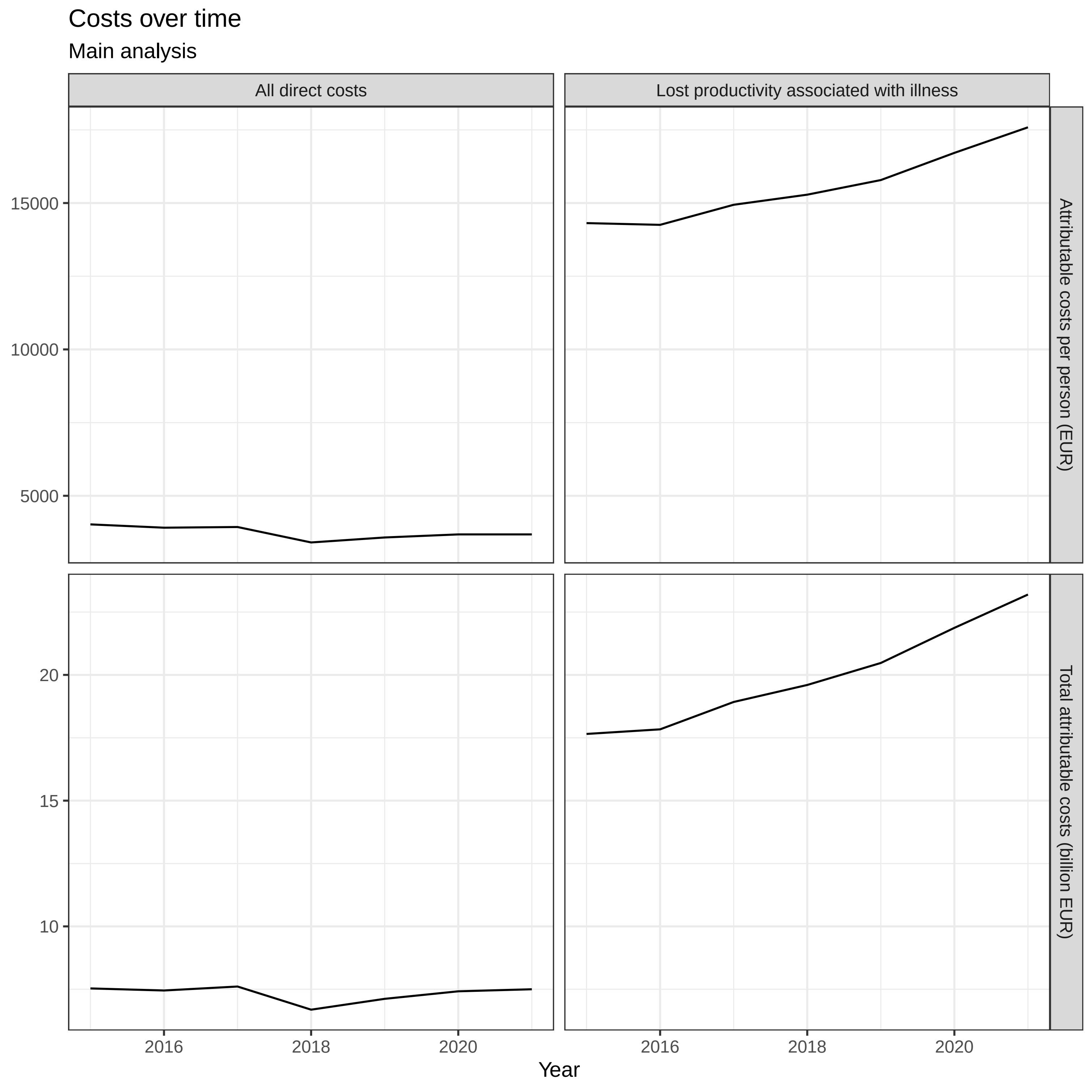
Costs over time
Main analysis
All direct costs
Lost productivity associated with illness
Attributable costs per person (EUR)
Total attributable costs (billion EUR)
15000
10000
5000
20
15
10
2016
2018
2020
Year
