## Appendix 8 for "Temporal trends in the occurrence, mortality, and cost of brain disorders in Denmark"

| | *CCI covariates included in model* | *No CCI covariates included in model* |
|---|---|---|
| Age 10-19 | -0.01 (-0.02 - -0.01) | -0.02 (-0.02 - -0.01) |
| Age 20-29 | 0.00 (-0.00 - 0.00) | -0.01 (-0.02 - -0.01) |
| Age 30-39 | 0.01 (0.00 - 0.01) | -0.00 (-0.01 - 0.00) |
| Age 40-49 | -0.00 (-0.00 - 0.00) | 0.00 (-0.00 - 0.01) |
| Age 50-59 | 0.03 (0.03 - 0.04) | 0.06 (0.06 - 0.07) |
| Age 60-69 | 0.11 (0.10 - 0.11) | 0.20 (0.20 - 0.20) |
| Age 70-79 | 0.23 (0.22 - 0.23) | 0.40 (0.40 - 0.41) |
| Age 80-89 | 0.75 (0.75 - 0.76) | 1.00 (0.99 - 1.01) |
| Age 90+ | 2.16 (2.14 - 2.17) | 2.39 (2.38 - 2.40) |
| Myocardial infarction | 0.06 (0.05 - 0.07) | |
| Diabetes without end-organ damage | 0.27 (0.27 - 0.28) | |
| Hemiplegia | 1.13 (1.11 - 1.16) | |
| Moderate to severe renal disease | 0.78 (0.77 - 0.79) | |
| Diabetes with end-organ damage | 0.42 (0.41 - 0.43) | |
| Solid cancer tumour | 0.35 (0.34 - 0.36) | |
| Leukaemia | 1.08 (1.06 - 1.11) | |
| Lymphoma | 0.97 (0.95 - 0.99) | |
| Moderate to severe liver disease | 0.64 (0.61 - 0.67) | |
| Metastatic solid tumour | 1.05 (1.03 - 1.07) | |
| AIDS | 0.21 (0.18 - 0.24) | |
| Congestive heart failure | 0.51 (0.50 - 0.52) | |
| Peripheral vascular disease | 0.30 (0.29 - 0.31) | |
| Cerebrovascular disease | 0.40 (0.39 - 0.41) | |
| Chronic pulmonary disease | 0.28 (0.28 - 0.29) | |
| Connective tissue disease | 0.34 (0.34 - 0.35) | |
| Ulcer disease | 0.34 (0.33 - 0.35) | |
| Mild liver disease | 0.29 (0.28 - 0.31) | |
| Alcohol abuse | 0.17 (0.16 - 0.17) | 0.22 (0.22 - 0.23) |
| Anxiety disorders | 0.08 (0.08 - 0.09) | 0.09 (0.09 - 0.09) |
| Bipolar disorder | 0.41 (0.40 - 0.42) | 0.40 (0.39 - 0.41) |
| Brain tumors | 0.29 (0.28 - 0.31) | 0.35 (0.33 - 0.36) |
| Cerebral palsy | 0.24 (0.22 - 0.26) | 0.50 (0.48 - 0.53) |
| Dementia | 1.10 (1.09 - 1.11) | 1.10 (1.09 - 1.11) |
| Depression | 0.14 (0.14 - 0.15) | 0.17 (0.17 - 0.18) |
| Developmental and behavioural disorders | 0.01 (0.00 - 0.01) | -0.01 (-0.01 - -0.00) |
| Drug abuse | 0.23 (0.22 - 0.24) | 0.24 (0.24 - 0.25) |
| Eating disorders | 0.41 (0.40 - 0.43) | 0.41 (0.40 - 0.43) |
| Epilepsy | 0.38 (0.37 - 0.38) | 0.43 (0.42 - 0.44) |
| Headache | 0.02 (0.01 - 0.02) | 0.03 (0.02 - 0.03) |
| Infections of the CNS | 0.23 (0.21 - 0.24) | 0.28 (0.27 - 0.30) |

| | *CCI covariates included in model* | *No CCI covariates included in model* |
|---|---|---|
| Intellectual disability | 0.23 (0.21 - 0.24) | 0.24 (0.23 - 0.25) |
| Multiple sclerosis | 0.72 (0.70 - 0.73) | 0.72 (0.71 - 0.74) |
| Neuromuscular disorders | 0.33 (0.31 - 0.35) | 0.43 (0.41 - 0.45) |
| Other neurodegenerative disorders | 0.62 (0.59 - 0.65) | 0.66 (0.63 - 0.68) |
| Parkinson’s disease | 0.36 (0.35 - 0.37) | 0.37 (0.36 - 0.38) |
| Personality disorders | 0.14 (0.14 - 0.15) | 0.13 (0.13 - 0.14) |
| Polyneuropathy | 0.21 (0.21 - 0.22) | 0.41 (0.40 - 0.42) |
| Schizophrenia spectrum disorders | 0.78 (0.77 - 0.79) | 0.77 (0.77 - 0.78) |
| Sleep disorders | 0.09 (0.08 - 0.09) | 0.12 (0.12 - 0.13) |
| Stress related disorders | 0.06 (0.06 - 0.07) | 0.06 (0.06 - 0.07) |
| Stroke | 0.21 (0.21 - 0.22) | 0.44 (0.44 - 0.45) |
| Traumatic brain injury | 0.07 (0.06 - 0.07) | 0.08 (0.07 - 0.08) |
| Male | -0.04 (-0.04 - -0.04) | -0.03 (-0.03 - -0.02) |

| | *CCI covariates included in model* | *No CCI covariates included in model* |
|---|---|---|
| Age 10-19 | -0.01 (-0.02 - -0.01) | -0.02 (-0.02 - -0.01) |
| Age 20-29 | -0.00 (-0.01 - 0.00) | -0.01 (-0.02 - -0.01) |
| Age 30-39 | 0.00 (-0.00 - 0.01) | -0.01 (-0.01 - -0.00) |
| Age 40-49 | -0.01 (-0.01 - -0.00) | -0.00 (-0.01 - -0.00) |
| Age 50-59 | 0.03 (0.02 - 0.03) | 0.06 (0.06 - 0.06) |
| Age 60-69 | 0.10 (0.10 - 0.10) | 0.19 (0.19 - 0.19) |
| Age 70-79 | 0.22 (0.21 - 0.22) | 0.38 (0.38 - 0.39) |
| Age 80-89 | 0.71 (0.70 - 0.71) | 0.94 (0.94 - 0.95) |
| Age 90+ | 2.07 (2.06 - 2.08) | 2.29 (2.28 - 2.31) |
| Myocardial infarction | 0.05 (0.04 - 0.06) | |
| Diabetes without end-organ damage | 0.27 (0.26 - 0.28) | |
| Hemiplegia | 1.07 (1.05 - 1.10) | |
| Moderate to severe renal disease | 0.71 (0.70 - 0.72) | |
| Diabetes with end-organ damage | 0.40 (0.39 - 0.41) | |
| Solid cancer tumour | 0.33 (0.33 - 0.34) | |
| Leukaemia | 0.93 (0.91 - 0.96) | |
| Lymphoma | 0.89 (0.88 - 0.91) | |
| Moderate to severe liver disease | 0.68 (0.65 - 0.71) | |
| Metastatic solid tumour | 0.97 (0.96 - 0.99) | |
| AIDS | 0.23 (0.20 - 0.26) | |
| Congestive heart failure | 0.50 (0.49 - 0.51) | |
| Peripheral vascular disease | 0.29 (0.28 - 0.30) | |
| Cerebrovascular disease | 0.34 (0.34 - 0.35) | |
| Chronic pulmonary disease | 0.27 (0.27 - 0.28) | |
| Connective tissue disease | 0.34 (0.33 - 0.34) | |
| Ulcer disease | 0.31 (0.30 - 0.32) | |
| Mild liver disease | 0.28 (0.26 - 0.29) | |
| Alcohol abuse | 0.17 (0.16 - 0.17) | 0.22 (0.22 - 0.23) |
| Anxiety disorders | 0.08 (0.07 - 0.08) | 0.08 (0.08 - 0.09) |
| Bipolar disorder | 0.38 (0.37 - 0.39) | 0.37 (0.36 - 0.39) |
| Brain tumors | 0.33 (0.32 - 0.35) | 0.39 (0.38 - 0.40) |
| Cerebral palsy | 0.25 (0.23 - 0.27) | 0.50 (0.48 - 0.52) |
| Dementia | 1.20 (1.19 - 1.21) | 1.20 (1.19 - 1.21) |
| Depression | 0.14 (0.14 - 0.14) | 0.17 (0.17 - 0.17) |
| Developmental and behavioural disorders | 0.02 (0.01 - 0.02) | 0.00 (-0.00 - 0.00) |
| Drug abuse | 0.24 (0.23 - 0.25) | 0.25 (0.24 - 0.26) |
| Eating disorders | 0.40 (0.39 - 0.42) | 0.40 (0.39 - 0.42) |
| Epilepsy | 0.38 (0.37 - 0.39) | 0.43 (0.42 - 0.44) |
| Headache | 0.02 (0.01 - 0.02) | 0.03 (0.02 - 0.03) |
| Infections of the CNS | 0.25 (0.24 - 0.26) | 0.30 (0.29 - 0.32) |

| | *CCI covariates included in model* | *No CCI covariates included in model* |
|---|---|---|
| Intellectual disability | 0.23 (0.21 - 0.24) | 0.24 (0.23 - 0.25) |
| Multiple sclerosis | 0.80 (0.79 - 0.82) | 0.81 (0.79 - 0.82) |
| Neuromuscular disorders | 0.34 (0.31 - 0.36) | 0.43 (0.41 - 0.45) |
| Other neurodegenerative disorders | 0.66 (0.64 - 0.69) | 0.70 (0.67 - 0.73) |
| Parkinson's disease | 0.40 (0.39 - 0.41) | 0.41 (0.40 - 0.42) |
| Personality disorders | 0.12 (0.11 - 0.13) | 0.11 (0.10 - 0.12) |
| Polyneuropathy | 0.22 (0.21 - 0.23) | 0.41 (0.41 - 0.42) |
| Schizophrenia spectrum disorders | 0.75 (0.75 - 0.76) | 0.75 (0.74 - 0.76) |
| Sleep disorders | 0.09 (0.09 - 0.10) | 0.13 (0.13 - 0.14) |
| Stress related disorders | 0.07 (0.07 - 0.08) | 0.08 (0.07 - 0.08) |
| Stroke | 0.24 (0.24 - 0.25) | 0.46 (0.45 - 0.46) |
| Traumatic brain injury | 0.07 (0.07 - 0.08) | 0.09 (0.08 - 0.09) |
| Male | -0.04 (-0.04 - -0.03) | -0.02 (-0.03 - -0.02) |

| | *CCI covariates included in model* | *No CCI covariates included in model* |
|---|---|---|
| Age 10-19 | -0.01 (-0.01 - -0.00) | -0.01 (-0.02 - -0.01) |
| Age 20-29 | 0.01 (0.00 - 0.01) | -0.01 (-0.01 - -0.00) |
| Age 30-39 | 0.01 (0.01 - 0.01) | 0.00 (-0.00 - 0.01) |
| Age 40-49 | -0.00 (-0.00 - 0.00) | 0.00 (-0.00 - 0.01) |
| Age 50-59 | 0.03 (0.03 - 0.04) | 0.07 (0.06 - 0.07) |
| Age 60-69 | 0.11 (0.11 - 0.11) | 0.20 (0.20 - 0.21) |
| Age 70-79 | 0.23 (0.22 - 0.23) | 0.40 (0.40 - 0.41) |
| Age 80-89 | 0.68 (0.68 - 0.69) | 0.93 (0.92 - 0.93) |
| Age 90+ | 2.01 (2.00 - 2.02) | 2.23 (2.22 - 2.25) |
| Myocardial infarction | 0.05 (0.04 - 0.06) | |
| Diabetes without end-organ damage | 0.28 (0.27 - 0.28) | |
| Hemiplegia | 1.16 (1.14 - 1.18) | |
| Moderate to severe renal disease | 0.73 (0.72 - 0.74) | |
| Diabetes with end-organ damage | 0.42 (0.41 - 0.43) | |
| Solid cancer tumour | 0.35 (0.35 - 0.36) | |
| Leukaemia | 0.95 (0.93 - 0.98) | |
| Lymphoma | 0.95 (0.94 - 0.97) | |
| Moderate to severe liver disease | 0.70 (0.67 - 0.73) | |
| Metastatic solid tumour | 0.98 (0.96 - 0.99) | |
| AIDS | 0.20 (0.17 - 0.24) | |
| Congestive heart failure | 0.48 (0.47 - 0.49) | |
| Peripheral vascular disease | 0.30 (0.29 - 0.30) | |
| Cerebrovascular disease | 0.30 (0.30 - 0.31) | |
| Chronic pulmonary disease | 0.28 (0.28 - 0.28) | |
| Connective tissue disease | 0.33 (0.32 - 0.34) | |
| Ulcer disease | 0.31 (0.30 - 0.32) | |
| Mild liver disease | 0.27 (0.26 - 0.28) | |
| Alcohol abuse | 0.17 (0.17 - 0.18) | 0.23 (0.23 - 0.24) |
| Anxiety disorders | 0.06 (0.06 - 0.07) | 0.07 (0.06 - 0.07) |
| Bipolar disorder | 0.34 (0.33 - 0.35) | 0.33 (0.32 - 0.35) |
| Brain tumors | 0.36 (0.35 - 0.37) | 0.43 (0.41 - 0.44) |
| Cerebral palsy | 0.22 (0.20 - 0.24) | 0.49 (0.47 - 0.51) |
| Dementia | 1.30 (1.30 - 1.31) | 1.31 (1.30 - 1.32) |
| Depression | 0.14 (0.14 - 0.14) | 0.17 (0.17 - 0.17) |
| Developmental and behavioural disorders | 0.02 (0.02 - 0.03) | 0.00 (-0.00 - 0.01) |
| Drug abuse | 0.25 (0.24 - 0.25) | 0.26 (0.25 - 0.27) |
| Eating disorders | 0.39 (0.38 - 0.41) | 0.39 (0.38 - 0.41) |
| Epilepsy | 0.38 (0.37 - 0.39) | 0.43 (0.42 - 0.44) |
| Headache | 0.02 (0.02 - 0.02) | 0.03 (0.03 - 0.03) |
| Infections of the CNS | 0.25 (0.24 - 0.26) | 0.31 (0.30 - 0.33) |

| | *CCI covariates included in model* | *No CCI covariates included in model* |
|---|---|---|
| Intellectual disability | 0.22 (0.21 - 0.23) | 0.23 (0.22 - 0.24) |
| Multiple sclerosis | 0.92 (0.90 - 0.93) | 0.92 (0.91 - 0.94) |
| Neuromuscular disorders | 0.32 (0.29 - 0.34) | 0.42 (0.40 - 0.45) |
| Other neurodegenerative disorders | 0.72 (0.69 - 0.75) | 0.77 (0.74 - 0.80) |
| Parkinson’s disease | 0.45 (0.44 - 0.46) | 0.46 (0.45 - 0.47) |
| Personality disorders | 0.12 (0.12 - 0.13) | 0.12 (0.11 - 0.12) |
| Polyneuropathy | 0.26 (0.25 - 0.27) | 0.47 (0.46 - 0.48) |
| Schizophrenia spectrum disorders | 0.78 (0.77 - 0.79) | 0.78 (0.77 - 0.79) |
| Sleep disorders | 0.11 (0.11 - 0.12) | 0.16 (0.15 - 0.16) |
| Stress related disorders | 0.08 (0.07 - 0.08) | 0.08 (0.08 - 0.08) |
| Stroke | 0.30 (0.29 - 0.31) | 0.51 (0.51 - 0.52) |
| Traumatic brain injury | 0.08 (0.08 - 0.08) | 0.09 (0.09 - 0.10) |
| Male | -0.04 (-0.04 - -0.04) | -0.03 (-0.03 - -0.03) |

| | *CCI covariates included in model* | *No CCI covariates included in model* |
|---|---|---|
| Age 10-19 | -0.01 (-0.01 - -0.00) | -0.01 (-0.01 - -0.01) |
| Age 20-29 | -0.00 (-0.01 - 0.00) | -0.01 (-0.01 - -0.01) |
| Age 30-39 | -0.01 (-0.01 - -0.00) | -0.01 (-0.02 - -0.01) |
| Age 40-49 | -0.02 (-0.02 - -0.02) | -0.02 (-0.02 - -0.02) |
| Age 50-59 | 0.01 (0.00 - 0.01) | 0.03 (0.03 - 0.03) |
| Age 60-69 | 0.08 (0.08 - 0.09) | 0.14 (0.14 - 0.15) |
| Age 70-79 | 0.19 (0.19 - 0.20) | 0.31 (0.30 - 0.31) |
| Age 80-89 | 0.64 (0.63 - 0.64) | 0.80 (0.80 - 0.81) |
| Age 90+ | 1.97 (1.96 - 1.98) | 2.13 (2.12 - 2.14) |
| Myocardial infarction | 0.04 (0.03 - 0.05) | |
| Diabetes without end-organ damage | 0.23 (0.22 - 0.23) | |
| Hemiplegia | 1.10 (1.08 - 1.13) | |
| Moderate to severe renal disease | 0.42 (0.41 - 0.42) | |
| Diabetes with end-organ damage | 0.32 (0.31 - 0.32) | |
| Solid cancer tumour | 0.18 (0.17 - 0.18) | |
| Leukaemia | 0.92 (0.90 - 0.94) | |
| Lymphoma | 0.51 (0.50 - 0.53) | |
| Moderate to severe liver disease | 0.67 (0.64 - 0.69) | |
| Metastatic solid tumour | 0.47 (0.45 - 0.48) | |
| AIDS | 0.04 (0.01 - 0.07) | |
| Congestive heart failure | 0.41 (0.40 - 0.42) | |
| Peripheral vascular disease | 0.24 (0.24 - 0.25) | |
| Cerebrovascular disease | 0.26 (0.26 - 0.27) | |
| Chronic pulmonary disease | 0.23 (0.22 - 0.23) | |
| Connective tissue disease | 0.11 (0.10 - 0.12) | |
| Ulcer disease | 0.30 (0.29 - 0.32) | |
| Mild liver disease | 0.22 (0.21 - 0.23) | |
| Alcohol abuse | 0.19 (0.19 - 0.20) | 0.25 (0.24 - 0.25) |
| Anxiety disorders | 0.05 (0.05 - 0.05) | 0.05 (0.05 - 0.06) |
| Bipolar disorder | 0.32 (0.31 - 0.33) | 0.31 (0.30 - 0.32) |
| Brain tumors | 0.32 (0.31 - 0.34) | 0.37 (0.35 - 0.38) |
| Cerebral palsy | 0.11 (0.09 - 0.13) | 0.37 (0.35 - 0.39) |
| Dementia | 1.45 (1.44 - 1.46) | 1.46 (1.45 - 1.47) |
| Depression | 0.14 (0.13 - 0.14) | 0.16 (0.16 - 0.16) |
| Developmental and behavioural disorders | 0.02 (0.02 - 0.03) | 0.01 (0.01 - 0.01) |
| Drug abuse | 0.20 (0.20 - 0.21) | 0.21 (0.20 - 0.22) |
| Eating disorders | 0.32 (0.31 - 0.33) | 0.32 (0.30 - 0.33) |
| Epilepsy | 0.33 (0.33 - 0.34) | 0.37 (0.37 - 0.38) |
| Headache | 0.00 (0.00 - 0.01) | 0.01 (0.01 - 0.01) |
| Infections of the CNS | 0.25 (0.24 - 0.26) | 0.30 (0.28 - 0.31) |

| | *CCI covariates included in model* | *No CCI covariates included in model* |
|---|---|---|
| Intellectual disability | 0.17 (0.16 - 0.18) | 0.18 (0.17 - 0.19) |
| Multiple sclerosis | 0.48 (0.46 - 0.49) | 0.48 (0.47 - 0.50) |
| Neuromuscular disorders | 0.25 (0.23 - 0.27) | 0.33 (0.31 - 0.34) |
| Other neurodegenerative disorders | 0.72 (0.69 - 0.74) | 0.77 (0.74 - 0.80) |
| Parkinson’s disease | 0.48 (0.47 - 0.49) | 0.49 (0.47 - 0.50) |
| Personality disorders | 0.09 (0.08 - 0.10) | 0.09 (0.08 - 0.09) |
| Polyneuropathy | 0.19 (0.18 - 0.20) | 0.35 (0.34 - 0.36) |
| Schizophrenia spectrum disorders | 0.69 (0.68 - 0.70) | 0.69 (0.68 - 0.70) |
| Sleep disorders | 0.09 (0.09 - 0.09) | 0.12 (0.12 - 0.12) |
| Stress related disorders | 0.07 (0.07 - 0.08) | 0.08 (0.07 - 0.08) |
| Stroke | 0.32 (0.32 - 0.33) | 0.52 (0.51 - 0.52) |
| Traumatic brain injury | 0.08 (0.07 - 0.08) | 0.09 (0.09 - 0.09) |
| Male | -0.03 (-0.03 - -0.03) | -0.02 (-0.02 - -0.02) |

| | *CCI covariates included in model* | *No CCI covariates included in model* |
|---|---|---|
| Age 10-19 | -0.01 (-0.01 - -0.01) | -0.01 (-0.02 - -0.01) |
| Age 20-29 | 0.01 (0.01 - 0.01) | 0.00 (-0.00 - 0.01) |
| Age 30-39 | 0.01 (0.00 - 0.01) | 0.00 (-0.00 - 0.00) |
| Age 40-49 | -0.01 (-0.02 - -0.01) | -0.01 (-0.01 - -0.00) |
| Age 50-59 | 0.01 (0.01 - 0.02) | 0.04 (0.04 - 0.05) |
| Age 60-69 | 0.09 (0.08 - 0.09) | 0.17 (0.17 - 0.17) |
| Age 70-79 | 0.21 (0.20 - 0.21) | 0.35 (0.35 - 0.36) |
| Age 80-89 | 0.62 (0.62 - 0.63) | 0.82 (0.82 - 0.83) |
| Age 90+ | 1.97 (1.96 - 1.98) | 2.16 (2.15 - 2.17) |
| Myocardial infarction | 0.03 (0.02 - 0.04) | |
| Diabetes without end-organ damage | 0.26 (0.25 - 0.26) | |
| Hemiplegia | 1.14 (1.12 - 1.16) | |
| Moderate to severe renal disease | 0.58 (0.57 - 0.59) | |
| Diabetes with end-organ damage | 0.39 (0.38 - 0.40) | |
| Solid cancer tumour | 0.28 (0.28 - 0.29) | |
| Leukaemia | 0.84 (0.82 - 0.86) | |
| Lymphoma | 0.87 (0.85 - 0.89) | |
| Moderate to severe liver disease | 0.66 (0.64 - 0.69) | |
| Metastatic solid tumour | 0.79 (0.78 - 0.81) | |
| AIDS | 0.12 (0.09 - 0.15) | |
| Congestive heart failure | 0.41 (0.41 - 0.42) | |
| Peripheral vascular disease | 0.26 (0.25 - 0.26) | |
| Cerebrovascular disease | 0.20 (0.19 - 0.20) | |
| Chronic pulmonary disease | 0.25 (0.25 - 0.26) | |
| Connective tissue disease | 0.22 (0.21 - 0.23) | |
| Ulcer disease | 0.27 (0.26 - 0.29) | |
| Mild liver disease | 0.23 (0.21 - 0.24) | |
| Alcohol abuse | 0.20 (0.20 - 0.21) | 0.26 (0.25 - 0.26) |
| Anxiety disorders | 0.04 (0.03 - 0.04) | 0.04 (0.04 - 0.04) |
| Bipolar disorder | 0.34 (0.33 - 0.35) | 0.34 (0.33 - 0.35) |
| Brain tumors | 0.37 (0.36 - 0.38) | 0.44 (0.42 - 0.45) |
| Cerebral palsy | 0.15 (0.13 - 0.17) | 0.40 (0.38 - 0.42) |
| Dementia | 1.61 (1.60 - 1.62) | 1.61 (1.60 - 1.62) |
| Depression | 0.14 (0.13 - 0.14) | 0.16 (0.16 - 0.16) |
| Developmental and behavioural disorders | 0.01 (0.01 - 0.01) | -0.01 (-0.01 - -0.00) |
| Drug abuse | 0.23 (0.23 - 0.24) | 0.24 (0.23 - 0.25) |
| Eating disorders | 0.36 (0.35 - 0.38) | 0.36 (0.35 - 0.38) |
| Epilepsy | 0.36 (0.35 - 0.36) | 0.40 (0.39 - 0.41) |
| Headache | 0.01 (0.00 - 0.01) | 0.01 (0.01 - 0.02) |
| Infections of the CNS | 0.27 (0.25 - 0.28) | 0.32 (0.31 - 0.34) |

| | *CCI covariates included in model* | *No CCI covariates included in model* |
|---|---|---|
| Intellectual disability | 0.16 (0.15 - 0.18) | 0.18 (0.17 - 0.19) |
| Multiple sclerosis | 0.81 (0.80 - 0.83) | 0.82 (0.80 - 0.83) |
| Neuromuscular disorders | 0.29 (0.27 - 0.31) | 0.39 (0.37 - 0.41) |
| Other neurodegenerative disorders | 0.77 (0.74 - 0.80) | 0.82 (0.80 - 0.85) |
| Parkinson’s disease | 0.56 (0.54 - 0.57) | 0.56 (0.55 - 0.57) |
| Personality disorders | 0.09 (0.09 - 0.10) | 0.09 (0.08 - 0.09) |
| Polyneuropathy | 0.25 (0.24 - 0.26) | 0.45 (0.44 - 0.46) |
| Schizophrenia spectrum disorders | 0.77 (0.76 - 0.77) | 0.77 (0.76 - 0.77) |
| Sleep disorders | 0.12 (0.12 - 0.12) | 0.16 (0.16 - 0.16) |
| Stress related disorders | 0.07 (0.06 - 0.07) | 0.07 (0.07 - 0.07) |
| Stroke | 0.40 (0.40 - 0.41) | 0.58 (0.58 - 0.59) |
| Traumatic brain injury | 0.08 (0.08 - 0.09) | 0.10 (0.09 - 0.10) |
| Male | -0.04 (-0.04 - -0.03) | -0.02 (-0.03 - -0.02) |

| | *CCI covariates included in model* | *No CCI covariates included in model* |
|---|---|---|
| Age 10-19 | -0.00 (-0.01 - 0.00) | -0.00 (-0.01 - 0.00) |
| Age 20-29 | 0.02 (0.01 - 0.02) | 0.01 (0.01 - 0.02) |
| Age 30-39 | 0.01 (0.01 - 0.01) | 0.01 (0.00 - 0.01) |
| Age 40-49 | -0.01 (-0.02 - -0.01) | -0.01 (-0.01 - -0.00) |
| Age 50-59 | 0.02 (0.01 - 0.02) | 0.05 (0.05 - 0.05) |
| Age 60-69 | 0.09 (0.09 - 0.10) | 0.18 (0.18 - 0.18) |
| Age 70-79 | 0.22 (0.22 - 0.23) | 0.38 (0.38 - 0.38) |
| Age 80-89 | 0.62 (0.62 - 0.63) | 0.84 (0.83 - 0.85) |
| Age 90+ | 1.96 (1.95 - 1.97) | 2.16 (2.15 - 2.17) |
| Myocardial infarction | 0.05 (0.04 - 0.06) | |
| Diabetes without end-organ damage | 0.26 (0.25 - 0.27) | |
| Hemiplegia | 1.15 (1.13 - 1.17) | |
| Moderate to severe renal disease | 0.63 (0.62 - 0.64) | |
| Diabetes with end-organ damage | 0.38 (0.37 - 0.39) | |
| Solid cancer tumour | 0.32 (0.31 - 0.32) | |
| Leukaemia | 0.89 (0.86 - 0.91) | |
| Lymphoma | 0.96 (0.94 - 0.97) | |
| Moderate to severe liver disease | 0.70 (0.67 - 0.72) | |
| Metastatic solid tumour | 0.88 (0.86 - 0.89) | |
| AIDS | 0.10 (0.07 - 0.13) | |
| Congestive heart failure | 0.43 (0.42 - 0.44) | |
| Peripheral vascular disease | 0.27 (0.27 - 0.28) | |
| Cerebrovascular disease | 0.14 (0.14 - 0.15) | |
| Chronic pulmonary disease | 0.24 (0.24 - 0.25) | |
| Connective tissue disease | 0.23 (0.22 - 0.23) | |
| Ulcer disease | 0.31 (0.30 - 0.32) | |
| Mild liver disease | 0.24 (0.23 - 0.25) | |
| Alcohol abuse | 0.21 (0.21 - 0.22) | 0.27 (0.26 - 0.27) |
| Anxiety disorders | 0.03 (0.03 - 0.04) | 0.03 (0.03 - 0.04) |
| Bipolar disorder | 0.36 (0.35 - 0.37) | 0.36 (0.35 - 0.37) |
| Brain tumors | 0.41 (0.40 - 0.42) | 0.48 (0.47 - 0.50) |
| Cerebral palsy | 0.12 (0.10 - 0.15) | 0.37 (0.35 - 0.39) |
| Dementia | 1.86 (1.85 - 1.87) | 1.86 (1.85 - 1.87) |
| Depression | 0.14 (0.14 - 0.14) | 0.16 (0.16 - 0.17) |
| Developmental and behavioural disorders | 0.01 (0.01 - 0.02) | -0.00 (-0.01 - 0.00) |
| Drug abuse | 0.21 (0.21 - 0.22) | 0.22 (0.21 - 0.23) |
| Eating disorders | 0.41 (0.40 - 0.43) | 0.41 (0.40 - 0.43) |
| Epilepsy | 0.38 (0.38 - 0.39) | 0.42 (0.42 - 0.43) |
| Headache | 0.00 (-0.00 - 0.00) | 0.01 (0.00 - 0.01) |
| Infections of the CNS | 0.26 (0.25 - 0.28) | 0.32 (0.30 - 0.33) |

| | *CCI covariates included in model* | *No CCI covariates included in model* |
|---|---|---|
| Intellectual disability | 0.19 (0.17 - 0.20) | 0.20 (0.19 - 0.21) |
| Multiple sclerosis | 0.86 (0.84 - 0.87) | 0.86 (0.85 - 0.88) |
| Neuromuscular disorders | 0.33 (0.30 - 0.35) | 0.42 (0.40 - 0.44) |
| Other neurodegenerative disorders | 0.83 (0.80 - 0.86) | 0.89 (0.86 - 0.92) |
| Parkinson's disease | 0.66 (0.65 - 0.68) | 0.67 (0.66 - 0.68) |
| Personality disorders | 0.10 (0.09 - 0.11) | 0.09 (0.09 - 0.10) |
| Polyneuropathy | 0.28 (0.28 - 0.29) | 0.51 (0.50 - 0.52) |
| Schizophrenia spectrum disorders | 0.84 (0.83 - 0.84) | 0.83 (0.83 - 0.84) |
| Sleep disorders | 0.14 (0.14 - 0.14) | 0.18 (0.18 - 0.18) |
| Stress related disorders | 0.07 (0.06 - 0.07) | 0.07 (0.07 - 0.08) |
| Stroke | 0.51 (0.50 - 0.51) | 0.67 (0.66 - 0.67) |
| Traumatic brain injury | 0.10 (0.10 - 0.11) | 0.11 (0.11 - 0.12) |
| Male | -0.04 (-0.04 - -0.04) | -0.03 (-0.03 - -0.02) |

| | *CCI covariates included in model* | *No CCI covariates included in model* |
|---|---|---|
| Age 10-19 | -0.00 (-0.00 - 0.00) | -0.00 (-0.00 - 0.00) |
| Age 20-29 | 0.02 (0.02 - 0.02) | 0.01 (0.01 - 0.02) |
| Age 30-39 | 0.01 (0.01 - 0.01) | 0.01 (0.00 - 0.01) |
| Age 40-49 | -0.01 (-0.02 - -0.01) | -0.01 (-0.01 - -0.00) |
| Age 50-59 | 0.01 (0.01 - 0.02) | 0.05 (0.04 - 0.05) |
| Age 60-69 | 0.09 (0.09 - 0.09) | 0.17 (0.17 - 0.18) |
| Age 70-79 | 0.23 (0.22 - 0.23) | 0.38 (0.38 - 0.38) |
| Age 80-89 | 0.62 (0.62 - 0.63) | 0.83 (0.82 - 0.83) |
| Age 90+ | 1.94 (1.93 - 1.95) | 2.13 (2.12 - 2.14) |
| Myocardial infarction | 0.04 (0.04 - 0.05) | |
| Diabetes without end-organ damage | 0.28 (0.27 - 0.28) | |
| Hemiplegia | 1.22 (1.20 - 1.24) | |
| Moderate to severe renal disease | 0.62 (0.61 - 0.63) | |
| Diabetes with end-organ damage | 0.37 (0.36 - 0.38) | |
| Solid cancer tumour | 0.29 (0.29 - 0.30) | |
| Leukaemia | 0.79 (0.77 - 0.81) | |
| Lymphoma | 0.95 (0.93 - 0.97) | |
| Moderate to severe liver disease | 0.66 (0.63 - 0.69) | |
| Metastatic solid tumour | 0.89 (0.87 - 0.91) | |
| AIDS | 0.09 (0.06 - 0.12) | |
| Congestive heart failure | 0.44 (0.43 - 0.44) | |
| Peripheral vascular disease | 0.27 (0.26 - 0.28) | |
| Cerebrovascular disease | 0.07 (0.06 - 0.08) | |
| Chronic pulmonary disease | 0.24 (0.24 - 0.25) | |
| Connective tissue disease | 0.22 (0.21 - 0.23) | |
| Ulcer disease | 0.32 (0.31 - 0.33) | |
| Mild liver disease | 0.23 (0.22 - 0.24) | |
| Alcohol abuse | 0.22 (0.22 - 0.23) | 0.28 (0.27 - 0.28) |
| Anxiety disorders | 0.03 (0.03 - 0.03) | 0.03 (0.03 - 0.04) |
| Bipolar disorder | 0.35 (0.34 - 0.36) | 0.34 (0.33 - 0.35) |
| Brain tumors | 0.49 (0.47 - 0.50) | 0.56 (0.55 - 0.58) |
| Cerebral palsy | 0.19 (0.17 - 0.21) | 0.44 (0.42 - 0.46) |
| Dementia | 2.09 (2.08 - 2.10) | 2.08 (2.07 - 2.09) |
| Depression | 0.14 (0.14 - 0.14) | 0.16 (0.16 - 0.17) |
| Developmental and behavioural disorders | 0.03 (0.02 - 0.03) | 0.01 (0.01 - 0.02) |
| Drug abuse | 0.19 (0.18 - 0.20) | 0.20 (0.19 - 0.21) |
| Eating disorders | 0.47 (0.46 - 0.49) | 0.47 (0.46 - 0.49) |
| Epilepsy | 0.36 (0.36 - 0.37) | 0.40 (0.39 - 0.41) |
| Headache | -0.00 (-0.01 - -0.00) | 0.00 (-0.00 - 0.00) |
| Infections of the CNS | 0.29 (0.27 - 0.30) | 0.35 (0.33 - 0.36) |

| | *CCI covariates included in model* | *No CCI covariates included in model* |
|---|---|---|
| Intellectual disability | 0.22 (0.21 - 0.24) | 0.24 (0.22 - 0.25) |
| Multiple sclerosis | 0.91 (0.90 - 0.93) | 0.92 (0.90 - 0.93) |
| Neuromuscular disorders | 0.32 (0.30 - 0.34) | 0.42 (0.40 - 0.45) |
| Other neurodegenerative disorders | 0.99 (0.96 - 1.02) | 1.06 (1.02 - 1.09) |
| Parkinson’s disease | 0.74 (0.73 - 0.76) | 0.75 (0.73 - 0.76) |
| Personality disorders | 0.09 (0.09 - 0.10) | 0.09 (0.08 - 0.10) |
| Polyneuropathy | 0.30 (0.29 - 0.31) | 0.53 (0.52 - 0.54) |
| Schizophrenia spectrum disorders | 0.87 (0.86 - 0.88) | 0.87 (0.86 - 0.88) |
| Sleep disorders | 0.15 (0.15 - 0.15) | 0.19 (0.19 - 0.19) |
| Stress related disorders | 0.07 (0.07 - 0.08) | 0.08 (0.07 - 0.08) |
| Stroke | 0.60 (0.59 - 0.61) | 0.72 (0.72 - 0.73) |
| Traumatic brain injury | 0.12 (0.11 - 0.12) | 0.13 (0.12 - 0.13) |
| Male | -0.04 (-0.04 - -0.04) | -0.03 (-0.03 - -0.03) |

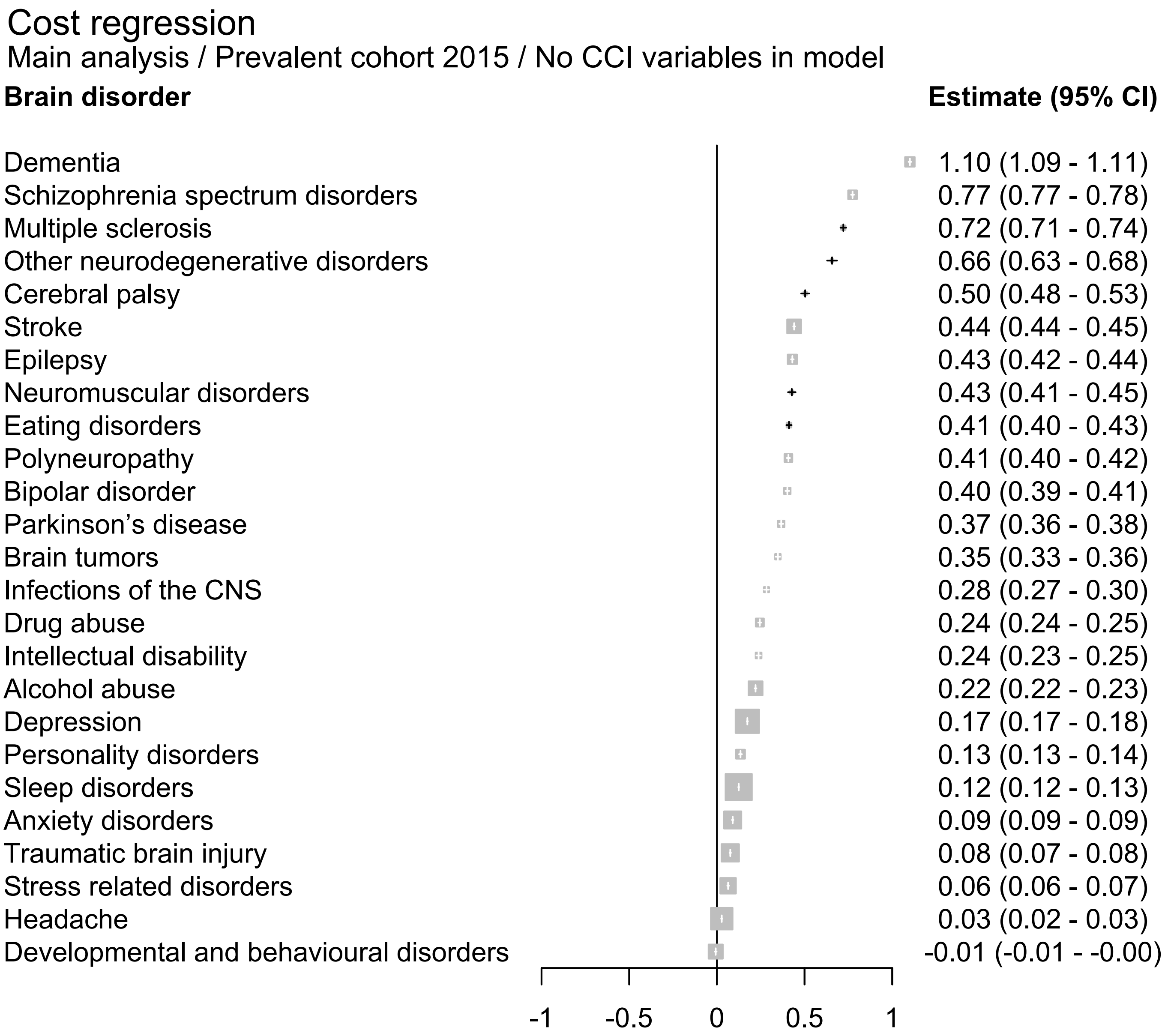
Cost regression
Main analysis / Prevalent cohort 2015 / No CCI variables in model
Brain disorder
Estimate (95% CI)
Dementia 1.10 (1.09 - 1.11)
Schizophrenia spectrum disorders 0.77 (0.77 - 0.78)
Multiple sclerosis 0.72 (0.71 - 0.74)
Other neurodegenerative disorders 0.66 (0.63 - 0.68)
Cerebral palsy 0.50 (0.48 - 0.53)
Stroke 0.44 (0.44 - 0.45)
Epilepsy 0.43 (0.42 - 0.44)
Neuromuscular disorders 0.43 (0.41 - 0.45)
Eating disorders 0.41 (0.40 - 0.43)
Polyneuropathy 0.41 (0.40 - 0.42)
Bipolar disorder 0.40 (0.39 - 0.41)
Parkinson's disease 0.37 (0.36 - 0.38)
Brain tumors 0.35 (0.33 - 0.36)
Infections of the CNS 0.28 (0.27 - 0.30)
Drug abuse 0.24 (0.24 - 0.25)
Intellectual disability 0.24 (0.23 - 0.25)
Alcohol abuse 0.22 (0.22 - 0.23)
Depression 0.17 (0.17 - 0.18)
Personality disorders 0.13 (0.13 - 0.14)
Sleep disorders 0.12 (0.12 - 0.13)
Anxiety disorders 0.09 (0.09 - 0.09)
Traumatic brain injury 0.08 (0.07 - 0.08)
Stress related disorders 0.06 (0.06 - 0.07)
Headache 0.03 (0.02 - 0.03)
Developmental and behavioural disorders -0.01 (-0.01 - -0.00)
-1
-0.5
0
0.5
1

### Cost regression

Main analysis / Prevalent cohort 2015 / CCI variables included in model

| **Brain disorder** | **Estimate (95% CI)** |
|---|---|
| Dementia | 1.10 (1.09 - 1.11) |
| Schizophrenia spectrum disorders | 0.78 (0.77 - 0.79) |
| Multiple sclerosis | 0.72 (0.70 - 0.73) |
| Other neurodegenerative disorders | 0.62 (0.59 - 0.65) |
| Eating disorders | 0.41 (0.40 - 0.43) |
| Bipolar disorder | 0.41 (0.40 - 0.42) |
| Epilepsy | 0.38 (0.37 - 0.38) |
| Parkinson’s disease | 0.36 (0.35 - 0.37) |
| Neuromuscular disorders | 0.33 (0.31 - 0.35) |
| Brain tumors | 0.29 (0.28 - 0.31) |
| Cerebral palsy | 0.24 (0.22 - 0.26) |
| Drug abuse | 0.23 (0.22 - 0.24) |
| Infections of the CNS | 0.23 (0.21 - 0.24) |
| Intellectual disability | 0.23 (0.21 - 0.24) |
| Polyneuropathy | 0.21 (0.21 - 0.22) |
| Stroke | 0.21 (0.21 - 0.22) |
| Alcohol abuse | 0.17 (0.16 - 0.17) |
| Depression | 0.14 (0.14 - 0.15) |
| Personality disorders | 0.14 (0.14 - 0.15) |
| Sleep disorders | 0.09 (0.08 - 0.09) |
| Anxiety disorders | 0.08 (0.08 - 0.09) |
| Traumatic brain injury | 0.07 (0.06 - 0.07) |
| Stress related disorders | 0.06 (0.06 - 0.07) |
| Headache | 0.02 (0.01 - 0.02) |
| Developmental and behavioural disorders | 0.01 (0.00 - 0.01) |

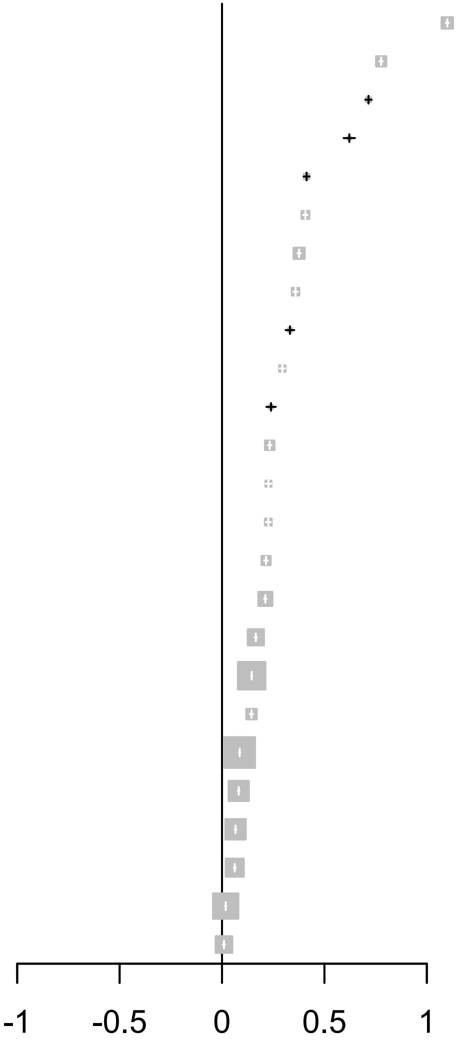

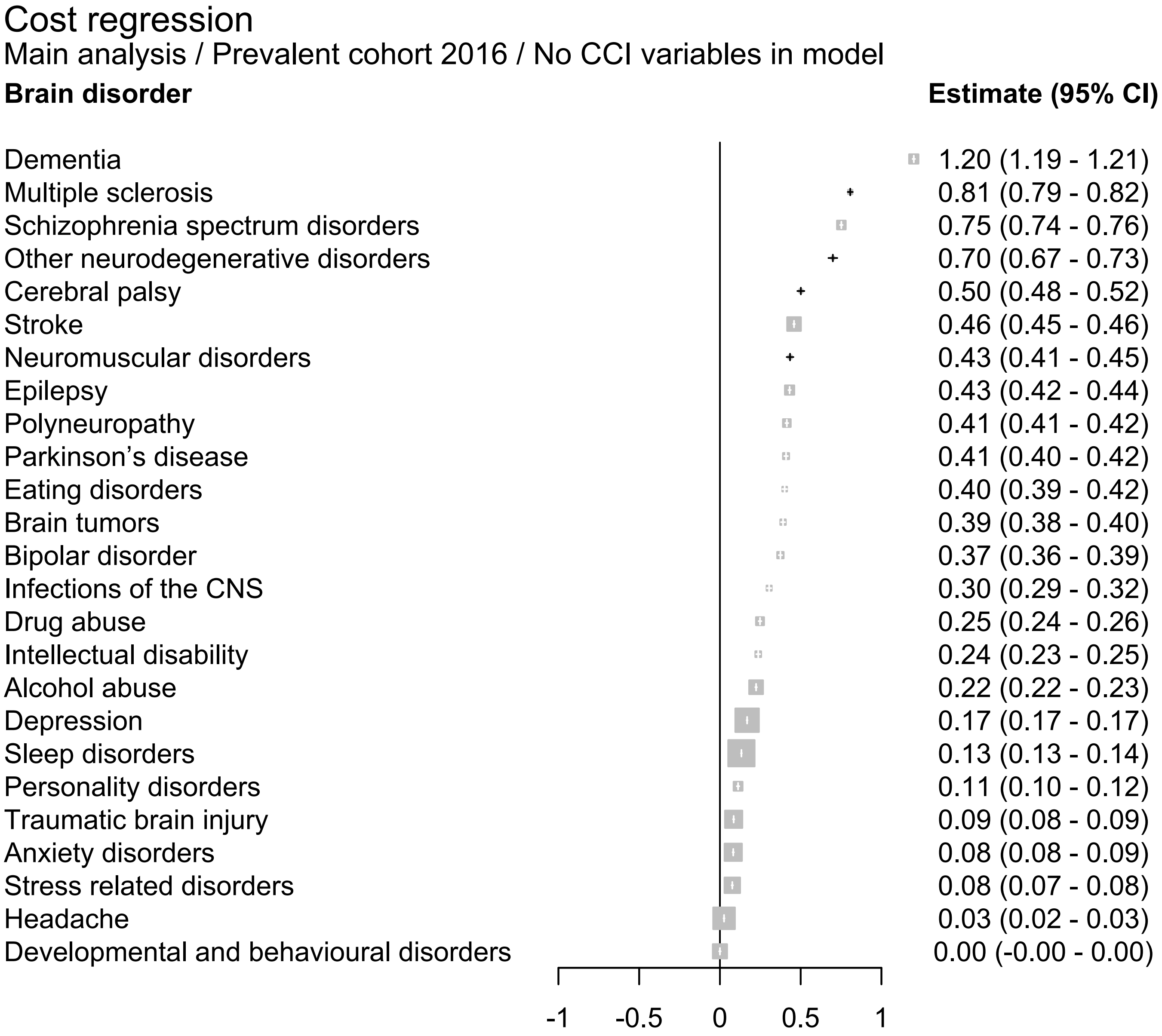
Cost regression
Main analysis / Prevalent cohort 2016 / No CCI variables in model
Brain disorder
Estimate (95% CI)
Dementia 1.20 (1.19 - 1.21)
Multiple sclerosis 0.81 (0.79 - 0.82)
Schizophrenia spectrum disorders 0.75 (0.74 - 0.76)
Other neurodegenerative disorders 0.70 (0.67 - 0.73)
Cerebral palsy 0.50 (0.48 - 0.52)
Stroke 0.46 (0.45 - 0.46)
Neuromuscular disorders 0.43 (0.41 - 0.45)
Epilepsy 0.43 (0.42 - 0.44)
Polyneuropathy 0.41 (0.41 - 0.42)
Parkinson’s disease 0.41 (0.40 - 0.42)
Eating disorders 0.40 (0.39 - 0.42)
Brain tumors 0.39 (0.38 - 0.40)
Bipolar disorder 0.37 (0.36 - 0.39)
Infections of the CNS 0.30 (0.29 - 0.32)
Drug abuse 0.25 (0.24 - 0.26)
Intellectual disability 0.24 (0.23 - 0.25)
Alcohol abuse 0.22 (0.22 - 0.23)
Depression 0.17 (0.17 - 0.17)
Sleep disorders 0.13 (0.13 - 0.14)
Personality disorders 0.11 (0.10 - 0.12)
Traumatic brain injury 0.09 (0.08 - 0.09)
Anxiety disorders 0.08 (0.08 - 0.09)
Stress related disorders 0.08 (0.07 - 0.08)
Headache 0.03 (0.02 - 0.03)
Developmental and behavioural disorders 0.00 (-0.00 - 0.00)
-1
-0.5
0
0.5
1

#### Cost regression

Main analysis / Prevalent cohort 2016 / CCI variables included in model

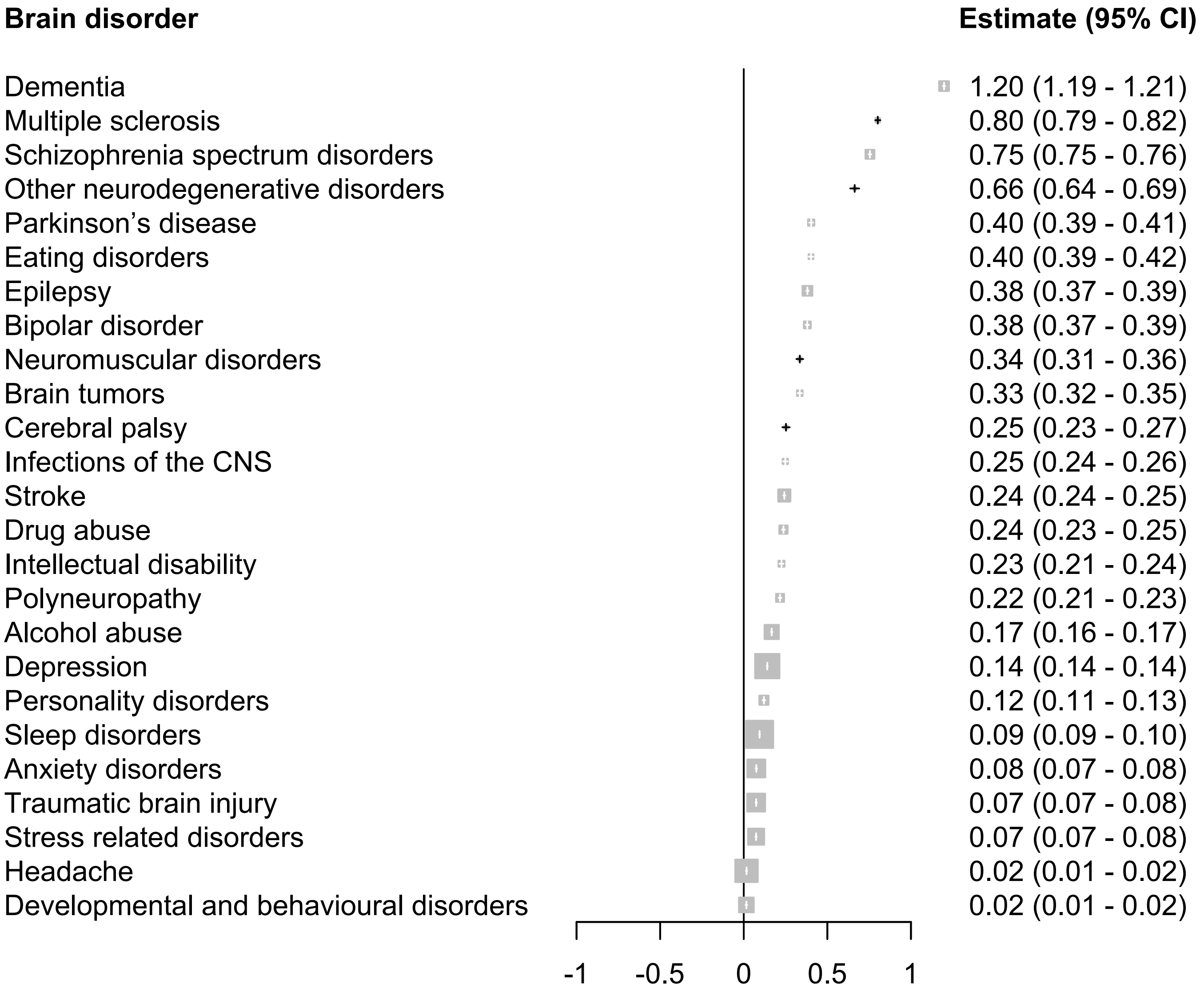

### Cost regression

#### Main analysis / Prevalent cohort 2017 / No CCI variables in model

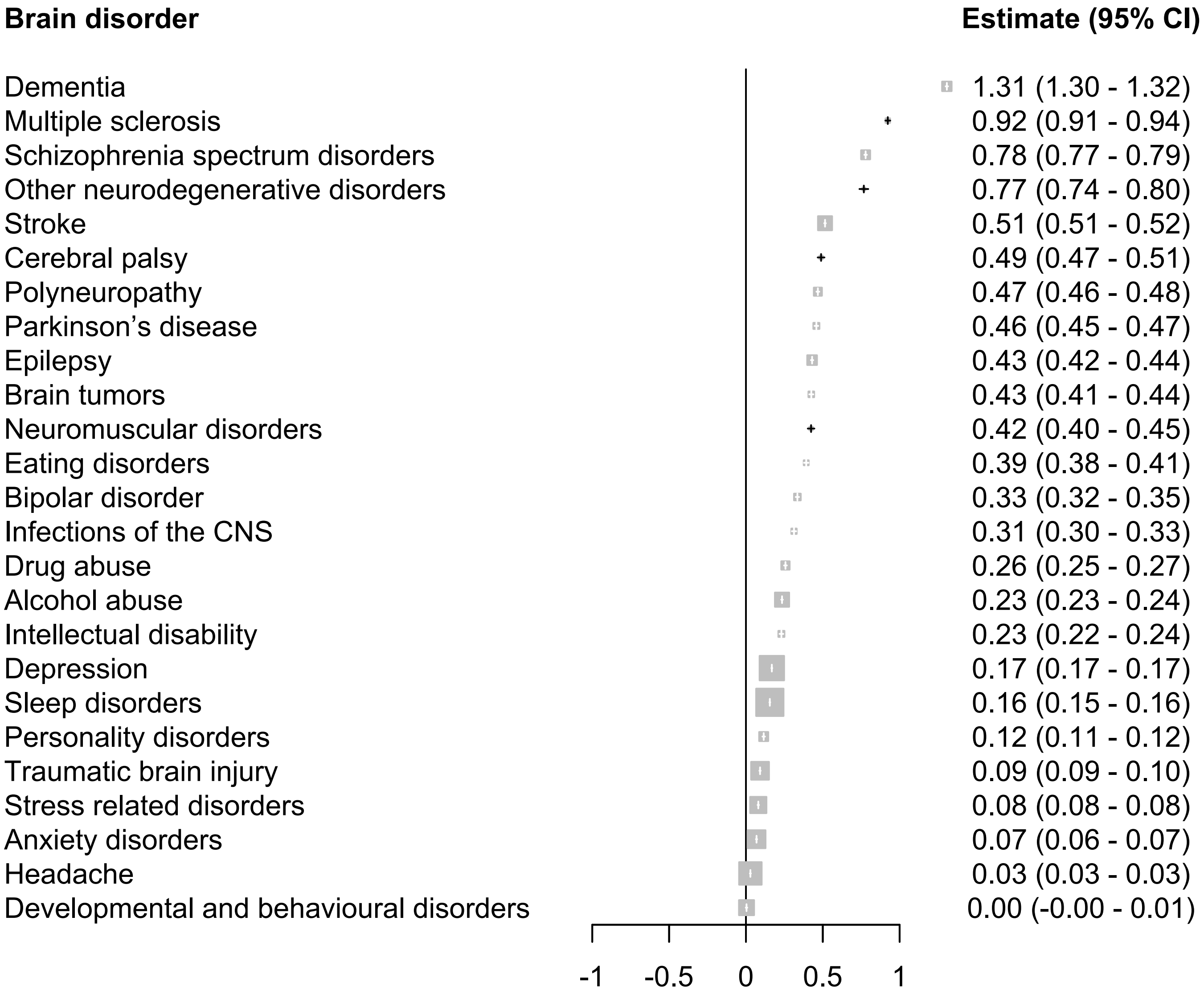

### Cost regression

Main analysis / Prevalent cohort 2017 / CCI variables included in model

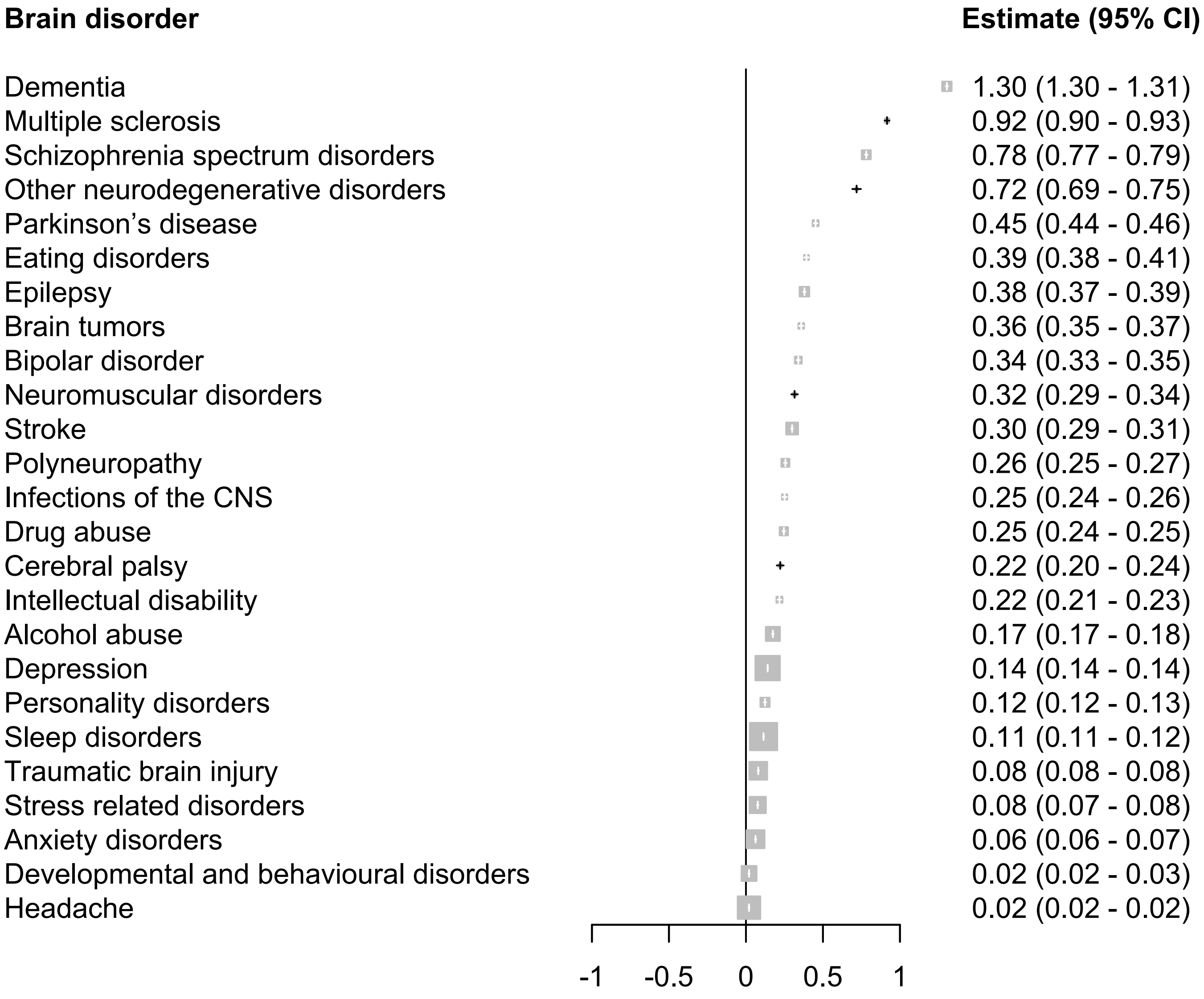

#### Cost regression

Main analysis / Prevalent cohort 2018 / No CCI variables in model

| **Brain disorder** | **Estimate (95% CI)** |
|---|---|
| Dementia | 1.46 (1.45 - 1.47) |
| Other neurodegenerative disorders | 0.77 (0.74 - 0.80) |
| Schizophrenia spectrum disorders | 0.69 (0.68 - 0.70) |
| Stroke | 0.52 (0.51 - 0.52) |
| Parkinson’s disease | 0.49 (0.47 - 0.50) |
| Multiple sclerosis | 0.48 (0.47 - 0.50) |
| Epilepsy | 0.37 (0.37 - 0.38) |
| Cerebral palsy | 0.37 (0.35 - 0.39) |
| Brain tumors | 0.37 (0.35 - 0.38) |
| Polyneuropathy | 0.35 (0.34 - 0.36) |
| Neuromuscular disorders | 0.33 (0.31 - 0.34) |
| Eating disorders | 0.32 (0.30 - 0.33) |
| Bipolar disorder | 0.31 (0.30 - 0.32) |
| Infections of the CNS | 0.30 (0.28 - 0.31) |
| Alcohol abuse | 0.25 (0.24 - 0.25) |
| Drug abuse | 0.21 (0.20 - 0.22) |
| Intellectual disability | 0.18 (0.17 - 0.19) |
| Depression | 0.16 (0.16 - 0.16) |
| Sleep disorders | 0.12 (0.12 - 0.12) |
| Traumatic brain injury | 0.09 (0.09 - 0.09) |
| Personality disorders | 0.09 (0.08 - 0.09) |
| Stress related disorders | 0.08 (0.07 - 0.08) |
| Anxiety disorders | 0.05 (0.05 - 0.06) |
| Developmental and behavioural disorders | 0.01 (0.01 - 0.01) |
| Headache | 0.01 (0.01 - 0.01) |

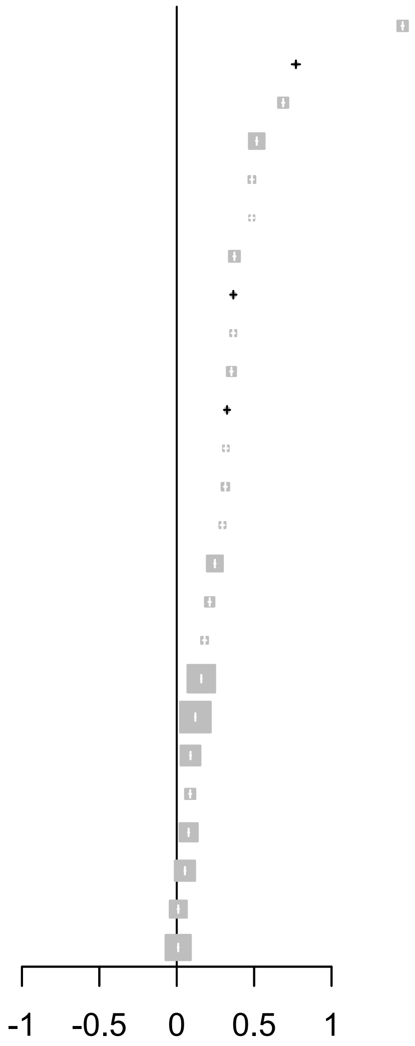

### Cost regression

Main analysis / Prevalent cohort 2018 / CCI variables included in model

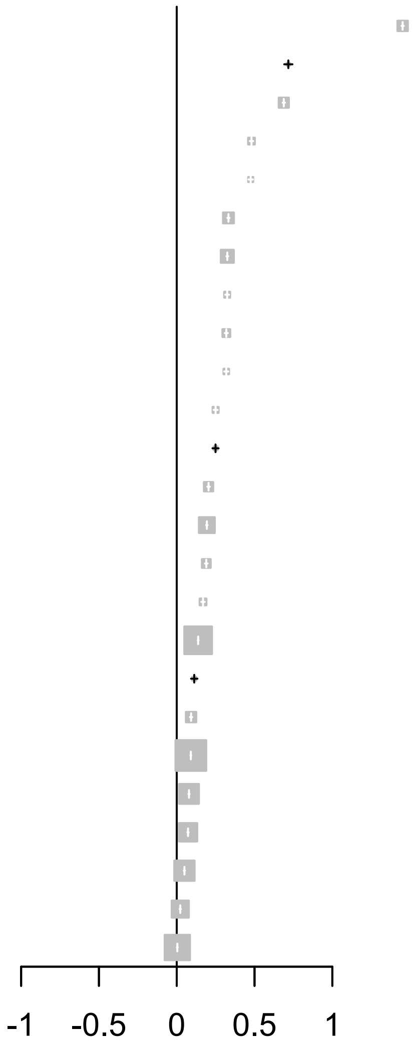


| Brain disorder | Estimate (95% CI) |
|---|---|
| Dementia | 1.45 (1.44 - 1.46) |
| Other neurodegenerative disorders | 0.72 (0.69 - 0.74) |
| Schizophrenia spectrum disorders | 0.69 (0.68 - 0.70) |
| Parkinson’s disease | 0.48 (0.47 - 0.49) |
| Multiple sclerosis | 0.48 (0.46 - 0.49) |
| Epilepsy | 0.33 (0.33 - 0.34) |
| Stroke | 0.32 (0.32 - 0.33) |
| Brain tumors | 0.32 (0.31 - 0.34) |
| Bipolar disorder | 0.32 (0.31 - 0.33) |
| Eating disorders | 0.32 (0.31 - 0.33) |
| Infections of the CNS | 0.25 (0.24 - 0.26) |
| Neuromuscular disorders | 0.25 (0.23 - 0.27) |
| Drug abuse | 0.20 (0.20 - 0.21) |
| Alcohol abuse | 0.19 (0.19 - 0.20) |
| Polyneuropathy | 0.19 (0.18 - 0.20) |
| Intellectual disability | 0.17 (0.16 - 0.18) |
| Depression | 0.14 (0.13 - 0.14) |
| Cerebral palsy | 0.11 (0.09 - 0.13) |
| Personality disorders | 0.09 (0.08 - 0.10) |
| Sleep disorders | 0.09 (0.09 - 0.09) |
| Traumatic brain injury | 0.08 (0.07 - 0.08) |
| Stress related disorders | 0.07 (0.07 - 0.08) |
| Anxiety disorders | 0.05 (0.05 - 0.05) |
| Developmental and behavioural disorders | 0.02 (0.02 - 0.03) |
| Headache | 0.00 (0.00 - 0.01) |

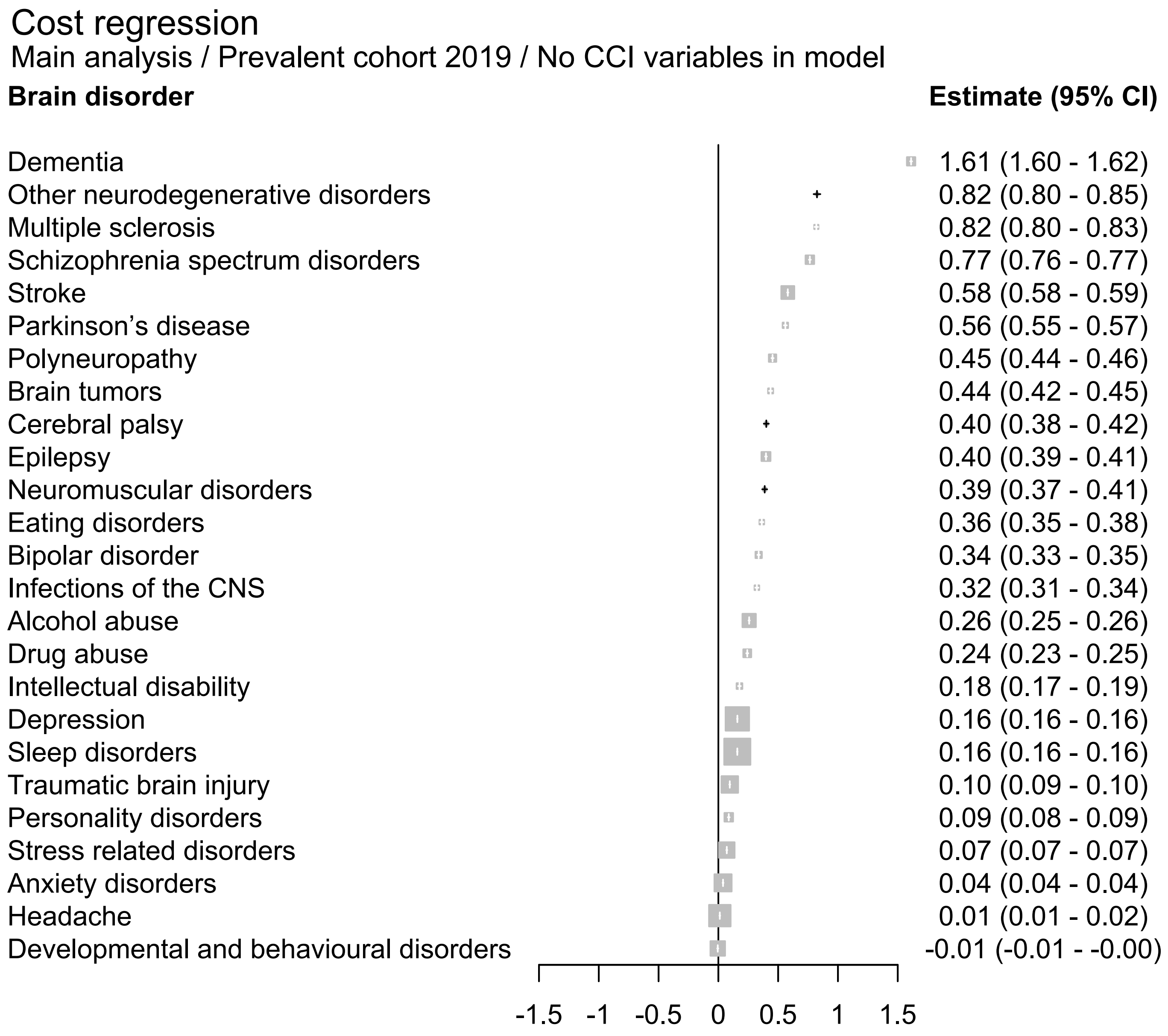
Cost regression
Main analysis / Prevalent cohort 2019 / No CCI variables in model
Brain disorder
Estimate (95% CI)
Dementia 1.61 (1.60 - 1.62)
Other neurodegenerative disorders 0.82 (0.80 - 0.85)
Multiple sclerosis 0.82 (0.80 - 0.83)
Schizophrenia spectrum disorders 0.77 (0.76 - 0.77)
Stroke 0.58 (0.58 - 0.59)
Parkinson’s disease 0.56 (0.55 - 0.57)
Polyneuropathy 0.45 (0.44 - 0.46)
Brain tumors 0.44 (0.42 - 0.45)
Cerebral palsy 0.40 (0.38 - 0.42)
Epilepsy 0.40 (0.39 - 0.41)
Neuromuscular disorders 0.39 (0.37 - 0.41)
Eating disorders 0.36 (0.35 - 0.38)
Bipolar disorder 0.34 (0.33 - 0.35)
Infections of the CNS 0.32 (0.31 - 0.34)
Alcohol abuse 0.26 (0.25 - 0.26)
Drug abuse 0.24 (0.23 - 0.25)
Intellectual disability 0.18 (0.17 - 0.19)
Depression 0.16 (0.16 - 0.16)
Sleep disorders 0.16 (0.16 - 0.16)
Traumatic brain injury 0.10 (0.09 - 0.10)
Personality disorders 0.09 (0.08 - 0.09)
Stress related disorders 0.07 (0.07 - 0.07)
Anxiety disorders 0.04 (0.04 - 0.04)
Headache 0.01 (0.01 - 0.02)
Developmental and behavioural disorders -0.01 (-0.01 - -0.00)
-1.5 -1 -0.5 0 0.5 1 1.5

### Cost regression

Main analysis / Prevalent cohort 2019 / CCI variables included in model

| Brain disorder | Estimate (95% CI) |
|---|---|
| Dementia | 1.61 (1.60 - 1.62) |
| Multiple sclerosis | 0.81 (0.80 - 0.83) |
| Other neurodegenerative disorders | 0.77 (0.74 - 0.80) |
| Schizophrenia spectrum disorders | 0.77 (0.76 - 0.77) |
| Parkinson’s disease | 0.56 (0.54 - 0.57) |
| Stroke | 0.40 (0.40 - 0.41) |
| Brain tumors | 0.37 (0.36 - 0.38) |
| Eating disorders | 0.36 (0.35 - 0.38) |
| Epilepsy | 0.36 (0.35 - 0.36) |
| Bipolar disorder | 0.34 (0.33 - 0.35) |
| Neuromuscular disorders | 0.29 (0.27 - 0.31) |
| Infections of the CNS | 0.27 (0.25 - 0.28) |
| Polyneuropathy | 0.25 (0.24 - 0.26) |
| Drug abuse | 0.23 (0.23 - 0.24) |
| Alcohol abuse | 0.20 (0.20 - 0.21) |
| Intellectual disability | 0.16 (0.15 - 0.18) |
| Cerebral palsy | 0.15 (0.13 - 0.17) |
| Depression | 0.14 (0.13 - 0.14) |
| Sleep disorders | 0.12 (0.12 - 0.12) |
| Personality disorders | 0.09 (0.09 - 0.10) |
| Traumatic brain injury | 0.08 (0.08 - 0.09) |
| Stress related disorders | 0.07 (0.06 - 0.07) |
| Anxiety disorders | 0.04 (0.03 - 0.04) |
| Developmental and behavioural disorders | 0.01 (0.01 - 0.01) |
| Headache | 0.01 (0.00 - 0.01) |

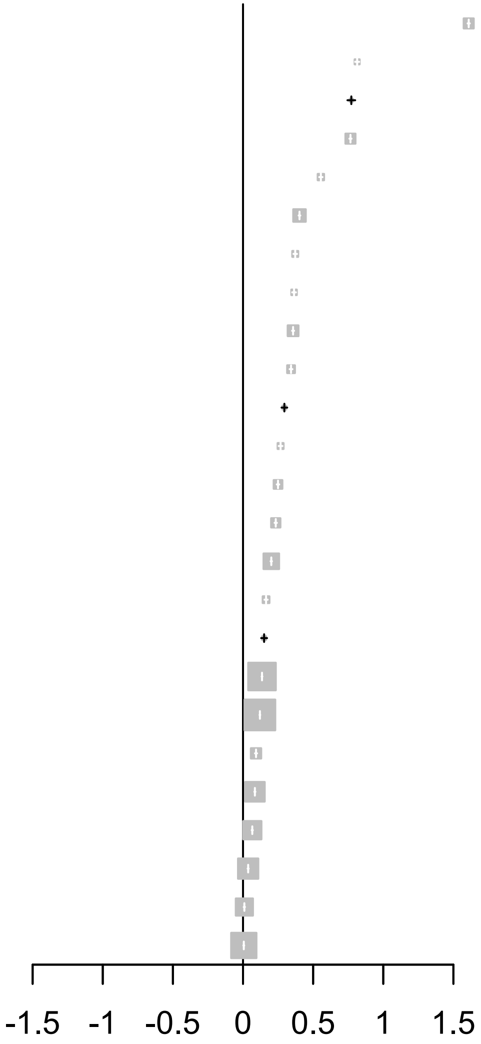

### Cost regression

Main analysis / Prevalent cohort 2020 / No CCI variables in model

| **Brain disorder** | **Estimate (95% CI)** |
|---|---|
| Dementia | 1.86 (1.85 - 1.87) |
| Other neurodegenerative disorders | 0.89 (0.86 - 0.92) |
| Multiple sclerosis | 0.86 (0.85 - 0.88) |
| Schizophrenia spectrum disorders | 0.83 (0.83 - 0.84) |
| Parkinson’s disease | 0.67 (0.66 - 0.68) |
| Stroke | 0.67 (0.66 - 0.67) |
| Polyneuropathy | 0.51 (0.50 - 0.52) |
| Brain tumors | 0.48 (0.47 - 0.50) |
| Epilepsy | 0.42 (0.42 - 0.43) |
| Neuromuscular disorders | 0.42 (0.40 - 0.44) |
| Eating disorders | 0.41 (0.40 - 0.43) |
| Cerebral palsy | 0.37 (0.35 - 0.39) |
| Bipolar disorder | 0.36 (0.35 - 0.37) |
| Infections of the CNS | 0.32 (0.30 - 0.33) |
| Alcohol abuse | 0.27 (0.26 - 0.27) |
| Drug abuse | 0.22 (0.21 - 0.23) |
| Intellectual disability | 0.20 (0.19 - 0.21) |
| Sleep disorders | 0.18 (0.18 - 0.18) |
| Depression | 0.16 (0.16 - 0.17) |
| Traumatic brain injury | 0.11 (0.11 - 0.12) |
| Personality disorders | 0.09 (0.09 - 0.10) |
| Stress related disorders | 0.07 (0.07 - 0.08) |
| Anxiety disorders | 0.03 (0.03 - 0.04) |
| Headache | 0.01 (0.00 - 0.01) |
| Developmental and behavioural disorders | -0.00 (-0.01 - 0.00) |

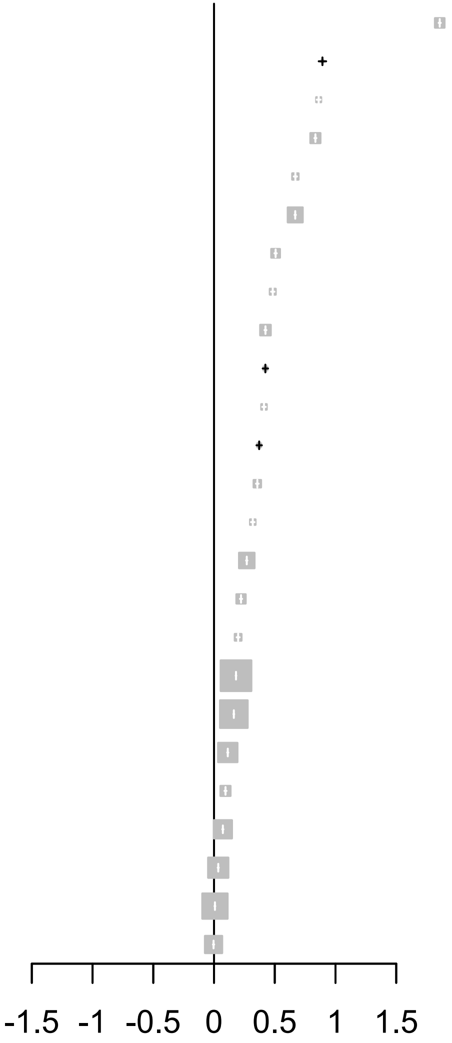

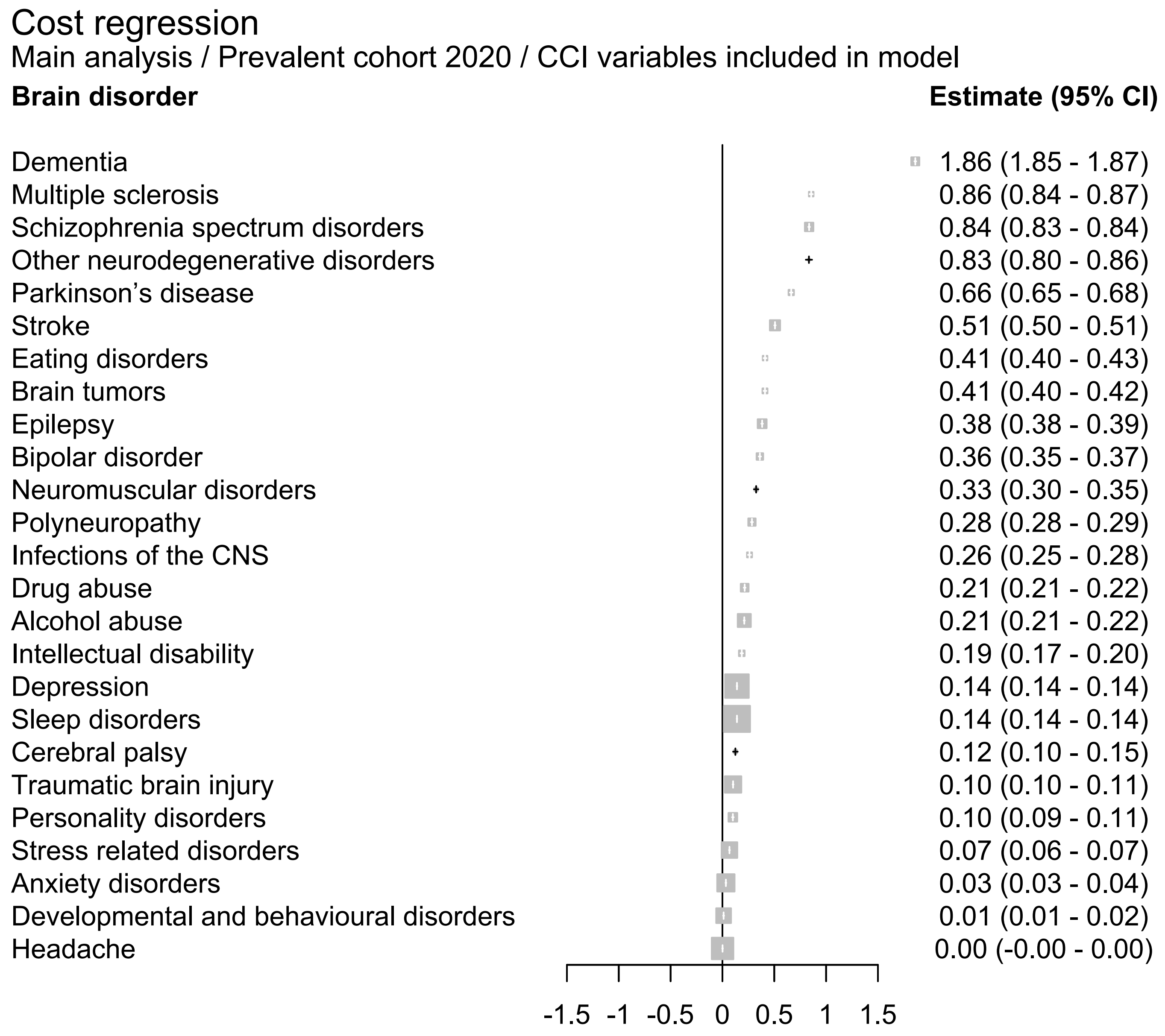
Cost regression
Main analysis / Prevalent cohort 2020 / CCI variables included in model
Brain disorder
Estimate (95% CI)
Dementia
1.86 (1.85 - 1.87)
Multiple sclerosis
0.86 (0.84 - 0.87)
Schizophrenia spectrum disorders
0.84 (0.83 - 0.84)
Other neurodegenerative disorders
0.83 (0.80 - 0.86)
Parkinson’s disease
0.66 (0.65 - 0.68)
Stroke
0.51 (0.50 - 0.51)
Eating disorders
0.41 (0.40 - 0.43)
Brain tumors
0.41 (0.40 - 0.42)
Epilepsy
0.38 (0.38 - 0.39)
Bipolar disorder
0.36 (0.35 - 0.37)
Neuromuscular disorders
0.33 (0.30 - 0.35)
Polyneuropathy
0.28 (0.28 - 0.29)
Infections of the CNS
0.26 (0.25 - 0.28)
Drug abuse
0.21 (0.21 - 0.22)
Alcohol abuse
0.21 (0.21 - 0.22)
Intellectual disability
0.19 (0.17 - 0.20)
Depression
0.14 (0.14 - 0.14)
Sleep disorders
0.14 (0.14 - 0.14)
Cerebral palsy
0.12 (0.10 - 0.15)
Traumatic brain injury
0.10 (0.10 - 0.11)
Personality disorders
0.10 (0.09 - 0.11)
Stress related disorders
0.07 (0.06 - 0.07)
Anxiety disorders
0.03 (0.03 - 0.04)
Developmental and behavioural disorders
0.01 (0.01 - 0.02)
Headache
0.00 (-0.00 - 0.00)
-1.5
-1
-0.5
0
0.5
1
1.5

#### Cost regression

Main analysis / Prevalent cohort 2021 / No CCI variables in model

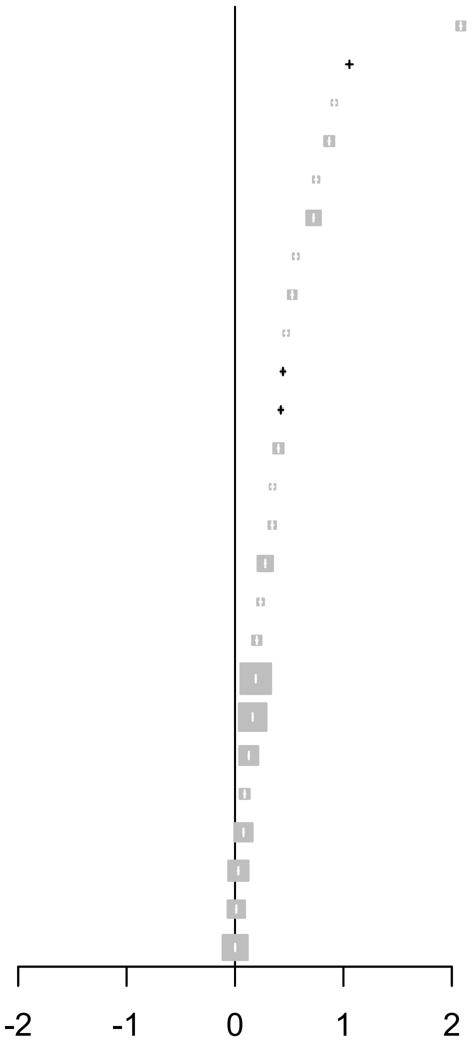


| **Brain disorder** | **Estimate (95% CI)** |
|---|---|
| Dementia | 2.08 (2.07 - 2.09) |
| Other neurodegenerative disorders | 1.06 (1.02 - 1.09) |
| Multiple sclerosis | 0.92 (0.90 - 0.93) |
| Schizophrenia spectrum disorders | 0.87 (0.86 - 0.88) |
| Parkinson’s disease | 0.75 (0.73 - 0.76) |
| Stroke | 0.72 (0.72 - 0.73) |
| Brain tumors | 0.56 (0.55 - 0.58) |
| Polyneuropathy | 0.53 (0.52 - 0.54) |
| Eating disorders | 0.47 (0.46 - 0.49) |
| Cerebral palsy | 0.44 (0.42 - 0.46) |
| Neuromuscular disorders | 0.42 (0.40 - 0.45) |
| Epilepsy | 0.40 (0.39 - 0.41) |
| Infections of the CNS | 0.35 (0.33 - 0.36) |
| Bipolar disorder | 0.34 (0.33 - 0.35) |
| Alcohol abuse | 0.28 (0.27 - 0.28) |
| Intellectual disability | 0.24 (0.22 - 0.25) |
| Drug abuse | 0.20 (0.19 - 0.21) |
| Sleep disorders | 0.19 (0.19 - 0.19) |
| Depression | 0.16 (0.16 - 0.17) |
| Traumatic brain injury | 0.13 (0.12 - 0.13) |
| Personality disorders | 0.09 (0.08 - 0.10) |
| Stress related disorders | 0.08 (0.07 - 0.08) |
| Anxiety disorders | 0.03 (0.03 - 0.04) |
| Developmental and behavioural disorders | 0.01 (0.01 - 0.02) |
| Headache | 0.00 (-0.00 - 0.00) |

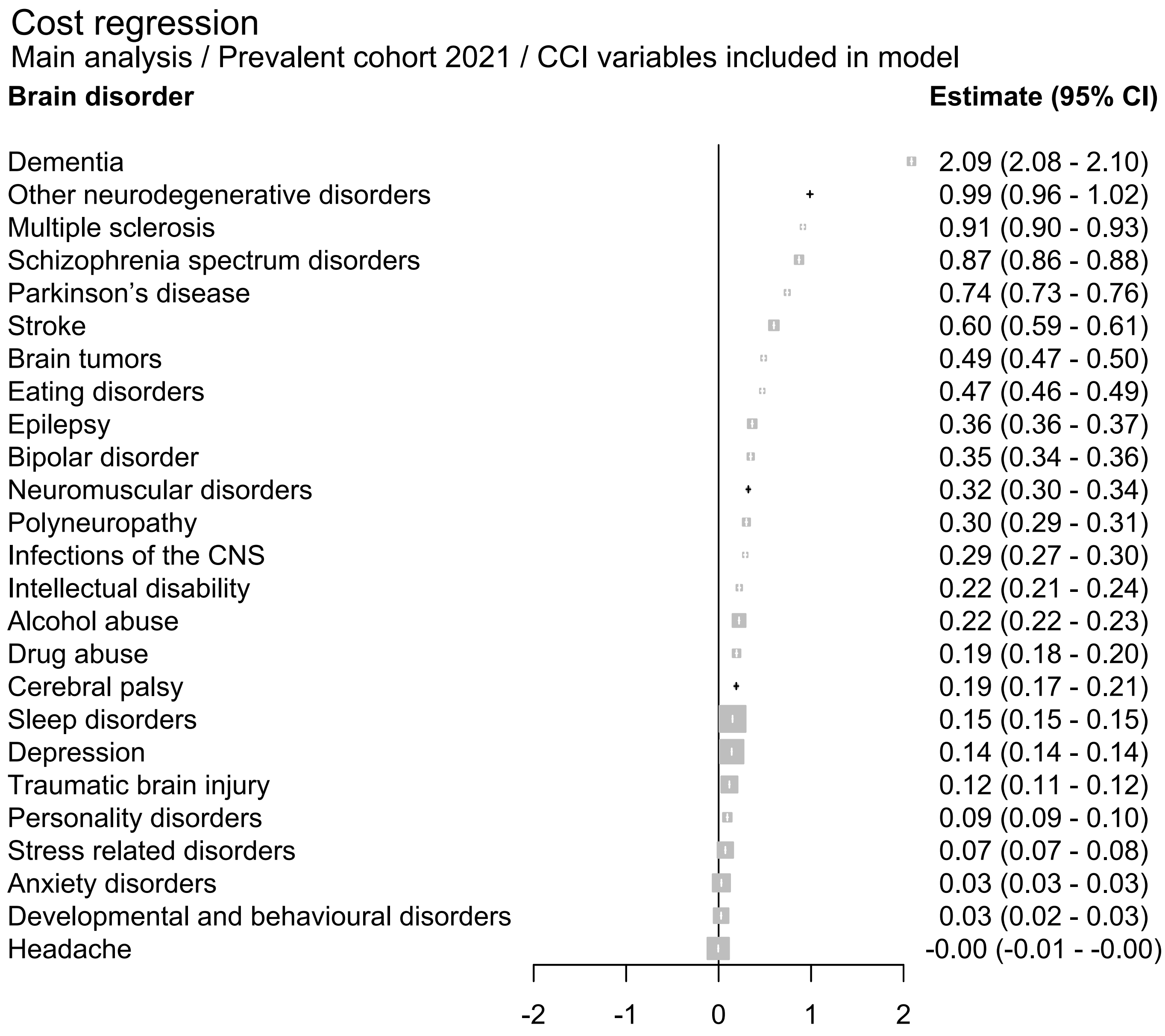
Cost regression
Main analysis / Prevalent cohort 2021 / CCI variables included in model
Brain disorder
Estimate (95% CI)
Dementia 2.09 (2.08 - 2.10)
Other neurodegenerative disorders 0.99 (0.96 - 1.02)
Multiple sclerosis 0.91 (0.90 - 0.93)
Schizophrenia spectrum disorders 0.87 (0.86 - 0.88)
Parkinson's disease 0.74 (0.73 - 0.76)
Stroke 0.60 (0.59 - 0.61)
Brain tumors 0.49 (0.47 - 0.50)
Eating disorders 0.47 (0.46 - 0.49)
Epilepsy 0.36 (0.36 - 0.37)
Bipolar disorder 0.35 (0.34 - 0.36)
Neuromuscular disorders 0.32 (0.30 - 0.34)
Polyneuropathy 0.30 (0.29 - 0.31)
Infections of the CNS 0.29 (0.27 - 0.30)
Intellectual disability 0.22 (0.21 - 0.24)
Alcohol abuse 0.22 (0.22 - 0.23)
Drug abuse 0.19 (0.18 - 0.20)
Cerebral palsy 0.19 (0.17 - 0.21)
Sleep disorders 0.15 (0.15 - 0.15)
Depression 0.14 (0.14 - 0.14)
Traumatic brain injury 0.12 (0.11 - 0.12)
Personality disorders 0.09 (0.09 - 0.10)
Stress related disorders 0.07 (0.07 - 0.08)
Anxiety disorders 0.03 (0.03 - 0.03)
Developmental and behavioural disorders 0.03 (0.02 - 0.03)
Headache -0.00 (-0.01 - -0.00)
-2
-1
0
1
2
