## Appendix 9 for "Temporal trends in the occurrence, mortality, and cost of brain disorders in Denmark"

| *Disease group* | *Persons, N* | *Men, N (%)%* | *Age, median (Q1-Q3)* | *CCI: 0, N (%)* | *CCI: 1-2, N (%)* | *CCI: +3, N (%)* |
|---|---|---|---|---|---|---|
| **Any brain disorder** | 399,064 | 197,809 (49.6) | 46.9 (22.0;68.5) | 303,420 (76.0) | 71,363 (17.9) | 24,281 (6.1) |
| **Alcohol abuse** | 37,402 | 25,507 (68.2) | 55.8 (42.7;66.7) | 25,298 (67.6) | 8,827 (23.6) | 3,277 (8.8) |
| **Anxiety disorders** | 54,013 | 20,298 (37.6) | 35.8 (22.3;56.4) | 38,479 (71.2) | 9,166 (17.0) | 6,368 (11.8) |
| **Bipolar disorder** | 9,977 | 4,118 (41.3) | 40.7 (28.5;54.9) | 7,849 (78.7) | 1,678 (16.8) | 450 (4.5) |
| **Brain tumors** | 9,739 | 4,352 (44.7) | 63.9 (50.6;72.8) | 6,034 (62.0) | 2,491 (25.6) | 1,214 (12.5) |
| **Cerebral palsy** | 2,333 | 1,301 (55.8) | 12.8 (2.5;52.7) | 1,443 (61.9) | 650 (27.9) | 240 (10.3) |
| **Dementia** | 43,711 | 17,897 (40.9) | 81.7 (75.2;86.9) | 20,079 (45.9) | 16,550 (37.9) | 7,082 (16.2) |
| **Depression** | 79,888 | 29,772 (37.3) | 45.5 (28.0;66.6) | 55,422 (69.4) | 16,880 (21.1) | 7,586 (9.5) |
| **Developmental and behavioural disorders** | 49,492 | 30,404 (61.4) | 14.1 (8.6;22.1) | 44,212 (89.3) | 4,863 (9.8) | 417 (0.8) |
| **Drug abuse** | 19,014 | 12,589 (66.2) | 26.9 (20.7;42.3) | 15,771 (82.9) | 2,563 (13.5) | 680 (3.6) |
| **Eating disorders** | 6,105 | 328 (5.4) | 19.8 (16.1;25.0) | 5,719 (93.7) | 359 (5.9) | 27 (0.4) |
| **Epilepsy** | 22,978 | 12,161 (52.9) | 52.3 (20.8;70.0) | 12,673 (55.2) | 6,964 (30.3) | 3,341 (14.5) |
| **Headache** | 38,756 | 12,818 (33.1) | 37.9 (23.8;50.4) | 31,562 (81.4) | 6,173 (15.9) | 1,021 (2.6) |
| **Infections of the CNS** | 9,641 | 5,002 (51.9) | 44.8 (22.2;64.2) | 6,973 (72.3) | 1,879 (19.5) | 789 (8.2) |
| **Intellectual disability** | 8,467 | 4,880 (57.6) | 17.3 (9.8;34.8) | 7,137 (84.3) | 1,158 (13.7) | 172 (2.0) |
| **Multiple sclerosis** | 4,124 | 1,342 (32.5) | 41.6 (32.2;51.8) | 3,488 (84.6) | 553 (13.4) | 83 (2.0) |
| **Neuromuscular disorders** | 3,373 | 1,757 (52.1) | 52.4 (33.0;67.3) | 2,087 (61.9) | 912 (27.0) | 374 (11.1) |
| **Other neurodegenerative disorders** | 1,662 | 866 (52.1) | 64.9 (52.5;72.6) | 1,018 (61.3) | 483 (29.1) | 161 (9.7) |
| **Parkinson’s disease** | 7,714 | 4,645 (60.2) | 74.6 (67.9;80.6) | 4,014 (52.0) | 2,693 (34.9) | 1,007 (13.1) |
| **Personality disorders** | 21,886 | 6,882 (31.4) | 27.3 (21.3;38.7) | 19,225 (87.8) | 2,349 (10.7) | 312 (1.4) |
| **Polyneuropathy** | 19,514 | 11,692 (59.9) | 65.5 (54.1;74.6) | 9,060 (46.4) | 6,864 (35.2) | 3,590 (18.4) |
| **Schizophrenia spectrum disorders** | 19,387 | 10,265 (52.9) | 29.8 (21.1;49.0) | 15,858 (81.8) | 2,787 (14.4) | 742 (3.8) |
| **Sleep disorders** | 42,020 | 29,409 (70.0) | 51.9 (39.8;62.2) | 29,634 (70.5) | 9,416 (22.4) | 2,970 (7.1) |
| **Stress related disorders** | 82,275 | 34,722 (42.2) | 34.7 (20.6;48.6) | 68,102 (82.8) | 11,606 (14.1) | 2,567 (3.1) |
| **Stroke** | 67,899 | 35,520 (52.3) | 71.8 (61.1;81.3) | 35,646 (52.5) | 22,412 (33.0) | 9,841 (14.5) |
| **Traumatic brain injury** | 65,287 | 35,621 (54.6) | 34.4 (15.8;63.3) | 49,977 (76.5) | 11,287 (17.3) | 4,023 (6.2) |

| *Disease group* | *Persons, N* | *Men, N (%)%* | *Age, median (Q1-Q3)* | *CCI: 0, N (%)* | *CCI: 1-2, N (%)* | *CCI: +3, N (%)* |
|---|---|---|---|---|---|---|
| **Any brain disorder** | 465,256 | 230,943 (49.6) | 47.6 (21.7;70.1) | 355,602 (76.4) | 83,181 (17.9) | 26,473 (5.7) |
| **Alcohol abuse** | 32,103 | 22,105 (68.9) | 57.8 (43.4;68.7) | 21,590 (67.3) | 7,721 (24.1) | 2,792 (8.7) |
| **Anxiety disorders** | 59,389 | 20,972 (35.3) | 31.1 (20.2;53.4) | 44,942 (75.7) | 9,630 (16.2) | 4,817 (8.1) |
| **Bipolar disorder** | 9,811 | 3,908 (39.8) | 37.7 (26.2;53.3) | 8,008 (81.6) | 1,480 (15.1) | 323 (3.3) |
| **Brain tumors** | 13,152 | 5,704 (43.4) | 65.1 (52.0;74.5) | 8,187 (62.2) | 3,516 (26.7) | 1,449 (11.0) |
| **Cerebral palsy** | 2,143 | 1,201 (56.0) | 12.8 (2.1;55.7) | 1,423 (66.4) | 514 (24.0) | 206 (9.6) |
| **Dementia** | 47,870 | 20,781 (43.4) | 80.9 (75.1;86.2) | 22,605 (47.2) | 17,893 (37.4) | 7,372 (15.4) |
| **Depression** | 73,822 | 28,176 (38.2) | 41.8 (24.4;63.9) | 54,140 (73.3) | 13,979 (18.9) | 5,703 (7.7) |
| **Developmental and behavioural disorders** | 66,975 | 39,172 (58.5) | 14.3 (9.2;22.0) | 60,531 (90.4) | 6,086 (9.1) | 358 (0.5) |
| **Drug abuse** | 20,930 | 14,153 (67.6) | 27.2 (21.2;40.4) | 17,517 (83.7) | 2,677 (12.8) | 736 (3.5) |
| **Eating disorders** | 6,916 | 427 (6.2) | 18.8 (15.2;24.3) | 6,427 (92.9) | 469 (6.8) | 20 (0.3) |
| **Epilepsy** | 22,387 | 12,074 (53.9) | 54.9 (19.6;72.5) | 12,340 (55.1) | 6,816 (30.4) | 3,231 (14.4) |
| **Headache** | 50,444 | 16,172 (32.1) | 37.6 (23.6;51.5) | 41,276 (81.8) | 7,908 (15.7) | 1,260 (2.5) |
| **Infections of the CNS** | 13,556 | 6,974 (51.4) | 45.1 (21.7;64.5) | 10,173 (75.0) | 2,465 (18.2) | 918 (6.8) |
| **Intellectual disability** | 7,689 | 4,564 (59.4) | 17.1 (8.7;31.7) | 6,572 (85.5) | 958 (12.5) | 159 (2.1) |
| **Multiple sclerosis** | 4,745 | 1,608 (33.9) | 41.3 (30.9;52.3) | 4,067 (85.7) | 571 (12.0) | 107 (2.3) |
| **Neuromuscular disorders** | 3,324 | 1,695 (51.0) | 54.9 (34.6;70.2) | 2,099 (63.1) | 887 (26.7) | 338 (10.2) |
| **Other neurodegenerative disorders** | 2,202 | 1,142 (51.9) | 67.3 (56.2;74.1) | 1,315 (59.7) | 706 (32.1) | 181 (8.2) |
| **Parkinson's disease** | 9,095 | 5,661 (62.2) | 75.1 (68.7;80.6) | 4,933 (54.2) | 3,013 (33.1) | 1,149 (12.6) |
| **Personality disorders** | 19,657 | 5,284 (26.9) | 26.6 (21.3;36.7) | 17,469 (88.9) | 1,973 (10.0) | 215 (1.1) |
| **Polyneuropathy** | 24,544 | 14,753 (60.1) | 67.7 (56.2;75.8) | 9,652 (39.3) | 9,049 (36.9) | 5,843 (23.8) |
| **Schizophrenia spectrum disorders** | 23,479 | 11,937 (50.8) | 27.0 (20.3;44.6) | 19,841 (84.5) | 2,954 (12.6) | 684 (2.9) |
| **Sleep disorders** | 61,506 | 40,598 (66.0) | 53.1 (40.9;63.4) | 42,965 (69.9) | 14,604 (23.7) | 3,937 (6.4) |
| **Stress related disorders** | 95,899 | 38,924 (40.6) | 32.2 (19.0;48.7) | 80,587 (84.0) | 12,702 (13.2) | 2,610 (2.7) |
| **Stroke** | 84,569 | 46,023 (54.4) | 72.9 (62.5;81.1) | 44,879 (53.1) | 27,405 (32.4) | 12,285 (14.5) |
| **Traumatic brain injury** | 78,140 | 41,290 (52.8) | 40.4 (17.6;70.3) | 56,927 (72.9) | 15,207 (19.5) | 6,006 (7.7) |

| *Disease group* | *Persons, N* | *Men, N (%)%* | *Age, median (Q1-Q3)* | *CCI: 0, N (%)* | *CCI: 1-2, N (%)* | *CCI: +3, N (%)* |
|---|---|---|---|---|---|---|
| **Any brain disorder** | 1,096,396 | 549,593 (50.1) | 46.1 (27.6;63.6) | 765,466 (69.8) | 248,408 (22.7) | 82,522 (7.5) |
| **Alcohol abuse** | 101,290 | 67,548 (66.7) | 56.4 (46.2;65.8) | 55,742 (55.0) | 30,396 (30.0) | 15,152 (15.0) |
| **Anxiety disorders** | 97,699 | 34,072 (34.9) | 41.2 (28.8;54.2) | 75,679 (77.5) | 17,304 (17.7) | 4,716 (4.8) |
| **Bipolar disorder** | 23,553 | 9,438 (40.1) | 52.2 (39.2;64.8) | 16,137 (68.5) | 5,551 (23.6) | 1,865 (7.9) |
| **Brain tumors** | 15,207 | 6,212 (40.8) | 62.6 (48.6;71.8) | 8,018 (52.7) | 4,842 (31.8) | 2,347 (15.4) |
| **Cerebral palsy** | 9,307 | 5,220 (56.1) | 22.1 (13.6;36.9) | 5,711 (61.4) | 2,809 (30.2) | 787 (8.5) |
| **Dementia** | 40,515 | 15,877 (39.2) | 81.6 (73.8;87.4) | 2,008 (5.0) | 26,081 (64.4) | 12,426 (30.7) |
| **Depression** | 182,108 | 64,687 (35.5) | 49.0 (35.6;64.0) | 124,296 (68.3) | 42,599 (23.4) | 15,213 (8.4) |
| **Developmental and behavioural disorders** | 104,812 | 69,744 (66.5) | 19.5 (14.2;25.9) | 94,179 (89.9) | 9,864 (9.4) | 769 (0.7) |
| **Drug abuse** | 50,253 | 32,451 (64.6) | 38.6 (28.4;50.8) | 36,698 (73.0) | 10,572 (21.0) | 2,983 (5.9) |
| **Eating disorders** | 16,080 | 760 (4.7) | 29.5 (23.0;36.9) | 14,238 (88.5) | 1,657 (10.3) | 185 (1.2) |
| **Epilepsy** | 73,576 | 37,737 (51.3) | 43.6 (24.7;62.1) | 49,052 (66.7) | 17,797 (24.2) | 6,727 (9.1) |
| **Headache** | 99,679 | 33,490 (33.6) | 45.3 (31.3;57.4) | 75,048 (75.3) | 20,215 (20.3) | 4,416 (4.4) |
| **Infections of the CNS** | 27,039 | 14,198 (52.5) | 43.2 (22.9;61.5) | 19,944 (73.8) | 5,276 (19.5) | 1,819 (6.7) |
| **Intellectual disability** | 27,861 | 16,256 (58.3) | 25.3 (17.0;45.0) | 22,449 (80.6) | 4,614 (16.6) | 798 (2.9) |
| **Multiple sclerosis** | 15,023 | 4,692 (31.2) | 52.2 (42.5;62.2) | 11,401 (75.9) | 2,959 (19.7) | 663 (4.4) |
| **Neuromuscular disorders** | 7,390 | 3,809 (51.5) | 51.2 (33.4;66.5) | 4,177 (56.5) | 2,279 (30.8) | 934 (12.6) |
| **Other neurodegenerative disorders** | 2,321 | 1,220 (52.6) | 59.7 (46.0;70.4) | 1,293 (55.7) | 764 (32.9) | 264 (11.4) |
| **Parkinson's disease** | 9,973 | 5,704 (57.2) | 74.8 (67.9;81.0) | 4,599 (46.1) | 3,809 (38.2) | 1,565 (15.7) |
| **Personality disorders** | 71,762 | 24,690 (34.4) | 39.9 (30.6;51.0) | 56,496 (78.7) | 12,594 (17.5) | 2,672 (3.7) |
| **Polyneuropathy** | 34,186 | 19,786 (57.9) | 66.5 (54.9;75.6) | 14,600 (42.7) | 12,045 (35.2) | 7,541 (22.1) |
| **Schizophrenia spectrum disorders** | 61,455 | 32,714 (53.2) | 45.1 (32.1;57.7) | 46,016 (74.9) | 12,155 (19.8) | 3,284 (5.3) |
| **Sleep disorders** | 75,151 | 54,958 (73.1) | 55.9 (43.9;66.0) | 49,143 (65.4) | 19,362 (25.8) | 6,646 (8.8) |
| **Stress related disorders** | 197,765 | 78,197 (39.5) | 42.3 (29.3;54.1) | 153,644 (77.7) | 35,710 (18.1) | 8,411 (4.3) |
| **Stroke** | 120,402 | 64,266 (53.4) | 70.9 (61.1;79.9) | 15,635 (13.0) | 71,213 (59.1) | 33,554 (27.9) |
| **Traumatic brain injury** | 250,352 | 143,335 (57.3) | 34.9 (22.2;53.3) | 201,928 (80.7) | 38,281 (15.3) | 10,143 (4.1) |

| *Disease group* | *Persons, N* | *Men, N (%)%* | *Age, median (Q1-Q3)* | *CCI: 0, N (%)* | *CCI: 1-2, N (%)* | *CCI: +3, N (%)* |
|---|---|---|---|---|---|---|
| **Any brain disorder** | 1,128,158 | 564,622 (50.0) | 46.3 (27.7;63.7) | 785,756 (69.6) | 256,039 (22.7) | 86,363 (7.7) |
| **Alcohol abuse** | 101,610 | 67,719 (66.6) | 56.9 (46.4;66.3) | 55,327 (54.5) | 30,720 (30.2) | 15,563 (15.3) |
| **Anxiety disorders** | 105,567 | 36,919 (35.0) | 40.9 (28.4;54.0) | 81,636 (77.3) | 18,740 (17.8) | 5,191 (4.9) |
| **Bipolar disorder** | 24,593 | 9,820 (39.9) | 51.8 (38.6;64.5) | 16,934 (68.9) | 5,724 (23.3) | 1,935 (7.9) |
| **Brain tumors** | 15,828 | 6,423 (40.6) | 62.9 (49.1;72.3) | 8,318 (52.6) | 5,076 (32.1) | 2,434 (15.4) |
| **Cerebral palsy** | 9,378 | 5,265 (56.1) | 22.6 (13.9;37.8) | 5,727 (61.1) | 2,834 (30.2) | 817 (8.7) |
| **Dementia** | 41,040 | 16,257 (39.6) | 81.6 (73.9;87.4) | 2,005 (4.9) | 26,059 (63.5) | 12,976 (31.6) |
| **Depression** | 189,551 | 67,264 (35.5) | 49.0 (35.4;63.6) | 129,849 (68.5) | 43,822 (23.1) | 15,880 (8.4) |
| **Developmental and behavioural disorders** | 113,574 | 74,875 (65.9) | 19.9 (14.4;26.4) | 102,004 (89.8) | 10,723 (9.4) | 847 (0.7) |
| **Drug abuse** | 52,126 | 33,898 (65.0) | 38.2 (28.3;50.6) | 38,308 (73.5) | 10,758 (20.6) | 3,060 (5.9) |
| **Eating disorders** | 16,915 | 802 (4.7) | 29.5 (23.0;37.1) | 14,997 (88.7) | 1,724 (10.2) | 194 (1.1) |
| **Epilepsy** | 73,759 | 37,821 (51.3) | 44.0 (25.0;62.7) | 48,838 (66.2) | 17,960 (24.3) | 6,961 (9.4) |
| **Headache** | 104,576 | 34,881 (33.4) | 45.3 (31.0;57.3) | 78,421 (75.0) | 21,448 (20.5) | 4,707 (4.5) |
| **Infections of the CNS** | 27,513 | 14,419 (52.4) | 43.8 (23.3;62.0) | 20,102 (73.1) | 5,430 (19.7) | 1,981 (7.2) |
| **Intellectual disability** | 28,590 | 16,703 (58.4) | 25.4 (17.4;44.8) | 23,050 (80.6) | 4,705 (16.5) | 835 (2.9) |
| **Multiple sclerosis** | 15,450 | 4,825 (31.2) | 52.4 (42.6;62.5) | 11,702 (75.7) | 3,024 (19.6) | 724 (4.7) |
| **Neuromuscular disorders** | 7,670 | 3,948 (51.5) | 51.6 (33.8;66.9) | 4,257 (55.5) | 2,412 (31.4) | 1,001 (13.1) |
| **Other neurodegenerative disorders** | 2,412 | 1,244 (51.6) | 60.6 (46.4;71.0) | 1,337 (55.4) | 787 (32.6) | 288 (11.9) |
| **Parkinson's disease** | 10,274 | 5,902 (57.4) | 74.9 (68.4;81.0) | 4,686 (45.6) | 3,901 (38.0) | 1,687 (16.4) |
| **Personality disorders** | 73,544 | 24,890 (33.8) | 39.7 (30.3;50.9) | 57,876 (78.7) | 12,912 (17.6) | 2,756 (3.7) |
| **Polyneuropathy** | 36,181 | 20,960 (57.9) | 66.9 (55.3;75.8) | 15,242 (42.1) | 12,743 (35.2) | 8,196 (22.7) |
| **Schizophrenia spectrum disorders** | 63,105 | 33,668 (53.4) | 44.7 (31.5;57.5) | 47,243 (74.9) | 12,437 (19.7) | 3,425 (5.4) |
| **Sleep disorders** | 83,508 | 60,601 (72.6) | 56.3 (44.2;66.3) | 53,949 (64.6) | 21,872 (26.2) | 7,687 (9.2) |
| **Stress related disorders** | 209,658 | 83,213 (39.7) | 42.4 (29.2;54.2) | 162,453 (77.5) | 38,160 (18.2) | 9,045 (4.3) |
| **Stroke** | 122,356 | 65,602 (53.6) | 71.2 (61.3;79.9) | 16,197 (13.2) | 71,758 (58.6) | 34,401 (28.1) |
| **Traumatic brain injury** | 248,060 | 141,490 (57.0) | 35.2 (22.4;54.0) | 198,925 (80.2) | 38,608 (15.6) | 10,527 (4.2) |

| *Disease group* | *Persons, N* | *Men, N (%)%* | *Age, median (Q1-Q3)* | *CCI: 0, N (%)* | *CCI: 1-2, N (%)* | *CCI: +3, N (%)* |
|---|---|---|---|---|---|---|
| **Any brain disorder** | 1,158,327 | 579,114 (50.0) | 46.5 (27.8;63.9) | 804,379 (69.4) | 263,745 (22.8) | 90,203 (7.8) |
| **Alcohol abuse** | 101,243 | 67,490 (66.7) | 57.3 (46.6;66.9) | 54,399 (53.7) | 30,904 (30.5) | 15,940 (15.7) |
| **Anxiety disorders** | 112,490 | 39,442 (35.1) | 40.7 (28.2;54.0) | 86,613 (77.0) | 20,250 (18.0) | 5,627 (5.0) |
| **Bipolar disorder** | 25,452 | 10,196 (40.1) | 51.5 (38.2;64.3) | 17,536 (68.9) | 5,918 (23.3) | 1,998 (7.9) |
| **Brain tumors** | 16,700 | 6,730 (40.3) | 63.3 (49.7;72.9) | 8,659 (51.9) | 5,379 (32.2) | 2,662 (15.9) |
| **Cerebral palsy** | 9,441 | 5,294 (56.1) | 22.8 (14.1;38.8) | 5,739 (60.8) | 2,837 (30.0) | 865 (9.2) |
| **Dementia** | 41,478 | 16,556 (39.9) | 81.5 (74.1;87.3) | 1,991 (4.8) | 26,254 (63.3) | 13,233 (31.9) |
| **Depression** | 195,527 | 69,405 (35.5) | 49.0 (35.3;63.4) | 133,966 (68.5) | 45,249 (23.1) | 16,312 (8.3) |
| **Developmental and behavioural disorders** | 122,351 | 80,013 (65.4) | 20.3 (14.7;26.9) | 109,692 (89.7) | 11,753 (9.6) | 906 (0.7) |
| **Drug abuse** | 53,651 | 35,118 (65.5) | 38.0 (28.3;50.5) | 39,570 (73.8) | 11,013 (20.5) | 3,068 (5.7) |
| **Eating disorders** | 17,512 | 851 (4.9) | 29.6 (23.2;37.4) | 15,513 (88.6) | 1,788 (10.2) | 211 (1.2) |
| **Epilepsy** | 73,679 | 37,793 (51.3) | 44.5 (25.2;63.3) | 48,563 (65.9) | 17,893 (24.3) | 7,223 (9.8) |
| **Headache** | 110,105 | 36,576 (33.2) | 45.3 (30.9;57.4) | 82,363 (74.8) | 22,725 (20.6) | 5,017 (4.6) |
| **Infections of the CNS** | 27,964 | 14,603 (52.2) | 44.5 (23.8;62.7) | 20,303 (72.6) | 5,581 (20.0) | 2,080 (7.4) |
| **Intellectual disability** | 29,377 | 17,153 (58.4) | 25.7 (17.8;44.5) | 23,629 (80.4) | 4,881 (16.6) | 867 (3.0) |
| **Multiple sclerosis** | 15,937 | 4,975 (31.2) | 52.6 (42.7;62.7) | 12,033 (75.5) | 3,143 (19.7) | 761 (4.8) |
| **Neuromuscular disorders** | 8,026 | 4,117 (51.3) | 51.9 (33.9;67.4) | 4,370 (54.4) | 2,585 (32.2) | 1,071 (13.3) |
| **Other neurodegenerative disorders** | 2,576 | 1,328 (51.6) | 61.7 (47.1;71.7) | 1,383 (53.7) | 860 (33.4) | 333 (12.9) |
| **Parkinson's disease** | 10,531 | 6,105 (58.0) | 75.1 (68.6;81.1) | 4,806 (45.6) | 3,972 (37.7) | 1,753 (16.6) |
| **Personality disorders** | 74,392 | 24,887 (33.5) | 39.6 (30.3;50.7) | 58,485 (78.6) | 13,102 (17.6) | 2,805 (3.8) |
| **Polyneuropathy** | 37,911 | 21,891 (57.7) | 67.4 (55.7;76.2) | 15,753 (41.6) | 13,369 (35.3) | 8,789 (23.2) |
| **Schizophrenia spectrum disorders** | 64,554 | 34,474 (53.4) | 44.3 (31.1;57.4) | 48,249 (74.7) | 12,784 (19.8) | 3,521 (5.5) |
| **Sleep disorders** | 91,951 | 66,152 (71.9) | 56.6 (44.6;66.7) | 58,703 (63.8) | 24,486 (26.6) | 8,762 (9.5) |
| **Stress related disorders** | 221,000 | 87,924 (39.8) | 42.4 (29.1;54.4) | 170,769 (77.3) | 40,490 (18.3) | 9,741 (4.4) |
| **Stroke** | 124,371 | 67,040 (53.9) | 71.5 (61.4;79.9) | 16,713 (13.4) | 72,293 (58.1) | 35,365 (28.4) |
| **Traumatic brain injury** | 245,649 | 139,704 (56.9) | 35.5 (22.6;54.7) | 195,607 (79.6) | 39,071 (15.9) | 10,971 (4.5) |

| *Disease group* | *Persons, N* | *Men, N (%)%* | *Age, median (Q1-Q3)* | *CCI: 0, N (%)* | *CCI: 1-2, N (%)* | *CCI: +3, N (%)* |
|---|---|---|---|---|---|---|
| **Any brain disorder** | 1,189,102 | 593,858 (49.9) | 46.6 (27.9;64.1) | 824,504 (69.3) | 271,082 (22.8) | 93,516 (7.9) |
| **Alcohol abuse** | 99,986 | 66,717 (66.7) | 57.7 (46.7;67.3) | 53,283 (53.3) | 30,711 (30.7) | 15,992 (16.0) |
| **Anxiety disorders** | 119,322 | 41,887 (35.1) | 40.5 (28.1;54.1) | 91,777 (76.9) | 21,618 (18.1) | 5,927 (5.0) |
| **Bipolar disorder** | 26,224 | 10,454 (39.9) | 51.3 (38.0;64.1) | 18,093 (69.0) | 6,121 (23.3) | 2,010 (7.7) |
| **Brain tumors** | 17,417 | 6,947 (39.9) | 63.8 (50.3;73.4) | 9,006 (51.7) | 5,621 (32.3) | 2,790 (16.0) |
| **Cerebral palsy** | 9,499 | 5,339 (56.2) | 23.2 (14.4;39.2) | 5,757 (60.6) | 2,869 (30.2) | 873 (9.2) |
| **Dementia** | 41,597 | 16,851 (40.5) | 81.3 (74.2;87.2) | 2,026 (4.9) | 26,266 (63.1) | 13,305 (32.0) |
| **Depression** | 200,929 | 71,354 (35.5) | 48.9 (35.1;63.2) | 138,030 (68.7) | 46,291 (23.0) | 16,608 (8.3) |
| **Developmental and behavioural disorders** | 131,906 | 85,636 (64.9) | 20.6 (14.9;27.3) | 118,070 (89.5) | 12,826 (9.7) | 1,010 (0.8) |
| **Drug abuse** | 55,330 | 36,426 (65.8) | 37.8 (28.4;50.4) | 41,016 (74.1) | 11,230 (20.3) | 3,084 (5.6) |
| **Eating disorders** | 18,261 | 913 (5.0) | 29.6 (23.2;37.4) | 16,149 (88.4) | 1,894 (10.4) | 218 (1.2) |
| **Epilepsy** | 73,595 | 37,746 (51.3) | 45.0 (25.5;63.7) | 48,318 (65.7) | 17,953 (24.4) | 7,324 (10.0) |
| **Headache** | 116,370 | 38,368 (33.0) | 45.5 (30.8;57.5) | 87,107 (74.9) | 23,910 (20.5) | 5,353 (4.6) |
| **Infections of the CNS** | 28,825 | 15,012 (52.1) | 44.8 (24.0;62.9) | 20,897 (72.5) | 5,809 (20.2) | 2,119 (7.4) |
| **Intellectual disability** | 30,214 | 17,639 (58.4) | 25.9 (18.3;44.2) | 24,320 (80.5) | 5,021 (16.6) | 873 (2.9) |
| **Multiple sclerosis** | 16,308 | 5,065 (31.1) | 52.9 (43.0;62.9) | 12,300 (75.4) | 3,207 (19.7) | 801 (4.9) |
| **Neuromuscular disorders** | 8,225 | 4,215 (51.2) | 52.5 (34.2;67.6) | 4,476 (54.4) | 2,662 (32.4) | 1,087 (13.2) |
| **Other neurodegenerative disorders** | 2,751 | 1,426 (51.8) | 62.4 (48.0;72.1) | 1,471 (53.5) | 920 (33.4) | 360 (13.1) |
| **Parkinson's disease** | 10,974 | 6,460 (58.9) | 75.2 (68.7;81.0) | 4,963 (45.2) | 4,152 (37.8) | 1,859 (16.9) |
| **Personality disorders** | 75,280 | 24,722 (32.8) | 39.6 (30.4;50.7) | 59,107 (78.5) | 13,426 (17.8) | 2,747 (3.6) |
| **Polyneuropathy** | 39,453 | 22,760 (57.7) | 67.7 (56.1;76.5) | 16,171 (41.0) | 13,961 (35.4) | 9,321 (23.6) |
| **Schizophrenia spectrum disorders** | 66,221 | 35,357 (53.4) | 43.9 (30.7;57.2) | 49,523 (74.8) | 13,133 (19.8) | 3,565 (5.4) |
| **Sleep disorders** | 100,921 | 71,952 (71.3) | 56.9 (45.0;67.1) | 63,795 (63.2) | 27,197 (26.9) | 9,929 (9.8) |
| **Stress related disorders** | 231,981 | 92,500 (39.9) | 42.4 (29.0;54.6) | 178,989 (77.2) | 42,682 (18.4) | 10,310 (4.4) |
| **Stroke** | 126,470 | 68,484 (54.2) | 71.8 (61.5;79.9) | 16,883 (13.3) | 73,293 (58.0) | 36,294 (28.7) |
| **Traumatic brain injury** | 243,264 | 137,623 (56.6) | 35.6 (22.8;55.3) | 192,591 (79.2) | 39,303 (16.2) | 11,370 (4.7) |

| *Disease group* | *Persons, N* | *Men, N (%)%* | *Age, median (Q1-Q3)* | *CCI: 0, N (%)* | *CCI: 1-2, N (%)* | *CCI: +3, N (%)* |
|---|---|---|---|---|---|---|
| **Any brain disorder** | 1,215,837 | 606,734 (49.9) | 46.8 (28.1;64.3) | 845,580 (69.5) | 275,650 (22.7) | 94,607 (7.8) |
| **Alcohol abuse** | 98,217 | 65,474 (66.7) | 58.0 (46.9;67.7) | 52,364 (53.3) | 30,150 (30.7) | 15,703 (16.0) |
| **Anxiety disorders** | 124,910 | 43,814 (35.1) | 40.4 (28.1;54.2) | 96,522 (77.3) | 22,419 (17.9) | 5,969 (4.8) |
| **Bipolar disorder** | 26,894 | 10,682 (39.7) | 51.2 (37.7;64.0) | 18,706 (69.6) | 6,204 (23.1) | 1,984 (7.4) |
| **Brain tumors** | 18,107 | 7,177 (39.6) | 64.1 (50.7;73.9) | 9,422 (52.0) | 5,807 (32.1) | 2,878 (15.9) |
| **Cerebral palsy** | 9,485 | 5,348 (56.4) | 23.8 (14.8;40.0) | 5,858 (61.8) | 2,785 (29.4) | 842 (8.9) |
| **Dementia** | 40,669 | 16,638 (40.9) | 81.4 (74.4;87.0) | 2,106 (5.2) | 25,705 (63.2) | 12,858 (31.6) |
| **Depression** | 205,152 | 72,871 (35.5) | 48.7 (34.9;62.9) | 142,246 (69.3) | 46,590 (22.7) | 16,316 (8.0) |
| **Developmental and behavioural disorders** | 140,654 | 90,810 (64.6) | 21.0 (15.2;27.9) | 126,063 (89.6) | 13,493 (9.6) | 1,098 (0.8) |
| **Drug abuse** | 56,762 | 37,609 (66.3) | 37.5 (28.6;50.2) | 42,444 (74.8) | 11,201 (19.7) | 3,117 (5.5) |
| **Eating disorders** | 18,751 | 944 (5.0) | 29.7 (23.4;37.5) | 16,624 (88.7) | 1,897 (10.1) | 230 (1.2) |
| **Epilepsy** | 72,675 | 37,199 (51.2) | 45.4 (25.8;64.0) | 47,787 (65.8) | 17,775 (24.5) | 7,113 (9.8) |
| **Headache** | 121,579 | 39,923 (32.8) | 45.6 (30.9;57.7) | 91,076 (74.9) | 24,945 (20.5) | 5,558 (4.6) |
| **Infections of the CNS** | 29,584 | 15,366 (51.9) | 45.1 (24.3;62.9) | 21,537 (72.8) | 5,885 (19.9) | 2,162 (7.3) |
| **Intellectual disability** | 30,696 | 17,941 (58.4) | 26.3 (18.7;44.0) | 24,834 (80.9) | 4,988 (16.2) | 874 (2.8) |
| **Multiple sclerosis** | 16,514 | 5,096 (30.9) | 53.2 (43.3;63.2) | 12,484 (75.6) | 3,217 (19.5) | 813 (4.9) |
| **Neuromuscular disorders** | 8,330 | 4,285 (51.4) | 53.0 (34.6;67.9) | 4,530 (54.4) | 2,708 (32.5) | 1,092 (13.1) |
| **Other neurodegenerative disorders** | 2,775 | 1,430 (51.5) | 62.9 (48.7;72.5) | 1,495 (53.9) | 923 (33.3) | 357 (12.9) |
| **Parkinson's disease** | 11,060 | 6,526 (59.0) | 75.5 (68.9;81.2) | 5,002 (45.2) | 4,203 (38.0) | 1,855 (16.8) |
| **Personality disorders** | 75,407 | 24,388 (32.3) | 39.7 (30.6;50.7) | 59,372 (78.7) | 13,341 (17.7) | 2,694 (3.6) |
| **Polyneuropathy** | 40,756 | 23,454 (57.5) | 68.2 (56.6;76.8) | 16,641 (40.8) | 14,414 (35.4) | 9,701 (23.8) |
| **Schizophrenia spectrum disorders** | 67,736 | 36,162 (53.4) | 43.6 (30.5;57.0) | 51,006 (75.3) | 13,199 (19.5) | 3,531 (5.2) |
| **Sleep disorders** | 110,441 | 78,071 (70.7) | 57.2 (45.4;67.5) | 69,561 (63.0) | 29,917 (27.1) | 10,963 (9.9) |
| **Stress related disorders** | 241,254 | 96,318 (39.9) | 42.5 (29.1;54.8) | 186,678 (77.4) | 44,016 (18.2) | 10,560 (4.4) |
| **Stroke** | 127,765 | 69,374 (54.3) | 72.1 (61.7;80.0) | 17,280 (13.5) | 73,939 (57.9) | 36,546 (28.6) |
| **Traumatic brain injury** | 240,748 | 135,613 (56.3) | 35.8 (23.0;55.9) | 190,246 (79.0) | 39,085 (16.2) | 11,417 (4.7) |

| *Disease group* | *Persons, N* | *Men, N (%)%* | *Age, median (Q1-Q3)* | *CCI: 0, N (%)* | *CCI: 1-2, N (%)* | *CCI: +3, N (%)* |
|---|---|---|---|---|---|---|
| **Any brain disorder** | 1,236,878 | 616,883 (49.9) | 47.0 (28.3;64.5) | 861,695 (69.7) | 279,731 (22.6) | 95,452 (7.7) |
| **Alcohol abuse** | 96,001 | 63,957 (66.6) | 58.3 (46.9;68.0) | 51,380 (53.5) | 29,337 (30.6) | 15,284 (15.9) |
| **Anxiety disorders** | 129,331 | 45,392 (35.1) | 40.1 (28.1;54.1) | 100,653 (77.8) | 22,780 (17.6) | 5,898 (4.6) |
| **Bipolar disorder** | 27,603 | 10,962 (39.7) | 50.7 (37.4;63.7) | 19,337 (70.1) | 6,274 (22.7) | 1,992 (7.2) |
| **Brain tumors** | 18,925 | 7,487 (39.6) | 64.4 (51.2;74.4) | 9,884 (52.2) | 6,142 (32.5) | 2,899 (15.3) |
| **Cerebral palsy** | 9,401 | 5,294 (56.3) | 24.0 (14.9;40.5) | 5,848 (62.2) | 2,724 (29.0) | 829 (8.8) |
| **Dementia** | 39,782 | 16,467 (41.4) | 81.3 (74.5;86.9) | 2,189 (5.5) | 25,225 (63.4) | 12,368 (31.1) |
| **Depression** | 207,645 | 73,769 (35.5) | 48.5 (34.7;62.6) | 145,460 (70.1) | 46,360 (22.3) | 15,825 (7.6) |
| **Developmental and behavioural disorders** | 148,992 | 95,666 (64.2) | 21.4 (15.5;28.4) | 133,655 (89.7) | 14,169 (9.5) | 1,168 (0.8) |
| **Drug abuse** | 58,041 | 38,653 (66.6) | 37.4 (28.8;49.9) | 43,791 (75.4) | 11,216 (19.3) | 3,034 (5.2) |
| **Eating disorders** | 19,236 | 983 (5.1) | 29.7 (23.5;37.6) | 17,089 (88.8) | 1,907 (9.9) | 240 (1.2) |
| **Epilepsy** | 71,785 | 36,739 (51.2) | 45.7 (25.8;64.5) | 47,253 (65.8) | 17,574 (24.5) | 6,958 (9.7) |
| **Headache** | 125,845 | 41,182 (32.7) | 45.9 (31.1;58.0) | 94,449 (75.1) | 25,718 (20.4) | 5,678 (4.5) |
| **Infections of the CNS** | 30,312 | 15,718 (51.9) | 45.5 (24.7;63.4) | 22,066 (72.8) | 6,027 (19.9) | 2,219 (7.3) |
| **Intellectual disability** | 30,981 | 18,122 (58.5) | 26.7 (19.2;43.8) | 25,170 (81.2) | 4,969 (16.0) | 842 (2.7) |
| **Multiple sclerosis** | 16,959 | 5,265 (31.0) | 53.4 (43.4;63.4) | 12,868 (75.9) | 3,264 (19.2) | 827 (4.9) |
| **Neuromuscular disorders** | 8,454 | 4,350 (51.5) | 53.7 (35.1;68.5) | 4,592 (54.3) | 2,750 (32.5) | 1,112 (13.2) |
| **Other neurodegenerative disorders** | 2,839 | 1,470 (51.8) | 63.3 (49.2;73.2) | 1,527 (53.8) | 961 (33.8) | 351 (12.4) |
| **Parkinson’s disease** | 11,244 | 6,602 (58.7) | 75.7 (69.1;81.3) | 5,121 (45.5) | 4,248 (37.8) | 1,875 (16.7) |
| **Personality disorders** | 75,678 | 24,089 (31.8) | 39.6 (30.7;50.5) | 59,896 (79.1) | 13,174 (17.4) | 2,608 (3.4) |
| **Polyneuropathy** | 42,756 | 24,640 (57.6) | 68.6 (57.0;77.2) | 16,913 (39.6) | 15,171 (35.5) | 10,672 (25.0) |
| **Schizophrenia spectrum disorders** | 69,161 | 36,887 (53.3) | 43.1 (30.3;56.8) | 52,424 (75.8) | 13,311 (19.2) | 3,426 (5.0) |
| **Sleep disorders** | 118,979 | 83,480 (70.2) | 57.6 (45.7;67.8) | 74,564 (62.7) | 32,585 (27.4) | 11,830 (9.9) |
| **Stress related disorders** | 249,655 | 99,728 (39.9) | 42.5 (29.1;55.1) | 193,863 (77.7) | 45,074 (18.1) | 10,718 (4.3) |
| **Stroke** | 129,226 | 70,494 (54.6) | 72.4 (61.9;80.0) | 17,664 (13.7) | 74,763 (57.9) | 36,799 (28.5) |
| **Traumatic brain injury** | 238,121 | 133,481 (56.1) | 36.0 (23.2;56.6) | 187,827 (78.9) | 38,940 (16.4) | 11,354 (4.8) |

| *Disease group* | *Persons, N* | *Men, N (%)%* | *Age, median (Q1-Q3)* | *CCI: 0, N (%)* | *CCI: 1-2, N (%)* | *CCI: +3, N (%)* |
|---|---|---|---|---|---|---|
| **Any brain disorder** | 1,253,321 | 623,843 (49.8) | 47.3 (28.5;64.7) | 874,220 (69.8) | 283,153 (22.6) | 95,948 (7.7) |
| **Alcohol abuse** | 93,703 | 62,357 (66.5) | 58.6 (47.0;68.4) | 50,357 (53.7) | 28,505 (30.4) | 14,841 (15.8) |
| **Anxiety disorders** | 133,731 | 46,696 (34.9) | 39.9 (28.1;54.2) | 104,686 (78.3) | 23,181 (17.3) | 5,864 (4.4) |
| **Bipolar disorder** | 28,154 | 11,149 (39.6) | 50.3 (37.0;63.5) | 19,833 (70.4) | 6,358 (22.6) | 1,963 (7.0) |
| **Brain tumors** | 19,590 | 7,717 (39.4) | 64.7 (51.5;74.9) | 10,308 (52.6) | 6,383 (32.6) | 2,899 (14.8) |
| **Cerebral palsy** | 9,351 | 5,277 (56.4) | 24.4 (15.2;40.9) | 5,887 (63.0) | 2,649 (28.3) | 815 (8.7) |
| **Dementia** | 38,842 | 16,186 (41.7) | 81.1 (74.7;86.7) | 2,290 (5.9) | 24,664 (63.5) | 11,888 (30.6) |
| **Depression** | 209,481 | 74,290 (35.5) | 48.4 (34.6;62.4) | 148,220 (70.8) | 45,992 (22.0) | 15,269 (7.3) |
| **Developmental and behavioural disorders** | 157,352 | 100,199 (63.7) | 21.9 (15.9;29.0) | 141,402 (89.9) | 14,722 (9.4) | 1,228 (0.8) |
| **Drug abuse** | 59,105 | 39,429 (66.7) | 37.3 (29.0;49.7) | 44,943 (76.0) | 11,174 (18.9) | 2,988 (5.1) |
| **Eating disorders** | 19,756 | 1,030 (5.2) | 29.8 (23.6;37.4) | 17,607 (89.1) | 1,929 (9.8) | 220 (1.1) |
| **Epilepsy** | 70,938 | 36,289 (51.2) | 46.0 (26.1;64.8) | 46,863 (66.1) | 17,289 (24.4) | 6,786 (9.6) |
| **Headache** | 129,702 | 42,276 (32.6) | 46.1 (31.3;58.3) | 97,473 (75.2) | 26,394 (20.3) | 5,835 (4.5) |
| **Infections of the CNS** | 30,346 | 15,638 (51.5) | 45.9 (24.8;63.9) | 22,098 (72.8) | 6,026 (19.9) | 2,222 (7.3) |
| **Intellectual disability** | 30,980 | 18,149 (58.6) | 27.1 (19.6;43.5) | 25,316 (81.7) | 4,877 (15.7) | 787 (2.5) |
| **Multiple sclerosis** | 17,343 | 5,373 (31.0) | 53.8 (43.6;63.7) | 13,184 (76.0) | 3,356 (19.4) | 803 (4.6) |
| **Neuromuscular disorders** | 8,606 | 4,409 (51.2) | 54.2 (35.5;69.0) | 4,643 (54.0) | 2,834 (32.9) | 1,129 (13.1) |
| **Other neurodegenerative disorders** | 2,857 | 1,454 (50.9) | 63.5 (49.4;73.5) | 1,565 (54.8) | 949 (33.2) | 343 (12.0) |
| **Parkinson's disease** | 11,232 | 6,619 (58.9) | 75.7 (69.2;81.2) | 5,184 (46.2) | 4,238 (37.7) | 1,810 (16.1) |
| **Personality disorders** | 75,905 | 23,686 (31.2) | 39.5 (30.9;50.4) | 60,350 (79.5) | 13,041 (17.2) | 2,514 (3.3) |
| **Polyneuropathy** | 44,576 | 25,804 (57.9) | 68.9 (57.5;77.4) | 17,121 (38.4) | 15,874 (35.6) | 11,581 (26.0) |
| **Schizophrenia spectrum disorders** | 70,593 | 37,517 (53.1) | 42.7 (30.2;56.7) | 53,783 (76.2) | 13,449 (19.1) | 3,361 (4.8) |
| **Sleep disorders** | 126,126 | 87,943 (69.7) | 58.0 (46.1;68.3) | 78,511 (62.2) | 35,001 (27.8) | 12,614 (10.0) |
| **Stress related disorders** | 255,788 | 101,929 (39.8) | 42.6 (29.2;55.5) | 199,245 (77.9) | 45,835 (17.9) | 10,708 (4.2) |
| **Stroke** | 131,100 | 71,736 (54.7) | 72.6 (62.2;80.2) | 17,988 (13.7) | 76,189 (58.1) | 36,923 (28.2) |
| **Traumatic brain injury** | 233,160 | 129,967 (55.7) | 36.3 (23.4;57.3) | 183,402 (78.7) | 38,511 (16.5) | 11,247 (4.8) |

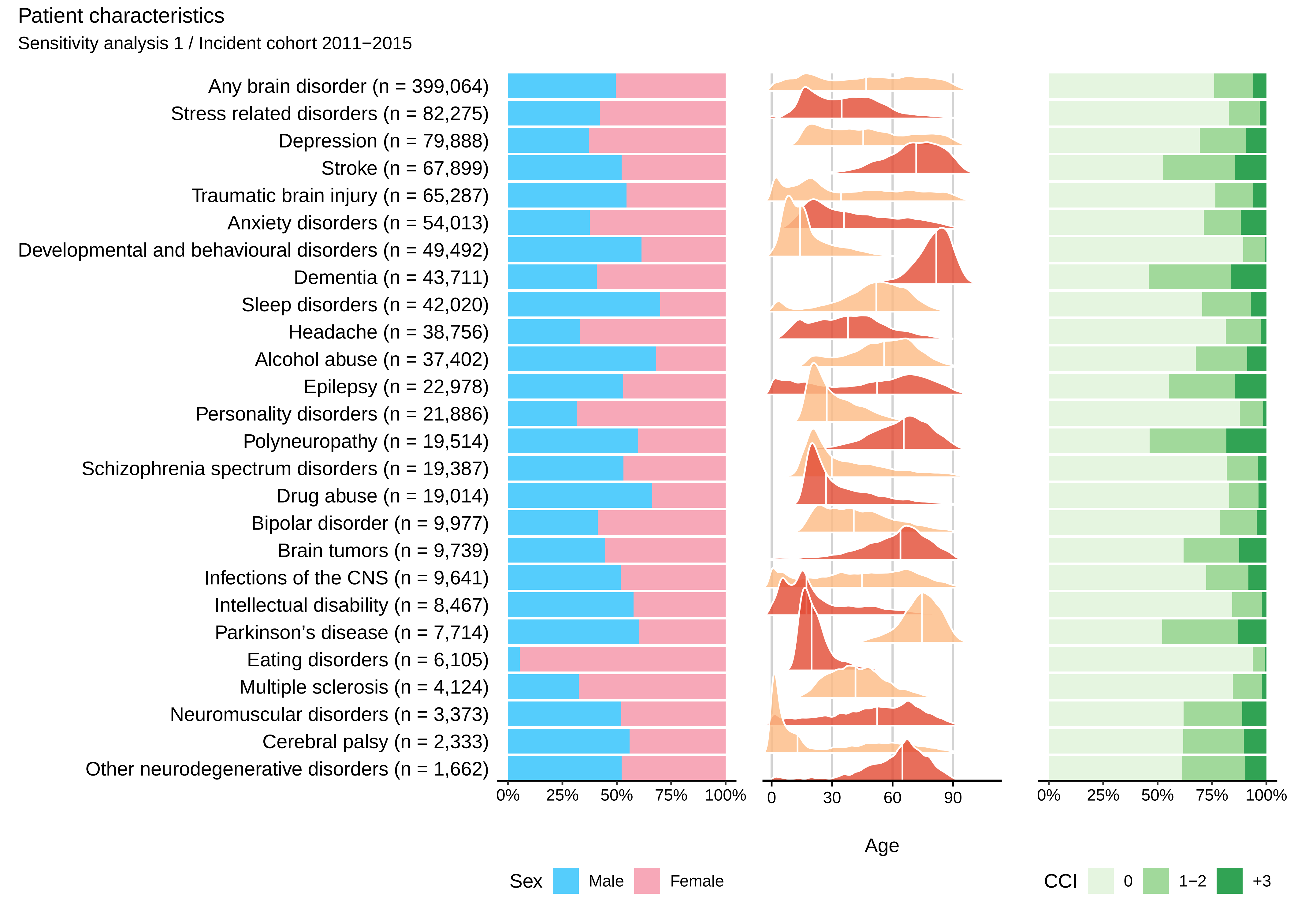

Patient characteristics
Sensitivity analysis 1 / Incident cohort 2011−2015
Any brain disorder (n = 399,064)
Stress related disorders (n = 82,275)
Depression (n = 79,888)
Stroke (n = 67,899)
Traumatic brain injury (n = 65,287)
Anxiety disorders (n = 54,013)
Developmental and behavioural disorders (n = 49,492)
Dementia (n = 43,711)
Sleep disorders (n = 42,020)
Headache (n = 38,756)
Alcohol abuse (n = 37,402)
Epilepsy (n = 22,978)
Personality disorders (n = 21,886)
Polyneuropathy (n = 19,514)
Schizophrenia spectrum disorders (n = 19,387)
Drug abuse (n = 19,014)
Bipolar disorder (n = 9,977)
Brain tumors (n = 9,739)
Infections of the CNS (n = 9,641)
Intellectual disability (n = 8,467)
Parkinson’s disease (n = 7,714)
Eating disorders (n = 6,105)
Multiple sclerosis (n = 4,124)
Neuromuscular disorders (n = 3,373)
Cerebral palsy (n = 2,333)
Other neurodegenerative disorders (n = 1,662)
0%
25%
50%
75%
100%
0
30
60
90
Age
Sex
Male
Female
CCI
0
1−2
+3

Patient characteristics

Sensitivity analysis 1 / Incident cohort 2016−2021

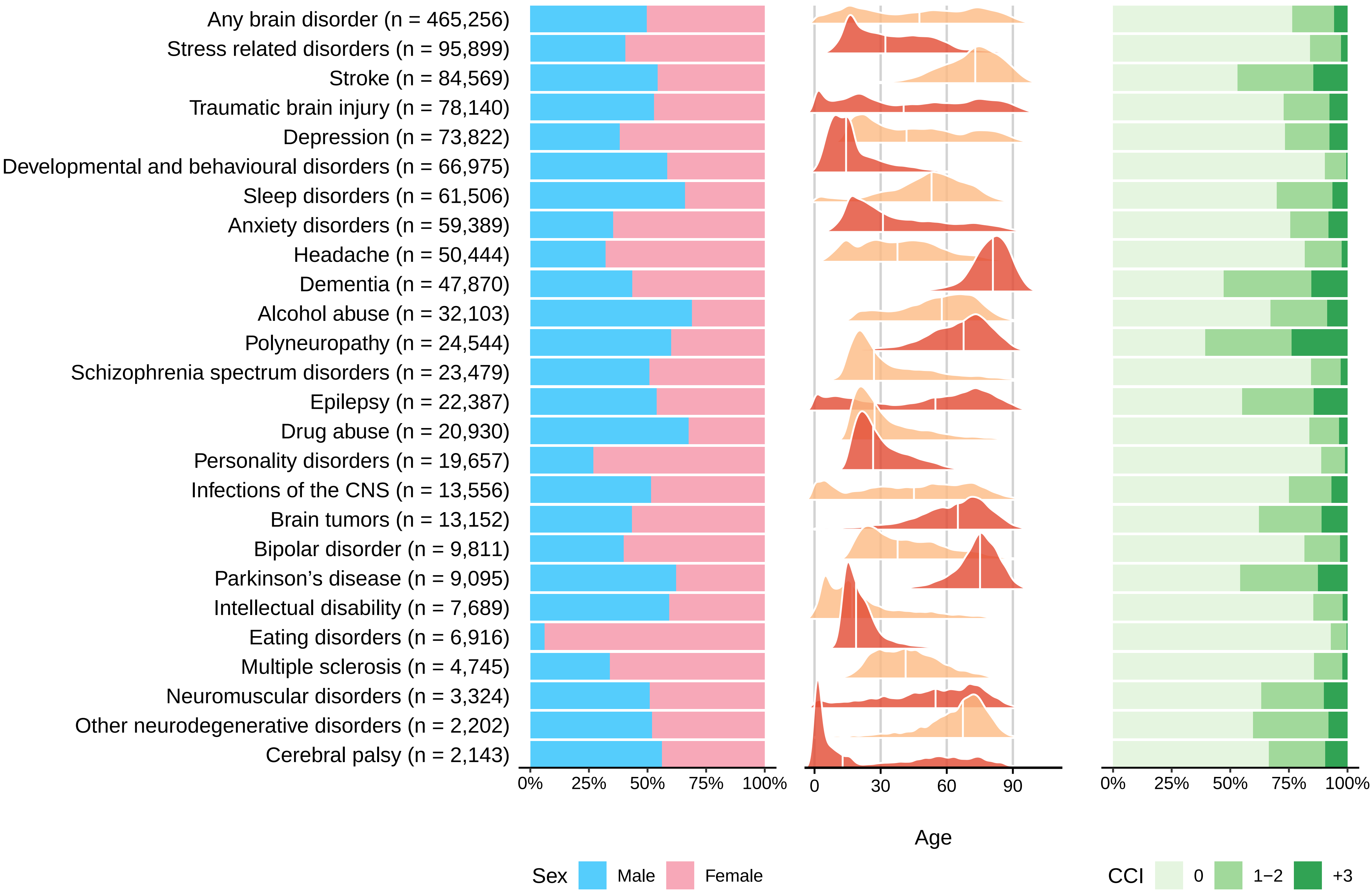

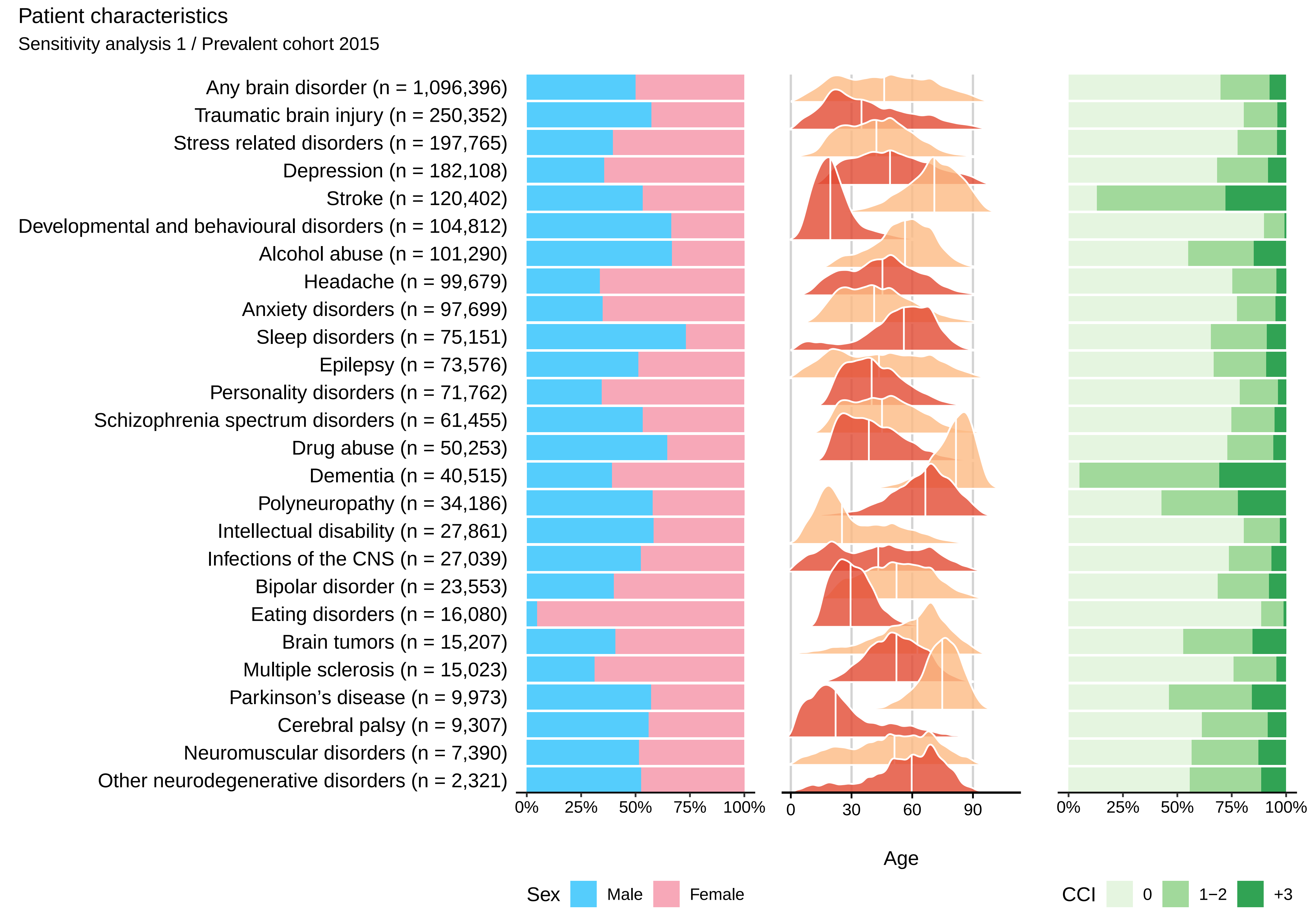

Patient characteristics
Sensitivity analysis 1 / Prevalent cohort 2015
Any brain disorder (n = 1,096,396)
Traumatic brain injury (n = 250,352)
Stress related disorders (n = 197,765)
Depression (n = 182,108)
Stroke (n = 120,402)
Developmental and behavioural disorders (n = 104,812)
Alcohol abuse (n = 101,290)
Headache (n = 99,679)
Anxiety disorders (n = 97,699)
Sleep disorders (n = 75,151)
Epilepsy (n = 73,576)
Personality disorders (n = 71,762)
Schizophrenia spectrum disorders (n = 61,455)
Drug abuse (n = 50,253)
Dementia (n = 40,515)
Polyneuropathy (n = 34,186)
Intellectual disability (n = 27,861)
Infections of the CNS (n = 27,039)
Bipolar disorder (n = 23,553)
Eating disorders (n = 16,080)
Brain tumors (n = 15,207)
Multiple sclerosis (n = 15,023)
Parkinson's disease (n = 9,973)
Cerebral palsy (n = 9,307)
Neuromuscular disorders (n = 7,390)
Other neurodegenerative disorders (n = 2,321)
0% 25% 50% 75% 100%
0 30 60 90
Age
0% 25% 50% 75% 100%
Sex Male Female
CCI 0 1−2 +3

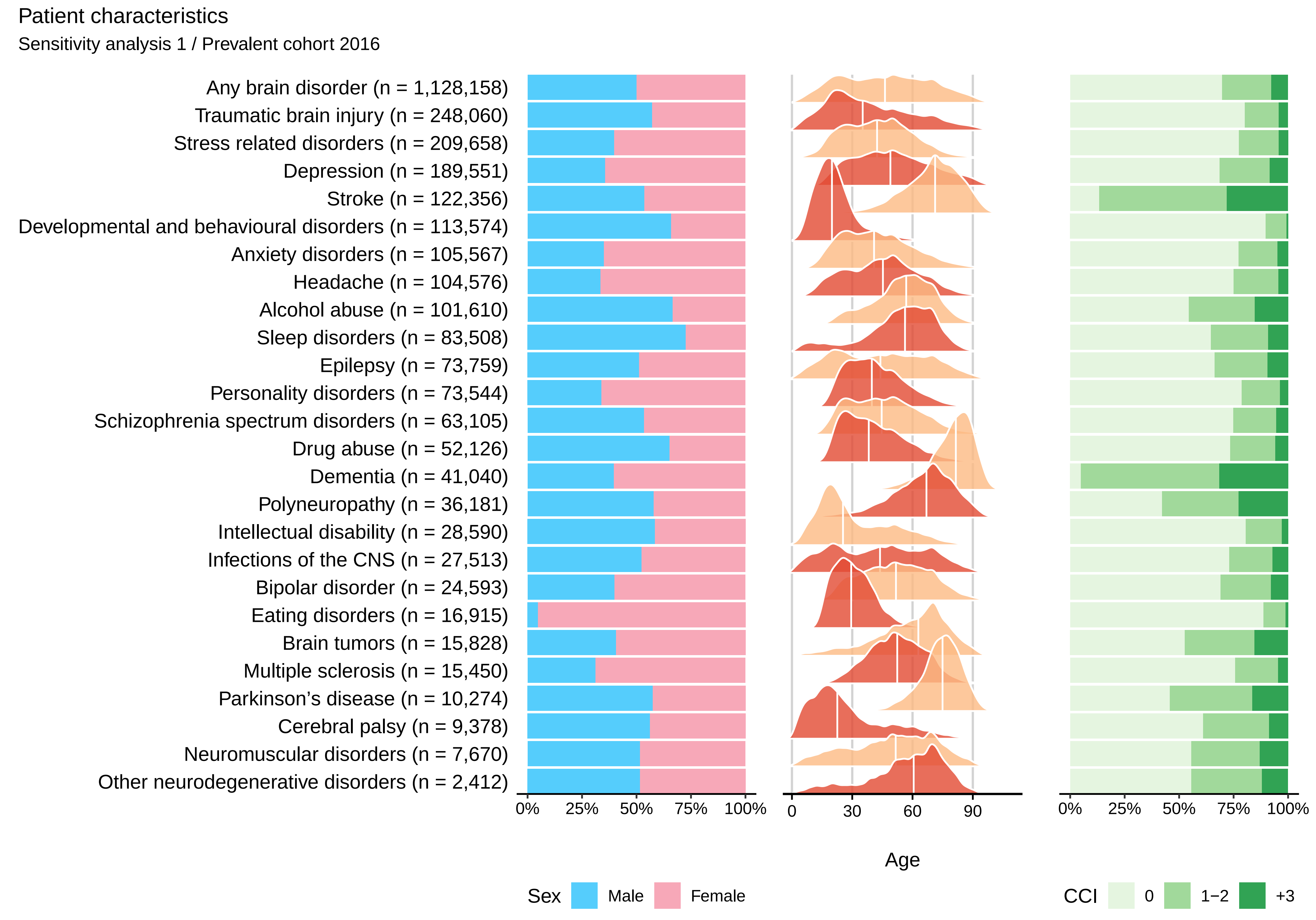

Patient characteristics
Sensitivity analysis 1 / Prevalent cohort 2016
Any brain disorder (n = 1,128,158)
Traumatic brain injury (n = 248,060)
Stress related disorders (n = 209,658)
Depression (n = 189,551)
Stroke (n = 122,356)
Developmental and behavioural disorders (n = 113,574)
Anxiety disorders (n = 105,567)
Headache (n = 104,576)
Alcohol abuse (n = 101,610)
Sleep disorders (n = 83,508)
Epilepsy (n = 73,759)
Personality disorders (n = 73,544)
Schizophrenia spectrum disorders (n = 63,105)
Drug abuse (n = 52,126)
Dementia (n = 41,040)
Polyneuropathy (n = 36,181)
Intellectual disability (n = 28,590)
Infections of the CNS (n = 27,513)
Bipolar disorder (n = 24,593)
Eating disorders (n = 16,915)
Brain tumors (n = 15,828)
Multiple sclerosis (n = 15,450)
Parkinson's disease (n = 10,274)
Cerebral palsy (n = 9,378)
Neuromuscular disorders (n = 7,670)
Other neurodegenerative disorders (n = 2,412)
0% 25% 50% 75% 100%
0 30 60 90
Age
0% 25% 50% 75% 100%
Sex Male Female
CCI 0 1−2 +3

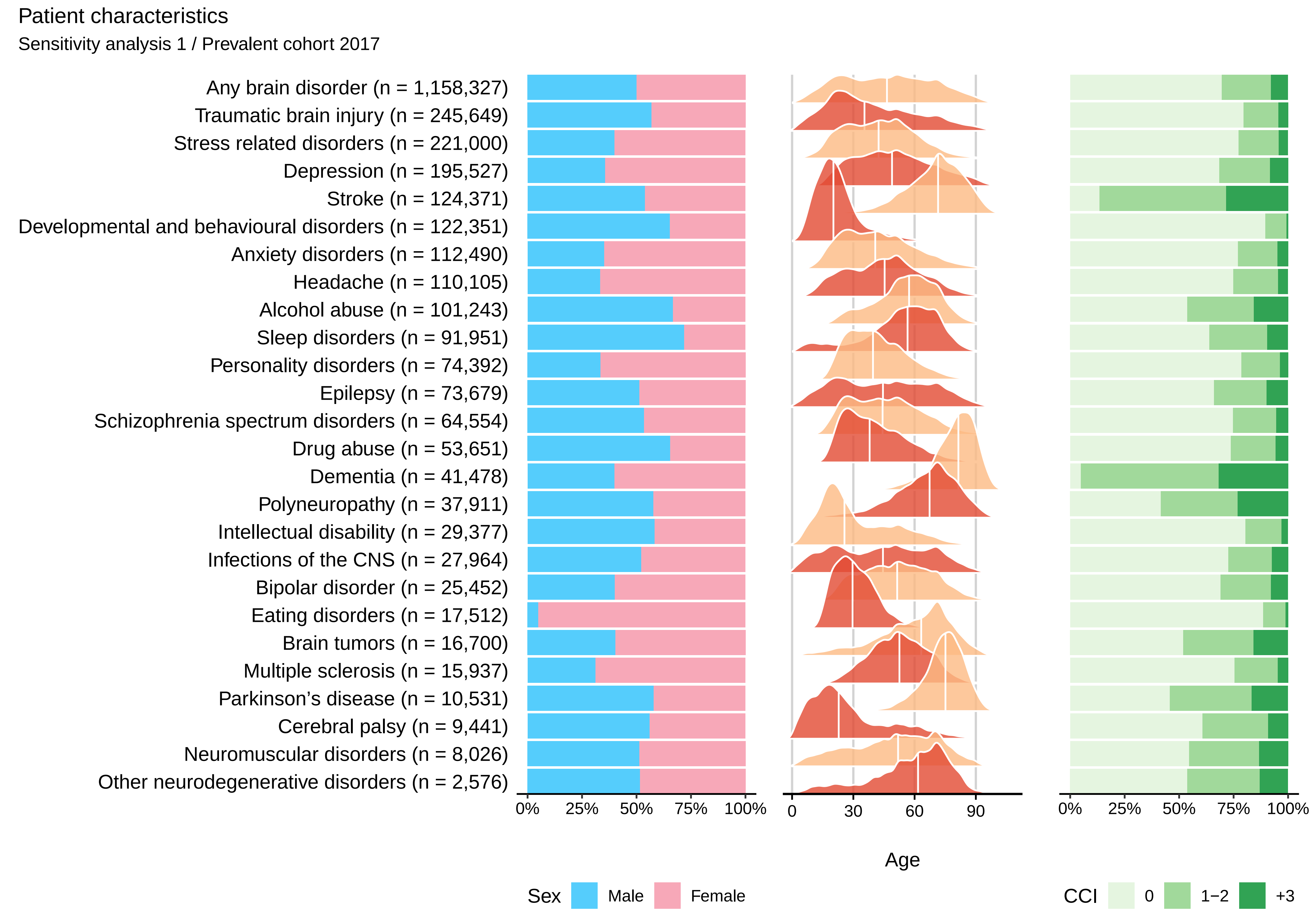

Patient characteristics
Sensitivity analysis 1 / Prevalent cohort 2017
Any brain disorder (n = 1,158,327)
Traumatic brain injury (n = 245,649)
Stress related disorders (n = 221,000)
Depression (n = 195,527)
Stroke (n = 124,371)
Developmental and behavioural disorders (n = 122,351)
Anxiety disorders (n = 112,490)
Headache (n = 110,105)
Alcohol abuse (n = 101,243)
Sleep disorders (n = 91,951)
Personality disorders (n = 74,392)
Epilepsy (n = 73,679)
Schizophrenia spectrum disorders (n = 64,554)
Drug abuse (n = 53,651)
Dementia (n = 41,478)
Polyneuropathy (n = 37,911)
Intellectual disability (n = 29,377)
Infections of the CNS (n = 27,964)
Bipolar disorder (n = 25,452)
Eating disorders (n = 17,512)
Brain tumors (n = 16,700)
Multiple sclerosis (n = 15,937)
Parkinson’s disease (n = 10,531)
Cerebral palsy (n = 9,441)
Neuromuscular disorders (n = 8,026)
Other neurodegenerative disorders (n = 2,576)
0% 25% 50% 75% 100%
0 30 60 90
Age
0% 25% 50% 75% 100%
Sex Male Female
CCI 0 1−2 +3

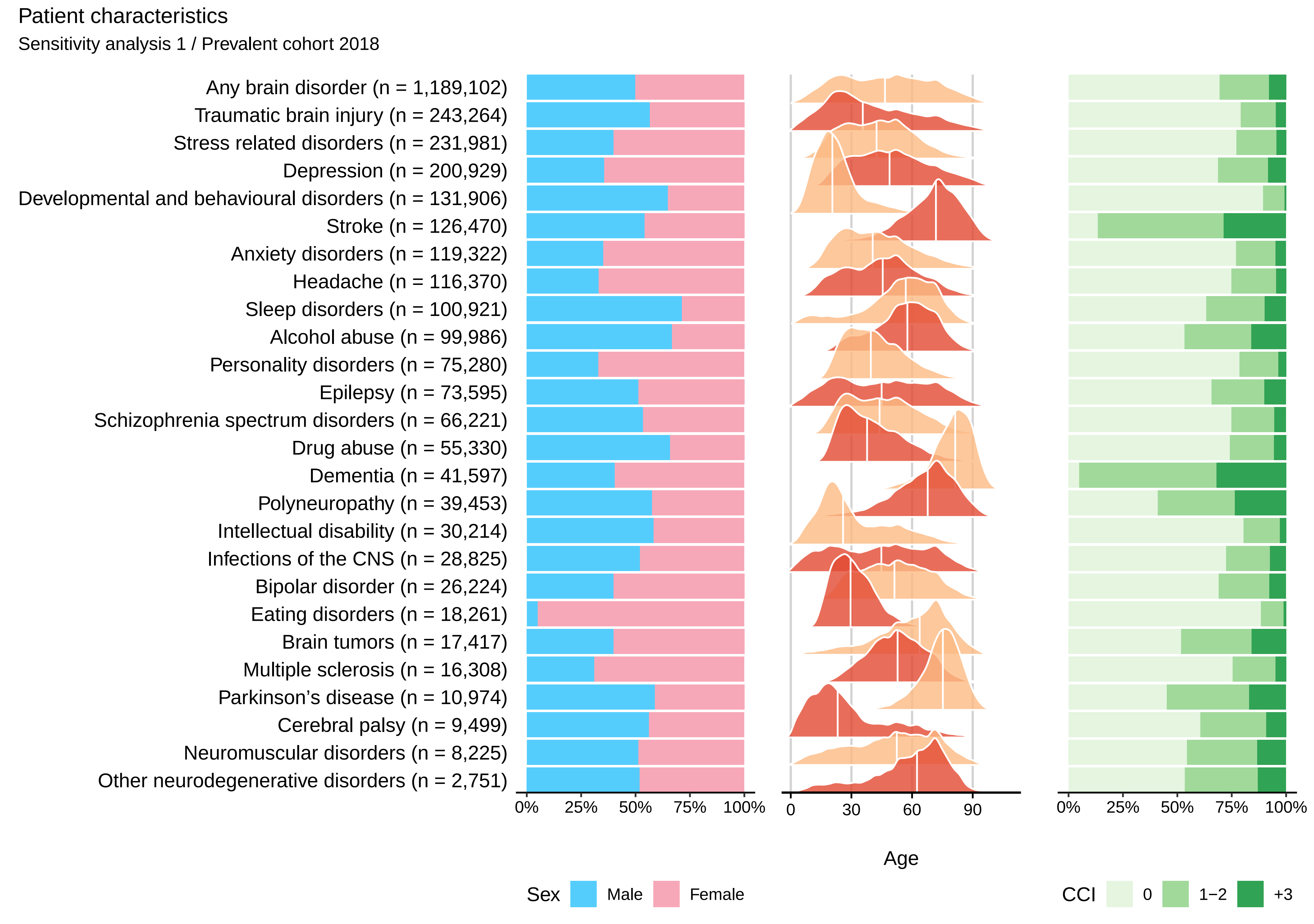

Patient characteristics
Sensitivity analysis 1 / Prevalent cohort 2018
Any brain disorder (n = 1,189,102)
Traumatic brain injury (n = 243,264)
Stress related disorders (n = 231,981)
Depression (n = 200,929)
Developmental and behavioural disorders (n = 131,906)
Stroke (n = 126,470)
Anxiety disorders (n = 119,322)
Headache (n = 116,370)
Sleep disorders (n = 100,921)
Alcohol abuse (n = 99,986)
Personality disorders (n = 75,280)
Epilepsy (n = 73,595)
Schizophrenia spectrum disorders (n = 66,221)
Drug abuse (n = 55,330)
Dementia (n = 41,597)
Polyneuropathy (n = 39,453)
Intellectual disability (n = 30,214)
Infections of the CNS (n = 28,825)
Bipolar disorder (n = 26,224)
Eating disorders (n = 18,261)
Brain tumors (n = 17,417)
Multiple sclerosis (n = 16,308)
Parkinson's disease (n = 10,974)
Cerebral palsy (n = 9,499)
Neuromuscular disorders (n = 8,225)
Other neurodegenerative disorders (n = 2,751)
0% 25% 50% 75% 100%
0 30 60 90
Age
0% 25% 50% 75% 100%
Sex Male Female
CCI 0 1−2 +3

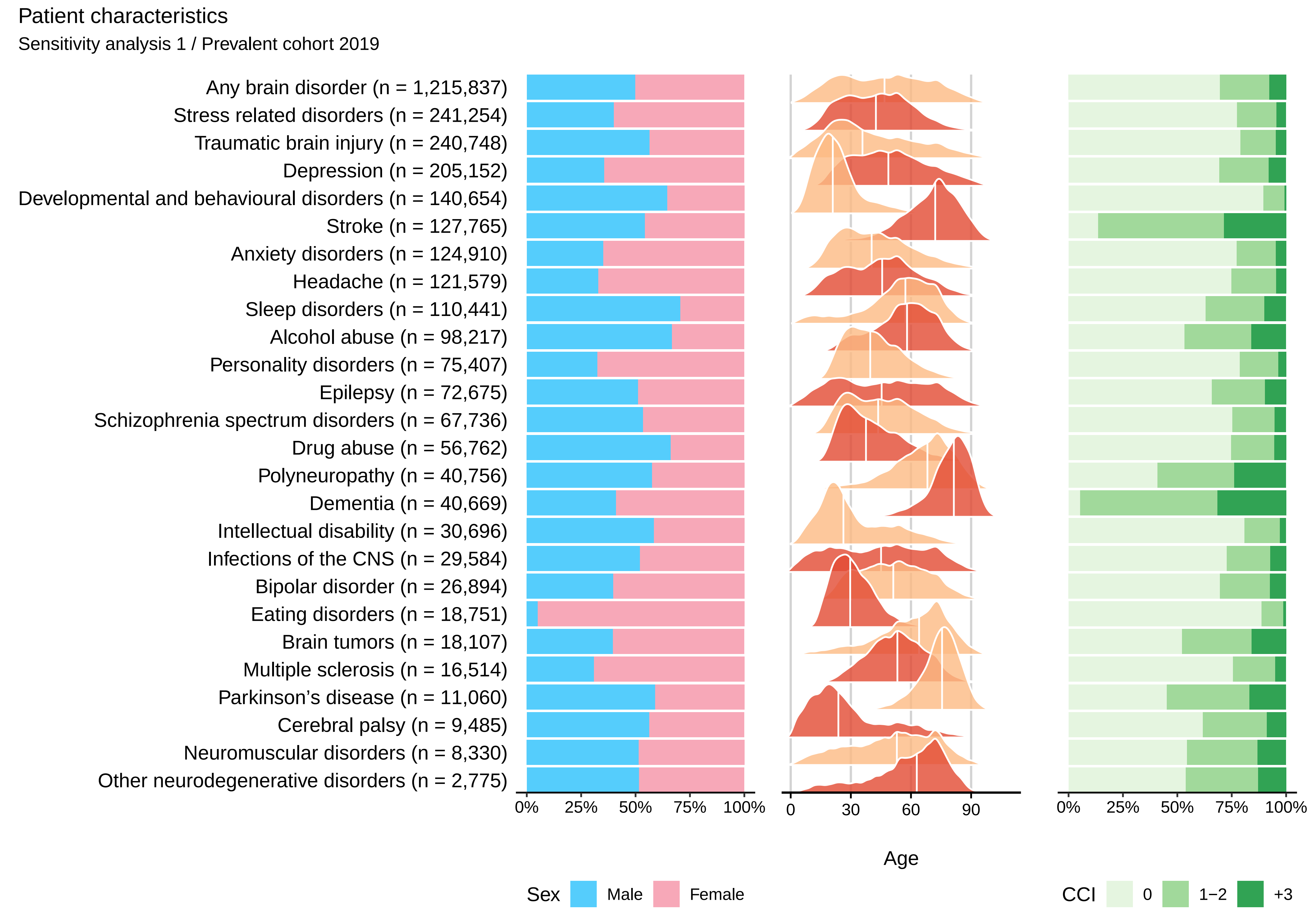
Patient characteristics
Sensitivity analysis 1 / Prevalent cohort 2019
Any brain disorder (n = 1,215,837)
Stress related disorders (n = 241,254)
Traumatic brain injury (n = 240,748)
Depression (n = 205,152)
Developmental and behavioural disorders (n = 140,654)
Stroke (n = 127,765)
Anxiety disorders (n = 124,910)
Headache (n = 121,579)
Sleep disorders (n = 110,441)
Alcohol abuse (n = 98,217)
Personality disorders (n = 75,407)
Epilepsy (n = 72,675)
Schizophrenia spectrum disorders (n = 67,736)
Drug abuse (n = 56,762)
Polyneuropathy (n = 40,756)
Dementia (n = 40,669)
Intellectual disability (n = 30,696)
Infections of the CNS (n = 29,584)
Bipolar disorder (n = 26,894)
Eating disorders (n = 18,751)
Brain tumors (n = 18,107)
Multiple sclerosis (n = 16,514)
Parkinson’s disease (n = 11,060)
Cerebral palsy (n = 9,485)
Neuromuscular disorders (n = 8,330)
Other neurodegenerative disorders (n = 2,775)
0% 25% 50% 75% 100%
0 30 60 90
Age
0% 25% 50% 75% 100%
Sex Male Female
CCI 0 1−2 +3

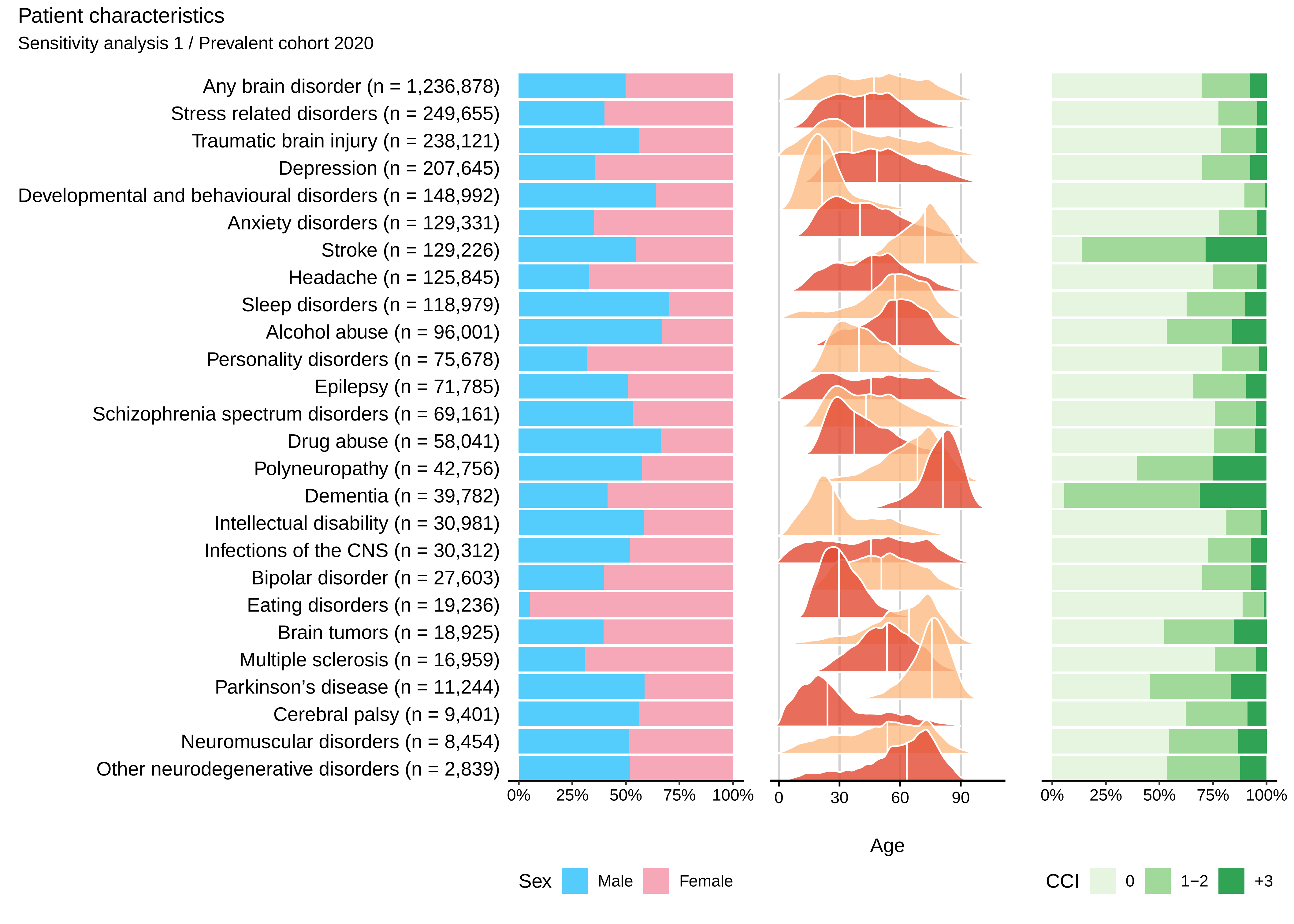

Patient characteristics
Sensitivity analysis 1 / Prevalent cohort 2020
Any brain disorder (n = 1,236,878)
Stress related disorders (n = 249,655)
Traumatic brain injury (n = 238,121)
Depression (n = 207,645)
Developmental and behavioural disorders (n = 148,992)
Anxiety disorders (n = 129,331)
Stroke (n = 129,226)
Headache (n = 125,845)
Sleep disorders (n = 118,979)
Alcohol abuse (n = 96,001)
Personality disorders (n = 75,678)
Epilepsy (n = 71,785)
Schizophrenia spectrum disorders (n = 69,161)
Drug abuse (n = 58,041)
Polyneuropathy (n = 42,756)
Dementia (n = 39,782)
Intellectual disability (n = 30,981)
Infections of the CNS (n = 30,312)
Bipolar disorder (n = 27,603)
Eating disorders (n = 19,236)
Brain tumors (n = 18,925)
Multiple sclerosis (n = 16,959)
Parkinson's disease (n = 11,244)
Cerebral palsy (n = 9,401)
Neuromuscular disorders (n = 8,454)
Other neurodegenerative disorders (n = 2,839)
0% 25% 50% 75% 100%
0 30 60 90
Age
0% 25% 50% 75% 100%
Sex Male Female
CCI 0 1–2 +3

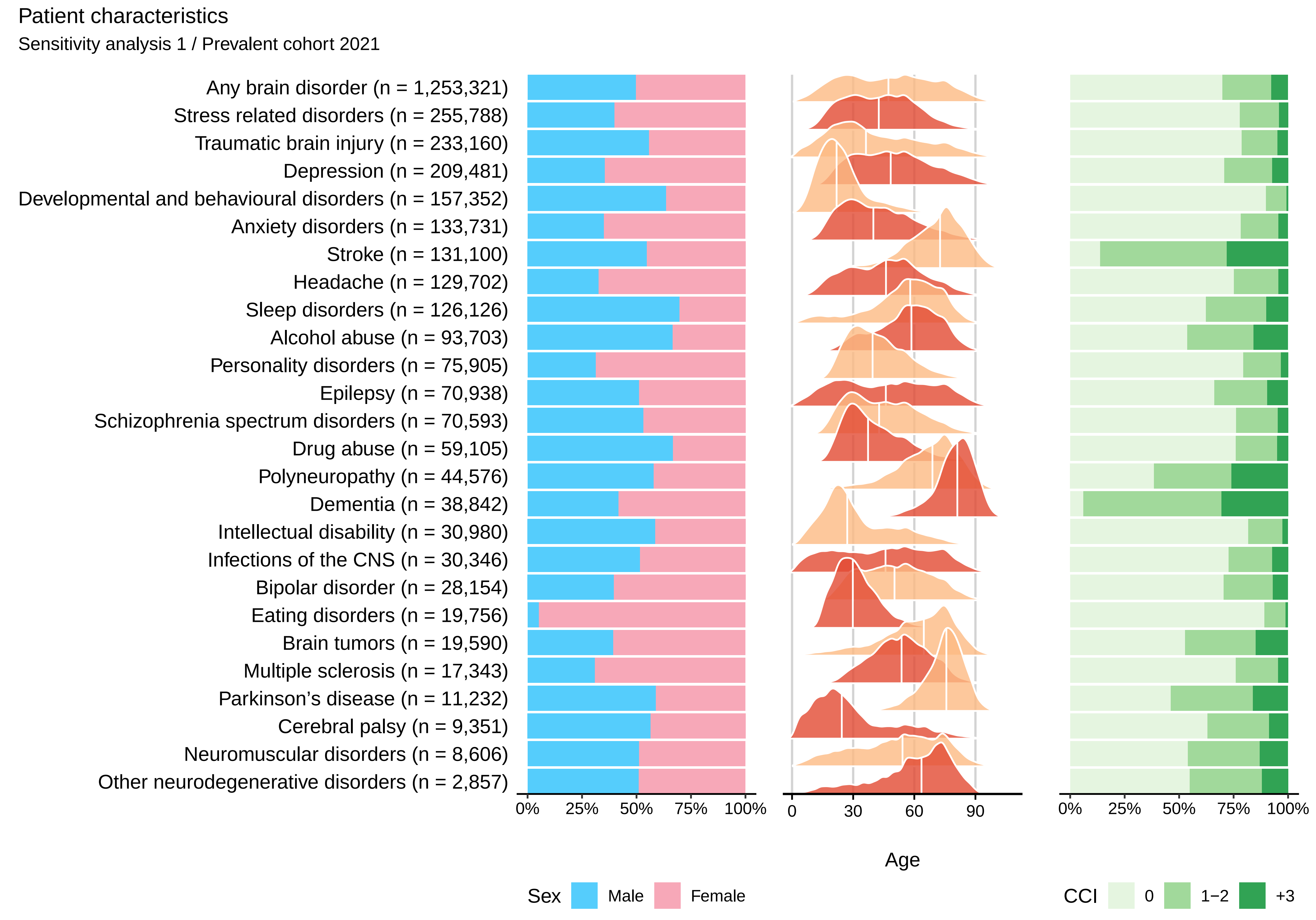

Patient characteristics
Sensitivity analysis 1 / Prevalent cohort 2021
Any brain disorder (n = 1,253,321)
Stress related disorders (n = 255,788)
Traumatic brain injury (n = 233,160)
Depression (n = 209,481)
Developmental and behavioural disorders (n = 157,352)
Anxiety disorders (n = 133,731)
Stroke (n = 131,100)
Headache (n = 129,702)
Sleep disorders (n = 126,126)
Alcohol abuse (n = 93,703)
Personality disorders (n = 75,905)
Epilepsy (n = 70,938)
Schizophrenia spectrum disorders (n = 70,593)
Drug abuse (n = 59,105)
Polyneuropathy (n = 44,576)
Dementia (n = 38,842)
Intellectual disability (n = 30,980)
Infections of the CNS (n = 30,346)
Bipolar disorder (n = 28,154)
Eating disorders (n = 19,756)
Brain tumors (n = 19,590)
Multiple sclerosis (n = 17,343)
Parkinson's disease (n = 11,232)
Cerebral palsy (n = 9,351)
Neuromuscular disorders (n = 8,606)
Other neurodegenerative disorders (n = 2,857)
0% 25% 50% 75% 100%
0 30 60 90
Age
0% 25% 50% 75% 100%
Sex Male Female
CCI 0 1−2 +3

### Concurrent diseases

#### Sensitivity analysis 1 − Incident cohort 2011−2015

| | Alcohol abuse | Anxiety disorders | Bipolar disorder | Brain tumors | Cerebral palsy | Dementia | Depression | Developmental and behavioural disorders | Drug abuse | Eating disorders | Epilepsy | Headache | Infections of the CNS | Intellectual disability | Multiple sclerosis | Neuromuscular disorders | Other neurodegenerative disorders | Parkinson's disease | Personality disorders | Polyneuropathy | Schizophrenia spectrum disorders | Sleep disorders | Stress related disorders | Stroke | Traumatic brain injury |
|---|---|---|---|---|---|---|---|---|---|---|---|---|---|---|---|---|---|---|---|---|---|---|---|---|---|
| Alcohol abuse | | 7.7 | 15.8 | 3.4 | 2.9 | 6.4 | 7.9 | 3.3 | 19.8 | 2 | 8.9 | 1.9 | 4.1 | 4.1 | 2.5 | 4.6 | 5.1 | 3.6 | 9.2 | 12.5 | 11.3 | 2.9 | 7.4 | 6.7 | 5.9 |
| Anxiety disorders | 6.7 | | 16.7 | 2.1 | 2 | 2.4 | 13.4 | 10.1 | 13.9 | 13.7 | 3.1 | 3.8 | 2.1 | 7.4 | 3.1 | 2.9 | 2.4 | 2.1 | 25 | 3 | 16 | 3.6 | 9.9 | 2 | 2.4 |
| Bipolar disorder | 2.4 | 2.2 | | 0.7 | 0.5 | 1.3 | 1.7 | 1.6 | 4.6 | 1.1 | 1.2 | 0.6 | 0.7 | 1.2 | 0.5 | 0.9 | 0.8 | 1.5 | 4.1 | 1.2 | 4.5 | 1 | 1.9 | 0.9 | 0.8 |
| Brain tumors | 0.3 | 0.9 | 0.4 | | 2 | 0.9 | 0.6 | 0.1 | 0.2 | 0.2 | 7 | 0.8 | 2.3 | 0.5 | 0.8 | 1.2 | 1.1 | 0.9 | 0.1 | 0.8 | 0.3 | 0.5 | 0.3 | 1.2 | 0.5 |
| Cerebral palsy | 0.1 | 0.2 | 0.2 | 0.2 | | 0.1 | 0.2 | 0.6 | 0.2 | 0.3 | 1.7 | 0.2 | 0.5 | 4.6 | 0.3 | 1 | 1.6 | 0.2 | 0.2 | 0.3 | 0.3 | 0.3 | 0.3 | 0.2 | 0.2 |
| Dementia | 2.6 | 1.1 | 2.5 | 1.8 | 2.6 | | 4.2 | 0.2 | 1 | 0.1 | 5.4 | 0.4 | 1.7 | 1.8 | 0.7 | 1.2 | 7.2 | 11.6 | 0.4 | 1.8 | 3 | 0.5 | 0.8 | 5.1 | 2.7 |
| Depression | 15.7 | 26.1 | 44.1 | 5 | 4.5 | 14.3 | | 11.8 | 23 | 20.1 | 8.4 | 7.1 | 4.8 | 8.1 | 5.7 | 6.1 | 7 | 10.8 | 38.8 | 8.6 | 26.5 | 7.6 | 21.9 | 6.9 | 5.9 |
| Developmental and behavioural disorders | 3.9 | 11.5 | 9.9 | 0.7 | 6.1 | 0.3 | 6 | | 20.1 | 9 | 4 | 2.5 | 1.5 | 39.8 | 1.7 | 2.1 | 1.1 | 0.4 | 17.3 | 0.6 | 15.1 | 3 | 9 | 0.3 | 3 |
| Drug abuse | 10 | 6.2 | 12.1 | 0.8 | 1.1 | 1.1 | 4.1 | 6.5 | | 2.8 | 3 | 1.5 | 1.5 | 4.3 | 1.2 | 1.6 | 1.1 | 0.8 | 13.6 | 1.8 | 17.8 | 1.4 | 5.6 | 1.2 | 2.1 |
| Eating disorders | 0.6 | 2.3 | 2.1 | 0.2 | 0.1 | 0 | 1.8 | 1.3 | 1.5 | | 0.4 | 0.8 | 0.5 | 0.7 | 0.5 | 0.4 | 0 | 0.1 | 4.3 | 0.2 | 1.7 | 0.3 | 1.3 | 0.1 | 0.4 |
| Epilepsy | 3.2 | 2.9 | 3.2 | 8.7 | 21.5 | 3.9 | 2.8 | 2.9 | 3.4 | 1.7 | | 2.6 | 3.9 | 15.1 | 2.3 | 3.6 | 4.2 | 3.6 | 2.9 | 3 | 3.9 | 2.2 | 2.5 | 3.3 | 3.1 |
| Headache | 1.9 | 3.8 | 3.4 | 3.2 | 1.5 | 1.3 | 4 | 1.8 | 2.8 | 2.5 | 3.3 | | 3.4 | 1.3 | 4.5 | 3.2 | 2 | 1.5 | 4 | 3 | 2.8 | 3.3 | 4 | 2.7 | 2.3 |
| Infections of the CNS | 0.7 | 0.7 | 0.6 | 1.4 | 2.9 | 0.7 | 0.7 | 0.6 | 0.8 | 0.6 | 2.3 | 1.3 | | 1 | 2 | 1.5 | 1.3 | 0.6 | 0.6 | 1.3 | 0.7 | 0.6 | 0.7 | 1 | 0.7 |
| Intellectual disability | 1 | 1.7 | 1.8 | 0.4 | 12.4 | 1 | 1 | 5.1 | 2.5 | 1 | 3.4 | 0.4 | 0.5 | | 0.4 | 1.2 | 1 | 0.5 | 2.1 | 0.2 | 4.1 | 0.8 | 1.6 | 0.4 | 0.8 |
| Multiple sclerosis | 0.2 | 0.3 | 0.4 | 0.6 | 0.7 | 0.3 | 0.3 | 0 | 0.2 | 0.1 | 0.8 | 0.4 | 0.5 | 0.1 | | 0.5 | 1.3 | 0.3 | 0.1 | 0.5 | 0.1 | 0.3 | 0.2 | 0.4 | 0.2 |
| Neuromuscular disorders | 0.2 | 0.2 | 0.2 | 0.4 | 0.9 | 0.2 | 0.2 | 0.1 | 0.2 | 0.1 | 0.3 | 0.2 | 0.4 | 0.6 | 0.6 | | 2.8 | 0.2 | 0.1 | 1.3 | 0.1 | 0.4 | 0.2 | 0.3 | 0.2 |
| Other neurodegenerative disorders | 0.1 | 0.1 | 0 | 0.1 | 1.3 | 0.3 | 0.1 | 0 | 0 | 0 | 0.1 | 0.1 | 0.1 | 0.2 | 0.5 | 0.8 | | 0.5 | 0 | 0.3 | 0.1 | 0.1 | 0.1 | 0.1 | 0.1 |
| Parkinson's disease | 0.3 | 0.3 | 0.3 | 0.6 | 0.9 | 3.9 | 0.7 | 0 | 0.2 | 0 | 1.1 | 0.1 | 0.3 | 0.1 | 0.2 | 1 | 2.4 | | 0.1 | 1.3 | 0.6 | 0.4 | 0.2 | 1 | 0.6 |
| Personality disorders | 5.6 | 10.5 | 16.8 | 1.2 | 1 | 1 | 6.5 | 7 | 14.7 | 10.8 | 2.3 | 2.9 | 1.7 | 5.2 | 2.8 | 2 | 1.4 | 1 | | 2 | 13.9 | 2.8 | 6.6 | 1.2 | 2 |
| Polyneuropathy | 2.9 | 1 | 1 | 1.5 | 1.6 | 2.6 | 1.6 | 0.2 | 1 | 0.2 | 1.9 | 0.8 | 1.8 | 0.3 | 3.1 | 6.8 | 8.2 | 3.4 | 0.5 | | 0.8 | 1.5 | 0.7 | 2.3 | 1.1 |
| Schizophrenia spectrum disorders | 6.7 | 5.7 | 15.4 | 1.6 | 1.6 | 2.9 | 4.1 | 3.9 | 17 | 3.4 | 3.1 | 1.6 | 1.5 | 8.8 | 1.5 | 1.5 | 2.5 | 2.4 | 9.2 | 1.7 | | 2 | 5.1 | 1.6 | 1.7 |
| Sleep disorders | 2.1 | 1.9 | 2.6 | 2.4 | 1.9 | 1.3 | 2.3 | 1.8 | 1.5 | 0.9 | 2.1 | 1.9 | 1.6 | 1.9 | 1 | 3.5 | 3.8 | 2.6 | 1.7 | 3.8 | 1.4 | | 1.9 | 2 | 1.2 |
| Stress related disorders | 15.9 | 21.2 | 32 | 4 | 5.2 | 3.6 | 23.2 | 15.3 | 30.4 | 16.6 | 6 | 8.1 | 4.7 | 14.2 | 5.6 | 4.7 | 4.9 | 3.4 | 40.5 | 5.8 | 30.1 | 6.6 | | 3.7 | 5.7 |
| Stroke | 6.7 | 3.2 | 3.1 | 9.6 | 14.2 | 17.3 | 6.8 | 0.5 | 2.1 | 0.4 | 21.2 | 4.3 | 6.8 | 1.9 | 3.3 | 5.3 | 7.7 | 11.9 | 1 | 8.7 | 2.7 | 4.2 | 2 | | 6.9 |
| Traumatic brain injury | 11.3 | 7.7 | 10 | 4.9 | 7.8 | 7.1 | 7.7 | 8.1 | 14.5 | 7.5 | 11.3 | 9.9 | 7.1 | 7.8 | 5.5 | 6 | 5.7 | 5.1 | 11.1 | 6.3 | 10.7 | 5.1 | 8.7 | 7.5 | |
| Myocardial infarction | 2.1 | 1.7 | 1 | 2.2 | 1.1 | 5 | 2.1 | 0.1 | 0.7 | 0 | 2.5 | 0.7 | 1.5 | 0.3 | 0.6 | 2.8 | 2.5 | 3.8 | 0.4 | 3.8 | 0.8 | 2.9 | 0.9 | 4.8 | 1.7 |
| Congestive heart failure | 3.2 | 2 | 1.4 | 2.3 | 1.4 | 8.2 | 2.9 | 0.2 | 1.1 | 0.1 | 3.5 | 0.7 | 1.9 | 0.6 | 0.4 | 3.8 | 2.4 | 4.8 | 0.3 | 5.7 | 1.4 | 3.5 | 0.9 | 6.7 | 2.5 |
| Peripheral vascular disease | 2.9 | 1.7 | 1 | 2.3 | 1.7 | 5.6 | 2.5 | 0.2 | 1.1 | 0.4 | 3.2 | 1 | 1.9 | 0.3 | 0.8 | 3 | 3.4 | 4 | 0.4 | 5.7 | 0.9 | 1.9 | 0.8 | 5.4 | 1.7 |
| Cerebrovascular disease | 6.7 | 4 | 3.8 | 9.9 | 15.1 | 20.8 | 7.8 | 0.6 | 2.6 | 0.3 | 22.4 | 4.9 | 6.5 | 2 | 3.8 | 6.7 | 10.5 | 15 | 1.2 | 10.6 | 3 | 5.4 | 2.5 | 11.7 | 6.6 |
| Chronic pulmonary disease | 8.8 | 8.3 | 6.2 | 6.3 | 8.1 | 10.9 | 8.1 | 6.6 | 6.1 | 3 | 8.3 | 5.8 | 6.3 | 6.4 | 3.7 | 10.1 | 11.3 | 7.9 | 4.8 | 11 | 5.8 | 9.1 | 5.7 | 9.6 | 6.5 |
| Connective tissue disease | 1.5 | 2 | 1.7 | 2.7 | 2.1 | 4.1 | 2.6 | 1.1 | 1.2 | 0.9 | 2.4 | 2.4 | 3.1 | 1.5 | 2 | 9 | 2.2 | 3 | 1.4 | 5.8 | 1.4 | 2.3 | 1.7 | 3.7 | 1.8 |
| Ulcer disease | 3.2 | 1.5 | 1.5 | 1.6 | 1.4 | 4.4 | 2.1 | 0.2 | 1.5 | 0.2 | 2.3 | 1 | 1.7 | 0.6 | 0.7 | 2 | 1.9 | 3 | 0.7 | 3.3 | 1.3 | 1.3 | 1 | 3.3 | 1.5 |
| Mild liver disease | 2.9 | 1.3 | 1.7 | 0.9 | 0.5 | 1.1 | 1.3 | 0.5 | 2.8 | 0.3 | 2 | 0.7 | 1.5 | 0.5 | 0.4 | 1.5 | 1.1 | 0.8 | 1.2 | 3.2 | 2 | 0.7 | 1.2 | 1.6 | 1.3 |
| Diabetes without end−organ damage | 5.9 | 4 | 3.9 | 5.3 | 3.1 | 10 | 5.3 | 0.8 | 2.8 | 0.9 | 6.3 | 2.3 | 4.5 | 2.2 | 2 | 5.7 | 5.1 | 8 | 1.9 | 16.3 | 3.1 | 7.9 | 3 | 9 | 3.6 |
| Hemiplegia | 0.2 | 0.2 | 0.2 | 0.5 | 14 | 0.3 | 0.3 | 0.2 | 0.2 | 0.1 | 1.3 | 0.3 | 0.8 | 1.8 | 1.2 | 1.2 | 2.4 | 0.4 | 0.1 | 0.5 | 0.2 | 0.5 | 0.3 | 0.3 | 0.3 |
| Moderate to severe renal disease | 1.9 | 1.5 | 1.4 | 1.7 | 1.5 | 4.2 | 2 | 0.4 | 1.2 | 0.4 | 2.8 | 0.9 | 2.2 | 0.9 | 0.5 | 4.4 | 1.9 | 3.2 | 0.6 | 5.1 | 1 | 2 | 1 | 4 | 1.6 |
| Diabetes with end−organ damage | 2.9 | 1.8 | 1.7 | 2.5 | 2 | 5.6 | 2.7 | 0.3 | 1.4 | 0.3 | 3.7 | 1 | 2.3 | 0.9 | 0.9 | 2.9 | 2.9 | 4.4 | 0.9 | 11.4 | 1.5 | 4.3 | 1.3 | 5.2 | 1.9 |
| Solid cancer tumour | 6.6 | 14.5 | 3.5 | 14.8 | 3.6 | 12.7 | 8.1 | 0.7 | 2 | 0.4 | 10.8 | 2.6 | 6.5 | 1.2 | 2.3 | 7.4 | 7.8 | 12.5 | 1 | 12.4 | 2.3 | 4.7 | 2.8 | 11.4 | 4.5 |
| Leukaemia | 0.1 | 0.3 | 0.2 | 0.5 | 0.1 | 0.3 | 0.3 | 0 | 0.1 | 0 | 0.4 | 0.1 | 0.8 | 0 | 0 | 0.7 | 0.4 | 0.4 | 0 | 0.7 | 0.1 | 0.2 | 0.2 | 0.4 | 0.2 |
| Lymphoma | 0.3 | 0.6 | 0.3 | 1.4 | 0.2 | 0.7 | 0.5 | 0 | 0.2 | 0 | 0.6 | 0.2 | 1 | 0.1 | 0.2 | 1.2 | 0.8 | 0.7 | 0.1 | 1.7 | 0.2 | 0.3 | 0.3 | 0.8 | 0.3 |
| Moderate to severe liver disease | 1.6 | 0.3 | 0.3 | 0.2 | 0.2 | 0.4 | 0.3 | 0.1 | 0.4 | 0 | 0.7 | 0.1 | 0.6 | 0.1 | 0.1 | 0.8 | 0.5 | 0.3 | 0.2 | 1 | 0.3 | 0.1 | 0.2 | 0.6 | 0.4 |
| Metastatic solid tumour | 0.6 | 5.1 | 0.3 | 4.6 | 0.6 | 0.7 | 1.8 | 0.3 | 0.2 | 0 | 2.1 | 0.3 | 0.9 | 0.1 | 0.3 | 0.8 | 0.7 | 0.8 | 0.1 | 1.5 | 0.2 | 0.8 | 0.3 | 1.3 | 0.4 |
| AIDS | 0.2 | 0.1 | 0.2 | 0.1 | 0 | 0.1 | 0.2 | 0 | 0.3 | 0 | 0.1 | 0.1 | 0.5 | 0.1 | 0 | 0.1 | 0 | 0.1 | 0.2 | 0.3 | 0.3 | 0.1 | 0.2 | 0.1 | 0.1 |

Percent: 40, 30, 20, 10, 0

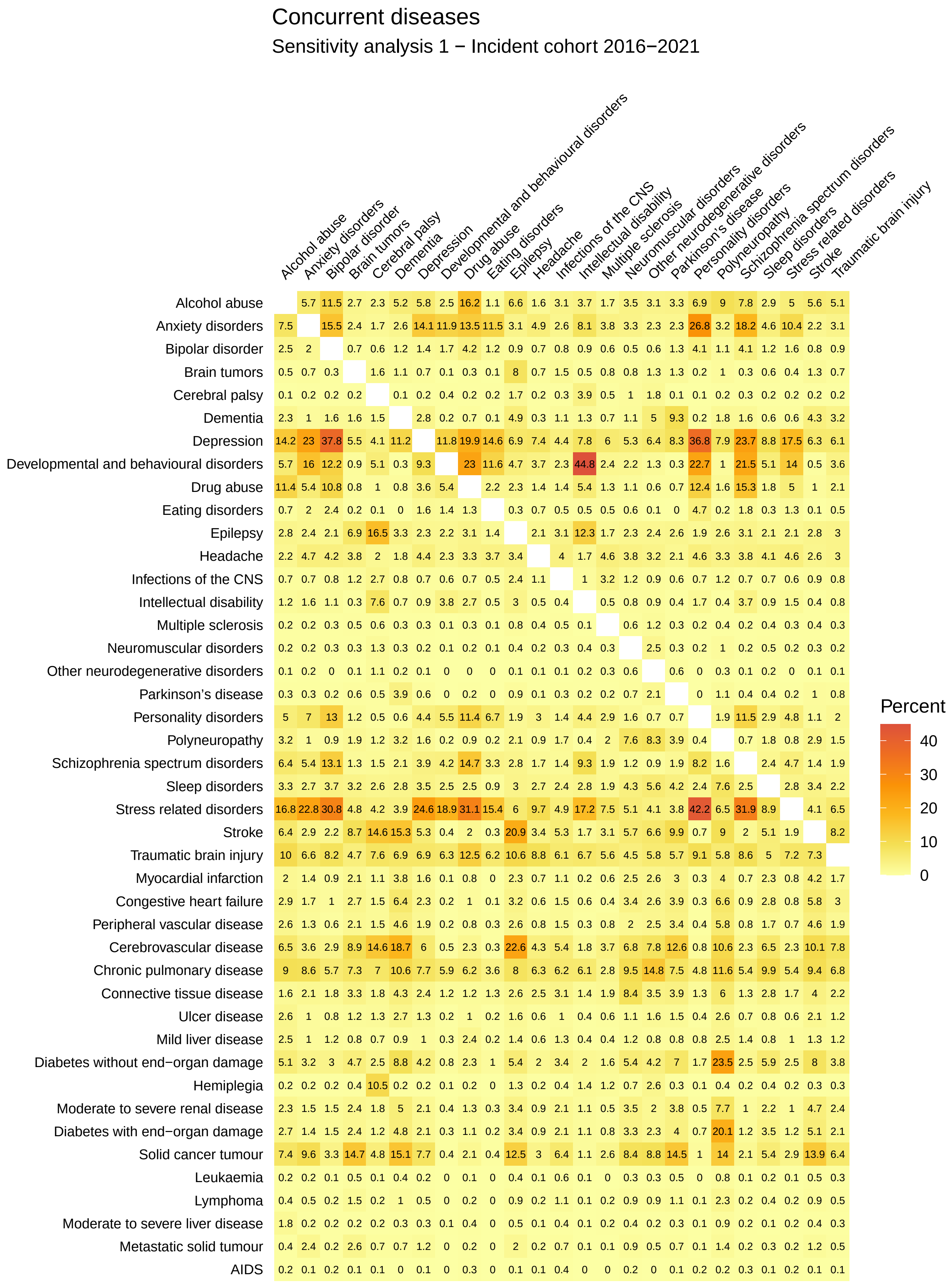
Concurrent diseases
Sensitivity analysis 1 − Incident cohort 2016−2021
Alcohol abuse
Anxiety disorders
Bipolar disorder
Brain tumors
Cerebral palsy
Dementia
Depression
Developmental and behavioural disorders
Drug abuse
Eating disorders
Epilepsy
Headache
Infections of the CNS
Intellectual disability
Multiple sclerosis
Neuromuscular disorders
Other neurodegenerative disorders
Parkinson's disease
Personality disorders
Polyneuropathy
Schizophrenia spectrum disorders
Sleep disorders
Stress related disorders
Stroke
Traumatic brain injury
Alcohol abuse 5.7 11.5 2.7 2.3 5.2 5.8 2.5 16.2 1.1 6.6 1.6 3.1 3.7 1.7 3.5 3.1 3.3 6.9 9 7.8 2.9 5 5.6 5.1
Anxiety disorders 7.5 15.5 2.4 1.7 2.6 14.1 11.9 13.5 11.5 3.1 4.9 2.6 8.1 3.8 3.3 2.3 2.3 26.8 3.2 18.2 4.6 10.4 2.2 3.1
Bipolar disorder 2.5 2 0.7 0.6 1.2 1.4 1.7 4.2 1.2 0.9 0.7 0.8 0.9 0.6 0.5 0.6 1.3 4.1 1.1 4.1 1.2 1.6 0.8 0.9
Brain tumors 0.5 0.7 0.3 1.6 1.1 0.7 0.1 0.3 0.1 8 0.7 1.5 0.5 0.8 0.8 1.3 1.3 0.2 1 0.3 0.6 0.4 1.3 0.7
Cerebral palsy 0.1 0.2 0.2 0.2 0.1 0.2 0.4 0.2 0.2 1.7 0.2 0.3 3.9 0.5 1 1.8 0.1 0.1 0.2 0.3 0.2 0.2 0.2 0.2
Dementia 2.3 1 1.6 1.6 1.5 2.8 0.2 0.7 0.1 4.9 0.3 1.1 1.3 0.7 1.1 5 9.3 0.2 1.8 1.6 0.6 0.6 4.3 3.2
Depression 14.2 23 37.8 5.5 4.1 11.2 11.8 19.9 14.6 6.9 7.4 4.4 7.8 6 5.3 6.4 8.3 36.8 7.9 23.7 8.8 17.5 6.3 6.1
Developmental and behavioural disorders 5.7 16 12.2 0.9 5.1 0.3 9.3 23 11.6 4.7 3.7 2.3 44.8 2.4 2.2 1.3 0.3 22.7 1 21.5 5.1 14 0.5 3.6
Drug abuse 11.4 5.4 10.8 0.8 1 0.8 3.6 5.4 2.2 2.3 1.4 1.4 5.4 1.3 1.1 0.6 0.7 12.4 1.6 15.3 1.8 5 1 2.1
Eating disorders 0.7 2 2.4 0.2 0.1 0 1.6 1.4 1.3 0.3 0.7 0.5 0.5 0.5 0.6 0.1 0 4.7 0.2 1.8 0.3 1.3 0.1 0.5
Epilepsy 2.8 2.4 2.1 6.9 16.5 3.3 2.3 2.2 3.1 1.4 2.1 3.1 12.3 1.7 2.3 2.4 2.6 1.9 2.6 3.1 2.1 2.1 2.8 3
Headache 2.2 4.7 4.2 3.8 2 1.8 4.4 2.3 3.3 3.7 3.4 4 1.7 4.6 3.8 3.2 2.1 4.6 3.3 3.8 4.1 4.6 2.6 3
Infections of the CNS 0.7 0.7 0.8 1.2 2.7 0.8 0.7 0.6 0.7 0.5 2.4 1.1 1 3.2 1.2 0.9 0.6 0.7 1.2 0.7 0.7 0.6 0.9 0.8
Intellectual disability 1.2 1.6 1.1 0.3 7.6 0.7 0.9 3.8 2.7 0.5 3 0.5 0.4 0.5 0.8 0.9 0.4 1.7 0.4 3.7 0.9 1.5 0.4 0.8
Multiple sclerosis 0.2 0.2 0.3 0.5 0.6 0.3 0.3 0.1 0.3 0.1 0.8 0.4 0.5 0.1 0.6 1.2 0.3 0.2 0.4 0.2 0.4 0.3 0.4 0.3
Neuromuscular disorders 0.2 0.2 0.3 0.3 1.3 0.3 0.2 0.1 0.2 0.1 0.4 0.2 0.3 0.4 0.3 2.5 0.3 0.2 1 0.2 0.5 0.2 0.3 0.2
Other neurodegenerative disorders 0.1 0.2 0 0.1 1.1 0.2 0.1 0 0 0 0.1 0.1 0.1 0.2 0.3 0.6 0.6 0 0.3 0.1 0.2 0 0.1 0.1
Parkinson's disease 0.3 0.3 0.2 0.6 0.5 3.9 0.6 0 0.2 0 0.9 0.1 0.3 0.2 0.2 0.7 2.1 0 1.1 0.4 0.4 0.2 1 0.8
Personality disorders 5 7 13 1.2 0.5 0.6 4.4 5.5 11.4 6.7 1.9 3 1.4 4.4 2.9 1.6 0.7 0.7 1.9 11.5 2.9 4.8 1.1 2
Polyneuropathy 3.2 1 0.9 1.9 1.2 3.2 1.6 0.2 0.9 0.2 2.1 0.9 1.7 0.4 2 7.6 8.3 3.9 0.4 0.7 1.8 0.8 2.9 1.5
Schizophrenia spectrum disorders 6.4 5.4 13.1 1.3 1.5 2.1 3.9 4.2 14.7 3.3 2.8 1.7 1.4 9.3 1.9 1.2 0.9 1.9 8.2 1.6 2.4 4.7 1.4 1.9
Sleep disorders 3.3 2.7 3.7 3.2 2.6 2.8 3.5 2.5 2.5 0.9 3 2.7 2.4 2.8 1.9 4.3 5.6 4.2 2.4 7.6 2.5 2.8 3.4 2.2
Stress related disorders 16.8 22.8 30.8 4.8 4.2 3.9 24.6 18.9 31.1 15.4 6 9.7 4.9 17.2 7.5 5.1 4.1 3.8 42.2 6.5 31.9 8.9 4.1 6.5
Stroke 6.4 2.9 2.2 8.7 14.6 15.3 5.3 0.4 2 0.3 20.9 3.4 5.3 1.7 3.1 5.7 6.6 9.9 0.7 9 2 5.1 1.9 8.2
Traumatic brain injury 10 6.6 8.2 4.7 7.6 6.9 6.9 6.3 12.5 6.2 10.6 8.8 6.1 6.7 5.6 4.5 5.8 5.7 9.1 5.8 8.6 5 7.2 7.3
Myocardial infarction 2 1.4 0.9 2.1 1.1 3.8 1.6 0.1 0.8 0 2.3 0.7 1.1 0.2 0.6 2.5 2.6 3 0.3 4 0.7 2.3 0.8 4.2 1.7
Congestive heart failure 2.9 1.7 1 2.7 1.5 6.4 2.3 0.2 1 0.1 3.2 0.6 1.5 0.6 0.4 3.4 2.6 3.9 0.3 6.6 0.9 2.8 0.8 5.8 3
Peripheral vascular disease 2.6 1.3 0.6 2.1 1.5 4.6 1.9 0.2 0.8 0.3 2.6 0.8 1.5 0.3 0.8 2 2.5 3.4 0.4 5.8 0.8 1.7 0.7 4.6 1.9
Cerebrovascular disease 6.5 3.6 2.9 8.9 14.6 18.7 6 0.5 2.3 0.3 22.6 4.3 5.4 1.8 3.7 6.8 7.8 12.6 0.8 10.6 2.3 6.5 2.3 10.1 7.8
Chronic pulmonary disease 9 8.6 5.7 7.3 7 10.6 7.7 5.9 6.2 3.6 8 6.3 6.2 6.1 2.8 9.5 14.8 7.5 4.8 11.6 5.4 9.9 5.4 9.4 6.8
Connective tissue disease 1.6 2.1 1.8 3.3 1.8 4.3 2.4 1.2 1.2 1.3 2.6 2.5 3.1 1.4 1.9 8.4 3.5 3.9 1.3 6 1.3 2.8 1.7 4 2.2
Ulcer disease 2.6 1 0.8 1.2 1.3 2.7 1.3 0.2 1 0.2 1.6 0.6 1 0.4 0.6 1.1 1.6 1.5 0.4 2.6 0.7 0.8 0.6 2.1 1.2
Mild liver disease 2.5 1 1.2 0.8 0.7 0.9 1 0.3 2.4 0.2 1.4 0.6 1.3 0.4 0.4 1.2 0.8 0.8 0.8 2.5 1.4 0.8 1 1.3 1.2
Diabetes without end−organ damage 5.1 3.2 3 4.7 2.5 8.8 4.2 0.8 2.3 1 5.4 2 3.4 2 1.6 5.4 4.2 7 1.7 23.5 2.5 5.9 2.5 8 3.8
Hemiplegia 0.2 0.2 0.2 0.4 10.5 0.2 0.2 0.1 0.2 0 1.3 0.2 0.4 1.4 1.2 0.7 2.6 0.3 0.1 0.4 0.2 0.4 0.2 0.3 0.3
Moderate to severe renal disease 2.3 1.5 1.5 2.4 1.8 5 2.1 0.4 1.3 0.3 3.4 0.9 2.1 1.1 0.5 3.5 2 3.8 0.5 7.7 1 2.2 1 4.7 2.4
Diabetes with end−organ damage 2.7 1.4 1.5 2.4 1.2 4.8 2.1 0.3 1.1 0.2 3.4 0.9 2.1 1.1 0.8 3.3 2.3 4 0.7 20.1 1.2 3.5 1.2 5.1 2.1
Solid cancer tumour 7.4 9.6 3.3 14.7 4.8 15.1 7.7 0.4 2.1 0.4 12.5 3 6.4 1.1 2.6 8.4 8.8 14.5 1 14 2.1 5.4 2.9 13.9 6.4
Leukaemia 0.2 0.2 0.1 0.5 0.1 0.4 0.2 0 0.1 0 0.4 0.1 0.6 0.1 0 0.3 0.3 0.5 0 0.8 0.1 0.2 0.1 0.5 0.3
Lymphoma 0.4 0.5 0.2 1.5 0.2 1 0.5 0 0.2 0 0.9 0.2 1.1 0.1 0.2 0.9 0.9 1.1 0.1 2.3 0.2 0.4 0.2 0.9 0.5
Moderate to severe liver disease 1.8 0.2 0.2 0.2 0.2 0.3 0.3 0.1 0.4 0 0.5 0.1 0.4 0.1 0.2 0.4 0.2 0.3 0.1 0.9 0.2 0.1 0.2 0.4 0.3
Metastatic solid tumour 0.4 2.4 0.2 2.6 0.7 0.7 1.2 0 0.2 0 2 0.2 0.7 0.1 0.1 0.9 0.5 0.7 0.1 1.4 0.2 0.3 0.2 1.2 0.5
AIDS 0.2 0.1 0.2 0.1 0.1 0 0.1 0 0.3 0 0.1 0.1 0.4 0 0 0.2 0 0.1 0.2 0.2 0.3 0.1 0.2 0.1 0.1
Percent
40
30
20
10
0

### Concurrent diseases

#### Sensitivity analysis 1 − Prevalent cohort 2015

| | Alcohol abuse | Anxiety disorders | Bipolar disorder | Brain tumors | Cerebral palsy | Dementia | Depression | Developmental and behavioural disorders | Drug abuse | Eating disorders | Epilepsy | Headache | Infections of the CNS | Intellectual disability | Multiple sclerosis | Neuromuscular disorders | Other neurodegenerative disorders | Parkinson's disease | Personality disorders | Polyneuropathy | Schizophrenia spectrum disorders | Sleep disorders | Stress related disorders | Stroke | Traumatic brain injury |
|---|---|---|---|---|---|---|---|---|---|---|---|---|---|---|---|---|---|---|---|---|---|---|---|---|---|
| Alcohol abuse | | 12.3 | 22 | 2.7 | 1.8 | 9.6 | 14.2 | 4.1 | 33.3 | 4.6 | 8.8 | 2.6 | 3 | 4.9 | 2.2 | 4.2 | 6.8 | 3.6 | 17.6 | 13.6 | 18.9 | 3.4 | 12.4 | 7.3 | 7.2 |
| Anxiety disorders | 11.9 | | 17.7 | 2.6 | 2.7 | 3.2 | 18.4 | 10.8 | 18.2 | 17.6 | 5.1 | 5.2 | 2.7 | 7.8 | 2.6 | 2.8 | 3.5 | 2.9 | 27.2 | 3.4 | 17.5 | 3.5 | 14.3 | 2.6 | 3.6 |
| Bipolar disorder | 5.1 | 4.3 | | 0.9 | 0.6 | 2.5 | 6.5 | 1.7 | 6.9 | 2.5 | 1.5 | 0.9 | 0.6 | 2.4 | 0.7 | 0.9 | 1.1 | 1.6 | 7.3 | 1.2 | 9.8 | 1.1 | 4 | 1.1 | 1.1 |
| Brain tumors | 0.4 | 0.4 | 0.6 | | 0.9 | 1 | 0.5 | 0.2 | 0.3 | 0.2 | 3.9 | 0.9 | 1.5 | 0.5 | 1 | 0.9 | 1 | 1.2 | 0.3 | 0.9 | 0.4 | 0.6 | 0.4 | 1.4 | 0.4 |
| Cerebral palsy | 0.2 | 0.3 | 0.2 | 0.6 | | 0.3 | 0.2 | 1 | 0.2 | 0.2 | 3.7 | 0.2 | 1.1 | 8.3 | 0.3 | 1.9 | 5.8 | 0.4 | 0.2 | 0.4 | 0.3 | 0.3 | 0.3 | 0.4 | 0.4 |
| Dementia | 3.8 | 1.3 | 4.2 | 2.6 | 1.1 | | 4.4 | 0.2 | 1.6 | 0.2 | 3.4 | 0.8 | 1.5 | 2.3 | 1.4 | 1.4 | 10.3 | 15.6 | 1.1 | 3.6 | 3 | 1 | 1.1 | 6.3 | 1.5 |
| Depression | 25.6 | 34.2 | 50.5 | 6 | 4 | 20 | | 11.8 | 30 | 28.9 | 9.4 | 9.8 | 5.2 | 9.6 | 5.9 | 6.3 | 9.7 | 12.9 | 44.8 | 9.9 | 30.4 | 8 | 31.4 | 9.9 | 7.2 |
| Developmental and behavioural disorders | 4.3 | 11.5 | 7.7 | 1.2 | 11.4 | 0.5 | 6.8 | | 16.9 | 9.4 | 7.3 | 2.6 | 3 | 37.4 | 0.6 | 3.2 | 2.2 | 0.5 | 13.2 | 0.8 | 11.1 | 2.7 | 8.9 | 0.5 | 4.3 |
| Drug abuse | 16.5 | 9.4 | 14.7 | 1.1 | 1.2 | 2 | 8.3 | 8.1 | | 5.3 | 3.7 | 2 | 1.7 | 4.2 | 1 | 1.5 | 1.2 | 1.3 | 18.1 | 2.3 | 22.1 | 1.3 | 9.1 | 1.4 | 3.8 |
| Eating disorders | 0.7 | 2.9 | 1.7 | 0.2 | 0.3 | 0.1 | 2.6 | 1.4 | 1.7 | | 0.6 | 0.7 | 0.5 | 0.7 | 0.3 | 0.4 | 0.3 | 0.1 | 5.2 | 0.3 | 1.9 | 0.2 | 1.9 | 0.1 | 0.6 |
| Epilepsy | 6.4 | 3.8 | 4.5 | 18.9 | 29.5 | 6.2 | 3.8 | 5.1 | 5.5 | 2.7 | | 3.9 | 6.2 | 21.4 | 3.5 | 4.3 | 6 | 4.8 | 4.3 | 3.9 | 5.8 | 2.7 | 3.4 | 8.3 | 4.3 |
| Headache | 2.6 | 5.3 | 3.7 | 6.2 | 2.1 | 1.9 | 5.4 | 2.5 | 4 | 4.4 | 5.3 | | 5.6 | 1.7 | 4.6 | 3.7 | 3.4 | 1.8 | 5.1 | 3.5 | 3.2 | 3.6 | 5.2 | 5.3 | 4.1 |
| Infections of the CNS | 0.8 | 0.8 | 0.7 | 2.7 | 3.1 | 1 | 0.8 | 0.8 | 0.9 | 0.8 | 2.3 | 1.5 | | 1.2 | 2.3 | 1.5 | 1.7 | 0.8 | 0.7 | 1.9 | 0.7 | 0.7 | 0.8 | 1.5 | 0.9 |
| Intellectual disability | 1.3 | 2.2 | 2.9 | 0.9 | 24.8 | 1.6 | 1.5 | 9.9 | 2.4 | 1.2 | 8.1 | 0.5 | 1.2 | | 0.3 | 2.7 | 2.8 | 0.8 | 2.4 | 0.5 | 5.9 | 0.9 | 2.1 | 0.5 | 1.1 |
| Multiple sclerosis | 0.3 | 0.4 | 0.4 | 1 | 0.5 | 0.5 | 0.5 | 0.1 | 0.3 | 0.3 | 0.7 | 0.7 | 1.3 | 0.2 | | 1 | 3.5 | 0.5 | 0.4 | 1.3 | 0.4 | 0.4 | 0.4 | 0.7 | 0.3 |
| Neuromuscular disorders | 0.3 | 0.2 | 0.3 | 0.4 | 1.5 | 0.3 | 0.3 | 0.2 | 0.2 | 0.2 | 0.4 | 0.3 | 0.4 | 0.7 | 0.5 | | 3 | 0.6 | 0.2 | 1.8 | 0.2 | 0.5 | 0.2 | 0.3 | 0.2 |
| Other neurodegenerative disorders | 0.2 | 0.1 | 0.1 | 0.2 | 1.4 | 0.6 | 0.1 | 0 | 0.1 | 0 | 0.2 | 0.1 | 0.1 | 0.2 | 0.5 | 0.9 | | 0.7 | 0.1 | 0.6 | 0.1 | 0.1 | 0.1 | 0.1 | 0.1 |
| Parkinson's disease | 0.4 | 0.3 | 0.7 | 0.8 | 0.4 | 3.9 | 0.7 | 0 | 0.3 | 0 | 0.6 | 0.2 | 0.3 | 0.3 | 0.3 | 0.7 | 3.1 | | 0.2 | 1.3 | 0.5 | 0.5 | 0.2 | 1 | 0.3 |
| Personality disorders | 12.5 | 20 | 22.4 | 1.3 | 1.4 | 1.9 | 17.6 | 9 | 25.9 | 23 | 4.2 | 3.7 | 1.9 | 6.3 | 2.1 | 2.3 | 2.2 | 1.4 | | 2.3 | 22.9 | 2.7 | 16.1 | 1.5 | 3.7 |
| Polyneuropathy | 4.6 | 1.2 | 1.8 | 2 | 1.5 | 3 | 1.8 | 0.3 | 1.5 | 0.6 | 1.8 | 1.2 | 2.5 | 0.6 | 3 | 8.5 | 8.4 | 4.4 | 1.1 | | 1.1 | 1.9 | 1.2 | 3 | 1 |
| Schizophrenia spectrum disorders | 11.4 | 11 | 25.6 | 1.6 | 2.3 | 4.6 | 10.3 | 6.5 | 27 | 7.3 | 4.8 | 2 | 1.6 | 12.9 | 1.6 | 1.7 | 3.5 | 3.2 | 19.6 | 2 | | 1.9 | 10 | 1.8 | 2.7 |
| Sleep disorders | 2.5 | 2.7 | 3.5 | 3 | 2.6 | 1.8 | 3.3 | 1.9 | 2 | 1 | 2.7 | 2.7 | 2 | 2.4 | 1.8 | 4.9 | 4 | 3.7 | 2.8 | 4.3 | 2.3 | | 2.6 | 3.1 | 1.6 |
| Stress related disorders | 24.1 | 29 | 33.5 | 5.2 | 5.7 | 5.6 | 34.1 | 16.8 | 35.8 | 23.5 | 9.3 | 10.3 | 5.8 | 14.6 | 5.8 | 5.5 | 7.7 | 4.5 | 44.3 | 6.7 | 32.1 | 6.9 | | 5.3 | 8.8 |
| Stroke | 8.7 | 3.3 | 5.4 | 11.2 | 5.7 | 18.6 | 6.6 | 0.5 | 3.5 | 0.7 | 13.6 | 6.4 | 6.5 | 2.1 | 5.8 | 5.4 | 6.2 | 12.2 | 2.5 | 10.5 | 3.5 | 5 | 3.2 | | 4.4 |
| Traumatic brain injury | 17.9 | 9.1 | 11.2 | 6.4 | 10.3 | 9.1 | 9.9 | 10.3 | 18.9 | 9.3 | 14.6 | 10.4 | 8 | 9.5 | 5.1 | 7.3 | 9.3 | 6.3 | 13.1 | 7.6 | 10.9 | 5.3 | 11.2 | 9 | |
| Myocardial infarction | 2.4 | 1.3 | 1.5 | 2 | 0.4 | 4.1 | 1.9 | 0.1 | 1.1 | 0.2 | 1.5 | 1.2 | 1.3 | 0.3 | 1.3 | 2.7 | 2.1 | 4 | 0.9 | 3.9 | 1 | 3.1 | 1.2 | 5 | 1.1 |
| Congestive heart failure | 3.6 | 1.5 | 2.1 | 2.4 | 0.6 | 6.8 | 2.5 | 0.2 | 1.3 | 0.2 | 2.1 | 1.1 | 1.5 | 0.8 | 1.1 | 4.6 | 2.3 | 5.6 | 0.9 | 6.1 | 1.5 | 3.7 | 1.2 | 7.2 | 1.3 |
| Peripheral vascular disease | 4.8 | 1.5 | 2.3 | 3.1 | 0.9 | 6.2 | 2.8 | 0.3 | 1.8 | 0.9 | 2.6 | 2 | 2.1 | 0.6 | 2 | 3.5 | 3.9 | 5.5 | 1.2 | 9.5 | 1.5 | 3.3 | 1.5 | 9.1 | 1.5 |
| Cerebrovascular disease | 9.8 | 3.9 | 6.3 | 13 | 6.2 | 24.6 | 7.8 | 0.6 | 3.9 | 0.8 | 14.9 | 7.2 | 6.8 | 2.3 | 5.8 | 6.9 | 9 | 16.9 | 2.9 | 13.3 | 4 | 6.3 | 3.7 | 79 | 4.9 |
| Chronic pulmonary disease | 12.2 | 7.6 | 8.6 | 6.5 | 8.4 | 11.1 | 8.7 | 5.5 | 8.9 | 4.4 | 7.3 | 6.8 | 6 | 6.1 | 4.5 | 15.4 | 15.3 | 8.1 | 7.5 | 12 | 7.4 | 10.6 | 6.9 | 10.7 | 5.6 |
| Connective tissue disease | 1.8 | 2 | 2.1 | 3.1 | 2 | 3.9 | 2.8 | 1.2 | 1.3 | 1.6 | 2.2 | 3.1 | 3.4 | 1.5 | 2.4 | 9.2 | 3 | 3.5 | 1.9 | 6.5 | 1.4 | 2.5 | 2 | 3.9 | 1.6 |
| Ulcer disease | 5.1 | 1.6 | 2.2 | 1.6 | 1.1 | 4.8 | 2.4 | 0.3 | 2.3 | 0.5 | 1.9 | 1.5 | 1.3 | 1 | 1.3 | 2.2 | 2.2 | 3.2 | 1.5 | 4 | 1.8 | 1.6 | 1.5 | 4 | 1.2 |
| Mild liver disease | 11.4 | 1.7 | 2.5 | 0.8 | 0.6 | 1.5 | 2 | 0.7 | 7.8 | 0.8 | 1.8 | 1 | 1.2 | 0.6 | 0.7 | 2.2 | 1.3 | 0.7 | 2.4 | 3.6 | 3 | 1 | 1.9 | 1.5 | 1.4 |
| Diabetes without end−organ damage | 8.5 | 4.3 | 6.8 | 6.1 | 1.7 | 11.6 | 6.4 | 1.1 | 4.2 | 1.7 | 4.7 | 3.7 | 4 | 3.3 | 3.4 | 6.8 | 5.8 | 9.2 | 4.5 | 16.8 | 6.3 | 10.7 | 4.2 | 12.4 | 3 |
| Hemiplegia | 0.4 | 0.2 | 0.3 | 2.6 | 24.9 | 0.5 | 0.4 | 0.3 | 0.4 | 0.1 | 2.4 | 0.3 | 1.7 | 3.1 | 2.2 | 3.1 | 7.9 | 0.8 | 0.2 | 1.2 | 0.3 | 0.6 | 0.3 | 1.2 | 0.4 |
| Moderate to severe renal disease | 2.7 | 1.2 | 3.1 | 2 | 1.2 | 4.2 | 2.1 | 0.5 | 1.6 | 0.9 | 2.1 | 1.3 | 1.9 | 1.1 | 1.1 | 3.9 | 1.6 | 3.3 | 1.1 | 5.9 | 1.5 | 2.5 | 1.2 | 5 | 1.1 |
| Diabetes with end−organ damage | 4.2 | 1.8 | 3 | 2.5 | 0.8 | 6 | 3 | 0.4 | 1.9 | 0.8 | 2.5 | 1.6 | 1.9 | 1.4 | 1.6 | 3.6 | 3 | 4.9 | 1.9 | 12.7 | 2.6 | 5.1 | 1.8 | 7.7 | 1.4 |
| Solid cancer tumour | 7.1 | 3.9 | 5.4 | 23.1 | 1.7 | 11.9 | 5.6 | 0.4 | 2.5 | 1.1 | 5.6 | 3.9 | 4.8 | 1.4 | 4.8 | 6.7 | 6.4 | 12.9 | 2.4 | 12.2 | 3.1 | 5.8 | 3.4 | 11.3 | 3.1 |
| Leukaemia | 0.2 | 0.1 | 0.2 | 0.6 | 0.1 | 0.3 | 0.2 | 0 | 0.1 | 0.1 | 0.2 | 0.1 | 0.3 | 0.1 | 0.1 | 0.3 | 0.4 | 0.5 | 0.1 | 0.6 | 0.1 | 0.2 | 0.1 | 0.3 | 0.1 |
| Lymphoma | 0.3 | 0.3 | 0.3 | 1.7 | 0.1 | 0.6 | 0.4 | 0.1 | 0.2 | 0.1 | 0.3 | 0.3 | 0.6 | 0.1 | 0.2 | 0.7 | 0.3 | 0.8 | 0.2 | 1.6 | 0.3 | 0.4 | 0.3 | 0.7 | 0.2 |
| Moderate to severe liver disease | 4.2 | 0.4 | 0.6 | 0.2 | 0.2 | 0.6 | 0.5 | 0.1 | 0.9 | 0.1 | 0.5 | 0.1 | 0.3 | 0.1 | 0.2 | 0.9 | 0.5 | 0.3 | 0.4 | 1.4 | 0.4 | 0.2 | 0.4 | 0.5 | 0.3 |
| Metastatic solid tumour | 0.8 | 0.6 | 0.6 | 3 | 0.2 | 0.7 | 0.6 | 0.1 | 0.3 | 0.1 | 0.7 | 0.4 | 0.5 | 0.2 | 0.5 | 0.6 | 0.5 | 0.8 | 0.2 | 1.4 | 0.3 | 0.5 | 0.4 | 1 | 0.3 |
| AIDS | 0.3 | 0.1 | 0.2 | 0.1 | 0 | 0.1 | 0.2 | 0.1 | 0.7 | 0.1 | 0.2 | 0.1 | 0.7 | 0.1 | 0 | 0.2 | 0 | 0.2 | 0.2 | 0.4 | 0.4 | 0.1 | 0.2 | 0.1 | 0.1 |

Percent: 0, 20, 40, 60

### Concurrent diseases

#### Sensitivity analysis 1 − Prevalent cohort 2016

| | Alcohol abuse | Anxiety disorders | Bipolar disorder | Brain tumors | Cerebral palsy | Dementia | Depression | Developmental and behavioural disorders | Drug abuse | Eating disorders | Epilepsy | Headache | Infections of the CNS | Intellectual disability | Multiple sclerosis | Neuromuscular disorders | Other neurodegenerative disorders | Parkinson's disease | Personality disorders | Polyneuropathy | Schizophrenia spectrum disorders | Sleep disorders | Stress related disorders | Stroke | Traumatic brain injury |
|---|---|---|---|---|---|---|---|---|---|---|---|---|---|---|---|---|---|---|---|---|---|---|---|---|---|
| Alcohol abuse | | 11.7 | 21.6 | 2.6 | 1.8 | 9.6 | 13.8 | 4.2 | 32.5 | 4.5 | 8.6 | 2.6 | 3 | 5 | 2.2 | 4 | 6.8 | 3.7 | 17 | 13.1 | 18.6 | 3.4 | 12 | 7.3 | 7.2 |
| Anxiety disorders | 12.2 | | 18.3 | 2.8 | 2.9 | 3.4 | 19.1 | 11.7 | 18.9 | 18.5 | 5.3 | 5.5 | 2.9 | 8.4 | 2.7 | 3.1 | 3.5 | 3 | 28.4 | 3.5 | 18.3 | 3.8 | 14.9 | 2.8 | 3.8 |
| Bipolar disorder | 5.2 | 4.3 | | 0.9 | 0.6 | 2.4 | 6.5 | 1.9 | 7 | 2.7 | 1.5 | 0.9 | 0.7 | 2.4 | 0.8 | 0.9 | 1.1 | 1.6 | 7.5 | 1.2 | 9.6 | 1.1 | 4 | 1 | 1.1 |
| Brain tumors | 0.4 | 0.4 | 0.6 | | 0.9 | 1 | 0.5 | 0.2 | 0.3 | 0.2 | 4 | 1 | 1.6 | 0.5 | 1 | 0.9 | 1.1 | 1.3 | 0.3 | 0.9 | 0.4 | 0.6 | 0.4 | 1.4 | 0.4 |
| Cerebral palsy | 0.2 | 0.3 | 0.2 | 0.6 | | 0.3 | 0.2 | 1 | 0.2 | 0.2 | 3.8 | 0.2 | 1.1 | 7.9 | 0.3 | 1.8 | 5.6 | 0.3 | 0.2 | 0.4 | 0.3 | 0.3 | 0.3 | 0.5 | 0.4 |
| Dementia | 3.9 | 1.3 | 4 | 2.5 | 1.3 | | 4.2 | 0.2 | 1.5 | 0.2 | 3.5 | 0.8 | 1.5 | 2.4 | 1.3 | 1.5 | 9.2 | 15.3 | 1 | 3.5 | 2.9 | 1 | 1.1 | 6.1 | 1.5 |
| Depression | 25.8 | 34.4 | 50 | 6.1 | 4.1 | 19.5 | | 12.4 | 30.1 | 29.4 | 9.6 | 10.1 | 5.5 | 9.8 | 5.9 | 6.4 | 9.6 | 12.8 | 45.5 | 9.9 | 30.6 | 8.3 | 31.6 | 9.8 | 7.4 |
| Developmental and behavioural disorders | 4.7 | 12.6 | 8.7 | 1.2 | 11.6 | 0.5 | 7.5 | | 18.4 | 10.2 | 7.6 | 2.8 | 3.1 | 38.7 | 0.8 | 3.3 | 2.2 | 0.5 | 14.4 | 0.8 | 12.2 | 2.9 | 9.7 | 0.5 | 4.6 |
| Drug abuse | 16.7 | 9.3 | 14.8 | 1.1 | 1.2 | 1.9 | 8.3 | 8.4 | | 5.3 | 3.8 | 2 | 1.7 | 4.6 | 1 | 1.5 | 1.4 | 1.3 | 18.3 | 2.2 | 22.5 | 1.4 | 9.1 | 1.5 | 3.9 |
| Eating disorders | 0.8 | 3 | 1.9 | 0.2 | 0.4 | 0.1 | 2.6 | 1.5 | 1.7 | | 0.6 | 0.7 | 0.5 | 0.7 | 0.3 | 0.4 | 0.2 | 0.1 | 5.3 | 0.3 | 2 | 0.3 | 1.9 | 0.1 | 0.6 |
| Epilepsy | 6.3 | 3.7 | 4.4 | 18.5 | 29.7 | 6.3 | 3.7 | 4.9 | 5.4 | 2.6 | | 3.8 | 6.2 | 21.1 | 3.5 | 4.2 | 5.7 | 4.6 | 4.2 | 3.7 | 5.7 | 2.6 | 3.4 | 8.2 | 4.3 |
| Headache | 2.6 | 5.4 | 3.9 | 6.3 | 2.3 | 2 | 5.6 | 2.6 | 4 | 4.5 | 5.3 | | 5.8 | 1.8 | 4.6 | 4 | 3.4 | 2 | 5.3 | 3.6 | 3.4 | 3.8 | 5.4 | 5.4 | 4.3 |
| Infections of the CNS | 0.8 | 0.8 | 0.7 | 2.8 | 3.2 | 1 | 0.8 | 0.8 | 0.9 | 0.8 | 2.3 | 1.5 | | 1.2 | 2.3 | 1.6 | 1.5 | 0.8 | 0.7 | 2 | 0.7 | 0.7 | 0.8 | 1.5 | 0.9 |
| Intellectual disability | 1.4 | 2.3 | 2.8 | 0.9 | 24.1 | 1.6 | 1.5 | 9.7 | 2.5 | 1.3 | 8.2 | 0.5 | 1.2 | | 0.3 | 2.6 | 2.7 | 0.7 | 2.5 | 0.5 | 5.9 | 0.9 | 2.1 | 0.5 | 1.1 |
| Multiple sclerosis | 0.3 | 0.4 | 0.5 | 1 | 0.5 | 0.5 | 0.5 | 0.1 | 0.3 | 0.3 | 0.7 | 0.7 | 1.3 | 0.2 | | 1 | 3.2 | 0.4 | 0.4 | 1.3 | 0.4 | 0.4 | 0.4 | 0.7 | 0.3 |
| Neuromuscular disorders | 0.3 | 0.2 | 0.3 | 0.4 | 1.5 | 0.3 | 0.3 | 0.2 | 0.2 | 0.2 | 0.4 | 0.3 | 0.4 | 0.7 | 0.5 | | 3.2 | 0.5 | 0.2 | 1.9 | 0.2 | 0.5 | 0.2 | 0.4 | 0.2 |
| Other neurodegenerative disorders | 0.2 | 0.1 | 0.1 | 0.2 | 1.4 | 0.5 | 0.1 | 0 | 0.1 | 0 | 0.2 | 0.1 | 0.1 | 0.2 | 0.5 | 1 | | 0.7 | 0.1 | 0.6 | 0.1 | 0.1 | 0.1 | 0.1 | 0.1 |
| Parkinson's disease | 0.4 | 0.3 | 0.7 | 0.8 | 0.4 | 3.8 | 0.7 | 0 | 0.3 | 0 | 0.6 | 0.2 | 0.3 | 0.3 | 0.3 | 0.7 | 2.9 | | 0.2 | 1.3 | 0.5 | 0.5 | 0.2 | 1 | 0.3 |
| Personality disorders | 12.3 | 19.8 | 22.4 | 1.3 | 1.4 | 1.8 | 17.6 | 9.4 | 25.8 | 23 | 4.2 | 3.8 | 1.9 | 6.5 | 2 | 2.3 | 2.2 | 1.3 | | 2.2 | 22.5 | 2.7 | 15.9 | 1.5 | 3.8 |
| Polyneuropathy | 4.7 | 1.2 | 1.8 | 2.1 | 1.5 | 3.1 | 1.9 | 0.3 | 1.5 | 0.5 | 1.8 | 1.2 | 2.6 | 0.6 | 3 | 8.8 | 8.6 | 4.6 | 1.1 | | 1.2 | 2.1 | 1.2 | 3.1 | 1.1 |
| Schizophrenia spectrum disorders | 11.6 | 10.9 | 24.8 | 1.5 | 2.3 | 4.4 | 10.2 | 6.8 | 27.2 | 7.4 | 4.9 | 2.1 | 1.6 | 13.1 | 1.6 | 1.7 | 3.3 | 3.2 | 19.3 | 2 | | 2 | 9.9 | 1.8 | 2.7 |
| Sleep disorders | 2.8 | 3 | 3.8 | 3.3 | 2.8 | 2 | 3.7 | 2.1 | 2.2 | 1.2 | 3 | 3 | 2.2 | 2.6 | 1.9 | 5.6 | 5 | 4 | 3.1 | 4.9 | 2.6 | | 2.9 | 3.4 | 1.8 |
| Stress related disorders | 24.7 | 29.7 | 34.4 | 5.3 | 6.3 | 5.7 | 35 | 18 | 36.6 | 24.1 | 9.6 | 10.8 | 6 | 15.5 | 5.9 | 5.9 | 7.9 | 4.7 | 45.4 | 6.9 | 33 | 7.3 | | 5.4 | 9.2 |
| Stroke | 8.8 | 3.2 | 5.2 | 11 | 6.2 | 18.2 | 6.3 | 0.5 | 3.4 | 0.7 | 13.6 | 6.3 | 6.6 | 2.1 | 5.8 | 5.6 | 6.6 | 12.2 | 2.5 | 10.5 | 3.5 | 5 | 3.2 | | 4.5 |
| Traumatic brain injury | 17.5 | 9 | 10.9 | 6.4 | 10.3 | 9.2 | 9.7 | 10.1 | 18.4 | 9 | 14.4 | 10.3 | 7.9 | 9.4 | 5.1 | 7.1 | 9 | 6.4 | 12.7 | 7.4 | 10.8 | 5.3 | 10.8 | 9.1 | |
| Myocardial infarction | 2.5 | 1.3 | 1.5 | 1.9 | 0.4 | 4.1 | 1.9 | 0.1 | 1.1 | 0.2 | 1.6 | 1.3 | 1.4 | 0.3 | 1.3 | 2.6 | 2.2 | 3.8 | 0.9 | 3.9 | 1 | 3.1 | 1.2 | 4.8 | 1.1 |
| Congestive heart failure | 3.7 | 1.5 | 2 | 2.4 | 0.7 | 6.8 | 2.4 | 0.2 | 1.3 | 0.2 | 2.2 | 1 | 1.5 | 0.7 | 1.1 | 4.7 | 2.2 | 5.7 | 0.9 | 6.1 | 1.5 | 3.8 | 1.1 | 7.2 | 1.3 |
| Peripheral vascular disease | 4.9 | 1.6 | 2.2 | 3.2 | 0.8 | 6.3 | 2.8 | 0.3 | 1.8 | 0.8 | 2.6 | 2 | 2.1 | 0.5 | 2.1 | 3.6 | 4.1 | 5.3 | 1.2 | 9.5 | 1.4 | 3.4 | 1.5 | 9.1 | 1.5 |
| Cerebrovascular disease | 9.9 | 3.8 | 6.2 | 12.6 | 6.9 | 24.2 | 7.6 | 0.6 | 3.9 | 0.8 | 14.9 | 7.1 | 6.9 | 2.3 | 5.8 | 6.9 | 9.2 | 16.8 | 2.9 | 13.2 | 4 | 6.3 | 3.7 | 78.4 | 5.1 |
| Chronic pulmonary disease | 12.5 | 7.6 | 8.6 | 6.7 | 8.5 | 11.4 | 8.8 | 5.5 | 8.9 | 4.4 | 7.5 | 7 | 6.1 | 6.2 | 4.6 | 16.1 | 15.7 | 8.7 | 7.6 | 12.2 | 7.5 | 10.9 | 7.1 | 11 | 5.7 |
| Connective tissue disease | 1.8 | 2.1 | 2.2 | 3.1 | 1.9 | 4 | 2.8 | 1.2 | 1.3 | 1.7 | 2.3 | 3.2 | 3.5 | 1.6 | 2.5 | 9.6 | 2.6 | 3.6 | 2 | 6.7 | 1.4 | 2.6 | 2.1 | 4 | 1.6 |
| Ulcer disease | 4.9 | 1.5 | 2 | 1.5 | 1.2 | 4.5 | 2.3 | 0.3 | 2.2 | 0.5 | 1.8 | 1.4 | 1.3 | 0.9 | 1.2 | 2.2 | 2 | 3.5 | 1.5 | 3.8 | 1.7 | 1.6 | 1.4 | 3.7 | 1.2 |
| Mild liver disease | 11.4 | 1.7 | 2.4 | 0.8 | 0.6 | 1.5 | 2 | 0.7 | 7.5 | 0.8 | 1.9 | 1 | 1.2 | 0.6 | 0.8 | 2.2 | 1.2 | 0.8 | 2.4 | 3.6 | 3 | 1 | 1.9 | 1.5 | 1.4 |
| Diabetes without end−organ damage | 8.6 | 4.3 | 6.8 | 6.1 | 2 | 11.8 | 6.4 | 1.1 | 4.2 | 1.6 | 4.9 | 3.8 | 4.1 | 3.3 | 3.4 | 7 | 5.8 | 9.2 | 4.5 | 17.5 | 6.4 | 11 | 4.3 | 12.6 | 3.1 |
| Hemiplegia | 0.4 | 0.2 | 0.3 | 2.5 | 24.6 | 0.5 | 0.4 | 0.3 | 0.3 | 0.1 | 2.4 | 0.3 | 1.7 | 3 | 2.2 | 3.1 | 7.2 | 0.8 | 0.2 | 1.2 | 0.3 | 0.6 | 0.3 | 1.2 | 0.5 |
| Moderate to severe renal disease | 2.9 | 1.3 | 3.2 | 2.1 | 1.3 | 4.6 | 2.2 | 0.5 | 1.7 | 0.8 | 2.2 | 1.4 | 2.1 | 1.2 | 1.3 | 4.2 | 2.2 | 3.7 | 1.2 | 6.1 | 1.6 | 2.7 | 1.3 | 5.2 | 1.2 |
| Diabetes with end−organ damage | 4.2 | 1.7 | 2.9 | 2.4 | 0.9 | 5.8 | 2.9 | 0.4 | 1.8 | 0.8 | 2.5 | 1.5 | 1.9 | 1.3 | 1.6 | 3.5 | 2.8 | 4.8 | 1.9 | 12.7 | 2.6 | 5.3 | 1.8 | 7.6 | 1.4 |
| Solid cancer tumour | 7.6 | 4.1 | 5.6 | 23.4 | 1.8 | 12.6 | 5.7 | 0.4 | 2.5 | 1.1 | 5.9 | 4.1 | 5.3 | 1.5 | 5 | 7.1 | 6.8 | 13.7 | 2.5 | 12.9 | 3.2 | 6.1 | 3.5 | 11.8 | 3.2 |
| Leukaemia | 0.2 | 0.1 | 0.2 | 0.6 | 0.1 | 0.3 | 0.2 | 0 | 0.1 | 0.1 | 0.2 | 0.1 | 0.4 | 0.1 | 0.1 | 0.3 | 0.5 | 0.4 | 0.1 | 0.7 | 0.1 | 0.2 | 0.1 | 0.4 | 0.1 |
| Lymphoma | 0.4 | 0.3 | 0.3 | 1.9 | 0.1 | 0.7 | 0.4 | 0.1 | 0.2 | 0.1 | 0.4 | 0.3 | 0.7 | 0.1 | 0.2 | 0.8 | 0.6 | 0.8 | 0.2 | 1.7 | 0.3 | 0.5 | 0.3 | 0.7 | 0.2 |
| Moderate to severe liver disease | 4.2 | 0.4 | 0.5 | 0.2 | 0.3 | 0.6 | 0.5 | 0.1 | 0.8 | 0.1 | 0.5 | 0.1 | 0.4 | 0.1 | 0.2 | 0.9 | 0.5 | 0.3 | 0.4 | 1.4 | 0.4 | 0.2 | 0.4 | 0.5 | 0.3 |
| Metastatic solid tumour | 0.9 | 0.6 | 0.5 | 2.9 | 0.2 | 0.7 | 0.6 | 0.1 | 0.2 | 0.1 | 0.7 | 0.4 | 0.6 | 0.2 | 0.5 | 0.6 | 0.6 | 0.9 | 0.2 | 1.6 | 0.3 | 0.6 | 0.4 | 1 | 0.3 |
| AIDS | 0.3 | 0.1 | 0.2 | 0.1 | 0 | 0.1 | 0.2 | 0.1 | 0.6 | 0.1 | 0.2 | 0.1 | 0.7 | 0.1 | 0 | 0.1 | 0 | 0.1 | 0.2 | 0.4 | 0.4 | 0.1 | 0.2 | 0.1 | 0.1 |

Percent: 0, 20, 40, 60

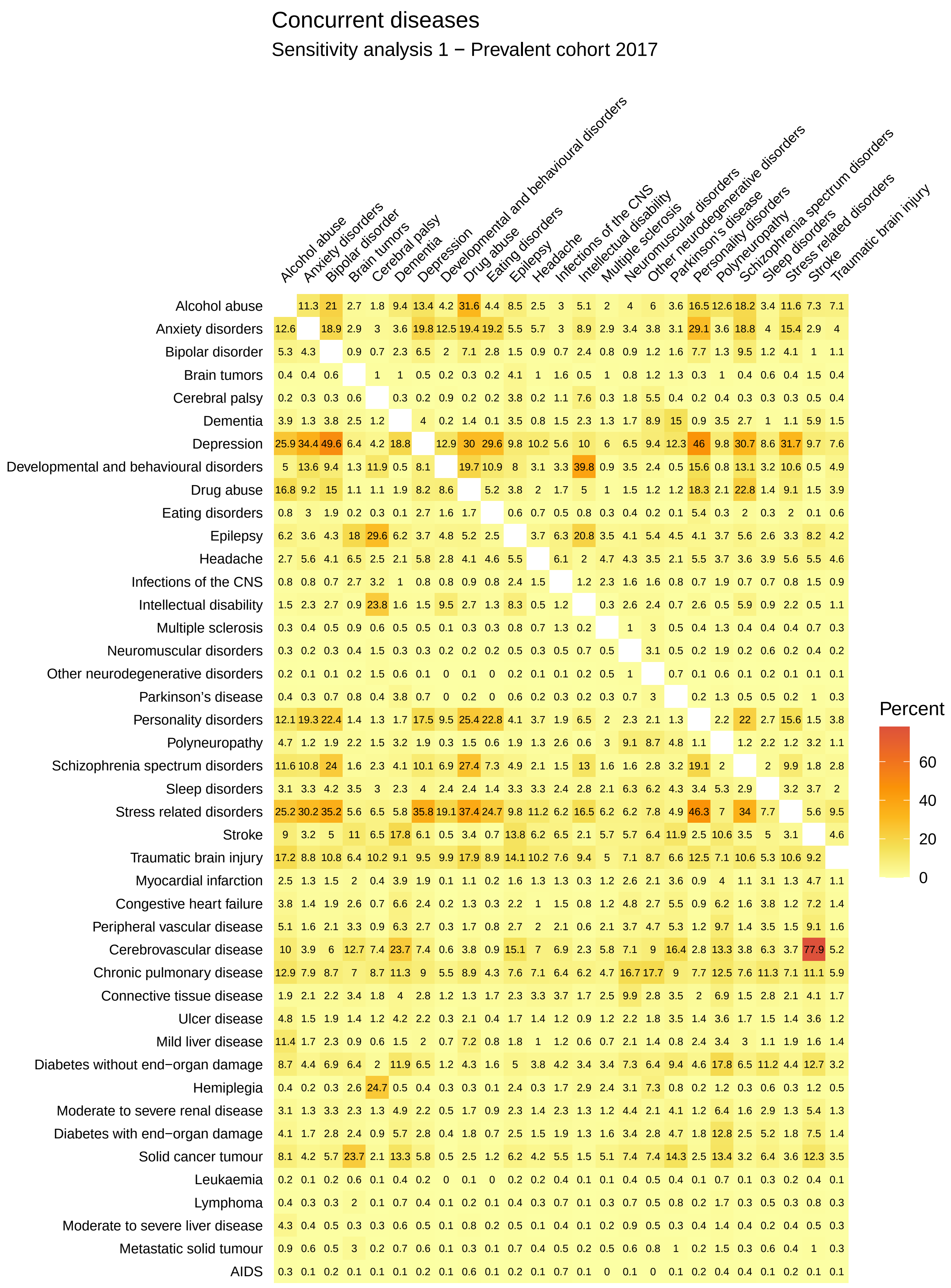
Concurrent diseases
Sensitivity analysis 1 − Prevalent cohort 2017
Alcohol abuse
Anxiety disorders
Bipolar disorder
Brain tumors
Cerebral palsy
Dementia
Depression
Developmental and behavioural disorders
Drug abuse
Eating disorders
Epilepsy
Headache
Infections of the CNS
Intellectual disability
Multiple sclerosis
Neuromuscular disorders
Other neurodegenerative disorders
Parkinson's disease
Personality disorders
Polyneuropathy
Schizophrenia spectrum disorders
Sleep disorders
Stress related disorders
Stroke
Traumatic brain injury
Myocardial infarction
Congestive heart failure
Peripheral vascular disease
Cerebrovascular disease
Chronic pulmonary disease
Connective tissue disease
Ulcer disease
Mild liver disease
Diabetes without end−organ damage
Hemiplegia
Moderate to severe renal disease
Diabetes with end−organ damage
Solid cancer tumour
Leukaemia
Lymphoma
Moderate to severe liver disease
Metastatic solid tumour
AIDS
Percent
60
40
20
0

### Concurrent diseases

#### Sensitivity analysis 1 − Prevalent cohort 2018

| | Alcohol abuse | Anxiety disorders | Bipolar disorder | Brain tumors | Cerebral palsy | Dementia | Depression | Developmental and behavioural disorders | Drug abuse | Eating disorders | Epilepsy | Headache | Infections of the CNS | Intellectual disability | Multiple sclerosis | Neuromuscular disorders | Other neurodegenerative disorders | Parkinson's disease | Personality disorders | Polyneuropathy | Schizophrenia spectrum disorders | Sleep disorders | Stress related disorders | Stroke | Traumatic brain injury |
|---|---|---|---|---|---|---|---|---|---|---|---|---|---|---|---|---|---|---|---|---|---|---|---|---|---|
| Alcohol abuse | | 10.8 | 20.6 | 2.7 | 1.9 | 9.1 | 12.9 | 4.2 | 30.8 | 4.2 | 8.2 | 2.4 | 3 | 5.1 | 2 | 3.9 | 6 | 3.6 | 15.9 | 12.1 | 17.7 | 3.4 | 11.1 | 7.2 | 6.9 |
| Anxiety disorders | 12.8 | | 19.4 | 3.1 | 3.2 | 3.6 | 20.3 | 13.2 | 19.8 | 19.8 | 5.6 | 6 | 3.1 | 9.1 | 3 | 3.6 | 4.1 | 3.1 | 29.9 | 3.7 | 19.5 | 4.2 | 15.9 | 3 | 4.2 |
| Bipolar disorder | 5.4 | 4.3 | | 0.9 | 0.7 | 2.3 | 6.4 | 2 | 7.1 | 2.9 | 1.4 | 0.9 | 0.8 | 2.3 | 0.8 | 1 | 1.2 | 1.6 | 7.7 | 1.3 | 9.3 | 1.2 | 4 | 1 | 1.1 |
| Brain tumors | 0.5 | 0.5 | 0.6 | | 1.1 | 1.1 | 0.6 | 0.2 | 0.3 | 0.2 | 4.2 | 1 | 1.6 | 0.5 | 1 | 0.8 | 1.2 | 1.4 | 0.3 | 1 | 0.4 | 0.7 | 0.4 | 1.5 | 0.5 |
| Cerebral palsy | 0.2 | 0.3 | 0.2 | 0.6 | | 0.3 | 0.2 | 0.9 | 0.2 | 0.2 | 3.8 | 0.2 | 1.1 | 7.5 | 0.4 | 1.8 | 5.2 | 0.4 | 0.2 | 0.4 | 0.3 | 0.3 | 0.3 | 0.5 | 0.4 |
| Dementia | 3.8 | 1.3 | 3.6 | 2.6 | 1.2 | | 3.7 | 0.2 | 1.3 | 0.1 | 3.5 | 0.8 | 1.4 | 2.2 | 1.2 | 1.7 | 8.5 | 15.1 | 0.9 | 3.5 | 2.5 | 1.1 | 1.1 | 5.8 | 1.6 |
| Depression | 25.9 | 34.2 | 48.9 | 6.6 | 4.3 | 18.1 | | 13.3 | 29.7 | 29.8 | 9.8 | 10.4 | 5.7 | 10.2 | 6 | 6.7 | 9.8 | 12.1 | 46.3 | 9.9 | 30.6 | 8.9 | 31.6 | 9.6 | 7.8 |
| Developmental and behavioural disorders | 5.5 | 14.6 | 10.3 | 1.3 | 12 | 0.5 | 8.7 | | 21 | 11.8 | 8.3 | 3.3 | 3.5 | 40.9 | 1 | 3.7 | 2.5 | 0.5 | 16.7 | 0.9 | 14.2 | 3.4 | 11.5 | 0.6 | 5.2 |
| Drug abuse | 17 | 9.2 | 15 | 1.1 | 1.1 | 1.8 | 8.2 | 8.8 | | 5.3 | 3.8 | 2 | 1.7 | 5.2 | 1.1 | 1.6 | 1.2 | 1.2 | 18.3 | 2.1 | 22.9 | 1.5 | 9.1 | 1.4 | 4 |
| Eating disorders | 0.8 | 3 | 2 | 0.2 | 0.4 | 0.1 | 2.7 | 1.6 | 1.7 | | 0.6 | 0.8 | 0.5 | 0.8 | 0.3 | 0.4 | 0.1 | 0.1 | 5.4 | 0.3 | 2 | 0.3 | 2 | 0.1 | 0.7 |
| Epilepsy | 6 | 3.5 | 4.1 | 17.6 | 29.5 | 6.1 | 3.6 | 4.6 | 5.1 | 2.5 | | 3.5 | 6.2 | 20.4 | 3.4 | 4 | 5.2 | 4.2 | 4 | 3.6 | 5.5 | 2.6 | 3.2 | 8.1 | 4.2 |
| Headache | 2.8 | 5.8 | 4.2 | 6.6 | 2.6 | 2.1 | 6 | 2.9 | 4.1 | 4.9 | 5.6 | | 6.3 | 2.1 | 4.9 | 4.4 | 3.9 | 2.2 | 5.8 | 3.9 | 3.8 | 4.1 | 5.9 | 5.6 | 4.8 |
| Infections of the CNS | 0.9 | 0.8 | 0.8 | 2.7 | 3.2 | 1 | 0.8 | 0.8 | 0.9 | 0.8 | 2.4 | 1.6 | | 1.1 | 2.4 | 1.6 | 1.6 | 0.8 | 0.7 | 1.9 | 0.7 | 0.7 | 0.8 | 1.5 | 0.9 |
| Intellectual disability | 1.5 | 2.3 | 2.7 | 0.8 | 23.7 | 1.6 | 1.5 | 9.4 | 2.8 | 1.3 | 8.4 | 0.6 | 1.2 | | 0.3 | 2.6 | 2.4 | 0.7 | 2.6 | 0.4 | 5.9 | 0.9 | 2.2 | 0.5 | 1.1 |
| Multiple sclerosis | 0.3 | 0.4 | 0.5 | 0.9 | 0.6 | 0.5 | 0.5 | 0.1 | 0.3 | 0.3 | 0.8 | 0.7 | 1.3 | 0.2 | | 1 | 2.8 | 0.5 | 0.4 | 1.2 | 0.4 | 0.4 | 0.4 | 0.7 | 0.3 |
| Neuromuscular disorders | 0.3 | 0.3 | 0.3 | 0.4 | 1.6 | 0.3 | 0.3 | 0.2 | 0.2 | 0.2 | 0.4 | 0.3 | 0.4 | 0.7 | 0.5 | | 3.1 | 0.5 | 0.2 | 1.9 | 0.2 | 0.5 | 0.2 | 0.4 | 0.2 |
| Other neurodegenerative disorders | 0.2 | 0.1 | 0.1 | 0.2 | 1.5 | 0.6 | 0.1 | 0.1 | 0.1 | 0 | 0.2 | 0.1 | 0.1 | 0.2 | 0.5 | 1 | | 0.8 | 0.1 | 0.6 | 0.1 | 0.2 | 0.1 | 0.1 | 0.1 |
| Parkinson's disease | 0.4 | 0.3 | 0.7 | 0.9 | 0.4 | 4 | 0.7 | 0 | 0.2 | 0 | 0.6 | 0.2 | 0.3 | 0.2 | 0.3 | 0.7 | 3.2 | | 0.2 | 1.4 | 0.5 | 0.5 | 0.2 | 1 | 0.3 |
| Personality disorders | 12 | 18.8 | 22.1 | 1.4 | 1.3 | 1.6 | 17.3 | 9.5 | 24.9 | 22.4 | 4.1 | 3.8 | 1.9 | 6.4 | 2 | 2.3 | 2 | 1.2 | | 2.1 | 21.5 | 2.8 | 15.3 | 1.5 | 3.8 |
| Polyneuropathy | 4.8 | 1.2 | 1.9 | 2.2 | 1.6 | 3.3 | 1.9 | 0.3 | 1.5 | 0.6 | 1.9 | 1.3 | 2.5 | 0.6 | 3 | 9.2 | 9.3 | 4.9 | 1.1 | | 1.1 | 2.3 | 1.2 | 3.3 | 1.2 |
| Schizophrenia spectrum disorders | 11.7 | 10.8 | 23.5 | 1.6 | 2.3 | 4 | 10.1 | 7.1 | 27.4 | 7.4 | 4.9 | 2.2 | 1.6 | 13 | 1.6 | 1.7 | 2.7 | 3.2 | 18.9 | 1.9 | | 2.1 | 10 | 1.8 | 2.8 |
| Sleep disorders | 3.4 | 3.6 | 4.6 | 3.8 | 3.3 | 2.7 | 4.4 | 2.6 | 2.7 | 1.5 | 3.5 | 3.5 | 2.6 | 3.1 | 2.3 | 6.7 | 6.1 | 4.7 | 3.7 | 5.9 | 3.2 | | 3.5 | 4 | 2.2 |
| Stress related disorders | 25.8 | 30.9 | 35.8 | 5.9 | 7 | 5.9 | 36.5 | 20.1 | 38.4 | 25.1 | 10.1 | 11.7 | 6.4 | 17 | 6.4 | 6.6 | 7.6 | 5.2 | 47 | 7.2 | 35 | 8 | | 5.7 | 9.8 |
| Stroke | 9.1 | 3.1 | 4.9 | 10.8 | 6.7 | 17.6 | 6 | 0.5 | 3.3 | 0.7 | 13.9 | 6.1 | 6.4 | 2.1 | 5.5 | 5.6 | 6.6 | 11.7 | 2.5 | 10.6 | 3.4 | 5.1 | 3.1 | | 4.8 |
| Traumatic brain injury | 16.9 | 8.6 | 10.5 | 6.4 | 9.9 | 9.2 | 9.4 | 9.5 | 17.4 | 8.7 | 13.9 | 10.1 | 7.6 | 9.1 | 4.9 | 6.9 | 8.9 | 6.9 | 12.2 | 7.1 | 10.5 | 5.3 | 10.3 | 9.2 | |
| Myocardial infarction | 2.5 | 1.3 | 1.5 | 2 | 0.4 | 3.8 | 1.8 | 0.2 | 1.1 | 0.2 | 1.6 | 1.3 | 1.3 | 0.3 | 1.2 | 2.5 | 2 | 3.4 | 0.9 | 4 | 1.1 | 3 | 1.3 | 4.5 | 1.1 |
| Congestive heart failure | 3.8 | 1.4 | 1.9 | 2.6 | 0.7 | 6.4 | 2.3 | 0.2 | 1.3 | 0.3 | 2.2 | 1 | 1.5 | 0.7 | 1.2 | 4.6 | 2.4 | 5.2 | 0.9 | 6.2 | 1.5 | 3.7 | 1.1 | 7.1 | 1.4 |
| Peripheral vascular disease | 5.2 | 1.6 | 2.1 | 3.3 | 1 | 6.2 | 2.7 | 0.3 | 1.7 | 0.9 | 2.8 | 1.9 | 2.1 | 0.6 | 2 | 3.7 | 4.2 | 5.3 | 1.2 | 9.9 | 1.4 | 3.6 | 1.5 | 9.1 | 1.7 |
| Cerebrovascular disease | 10.1 | 3.8 | 5.9 | 12.5 | 7.6 | 23.3 | 7.2 | 0.6 | 3.7 | 0.9 | 15.1 | 6.8 | 6.8 | 2.3 | 5.6 | 6.8 | 8.8 | 16.1 | 2.7 | 13.3 | 3.7 | 6.4 | 3.6 | 77.8 | 5.3 |
| Chronic pulmonary disease | 13.1 | 8 | 8.7 | 7.3 | 8.8 | 11.1 | 9 | 5.5 | 8.9 | 4.3 | 7.7 | 7.3 | 6.5 | 6.3 | 5 | 16.9 | 17.7 | 8.9 | 7.8 | 12.8 | 7.7 | 11.5 | 7.3 | 11.2 | 6 |
| Connective tissue disease | 1.9 | 2.2 | 2.2 | 3.4 | 1.8 | 4 | 2.8 | 1.3 | 1.3 | 1.8 | 2.4 | 3.3 | 3.7 | 1.6 | 2.5 | 10 | 2.9 | 3.7 | 2 | 7 | 1.5 | 2.9 | 2.2 | 4.2 | 1.8 |
| Ulcer disease | 4.6 | 1.4 | 1.9 | 1.4 | 1.3 | 4 | 2 | 0.3 | 2 | 0.4 | 1.7 | 1.3 | 1.2 | 0.9 | 1.1 | 2.1 | 1.9 | 3.2 | 1.4 | 3.5 | 1.6 | 1.5 | 1.3 | 3.4 | 1.1 |
| Mild liver disease | 11.5 | 1.7 | 2.3 | 1 | 0.5 | 1.4 | 2 | 0.7 | 7 | 0.8 | 1.8 | 1 | 1.2 | 0.6 | 0.7 | 2 | 1.5 | 0.8 | 2.3 | 3.5 | 3 | 1.1 | 1.9 | 1.6 | 1.4 |
| Diabetes without end−organ damage | 8.7 | 4.4 | 6.9 | 6.4 | 2 | 12.2 | 6.5 | 1.2 | 4.2 | 1.7 | 5.1 | 3.7 | 4.1 | 3.4 | 3.4 | 7.2 | 6.8 | 9.5 | 4.5 | 18.1 | 6.5 | 11.4 | 4.4 | 12.7 | 3.2 |
| Hemiplegia | 0.4 | 0.2 | 0.3 | 2.5 | 24.5 | 0.5 | 0.4 | 0.3 | 0.3 | 0.1 | 2.4 | 0.3 | 1.7 | 2.8 | 2.4 | 3 | 7.5 | 0.8 | 0.2 | 1.2 | 0.3 | 0.6 | 0.3 | 1.3 | 0.5 |
| Moderate to severe renal disease | 3.3 | 1.4 | 3.3 | 2.5 | 1.4 | 5 | 2.2 | 0.5 | 1.7 | 0.9 | 2.4 | 1.5 | 2.3 | 1.3 | 1.3 | 4.1 | 2.2 | 4.1 | 1.2 | 6.7 | 1.6 | 3 | 1.4 | 5.6 | 1.4 |
| Diabetes with end−organ damage | 4.1 | 1.7 | 2.8 | 2.5 | 0.9 | 5.7 | 2.8 | 0.4 | 1.8 | 0.8 | 2.5 | 1.5 | 1.8 | 1.3 | 1.5 | 3.4 | 3.1 | 4.7 | 1.8 | 13.1 | 2.5 | 5.4 | 1.8 | 7.4 | 1.5 |
| Solid cancer tumour | 8.4 | 4.2 | 5.7 | 23.1 | 2.2 | 13.6 | 5.9 | 0.5 | 2.5 | 1.3 | 6.4 | 4.3 | 5.7 | 1.5 | 5.4 | 7.6 | 7.3 | 15.1 | 2.6 | 13.8 | 3.2 | 6.7 | 3.6 | 12.7 | 3.7 |
| Leukaemia | 0.2 | 0.1 | 0.2 | 0.6 | 0.1 | 0.3 | 0.2 | 0 | 0.1 | 0 | 0.2 | 0.2 | 0.4 | 0.1 | 0.1 | 0.3 | 0.5 | 0.4 | 0.1 | 0.7 | 0.1 | 0.3 | 0.2 | 0.4 | 0.1 |
| Lymphoma | 0.4 | 0.3 | 0.3 | 2 | 0.1 | 0.8 | 0.4 | 0.1 | 0.2 | 0.1 | 0.4 | 0.3 | 0.8 | 0.1 | 0.3 | 0.7 | 0.7 | 1 | 0.2 | 1.8 | 0.3 | 0.5 | 0.3 | 0.8 | 0.3 |
| Moderate to severe liver disease | 4.3 | 0.4 | 0.5 | 0.3 | 0.3 | 0.5 | 0.5 | 0.1 | 0.8 | 0.1 | 0.5 | 0.2 | 0.3 | 0.1 | 0.2 | 0.8 | 0.5 | 0.3 | 0.4 | 1.4 | 0.5 | 0.2 | 0.4 | 0.5 | 0.3 |
| Metastatic solid tumour | 0.8 | 0.6 | 0.5 | 3 | 0.2 | 0.7 | 0.6 | 0 | 0.2 | 0.1 | 0.7 | 0.3 | 0.5 | 0.2 | 0.5 | 0.8 | 0.7 | 0.9 | 0.2 | 1.5 | 0.3 | 0.6 | 0.4 | 1 | 0.3 |
| AIDS | 0.3 | 0.1 | 0.2 | 0.1 | 0.1 | 0.1 | 0.2 | 0.1 | 0.6 | 0.1 | 0.2 | 0.1 | 0.7 | 0.1 | 0 | 0.2 | 0 | 0.2 | 0.2 | 0.4 | 0.4 | 0.1 | 0.2 | 0.1 | 0.1 |

Percent: 0, 20, 40, 60

### Concurrent diseases

#### Sensitivity analysis 1 − Prevalent cohort 2019

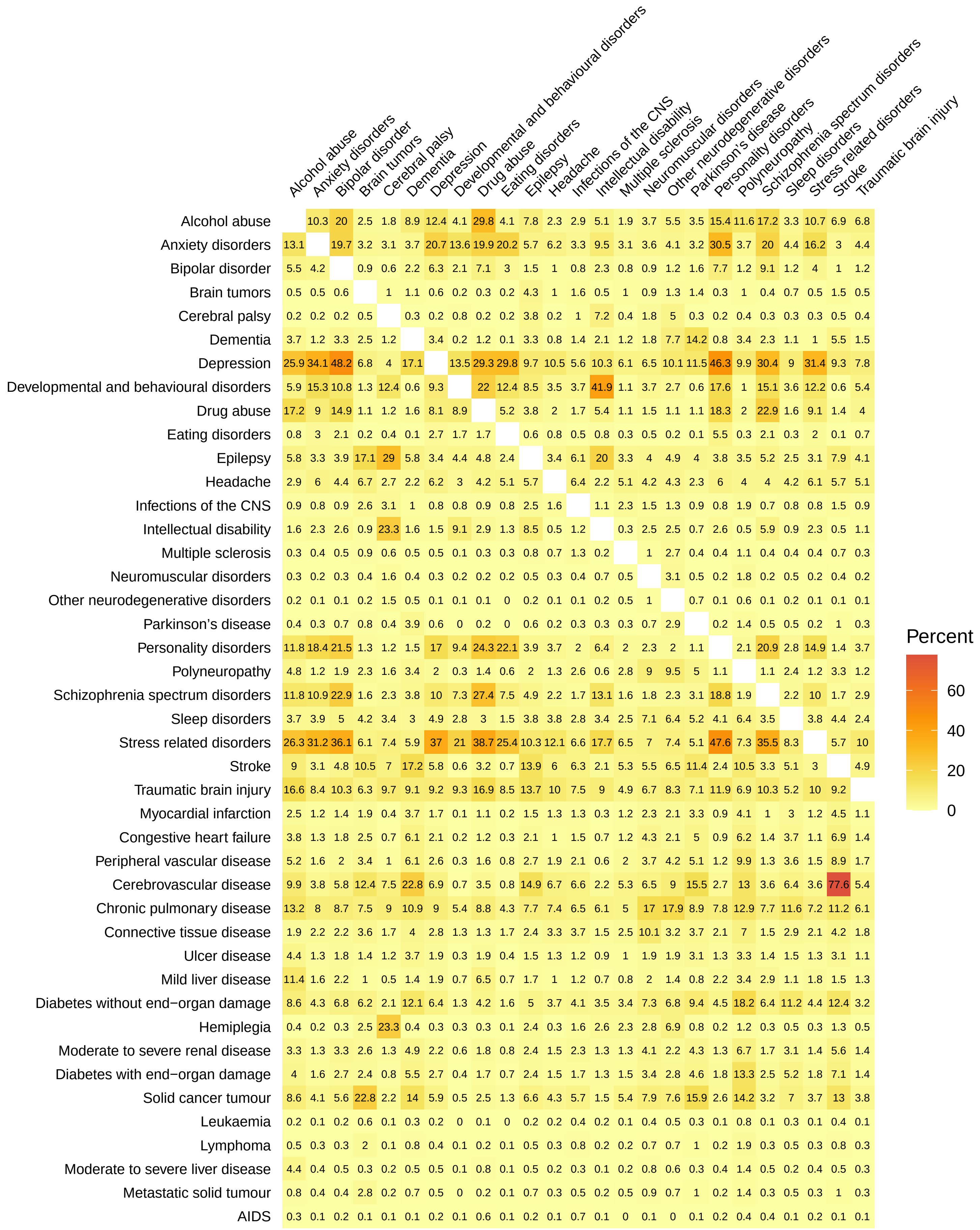


| | Alcohol abuse | Anxiety disorders | Bipolar disorder | Brain tumors | Cerebral palsy | Dementia | Depression | Developmental and behavioural disorders | Drug abuse | Eating disorders | Epilepsy | Headache | Infections of the CNS | Intellectual disability | Multiple sclerosis | Neuromuscular disorders | Other neurodegenerative disorders | Parkinson's disease | Personality disorders | Polyneuropathy | Schizophrenia spectrum disorders | Sleep disorders | Stress related disorders | Stroke | Traumatic brain injury |
|---|---|---|---|---|---|---|---|---|---|---|---|---|---|---|---|---|---|---|---|---|---|---|---|---|---|
| Alcohol abuse | | 10.3 | 20 | 2.5 | 1.8 | 8.9 | 12.4 | 4.1 | 29.8 | 4.1 | 7.8 | 2.3 | 2.9 | 5.1 | 1.9 | 3.7 | 5.5 | 3.5 | 15.4 | 11.6 | 17.2 | 3.3 | 10.7 | 6.9 | 6.8 |
| Anxiety disorders | 13.1 | | 19.7 | 3.2 | 3.1 | 3.7 | 20.7 | 13.6 | 19.9 | 20.2 | 5.7 | 6.2 | 3.3 | 9.5 | 3.1 | 3.6 | 4.1 | 3.2 | 30.5 | 3.7 | 20 | 4.4 | 16.2 | 3 | 4.4 |
| Bipolar disorder | 5.5 | 4.2 | | 0.9 | 0.6 | 2.2 | 6.3 | 2.1 | 7.1 | 3 | 1.5 | 1 | 0.8 | 2.3 | 0.8 | 0.9 | 1.2 | 1.6 | 7.7 | 1.2 | 9.1 | 1.2 | 4 | 1 | 1.2 |
| Brain tumors | 0.5 | 0.5 | 0.6 | | 1 | 1.1 | 0.6 | 0.2 | 0.3 | 0.2 | 4.3 | 1 | 1.6 | 0.5 | 1 | 0.9 | 1.3 | 1.4 | 0.3 | 1 | 0.4 | 0.7 | 0.5 | 1.5 | 0.5 |
| Cerebral palsy | 0.2 | 0.2 | 0.2 | 0.5 | | 0.3 | 0.2 | 0.8 | 0.2 | 0.2 | 3.8 | 0.2 | 1 | 7.2 | 0.4 | 1.8 | 5 | 0.3 | 0.2 | 0.4 | 0.3 | 0.3 | 0.3 | 0.5 | 0.4 |
| Dementia | 3.7 | 1.2 | 3.3 | 2.5 | 1.2 | | 3.4 | 0.2 | 1.2 | 0.1 | 3.3 | 0.8 | 1.4 | 2.1 | 1.2 | 1.8 | 7.7 | 14.2 | 0.8 | 3.4 | 2.3 | 1.1 | 1 | 5.5 | 1.5 |
| Depression | 25.9 | 34.1 | 48.2 | 6.8 | 4 | 17.1 | | 13.5 | 29.3 | 29.8 | 9.7 | 10.5 | 5.6 | 10.3 | 6.1 | 6.5 | 10.1 | 11.5 | 46.3 | 9.9 | 30.4 | 9 | 31.4 | 9.3 | 7.8 |
| Developmental and behavioural disorders | 5.9 | 15.3 | 10.8 | 1.3 | 12.4 | 0.6 | 9.3 | | 22 | 12.4 | 8.5 | 3.5 | 3.7 | 41.9 | 1.1 | 3.7 | 2.7 | 0.6 | 17.6 | 1 | 15.1 | 3.6 | 12.2 | 0.6 | 5.4 |
| Drug abuse | 17.2 | 9 | 14.9 | 1.1 | 1.2 | 1.6 | 8.1 | 8.9 | | 5.2 | 3.8 | 2 | 1.7 | 5.4 | 1.1 | 1.5 | 1.1 | 1.1 | 18.3 | 2 | 22.9 | 1.6 | 9.1 | 1.4 | 4 |
| Eating disorders | 0.8 | 3 | 2.1 | 0.2 | 0.4 | 0.1 | 2.7 | 1.7 | 1.7 | | 0.6 | 0.8 | 0.5 | 0.8 | 0.3 | 0.5 | 0.2 | 0.1 | 5.5 | 0.3 | 2.1 | 0.3 | 2 | 0.1 | 0.7 |
| Epilepsy | 5.8 | 3.3 | 3.9 | 17.1 | 29 | 5.8 | 3.4 | 4.4 | 4.8 | 2.4 | | 3.4 | 6.1 | 20 | 3.3 | 4 | 4.9 | 4 | 3.8 | 3.5 | 5.2 | 2.5 | 3.1 | 7.9 | 4.1 |
| Headache | 2.9 | 6 | 4.4 | 6.7 | 2.7 | 2.2 | 6.2 | 3 | 4.2 | 5.1 | 5.7 | | 6.4 | 2.2 | 5.1 | 4.2 | 4.3 | 2.3 | 6 | 4 | 4 | 4.2 | 6.1 | 5.7 | 5.1 |
| Infections of the CNS | 0.9 | 0.8 | 0.9 | 2.6 | 3.1 | 1 | 0.8 | 0.8 | 0.9 | 0.8 | 2.5 | 1.6 | | 1.1 | 2.3 | 1.5 | 1.3 | 0.9 | 0.8 | 1.9 | 0.7 | 0.8 | 0.8 | 1.5 | 0.9 |
| Intellectual disability | 1.6 | 2.3 | 2.6 | 0.9 | 23.3 | 1.6 | 1.5 | 9.1 | 2.9 | 1.3 | 8.5 | 0.5 | 1.2 | | 0.3 | 2.5 | 2.5 | 0.7 | 2.6 | 0.5 | 5.9 | 0.9 | 2.3 | 0.5 | 1.1 |
| Multiple sclerosis | 0.3 | 0.4 | 0.5 | 0.9 | 0.6 | 0.5 | 0.5 | 0.1 | 0.3 | 0.3 | 0.8 | 0.7 | 1.3 | 0.2 | | 1 | 2.7 | 0.4 | 0.4 | 1.1 | 0.4 | 0.4 | 0.4 | 0.7 | 0.3 |
| Neuromuscular disorders | 0.3 | 0.2 | 0.3 | 0.4 | 1.6 | 0.4 | 0.3 | 0.2 | 0.2 | 0.2 | 0.5 | 0.3 | 0.4 | 0.7 | 0.5 | | 3.1 | 0.5 | 0.2 | 1.8 | 0.2 | 0.5 | 0.2 | 0.4 | 0.2 |
| Other neurodegenerative disorders | 0.2 | 0.1 | 0.1 | 0.2 | 1.5 | 0.5 | 0.1 | 0.1 | 0.1 | 0 | 0.2 | 0.1 | 0.1 | 0.2 | 0.5 | 1 | | 0.7 | 0.1 | 0.6 | 0.1 | 0.2 | 0.1 | 0.1 | 0.1 |
| Parkinson's disease | 0.4 | 0.3 | 0.7 | 0.8 | 0.4 | 3.9 | 0.6 | 0 | 0.2 | 0 | 0.6 | 0.2 | 0.3 | 0.3 | 0.3 | 0.7 | 2.9 | | 0.2 | 1.4 | 0.5 | 0.5 | 0.2 | 1 | 0.3 |
| Personality disorders | 11.8 | 18.4 | 21.5 | 1.3 | 1.2 | 1.5 | 17 | 9.4 | 24.3 | 22.1 | 3.9 | 3.7 | 2 | 6.4 | 2 | 2.3 | 2 | 1.1 | | 2.1 | 20.9 | 2.8 | 14.9 | 1.4 | 3.7 |
| Polyneuropathy | 4.8 | 1.2 | 1.9 | 2.3 | 1.6 | 3.4 | 2 | 0.3 | 1.4 | 0.6 | 2 | 1.3 | 2.6 | 0.6 | 2.8 | 9 | 9.5 | 5 | 1.1 | | 1.1 | 2.4 | 1.2 | 3.3 | 1.2 |
| Schizophrenia spectrum disorders | 11.8 | 10.9 | 22.9 | 1.6 | 2.3 | 3.8 | 10 | 7.3 | 27.4 | 7.5 | 4.9 | 2.2 | 1.7 | 13.1 | 1.6 | 1.8 | 2.3 | 3.1 | 18.8 | 1.9 | | 2.2 | 10 | 1.7 | 2.9 |
| Sleep disorders | 3.7 | 3.9 | 5 | 4.2 | 3.4 | 3 | 4.9 | 2.8 | 3 | 1.5 | 3.8 | 3.8 | 2.8 | 3.4 | 2.5 | 7.1 | 6.4 | 5.2 | 4.1 | 6.4 | 3.5 | | 3.8 | 4.4 | 2.4 |
| Stress related disorders | 26.3 | 31.2 | 36.1 | 6.1 | 7.4 | 5.9 | 37 | 21 | 38.7 | 25.4 | 10.3 | 12.1 | 6.6 | 17.7 | 6.5 | 7 | 7.4 | 5.1 | 47.6 | 7.3 | 35.5 | 8.3 | | 5.7 | 10 |
| Stroke | 9 | 3.1 | 4.8 | 10.5 | 7 | 17.2 | 5.8 | 0.6 | 3.2 | 0.7 | 13.9 | 6 | 6.3 | 2.1 | 5.3 | 5.5 | 6.5 | 11.4 | 2.4 | 10.5 | 3.3 | 5.1 | 3 | | 4.9 |
| Traumatic brain injury | 16.6 | 8.4 | 10.3 | 6.3 | 9.7 | 9.1 | 9.2 | 9.3 | 16.9 | 8.5 | 13.7 | 10 | 7.5 | 9 | 4.9 | 6.7 | 8.3 | 7.1 | 11.9 | 6.9 | 10.3 | 5.2 | 10 | 9.2 | |
| Myocardial infarction | 2.5 | 1.2 | 1.4 | 1.9 | 0.4 | 3.7 | 1.7 | 0.1 | 1.1 | 0.2 | 1.5 | 1.3 | 1.3 | 0.3 | 1.2 | 2.3 | 2.1 | 3.3 | 0.9 | 4.1 | 1 | 3 | 1.2 | 4.5 | 1.1 |
| Congestive heart failure | 3.8 | 1.3 | 1.8 | 2.5 | 0.7 | 6.1 | 2.1 | 0.2 | 1.2 | 0.3 | 2.1 | 1 | 1.5 | 0.7 | 1.2 | 4.3 | 2.1 | 5 | 0.9 | 6.2 | 1.4 | 3.7 | 1.1 | 6.9 | 1.4 |
| Peripheral vascular disease | 5.2 | 1.6 | 2 | 3.4 | 1 | 6.1 | 2.6 | 0.3 | 1.6 | 0.8 | 2.7 | 1.9 | 2.1 | 0.6 | 2 | 3.7 | 4.2 | 5.1 | 1.2 | 9.9 | 1.3 | 3.6 | 1.5 | 8.9 | 1.7 |
| Cerebrovascular disease | 9.9 | 3.8 | 5.8 | 12.4 | 7.5 | 22.8 | 6.9 | 0.7 | 3.5 | 0.8 | 14.9 | 6.7 | 6.6 | 2.2 | 5.3 | 6.5 | 9 | 15.5 | 2.7 | 13 | 3.6 | 6.4 | 3.6 | 77.6 | 5.4 |
| Chronic pulmonary disease | 13.2 | 8 | 8.7 | 7.5 | 9 | 10.9 | 9 | 5.4 | 8.8 | 4.3 | 7.7 | 7.4 | 6.5 | 6.1 | 5 | 17 | 17.9 | 8.9 | 7.8 | 12.9 | 7.7 | 11.6 | 7.2 | 11.2 | 6.1 |
| Connective tissue disease | 1.9 | 2.2 | 2.2 | 3.6 | 1.7 | 4 | 2.8 | 1.3 | 1.3 | 1.7 | 2.4 | 3.3 | 3.7 | 1.5 | 2.5 | 10.1 | 3.2 | 3.7 | 2.1 | 7 | 1.5 | 2.9 | 2.1 | 4.2 | 1.8 |
| Ulcer disease | 4.4 | 1.3 | 1.8 | 1.4 | 1.2 | 3.7 | 1.9 | 0.3 | 1.9 | 0.4 | 1.5 | 1.3 | 1.2 | 0.9 | 1 | 1.9 | 1.9 | 3.1 | 1.3 | 3.3 | 1.4 | 1.5 | 1.3 | 3.1 | 1.1 |
| Mild liver disease | 11.4 | 1.6 | 2.2 | 1 | 0.5 | 1.4 | 1.9 | 0.7 | 6.5 | 0.7 | 1.7 | 1 | 1.2 | 0.7 | 0.8 | 2 | 1.4 | 0.8 | 2.2 | 3.4 | 2.9 | 1.1 | 1.8 | 1.5 | 1.3 |
| Diabetes without end−organ damage | 8.6 | 4.3 | 6.8 | 6.2 | 2.1 | 12.1 | 6.4 | 1.3 | 4.2 | 1.6 | 5 | 3.7 | 4.1 | 3.5 | 3.4 | 7.3 | 6.8 | 9.4 | 4.5 | 18.2 | 6.4 | 11.2 | 4.4 | 12.4 | 3.2 |
| Hemiplegia | 0.4 | 0.2 | 0.3 | 2.5 | 23.3 | 0.4 | 0.3 | 0.3 | 0.3 | 0.1 | 2.4 | 0.3 | 1.6 | 2.6 | 2.3 | 2.8 | 6.9 | 0.8 | 0.2 | 1.2 | 0.3 | 0.5 | 0.3 | 1.3 | 0.5 |
| Moderate to severe renal disease | 3.3 | 1.3 | 3.3 | 2.6 | 1.3 | 4.9 | 2.2 | 0.6 | 1.8 | 0.8 | 2.4 | 1.5 | 2.3 | 1.3 | 1.3 | 4.1 | 2.2 | 4.3 | 1.3 | 6.7 | 1.7 | 3.1 | 1.4 | 5.6 | 1.4 |
| Diabetes with end−organ damage | 4 | 1.6 | 2.7 | 2.4 | 0.8 | 5.5 | 2.7 | 0.4 | 1.7 | 0.7 | 2.4 | 1.5 | 1.7 | 1.3 | 1.5 | 3.4 | 2.8 | 4.6 | 1.8 | 13.3 | 2.5 | 5.2 | 1.8 | 7.1 | 1.4 |
| Solid cancer tumour | 8.6 | 4.1 | 5.6 | 22.8 | 2.2 | 14 | 5.9 | 0.5 | 2.5 | 1.3 | 6.6 | 4.3 | 5.7 | 1.5 | 5.4 | 7.9 | 7.6 | 15.9 | 2.6 | 14.2 | 3.2 | 7 | 3.7 | 13 | 3.8 |
| Leukaemia | 0.2 | 0.1 | 0.2 | 0.6 | 0.1 | 0.3 | 0.2 | 0 | 0.1 | 0 | 0.2 | 0.2 | 0.4 | 0.2 | 0.1 | 0.4 | 0.5 | 0.3 | 0.1 | 0.8 | 0.1 | 0.3 | 0.1 | 0.4 | 0.1 |
| Lymphoma | 0.5 | 0.3 | 0.3 | 2 | 0.1 | 0.8 | 0.4 | 0.1 | 0.2 | 0.1 | 0.5 | 0.3 | 0.8 | 0.2 | 0.2 | 0.7 | 0.7 | 1 | 0.2 | 1.9 | 0.3 | 0.5 | 0.3 | 0.8 | 0.3 |
| Moderate to severe liver disease | 4.4 | 0.4 | 0.5 | 0.3 | 0.2 | 0.5 | 0.5 | 0.1 | 0.8 | 0.1 | 0.5 | 0.2 | 0.3 | 0.1 | 0.2 | 0.8 | 0.6 | 0.3 | 0.4 | 1.4 | 0.5 | 0.2 | 0.4 | 0.5 | 0.3 |
| Metastatic solid tumour | 0.8 | 0.4 | 0.4 | 2.8 | 0.2 | 0.7 | 0.5 | 0 | 0.2 | 0.1 | 0.7 | 0.3 | 0.5 | 0.2 | 0.5 | 0.9 | 0.7 | 1 | 0.2 | 1.4 | 0.3 | 0.5 | 0.3 | 1 | 0.3 |
| AIDS | 0.3 | 0.1 | 0.2 | 0.1 | 0.1 | 0.1 | 0.2 | 0.1 | 0.6 | 0.1 | 0.2 | 0.1 | 0.7 | 0.1 | 0 | 0.1 | 0 | 0.1 | 0.2 | 0.4 | 0.4 | 0.1 | 0.2 | 0.1 | 0.1 |

#### Concurrent diseases

##### Sensitivity analysis 1 − Prevalent cohort 2020

| | Alcohol abuse | Anxiety disorders | Bipolar disorder | Brain tumors | Cerebral palsy | Dementia | Depression | Developmental and behavioural disorders | Drug abuse | Eating disorders | Epilepsy | Headache | Infections of the CNS | Intellectual disability | Multiple sclerosis | Neuromuscular disorders | Other neurodegenerative disorders | Parkinson's disease | Personality disorders | Polyneuropathy | Schizophrenia spectrum disorders | Sleep disorders | Stress related disorders | Stroke | Traumatic brain injury |
|---|---|---|---|---|---|---|---|---|---|---|---|---|---|---|---|---|---|---|---|---|---|---|---|---|---|
| Alcohol abuse | | 9.9 | 19.5 | 2.4 | 1.7 | 8.6 | 11.9 | 4.1 | 29.1 | 4 | 7.5 | 2.3 | 2.8 | 5.1 | 1.9 | 3.6 | 4.6 | 3.3 | 14.9 | 10.8 | 16.6 | 3.2 | 10.3 | 6.6 | 6.6 |
| Anxiety disorders | 13.3 | | 19.9 | 3.1 | 3.2 | 3.6 | 21.1 | 14.3 | 20.1 | 20.5 | 5.8 | 6.4 | 3.2 | 9.9 | 3.1 | 3.7 | 3.8 | 3.3 | 31.1 | 3.7 | 20.6 | 4.5 | 16.5 | 3 | 4.5 |
| Bipolar disorder | 5.6 | 4.2 | | 0.9 | 0.7 | 2.2 | 6.3 | 2.2 | 7.1 | 3.1 | 1.4 | 1 | 0.9 | 2.2 | 0.8 | 0.9 | 1.1 | 1.6 | 7.7 | 1.2 | 8.9 | 1.2 | 4 | 1 | 1.2 |
| Brain tumors | 0.5 | 0.5 | 0.6 | | 1 | 1.2 | 0.6 | 0.2 | 0.3 | 0.2 | 4.3 | 1 | 1.6 | 0.5 | 1.1 | 0.9 | 1.2 | 1.4 | 0.3 | 1.1 | 0.4 | 0.7 | 0.5 | 1.5 | 0.5 |
| Cerebral palsy | 0.2 | 0.2 | 0.2 | 0.5 | | 0.3 | 0.2 | 0.8 | 0.2 | 0.2 | 3.8 | 0.2 | 1 | 6.9 | 0.3 | 1.8 | 5.1 | 0.3 | 0.1 | 0.4 | 0.3 | 0.3 | 0.3 | 0.5 | 0.4 |
| Dementia | 3.6 | 1.1 | 3.1 | 2.5 | 1.1 | | 3.1 | 0.1 | 1.1 | 0.1 | 3.2 | 0.8 | 1.4 | 2 | 1.1 | 1.6 | 7 | 13.7 | 0.8 | 3.3 | 2.1 | 1.1 | 0.9 | 5.2 | 1.5 |
| Depression | 25.8 | 34 | 47.4 | 6.6 | 4 | 16.1 | | 14 | 29 | 29.8 | 9.6 | 10.6 | 5.6 | 10.5 | 6 | 6.4 | 9.5 | 10.9 | 46.5 | 9.6 | 30.4 | 9.2 | 31.3 | 9 | 7.8 |
| Developmental and behavioural disorders | 6.4 | 16.5 | 11.6 | 1.4 | 12.7 | 0.6 | 10.1 | | 23.2 | 13.2 | 8.8 | 3.7 | 3.8 | 42.9 | 1.2 | 3.9 | 2.7 | 0.6 | 18.8 | 1.1 | 16.3 | 3.9 | 13.2 | 0.7 | 5.7 |
| Drug abuse | 17.6 | 9 | 15 | 1 | 1.3 | 1.6 | 8.1 | 9 | | 5.2 | 3.8 | 2 | 1.6 | 5.7 | 1.1 | 1.5 | 1 | 1.1 | 18.4 | 1.9 | 23.1 | 1.6 | 9.1 | 1.4 | 4 |
| Eating disorders | 0.8 | 3.1 | 2.2 | 0.2 | 0.4 | 0.1 | 2.8 | 1.7 | 1.7 | | 0.6 | 0.8 | 0.5 | 0.8 | 0.3 | 0.5 | 0.2 | 0 | 5.5 | 0.3 | 2.1 | 0.3 | 2 | 0.1 | 0.7 |
| Epilepsy | 5.6 | 3.2 | 3.8 | 16.4 | 28.7 | 5.7 | 3.3 | 4.2 | 4.7 | 2.3 | | 3.3 | 5.9 | 19.6 | 3.2 | 4 | 4.9 | 3.8 | 3.6 | 3.4 | 5.1 | 2.5 | 3 | 7.7 | 4.1 |
| Headache | 2.9 | 6.2 | 4.6 | 6.8 | 2.8 | 2.4 | 6.4 | 3.1 | 4.3 | 5.3 | 5.8 | | 6.6 | 2.2 | 5.1 | 4.4 | 4.5 | 2.2 | 6.2 | 4.1 | 4.1 | 4.3 | 6.2 | 5.7 | 5.2 |
| Infections of the CNS | 0.9 | 0.8 | 0.9 | 2.6 | 3.1 | 1.1 | 0.8 | 0.8 | 0.8 | 0.8 | 2.5 | 1.6 | | 1.1 | 2.4 | 1.5 | 1.3 | 0.8 | 0.8 | 1.9 | 0.7 | 0.8 | 0.8 | 1.5 | 0.9 |
| Intellectual disability | 1.6 | 2.4 | 2.5 | 0.8 | 22.8 | 1.6 | 1.6 | 8.9 | 3.1 | 1.3 | 8.5 | 0.6 | 1.1 | | 0.3 | 2.4 | 2.4 | 0.7 | 2.6 | 0.5 | 5.9 | 1 | 2.3 | 0.5 | 1.2 |
| Multiple sclerosis | 0.3 | 0.4 | 0.5 | 1 | 0.6 | 0.5 | 0.5 | 0.1 | 0.3 | 0.3 | 0.8 | 0.7 | 1.3 | 0.2 | | 1 | 2.4 | 0.5 | 0.4 | 1.1 | 0.4 | 0.4 | 0.4 | 0.7 | 0.3 |
| Neuromuscular disorders | 0.3 | 0.2 | 0.3 | 0.4 | 1.6 | 0.3 | 0.3 | 0.2 | 0.2 | 0.2 | 0.5 | 0.3 | 0.4 | 0.6 | 0.5 | | 2.9 | 0.5 | 0.2 | 1.8 | 0.2 | 0.5 | 0.2 | 0.4 | 0.2 |
| Other neurodegenerative disorders | 0.1 | 0.1 | 0.1 | 0.2 | 1.5 | 0.5 | 0.1 | 0.1 | 0 | 0 | 0.2 | 0.1 | 0.1 | 0.2 | 0.4 | 1 | | 0.8 | 0.1 | 0.6 | 0.1 | 0.2 | 0.1 | 0.1 | 0.1 |
| Parkinson's disease | 0.4 | 0.3 | 0.7 | 0.9 | 0.4 | 3.9 | 0.6 | 0 | 0.2 | 0 | 0.6 | 0.2 | 0.3 | 0.2 | 0.3 | 0.7 | 3.2 | | 0.2 | 1.4 | 0.5 | 0.5 | 0.2 | 1 | 0.3 |
| Personality disorders | 11.7 | 18.2 | 21.2 | 1.3 | 1.2 | 1.4 | 17 | 9.5 | 24 | 21.7 | 3.8 | 3.7 | 1.9 | 6.4 | 2 | 2.1 | 1.9 | 1.1 | | 2 | 20.4 | 2.8 | 14.6 | 1.4 | 3.7 |
| Polyneuropathy | 4.8 | 1.2 | 1.8 | 2.4 | 1.7 | 3.5 | 2 | 0.3 | 1.4 | 0.6 | 2 | 1.4 | 2.6 | 0.6 | 2.7 | 9.1 | 9.3 | 5.2 | 1.1 | | 1.1 | 2.5 | 1.3 | 3.5 | 1.2 |
| Schizophrenia spectrum disorders | 12 | 11 | 22.2 | 1.5 | 2.3 | 3.6 | 10.1 | 7.6 | 27.5 | 7.6 | 4.9 | 2.2 | 1.7 | 13.3 | 1.6 | 1.8 | 2.1 | 3 | 18.6 | 1.8 | | 2.2 | 10 | 1.7 | 2.9 |
| Sleep disorders | 4 | 4.1 | 5.4 | 4.4 | 3.8 | 3.4 | 5.2 | 3.1 | 3.3 | 1.6 | 4.1 | 4.1 | 3 | 3.7 | 2.8 | 7.5 | 7 | 5.5 | 4.4 | 7 | 3.8 | | 4.1 | 4.9 | 2.6 |
| Stress related disorders | 26.7 | 31.8 | 36.4 | 6.3 | 7.7 | 5.9 | 37.6 | 22.1 | 39.2 | 25.9 | 10.5 | 12.3 | 6.6 | 18.3 | 6.6 | 7.2 | 7 | 5.2 | 48.2 | 7.4 | 36.3 | 8.6 | | 5.8 | 10.2 |
| Stroke | 8.9 | 3 | 4.7 | 10.3 | 7.1 | 16.9 | 5.6 | 0.6 | 3.1 | 0.7 | 13.8 | 5.8 | 6.2 | 2 | 5.2 | 5.7 | 6.1 | 11.2 | 2.3 | 10.5 | 3.2 | 5.3 | 3 | | 5 |
| Traumatic brain injury | 16.3 | 8.3 | 10.1 | 6.4 | 9.5 | 9.1 | 9 | 9.1 | 16.5 | 8.4 | 13.5 | 9.9 | 7.4 | 8.9 | 4.9 | 6.6 | 7.9 | 7.4 | 11.6 | 6.9 | 10 | 5.2 | 9.8 | 9.2 | |
| Myocardial infarction | 2.5 | 1.2 | 1.4 | 1.9 | 0.5 | 3.5 | 1.7 | 0.2 | 1.1 | 0.2 | 1.5 | 1.2 | 1.2 | 0.3 | 1.2 | 2.3 | 2.1 | 3.4 | 0.9 | 4.1 | 1 | 2.9 | 1.2 | 4.3 | 1.1 |
| Congestive heart failure | 3.7 | 1.3 | 1.7 | 2.5 | 0.7 | 5.8 | 2.1 | 0.2 | 1.2 | 0.2 | 2.1 | 1 | 1.5 | 0.7 | 1.1 | 4.5 | 2.1 | 5 | 0.8 | 6.4 | 1.3 | 3.7 | 1.1 | 6.8 | 1.4 |
| Peripheral vascular disease | 5.2 | 1.5 | 2 | 3.2 | 1 | 5.9 | 2.5 | 0.3 | 1.6 | 0.8 | 2.7 | 1.9 | 2.2 | 0.6 | 1.9 | 3.5 | 3.8 | 5.1 | 1.1 | 10.1 | 1.3 | 3.6 | 1.5 | 8.7 | 1.7 |
| Cerebrovascular disease | 9.7 | 3.6 | 5.6 | 12 | 7.5 | 22.3 | 6.5 | 0.7 | 3.4 | 0.8 | 14.7 | 6.5 | 6.4 | 2.2 | 5.1 | 6.5 | 8.3 | 15.2 | 2.6 | 13 | 3.5 | 6.6 | 3.4 | 77.2 | 5.4 |
| Chronic pulmonary disease | 13.1 | 7.8 | 8.6 | 7.5 | 9 | 10.7 | 8.8 | 5.2 | 8.6 | 4.2 | 7.7 | 7.4 | 6.4 | 6.1 | 5 | 17 | 17.7 | 8.6 | 7.6 | 12.9 | 7.7 | 11.7 | 7.1 | 11.1 | 6.1 |
| Connective tissue disease | 1.9 | 2.2 | 2.3 | 3.7 | 1.7 | 4.1 | 2.8 | 1.2 | 1.3 | 1.8 | 2.4 | 3.3 | 3.7 | 1.5 | 2.4 | 9.9 | 3.3 | 3.8 | 2 | 7.1 | 1.4 | 3 | 2.2 | 4.3 | 1.8 |
| Ulcer disease | 4.3 | 1.2 | 1.6 | 1.3 | 1.1 | 3.4 | 1.8 | 0.3 | 1.8 | 0.5 | 1.5 | 1.2 | 1.1 | 0.8 | 1 | 1.8 | 2.1 | 2.9 | 1.2 | 3.1 | 1.3 | 1.4 | 1.2 | 2.9 | 1 |
| Mild liver disease | 11.3 | 1.6 | 2.2 | 1.1 | 0.6 | 1.4 | 1.9 | 0.7 | 6.2 | 0.7 | 1.7 | 1 | 1.2 | 0.7 | 0.9 | 2 | 1.3 | 0.9 | 2.1 | 3.3 | 2.8 | 1.2 | 1.7 | 1.5 | 1.3 |
| Diabetes without end−organ damage | 8.4 | 4.2 | 6.6 | 6 | 2.2 | 11.9 | 6.2 | 1.3 | 4 | 1.6 | 4.9 | 3.5 | 3.9 | 3.4 | 3.4 | 7.2 | 6.3 | 9.1 | 4.4 | 19.9 | 6.2 | 11 | 4.3 | 12 | 3.2 |
| Hemiplegia | 0.4 | 0.2 | 0.2 | 2.4 | 22.5 | 0.4 | 0.3 | 0.3 | 0.3 | 0.1 | 2.4 | 0.3 | 1.6 | 2.5 | 2.3 | 2.7 | 6.7 | 0.8 | 0.2 | 1.1 | 0.3 | 0.5 | 0.3 | 1.3 | 0.5 |
| Moderate to severe renal disease | 3.4 | 1.3 | 3.3 | 2.6 | 1.4 | 5 | 2.2 | 0.6 | 1.8 | 0.8 | 2.5 | 1.5 | 2.3 | 1.3 | 1.4 | 4 | 2.5 | 4.5 | 1.2 | 7.2 | 1.6 | 3.2 | 1.4 | 5.7 | 1.4 |
| Diabetes with end−organ damage | 3.9 | 1.6 | 2.6 | 2.4 | 0.9 | 5.3 | 2.7 | 0.4 | 1.7 | 0.8 | 2.4 | 1.5 | 1.8 | 1.3 | 1.5 | 3.4 | 2.8 | 4.5 | 1.8 | 15.8 | 2.4 | 5.3 | 1.8 | 6.9 | 1.5 |
| Solid cancer tumour | 8.7 | 3.9 | 5.6 | 22.7 | 2.3 | 13.9 | 5.8 | 0.5 | 2.5 | 1.3 | 6.7 | 4.4 | 5.9 | 1.5 | 5.5 | 8.3 | 8 | 16 | 2.6 | 14.4 | 3.1 | 7.2 | 3.7 | 13.2 | 3.9 |
| Leukaemia | 0.2 | 0.1 | 0.2 | 0.5 | 0.1 | 0.4 | 0.2 | 0 | 0.1 | 0 | 0.2 | 0.2 | 0.4 | 0.2 | 0.1 | 0.4 | 0.4 | 0.4 | 0.1 | 0.8 | 0.1 | 0.3 | 0.1 | 0.4 | 0.1 |
| Lymphoma | 0.5 | 0.3 | 0.3 | 2.1 | 0.1 | 0.8 | 0.4 | 0.1 | 0.2 | 0.1 | 0.5 | 0.3 | 0.8 | 0.1 | 0.3 | 0.7 | 1 | 1 | 0.2 | 1.9 | 0.3 | 0.5 | 0.3 | 0.9 | 0.3 |
| Moderate to severe liver disease | 4.4 | 0.4 | 0.5 | 0.3 | 0.2 | 0.5 | 0.4 | 0.1 | 0.8 | 0.1 | 0.5 | 0.2 | 0.3 | 0.1 | 0.2 | 0.8 | 0.6 | 0.3 | 0.3 | 1.3 | 0.4 | 0.2 | 0.4 | 0.5 | 0.3 |
| Metastatic solid tumour | 0.8 | 0.4 | 0.4 | 2.5 | 0.2 | 0.6 | 0.5 | 0 | 0.2 | 0.1 | 0.7 | 0.3 | 0.4 | 0.1 | 0.4 | 0.8 | 0.7 | 0.8 | 0.2 | 1.4 | 0.2 | 0.5 | 0.3 | 0.9 | 0.3 |
| AIDS | 0.3 | 0.1 | 0.2 | 0.1 | 0.1 | 0.1 | 0.2 | 0.1 | 0.5 | 0.1 | 0.2 | 0.1 | 0.7 | 0.1 | 0 | 0.2 | 0 | 0.1 | 0.2 | 0.4 | 0.3 | 0.1 | 0.2 | 0.2 | 0.1 |

Percent
60
40
20
0

### Concurrent diseases

#### Sensitivity analysis 1 − Prevalent cohort 2021

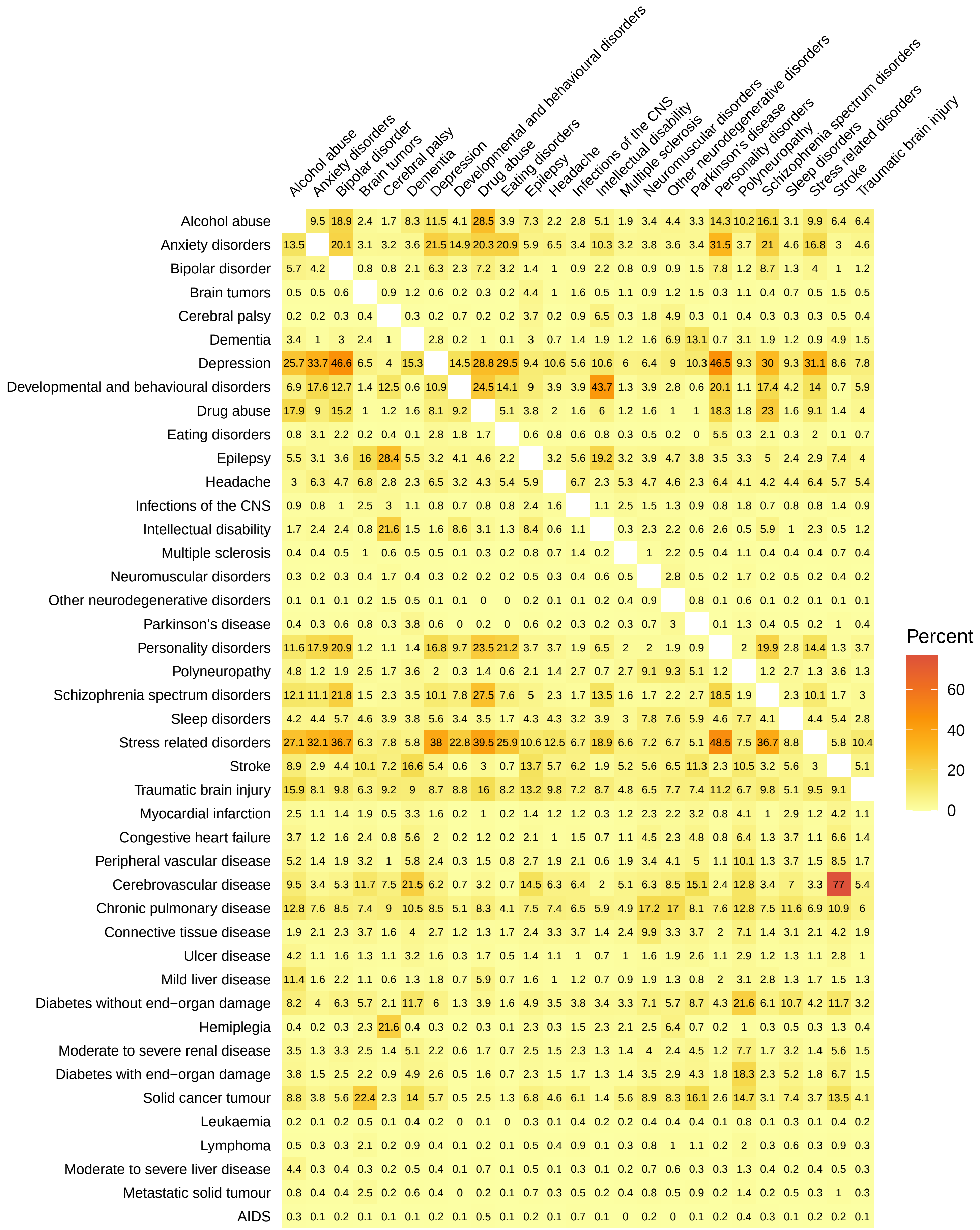

***Sensitivity analysis 1 / Incident cohort 2011-2015***

| | *Incidence, N* | *Incidence rate per 100,000 person-years (95% CI)* |
|---|---|---|
| Any brain disorder | 399,064 | 1,408 (1,404-1,412) |
| Alcohol abuse | 37,402 | 132 (131-133) |
| Anxiety disorders | 54,013 | 191 (189-192) |
| Bipolar disorder | 9,977 | 35 (35-36) |
| Brain tumors | 9,739 | 34 (34-35) |
| Cerebral palsy | 2,333 | 8 (8-9) |
| Dementia | 43,711 | 154 (153-156) |
| Depression | 79,888 | 282 (280-284) |
| Developmental and behavioural disorders | 49,492 | 175 (173-176) |
| Drug abuse | 19,014 | 67 (66-68) |
| Eating disorders | 6,105 | 22 (21-22) |
| Epilepsy | 22,978 | 81 (80-82) |
| Headache | 38,756 | 137 (135-138) |
| Infections of the CNS | 9,641 | 34 (33-35) |
| Intellectual disability | 8,467 | 30 (29-31) |
| Multiple sclerosis | 4,124 | 15 (14-15) |
| Neuromuscular disorders | 3,373 | 12 (12-12) |
| Other neurodegenerative disorders | 1,662 | 6 (6-6) |
| Parkinson’s disease | 7,714 | 27 (27-28) |
| Personality disorders | 21,886 | 77 (76-78) |
| Polyneuropathy | 19,514 | 69 (68-70) |
| Schizophrenia spectrum disorders | 19,387 | 68 (67-69) |
| Sleep disorders | 42,020 | 148 (147-150) |
| Stress related disorders | 82,275 | 290 (288-292) |
| Stroke | 67,899 | 240 (238-241) |
| Traumatic brain injury | 65,287 | 230 (229-232) |

***Sensitivity analysis 1 / Incident cohort 2016-2021***

| | *Incidence, N* | *Incidence rate per 100,000 person-years (95% CI)* |
|---|---|---|
| Any brain disorder | 465,256 | 1,331 (1,327-1,335) |
| Alcohol abuse | 32,103 | 92 (91-93) |
| Anxiety disorders | 59,389 | 170 (169-171) |
| Bipolar disorder | 9,811 | 28 (28-29) |
| Brain tumors | 13,152 | 38 (37-38) |
| Cerebral palsy | 2,143 | 6 (6-6) |
| Dementia | 47,870 | 137 (136-138) |
| Depression | 73,822 | 211 (210-213) |
| Developmental and behavioural disorders | 66,975 | 192 (190-193) |
| Drug abuse | 20,930 | 60 (59-61) |
| Eating disorders | 6,916 | 20 (19-20) |
| Epilepsy | 22,387 | 64 (63-65) |
| Headache | 50,444 | 144 (143-146) |
| Infections of the CNS | 13,556 | 39 (38-39) |
| Intellectual disability | 7,689 | 22 (22-22) |
| Multiple sclerosis | 4,745 | 14 (13-14) |
| Neuromuscular disorders | 3,324 | 10 (9-10) |
| Other neurodegenerative disorders | 2,202 | 6 (6-7) |
| Parkinson’s disease | 9,095 | 26 (25-27) |
| Personality disorders | 19,657 | 56 (55-57) |
| Polyneuropathy | 24,544 | 70 (69-71) |
| Schizophrenia spectrum disorders | 23,479 | 67 (66-68) |
| Sleep disorders | 61,506 | 176 (175-177) |
| Stress related disorders | 95,899 | 274 (273-276) |
| Stroke | 84,569 | 242 (240-244) |
| Traumatic brain injury | 78,140 | 224 (222-225) |

***Sensitivity analysis 1 / Prevalent cohort 2015***

| | *Prevalence, N (%)* | *Prevalence per 100,000 persons (95% CI)* |
|---|---|---|
| Any brain disorder | 1,096,530 (19.2) | 19,219 (19,183-19,255) |
| Alcohol abuse | 101,318 (1.8) | 1,776 (1,765-1,787) |
| Anxiety disorders | 97,734 (1.7) | 1,713 (1,702-1,724) |
| Bipolar disorder | 23,556 (0.4) | 413 (408-418) |
| Brain tumors | 15,212 (0.3) | 267 (262-271) |
| Cerebral palsy | 9,308 (0.2) | 163 (160-166) |
| Dementia | 40,536 (0.7) | 710 (704-717) |
| Depression | 182,143 (3.2) | 3,192 (3,178-3,207) |
| Developmental and behavioural disorders | 104,848 (1.8) | 1,838 (1,827-1,849) |
| Drug abuse | 50,264 (0.9) | 881 (873-889) |
| Eating disorders | 16,081 (0.3) | 282 (278-286) |
| Epilepsy | 73,582 (1.3) | 1,290 (1,280-1,299) |
| Headache | 99,688 (1.7) | 1,747 (1,736-1,758) |
| Infections of the CNS | 27,043 (0.5) | 474 (468-480) |
| Intellectual disability | 27,868 (0.5) | 488 (483-494) |
| Multiple sclerosis | 15,023 (0.3) | 263 (259-268) |
| Neuromuscular disorders | 7,391 (0.1) | 130 (127-133) |
| Other neurodegenerative disorders | 2,321 (0.0) | 41 (39-42) |
| Parkinson’s disease | 9,974 (0.2) | 175 (171-178) |
| Personality disorders | 71,766 (1.3) | 1,258 (1,249-1,267) |
| Polyneuropathy | 34,187 (0.6) | 599 (593-606) |
| Schizophrenia spectrum disorders | 61,462 (1.1) | 1,077 (1,069-1,086) |
| Sleep disorders | 75,153 (1.3) | 1,317 (1,308-1,327) |
| Stress related disorders | 197,786 (3.5) | 3,467 (3,451-3,482) |
| Stroke | 120,435 (2.1) | 2,111 (2,099-2,123) |
| Traumatic brain injury | 250,395 (4.4) | 4,389 (4,371-4,406) |

***Sensitivity analysis 1 / Prevalent cohort 2016***

| | *Prevalence, N (%)* | *Prevalence per 100,000 persons (95% CI)* |
|---|---|---|
| Any brain disorder | 1,128,297 (19.6) | 19,580 (19,544-19,616) |
| Alcohol abuse | 101,636 (1.8) | 1,764 (1,753-1,775) |
| Anxiety disorders | 105,592 (1.8) | 1,832 (1,821-1,843) |
| Bipolar disorder | 24,602 (0.4) | 427 (422-432) |
| Brain tumors | 15,828 (0.3) | 275 (270-279) |
| Cerebral palsy | 9,378 (0.2) | 163 (159-166) |
| Dementia | 41,076 (0.7) | 713 (706-720) |
| Depression | 189,582 (3.3) | 3,290 (3,275-3,305) |
| Developmental and behavioural disorders | 113,617 (2.0) | 1,972 (1,960-1,983) |
| Drug abuse | 52,150 (0.9) | 905 (897-913) |
| Eating disorders | 16,915 (0.3) | 294 (289-298) |
| Epilepsy | 73,771 (1.3) | 1,280 (1,271-1,289) |
| Headache | 104,585 (1.8) | 1,815 (1,804-1,826) |
| Infections of the CNS | 27,514 (0.5) | 477 (472-483) |
| Intellectual disability | 28,593 (0.5) | 496 (490-502) |
| Multiple sclerosis | 15,450 (0.3) | 268 (264-272) |
| Neuromuscular disorders | 7,670 (0.1) | 133 (130-136) |
| Other neurodegenerative disorders | 2,412 (0.0) | 42 (40-44) |
| Parkinson’s disease | 10,275 (0.2) | 178 (175-182) |
| Personality disorders | 73,566 (1.3) | 1,277 (1,267-1,286) |
| Polyneuropathy | 36,182 (0.6) | 628 (621-634) |
| Schizophrenia spectrum disorders | 63,115 (1.1) | 1,095 (1,087-1,104) |
| Sleep disorders | 83,512 (1.4) | 1,449 (1,439-1,459) |
| Stress related disorders | 209,698 (3.6) | 3,639 (3,623-3,655) |
| Stroke | 122,391 (2.1) | 2,124 (2,112-2,136) |
| Traumatic brain injury | 248,104 (4.3) | 4,305 (4,289-4,322) |

***Sensitivity analysis 1 / Prevalent cohort 2017***

| | *Prevalence, N (%)* | *Prevalence per 100,000 persons (95% CI)* |
|---|---|---|
| Any brain disorder | 1,158,443 (20.0) | 20,035 (19,998-20,071) |
| Alcohol abuse | 101,265 (1.8) | 1,751 (1,741-1,762) |
| Anxiety disorders | 112,507 (1.9) | 1,946 (1,934-1,957) |
| Bipolar disorder | 25,454 (0.4) | 440 (435-446) |
| Brain tumors | 16,702 (0.3) | 289 (285-293) |
| Cerebral palsy | 9,442 (0.2) | 163 (160-167) |
| Dementia | 41,484 (0.7) | 717 (711-724) |
| Depression | 195,542 (3.4) | 3,382 (3,367-3,397) |
| Developmental and behavioural disorders | 122,363 (2.1) | 2,116 (2,104-2,128) |
| Drug abuse | 53,667 (0.9) | 928 (920-936) |
| Eating disorders | 17,512 (0.3) | 303 (298-307) |
| Epilepsy | 73,701 (1.3) | 1,275 (1,265-1,284) |
| Headache | 110,114 (1.9) | 1,904 (1,893-1,916) |
| Infections of the CNS | 27,970 (0.5) | 484 (478-489) |
| Intellectual disability | 29,379 (0.5) | 508 (502-514) |
| Multiple sclerosis | 15,940 (0.3) | 276 (271-280) |
| Neuromuscular disorders | 8,027 (0.1) | 139 (136-142) |
| Other neurodegenerative disorders | 2,576 (0.0) | 45 (43-46) |
| Parkinson’s disease | 10,531 (0.2) | 182 (179-186) |
| Personality disorders | 74,393 (1.3) | 1,287 (1,277-1,296) |
| Polyneuropathy | 37,920 (0.7) | 656 (649-662) |
| Schizophrenia spectrum disorders | 64,558 (1.1) | 1,117 (1,108-1,125) |
| Sleep disorders | 91,952 (1.6) | 1,590 (1,580-1,601) |
| Stress related disorders | 221,022 (3.8) | 3,822 (3,807-3,838) |
| Stroke | 124,390 (2.2) | 2,151 (2,139-2,163) |
| Traumatic brain injury | 245,696 (4.2) | 4,249 (4,232-4,266) |

***Sensitivity analysis 1 / Prevalent cohort 2018***

| | *Prevalence, N (%)* | *Prevalence per 100,000 persons (95% CI)* |
|---|---|---|
| Any brain disorder | 1,189,228 (20.5) | 20,471 (20,435-20,508) |
| Alcohol abuse | 100,003 (1.7) | 1,721 (1,711-1,732) |
| Anxiety disorders | 119,340 (2.1) | 2,054 (2,043-2,066) |
| Bipolar disorder | 26,227 (0.5) | 451 (446-457) |
| Brain tumors | 17,420 (0.3) | 300 (295-304) |
| Cerebral palsy | 9,501 (0.2) | 164 (160-167) |
| Dementia | 41,602 (0.7) | 716 (709-723) |
| Depression | 200,949 (3.5) | 3,459 (3,444-3,474) |
| Developmental and behavioural disorders | 131,925 (2.3) | 2,271 (2,259-2,283) |
| Drug abuse | 55,340 (1.0) | 953 (945-961) |
| Eating disorders | 18,261 (0.3) | 314 (310-319) |
| Epilepsy | 73,601 (1.3) | 1,267 (1,258-1,276) |
| Headache | 116,379 (2.0) | 2,003 (1,992-2,015) |
| Infections of the CNS | 28,831 (0.5) | 496 (491-502) |
| Intellectual disability | 30,217 (0.5) | 520 (514-526) |
| Multiple sclerosis | 16,308 (0.3) | 281 (276-285) |
| Neuromuscular disorders | 8,225 (0.1) | 142 (139-145) |
| Other neurodegenerative disorders | 2,752 (0.0) | 47 (46-49) |
| Parkinson’s disease | 10,976 (0.2) | 189 (185-193) |
| Personality disorders | 75,283 (1.3) | 1,296 (1,287-1,305) |
| Polyneuropathy | 39,453 (0.7) | 679 (672-686) |
| Schizophrenia spectrum disorders | 66,225 (1.1) | 1,140 (1,131-1,149) |
| Sleep disorders | 100,924 (1.7) | 1,737 (1,727-1,748) |
| Stress related disorders | 232,012 (4.0) | 3,994 (3,978-4,010) |
| Stroke | 126,495 (2.2) | 2,177 (2,166-2,190) |
| Traumatic brain injury | 243,321 (4.2) | 4,189 (4,172-4,205) |

***Sensitivity analysis 1 / Prevalent cohort 2019***

| | *Prevalence, N (%)* | *Prevalence per 100,000 persons (95% CI)* |
|---|---|---|
| Any brain disorder | 1,215,960 (20.9) | 20,860 (20,823-20,897) |
| Alcohol abuse | 98,227 (1.7) | 1,685 (1,675-1,696) |
| Anxiety disorders | 124,921 (2.1) | 2,143 (2,131-2,155) |
| Bipolar disorder | 26,897 (0.5) | 461 (456-467) |
| Brain tumors | 18,110 (0.3) | 311 (306-315) |
| Cerebral palsy | 9,485 (0.2) | 163 (159-166) |
| Dementia | 40,670 (0.7) | 698 (691-705) |
| Depression | 205,160 (3.5) | 3,520 (3,504-3,535) |
| Developmental and behavioural disorders | 140,656 (2.4) | 2,413 (2,400-2,426) |
| Drug abuse | 56,779 (1.0) | 974 (966-982) |
| Eating disorders | 18,751 (0.3) | 322 (317-326) |
| Epilepsy | 72,679 (1.2) | 1,247 (1,238-1,256) |
| Headache | 121,587 (2.1) | 2,086 (2,074-2,098) |
| Infections of the CNS | 29,588 (0.5) | 508 (502-513) |
| Intellectual disability | 30,697 (0.5) | 527 (521-533) |
| Multiple sclerosis | 16,515 (0.3) | 283 (279-288) |
| Neuromuscular disorders | 8,330 (0.1) | 143 (140-146) |
| Other neurodegenerative disorders | 2,775 (0.0) | 48 (46-49) |
| Parkinson’s disease | 11,060 (0.2) | 190 (186-193) |
| Personality disorders | 75,410 (1.3) | 1,294 (1,284-1,303) |
| Polyneuropathy | 40,756 (0.7) | 699 (692-706) |
| Schizophrenia spectrum disorders | 67,745 (1.2) | 1,162 (1,153-1,171) |
| Sleep disorders | 110,442 (1.9) | 1,895 (1,883-1,906) |
| Stress related disorders | 241,284 (4.1) | 4,139 (4,123-4,156) |
| Stroke | 127,804 (2.2) | 2,192 (2,180-2,205) |
| Traumatic brain injury | 240,801 (4.1) | 4,131 (4,114-4,147) |

***Sensitivity analysis 1 / Prevalent cohort 2020***

| | *Prevalence, N (%)* | *Prevalence per 100,000 persons (95% CI)* |
|---|---|---|
| Any brain disorder | 1,236,961 (21.1) | 21,103 (21,066-21,140) |
| Alcohol abuse | 96,009 (1.6) | 1,638 (1,628-1,648) |
| Anxiety disorders | 129,338 (2.2) | 2,207 (2,195-2,219) |
| Bipolar disorder | 27,604 (0.5) | 471 (465-477) |
| Brain tumors | 18,927 (0.3) | 323 (318-328) |
| Cerebral palsy | 9,403 (0.2) | 160 (157-164) |
| Dementia | 39,784 (0.7) | 679 (672-685) |
| Depression | 207,650 (3.5) | 3,543 (3,527-3,558) |
| Developmental and behavioural disorders | 148,992 (2.5) | 2,542 (2,529-2,555) |
| Drug abuse | 58,052 (1.0) | 990 (982-998) |
| Eating disorders | 19,236 (0.3) | 328 (324-333) |
| Epilepsy | 71,789 (1.2) | 1,225 (1,216-1,234) |
| Headache | 125,850 (2.1) | 2,147 (2,135-2,159) |
| Infections of the CNS | 30,314 (0.5) | 517 (511-523) |
| Intellectual disability | 30,982 (0.5) | 529 (523-534) |
| Multiple sclerosis | 16,960 (0.3) | 289 (285-294) |
| Neuromuscular disorders | 8,454 (0.1) | 144 (141-147) |
| Other neurodegenerative disorders | 2,839 (0.0) | 48 (47-50) |
| Parkinson’s disease | 11,245 (0.2) | 192 (188-195) |
| Personality disorders | 75,678 (1.3) | 1,291 (1,282-1,300) |
| Polyneuropathy | 42,758 (0.7) | 729 (723-736) |
| Schizophrenia spectrum disorders | 69,162 (1.2) | 1,180 (1,171-1,189) |
| Sleep disorders | 118,983 (2.0) | 2,030 (2,018-2,041) |
| Stress related disorders | 249,672 (4.3) | 4,259 (4,243-4,276) |
| Stroke | 129,247 (2.2) | 2,205 (2,193-2,217) |
| Traumatic brain injury | 238,172 (4.1) | 4,063 (4,047-4,080) |

***Sensitivity analysis 1 / Prevalent cohort 2021***

| | *Prevalence, N (%)* | *Prevalence per 100,000 persons (95% CI)* |
|---|---|---|
| Any brain disorder | 1,253,399 (21.4) | 21,401 (21,364-21,439) |
| Alcohol abuse | 93,710 (1.6) | 1,600 (1,590-1,610) |
| Anxiety disorders | 133,733 (2.3) | 2,283 (2,271-2,296) |
| Bipolar disorder | 28,154 (0.5) | 481 (475-486) |
| Brain tumors | 19,592 (0.3) | 335 (330-339) |
| Cerebral palsy | 9,351 (0.2) | 160 (156-163) |
| Dementia | 38,844 (0.7) | 663 (657-670) |
| Depression | 209,483 (3.6) | 3,577 (3,562-3,592) |
| Developmental and behavioural disorders | 157,353 (2.7) | 2,687 (2,673-2,700) |
| Drug abuse | 59,114 (1.0) | 1,009 (1,001-1,018) |
| Eating disorders | 19,756 (0.3) | 337 (333-342) |
| Epilepsy | 70,944 (1.2) | 1,211 (1,202-1,220) |
| Headache | 129,706 (2.2) | 2,215 (2,203-2,227) |
| Infections of the CNS | 30,347 (0.5) | 518 (512-524) |
| Intellectual disability | 30,980 (0.5) | 529 (523-535) |
| Multiple sclerosis | 17,343 (0.3) | 296 (292-301) |
| Neuromuscular disorders | 8,606 (0.1) | 147 (144-150) |
| Other neurodegenerative disorders | 2,857 (0.0) | 49 (47-51) |
| Parkinson’s disease | 11,232 (0.2) | 192 (188-195) |
| Personality disorders | 75,906 (1.3) | 1,296 (1,287-1,305) |
| Polyneuropathy | 44,577 (0.8) | 761 (754-768) |
| Schizophrenia spectrum disorders | 70,597 (1.2) | 1,205 (1,197-1,214) |
| Sleep disorders | 126,128 (2.2) | 2,154 (2,142-2,165) |
| Stress related disorders | 255,796 (4.4) | 4,368 (4,351-4,385) |
| Stroke | 131,130 (2.2) | 2,239 (2,227-2,251) |
| Traumatic brain injury | 233,203 (4.0) | 3,982 (3,966-3,998) |

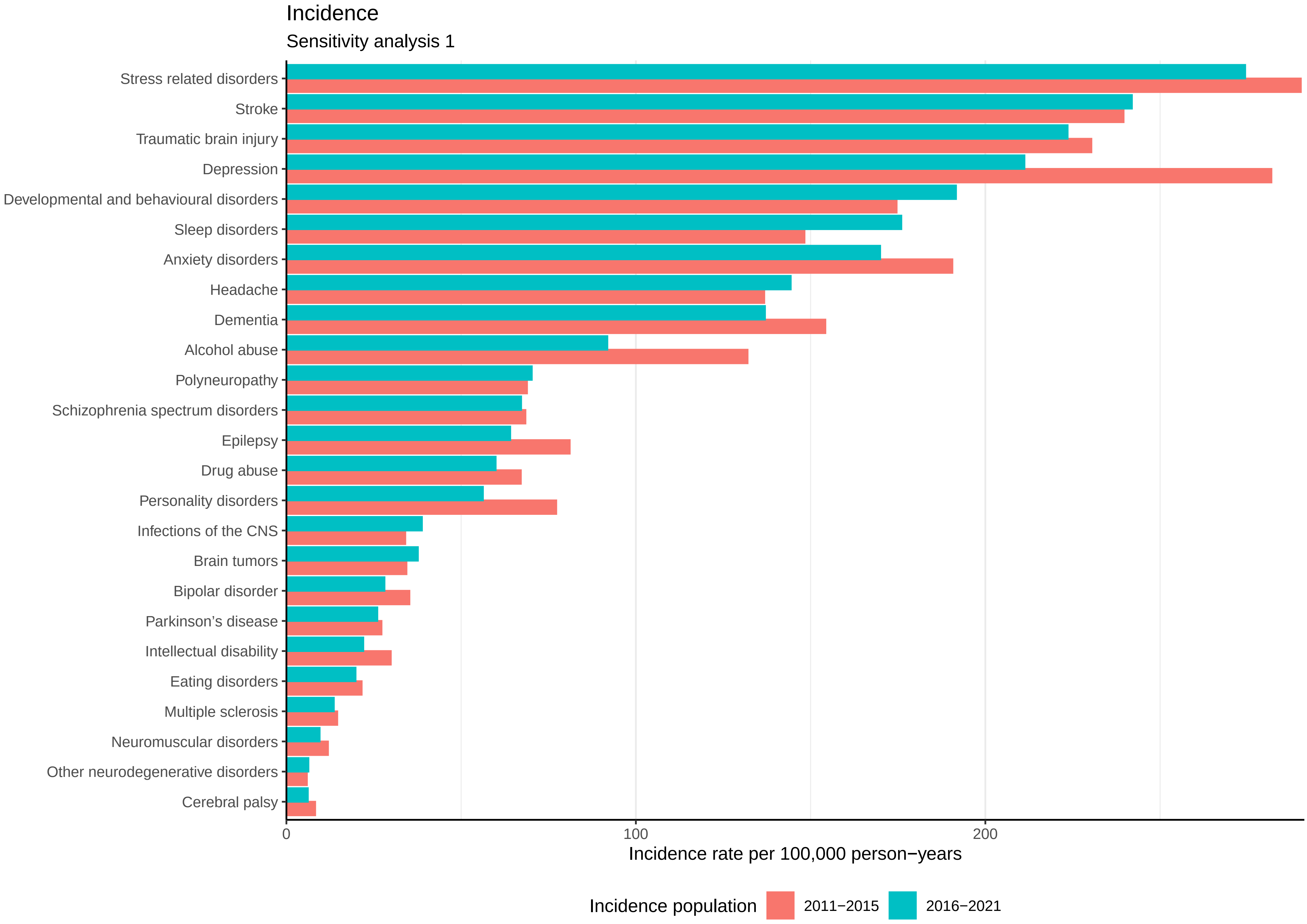

Incidence
Sensitivity analysis 1
Stress related disorders
Stroke
Traumatic brain injury
Depression
Developmental and behavioural disorders
Sleep disorders
Anxiety disorders
Headache
Dementia
Alcohol abuse
Polyneuropathy
Schizophrenia spectrum disorders
Epilepsy
Drug abuse
Personality disorders
Infections of the CNS
Brain tumors
Bipolar disorder
Parkinson’s disease
Intellectual disability
Eating disorders
Multiple sclerosis
Neuromuscular disorders
Other neurodegenerative disorders
Cerebral palsy
0
100
200
Incidence rate per 100,000 person−years
Incidence population
2011−2015
2016−2021

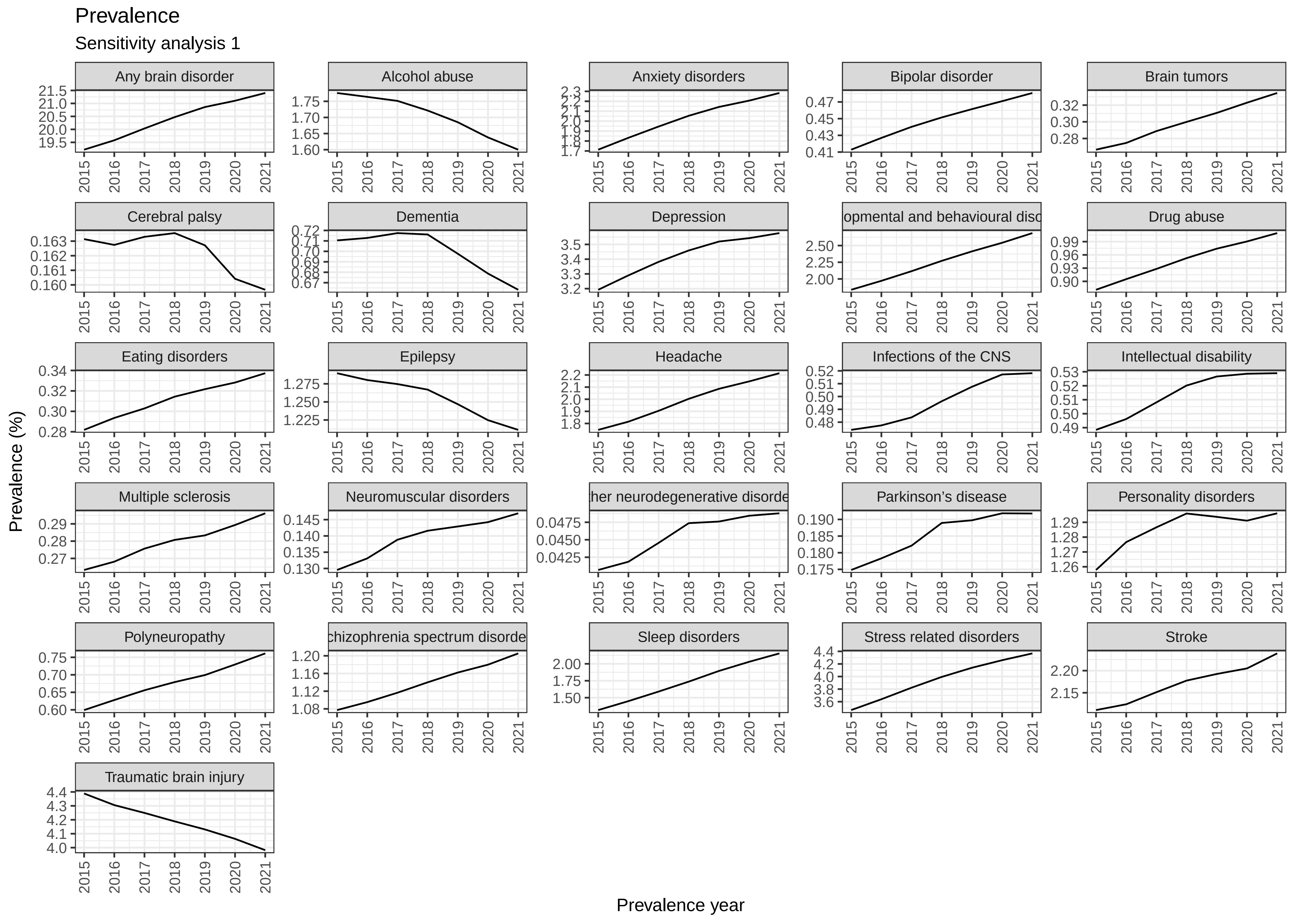

Prevalence
Sensitivity analysis 1
Any brain disorder
Alcohol abuse
Anxiety disorders
Bipolar disorder
Brain tumors
Cerebral palsy
Dementia
Depression
opmental and behavioural diso
Drug abuse
Eating disorders
Epilepsy
Headache
Infections of the CNS
Intellectual disability
Multiple sclerosis
Neuromuscular disorders
her neurodegenerative disorde
Parkinson's disease
Personality disorders
Polyneuropathy
chizophrenia spectrum disorde
Sleep disorders
Stress related disorders
Stroke
Traumatic brain injury
Prevalence (%)
Prevalence year

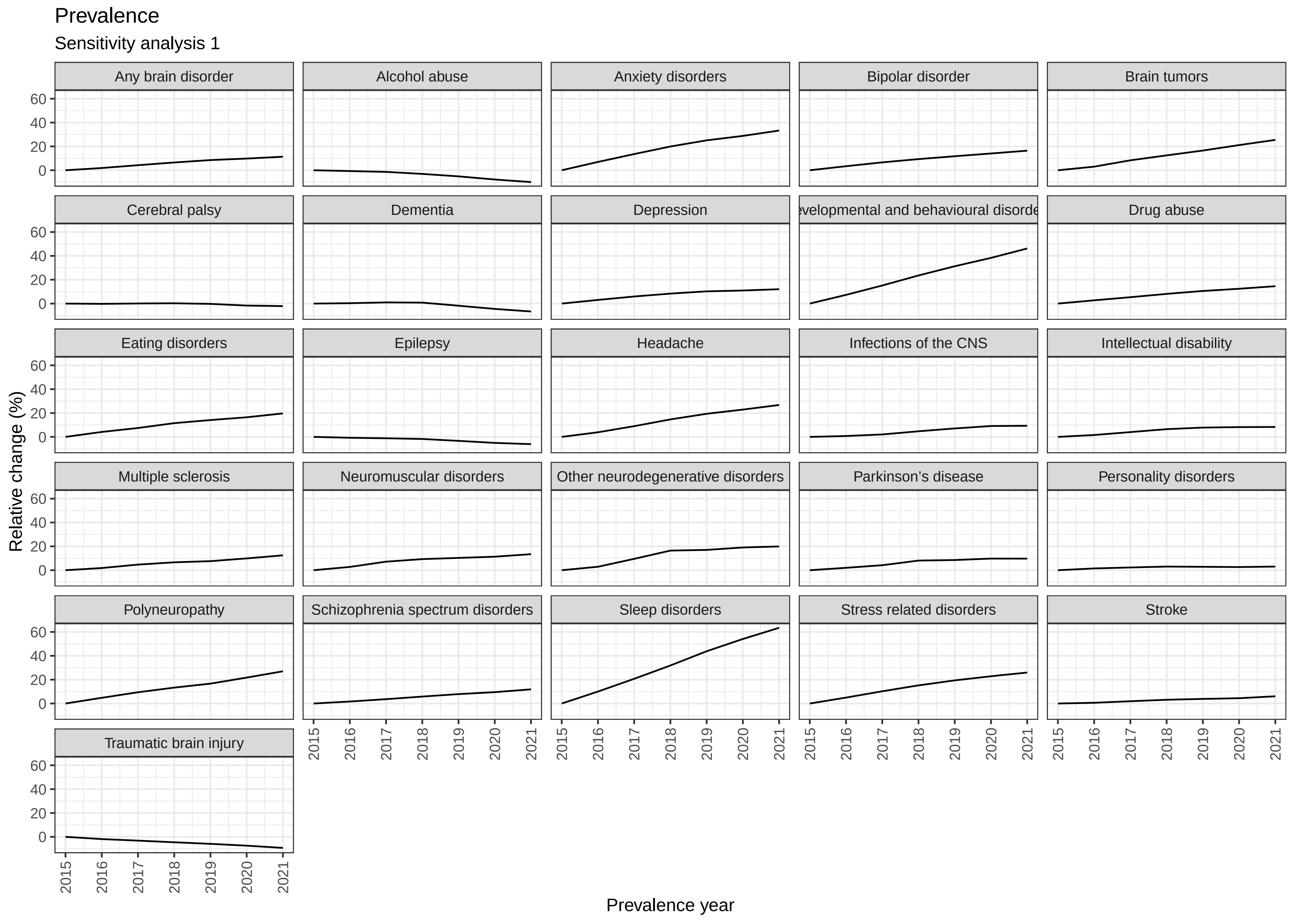

Prevalence
Sensitivity analysis 1
Any brain disorder
Alcohol abuse
Anxiety disorders
Bipolar disorder
Brain tumors
Cerebral palsy
Dementia
Depression
evelopmental and behavioural disorde
Drug abuse
Eating disorders
Epilepsy
Headache
Infections of the CNS
Intellectual disability
Multiple sclerosis
Neuromuscular disorders
Other neurodegenerative disorders
Parkinson's disease
Personality disorders
Polyneuropathy
Schizophrenia spectrum disorders
Sleep disorders
Stress related disorders
Stroke
Traumatic brain injury
Relative change (%)
Prevalence year

***Sensitivity analysis 1 / Incident cohort 2011-2015***

| *Brain disorder* | *Patient group* | | *Comparison group* | | |
|---|---|---|---|---|---|
| | *Persons, N* | *Deaths, N(%)* | *Persons, N* | *Deaths, N(%)* | *Adjusted HR (95% CI)* |
| Any brain disorder | 397,926 | 29,619 (7.4) | 3,971,775 | 55,854 (1.4) | 5.5 (5.4-5.6) |
| Alcohol abuse | 37,286 | 3,631 (9.7) | 372,701 | 4,282 (1.1) | 7.8 (7.4-8.1) |
| Anxiety disorders | 53,884 | 7,332 (13.6) | 536,764 | 4,790 (0.9) | 12.4 (11.9-12.8) |
| Bipolar disorder | 9,940 | 269 (2.7) | 99,082 | 736 (0.7) | 3.1 (2.7-3.6) |
| Brain tumors | 9,721 | 2,293 (23.6) | 97,173 | 2,030 (2.1) | 13.2 (12.4-14.0) |
| Cerebral palsy | 2,331 | 144 (6.2) | 23,257 | 179 (0.8) | 6.8 (5.4-8.5) |
| Dementia | 43,694 | 8,702 (19.9) | 436,871 | 30,006 (6.9) | 3.2 (3.1-3.3) |
| Depression | 79,731 | 6,194 (7.8) | 794,463 | 13,309 (1.7) | 4.1 (3.9-4.2) |
| Developmental and behavioural disorders | 49,383 | 303 (0.6) | 492,311 | 391 (0.1) | 6.6 (5.7-7.7) |
| Drug abuse | 18,915 | 505 (2.7) | 188,421 | 796 (0.4) | 5.2 (4.6-5.8) |
| Eating disorders | 6,087 | 14 (0.2) | 60,492 | 32 (0.1) | 4.9 (2.6-9.4) |
| Epilepsy | 22,907 | 2,776 (12.1) | 229,083 | 4,167 (1.8) | 5.2 (4.9-5.5) |
| Headache | 38,649 | 212 (0.5) | 385,362 | 1,714 (0.4) | 1.0 (0.8-1.1) |
| Infections of the CNS | 9,609 | 756 (7.9) | 96,007 | 1,044 (1.1) | 7.2 (6.5-7.9) |
| Intellectual disability | 8,450 | 181 (2.1) | 84,262 | 199 (0.2) | 8.9 (7.3-10.9) |
| Multiple sclerosis | 4,110 | 59 (1.4) | 40,976 | 152 (0.4) | 3.5 (2.6-4.8) |
| Neuromuscular disorders | 3,368 | 293 (8.7) | 33,627 | 447 (1.3) | 5.7 (4.9-6.7) |
| Other neurodegenerative disorders | 1,660 | 402 (24.2) | 16,589 | 326 (2.0) | 14.3 (12.4-16.6) |
| Parkinson’s disease | 7,712 | 921 (11.9) | 77,084 | 3,397 (4.4) | 2.8 (2.6-3.1) |
| Personality disorders | 21,832 | 153 (0.7) | 216,677 | 369 (0.2) | 3.1 (2.6-3.8) |
| Polyneuropathy | 19,498 | 1,087 (5.6) | 194,792 | 4,868 (2.5) | 1.7 (1.6-1.8) |
| Schizophrenia spectrum disorders | 19,269 | 593 (3.1) | 192,355 | 1,686 (0.9) | 3.4 (3.1-3.7) |
| Sleep disorders | 41,958 | 759 (1.8) | 418,706 | 3,468 (0.8) | 1.5 (1.4-1.7) |
| Stress related disorders | 81,999 | 1,243 (1.5) | 817,602 | 3,371 (0.4) | 3.0 (2.8-3.2) |
| Stroke | 67,848 | 14,052 (20.7) | 678,201 | 28,724 (4.2) | 5.6 (5.5-5.7) |
| Traumatic brain injury | 65,087 | 3,847 (5.9) | 649,752 | 10,893 (1.7) | 3.6 (3.5-3.7) |

***Sensitivity analysis 1 / Incident cohort 2016-2021***

| *Brain disorder* | *Patient group* | | *Comparison group* | | |
|---|---|---|---|---|---|
| | *Persons, N* | *Deaths, N(%)* | *Persons, N* | *Deaths, N(%)* | *Adjusted HR (95% CI)* |
| Any brain disorder | 463,442 | 29,265 (6.3) | 4,625,490 | 63,359 (1.4) | 4.9 (4.8-4.9) |
| Alcohol abuse | 31,920 | 3,313 (10.4) | 319,623 | 3,785 (1.2) | 8.5 (8.1-8.9) |
| Anxiety disorders | 59,202 | 5,390 (9.1) | 589,333 | 4,986 (0.8) | 9.6 (9.2-10.0) |
| Bipolar disorder | 9,749 | 189 (1.9) | 97,228 | 522 (0.5) | 3.4 (2.9-4.0) |
| Brain tumors | 13,134 | 2,783 (21.2) | 131,180 | 2,900 (2.2) | 11.3 (10.8-12.0) |
| Cerebral palsy | 2,133 | 142 (6.7) | 21,332 | 158 (0.7) | 8.1 (6.4-10.2) |
| Dementia | 47,846 | 7,939 (16.6) | 478,494 | 31,038 (6.5) | 2.8 (2.7-2.9) |
| Depression | 73,587 | 4,779 (6.5) | 732,940 | 10,431 (1.4) | 4.3 (4.1-4.4) |
| Developmental and behavioural disorders | 66,777 | 205 (0.3) | 665,250 | 461 (0.1) | 4.0 (3.4-4.7) |
| Drug abuse | 20,807 | 555 (2.7) | 207,056 | 710 (0.3) | 6.3 (5.7-7.1) |
| Eating disorders | 6,884 | 18 (0.3) | 68,511 | 25 (0.0) | 8.5 (4.5-16.0) |
| Epilepsy | 22,295 | 2,772 (12.4) | 222,957 | 4,181 (1.9) | 5.6 (5.3-5.9) |
| Headache | 50,252 | 255 (0.5) | 500,913 | 2,095 (0.4) | 1.0 (0.9-1.2) |
| Infections of the CNS | 13,514 | 820 (6.1) | 134,841 | 1,433 (1.1) | 6.0 (5.5-6.5) |
| Intellectual disability | 7,663 | 135 (1.8) | 76,381 | 130 (0.2) | 10.3 (8.1-13.1) |
| Multiple sclerosis | 4,722 | 36 (0.8) | 47,101 | 165 (0.4) | 2.1 (1.5-3.0) |
| Neuromuscular disorders | 3,318 | 188 (5.7) | 33,113 | 473 (1.4) | 3.7 (3.1-4.4) |
| Other neurodegenerative disorders | 2,201 | 552 (25.1) | 21,967 | 401 (1.8) | 16.3 (14.3-18.5) |
| Parkinson's disease | 9,090 | 912 (10.0) | 90,877 | 3,742 (4.1) | 2.6 (2.4-2.8) |
| Personality disorders | 19,604 | 75 (0.4) | 194,270 | 228 (0.1) | 2.8 (2.2-3.6) |
| Polyneuropathy | 24,518 | 1,277 (5.2) | 244,940 | 6,056 (2.5) | 1.5 (1.4-1.6) |
| Schizophrenia spectrum disorders | 23,308 | 509 (2.2) | 232,607 | 1,285 (0.6) | 3.8 (3.4-4.2) |
| Sleep disorders | 61,408 | 748 (1.2) | 612,249 | 5,496 (0.9) | 1.0 (1.0-1.1) |
| Stress related disorders | 95,445 | 1,156 (1.2) | 951,812 | 3,503 (0.4) | 2.8 (2.6-3.0) |
| Stroke | 84,507 | 15,736 (18.6) | 844,641 | 35,216 (4.2) | 5.1 (5.0-5.2) |
| Traumatic brain injury | 77,847 | 5,720 (7.3) | 776,847 | 16,506 (2.1) | 3.6 (3.5-3.7) |

#### Mortality

Sensitivity analysis 1 / Incident cohort 2011-2015

| Brain disorder | Patient group -Deaths (%) | Comparison group- Deaths (%) | Adjusted hazard ratio (95% CI) |
|---|---|---|---|
| Any brain disorder | 29,619 (7.4) | 55,854 (1.4) | 5.5 (5.4-5.6) |
| Other neurodegenerative disorders | 402 (24.2) | 326 (2.0) | 14.3 (12.4-16.6) |
| Brain tumors | 2,293 (23.6) | 2,030 (2.1) | 13.2 (12.4-14.0) |
| Anxiety disorders | 7,332 (13.6) | 4,790 (0.9) | 12.4 (11.9-12.8) |
| Intellectual disability | 181 (2.1) | 199 (0.2) | 8.9 (7.3-10.9) |
| Alcohol abuse | 3,631 (9.7) | 4,282 (1.1) | 7.8 (7.4-8.1) |
| Infections of the CNS | 756 (7.9) | 1,044 (1.1) | 7.2 (6.5-7.9) |
| Cerebral palsy | 144 (6.2) | 179 (0.8) | 6.8 (5.4-8.5) |
| Developmental and behavioural disorders | 303 (0.6) | 391 (0.1) | 6.6 (5.7-7.7) |
| Neuromuscular disorders | 293 (8.7) | 447 (1.3) | 5.7 (4.9-6.7) |
| Stroke | 14,052 (20.7) | 28,724 (4.2) | 5.6 (5.5-5.7) |
| Epilepsy | 2,776 (12.1) | 4,167 (1.8) | 5.2 (4.9-5.5) |
| Drug abuse | 505 (2.7) | 796 (0.4) | 5.2 (4.6-5.8) |
| Eating disorders | 14 (0.2) | 32 (0.1) | 4.9 (2.6-9.4) |
| Depression | 6,194 (7.8) | 13,309 (1.7) | 4.1 (3.9-4.2) |
| Traumatic brain injury | 3,847 (5.9) | 10,893 (1.7) | 3.6 (3.5-3.7) |
| Multiple sclerosis | 59 (1.4) | 152 (0.4) | 3.5 (2.6-4.8) |
| Schizophrenia spectrum disorders | 593 (3.1) | 1,686 (0.9) | 3.4 (3.1-3.7) |
| Dementia | 8,702 (19.9) | 30,006 (6.9) | 3.2 (3.1-3.3) |
| Personality disorders | 153 (0.7) | 369 (0.2) | 3.1 (2.6-3.8) |
| Bipolar disorder | 269 (2.7) | 736 (0.7) | 3.1 (2.7-3.6) |
| Stress related disorders | 1,243 (1.5) | 3,371 (0.4) | 3.0 (2.8-3.2) |
| Parkinson’s disease | 921 (11.9) | 3,397 (4.4) | 2.8 (2.6-3.1) |
| Polyneuropathy | 1,087 (5.6) | 4,868 (2.5) | 1.7 (1.6-1.8) |
| Sleep disorders | 759 (1.8) | 3,468 (0.8) | 1.5 (1.4-1.7) |
| Headache | 212 (0.5) | 1,714 (0.4) | 1.0 (0.8-1.1) |

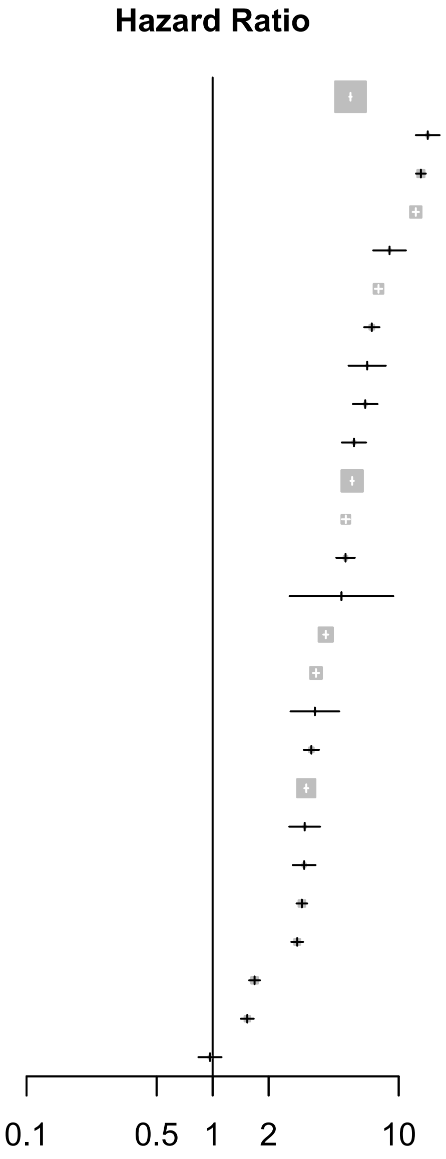

### Mortality

Sensitivity analysis 1 / Incident cohort 2016-2021

| Brain disorder | Patient group -Deaths (%) | Comparison group- Deaths (%) | Adjusted hazard ratio (95% CI) |
|---|---|---|---|
| Any brain disorder | 29,265 (6.3) | 63,359 (1.4) | 4.9 (4.8-4.9) |
| Other neurodegenerative disorders | 552 (25.1) | 401 (1.8) | 16.3 (14.3-18.5) |
| Brain tumors | 2,783 (21.2) | 2,900 (2.2) | 11.3 (10.8-12.0) |
| Intellectual disability | 135 (1.8) | 130 (0.2) | 10.3 (8.1-13.1) |
| Anxiety disorders | 5,390 (9.1) | 4,986 (0.8) | 9.6 (9.2-10.0) |
| Eating disorders | 18 (0.3) | 25 (0.0) | 8.5 (4.5-16.0) |
| Alcohol abuse | 3,313 (10.4) | 3,785 (1.2) | 8.5 (8.1-8.9) |
| Cerebral palsy | 142 (6.7) | 158 (0.7) | 8.1 (6.4-10.2) |
| Drug abuse | 555 (2.7) | 710 (0.3) | 6.3 (5.7-7.1) |
| Infections of the CNS | 820 (6.1) | 1,433 (1.1) | 6.0 (5.5-6.5) |
| Epilepsy | 2,772 (12.4) | 4,181 (1.9) | 5.6 (5.3-5.9) |
| Stroke | 15,736 (18.6) | 35,216 (4.2) | 5.1 (5.0-5.2) |
| Depression | 4,779 (6.5) | 10,431 (1.4) | 4.3 (4.1-4.4) |
| Developmental and behavioural disorders | 205 (0.3) | 461 (0.1) | 4.0 (3.4-4.7) |
| Schizophrenia spectrum disorders | 509 (2.2) | 1,285 (0.6) | 3.8 (3.4-4.2) |
| Neuromuscular disorders | 188 (5.7) | 473 (1.4) | 3.7 (3.1-4.4) |
| Traumatic brain injury | 5,720 (7.3) | 16,506 (2.1) | 3.6 (3.5-3.7) |
| Bipolar disorder | 189 (1.9) | 522 (0.5) | 3.4 (2.9-4.0) |
| Personality disorders | 75 (0.4) | 228 (0.1) | 2.8 (2.2-3.6) |
| Stress related disorders | 1,156 (1.2) | 3,503 (0.4) | 2.8 (2.6-3.0) |
| Dementia | 7,939 (16.6) | 31,038 (6.5) | 2.8 (2.7-2.9) |
| Parkinson’s disease | 912 (10.0) | 3,742 (4.1) | 2.6 (2.4-2.8) |
| Multiple sclerosis | 36 (0.8) | 165 (0.4) | 2.1 (1.5-3.0) |
| Polyneuropathy | 1,277 (5.2) | 6,056 (2.5) | 1.5 (1.4-1.6) |
| Sleep disorders | 748 (1.2) | 5,496 (0.9) | 1.0 (1.0-1.1) |
| Headache | 255 (0.5) | 2,095 (0.4) | 1.0 (0.9-1.2) |

**Hazard Ratio**

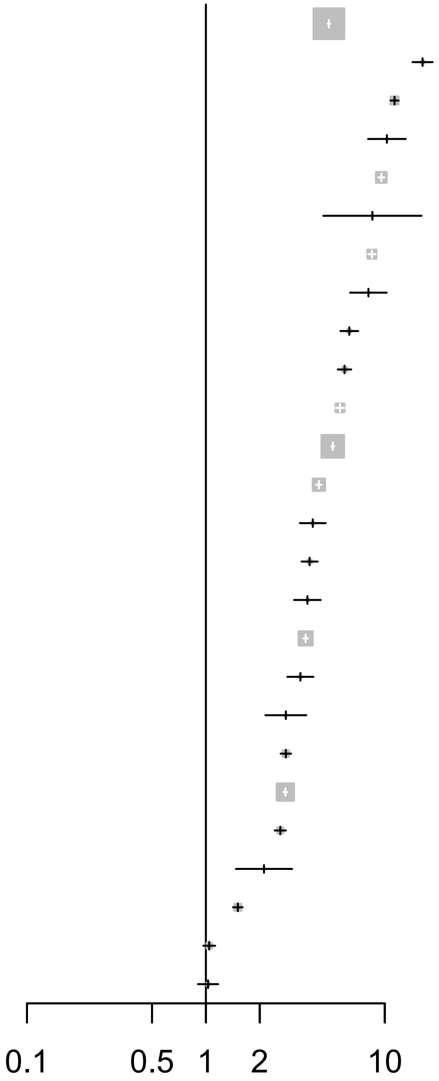

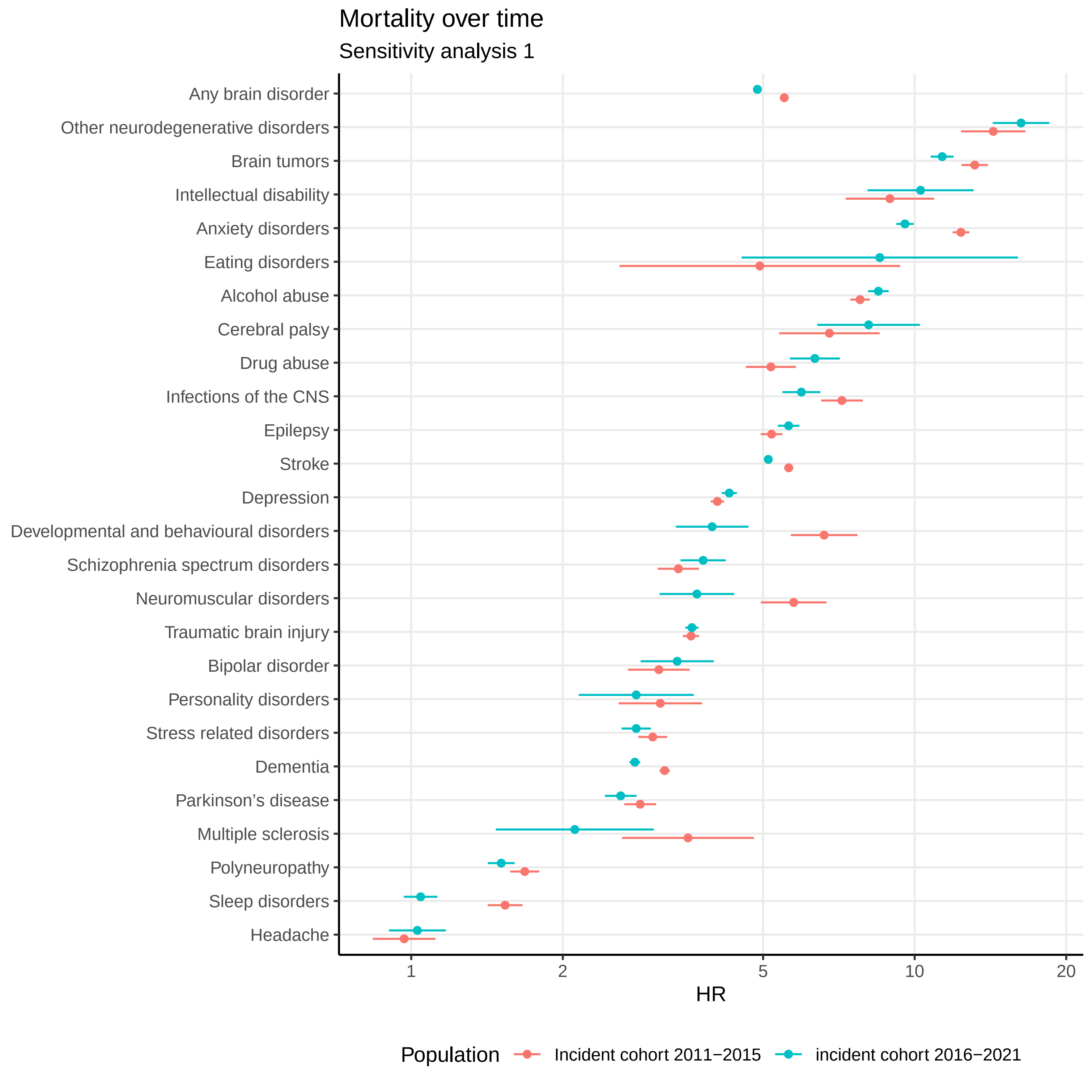

Mortality over time
Sensitivity analysis 1
Any brain disorder
Other neurodegenerative disorders
Brain tumors
Intellectual disability
Anxiety disorders
Eating disorders
Alcohol abuse
Cerebral palsy
Drug abuse
Infections of the CNS
Epilepsy
Stroke
Depression
Developmental and behavioural disorders
Schizophrenia spectrum disorders
Neuromuscular disorders
Traumatic brain injury
Bipolar disorder
Personality disorders
Stress related disorders
Dementia
Parkinson’s disease
Multiple sclerosis
Polyneuropathy
Sleep disorders
Headache
1
2
5
10
20
HR
Population
Incident cohort 2011−2015
incident cohort 2016−2021

***Sensitivity analysis 1 / Incident cohort 2011-2015***

| *Brain disorder* | *Cost component* | *Actual costs (EUR)* | *Actual costs per person* | *Attributable costs (EUR)* | *Attributable costs per person* |
|---|---|---|---|---|---|
| Any brain disorder | Primary sector | 176,853,349 | 469 | 67,283,811 | 178 |
| | Hospital care | 5,621,629,182 | 14,908 | 4,882,739,614 | 12,948 |
| | Filled prescriptions | 180,607,378 | 479 | 100,395,562 | 266 |
| | Home care | 341,944,826 | 907 | 247,688,826 | 657 |
| | Nursing home / sheltered accomodation | 724,892,288 | 1,922 | 560,459,633 | 1,486 |
| | Lost productivity associated with illness | . | . | 2,986,784,517 | 15,534 |
| | Total cost (excl. lost productivity) | 7,045,927,023 | 18,685 | 5,858,567,446 | 15,536 |
| Alcohol abuse | Primary sector | 16,800,862 | 483 | 5,642,527 | 162 |
| | Hospital care | 762,797,168 | 21,913 | 675,731,752 | 19,412 |
| | Filled prescriptions | 22,178,264 | 637 | 12,020,706 | 345 |
| | Home care | 35,021,998 | 1,006 | 27,874,190 | 801 |
| | Nursing home / sheltered accomodation | 57,752,555 | 1,659 | 42,756,650 | 1,228 |
| | Lost productivity associated with illness | . | . | 746,201,114 | 32,442 |
| | Total cost (excl. lost productivity) | 894,550,847 | 25,698 | 764,025,825 | 21,949 |
| Anxiety disorders | Primary sector | 25,714,491 | 532 | 12,332,800 | 255 |
| | Hospital care | 866,144,069 | 17,929 | 774,676,734 | 16,035 |
| | Filled prescriptions | 34,375,197 | 712 | 24,872,902 | 515 |
| | Home care | 26,384,502 | 546 | 18,708,830 | 387 |
| | Nursing home / sheltered accomodation | 39,871,728 | 825 | 21,475,796 | 445 |
| | Lost productivity associated with illness | . | . | 776,939,071 | 22,758 |
| | Total cost (excl. lost productivity) | 992,489,986 | 20,544 | 852,067,063 | 17,637 |
| Bipolar disorder | Primary sector | 5,048,620 | 515 | 2,092,910 | 214 |
| | Hospital care | 291,626,398 | 29,773 | 271,281,867 | 27,696 |
| | Filled prescriptions | 9,739,617 | 994 | 7,534,847 | 769 |
| | Home care | 4,919,598 | 502 | 3,330,917 | 340 |
| | Nursing home / sheltered accomodation | 12,397,228 | 1,266 | 8,490,507 | 867 |
| | Lost productivity associated with illness | . | . | 235,864,828 | 29,198 |
| | Total cost (excl. lost productivity) | 323,731,460 | 33,051 | 292,731,048 | 29,886 |
| Brain tumors | Primary sector | 4,761,849 | 574 | 1,573,904 | 190 |
| | Hospital care | 412,782,835 | 49,756 | 387,553,840 | 46,715 |
| | Filled prescriptions | 5,507,517 | 664 | 2,409,631 | 290 |
| | Home care | 11,769,533 | 1,419 | 8,296,526 | 1,000 |
| | Nursing home / sheltered accomodation | 11,569,206 | 1,395 | 3,049,168 | 368 |

***Sensitivity analysis 1 / Incident cohort 2011-2015***

| *Brain disorder* | *Cost component* | *Actual costs (EUR)* | *Actual costs per person* | *Attributable costs (EUR)* | *Attributable costs per person* |
|---|---|---|---|---|---|
| | Lost productivity associated with illness | . | . | 21,421,603 | 5,785 |
| | Total cost (excl. lost productivity) | 446,390,939 | 53,807 | 402,883,069 | 48,562 |
| Cerebral palsy | Primary sector | 1,456,451 | 652 | 845,935 | 379 |
| | Hospital care | 39,147,738 | 17,536 | 35,688,905 | 15,987 |
| | Filled prescriptions | 1,595,967 | 715 | 1,231,595 | 552 |
| | Home care | 5,813,039 | 2,604 | 5,536,717 | 2,480 |
| | Nursing home / sheltered accomodation | 8,193,681 | 3,670 | 7,323,138 | 3,280 |
| | Lost productivity associated with illness | . | . | 23,039,761 | 36,864 |
| | Total cost (excl. lost productivity) | 56,206,876 | 25,178 | 50,626,290 | 22,678 |
| Dementia | Primary sector | 23,804,780 | 624 | 3,746,014 | 98 |
| | Hospital care | 527,936,178 | 13,839 | 340,200,609 | 8,918 |
| | Filled prescriptions | 40,658,161 | 1,066 | 18,198,391 | 477 |
| | Home care | 206,835,530 | 5,422 | 138,320,965 | 3,626 |
| | Nursing home / sheltered accomodation | 744,870,688 | 19,526 | 598,416,978 | 15,687 |
| | Lost productivity associated with illness | . | . | 124,385,679 | 55,184 |
| | Total cost (excl. lost productivity) | 1,544,105,337 | 40,477 | 1,098,882,957 | 28,806 |
| Depression | Primary sector | 44,845,471 | 593 | 20,397,271 | 270 |
| | Hospital care | 1,559,796,854 | 20,620 | 1,383,506,873 | 18,290 |
| | Filled prescriptions | 55,054,376 | 728 | 35,194,387 | 465 |
| | Home care | 97,920,749 | 1,294 | 69,194,132 | 915 |
| | Nursing home / sheltered accomodation | 211,781,913 | 2,800 | 137,194,516 | 1,814 |
| | Lost productivity associated with illness | . | . | 1,286,267,581 | 25,569 |
| | Total cost (excl. lost productivity) | 1,969,399,363 | 26,035 | 1,645,487,178 | 21,753 |
| Developmental and behavioural disorders | Primary sector | 12,951,102 | 263 | 4,909,947 | 100 |
| | Hospital care | 591,506,940 | 12,016 | 545,568,403 | 11,082 |
| | Filled prescriptions | 28,137,145 | 572 | 24,402,044 | 496 |
| | Home care | 2,241,382 | 46 | 1,476,502 | 30 |
| | Nursing home / sheltered accomodation | 5,180,705 | 105 | 3,999,197 | 81 |
| | Lost productivity associated with illness | . | . | 397,667,715 | 25,399 |
| | Total cost (excl. lost productivity) | 640,017,273 | 13,001 | 580,356,093 | 11,789 |
| Drug abuse | Primary sector | 7,220,957 | 387 | 3,097,987 | 166 |
| | Hospital care | 433,879,771 | 23,281 | 406,432,643 | 21,808 |
| | Filled prescriptions | 12,888,395 | 692 | 10,064,936 | 540 |

*Sensitivity analysis 1 / Incident cohort 2011-2015*

| *Brain disorder* | *Cost component* | *Actual costs (EUR)* | *Actual costs per person* | *Attributable costs (EUR)* | *Attributable costs per person* |
|---|---|---|---|---|---|
| | Home care | 8,514,277 | 457 | 6,799,542 | 365 |
| | Nursing home / sheltered accomodation | 11,715,821 | 629 | 7,645,762 | 410 |
| | Lost productivity associated with illness | . | . | 422,672,229 | 27,727 |
| | Total cost (excl. lost productivity) | 474,219,221 | 25,445 | 434,040,870 | 23,289 |
| Eating disorders | Primary sector | 2,105,606 | 346 | 643,700 | 106 |
| | Hospital care | 220,014,012 | 36,140 | 210,934,156 | 34,649 |
| | Filled prescriptions | 1,384,863 | 227 | 630,977 | 104 |
| | Home care | 338,539 | 56 | 247,205 | 41 |
| | Nursing home / sheltered accomodation | 447,829 | 74 | 374,603 | 62 |
| | Lost productivity associated with illness | . | . | 26,789,067 | 7,268 |
| | Total cost (excl. lost productivity) | 224,290,850 | 36,843 | 212,830,640 | 34,960 |
| Epilepsy | Primary sector | 11,405,817 | 540 | 4,595,806 | 218 |
| | Hospital care | 448,947,402 | 21,253 | 395,207,916 | 18,709 |
| | Filled prescriptions | 17,250,052 | 817 | 11,319,395 | 536 |
| | Home care | 39,717,638 | 1,880 | 31,910,613 | 1,511 |
| | Nursing home / sheltered accomodation | 99,465,592 | 4,709 | 79,519,465 | 3,764 |
| | Lost productivity associated with illness | . | . | 188,581,749 | 21,759 |
| | Total cost (excl. lost productivity) | 616,786,502 | 29,198 | 522,553,195 | 24,737 |
| Headache | Primary sector | 18,528,152 | 480 | 7,685,090 | 199 |
| | Hospital care | 253,323,636 | 6,566 | 181,062,932 | 4,693 |
| | Filled prescriptions | 13,436,278 | 348 | 5,746,577 | 149 |
| | Home care | 4,568,795 | 118 | 682,888 | 18 |
| | Nursing home / sheltered accomodation | 5,950,963 | 154 | -1,973,052 | -51 |
| | Lost productivity associated with illness | . | . | 155,956,744 | 5,475 |
| | Total cost (excl. lost productivity) | 295,807,824 | 7,668 | 193,204,435 | 5,008 |
| Infections of the CNS | Primary sector | 4,042,719 | 447 | 1,273,453 | 141 |
| | Hospital care | 247,282,556 | 27,362 | 227,147,277 | 25,134 |
| | Filled prescriptions | 3,836,493 | 425 | 1,668,080 | 185 |
| | Home care | 7,926,142 | 877 | 5,939,782 | 657 |
| | Nursing home / sheltered accomodation | 10,239,371 | 1,133 | 5,748,740 | 636 |
| | Lost productivity associated with illness | . | . | 9,800,043 | 2,020 |
| | Total cost (excl. lost productivity) | 273,327,281 | 30,244 | 241,777,332 | 26,753 |
| Intellectual disability | Primary sector | 3,255,411 | 390 | 1,538,119 | 184 |
| | Hospital care | 129,888,412 | 15,573 | 119,255,543 | 14,299 |
| | Filled prescriptions | 5,653,343 | 678 | 4,618,089 | 554 |
| | Home care | 4,159,806 | 499 | 3,773,353 | 452 |
| | Nursing home / sheltered accomodation | 7,661,621 | 919 | 6,806,182 | 816 |
| | Lost productivity associated with illness | . | . | 119,066,938 | 34,175 |
| | Total cost (excl. lost productivity) | 150,618,592 | 18,059 | 135,991,286 | 16,305 |

*Sensitivity analysis 1 / Incident cohort 2011-2015*

| *Brain disorder* | *Cost component* | *Actual costs (EUR)* | *Actual costs per person* | *Attributable costs (EUR)* | *Attributable costs per person* |
|---|---|---|---|---|---|
| Multiple sclerosis | Primary sector | 2,434,817 | 597 | 1,184,244 | 290 |
| | Hospital care | 76,976,497 | 18,866 | 68,814,329 | 16,865 |
| | Filled prescriptions | 1,569,360 | 385 | 699,671 | 171 |
| | Home care | 2,821,710 | 692 | 2,551,724 | 625 |
| | Nursing home / sheltered accomodation | 1,972,382 | 483 | 1,544,839 | 379 |
| | Lost productivity associated with illness | . | . | 32,462,559 | 8,812 |
| | Total cost (excl. lost productivity) | 85,774,765 | 21,022 | 74,794,807 | 18,331 |
| Neuromuscular disorders | Primary sector | 1,760,759 | 557 | 721,167 | 228 |
| | Hospital care | 73,254,387 | 23,194 | 65,296,441 | 20,674 |
| | Filled prescriptions | 2,097,662 | 664 | 1,198,621 | 380 |
| | Home care | 2,978,477 | 943 | 2,146,196 | 680 |
| | Nursing home / sheltered accomodation | 2,672,693 | 846 | 652,864 | 207 |
| | Lost productivity associated with illness | . | . | 15,526,631 | 8,996 |
| | Total cost (excl. lost productivity) | 82,763,979 | 26,205 | 70,015,290 | 22,168 |
| Other neurodegenerative disorders | Primary sector | 1,192,229 | 840 | 649,289 | 457 |
| | Hospital care | 34,000,616 | 23,945 | 29,601,679 | 20,847 |
| | Filled prescriptions | 2,035,186 | 1,433 | 1,504,529 | 1,060 |
| | Home care | 8,256,132 | 5,814 | 7,734,407 | 5,447 |
| | Nursing home / sheltered accomodation | 7,149,726 | 5,035 | 5,854,273 | 4,123 |
| | Lost productivity associated with illness | . | . | 13,998,932 | 24,821 |
| | Total cost (excl. lost productivity) | 52,633,889 | 37,067 | 45,344,176 | 31,933 |
| Parkinson's disease | Primary sector | 6,797,397 | 955 | 3,379,122 | 475 |
| | Hospital care | 106,491,060 | 14,954 | 74,468,088 | 10,457 |
| | Filled prescriptions | 11,603,646 | 1,629 | 7,916,738 | 1,112 |
| | Home care | 30,912,373 | 4,341 | 24,548,892 | 3,447 |
| | Nursing home / sheltered accomodation | 53,847,491 | 7,562 | 36,204,375 | 5,084 |
| | Lost productivity associated with illness | . | . | 10,867,266 | 9,888 |
| | Total cost (excl. lost productivity) | 209,651,968 | 29,441 | 146,517,215 | 20,575 |
| Personality disorders | Primary sector | 9,545,569 | 438 | 3,904,986 | 179 |
| | Hospital care | 446,637,130 | 20,510 | 413,666,858 | 18,996 |
| | Filled prescriptions | 11,972,493 | 550 | 8,791,843 | 404 |
| | Home care | 2,264,370 | 104 | 1,279,220 | 59 |
| | Nursing home / sheltered accomodation | 6,441,433 | 296 | 5,170,930 | 237 |
| | Lost productivity associated with illness | . | . | 495,764,001 | 25,530 |
| | Total cost (excl. lost productivity) | 476,860,997 | 21,898 | 432,813,836 | 19,875 |
| Polyneuropathy | Primary sector | 12,769,046 | 676 | 5,208,213 | 276 |
| | Hospital care | 317,261,896 | 16,796 | 250,719,443 | 13,273 |
| | Filled prescriptions | 18,651,987 | 987 | 10,993,641 | 582 |
| | Home care | 25,758,093 | 1,364 | 15,230,758 | 806 |

***Sensitivity analysis 1 / Incident cohort 2011-2015***

| *Brain disorder* | *Cost component* | *Actual costs (EUR)* | *Actual costs per person* | *Attributable costs (EUR)* | *Attributable costs per person* |
|---|---|---|---|---|---|
| | Nursing home / sheltered accomodation | 22,875,427 | 1,211 | -3,837,710 | -203 |
| | Lost productivity associated with illness | . | . | 165,406,236 | 20,421 |
| | Total cost (excl. lost productivity) | 397,316,449 | 21,035 | 278,314,345 | 14,734 |
| Schizophrenia spectrum disorders | Primary sector | 7,166,569 | 378 | 2,361,166 | 125 |
| | Hospital care | 856,720,642 | 45,175 | 824,370,820 | 43,469 |
| | Filled prescriptions | 14,817,946 | 781 | 11,363,320 | 599 |
| | Home care | 11,054,388 | 583 | 7,324,063 | 386 |
| | Nursing home / sheltered accomodation | 39,060,090 | 2,060 | 28,979,372 | 1,528 |
| | Lost productivity associated with illness | . | . | 438,924,509 | 30,703 |
| | Total cost (excl. lost productivity) | 928,819,634 | 48,977 | 874,398,741 | 46,107 |
| Sleep disorders | Primary sector | 20,699,916 | 499 | 8,418,591 | 203 |
| | Hospital care | 316,560,607 | 7,628 | 220,116,187 | 5,304 |
| | Filled prescriptions | 25,864,544 | 623 | 15,131,504 | 365 |
| | Home care | 13,151,267 | 317 | 6,704,741 | 162 |
| | Nursing home / sheltered accomodation | 10,406,572 | 251 | -1,208,601 | -29 |
| | Lost productivity associated with illness | . | . | 253,321,932 | 8,766 |
| | Total cost (excl. lost productivity) | 386,682,905 | 9,317 | 249,162,422 | 6,004 |
| Stress related disorders | Primary sector | 40,181,998 | 494 | 19,320,599 | 237 |
| | Hospital care | 1,268,153,485 | 15,580 | 1,133,697,776 | 13,928 |
| | Filled prescriptions | 36,442,053 | 448 | 22,372,353 | 275 |
| | Home care | 19,644,954 | 241 | 11,939,881 | 147 |
| | Nursing home / sheltered accomodation | 44,277,134 | 544 | 28,989,774 | 356 |
| | Lost productivity associated with illness | . | . | 1,390,144,263 | 23,151 |
| | Total cost (excl. lost productivity) | 1,408,699,623 | 17,306 | 1,216,320,383 | 14,943 |
| Stroke | Primary sector | 37,626,711 | 664 | 13,182,929 | 233 |
| | Hospital care | 1,975,042,560 | 34,838 | 1,759,528,558 | 31,037 |
| | Filled prescriptions | 47,119,081 | 831 | 21,300,452 | 376 |
| | Home care | 145,036,426 | 2,558 | 98,926,906 | 1,745 |
| | Nursing home / sheltered accomodation | 307,135,751 | 5,418 | 176,341,087 | 3,111 |
| | Lost productivity associated with illness | . | . | 317,767,914 | 17,826 |
| | Total cost (excl. lost productivity) | 2,511,960,528 | 44,309 | 2,069,279,933 | 36,500 |
| Traumatic brain injury | Primary sector | 25,347,301 | 407 | 7,722,594 | 124 |
| | Hospital care | 748,966,854 | 12,012 | 620,444,585 | 9,951 |
| | Filled prescriptions | 21,828,462 | 350 | 7,693,279 | 123 |
| | Home care | 52,719,785 | 846 | 31,439,821 | 504 |
| | Nursing home / sheltered accomodation | 129,284,895 | 2,074 | 69,044,330 | 1,107 |
| | Lost productivity associated with illness | . | . | 253,094,896 | 8,583 |
| | Total cost (excl. lost productivity) | 978,147,296 | 15,688 | 736,344,609 | 11,810 |

***Sensitivity analysis 1 / Incident cohort 2016-2021***

| *Brain disorder* | *Cost component* | *Actual costs (EUR)* | *Actual costs per person* | *Attributable costs (EUR)* | *Attributable costs per person* |
|---|---|---|---|---|---|
| Any brain disorder | Primary sector | 202,819,395 | 457 | 75,832,478 | 171 |
| | Hospital care | 5,438,101,003 | 12,261 | 4,645,771,280 | 10,474 |
| | Filled prescriptions | 195,444,156 | 441 | 97,437,296 | 220 |
| | Home care | 318,322,336 | 718 | 235,316,220 | 531 |
| | Nursing home / sheltered accomodation | 752,588,813 | 1,697 | 580,248,512 | 1,308 |
| | Lost productivity associated with illness | . | . | 3,436,398,908 | 15,801 |
| | Total cost (excl. lost productivity) | 6,907,275,704 | 15,573 | 5,634,605,785 | 12,704 |
| Alcohol abuse | Primary sector | 14,179,533 | 477 | 4,567,053 | 154 |
| | Hospital care | 586,429,282 | 19,728 | 516,157,182 | 17,364 |
| | Filled prescriptions | 17,241,234 | 580 | 8,288,435 | 279 |
| | Home care | 31,210,426 | 1,050 | 25,579,716 | 861 |
| | Nursing home / sheltered accomodation | 55,981,612 | 1,883 | 43,121,539 | 1,451 |
| | Lost productivity associated with illness | . | . | 684,985,783 | 36,976 |
| | Total cost (excl. lost productivity) | 705,042,087 | 23,718 | 597,713,924 | 20,108 |
| Anxiety disorders | Primary sector | 27,802,940 | 504 | 13,082,256 | 237 |
| | Hospital care | 710,729,474 | 12,880 | 619,028,605 | 11,218 |
| | Filled prescriptions | 27,419,172 | 497 | 17,184,589 | 311 |
| | Home care | 25,231,780 | 457 | 17,879,553 | 324 |
| | Nursing home / sheltered accomodation | 48,823,904 | 885 | 27,544,531 | 499 |
| | Lost productivity associated with illness | . | . | 843,225,955 | 22,972 |
| | Total cost (excl. lost productivity) | 840,007,271 | 15,223 | 694,719,534 | 12,590 |
| Bipolar disorder | Primary sector | 4,505,389 | 466 | 1,708,604 | 177 |
| | Hospital care | 207,023,719 | 21,422 | 189,105,348 | 19,568 |
| | Filled prescriptions | 5,622,801 | 582 | 3,629,079 | 376 |
| | Home care | 3,108,499 | 322 | 2,075,982 | 215 |
| | Nursing home / sheltered accomodation | 9,835,862 | 1,018 | 7,402,189 | 766 |
| | Lost productivity associated with illness | . | . | 209,403,650 | 25,855 |
| | Total cost (excl. lost productivity) | 230,096,270 | 23,810 | 203,921,202 | 21,101 |
| Brain tumors | Primary sector | 6,619,549 | 581 | 2,149,303 | 189 |
| | Hospital care | 472,356,911 | 41,443 | 439,391,085 | 38,550 |
| | Filled prescriptions | 7,621,185 | 669 | 3,258,708 | 286 |
| | Home care | 13,442,208 | 1,179 | 9,245,014 | 811 |
| | Nursing home / sheltered accomodation | 17,586,335 | 1,543 | 5,787,352 | 508 |

*Sensitivity analysis 1 / Incident cohort 2016-2021*

| *Brain disorder* | *Cost component* | *Actual costs (EUR)* | *Actual costs per person* | *Attributable costs (EUR)* | *Attributable costs per person* |
|---|---|---|---|---|---|
| | Lost productivity associated with illness | . | . | 28,162,864 | 5,665 |
| | Total cost (excl. lost productivity) | 517,626,188 | 45,414 | 459,831,462 | 40,344 |
| Cerebral palsy | Primary sector | 1,297,043 | 636 | 753,711 | 369 |
| | Hospital care | 26,925,179 | 13,199 | 23,747,177 | 11,641 |
| | Filled prescriptions | 1,487,241 | 729 | 1,145,156 | 561 |
| | Home care | 3,320,685 | 1,628 | 3,073,098 | 1,506 |
| | Nursing home / sheltered accomodation | 6,258,268 | 3,068 | 5,681,115 | 2,785 |
| | Lost productivity associated with illness | . | . | 19,611,702 | 36,318 |
| | Total cost (excl. lost productivity) | 39,288,417 | 19,260 | 34,400,257 | 16,863 |
| Dementia | Primary sector | 28,065,234 | 652 | 4,809,100 | 112 |
| | Hospital care | 427,912,047 | 9,944 | 234,476,080 | 5,449 |
| | Filled prescriptions | 40,242,830 | 935 | 13,398,016 | 311 |
| | Home care | 187,479,461 | 4,357 | 131,372,128 | 3,053 |
| | Nursing home / sheltered accomodation | 698,974,899 | 16,244 | 562,863,985 | 13,080 |
| | Lost productivity associated with illness | . | . | 85,581,771 | 41,504 |
| | Total cost (excl. lost productivity) | 1,382,674,470 | 32,132 | 946,919,310 | 22,006 |
| Depression | Primary sector | 39,325,570 | 558 | 17,710,317 | 251 |
| | Hospital care | 1,114,754,467 | 15,817 | 974,917,725 | 13,833 |
| | Filled prescriptions | 38,662,587 | 549 | 21,536,108 | 306 |
| | Home care | 60,353,210 | 856 | 42,883,327 | 608 |
| | Nursing home / sheltered accomodation | 147,801,337 | 2,097 | 92,969,447 | 1,319 |
| | Lost productivity associated with illness | . | . | 1,183,204,802 | 25,002 |
| | Total cost (excl. lost productivity) | 1,400,897,171 | 19,878 | 1,150,016,924 | 16,318 |
| Developmental and behavioural disorders | Primary sector | 16,865,766 | 253 | 6,346,701 | 95 |
| | Hospital care | 563,622,365 | 8,443 | 509,541,088 | 7,633 |
| | Filled prescriptions | 26,649,824 | 399 | 21,773,311 | 326 |
| | Home care | 2,437,363 | 37 | 1,590,142 | 24 |
| | Nursing home / sheltered accomodation | 8,433,770 | 126 | 6,949,465 | 104 |
| | Lost productivity associated with illness | . | . | 559,606,470 | 26,818 |
| | Total cost (excl. lost productivity) | 618,009,088 | 9,258 | 546,200,706 | 8,182 |
| Drug abuse | Primary sector | 7,495,848 | 366 | 3,136,698 | 153 |
| | Hospital care | 372,702,940 | 18,178 | 345,895,246 | 16,871 |
| | Filled prescriptions | 9,196,469 | 449 | 6,378,121 | 311 |

***Sensitivity analysis 1 / Incident cohort 2016-2021***

| *Brain disorder* | *Cost component* | *Actual costs (EUR)* | *Actual costs per person* | *Attributable costs (EUR)* | *Attributable costs per person* |
|---|---|---|---|---|---|
| | Home care | 6,268,005 | 306 | 5,004,558 | 244 |
| | Nursing home / sheltered accomodation | 9,032,206 | 441 | 6,153,600 | 300 |
| | Lost productivity associated with illness | . | . | 493,583,949 | 28,922 |
| | Total cost (excl. lost productivity) | 404,695,468 | 19,739 | 366,568,224 | 17,879 |
| Eating disorders | Primary sector | 2,093,952 | 304 | 528,475 | 77 |
| | Hospital care | 212,916,135 | 30,893 | 204,351,898 | 29,650 |
| | Filled prescriptions | 1,136,640 | 165 | 359,080 | 52 |
| | Home care | 156,002 | 23 | 94,195 | 14 |
| | Nursing home / sheltered accomodation | 66,796 | 10 | 30,619 | 4 |
| | Lost productivity associated with illness | . | . | 28,913,795 | 7,595 |
| | Total cost (excl. lost productivity) | 216,369,525 | 31,394 | 205,364,267 | 29,797 |
| Epilepsy | Primary sector | 10,760,119 | 523 | 4,092,998 | 199 |
| | Hospital care | 387,367,955 | 18,839 | 339,508,822 | 16,511 |
| | Filled prescriptions | 17,019,350 | 828 | 10,977,862 | 534 |
| | Home care | 35,132,939 | 1,709 | 28,775,364 | 1,399 |
| | Nursing home / sheltered accomodation | 99,953,751 | 4,861 | 81,116,665 | 3,945 |
| | Lost productivity associated with illness | . | . | 166,840,922 | 22,084 |
| | Total cost (excl. lost productivity) | 550,234,115 | 26,760 | 464,471,711 | 22,589 |
| Headache | Primary sector | 23,390,049 | 466 | 9,606,632 | 191 |
| | Hospital care | 268,015,062 | 5,337 | 182,688,626 | 3,638 |
| | Filled prescriptions | 15,840,205 | 315 | 6,222,434 | 124 |
| | Home care | 5,283,893 | 105 | 870,757 | 17 |
| | Nursing home / sheltered accomodation | 7,372,170 | 147 | -3,486,202 | -69 |
| | Lost productivity associated with illness | . | . | 220,445,248 | 6,058 |
| | Total cost (excl. lost productivity) | 319,901,380 | 6,370 | 195,902,248 | 3,901 |
| Infections of the CNS | Primary sector | 5,310,168 | 412 | 1,486,209 | 115 |
| | Hospital care | 268,299,498 | 20,812 | 243,026,377 | 18,852 |
| | Filled prescriptions | 4,955,136 | 384 | 1,887,410 | 146 |
| | Home care | 6,592,363 | 511 | 4,384,010 | 340 |
| | Nursing home / sheltered accomodation | 11,359,994 | 881 | 5,621,366 | 436 |
| | Lost productivity associated with illness | . | . | -8,026,967 | -1,179 |
| | Total cost (excl. lost productivity) | 296,517,160 | 23,001 | 256,405,371 | 19,890 |
| Intellectual disability | Primary sector | 2,671,418 | 352 | 1,187,835 | 157 |
| | Hospital care | 98,695,951 | 13,016 | 90,091,305 | 11,881 |
| | Filled prescriptions | 3,999,727 | 527 | 3,169,895 | 418 |
| | Home care | 2,656,661 | 350 | 2,388,854 | 315 |
| | Nursing home / sheltered accomodation | 6,166,896 | 813 | 5,593,717 | 738 |
| | Lost productivity associated with illness | . | . | 117,271,912 | 37,288 |
| | Total cost (excl. lost productivity) | 114,190,653 | 15,060 | 102,431,605 | 13,509 |

*Sensitivity analysis 1 / Incident cohort 2016-2021*

| *Brain disorder* | *Cost component* | *Actual costs (EUR)* | *Actual costs per person* | *Attributable costs (EUR)* | *Attributable costs per person* |
|---|---|---|---|---|---|
| Multiple sclerosis | Primary sector | 2,785,855 | 591 | 1,403,061 | 298 |
| | Hospital care | 58,963,034 | 12,508 | 50,423,721 | 10,697 |
| | Filled prescriptions | 1,423,027 | 302 | 460,620 | 98 |
| | Home care | 2,918,357 | 619 | 2,656,066 | 563 |
| | Nursing home / sheltered accomodation | 1,607,313 | 341 | 1,056,916 | 224 |
| | Lost productivity associated with illness | . | . | 33,746,491 | 7,946 |
| | Total cost (excl. lost productivity) | 67,697,585 | 14,361 | 56,000,384 | 11,880 |
| Neuromuscular disorders | Primary sector | 1,828,515 | 572 | 746,728 | 234 |
| | Hospital care | 52,102,956 | 16,308 | 44,324,046 | 13,873 |
| | Filled prescriptions | 2,230,134 | 698 | 1,245,115 | 390 |
| | Home care | 3,195,135 | 1,000 | 2,438,453 | 763 |
| | Nursing home / sheltered accomodation | 2,751,462 | 861 | 768,070 | 240 |
| | Lost productivity associated with illness | . | . | 21,601,672 | 12,684 |
| | Total cost (excl. lost productivity) | 62,108,203 | 19,439 | 49,522,412 | 15,500 |
| Other neurodegenerative disorders | Primary sector | 1,604,733 | 854 | 863,433 | 460 |
| | Hospital care | 37,978,494 | 20,213 | 32,571,669 | 17,336 |
| | Filled prescriptions | 2,106,153 | 1,121 | 1,343,471 | 715 |
| | Home care | 8,097,406 | 4,310 | 7,534,059 | 4,010 |
| | Nursing home / sheltered accomodation | 6,200,714 | 3,300 | 4,598,224 | 2,447 |
| | Lost productivity associated with illness | . | . | 19,179,622 | 27,676 |
| | Total cost (excl. lost productivity) | 55,987,500 | 29,798 | 46,910,856 | 24,967 |
| Parkinson’s disease | Primary sector | 8,928,321 | 1,047 | 4,780,154 | 560 |
| | Hospital care | 100,005,288 | 11,722 | 65,602,212 | 7,689 |
| | Filled prescriptions | 10,429,685 | 1,222 | 5,896,822 | 691 |
| | Home care | 30,484,116 | 3,573 | 24,453,221 | 2,866 |
| | Nursing home / sheltered accomodation | 61,009,820 | 7,151 | 43,103,009 | 5,052 |
| | Lost productivity associated with illness | . | . | 18,188,206 | 14,848 |
| | Total cost (excl. lost productivity) | 210,857,230 | 24,715 | 143,835,417 | 16,859 |
| Personality disorders | Primary sector | 7,981,971 | 408 | 2,955,356 | 151 |
| | Hospital care | 316,134,945 | 16,142 | 288,749,568 | 14,744 |
| | Filled prescriptions | 6,360,619 | 325 | 3,740,994 | 191 |
| | Home care | 1,257,176 | 64 | 665,197 | 34 |
| | Nursing home / sheltered accomodation | 3,192,780 | 163 | 2,272,705 | 116 |
| | Lost productivity associated with illness | . | . | 447,155,638 | 25,537 |
| | Total cost (excl. lost productivity) | 334,927,491 | 17,101 | 298,383,818 | 15,235 |
| Polyneuropathy | Primary sector | 15,838,848 | 664 | 6,071,306 | 255 |
| | Hospital care | 339,716,381 | 14,250 | 262,684,035 | 11,018 |
| | Filled prescriptions | 27,308,714 | 1,145 | 17,063,302 | 716 |
| | Home care | 29,054,869 | 1,219 | 18,211,131 | 764 |

***Sensitivity analysis 1 / Incident cohort 2016-2021***

| *Brain disorder* | *Cost component* | *Actual costs (EUR)* | *Actual costs per person* | *Attributable costs (EUR)* | *Attributable costs per person* |
|---|---|---|---|---|---|
| | Nursing home / sheltered accomodation | 27,230,000 | 1,142 | -4,801,372 | -201 |
| | Lost productivity associated with illness | . | . | 213,994,223 | 23,265 |
| | Total cost (excl. lost productivity) | 439,148,812 | 18,420 | 299,228,402 | 12,551 |
| Schizophrenia spectrum disorders | Primary sector | 7,896,094 | 342 | 2,342,770 | 102 |
| | Hospital care | 738,618,574 | 32,004 | 705,734,471 | 30,579 |
| | Filled prescriptions | 9,096,656 | 394 | 5,339,786 | 231 |
| | Home care | 8,568,347 | 371 | 6,093,712 | 264 |
| | Nursing home / sheltered accomodation | 31,937,880 | 1,384 | 24,611,250 | 1,066 |
| | Lost productivity associated with illness | . | . | 546,581,670 | 31,308 |
| | Total cost (excl. lost productivity) | 796,117,552 | 34,495 | 744,121,989 | 32,242 |
| Sleep disorders | Primary sector | 29,662,657 | 486 | 11,469,913 | 188 |
| | Hospital care | 433,714,614 | 7,104 | 302,165,544 | 4,949 |
| | Filled prescriptions | 36,903,473 | 604 | 20,777,545 | 340 |
| | Home care | 19,967,649 | 327 | 10,719,669 | 176 |
| | Nursing home / sheltered accomodation | 17,071,079 | 280 | -4,454,280 | -73 |
| | Lost productivity associated with illness | . | . | 512,291,974 | 12,258 |
| | Total cost (excl. lost productivity) | 537,319,471 | 8,801 | 340,678,391 | 5,580 |
| Stress related disorders | Primary sector | 44,795,231 | 472 | 21,607,665 | 228 |
| | Hospital care | 1,130,404,785 | 11,902 | 997,159,214 | 10,499 |
| | Filled prescriptions | 32,112,617 | 338 | 16,670,047 | 176 |
| | Home care | 17,577,057 | 185 | 10,945,705 | 115 |
| | Nursing home / sheltered accomodation | 42,952,550 | 452 | 27,057,155 | 285 |
| | Lost productivity associated with illness | . | . | 1,609,346,724 | 24,069 |
| | Total cost (excl. lost productivity) | 1,267,842,241 | 13,349 | 1,073,439,785 | 11,303 |
| Stroke | Primary sector | 47,943,785 | 664 | 16,041,842 | 222 |
| | Hospital care | 2,278,022,053 | 31,559 | 2,025,467,612 | 28,060 |
| | Filled prescriptions | 61,869,551 | 857 | 27,466,642 | 381 |
| | Home care | 146,375,480 | 2,028 | 99,564,232 | 1,379 |
| | Nursing home / sheltered accomodation | 351,525,262 | 4,870 | 198,121,613 | 2,745 |
| | Lost productivity associated with illness | . | . | 397,787,289 | 19,305 |
| | Total cost (excl. lost productivity) | 2,885,736,132 | 39,978 | 2,366,661,941 | 32,787 |
| Traumatic brain injury | Primary sector | 32,415,021 | 439 | 10,223,489 | 138 |
| | Hospital care | 817,072,902 | 11,062 | 664,692,995 | 8,999 |
| | Filled prescriptions | 30,059,574 | 407 | 11,194,759 | 152 |
| | Home care | 66,356,898 | 898 | 40,531,444 | 549 |
| | Nursing home / sheltered accomodation | 202,398,081 | 2,740 | 118,517,415 | 1,605 |
| | Lost productivity associated with illness | . | . | 228,679,715 | 6,885 |
| | Total cost (excl. lost productivity) | 1,148,302,476 | 15,546 | 845,160,101 | 11,442 |

***Sensitivity analysis 1 / Prevalent cohort 2015***

| *Brain disorder* | *Cost component* | *Actual costs (EUR)* | *Actual costs per person* | *Attributable costs (EUR)* | *Attributable costs per person* |
|---|---|---|---|---|---|
| Any brain disorder | Primary sector | 448,456,035 | 414 | 140,252,169 | 130 |
| | Hospital care | 4,808,174,453 | 4,443 | 2,835,931,776 | 2,621 |
| | Filled prescriptions | 542,264,455 | 501 | 325,819,552 | 301 |
| | Home care | 735,597,282 | 680 | 557,508,761 | 515 |
| | Nursing home / sheltered accomodation | 2,385,698,182 | 2,205 | 2,055,681,659 | 1,900 |
| | Lost productivity associated with illness | . | . | 12,434,546,903 | 17,609 |
| | Total cost (excl. lost productivity) | 8,920,190,407 | 8,243 | 5,915,193,916 | 5,466 |
| Alcohol abuse | Primary sector | 42,141,763 | 426 | 9,521,213 | 96 |
| | Hospital care | 817,168,001 | 8,261 | 569,837,862 | 5,760 |
| | Filled prescriptions | 70,638,983 | 714 | 41,703,952 | 422 |
| | Home care | 90,987,785 | 920 | 73,537,699 | 743 |
| | Nursing home / sheltered accomodation | 296,805,638 | 3,000 | 258,365,948 | 2,612 |
| | Lost productivity associated with illness | . | . | 2,492,580,115 | 35,993 |
| | Total cost (excl. lost productivity) | 1,317,742,170 | 13,321 | 952,966,675 | 9,633 |
| Anxiety disorders | Primary sector | 45,843,298 | 474 | 16,842,581 | 174 |
| | Hospital care | 579,812,553 | 5,989 | 389,263,846 | 4,021 |
| | Filled prescriptions | 58,882,811 | 608 | 38,307,948 | 396 |
| | Home care | 34,419,489 | 356 | 21,916,164 | 226 |
| | Nursing home / sheltered accomodation | 97,514,846 | 1,007 | 66,728,142 | 689 |
| | Lost productivity associated with illness | . | . | 1,776,566,819 | 22,248 |
| | Total cost (excl. lost productivity) | 816,472,997 | 8,433 | 533,058,682 | 5,506 |
| Bipolar disorder | Primary sector | 12,494,760 | 538 | 4,475,058 | 193 |
| | Hospital care | 254,925,156 | 10,973 | 197,930,511 | 8,520 |
| | Filled prescriptions | 23,323,541 | 1,004 | 16,571,244 | 713 |
| | Home care | 18,766,249 | 808 | 13,255,737 | 571 |
| | Nursing home / sheltered accomodation | 72,119,434 | 3,104 | 58,674,855 | 2,526 |
| | Lost productivity associated with illness | . | . | 547,038,680 | 32,128 |
| | Total cost (excl. lost productivity) | 381,629,140 | 16,427 | 290,907,405 | 12,522 |
| Brain tumors | Primary sector | 7,757,673 | 526 | 2,015,312 | 137 |
| | Hospital care | 118,501,141 | 8,032 | 73,289,553 | 4,967 |
| | Filled prescriptions | 9,030,374 | 612 | 3,547,087 | 240 |
| | Home care | 17,335,029 | 1,175 | 11,053,545 | 749 |
| | Nursing home / sheltered accomodation | 32,913,778 | 2,231 | 15,406,080 | 1,044 |

***Sensitivity analysis 1 / Prevalent cohort 2015***

| *Brain disorder* | *Cost component* | *Actual costs (EUR)* | *Actual costs per person* | *Attributable costs (EUR)* | *Attributable costs per person* |
|---|---|---|---|---|---|
| | Lost productivity associated with illness | . | . | 72,075,182 | 9,732 |
| | Total cost (excl. lost productivity) | 185,537,995 | 12,575 | 105,311,577 | 7,138 |
| Cerebral palsy | Primary sector | 6,384,551 | 691 | 4,454,678 | 482 |
| | Hospital care | 46,844,503 | 5,073 | 33,590,212 | 3,638 |
| | Filled prescriptions | 6,712,756 | 727 | 5,494,374 | 595 |
| | Home care | 14,876,674 | 1,611 | 14,502,854 | 1,571 |
| | Nursing home / sheltered accomodation | 11,975,865 | 1,297 | 11,025,560 | 1,194 |
| | Lost productivity associated with illness | . | . | 145,308,016 | 27,885 |
| | Total cost (excl. lost productivity) | 86,794,348 | 9,400 | 69,067,678 | 7,480 |
| Dementia | Primary sector | 20,927,844 | 571 | 1,734,414 | 47 |
| | Hospital care | 217,433,694 | 5,935 | 48,260,140 | 1,317 |
| | Filled prescriptions | 36,063,123 | 984 | 14,663,983 | 400 |
| | Home care | 156,559,454 | 4,274 | 91,651,499 | 2,502 |
| | Nursing home / sheltered accomodation | 1,285,358,309 | 35,086 | 1,135,287,102 | 30,990 |
| | Lost productivity associated with illness | . | . | 133,786,524 | 39,130 |
| | Total cost (excl. lost productivity) | 1,716,342,424 | 46,851 | 1,291,597,139 | 35,256 |
| Depression | Primary sector | 90,820,646 | 507 | 30,436,635 | 170 |
| | Hospital care | 1,162,793,869 | 6,485 | 746,736,923 | 4,165 |
| | Filled prescriptions | 123,953,347 | 691 | 75,891,707 | 423 |
| | Home care | 154,685,378 | 863 | 98,926,557 | 552 |
| | Nursing home / sheltered accomodation | 575,922,436 | 3,212 | 424,854,561 | 2,370 |
| | Lost productivity associated with illness | . | . | 3,373,129,112 | 25,422 |
| | Total cost (excl. lost productivity) | 2,108,175,676 | 11,758 | 1,376,846,382 | 7,679 |
| Developmental and behavioural disorders | Primary sector | 26,506,527 | 253 | 9,103,490 | 87 |
| | Hospital care | 422,387,304 | 4,036 | 316,607,313 | 3,025 |
| | Filled prescriptions | 54,484,190 | 521 | 45,962,884 | 439 |
| | Home care | 5,910,921 | 56 | 4,479,864 | 43 |
| | Nursing home / sheltered accomodation | 9,854,977 | 94 | 7,695,190 | 74 |
| | Lost productivity associated with illness | . | . | 985,130,268 | 16,634 |
| | Total cost (excl. lost productivity) | 519,143,918 | 4,961 | 383,848,741 | 3,668 |
| Drug abuse | Primary sector | 18,328,996 | 369 | 5,781,175 | 116 |
| | Hospital care | 478,724,279 | 9,628 | 396,584,806 | 7,976 |
| | Filled prescriptions | 39,623,932 | 797 | 30,731,875 | 618 |

*Sensitivity analysis 1 / Prevalent cohort 2015*

| *Brain disorder* | *Cost component* | *Actual costs (EUR)* | *Actual costs per person* | *Attributable costs (EUR)* | *Attributable costs per person* |
|---|---|---|---|---|---|
| | Home care | 22,399,478 | 450 | 17,811,701 | 358 |
| | Nursing home / sheltered accomodation | 57,372,782 | 1,154 | 46,606,664 | 937 |
| | Lost productivity associated with illness | . | . | 1,556,718,369 | 34,847 |
| | Total cost (excl. lost productivity) | 616,449,467 | 12,398 | 497,516,220 | 10,006 |
| Eating disorders | Primary sector | 6,849,665 | 427 | 2,044,755 | 127 |
| | Hospital care | 141,375,424 | 8,813 | 112,480,191 | 7,012 |
| | Filled prescriptions | 5,134,371 | 320 | 2,668,374 | 166 |
| | Home care | 1,075,080 | 67 | 848,809 | 53 |
| | Nursing home / sheltered accomodation | 1,647,012 | 103 | 1,405,236 | 88 |
| | Lost productivity associated with illness | . | . | 95,944,937 | 6,606 |
| | Total cost (excl. lost productivity) | 156,081,552 | 9,730 | 119,447,365 | 7,446 |
| Epilepsy | Primary sector | 31,967,438 | 441 | 10,405,193 | 144 |
| | Hospital care | 414,148,572 | 5,718 | 254,853,362 | 3,518 |
| | Filled prescriptions | 57,623,808 | 796 | 40,260,686 | 556 |
| | Home care | 75,564,134 | 1,043 | 60,279,350 | 832 |
| | Nursing home / sheltered accomodation | 207,973,683 | 2,871 | 166,402,529 | 2,297 |
| | Lost productivity associated with illness | . | . | 885,587,468 | 19,216 |
| | Total cost (excl. lost productivity) | 787,277,635 | 10,869 | 532,201,119 | 7,347 |
| Headache | Primary sector | 45,187,929 | 455 | 14,300,527 | 144 |
| | Hospital care | 362,685,642 | 3,653 | 152,544,561 | 1,536 |
| | Filled prescriptions | 42,239,500 | 425 | 18,915,833 | 191 |
| | Home care | 19,566,443 | 197 | 5,663,250 | 57 |
| | Nursing home / sheltered accomodation | 40,451,679 | 407 | 6,934,537 | 70 |
| | Lost productivity associated with illness | . | . | 538,243,877 | 6,951 |
| | Total cost (excl. lost productivity) | 510,131,193 | 5,138 | 198,358,709 | 1,998 |
| Infections of the CNS | Primary sector | 9,852,370 | 367 | 2,010,928 | 75 |
| | Hospital care | 103,987,010 | 3,878 | 46,391,086 | 1,730 |
| | Filled prescriptions | 9,764,459 | 364 | 3,348,031 | 125 |
| | Home care | 13,435,294 | 501 | 8,049,559 | 300 |
| | Nursing home / sheltered accomodation | 29,885,259 | 1,115 | 16,071,785 | 599 |
| | Lost productivity associated with illness | . | . | 34,716,969 | 2,101 |
| | Total cost (excl. lost productivity) | 166,924,392 | 6,225 | 75,871,389 | 2,830 |
| Intellectual disability | Primary sector | 11,344,833 | 410 | 5,183,540 | 187 |
| | Hospital care | 161,192,376 | 5,827 | 119,253,145 | 4,311 |
| | Filled prescriptions | 21,750,057 | 786 | 17,593,509 | 636 |
| | Home care | 14,661,044 | 530 | 13,156,529 | 476 |
| | Nursing home / sheltered accomodation | 31,524,079 | 1,140 | 28,369,156 | 1,025 |
| | Lost productivity associated with illness | . | . | 610,349,050 | 34,081 |
| | Total cost (excl. lost productivity) | 240,472,390 | 8,693 | 183,555,879 | 6,635 |

*Sensitivity analysis 1 / Prevalent cohort 2015*

| *Brain disorder* | *Cost component* | *Actual costs (EUR)* | *Actual costs per person* | *Attributable costs (EUR)* | *Attributable costs per person* |
|---|---|---|---|---|---|
| Multiple sclerosis | Primary sector | 12,096,775 | 813 | 6,926,721 | 466 |
| | Hospital care | 106,970,567 | 7,192 | 70,569,113 | 4,744 |
| | Filled prescriptions | 9,912,316 | 666 | 5,638,864 | 379 |
| | Home care | 54,459,504 | 3,661 | 52,280,198 | 3,515 |
| | Nursing home / sheltered accomodation | 34,795,208 | 2,339 | 30,102,266 | 2,024 |
| | Lost productivity associated with illness | . | . | 221,303,492 | 18,942 |
| | Total cost (excl. lost productivity) | 218,234,370 | 14,672 | 165,517,162 | 11,128 |
| Neuromuscular disorders | Primary sector | 4,258,791 | 585 | 1,888,279 | 259 |
| | Hospital care | 52,907,446 | 7,264 | 34,985,732 | 4,803 |
| | Filled prescriptions | 4,561,800 | 626 | 2,504,524 | 344 |
| | Home care | 9,557,772 | 1,312 | 7,641,399 | 1,049 |
| | Nursing home / sheltered accomodation | 9,793,012 | 1,344 | 4,778,770 | 656 |
| | Lost productivity associated with illness | . | . | 67,916,449 | 15,157 |
| | Total cost (excl. lost productivity) | 81,078,821 | 11,131 | 51,798,705 | 7,111 |
| Other neurodegenerative disorders | Primary sector | 1,783,261 | 805 | 976,422 | 441 |
| | Hospital care | 19,616,092 | 8,853 | 12,971,523 | 5,854 |
| | Filled prescriptions | 2,096,424 | 946 | 1,342,625 | 606 |
| | Home care | 10,838,340 | 4,891 | 10,162,141 | 4,586 |
| | Nursing home / sheltered accomodation | 18,277,824 | 8,249 | 16,657,982 | 7,518 |
| | Lost productivity associated with illness | . | . | 37,795,846 | 32,332 |
| | Total cost (excl. lost productivity) | 52,611,943 | 23,744 | 42,110,693 | 19,005 |
| Parkinson’s disease | Primary sector | 9,292,070 | 989 | 4,710,488 | 501 |
| | Hospital care | 79,717,761 | 8,484 | 38,529,809 | 4,101 |
| | Filled prescriptions | 29,253,611 | 3,113 | 24,414,507 | 2,598 |
| | Home care | 50,079,377 | 5,330 | 41,647,093 | 4,432 |
| | Nursing home / sheltered accomodation | 137,170,851 | 14,599 | 112,350,495 | 11,957 |
| | Lost productivity associated with illness | . | . | 33,256,111 | 21,581 |
| | Total cost (excl. lost productivity) | 305,513,670 | 32,516 | 221,652,391 | 23,590 |
| Personality disorders | Primary sector | 33,070,646 | 463 | 11,882,689 | 166 |
| | Hospital care | 545,135,869 | 7,636 | 413,207,997 | 5,788 |
| | Filled prescriptions | 48,098,687 | 674 | 34,031,908 | 477 |
| | Home care | 19,128,288 | 268 | 13,895,600 | 195 |
| | Nursing home / sheltered accomodation | 51,390,311 | 720 | 41,701,088 | 584 |
| | Lost productivity associated with illness | . | . | 1,949,975,843 | 29,862 |
| | Total cost (excl. lost productivity) | 696,823,800 | 9,761 | 514,719,282 | 7,210 |
| Polyneuropathy | Primary sector | 20,445,880 | 613 | 6,749,603 | 202 |
| | Hospital care | 282,123,746 | 8,458 | 167,345,797 | 5,017 |
| | Filled prescriptions | 31,450,051 | 943 | 17,727,836 | 531 |
| | Home care | 56,400,109 | 1,691 | 37,262,313 | 1,117 |

***Sensitivity analysis 1 / Prevalent cohort 2015***

| *Brain disorder* | *Cost component* | *Actual costs (EUR)* | *Actual costs per person* | *Attributable costs (EUR)* | *Attributable costs per person* |
|---|---|---|---|---|---|
| | Nursing home / sheltered accomodation | 97,916,044 | 2,936 | 41,703,174 | 1,250 |
| | Lost productivity associated with illness | . | . | 273,079,524 | 19,053 |
| | Total cost (excl. lost productivity) | 488,335,829 | 14,641 | 270,788,723 | 8,118 |
| Schizophrenia spectrum disorders | Primary sector | 24,982,333 | 411 | 7,253,027 | 119 |
| | Hospital care | 788,327,352 | 12,982 | 670,576,588 | 11,043 |
| | Filled prescriptions | 60,694,950 | 1,000 | 47,138,950 | 776 |
| | Home care | 42,530,160 | 700 | 33,410,079 | 550 |
| | Nursing home / sheltered accomodation | 157,124,464 | 2,588 | 134,872,262 | 2,221 |
| | Lost productivity associated with illness | . | . | 1,974,644,879 | 39,393 |
| | Total cost (excl. lost productivity) | 1,073,659,259 | 17,681 | 893,250,907 | 14,710 |
| Sleep disorders | Primary sector | 36,140,580 | 484 | 12,699,357 | 170 |
| | Hospital care | 341,271,539 | 4,572 | 148,798,868 | 1,994 |
| | Filled prescriptions | 48,553,041 | 650 | 27,269,000 | 365 |
| | Home care | 23,126,389 | 310 | 9,343,990 | 125 |
| | Nursing home / sheltered accomodation | 34,367,123 | 460 | 3,088,132 | 41 |
| | Lost productivity associated with illness | . | . | 424,823,919 | 9,041 |
| | Total cost (excl. lost productivity) | 483,458,672 | 6,477 | 201,199,347 | 2,696 |
| Stress related disorders | Primary sector | 84,634,253 | 430 | 27,754,131 | 141 |
| | Hospital care | 1,100,397,240 | 5,597 | 738,857,247 | 3,758 |
| | Filled prescriptions | 95,197,887 | 484 | 55,193,558 | 281 |
| | Home care | 54,515,050 | 277 | 34,997,101 | 178 |
| | Nursing home / sheltered accomodation | 145,113,941 | 738 | 103,561,313 | 527 |
| | Lost productivity associated with illness | . | . | 3,673,553,783 | 22,402 |
| | Total cost (excl. lost productivity) | 1,479,858,371 | 7,527 | 960,363,351 | 4,885 |
| Stroke | Primary sector | 70,179,238 | 606 | 18,984,472 | 164 |
| | Hospital care | 780,767,736 | 6,741 | 346,000,200 | 2,987 |
| | Filled prescriptions | 92,869,339 | 802 | 40,159,380 | 347 |
| | Home care | 275,192,054 | 2,376 | 187,437,973 | 1,618 |
| | Nursing home / sheltered accomodation | 791,410,704 | 6,833 | 526,429,523 | 4,545 |
| | Lost productivity associated with illness | . | . | 703,911,939 | 19,601 |
| | Total cost (excl. lost productivity) | 2,010,419,071 | 17,358 | 1,119,011,548 | 9,662 |
| Traumatic brain injury | Primary sector | 79,123,822 | 319 | 14,479,315 | 58 |
| | Hospital care | 785,847,190 | 3,164 | 326,573,043 | 1,315 |
| | Filled prescriptions | 75,622,334 | 304 | 26,433,222 | 106 |
| | Home care | 97,360,991 | 392 | 53,752,879 | 216 |
| | Nursing home / sheltered accomodation | 303,522,869 | 1,222 | 184,449,292 | 743 |
| | Lost productivity associated with illness | . | . | 1,424,052,807 | 8,256 |
| | Total cost (excl. lost productivity) | 1,341,477,205 | 5,401 | 605,687,752 | 2,438 |

***Sensitivity analysis 1 / Prevalent cohort 2016***

| *Brain disorder* | *Cost component* | *Actual costs (EUR)* | *Actual costs per person* | *Attributable costs (EUR)* | *Attributable costs per person* |
|---|---|---|---|---|---|
| Any brain disorder | Primary sector | 449,716,546 | 404 | 141,071,688 | 127 |
| | Hospital care | 4,846,869,930 | 4,352 | 2,848,955,745 | 2,558 |
| | Filled prescriptions | 537,502,002 | 483 | 316,717,549 | 284 |
| | Home care | 653,814,206 | 587 | 498,277,489 | 447 |
| | Nursing home / sheltered accomodation | 2,404,030,620 | 2,159 | 2,069,199,266 | 1,858 |
| | Lost productivity associated with illness | . | . | 12,575,037,988 | 17,322 |
| | Total cost (excl. lost productivity) | 8,891,933,304 | 7,984 | 5,874,221,736 | 5,275 |
| Alcohol abuse | Primary sector | 40,948,710 | 413 | 8,909,454 | 90 |
| | Hospital care | 814,623,637 | 8,220 | 567,529,883 | 5,727 |
| | Filled prescriptions | 68,599,014 | 692 | 39,459,336 | 398 |
| | Home care | 75,862,765 | 765 | 59,585,000 | 601 |
| | Nursing home / sheltered accomodation | 306,234,211 | 3,090 | 267,086,654 | 2,695 |
| | Lost productivity associated with illness | . | . | 2,456,380,142 | 36,034 |
| | Total cost (excl. lost productivity) | 1,306,268,337 | 13,181 | 942,570,328 | 9,511 |
| Anxiety disorders | Primary sector | 47,565,428 | 455 | 17,465,298 | 167 |
| | Hospital care | 601,915,096 | 5,757 | 402,561,312 | 3,850 |
| | Filled prescriptions | 56,808,649 | 543 | 35,405,432 | 339 |
| | Home care | 31,860,683 | 305 | 20,200,321 | 193 |
| | Nursing home / sheltered accomodation | 103,099,690 | 986 | 69,539,775 | 665 |
| | Lost productivity associated with illness | . | . | 1,930,989,894 | 22,471 |
| | Total cost (excl. lost productivity) | 841,249,545 | 8,046 | 545,172,137 | 5,214 |
| Bipolar disorder | Primary sector | 12,532,769 | 517 | 4,456,832 | 184 |
| | Hospital care | 256,143,830 | 10,558 | 197,748,917 | 8,151 |
| | Filled prescriptions | 21,175,765 | 873 | 14,353,399 | 592 |
| | Home care | 15,494,002 | 639 | 10,902,020 | 449 |
| | Nursing home / sheltered accomodation | 68,965,303 | 2,843 | 54,993,964 | 2,267 |
| | Lost productivity associated with illness | . | . | 580,777,411 | 32,451 |
| | Total cost (excl. lost productivity) | 374,311,669 | 15,429 | 282,455,132 | 11,643 |
| Brain tumors | Primary sector | 7,838,703 | 510 | 1,941,205 | 126 |
| | Hospital care | 123,575,004 | 8,035 | 77,329,980 | 5,028 |
| | Filled prescriptions | 9,121,248 | 593 | 3,440,563 | 224 |
| | Home care | 15,233,128 | 991 | 9,636,878 | 627 |
| | Nursing home / sheltered accomodation | 34,615,194 | 2,251 | 16,262,784 | 1,057 |

***Sensitivity analysis 1 / Prevalent cohort 2016***

| *Brain disorder* | *Cost component* | *Actual costs (EUR)* | *Actual costs per person* | *Attributable costs (EUR)* | *Attributable costs per person* |
|---|---|---|---|---|---|
| | Lost productivity associated with illness | . | . | 76,567,290 | 10,080 |
| | Total cost (excl. lost productivity) | 190,383,277 | 12,379 | 108,611,409 | 7,062 |
| Cerebral palsy | Primary sector | 6,447,493 | 693 | 4,525,552 | 487 |
| | Hospital care | 48,917,348 | 5,259 | 36,144,883 | 3,886 |
| | Filled prescriptions | 6,458,876 | 694 | 5,198,132 | 559 |
| | Home care | 14,288,704 | 1,536 | 13,901,891 | 1,495 |
| | Nursing home / sheltered accomodation | 12,929,498 | 1,390 | 11,831,077 | 1,272 |
| | Lost productivity associated with illness | . | . | 161,837,806 | 30,616 |
| | Total cost (excl. lost productivity) | 89,041,918 | 9,573 | 71,601,536 | 7,698 |
| Dementia | Primary sector | 19,864,913 | 535 | 864,528 | 23 |
| | Hospital care | 215,852,658 | 5,818 | 44,657,261 | 1,204 |
| | Filled prescriptions | 35,612,794 | 960 | 13,941,109 | 376 |
| | Home care | 138,178,750 | 3,725 | 81,904,228 | 2,208 |
| | Nursing home / sheltered accomodation | 1,298,031,124 | 34,988 | 1,148,351,085 | 30,954 |
| | Lost productivity associated with illness | . | . | 134,012,784 | 40,268 |
| | Total cost (excl. lost productivity) | 1,707,540,240 | 46,027 | 1,289,718,212 | 34,764 |
| Depression | Primary sector | 91,282,560 | 489 | 30,560,585 | 164 |
| | Hospital care | 1,158,800,146 | 6,208 | 735,274,307 | 3,939 |
| | Filled prescriptions | 118,387,273 | 634 | 69,671,141 | 373 |
| | Home care | 130,370,797 | 698 | 81,531,667 | 437 |
| | Nursing home / sheltered accomodation | 569,025,822 | 3,049 | 418,114,139 | 2,240 |
| | Lost productivity associated with illness | . | . | 3,610,692,063 | 25,983 |
| | Total cost (excl. lost productivity) | 2,067,866,598 | 11,079 | 1,335,151,839 | 7,153 |
| Developmental and behavioural disorders | Primary sector | 28,695,848 | 253 | 10,147,075 | 89 |
| | Hospital care | 438,547,122 | 3,868 | 325,788,914 | 2,873 |
| | Filled prescriptions | 55,369,525 | 488 | 46,571,999 | 411 |
| | Home care | 5,111,676 | 45 | 3,698,475 | 33 |
| | Nursing home / sheltered accomodation | 9,963,153 | 88 | 7,600,502 | 67 |
| | Lost productivity associated with illness | . | . | 1,108,216,391 | 16,822 |
| | Total cost (excl. lost productivity) | 537,687,324 | 4,742 | 393,806,964 | 3,473 |
| Drug abuse | Primary sector | 18,180,319 | 353 | 5,601,106 | 109 |
| | Hospital care | 477,895,897 | 9,268 | 396,305,547 | 7,686 |
| | Filled prescriptions | 37,602,694 | 729 | 28,712,494 | 557 |

***Sensitivity analysis 1 / Prevalent cohort 2016***

| *Brain disorder* | *Cost component* | *Actual costs (EUR)* | *Actual costs per person* | *Attributable costs (EUR)* | *Attributable costs per person* |
|---|---|---|---|---|---|
| | Home care | 18,275,141 | 354 | 14,154,470 | 275 |
| | Nursing home / sheltered accomodation | 57,429,916 | 1,114 | 47,160,336 | 915 |
| | Lost productivity associated with illness | . | . | 1,603,031,944 | 34,579 |
| | Total cost (excl. lost productivity) | 609,383,968 | 11,818 | 491,933,952 | 9,540 |
| Eating disorders | Primary sector | 6,886,547 | 408 | 2,039,170 | 121 |
| | Hospital care | 136,007,704 | 8,057 | 107,011,167 | 6,339 |
| | Filled prescriptions | 4,941,165 | 293 | 2,523,560 | 149 |
| | Home care | 917,025 | 54 | 636,268 | 38 |
| | Nursing home / sheltered accomodation | 1,747,417 | 104 | 1,368,999 | 81 |
| | Lost productivity associated with illness | . | . | 113,554,451 | 7,395 |
| | Total cost (excl. lost productivity) | 150,499,857 | 8,915 | 113,579,163 | 6,728 |
| Epilepsy | Primary sector | 31,378,102 | 432 | 10,190,485 | 140 |
| | Hospital care | 413,962,098 | 5,701 | 256,385,441 | 3,531 |
| | Filled prescriptions | 55,668,245 | 767 | 38,349,315 | 528 |
| | Home care | 68,787,102 | 947 | 55,008,941 | 758 |
| | Nursing home / sheltered accomodation | 214,445,994 | 2,953 | 173,464,760 | 2,389 |
| | Lost productivity associated with illness | . | . | 890,618,645 | 19,438 |
| | Total cost (excl. lost productivity) | 784,241,540 | 10,800 | 533,398,941 | 7,346 |
| Headache | Primary sector | 45,671,958 | 438 | 14,225,946 | 137 |
| | Hospital care | 374,631,800 | 3,596 | 158,901,728 | 1,525 |
| | Filled prescriptions | 42,293,008 | 406 | 18,560,118 | 178 |
| | Home care | 16,540,058 | 159 | 3,910,077 | 38 |
| | Nursing home / sheltered accomodation | 43,501,204 | 418 | 9,666,439 | 93 |
| | Lost productivity associated with illness | . | . | 554,186,768 | 6,821 |
| | Total cost (excl. lost productivity) | 522,638,027 | 5,017 | 205,264,308 | 1,971 |
| Infections of the CNS | Primary sector | 9,810,554 | 360 | 1,961,549 | 72 |
| | Hospital care | 103,238,460 | 3,784 | 45,005,475 | 1,649 |
| | Filled prescriptions | 9,535,830 | 349 | 3,034,739 | 111 |
| | Home care | 12,651,977 | 464 | 7,680,676 | 281 |
| | Nursing home / sheltered accomodation | 31,203,541 | 1,144 | 17,078,167 | 626 |
| | Lost productivity associated with illness | . | . | 33,574,864 | 2,013 |
| | Total cost (excl. lost productivity) | 166,440,362 | 6,100 | 74,760,606 | 2,740 |
| Intellectual disability | Primary sector | 11,503,428 | 405 | 5,362,465 | 189 |
| | Hospital care | 157,326,502 | 5,544 | 114,619,260 | 4,039 |
| | Filled prescriptions | 20,932,786 | 738 | 16,786,719 | 592 |
| | Home care | 13,732,896 | 484 | 12,388,652 | 437 |
| | Nursing home / sheltered accomodation | 31,811,639 | 1,121 | 28,658,365 | 1,010 |
| | Lost productivity associated with illness | . | . | 648,278,739 | 34,573 |
| | Total cost (excl. lost productivity) | 235,307,250 | 8,292 | 177,815,461 | 6,266 |

***Sensitivity analysis 1 / Prevalent cohort 2016***

| *Brain disorder* | *Cost component* | *Actual costs (EUR)* | *Actual costs per person* | *Attributable costs (EUR)* | *Attributable costs per person* |
|---|---|---|---|---|---|
| Multiple sclerosis | Primary sector | 12,140,477 | 793 | 6,927,488 | 452 |
| | Hospital care | 123,288,244 | 8,051 | 87,295,531 | 5,700 |
| | Filled prescriptions | 9,703,236 | 634 | 5,395,720 | 352 |
| | Home care | 51,717,917 | 3,377 | 49,764,875 | 3,250 |
| | Nursing home / sheltered accomodation | 34,616,874 | 2,260 | 29,910,332 | 1,953 |
| | Lost productivity associated with illness | . | . | 222,178,465 | 18,603 |
| | Total cost (excl. lost productivity) | 231,466,748 | 15,115 | 179,293,945 | 11,708 |
| Neuromuscular disorders | Primary sector | 4,322,743 | 572 | 1,900,437 | 252 |
| | Hospital care | 54,329,416 | 7,192 | 35,574,802 | 4,709 |
| | Filled prescriptions | 4,806,337 | 636 | 2,665,396 | 353 |
| | Home care | 8,885,283 | 1,176 | 7,169,700 | 949 |
| | Nursing home / sheltered accomodation | 10,623,765 | 1,406 | 5,396,726 | 714 |
| | Lost productivity associated with illness | . | . | 72,243,890 | 15,620 |
| | Total cost (excl. lost productivity) | 82,967,543 | 10,983 | 52,707,061 | 6,977 |
| Other neurodegenerative disorders | Primary sector | 1,823,492 | 789 | 985,465 | 426 |
| | Hospital care | 19,377,644 | 8,385 | 12,688,506 | 5,491 |
| | Filled prescriptions | 2,515,727 | 1,089 | 1,710,139 | 740 |
| | Home care | 10,494,752 | 4,541 | 9,865,208 | 4,269 |
| | Nursing home / sheltered accomodation | 18,390,201 | 7,958 | 16,407,481 | 7,100 |
| | Lost productivity associated with illness | . | . | 39,674,730 | 33,340 |
| | Total cost (excl. lost productivity) | 52,601,816 | 22,763 | 41,656,800 | 18,026 |
| Parkinson’s disease | Primary sector | 9,372,688 | 969 | 4,760,811 | 492 |
| | Hospital care | 80,598,805 | 8,337 | 38,328,136 | 3,965 |
| | Filled prescriptions | 29,249,352 | 3,026 | 24,237,505 | 2,507 |
| | Home care | 44,310,887 | 4,583 | 36,615,539 | 3,787 |
| | Nursing home / sheltered accomodation | 142,509,305 | 14,741 | 118,023,265 | 12,208 |
| | Lost productivity associated with illness | . | . | 35,260,649 | 22,941 |
| | Total cost (excl. lost productivity) | 306,041,037 | 31,656 | 221,965,257 | 22,960 |
| Personality disorders | Primary sector | 32,965,469 | 451 | 11,982,643 | 164 |
| | Hospital care | 531,867,399 | 7,272 | 400,779,841 | 5,480 |
| | Filled prescriptions | 44,111,804 | 603 | 30,484,603 | 417 |
| | Home care | 15,915,204 | 218 | 11,433,643 | 156 |
| | Nursing home / sheltered accomodation | 49,968,684 | 683 | 40,373,420 | 552 |
| | Lost productivity associated with illness | . | . | 2,010,511,407 | 30,000 |
| | Total cost (excl. lost productivity) | 674,828,560 | 9,227 | 495,054,150 | 6,769 |
| Polyneuropathy | Primary sector | 21,305,752 | 604 | 7,081,272 | 201 |
| | Hospital care | 294,419,525 | 8,351 | 173,157,486 | 4,911 |
| | Filled prescriptions | 33,021,912 | 937 | 18,489,107 | 524 |
| | Home care | 47,173,402 | 1,338 | 29,239,959 | 829 |

*Sensitivity analysis 1 / Prevalent cohort 2016*

| *Brain disorder* | *Cost component* | *Actual costs (EUR)* | *Actual costs per person* | *Attributable costs (EUR)* | *Attributable costs per person* |
|---|---|---|---|---|---|
| | Nursing home / sheltered accomodation | 104,181,244 | 2,955 | 44,064,432 | 1,250 |
| | Lost productivity associated with illness | . | . | 311,889,633 | 21,030 |
| | Total cost (excl. lost productivity) | 500,101,835 | 14,185 | 272,032,256 | 7,716 |
| Schizophrenia spectrum disorders | Primary sector | 24,486,063 | 393 | 6,875,196 | 110 |
| | Hospital care | 774,136,108 | 12,421 | 656,408,245 | 10,532 |
| | Filled prescriptions | 55,506,240 | 891 | 42,038,613 | 675 |
| | Home care | 33,831,782 | 543 | 25,735,450 | 413 |
| | Nursing home / sheltered accomodation | 157,207,224 | 2,522 | 135,730,255 | 2,178 |
| | Lost productivity associated with illness | . | . | 2,044,342,404 | 39,673 |
| | Total cost (excl. lost productivity) | 1,045,167,418 | 16,770 | 866,787,759 | 13,908 |
| Sleep disorders | Primary sector | 39,469,292 | 476 | 13,819,640 | 167 |
| | Hospital care | 378,203,971 | 4,561 | 166,282,622 | 2,005 |
| | Filled prescriptions | 53,672,719 | 647 | 29,835,865 | 360 |
| | Home care | 22,850,137 | 276 | 9,118,700 | 110 |
| | Nursing home / sheltered accomodation | 40,643,876 | 490 | 5,896,222 | 71 |
| | Lost productivity associated with illness | . | . | 503,197,195 | 9,710 |
| | Total cost (excl. lost productivity) | 534,839,995 | 6,450 | 224,953,049 | 2,713 |
| Stress related disorders | Primary sector | 87,251,568 | 419 | 29,135,320 | 140 |
| | Hospital care | 1,147,583,890 | 5,507 | 773,003,850 | 3,709 |
| | Filled prescriptions | 93,812,681 | 450 | 52,805,538 | 253 |
| | Home care | 48,830,675 | 234 | 30,935,749 | 148 |
| | Nursing home / sheltered accomodation | 148,257,989 | 711 | 104,312,084 | 501 |
| | Lost productivity associated with illness | . | . | 3,992,221,781 | 22,975 |
| | Total cost (excl. lost productivity) | 1,525,736,802 | 7,321 | 990,192,540 | 4,751 |
| Stroke | Primary sector | 69,225,933 | 587 | 18,085,898 | 153 |
| | Hospital care | 763,872,390 | 6,475 | 320,635,702 | 2,718 |
| | Filled prescriptions | 94,213,532 | 799 | 40,483,309 | 343 |
| | Home care | 247,961,522 | 2,102 | 171,604,905 | 1,455 |
| | Nursing home / sheltered accomodation | 784,623,526 | 6,651 | 515,459,429 | 4,369 |
| | Lost productivity associated with illness | . | . | 721,110,082 | 19,960 |
| | Total cost (excl. lost productivity) | 1,959,896,904 | 16,614 | 1,066,269,244 | 9,039 |
| Traumatic brain injury | Primary sector | 76,708,884 | 312 | 13,987,701 | 57 |
| | Hospital care | 780,478,877 | 3,172 | 328,540,164 | 1,335 |
| | Filled prescriptions | 73,524,724 | 299 | 25,374,687 | 103 |
| | Home care | 86,861,067 | 353 | 48,611,210 | 198 |
| | Nursing home / sheltered accomodation | 312,247,349 | 1,269 | 189,498,735 | 770 |
| | Lost productivity associated with illness | . | . | 964,500,682 | 5,671 |
| | Total cost (excl. lost productivity) | 1,329,820,901 | 5,405 | 606,012,498 | 2,463 |

***Sensitivity analysis 1 / Prevalent cohort 2017***

| Brain disorder | Cost component | Actual costs (EUR) | Actual costs per person | Attributable costs (EUR) | Attributable costs per person |
|---|---|---|---|---|---|
| Any brain disorder | Primary sector | 467,173,110 | 409 | 146,557,741 | 128 |
| | Hospital care | 5,160,156,941 | 4,513 | 2,943,939,286 | 2,575 |
| | Filled prescriptions | 527,455,141 | 461 | 303,324,768 | 265 |
| | Home care | 693,245,267 | 606 | 537,883,611 | 470 |
| | Nursing home / sheltered accomodation | 2,404,038,812 | 2,102 | 2,069,523,046 | 1,810 |
| | Lost productivity associated with illness | . | . | 13,642,739,073 | 18,326 |
| | Total cost (excl. lost productivity) | 9,252,069,271 | 8,092 | 6,001,228,451 | 5,248 |
| Alcohol abuse | Primary sector | 41,423,093 | 420 | 8,795,091 | 89 |
| | Hospital care | 829,442,261 | 8,404 | 561,936,894 | 5,694 |
| | Filled prescriptions | 64,339,368 | 652 | 35,473,908 | 359 |
| | Home care | 87,211,227 | 884 | 69,537,365 | 705 |
| | Nursing home / sheltered accomodation | 312,983,789 | 3,171 | 272,979,135 | 2,766 |
| | Lost productivity associated with illness | . | . | 2,466,656,705 | 36,946 |
| | Total cost (excl. lost productivity) | 1,335,399,738 | 13,530 | 948,722,392 | 9,613 |
| Anxiety disorders | Primary sector | 50,726,737 | 456 | 18,410,943 | 165 |
| | Hospital care | 649,230,523 | 5,830 | 420,175,730 | 3,773 |
| | Filled prescriptions | 55,258,807 | 496 | 33,072,818 | 297 |
| | Home care | 36,171,248 | 325 | 23,340,953 | 210 |
| | Nursing home / sheltered accomodation | 110,505,048 | 992 | 75,792,286 | 681 |
| | Lost productivity associated with illness | . | . | 2,076,411,323 | 22,732 |
| | Total cost (excl. lost productivity) | 901,892,363 | 8,099 | 570,792,730 | 5,125 |
| Bipolar disorder | Primary sector | 12,972,365 | 516 | 4,536,927 | 181 |
| | Hospital care | 254,897,276 | 10,147 | 189,923,795 | 7,560 |
| | Filled prescriptions | 19,103,216 | 760 | 12,273,302 | 489 |
| | Home care | 16,763,180 | 667 | 12,035,513 | 479 |
| | Nursing home / sheltered accomodation | 68,372,748 | 2,722 | 55,079,262 | 2,193 |
| | Lost productivity associated with illness | . | . | 618,237,500 | 33,057 |
| | Total cost (excl. lost productivity) | 372,108,785 | 14,813 | 273,848,799 | 10,901 |
| Brain tumors | Primary sector | 8,482,543 | 523 | 2,128,680 | 131 |
| | Hospital care | 134,793,436 | 8,310 | 82,470,846 | 5,084 |
| | Filled prescriptions | 9,133,586 | 563 | 3,173,158 | 196 |
| | Home care | 17,086,753 | 1,053 | 10,874,397 | 670 |
| | Nursing home / sheltered accomodation | 39,047,697 | 2,407 | 19,659,325 | 1,212 |

*Sensitivity analysis 1 / Prevalent cohort 2017*

| *Brain disorder* | *Cost component* | *Actual costs (EUR)* | *Actual costs per person* | *Attributable costs (EUR)* | *Attributable costs per person* |
|---|---|---|---|---|---|
| | Lost productivity associated with illness | . | . | 79,220,419 | 10,010 |
| | Total cost (excl. lost productivity) | 208,544,015 | 12,857 | 118,306,406 | 7,294 |
| Cerebral palsy | Primary sector | 6,380,318 | 681 | 4,417,396 | 472 |
| | Hospital care | 50,135,694 | 5,352 | 35,863,742 | 3,829 |
| | Filled prescriptions | 6,589,978 | 704 | 5,324,389 | 568 |
| | Home care | 15,351,078 | 1,639 | 14,894,263 | 1,590 |
| | Nursing home / sheltered accomodation | 13,175,468 | 1,407 | 12,086,909 | 1,290 |
| | Lost productivity associated with illness | . | . | 150,642,265 | 28,295 |
| | Total cost (excl. lost productivity) | 91,632,535 | 9,782 | 72,586,700 | 7,749 |
| Dementia | Primary sector | 20,673,085 | 551 | 987,287 | 26 |
| | Hospital care | 216,132,490 | 5,760 | 33,950,496 | 905 |
| | Filled prescriptions | 33,716,972 | 899 | 11,925,941 | 318 |
| | Home care | 145,821,021 | 3,886 | 90,392,489 | 2,409 |
| | Nursing home / sheltered accomodation | 1,289,077,898 | 34,355 | 1,143,094,134 | 30,464 |
| | Lost productivity associated with illness | . | . | 134,202,294 | 40,779 |
| | Total cost (excl. lost productivity) | 1,705,421,465 | 45,451 | 1,280,350,348 | 34,122 |
| Depression | Primary sector | 94,718,014 | 492 | 31,507,776 | 164 |
| | Hospital care | 1,219,120,751 | 6,329 | 750,173,459 | 3,895 |
| | Filled prescriptions | 112,528,292 | 584 | 63,561,867 | 330 |
| | Home care | 134,896,593 | 700 | 86,653,883 | 450 |
| | Nursing home / sheltered accomodation | 557,024,961 | 2,892 | 410,977,877 | 2,134 |
| | Lost productivity associated with illness | . | . | 3,792,636,470 | 26,296 |
| | Total cost (excl. lost productivity) | 2,118,288,610 | 10,997 | 1,342,874,863 | 6,972 |
| Developmental and behavioural disorders | Primary sector | 31,301,603 | 256 | 11,166,245 | 91 |
| | Hospital care | 478,196,496 | 3,915 | 348,395,967 | 2,852 |
| | Filled prescriptions | 55,503,320 | 454 | 45,908,811 | 376 |
| | Home care | 5,503,271 | 45 | 3,876,038 | 32 |
| | Nursing home / sheltered accomodation | 9,915,262 | 81 | 7,775,024 | 64 |
| | Lost productivity associated with illness | . | . | 1,256,321,307 | 17,304 |
| | Total cost (excl. lost productivity) | 580,419,952 | 4,752 | 417,122,085 | 3,415 |
| Drug abuse | Primary sector | 18,577,786 | 350 | 5,659,942 | 107 |
| | Hospital care | 507,504,604 | 9,558 | 416,804,365 | 7,850 |
| | Filled prescriptions | 35,061,752 | 660 | 26,188,601 | 493 |

***Sensitivity analysis 1 / Prevalent cohort 2017***

| *Brain disorder* | *Cost component* | *Actual costs (EUR)* | *Actual costs per person* | *Attributable costs (EUR)* | *Attributable costs per person* |
|---|---|---|---|---|---|
| | Home care | 22,343,486 | 421 | 18,173,246 | 342 |
| | Nursing home / sheltered accomodation | 56,802,073 | 1,070 | 46,456,581 | 875 |
| | Lost productivity associated with illness | . | . | 1,671,568,718 | 34,956 |
| | Total cost (excl. lost productivity) | 640,289,700 | 12,059 | 513,282,736 | 9,667 |
| Eating disorders | Primary sector | 7,001,602 | 401 | 1,955,999 | 112 |
| | Hospital care | 131,813,062 | 7,542 | 98,816,164 | 5,654 |
| | Filled prescriptions | 4,919,128 | 281 | 2,418,640 | 138 |
| | Home care | 875,276 | 50 | 557,572 | 32 |
| | Nursing home / sheltered accomodation | 1,691,543 | 97 | 1,425,374 | 82 |
| | Lost productivity associated with illness | . | . | 97,170,927 | 6,054 |
| | Total cost (excl. lost productivity) | 146,300,611 | 8,371 | 105,173,750 | 6,018 |
| Epilepsy | Primary sector | 31,545,596 | 435 | 10,043,122 | 138 |
| | Hospital care | 419,602,293 | 5,786 | 249,824,842 | 3,445 |
| | Filled prescriptions | 54,419,801 | 750 | 37,278,944 | 514 |
| | Home care | 73,487,350 | 1,013 | 59,432,814 | 820 |
| | Nursing home / sheltered accomodation | 215,913,202 | 2,977 | 173,086,601 | 2,387 |
| | Lost productivity associated with illness | . | . | 899,463,243 | 19,790 |
| | Total cost (excl. lost productivity) | 794,968,242 | 10,963 | 529,666,321 | 7,304 |
| Headache | Primary sector | 48,675,253 | 444 | 15,200,568 | 139 |
| | Hospital care | 409,490,374 | 3,734 | 164,112,890 | 1,497 |
| | Filled prescriptions | 42,037,403 | 383 | 17,615,032 | 161 |
| | Home care | 19,960,922 | 182 | 6,210,413 | 57 |
| | Nursing home / sheltered accomodation | 44,616,156 | 407 | 9,399,287 | 86 |
| | Lost productivity associated with illness | . | . | 562,924,098 | 6,588 |
| | Total cost (excl. lost productivity) | 564,780,108 | 5,150 | 212,538,190 | 1,938 |
| Infections of the CNS | Primary sector | 10,191,318 | 368 | 2,080,257 | 75 |
| | Hospital care | 112,360,780 | 4,053 | 47,876,407 | 1,727 |
| | Filled prescriptions | 9,651,747 | 348 | 3,078,485 | 111 |
| | Home care | 13,333,165 | 481 | 7,672,234 | 277 |
| | Nursing home / sheltered accomodation | 30,311,243 | 1,093 | 15,565,037 | 561 |
| | Lost productivity associated with illness | . | . | 41,069,261 | 2,448 |
| | Total cost (excl. lost productivity) | 175,848,253 | 6,343 | 76,272,421 | 2,751 |
| Intellectual disability | Primary sector | 11,797,744 | 405 | 5,440,422 | 187 |
| | Hospital care | 167,175,413 | 5,733 | 120,618,946 | 4,136 |
| | Filled prescriptions | 20,315,116 | 697 | 16,193,266 | 555 |
| | Home care | 13,272,438 | 455 | 11,860,937 | 407 |
| | Nursing home / sheltered accomodation | 32,892,773 | 1,128 | 29,710,288 | 1,019 |
| | Lost productivity associated with illness | . | . | 670,902,577 | 34,092 |
| | Total cost (excl. lost productivity) | 245,453,485 | 8,417 | 183,823,859 | 6,303 |

***Sensitivity analysis 1 / Prevalent cohort 2017***

| *Brain disorder* | *Cost component* | *Actual costs (EUR)* | *Actual costs per person* | *Attributable costs (EUR)* | *Attributable costs per person* |
|---|---|---|---|---|---|
| Multiple sclerosis | Primary sector | 12,397,569 | 785 | 6,967,157 | 441 |
| | Hospital care | 145,794,571 | 9,235 | 104,949,289 | 6,648 |
| | Filled prescriptions | 9,408,996 | 596 | 5,077,958 | 322 |
| | Home care | 56,412,110 | 3,573 | 54,335,268 | 3,442 |
| | Nursing home / sheltered accomodation | 35,485,226 | 2,248 | 30,281,469 | 1,918 |
| | Lost productivity associated with illness | . | . | 238,151,891 | 19,398 |
| | Total cost (excl. lost productivity) | 259,498,473 | 16,437 | 201,611,141 | 12,770 |
| Neuromuscular disorders | Primary sector | 4,605,422 | 584 | 2,048,190 | 260 |
| | Hospital care | 59,051,246 | 7,487 | 38,301,340 | 4,856 |
| | Filled prescriptions | 4,929,441 | 625 | 2,742,879 | 348 |
| | Home care | 9,907,232 | 1,256 | 8,093,928 | 1,026 |
| | Nursing home / sheltered accomodation | 11,381,630 | 1,443 | 6,135,090 | 778 |
| | Lost productivity associated with illness | . | . | 77,031,541 | 16,062 |
| | Total cost (excl. lost productivity) | 89,874,971 | 11,394 | 57,321,427 | 7,267 |
| Other neurodegenerative disorders | Primary sector | 2,030,948 | 823 | 1,116,297 | 452 |
| | Hospital care | 20,690,696 | 8,380 | 12,873,372 | 5,214 |
| | Filled prescriptions | 2,349,666 | 952 | 1,515,092 | 614 |
| | Home care | 12,087,817 | 4,895 | 11,379,131 | 4,608 |
| | Nursing home / sheltered accomodation | 19,413,151 | 7,862 | 17,455,273 | 7,069 |
| | Lost productivity associated with illness | . | . | 40,816,353 | 32,758 |
| | Total cost (excl. lost productivity) | 56,572,278 | 22,911 | 44,339,164 | 17,957 |
| Parkinson’s disease | Primary sector | 9,966,333 | 1,003 | 5,125,162 | 516 |
| | Hospital care | 84,874,232 | 8,539 | 39,411,405 | 3,965 |
| | Filled prescriptions | 29,816,783 | 3,000 | 24,740,588 | 2,489 |
| | Home care | 48,574,837 | 4,887 | 40,524,400 | 4,077 |
| | Nursing home / sheltered accomodation | 142,289,381 | 14,316 | 117,719,405 | 11,844 |
| | Lost productivity associated with illness | . | . | 36,635,355 | 23,992 |
| | Total cost (excl. lost productivity) | 315,521,566 | 31,744 | 227,520,960 | 22,891 |
| Personality disorders | Primary sector | 33,199,105 | 449 | 11,856,064 | 160 |
| | Hospital care | 557,835,160 | 7,538 | 413,350,993 | 5,585 |
| | Filled prescriptions | 40,204,021 | 543 | 26,749,465 | 361 |
| | Home care | 16,988,795 | 230 | 12,470,122 | 169 |
| | Nursing home / sheltered accomodation | 48,175,896 | 651 | 39,280,584 | 531 |
| | Lost productivity associated with illness | . | . | 2,063,778,940 | 30,352 |
| | Total cost (excl. lost productivity) | 696,402,977 | 9,410 | 503,707,228 | 6,806 |
| Polyneuropathy | Primary sector | 22,782,733 | 616 | 7,530,819 | 204 |
| | Hospital care | 319,633,253 | 8,645 | 183,427,805 | 4,961 |
| | Filled prescriptions | 33,717,069 | 912 | 18,599,313 | 503 |
| | Home care | 52,763,606 | 1,427 | 32,908,455 | 890 |

*Sensitivity analysis 1 / Prevalent cohort 2017*

| *Brain disorder* | *Cost component* | *Actual costs (EUR)* | *Actual costs per person* | *Attributable costs (EUR)* | *Attributable costs per person* |
|---|---|---|---|---|---|
| | Nursing home / sheltered accomodation | 109,511,506 | 2,962 | 46,723,166 | 1,264 |
| | Lost productivity associated with illness | . | . | 317,418,673 | 20,821 |
| | Total cost (excl. lost productivity) | 538,408,166 | 14,562 | 289,189,558 | 7,822 |
| Schizophrenia spectrum disorders | Primary sector | 24,948,590 | 391 | 6,872,605 | 108 |
| | Hospital care | 802,997,585 | 12,590 | 673,148,631 | 10,554 |
| | Filled prescriptions | 50,592,377 | 793 | 37,266,109 | 584 |
| | Home care | 36,944,922 | 579 | 28,731,568 | 450 |
| | Nursing home / sheltered accomodation | 155,479,972 | 2,438 | 133,690,831 | 2,096 |
| | Lost productivity associated with illness | . | . | 2,110,262,849 | 39,902 |
| | Total cost (excl. lost productivity) | 1,070,963,446 | 16,791 | 879,709,745 | 13,792 |
| Sleep disorders | Primary sector | 44,150,427 | 483 | 15,321,327 | 168 |
| | Hospital care | 449,574,949 | 4,923 | 198,155,063 | 2,170 |
| | Filled prescriptions | 58,460,247 | 640 | 32,427,917 | 355 |
| | Home care | 30,322,036 | 332 | 14,245,762 | 156 |
| | Nursing home / sheltered accomodation | 45,670,614 | 500 | 5,296,787 | 58 |
| | Lost productivity associated with illness | . | . | 590,056,645 | 10,424 |
| | Total cost (excl. lost productivity) | 628,178,273 | 6,879 | 265,446,856 | 2,907 |
| Stress related disorders | Primary sector | 92,297,687 | 420 | 30,613,081 | 139 |
| | Hospital care | 1,241,798,002 | 5,653 | 816,311,387 | 3,716 |
| | Filled prescriptions | 92,217,305 | 420 | 49,972,991 | 227 |
| | Home care | 53,513,887 | 244 | 33,063,787 | 151 |
| | Nursing home / sheltered accomodation | 156,807,707 | 714 | 111,894,019 | 509 |
| | Lost productivity associated with illness | . | . | 4,293,710,407 | 23,470 |
| | Total cost (excl. lost productivity) | 1,636,634,587 | 7,450 | 1,041,855,265 | 4,743 |
| Stroke | Primary sector | 71,374,240 | 596 | 18,280,149 | 153 |
| | Hospital care | 819,609,080 | 6,841 | 341,511,075 | 2,850 |
| | Filled prescriptions | 93,220,609 | 778 | 39,380,731 | 329 |
| | Home care | 259,556,363 | 2,166 | 181,780,907 | 1,517 |
| | Nursing home / sheltered accomodation | 781,619,583 | 6,524 | 515,170,503 | 4,300 |
| | Lost productivity associated with illness | . | . | 755,926,566 | 20,771 |
| | Total cost (excl. lost productivity) | 2,025,379,873 | 16,905 | 1,096,123,365 | 9,149 |
| Traumatic brain injury | Primary sector | 77,363,391 | 318 | 14,168,893 | 58 |
| | Hospital care | 815,751,905 | 3,350 | 333,081,514 | 1,368 |
| | Filled prescriptions | 71,390,224 | 293 | 23,938,889 | 98 |
| | Home care | 91,890,315 | 377 | 52,731,330 | 217 |
| | Nursing home / sheltered accomodation | 319,311,375 | 1,311 | 197,009,296 | 809 |
| | Lost productivity associated with illness | . | . | 1,338,220,051 | 7,985 |
| | Total cost (excl. lost productivity) | 1,375,707,208 | 5,649 | 620,929,921 | 2,550 |

***Sensitivity analysis 1 / Prevalent cohort 2018***

| *Brain disorder* | *Cost component* | *Actual costs (EUR)* | *Actual costs per person* | *Attributable costs (EUR)* | *Attributable costs per person* |
|---|---|---|---|---|---|
| Any brain disorder | Primary sector | 483,920,171 | 412 | 159,759,125 | 136 |
| | Hospital care | 4,004,686,351 | 3,413 | 2,380,698,569 | 2,029 |
| | Filled prescriptions | 539,685,601 | 460 | 310,441,108 | 265 |
| | Home care | 588,116,405 | 501 | 456,036,335 | 389 |
| | Nursing home / sheltered accomodation | 2,402,223,128 | 2,047 | 2,077,862,731 | 1,771 |
| | Lost productivity associated with illness | . | . | 14,262,906,120 | 18,682 |
| | Total cost (excl. lost productivity) | 8,018,631,657 | 6,834 | 5,384,797,868 | 4,589 |
| Alcohol abuse | Primary sector | 41,841,250 | 430 | 9,889,447 | 102 |
| | Hospital care | 698,427,918 | 7,173 | 509,626,027 | 5,234 |
| | Filled prescriptions | 63,102,414 | 648 | 34,501,059 | 354 |
| | Home care | 72,442,056 | 744 | 57,541,610 | 591 |
| | Nursing home / sheltered accomodation | 316,129,097 | 3,247 | 276,084,310 | 2,836 |
| | Lost productivity associated with illness | . | . | 2,415,652,023 | 37,280 |
| | Total cost (excl. lost productivity) | 1,191,942,734 | 12,242 | 887,642,452 | 9,117 |
| Anxiety disorders | Primary sector | 53,842,742 | 455 | 19,975,782 | 169 |
| | Hospital care | 539,505,947 | 4,564 | 366,041,250 | 3,097 |
| | Filled prescriptions | 56,898,042 | 481 | 33,449,057 | 283 |
| | Home care | 31,774,722 | 269 | 19,913,421 | 168 |
| | Nursing home / sheltered accomodation | 115,022,832 | 973 | 78,967,124 | 668 |
| | Lost productivity associated with illness | . | . | 2,240,629,033 | 23,124 |
| | Total cost (excl. lost productivity) | 797,044,286 | 6,743 | 518,346,633 | 4,385 |
| Bipolar disorder | Primary sector | 13,565,445 | 524 | 4,965,647 | 192 |
| | Hospital care | 213,928,662 | 8,263 | 165,890,328 | 6,407 |
| | Filled prescriptions | 19,355,545 | 748 | 12,369,442 | 478 |
| | Home care | 14,609,825 | 564 | 10,599,205 | 409 |
| | Nursing home / sheltered accomodation | 67,085,974 | 2,591 | 54,040,041 | 2,087 |
| | Lost productivity associated with illness | . | . | 641,956,340 | 33,164 |
| | Total cost (excl. lost productivity) | 328,545,451 | 12,690 | 247,864,662 | 9,573 |
| Brain tumors | Primary sector | 8,942,234 | 529 | 2,285,582 | 135 |
| | Hospital care | 97,371,986 | 5,757 | 57,402,040 | 3,394 |
| | Filled prescriptions | 9,709,236 | 574 | 3,395,762 | 201 |
| | Home care | 14,807,501 | 876 | 8,962,969 | 530 |
| | Nursing home / sheltered accomodation | 39,750,948 | 2,350 | 19,471,173 | 1,151 |

***Sensitivity analysis 1 / Prevalent cohort 2018***

| *Brain disorder* | *Cost component* | *Actual costs (EUR)* | *Actual costs per person* | *Attributable costs (EUR)* | *Attributable costs per person* |
|---|---|---|---|---|---|
| | Lost productivity associated with illness | . | . | 82,665,785 | 10,189 |
| | Total cost (excl. lost productivity) | 170,581,907 | 10,086 | 91,517,527 | 5,411 |
| Cerebral palsy | Primary sector | 6,677,649 | 709 | 4,703,953 | 499 |
| | Hospital care | 35,189,192 | 3,734 | 24,845,965 | 2,636 |
| | Filled prescriptions | 6,862,500 | 728 | 5,587,299 | 593 |
| | Home care | 12,161,833 | 1,291 | 11,780,462 | 1,250 |
| | Nursing home / sheltered accomodation | 13,814,864 | 1,466 | 12,761,906 | 1,354 |
| | Lost productivity associated with illness | . | . | 158,461,644 | 29,086 |
| | Total cost (excl. lost productivity) | 74,706,038 | 7,927 | 59,679,587 | 6,333 |
| Dementia | Primary sector | 23,202,093 | 621 | 3,251,498 | 87 |
| | Hospital care | 184,892,081 | 4,951 | 45,219,248 | 1,211 |
| | Filled prescriptions | 33,305,745 | 892 | 11,268,079 | 302 |
| | Home care | 123,392,767 | 3,304 | 77,777,365 | 2,083 |
| | Nursing home / sheltered accomodation | 1,276,360,552 | 34,177 | 1,134,893,721 | 30,389 |
| | Lost productivity associated with illness | . | . | 133,686,665 | 41,934 |
| | Total cost (excl. lost productivity) | 1,641,153,237 | 43,945 | 1,272,409,911 | 34,071 |
| Depression | Primary sector | 98,057,461 | 496 | 33,879,176 | 171 |
| | Hospital care | 983,740,505 | 4,971 | 642,799,805 | 3,248 |
| | Filled prescriptions | 113,329,958 | 573 | 63,470,364 | 321 |
| | Home care | 112,727,443 | 570 | 71,920,741 | 363 |
| | Nursing home / sheltered accomodation | 538,510,829 | 2,721 | 397,752,648 | 2,010 |
| | Lost productivity associated with illness | . | . | 3,990,469,523 | 26,778 |
| | Total cost (excl. lost productivity) | 1,846,366,197 | 9,331 | 1,209,822,733 | 6,114 |
| Developmental and behavioural disorders | Primary sector | 34,441,953 | 262 | 12,674,301 | 96 |
| | Hospital care | 414,139,550 | 3,145 | 310,106,242 | 2,355 |
| | Filled prescriptions | 59,018,408 | 448 | 48,529,066 | 369 |
| | Home care | 4,812,782 | 37 | 3,498,199 | 27 |
| | Nursing home / sheltered accomodation | 10,328,222 | 78 | 8,152,647 | 62 |
| | Lost productivity associated with illness | . | . | 1,442,273,414 | 17,964 |
| | Total cost (excl. lost productivity) | 522,740,915 | 3,970 | 382,960,456 | 2,908 |
| Drug abuse | Primary sector | 19,319,591 | 353 | 6,250,481 | 114 |
| | Hospital care | 423,965,982 | 7,751 | 357,663,504 | 6,538 |
| | Filled prescriptions | 34,180,385 | 625 | 25,232,505 | 461 |

***Sensitivity analysis 1 / Prevalent cohort 2018***

| *Brain disorder* | *Cost component* | *Actual costs (EUR)* | *Actual costs per person* | *Attributable costs (EUR)* | *Attributable costs per person* |
|---|---|---|---|---|---|
| | Home care | 18,322,711 | 335 | 14,850,833 | 271 |
| | Nursing home / sheltered accomodation | 55,569,282 | 1,016 | 45,598,697 | 834 |
| | Lost productivity associated with illness | . | . | 1,753,730,524 | 35,516 |
| | Total cost (excl. lost productivity) | 551,357,950 | 10,079 | 449,596,019 | 8,219 |
| Eating disorders | Primary sector | 7,318,119 | 402 | 2,120,029 | 116 |
| | Hospital care | 113,902,664 | 6,252 | 88,815,971 | 4,875 |
| | Filled prescriptions | 5,183,680 | 285 | 2,606,255 | 143 |
| | Home care | 820,604 | 45 | 596,004 | 33 |
| | Nursing home / sheltered accomodation | 1,392,430 | 76 | 1,180,157 | 65 |
| | Lost productivity associated with illness | . | . | 104,528,921 | 6,247 |
| | Total cost (excl. lost productivity) | 128,617,497 | 7,060 | 95,318,416 | 5,232 |
| Epilepsy | Primary sector | 32,386,003 | 448 | 10,998,232 | 152 |
| | Hospital care | 317,022,357 | 4,383 | 192,457,837 | 2,661 |
| | Filled prescriptions | 55,553,373 | 768 | 38,452,285 | 532 |
| | Home care | 63,348,912 | 876 | 51,085,654 | 706 |
| | Nursing home / sheltered accomodation | 216,343,387 | 2,991 | 174,057,781 | 2,406 |
| | Lost productivity associated with illness | . | . | 904,746,942 | 20,057 |
| | Total cost (excl. lost productivity) | 684,654,032 | 9,465 | 467,051,789 | 6,457 |
| Headache | Primary sector | 51,105,283 | 441 | 16,124,248 | 139 |
| | Hospital care | 311,160,003 | 2,685 | 125,767,439 | 1,085 |
| | Filled prescriptions | 43,658,452 | 377 | 17,896,439 | 154 |
| | Home care | 17,204,650 | 148 | 4,540,270 | 39 |
| | Nursing home / sheltered accomodation | 47,233,606 | 408 | 9,783,093 | 84 |
| | Lost productivity associated with illness | . | . | 641,589,172 | 7,106 |
| | Total cost (excl. lost productivity) | 470,361,994 | 4,059 | 174,111,488 | 1,502 |
| Infections of the CNS | Primary sector | 10,572,212 | 370 | 2,243,835 | 79 |
| | Hospital care | 83,045,424 | 2,907 | 35,044,413 | 1,227 |
| | Filled prescriptions | 10,108,345 | 354 | 3,326,915 | 116 |
| | Home care | 10,877,046 | 381 | 6,382,030 | 223 |
| | Nursing home / sheltered accomodation | 29,837,997 | 1,045 | 15,180,579 | 531 |
| | Lost productivity associated with illness | . | . | 39,920,221 | 2,320 |
| | Total cost (excl. lost productivity) | 144,441,025 | 5,056 | 62,177,772 | 2,177 |
| Intellectual disability | Primary sector | 12,428,024 | 414 | 5,917,646 | 197 |
| | Hospital care | 132,991,928 | 4,433 | 97,243,080 | 3,242 |
| | Filled prescriptions | 21,295,618 | 710 | 17,075,564 | 569 |
| | Home care | 10,418,040 | 347 | 9,269,623 | 309 |
| | Nursing home / sheltered accomodation | 31,905,742 | 1,064 | 28,934,170 | 965 |
| | Lost productivity associated with illness | . | . | 709,706,666 | 34,305 |
| | Total cost (excl. lost productivity) | 209,039,353 | 6,968 | 158,440,083 | 5,282 |

***Sensitivity analysis 1 / Prevalent cohort 2018***

| *Brain disorder* | *Cost component* | *Actual costs (EUR)* | *Actual costs per person* | *Attributable costs (EUR)* | *Attributable costs per person* |
|---|---|---|---|---|---|
| Multiple sclerosis | Primary sector | 12,896,279 | 799 | 7,402,099 | 458 |
| | Hospital care | 52,727,508 | 3,265 | 23,438,271 | 1,451 |
| | Filled prescriptions | 9,744,677 | 603 | 5,355,018 | 332 |
| | Home care | 49,512,601 | 3,066 | 47,660,112 | 2,951 |
| | Nursing home / sheltered accomodation | 35,798,623 | 2,217 | 30,784,193 | 1,906 |
| | Lost productivity associated with illness | . | . | 240,059,611 | 19,229 |
| | Total cost (excl. lost productivity) | 160,679,687 | 9,949 | 114,639,693 | 7,099 |
| Neuromuscular disorders | Primary sector | 4,764,141 | 588 | 2,142,161 | 265 |
| | Hospital care | 43,334,226 | 5,352 | 27,823,381 | 3,436 |
| | Filled prescriptions | 5,106,274 | 631 | 2,807,728 | 347 |
| | Home care | 8,803,527 | 1,087 | 7,031,616 | 868 |
| | Nursing home / sheltered accomodation | 11,622,769 | 1,435 | 6,132,038 | 757 |
| | Lost productivity associated with illness | . | . | 83,340,904 | 16,946 |
| | Total cost (excl. lost productivity) | 73,630,937 | 9,094 | 45,936,925 | 5,673 |
| Other neurodegenerative disorders | Primary sector | 2,255,326 | 862 | 1,289,717 | 493 |
| | Hospital care | 18,950,656 | 7,245 | 13,084,363 | 5,002 |
| | Filled prescriptions | 2,768,930 | 1,059 | 1,841,133 | 704 |
| | Home care | 10,836,396 | 4,143 | 10,078,582 | 3,853 |
| | Nursing home / sheltered accomodation | 19,832,204 | 7,582 | 17,652,162 | 6,749 |
| | Lost productivity associated with illness | . | . | 43,450,408 | 34,025 |
| | Total cost (excl. lost productivity) | 54,643,512 | 20,890 | 43,945,958 | 16,801 |
| Parkinson's disease | Primary sector | 10,968,735 | 1,063 | 5,921,132 | 574 |
| | Hospital care | 73,131,490 | 7,090 | 37,793,915 | 3,664 |
| | Filled prescriptions | 30,931,226 | 2,999 | 25,631,266 | 2,485 |
| | Home care | 41,402,846 | 4,014 | 34,747,332 | 3,369 |
| | Nursing home / sheltered accomodation | 146,658,238 | 14,219 | 121,424,336 | 11,773 |
| | Lost productivity associated with illness | . | . | 39,077,692 | 24,655 |
| | Total cost (excl. lost productivity) | 303,092,535 | 29,386 | 225,517,982 | 21,865 |
| Personality disorders | Primary sector | 33,642,105 | 449 | 12,305,867 | 164 |
| | Hospital care | 446,383,220 | 5,963 | 343,533,728 | 4,589 |
| | Filled prescriptions | 38,802,761 | 518 | 25,263,779 | 337 |
| | Home care | 14,255,503 | 190 | 10,295,405 | 138 |
| | Nursing home / sheltered accomodation | 45,041,300 | 602 | 36,389,628 | 486 |
| | Lost productivity associated with illness | . | . | 2,136,089,726 | 30,962 |
| | Total cost (excl. lost productivity) | 578,124,890 | 7,723 | 427,788,408 | 5,715 |
| Polyneuropathy | Primary sector | 24,272,920 | 631 | 8,364,047 | 218 |
| | Hospital care | 238,981,435 | 6,216 | 132,872,595 | 3,456 |
| | Filled prescriptions | 35,012,746 | 911 | 19,064,446 | 496 |
| | Home care | 45,768,699 | 1,190 | 28,583,768 | 743 |

***Sensitivity analysis 1 / Prevalent cohort 2018***

| *Brain disorder* | *Cost component* | *Actual costs (EUR)* | *Actual costs per person* | *Attributable costs (EUR)* | *Attributable costs per person* |
|---|---|---|---|---|---|
| | Nursing home / sheltered accomodation | 112,979,243 | 2,939 | 46,837,980 | 1,218 |
| | Lost productivity associated with illness | . | . | 318,693,163 | 20,542 |
| | Total cost (excl. lost productivity) | 457,015,042 | 11,887 | 235,722,836 | 6,131 |
| Schizophrenia spectrum disorders | Primary sector | 25,769,877 | 394 | 7,481,320 | 114 |
| | Hospital care | 663,285,873 | 10,142 | 568,298,660 | 8,690 |
| | Filled prescriptions | 49,977,095 | 764 | 36,521,716 | 558 |
| | Home care | 30,499,810 | 466 | 23,673,425 | 362 |
| | Nursing home / sheltered accomodation | 151,166,569 | 2,312 | 129,483,721 | 1,980 |
| | Lost productivity associated with illness | . | . | 2,208,142,318 | 40,705 |
| | Total cost (excl. lost productivity) | 920,699,224 | 14,079 | 765,458,842 | 11,705 |
| Sleep disorders | Primary sector | 48,051,292 | 480 | 16,537,932 | 165 |
| | Hospital care | 358,949,626 | 3,582 | 155,807,184 | 1,555 |
| | Filled prescriptions | 64,991,380 | 649 | 36,342,151 | 363 |
| | Home care | 28,792,614 | 287 | 13,246,720 | 132 |
| | Nursing home / sheltered accomodation | 52,408,430 | 523 | 5,360,580 | 54 |
| | Lost productivity associated with illness | . | . | 673,938,472 | 10,929 |
| | Total cost (excl. lost productivity) | 553,193,342 | 5,521 | 227,294,567 | 2,269 |
| Stress related disorders | Primary sector | 95,942,081 | 416 | 32,077,477 | 139 |
| | Hospital care | 1,023,783,444 | 4,441 | 710,009,694 | 3,080 |
| | Filled prescriptions | 95,082,557 | 412 | 51,139,678 | 222 |
| | Home care | 49,220,692 | 214 | 31,853,990 | 138 |
| | Nursing home / sheltered accomodation | 163,095,592 | 707 | 115,372,526 | 500 |
| | Lost productivity associated with illness | . | . | 4,585,962,832 | 23,926 |
| | Total cost (excl. lost productivity) | 1,427,124,366 | 6,190 | 940,453,364 | 4,079 |
| Stroke | Primary sector | 73,857,640 | 607 | 19,973,173 | 164 |
| | Hospital care | 662,718,763 | 5,447 | 303,721,813 | 2,496 |
| | Filled prescriptions | 95,413,657 | 784 | 40,749,119 | 335 |
| | Home care | 218,681,150 | 1,797 | 151,957,416 | 1,249 |
| | Nursing home / sheltered accomodation | 780,648,306 | 6,416 | 518,795,761 | 4,264 |
| | Lost productivity associated with illness | . | . | 765,078,403 | 20,895 |
| | Total cost (excl. lost productivity) | 1,831,319,515 | 15,052 | 1,035,197,282 | 8,509 |
| Traumatic brain injury | Primary sector | 77,897,065 | 323 | 15,355,652 | 64 |
| | Hospital care | 630,144,399 | 2,614 | 276,101,637 | 1,145 |
| | Filled prescriptions | 71,465,571 | 296 | 24,076,915 | 100 |
| | Home care | 77,949,648 | 323 | 43,422,757 | 180 |
| | Nursing home / sheltered accomodation | 325,058,168 | 1,349 | 201,331,909 | 835 |
| | Lost productivity associated with illness | . | . | 1,320,599,854 | 8,018 |
| | Total cost (excl. lost productivity) | 1,182,514,852 | 4,906 | 560,288,870 | 2,324 |

***Sensitivity analysis 1 / Prevalent cohort 2019***

| *Brain disorder* | *Cost component* | *Actual costs (EUR)* | *Actual costs per person* | *Attributable costs (EUR)* | *Attributable costs per person* |
|---|---|---|---|---|---|
| Any brain disorder | Primary sector | 495,074,012 | 412 | 163,498,812 | 136 |
| | Hospital care | 4,545,426,691 | 3,786 | 2,601,973,271 | 2,167 |
| | Filled prescriptions | 574,350,436 | 478 | 322,789,575 | 269 |
| | Home care | 675,854,496 | 563 | 524,700,320 | 437 |
| | Nursing home / sheltered accomodation | 2,418,115,399 | 2,014 | 2,091,862,134 | 1,742 |
| | Lost productivity associated with illness | . | . | 15,002,993,946 | 19,203 |
| | Total cost (excl. lost productivity) | 8,708,821,034 | 7,254 | 5,704,824,112 | 4,752 |
| Alcohol abuse | Primary sector | 40,757,650 | 426 | 9,444,790 | 99 |
| | Hospital care | 733,601,026 | 7,667 | 514,700,553 | 5,379 |
| | Filled prescriptions | 64,058,894 | 669 | 33,522,831 | 350 |
| | Home care | 83,844,255 | 876 | 66,970,615 | 700 |
| | Nursing home / sheltered accomodation | 314,431,463 | 3,286 | 273,429,403 | 2,858 |
| | Lost productivity associated with illness | . | . | 2,415,842,721 | 38,468 |
| | Total cost (excl. lost productivity) | 1,236,693,288 | 12,925 | 898,068,192 | 9,386 |
| Anxiety disorders | Primary sector | 56,717,871 | 458 | 21,003,105 | 170 |
| | Hospital care | 606,594,255 | 4,897 | 395,544,667 | 3,193 |
| | Filled prescriptions | 59,278,503 | 479 | 33,456,756 | 270 |
| | Home care | 37,882,366 | 306 | 23,951,553 | 193 |
| | Nursing home / sheltered accomodation | 117,952,207 | 952 | 80,294,486 | 648 |
| | Lost productivity associated with illness | . | . | 2,400,036,455 | 23,554 |
| | Total cost (excl. lost productivity) | 878,425,203 | 7,091 | 554,250,568 | 4,474 |
| Bipolar disorder | Primary sector | 13,880,330 | 522 | 5,115,623 | 193 |
| | Hospital care | 232,446,164 | 8,750 | 175,707,574 | 6,614 |
| | Filled prescriptions | 20,760,034 | 781 | 13,200,694 | 497 |
| | Home care | 16,794,774 | 632 | 12,015,913 | 452 |
| | Nursing home / sheltered accomodation | 67,436,340 | 2,538 | 54,345,160 | 2,046 |
| | Lost productivity associated with illness | . | . | 668,130,527 | 33,524 |
| | Total cost (excl. lost productivity) | 351,317,642 | 13,224 | 260,384,964 | 9,801 |
| Brain tumors | Primary sector | 9,368,759 | 532 | 2,462,769 | 140 |
| | Hospital care | 119,907,917 | 6,811 | 71,015,907 | 4,034 |
| | Filled prescriptions | 10,730,418 | 610 | 3,668,127 | 208 |
| | Home care | 15,827,764 | 899 | 8,869,198 | 504 |
| | Nursing home / sheltered accomodation | 41,350,699 | 2,349 | 19,723,207 | 1,120 |

***Sensitivity analysis 1 / Prevalent cohort 2019***

| *Brain disorder* | *Cost component* | *Actual costs (EUR)* | *Actual costs per person* | *Attributable costs (EUR)* | *Attributable costs per person* |
|---|---|---|---|---|---|
| | Lost productivity associated with illness | . | . | 91,036,076 | 10,921 |
| | Total cost (excl. lost productivity) | 197,185,557 | 11,201 | 105,739,209 | 6,006 |
| Cerebral palsy | Primary sector | 6,792,338 | 723 | 4,788,060 | 509 |
| | Hospital care | 40,595,806 | 4,318 | 28,477,461 | 3,029 |
| | Filled prescriptions | 7,624,181 | 811 | 6,248,096 | 665 |
| | Home care | 14,400,185 | 1,532 | 13,876,546 | 1,476 |
| | Nursing home / sheltered accomodation | 14,135,402 | 1,504 | 12,750,345 | 1,356 |
| | Lost productivity associated with illness | . | . | 164,320,670 | 29,731 |
| | Total cost (excl. lost productivity) | 83,547,912 | 8,887 | 66,140,508 | 7,036 |
| Dementia | Primary sector | 23,117,109 | 629 | 3,667,239 | 100 |
| | Hospital care | 172,271,706 | 4,686 | 16,018,318 | 436 |
| | Filled prescriptions | 34,941,804 | 950 | 11,769,269 | 320 |
| | Home care | 129,228,696 | 3,515 | 77,921,814 | 2,120 |
| | Nursing home / sheltered accomodation | 1,266,879,046 | 34,461 | 1,130,208,956 | 30,743 |
| | Lost productivity associated with illness | . | . | 129,874,461 | 43,205 |
| | Total cost (excl. lost productivity) | 1,626,438,361 | 44,242 | 1,239,585,596 | 33,719 |
| Depression | Primary sector | 99,580,005 | 492 | 34,168,180 | 169 |
| | Hospital care | 1,054,991,894 | 5,215 | 649,115,612 | 3,209 |
| | Filled prescriptions | 117,884,915 | 583 | 64,356,500 | 318 |
| | Home care | 128,162,254 | 634 | 83,240,160 | 411 |
| | Nursing home / sheltered accomodation | 516,829,754 | 2,555 | 381,392,051 | 1,885 |
| | Lost productivity associated with illness | . | . | 4,156,483,484 | 27,116 |
| | Total cost (excl. lost productivity) | 1,917,448,822 | 9,479 | 1,212,272,502 | 5,993 |
| Developmental and behavioural disorders | Primary sector | 38,040,650 | 271 | 14,118,558 | 101 |
| | Hospital care | 449,769,678 | 3,203 | 323,347,332 | 2,303 |
| | Filled prescriptions | 58,433,140 | 416 | 46,347,518 | 330 |
| | Home care | 5,992,198 | 43 | 4,219,802 | 30 |
| | Nursing home / sheltered accomodation | 11,744,388 | 84 | 9,042,164 | 64 |
| | Lost productivity associated with illness | . | . | 1,626,635,883 | 18,565 |
| | Total cost (excl. lost productivity) | 563,980,054 | 4,017 | 397,075,375 | 2,828 |
| Drug abuse | Primary sector | 19,789,924 | 352 | 6,326,591 | 113 |
| | Hospital care | 471,099,178 | 8,389 | 391,243,417 | 6,967 |
| | Filled prescriptions | 33,728,109 | 601 | 24,119,731 | 430 |

***Sensitivity analysis 1 / Prevalent cohort 2019***

| *Brain disorder* | *Cost component* | *Actual costs (EUR)* | *Actual costs per person* | *Attributable costs (EUR)* | *Attributable costs per person* |
|---|---|---|---|---|---|
| | Home care | 20,053,900 | 357 | 15,913,293 | 283 |
| | Nursing home / sheltered accomodation | 54,222,223 | 966 | 44,712,056 | 796 |
| | Lost productivity associated with illness | . | . | 1,853,830,743 | 36,445 |
| | Total cost (excl. lost productivity) | 598,893,333 | 10,665 | 482,315,088 | 8,589 |
| Eating disorders | Primary sector | 7,690,183 | 411 | 2,269,033 | 121 |
| | Hospital care | 126,872,432 | 6,780 | 96,893,385 | 5,178 |
| | Filled prescriptions | 5,459,941 | 292 | 2,742,406 | 147 |
| | Home care | 964,445 | 52 | 608,331 | 33 |
| | Nursing home / sheltered accomodation | 1,347,059 | 72 | 1,050,035 | 56 |
| | Lost productivity associated with illness | . | . | 112,998,470 | 6,567 |
| | Total cost (excl. lost productivity) | 142,334,060 | 7,606 | 103,563,191 | 5,534 |
| Epilepsy | Primary sector | 32,257,004 | 451 | 11,062,092 | 155 |
| | Hospital care | 341,737,537 | 4,783 | 198,585,932 | 2,780 |
| | Filled prescriptions | 61,114,254 | 855 | 43,002,491 | 602 |
| | Home care | 73,263,797 | 1,025 | 59,514,410 | 833 |
| | Nursing home / sheltered accomodation | 211,368,532 | 2,958 | 168,821,653 | 2,363 |
| | Lost productivity associated with illness | . | . | 927,561,692 | 20,846 |
| | Total cost (excl. lost productivity) | 719,741,124 | 10,074 | 480,986,578 | 6,732 |
| Headache | Primary sector | 53,354,813 | 441 | 16,592,790 | 137 |
| | Hospital care | 382,548,408 | 3,160 | 157,685,264 | 1,302 |
| | Filled prescriptions | 46,705,249 | 386 | 18,001,609 | 149 |
| | Home care | 20,511,079 | 169 | 5,525,956 | 46 |
| | Nursing home / sheltered accomodation | 49,965,953 | 413 | 10,934,086 | 90 |
| | Lost productivity associated with illness | . | . | 671,740,513 | 7,108 |
| | Total cost (excl. lost productivity) | 553,085,502 | 4,568 | 208,739,704 | 1,724 |
| Infections of the CNS | Primary sector | 10,872,925 | 370 | 2,311,049 | 79 |
| | Hospital care | 101,306,803 | 3,451 | 43,991,630 | 1,499 |
| | Filled prescriptions | 11,162,883 | 380 | 3,703,720 | 126 |
| | Home care | 12,195,677 | 415 | 6,942,723 | 237 |
| | Nursing home / sheltered accomodation | 29,743,474 | 1,013 | 14,165,021 | 483 |
| | Lost productivity associated with illness | . | . | 40,605,928 | 2,302 |
| | Total cost (excl. lost productivity) | 165,281,762 | 5,630 | 71,114,142 | 2,423 |
| Intellectual disability | Primary sector | 12,917,146 | 424 | 6,221,417 | 204 |
| | Hospital care | 141,934,339 | 4,659 | 101,450,809 | 3,330 |
| | Filled prescriptions | 23,061,708 | 757 | 18,500,927 | 607 |
| | Home care | 11,994,914 | 394 | 10,610,541 | 348 |
| | Nursing home / sheltered accomodation | 32,176,288 | 1,056 | 28,966,680 | 951 |
| | Lost productivity associated with illness | . | . | 747,955,821 | 34,797 |
| | Total cost (excl. lost productivity) | 222,084,396 | 7,290 | 165,750,374 | 5,441 |

***Sensitivity analysis 1 / Prevalent cohort 2019***

| *Brain disorder* | *Cost component* | *Actual costs (EUR)* | *Actual costs per person* | *Attributable costs (EUR)* | *Attributable costs per person* |
|---|---|---|---|---|---|
| Multiple sclerosis | Primary sector | 13,095,724 | 800 | 7,539,775 | 461 |
| | Hospital care | 113,935,899 | 6,964 | 79,381,459 | 4,852 |
| | Filled prescriptions | 10,232,810 | 625 | 5,459,566 | 334 |
| | Home care | 57,231,639 | 3,498 | 55,124,212 | 3,369 |
| | Nursing home / sheltered accomodation | 35,586,891 | 2,175 | 30,230,593 | 1,848 |
| | Lost productivity associated with illness | . | . | 254,059,590 | 20,234 |
| | Total cost (excl. lost productivity) | 230,082,963 | 14,064 | 177,735,605 | 10,864 |
| Neuromuscular disorders | Primary sector | 4,922,963 | 599 | 2,246,813 | 274 |
| | Hospital care | 48,459,643 | 5,901 | 30,198,958 | 3,677 |
| | Filled prescriptions | 5,436,278 | 662 | 2,958,835 | 360 |
| | Home care | 10,730,744 | 1,307 | 9,011,039 | 1,097 |
| | Nursing home / sheltered accomodation | 11,941,626 | 1,454 | 6,471,262 | 788 |
| | Lost productivity associated with illness | . | . | 81,639,311 | 16,433 |
| | Total cost (excl. lost productivity) | 81,491,255 | 9,923 | 50,886,906 | 6,196 |
| Other neurodegenerative disorders | Primary sector | 2,239,875 | 846 | 1,255,248 | 474 |
| | Hospital care | 19,665,410 | 7,427 | 12,347,919 | 4,664 |
| | Filled prescriptions | 2,411,357 | 911 | 1,409,403 | 532 |
| | Home care | 11,776,137 | 4,448 | 11,079,329 | 4,184 |
| | Nursing home / sheltered accomodation | 19,935,339 | 7,529 | 17,594,032 | 6,645 |
| | Lost productivity associated with illness | . | . | 43,033,217 | 34,126 |
| | Total cost (excl. lost productivity) | 56,028,118 | 21,161 | 43,685,931 | 16,499 |
| Parkinson’s disease | Primary sector | 11,249,123 | 1,080 | 6,237,034 | 599 |
| | Hospital care | 77,539,118 | 7,442 | 35,848,208 | 3,441 |
| | Filled prescriptions | 33,063,987 | 3,173 | 27,263,188 | 2,617 |
| | Home care | 46,940,769 | 4,505 | 39,251,186 | 3,767 |
| | Nursing home / sheltered accomodation | 149,545,603 | 14,353 | 124,277,655 | 11,928 |
| | Lost productivity associated with illness | . | . | 41,107,128 | 26,385 |
| | Total cost (excl. lost productivity) | 318,338,600 | 30,554 | 232,877,270 | 22,351 |
| Personality disorders | Primary sector | 33,815,104 | 451 | 12,263,434 | 163 |
| | Hospital care | 482,110,489 | 6,424 | 360,346,802 | 4,801 |
| | Filled prescriptions | 38,555,775 | 514 | 24,295,948 | 324 |
| | Home care | 17,147,855 | 228 | 12,898,962 | 172 |
| | Nursing home / sheltered accomodation | 43,866,693 | 584 | 35,316,343 | 471 |
| | Lost productivity associated with illness | . | . | 2,199,066,934 | 31,703 |
| | Total cost (excl. lost productivity) | 615,495,917 | 8,201 | 445,121,488 | 5,931 |
| Polyneuropathy | Primary sector | 24,773,321 | 623 | 8,437,621 | 212 |
| | Hospital care | 293,964,675 | 7,397 | 167,936,770 | 4,226 |
| | Filled prescriptions | 38,239,584 | 962 | 20,410,306 | 514 |
| | Home care | 55,610,277 | 1,399 | 35,214,276 | 886 |

*Sensitivity analysis 1 / Prevalent cohort 2019*

| *Brain disorder* | *Cost component* | *Actual costs (EUR)* | *Actual costs per person* | *Attributable costs (EUR)* | *Attributable costs per person* |
|---|---|---|---|---|---|
| | Nursing home / sheltered accomodation | 119,388,118 | 3,004 | 49,106,312 | 1,236 |
| | Lost productivity associated with illness | . | . | 332,983,353 | 21,246 |
| | Total cost (excl. lost productivity) | 531,975,975 | 13,386 | 281,105,285 | 7,074 |
| Schizophrenia spectrum disorders | Primary sector | 26,218,851 | 392 | 7,568,236 | 113 |
| | Hospital care | 748,156,431 | 11,181 | 637,426,346 | 9,526 |
| | Filled prescriptions | 52,071,729 | 778 | 37,696,218 | 563 |
| | Home care | 34,786,593 | 520 | 26,838,808 | 401 |
| | Nursing home / sheltered accomodation | 147,669,845 | 2,207 | 127,249,541 | 1,902 |
| | Lost productivity associated with illness | . | . | 2,301,619,956 | 41,307 |
| | Total cost (excl. lost productivity) | 1,008,903,449 | 15,078 | 836,779,148 | 12,505 |
| Sleep disorders | Primary sector | 51,472,027 | 470 | 17,117,647 | 156 |
| | Hospital care | 453,634,773 | 4,139 | 196,882,401 | 1,796 |
| | Filled prescriptions | 75,372,317 | 688 | 41,770,113 | 381 |
| | Home care | 37,162,900 | 339 | 16,959,140 | 155 |
| | Nursing home / sheltered accomodation | 58,521,132 | 534 | 5,737,964 | 52 |
| | Lost productivity associated with illness | . | . | 776,290,344 | 11,561 |
| | Total cost (excl. lost productivity) | 676,163,148 | 6,169 | 278,467,266 | 2,540 |
| Stress related disorders | Primary sector | 100,138,553 | 418 | 33,493,652 | 140 |
| | Hospital care | 1,128,853,386 | 4,707 | 747,721,653 | 3,118 |
| | Filled prescriptions | 100,442,361 | 419 | 51,809,982 | 216 |
| | Home care | 54,982,408 | 229 | 34,915,184 | 146 |
| | Nursing home / sheltered accomodation | 166,099,746 | 693 | 118,415,353 | 494 |
| | Lost productivity associated with illness | . | . | 4,855,383,041 | 24,359 |
| | Total cost (excl. lost productivity) | 1,550,516,455 | 6,466 | 986,355,824 | 4,113 |
| Stroke | Primary sector | 73,797,161 | 599 | 19,756,670 | 160 |
| | Hospital care | 716,324,131 | 5,814 | 295,242,088 | 2,396 |
| | Filled prescriptions | 102,014,507 | 828 | 41,919,781 | 340 |
| | Home care | 250,600,009 | 2,034 | 175,021,888 | 1,421 |
| | Nursing home / sheltered accomodation | 778,256,871 | 6,317 | 516,499,312 | 4,192 |
| | Lost productivity associated with illness | . | . | 807,300,752 | 22,025 |
| | Total cost (excl. lost productivity) | 1,920,992,679 | 15,593 | 1,048,439,739 | 8,510 |
| Traumatic brain injury | Primary sector | 78,156,172 | 328 | 15,566,721 | 65 |
| | Hospital care | 683,939,216 | 2,867 | 277,548,723 | 1,163 |
| | Filled prescriptions | 74,523,148 | 312 | 24,096,354 | 101 |
| | Home care | 91,775,682 | 385 | 52,854,023 | 222 |
| | Nursing home / sheltered accomodation | 327,836,937 | 1,374 | 204,907,746 | 859 |
| | Lost productivity associated with illness | . | . | 1,327,346,293 | 8,194 |
| | Total cost (excl. lost productivity) | 1,256,231,154 | 5,266 | 574,973,567 | 2,410 |

***Sensitivity analysis 1 / Prevalent cohort 2020***

| *Brain disorder* | *Cost component* | *Actual costs (EUR)* | *Actual costs per person* | *Attributable costs (EUR)* | *Attributable costs per person* |
|---|---|---|---|---|---|
| Any brain disorder | Primary sector | 505,522,130 | 414 | 163,643,730 | 134 |
| | Hospital care | 4,822,841,224 | 3,946 | 2,747,709,783 | 2,248 |
| | Filled prescriptions | 602,047,245 | 493 | 338,459,855 | 277 |
| | Home care | 756,927,847 | 619 | 586,093,768 | 480 |
| | Nursing home / sheltered accomodation | 2,422,006,390 | 1,982 | 2,096,028,831 | 1,715 |
| | Lost productivity associated with illness | . | . | 16,009,198,147 | 20,159 |
| | Total cost (excl. lost productivity) | 9,109,344,836 | 7,454 | 5,931,935,967 | 4,854 |
| Alcohol abuse | Primary sector | 39,706,134 | 424 | 8,788,785 | 94 |
| | Hospital care | 746,047,452 | 7,970 | 517,372,503 | 5,527 |
| | Filled prescriptions | 63,151,078 | 675 | 32,340,577 | 345 |
| | Home care | 97,224,684 | 1,039 | 78,805,645 | 842 |
| | Nursing home / sheltered accomodation | 313,229,759 | 3,346 | 271,579,789 | 2,901 |
| | Lost productivity associated with illness | . | . | 2,445,512,306 | 40,365 |
| | Total cost (excl. lost productivity) | 1,259,359,107 | 13,454 | 908,887,299 | 9,710 |
| Anxiety disorders | Primary sector | 58,291,592 | 454 | 21,043,862 | 164 |
| | Hospital care | 641,005,430 | 4,992 | 411,717,510 | 3,206 |
| | Filled prescriptions | 62,953,051 | 490 | 35,573,626 | 277 |
| | Home care | 43,603,263 | 340 | 28,015,810 | 218 |
| | Nursing home / sheltered accomodation | 119,822,924 | 933 | 81,562,060 | 635 |
| | Lost productivity associated with illness | . | . | 2,610,558,944 | 24,675 |
| | Total cost (excl. lost productivity) | 925,676,260 | 7,209 | 577,912,869 | 4,501 |
| Bipolar disorder | Primary sector | 14,245,873 | 523 | 5,231,921 | 192 |
| | Hospital care | 244,625,207 | 8,976 | 184,175,698 | 6,758 |
| | Filled prescriptions | 22,657,895 | 831 | 14,834,731 | 544 |
| | Home care | 18,954,277 | 696 | 13,855,162 | 508 |
| | Nursing home / sheltered accomodation | 66,921,084 | 2,456 | 54,168,645 | 1,988 |
| | Lost productivity associated with illness | . | . | 712,854,906 | 34,675 |
| | Total cost (excl. lost productivity) | 367,404,336 | 13,482 | 272,266,157 | 9,991 |
| Brain tumors | Primary sector | 9,856,721 | 535 | 2,489,511 | 135 |
| | Hospital care | 129,386,568 | 7,023 | 76,105,017 | 4,131 |
| | Filled prescriptions | 11,664,009 | 633 | 4,068,806 | 221 |
| | Home care | 17,580,223 | 954 | 9,391,562 | 510 |
| | Nursing home / sheltered accomodation | 43,668,615 | 2,370 | 20,767,168 | 1,127 |

***Sensitivity analysis 1 / Prevalent cohort 2020***

| *Brain disorder* | *Cost component* | *Actual costs (EUR)* | *Actual costs per person* | *Attributable costs (EUR)* | *Attributable costs per person* |
|---|---|---|---|---|---|
| | Lost productivity associated with illness | . | . | 94,128,349 | 10,929 |
| | Total cost (excl. lost productivity) | 212,156,136 | 11,516 | 112,822,064 | 6,124 |
| Cerebral palsy | Primary sector | 6,169,094 | 661 | 4,154,273 | 445 |
| | Hospital care | 38,091,301 | 4,079 | 25,368,388 | 2,717 |
| | Filled prescriptions | 8,170,309 | 875 | 6,736,395 | 721 |
| | Home care | 15,798,819 | 1,692 | 15,135,819 | 1,621 |
| | Nursing home / sheltered accomodation | 13,885,651 | 1,487 | 12,513,780 | 1,340 |
| | Lost productivity associated with illness | . | . | 171,280,918 | 31,057 |
| | Total cost (excl. lost productivity) | 82,115,174 | 8,794 | 63,908,654 | 6,844 |
| Dementia | Primary sector | 23,607,318 | 654 | 4,127,332 | 114 |
| | Hospital care | 179,539,066 | 4,977 | 18,710,574 | 519 |
| | Filled prescriptions | 37,427,467 | 1,038 | 14,198,138 | 394 |
| | Home care | 146,343,774 | 4,057 | 90,659,586 | 2,513 |
| | Nursing home / sheltered accomodation | 1,238,910,926 | 34,345 | 1,107,997,403 | 30,716 |
| | Lost productivity associated with illness | . | . | 130,166,509 | 44,823 |
| | Total cost (excl. lost productivity) | 1,625,828,550 | 45,071 | 1,235,693,033 | 34,256 |
| Depression | Primary sector | 100,156,284 | 488 | 33,622,685 | 164 |
| | Hospital care | 1,107,926,230 | 5,402 | 678,195,933 | 3,307 |
| | Filled prescriptions | 121,795,611 | 594 | 66,507,488 | 324 |
| | Home care | 143,011,346 | 697 | 93,901,250 | 458 |
| | Nursing home / sheltered accomodation | 492,849,037 | 2,403 | 361,513,589 | 1,763 |
| | Lost productivity associated with illness | . | . | 4,441,829,013 | 28,451 |
| | Total cost (excl. lost productivity) | 1,965,738,507 | 9,585 | 1,233,740,943 | 6,015 |
| Developmental and behavioural disorders | Primary sector | 41,222,481 | 277 | 14,885,602 | 100 |
| | Hospital care | 501,340,835 | 3,371 | 361,811,365 | 2,432 |
| | Filled prescriptions | 56,674,302 | 381 | 42,848,496 | 288 |
| | Home care | 6,224,062 | 42 | 4,246,153 | 29 |
| | Nursing home / sheltered accomodation | 13,012,258 | 87 | 10,213,301 | 69 |
| | Lost productivity associated with illness | . | . | 1,882,656,997 | 19,783 |
| | Total cost (excl. lost productivity) | 618,473,938 | 4,158 | 434,004,918 | 2,918 |
| Drug abuse | Primary sector | 20,187,653 | 351 | 6,433,158 | 112 |
| | Hospital care | 484,600,357 | 8,428 | 399,690,375 | 6,951 |
| | Filled prescriptions | 33,256,871 | 578 | 23,194,713 | 403 |

***Sensitivity analysis 1 / Prevalent cohort 2020***

| *Brain disorder* | *Cost component* | *Actual costs (EUR)* | *Actual costs per person* | *Attributable costs (EUR)* | *Attributable costs per person* |
|---|---|---|---|---|---|
| | Home care | 22,672,287 | 394 | 18,143,544 | 316 |
| | Nursing home / sheltered accomodation | 52,439,937 | 912 | 42,764,620 | 744 |
| | Lost productivity associated with illness | . | . | 1,969,949,782 | 37,758 |
| | Total cost (excl. lost productivity) | 613,157,105 | 10,663 | 490,226,411 | 8,525 |
| Eating disorders | Primary sector | 8,190,029 | 427 | 2,502,753 | 130 |
| | Hospital care | 138,766,605 | 7,232 | 106,576,793 | 5,555 |
| | Filled prescriptions | 5,673,053 | 296 | 2,663,847 | 139 |
| | Home care | 1,157,475 | 60 | 790,670 | 41 |
| | Nursing home / sheltered accomodation | 1,635,669 | 85 | 1,394,739 | 73 |
| | Lost productivity associated with illness | . | . | 123,912,388 | 7,056 |
| | Total cost (excl. lost productivity) | 155,422,833 | 8,100 | 113,928,802 | 5,938 |
| Epilepsy | Primary sector | 31,662,738 | 448 | 10,394,892 | 147 |
| | Hospital care | 349,889,796 | 4,951 | 201,253,447 | 2,848 |
| | Filled prescriptions | 67,960,740 | 962 | 49,437,916 | 700 |
| | Home care | 77,948,168 | 1,103 | 62,528,479 | 885 |
| | Nursing home / sheltered accomodation | 210,401,036 | 2,977 | 168,511,479 | 2,385 |
| | Lost productivity associated with illness | . | . | 947,912,706 | 21,743 |
| | Total cost (excl. lost productivity) | 737,862,478 | 10,441 | 492,126,214 | 6,964 |
| Headache | Primary sector | 55,999,070 | 447 | 17,640,918 | 141 |
| | Hospital care | 413,297,667 | 3,297 | 167,959,137 | 1,340 |
| | Filled prescriptions | 49,992,769 | 399 | 19,453,163 | 155 |
| | Home care | 25,133,902 | 201 | 7,629,050 | 61 |
| | Nursing home / sheltered accomodation | 53,196,542 | 424 | 12,991,567 | 104 |
| | Lost productivity associated with illness | . | . | 745,630,924 | 7,633 |
| | Total cost (excl. lost productivity) | 597,619,950 | 4,768 | 225,673,835 | 1,800 |
| Infections of the CNS | Primary sector | 11,299,209 | 376 | 2,440,002 | 81 |
| | Hospital care | 107,914,921 | 3,589 | 45,689,204 | 1,520 |
| | Filled prescriptions | 11,715,444 | 390 | 3,896,611 | 130 |
| | Home care | 14,125,646 | 470 | 7,792,495 | 259 |
| | Nursing home / sheltered accomodation | 32,695,325 | 1,087 | 16,416,198 | 546 |
| | Lost productivity associated with illness | . | . | 49,432,026 | 2,756 |
| | Total cost (excl. lost productivity) | 177,750,546 | 5,912 | 76,234,509 | 2,535 |
| Intellectual disability | Primary sector | 12,642,144 | 411 | 5,785,592 | 188 |
| | Hospital care | 154,701,141 | 5,026 | 112,194,630 | 3,645 |
| | Filled prescriptions | 24,203,736 | 786 | 19,473,266 | 633 |
| | Home care | 12,148,694 | 395 | 10,485,241 | 341 |
| | Nursing home / sheltered accomodation | 31,979,890 | 1,039 | 29,010,532 | 943 |
| | Lost productivity associated with illness | . | . | 791,405,272 | 35,810 |
| | Total cost (excl. lost productivity) | 235,675,606 | 7,657 | 176,949,262 | 5,749 |

***Sensitivity analysis 1 / Prevalent cohort 2020***

| *Brain disorder* | *Cost component* | *Actual costs (EUR)* | *Actual costs per person* | *Attributable costs (EUR)* | *Attributable costs per person* |
|---|---|---|---|---|---|
| Multiple sclerosis | Primary sector | 12,714,467 | 757 | 7,010,644 | 417 |
| | Hospital care | 122,232,986 | 7,275 | 84,537,895 | 5,031 |
| | Filled prescriptions | 10,499,177 | 625 | 5,541,921 | 330 |
| | Home care | 58,619,714 | 3,489 | 56,077,799 | 3,338 |
| | Nursing home / sheltered accomodation | 36,552,707 | 2,176 | 30,745,732 | 1,830 |
| | Lost productivity associated with illness | . | . | 265,403,104 | 20,657 |
| | Total cost (excl. lost productivity) | 240,619,052 | 14,321 | 183,913,990 | 10,946 |
| Neuromuscular disorders | Primary sector | 4,843,482 | 580 | 2,092,840 | 251 |
| | Hospital care | 52,611,576 | 6,304 | 32,881,188 | 3,940 |
| | Filled prescriptions | 5,573,527 | 668 | 2,968,406 | 356 |
| | Home care | 11,993,566 | 1,437 | 9,768,526 | 1,170 |
| | Nursing home / sheltered accomodation | 11,784,298 | 1,412 | 5,824,288 | 698 |
| | Lost productivity associated with illness | . | . | 88,920,253 | 17,777 |
| | Total cost (excl. lost productivity) | 86,806,449 | 10,401 | 53,535,248 | 6,415 |
| Other neurodegenerative disorders | Primary sector | 2,207,663 | 812 | 1,186,319 | 437 |
| | Hospital care | 19,406,435 | 7,142 | 11,519,021 | 4,239 |
| | Filled prescriptions | 2,431,548 | 895 | 1,372,364 | 505 |
| | Home care | 13,646,249 | 5,022 | 12,697,317 | 4,673 |
| | Nursing home / sheltered accomodation | 19,702,392 | 7,251 | 17,172,791 | 6,320 |
| | Lost productivity associated with illness | . | . | 44,145,878 | 34,543 |
| | Total cost (excl. lost productivity) | 57,394,286 | 21,123 | 43,947,812 | 16,174 |
| Parkinson’s disease | Primary sector | 11,290,093 | 1,071 | 6,135,543 | 582 |
| | Hospital care | 83,135,359 | 7,887 | 39,025,524 | 3,702 |
| | Filled prescriptions | 34,394,161 | 3,263 | 28,432,012 | 2,697 |
| | Home care | 56,638,628 | 5,373 | 48,082,935 | 4,562 |
| | Nursing home / sheltered accomodation | 155,515,261 | 14,754 | 130,491,549 | 12,380 |
| | Lost productivity associated with illness | . | . | 42,185,870 | 27,287 |
| | Total cost (excl. lost productivity) | 340,973,502 | 32,349 | 252,167,562 | 23,924 |
| Personality disorders | Primary sector | 33,975,590 | 451 | 12,182,256 | 162 |
| | Hospital care | 502,831,407 | 6,675 | 374,265,427 | 4,969 |
| | Filled prescriptions | 39,277,368 | 521 | 24,597,850 | 327 |
| | Home care | 18,438,288 | 245 | 13,608,511 | 181 |
| | Nursing home / sheltered accomodation | 41,239,372 | 547 | 32,910,551 | 437 |
| | Lost productivity associated with illness | . | . | 2,326,606,374 | 33,384 |
| | Total cost (excl. lost productivity) | 635,762,025 | 8,440 | 457,564,595 | 6,074 |
| Polyneuropathy | Primary sector | 26,044,024 | 625 | 8,604,015 | 207 |
| | Hospital care | 328,288,566 | 7,881 | 187,951,209 | 4,512 |
| | Filled prescriptions | 42,502,049 | 1,020 | 23,365,927 | 561 |
| | Home care | 66,337,024 | 1,593 | 42,022,162 | 1,009 |

***Sensitivity analysis 1 / Prevalent cohort 2020***

| *Brain disorder* | *Cost component* | *Actual costs (EUR)* | *Actual costs per person* | *Attributable costs (EUR)* | *Attributable costs per person* |
|---|---|---|---|---|---|
| | Nursing home / sheltered accomodation | 125,629,582 | 3,016 | 53,376,940 | 1,281 |
| | Lost productivity associated with illness | . | . | 360,702,801 | 22,426 |
| | Total cost (excl. lost productivity) | 588,801,244 | 14,135 | 315,320,253 | 7,570 |
| Schizophrenia spectrum disorders | Primary sector | 26,730,454 | 391 | 7,696,980 | 113 |
| | Hospital care | 795,722,386 | 11,630 | 678,553,990 | 9,918 |
| | Filled prescriptions | 53,372,949 | 780 | 38,387,817 | 561 |
| | Home care | 40,455,340 | 591 | 32,153,294 | 470 |
| | Nursing home / sheltered accomodation | 144,019,153 | 2,105 | 124,314,379 | 1,817 |
| | Lost productivity associated with illness | . | . | 2,436,526,103 | 42,697 |
| | Total cost (excl. lost productivity) | 1,060,300,282 | 15,498 | 881,106,458 | 12,878 |
| Sleep disorders | Primary sector | 55,307,141 | 468 | 17,786,835 | 151 |
| | Hospital care | 511,790,509 | 4,333 | 218,236,741 | 1,848 |
| | Filled prescriptions | 83,402,765 | 706 | 45,734,728 | 387 |
| | Home care | 43,291,926 | 367 | 18,444,395 | 156 |
| | Nursing home / sheltered accomodation | 66,789,592 | 565 | 7,868,424 | 67 |
| | Lost productivity associated with illness | . | . | 903,545,656 | 12,631 |
| | Total cost (excl. lost productivity) | 760,581,933 | 6,440 | 308,071,123 | 2,608 |
| Stress related disorders | Primary sector | 103,501,010 | 417 | 34,071,946 | 137 |
| | Hospital care | 1,222,075,122 | 4,923 | 810,957,052 | 3,267 |
| | Filled prescriptions | 106,128,467 | 427 | 54,489,922 | 219 |
| | Home care | 61,720,983 | 249 | 39,016,739 | 157 |
| | Nursing home / sheltered accomodation | 165,957,961 | 668 | 118,370,169 | 477 |
| | Lost productivity associated with illness | . | . | 5,291,058,843 | 25,702 |
| | Total cost (excl. lost productivity) | 1,659,383,544 | 6,684 | 1,056,905,828 | 4,257 |
| Stroke | Primary sector | 74,799,977 | 600 | 19,056,471 | 153 |
| | Hospital care | 773,669,068 | 6,201 | 325,105,032 | 2,606 |
| | Filled prescriptions | 106,000,850 | 850 | 43,549,998 | 349 |
| | Home care | 276,332,892 | 2,215 | 190,336,505 | 1,526 |
| | Nursing home / sheltered accomodation | 777,477,161 | 6,231 | 515,280,183 | 4,130 |
| | Lost productivity associated with illness | . | . | 830,653,727 | 22,702 |
| | Total cost (excl. lost productivity) | 2,008,279,948 | 16,096 | 1,093,328,190 | 8,763 |
| Traumatic brain injury | Primary sector | 78,620,857 | 333 | 15,562,017 | 66 |
| | Hospital care | 714,289,064 | 3,027 | 290,750,882 | 1,232 |
| | Filled prescriptions | 77,031,470 | 326 | 24,912,797 | 106 |
| | Home care | 101,951,726 | 432 | 56,818,095 | 241 |
| | Nursing home / sheltered accomodation | 334,168,093 | 1,416 | 209,592,089 | 888 |
| | Lost productivity associated with illness | . | . | 1,324,828,434 | 8,332 |
| | Total cost (excl. lost productivity) | 1,306,061,210 | 5,535 | 597,635,880 | 2,533 |

***Sensitivity analysis 1 / Prevalent cohort 2021***

| *Brain disorder* | *Cost component* | *Actual costs (EUR)* | *Actual costs per person* | *Attributable costs (EUR)* | *Attributable costs per person* |
|---|---|---|---|---|---|
| Any brain disorder | Primary sector | 526,583,120 | 425 | 170,984,512 | 138 |
| | Hospital care | 4,853,145,085 | 3,921 | 2,764,018,283 | 2,233 |
| | Filled prescriptions | 590,126,739 | 477 | 327,515,018 | 265 |
| | Home care | 850,190,539 | 687 | 660,586,391 | 534 |
| | Nursing home / sheltered accomodation | 2,380,786,553 | 1,924 | 2,054,698,560 | 1,660 |
| | Lost productivity associated with illness | . | . | 16,894,841,529 | 21,035 |
| | Total cost (excl. lost productivity) | 9,200,832,036 | 7,434 | 5,977,802,764 | 4,830 |
| Alcohol abuse | Primary sector | 39,668,863 | 435 | 8,683,992 | 95 |
| | Hospital care | 720,362,469 | 7,898 | 498,479,410 | 5,465 |
| | Filled prescriptions | 58,922,339 | 646 | 29,332,080 | 322 |
| | Home care | 102,773,040 | 1,127 | 82,232,190 | 902 |
| | Nursing home / sheltered accomodation | 310,084,241 | 3,400 | 268,033,581 | 2,939 |
| | Lost productivity associated with illness | . | . | 2,415,672,691 | 41,409 |
| | Total cost (excl. lost productivity) | 1,231,810,951 | 13,505 | 886,761,253 | 9,722 |
| Anxiety disorders | Primary sector | 61,685,680 | 465 | 21,896,276 | 165 |
| | Hospital care | 656,498,764 | 4,944 | 420,988,923 | 3,170 |
| | Filled prescriptions | 60,468,521 | 455 | 32,660,569 | 246 |
| | Home care | 48,828,721 | 368 | 30,571,888 | 230 |
| | Nursing home / sheltered accomodation | 123,680,936 | 931 | 84,850,443 | 639 |
| | Lost productivity associated with illness | . | . | 2,786,193,330 | 25,410 |
| | Total cost (excl. lost productivity) | 951,162,622 | 7,163 | 590,968,099 | 4,450 |
| Bipolar disorder | Primary sector | 14,731,866 | 530 | 5,338,914 | 192 |
| | Hospital care | 240,772,498 | 8,656 | 180,008,692 | 6,471 |
| | Filled prescriptions | 21,574,129 | 776 | 13,879,728 | 499 |
| | Home care | 20,915,704 | 752 | 15,269,431 | 549 |
| | Nursing home / sheltered accomodation | 64,740,961 | 2,327 | 52,667,087 | 1,893 |
| | Lost productivity associated with illness | . | . | 751,409,705 | 35,534 |
| | Total cost (excl. lost productivity) | 362,735,160 | 13,040 | 267,163,853 | 9,605 |
| Brain tumors | Primary sector | 10,321,932 | 541 | 2,514,106 | 132 |
| | Hospital care | 136,319,065 | 7,151 | 80,454,027 | 4,220 |
| | Filled prescriptions | 11,154,889 | 585 | 3,455,402 | 181 |
| | Home care | 21,286,988 | 1,117 | 11,836,998 | 621 |
| | Nursing home / sheltered accomodation | 43,788,764 | 2,297 | 19,922,797 | 1,045 |

*Sensitivity analysis 1 / Prevalent cohort 2021*

| Brain disorder | Cost component | Actual costs (EUR) | Actual costs per person | Attributable costs (EUR) | Attributable costs per person |
|---|---|---|---|---|---|
| | Lost productivity associated with illness | . | . | 101,096,230 | 11,443 |
| | Total cost (excl. lost productivity) | 222,871,638 | 11,691 | 118,183,331 | 6,199 |
| Cerebral palsy | Primary sector | 6,526,579 | 703 | 4,433,803 | 478 |
| | Hospital care | 39,715,357 | 4,279 | 27,067,968 | 2,916 |
| | Filled prescriptions | 8,172,995 | 881 | 6,814,448 | 734 |
| | Home care | 20,839,416 | 2,245 | 20,237,373 | 2,180 |
| | Nursing home / sheltered accomodation | 14,122,589 | 1,522 | 12,869,341 | 1,387 |
| | Lost productivity associated with illness | . | . | 178,819,263 | 32,365 |
| | Total cost (excl. lost productivity) | 89,376,936 | 9,630 | 71,422,933 | 7,695 |
| Dementia | Primary sector | 24,426,767 | 699 | 5,278,511 | 151 |
| | Hospital care | 168,102,118 | 4,812 | 14,250,168 | 408 |
| | Filled prescriptions | 34,790,501 | 996 | 13,069,909 | 374 |
| | Home care | 150,957,954 | 4,321 | 94,290,876 | 2,699 |
| | Nursing home / sheltered accomodation | 1,179,874,327 | 33,776 | 1,057,403,251 | 30,270 |
| | Lost productivity associated with illness | . | . | 126,935,480 | 46,445 |
| | Total cost (excl. lost productivity) | 1,558,151,666 | 44,605 | 1,184,292,715 | 33,903 |
| Depression | Primary sector | 102,808,212 | 497 | 33,979,810 | 164 |
| | Hospital care | 1,111,055,481 | 5,369 | 685,987,272 | 3,315 |
| | Filled prescriptions | 115,432,342 | 558 | 61,161,493 | 296 |
| | Home care | 149,046,599 | 720 | 96,625,295 | 467 |
| | Nursing home / sheltered accomodation | 464,967,878 | 2,247 | 343,317,855 | 1,659 |
| | Lost productivity associated with illness | . | . | 4,637,310,528 | 29,274 |
| | Total cost (excl. lost productivity) | 1,943,310,512 | 9,390 | 1,221,071,725 | 5,900 |
| Developmental and behavioural disorders | Primary sector | 45,092,914 | 287 | 16,164,909 | 103 |
| | Hospital care | 548,907,463 | 3,495 | 398,160,065 | 2,535 |
| | Filled prescriptions | 59,505,468 | 379 | 44,995,824 | 286 |
| | Home care | 11,433,192 | 73 | 8,340,697 | 53 |
| | Nursing home / sheltered accomodation | 13,720,761 | 87 | 11,031,098 | 70 |
| | Lost productivity associated with illness | . | . | 2,143,489,806 | 20,810 |
| | Total cost (excl. lost productivity) | 678,659,798 | 4,321 | 478,692,593 | 3,048 |
| Drug abuse | Primary sector | 20,867,594 | 356 | 6,366,968 | 109 |
| | Hospital care | 484,665,245 | 8,278 | 397,944,420 | 6,797 |
| | Filled prescriptions | 32,710,869 | 559 | 22,626,054 | 386 |

*Sensitivity analysis 1 / Prevalent cohort 2021*

| *Brain disorder* | *Cost component* | *Actual costs (EUR)* | *Actual costs per person* | *Attributable costs (EUR)* | *Attributable costs per person* |
|---|---|---|---|---|---|
| | Home care | 25,260,593 | 431 | 20,127,710 | 344 |
| | Nursing home / sheltered accomodation | 51,740,466 | 884 | 42,753,737 | 730 |
| | Lost productivity associated with illness | . | . | 2,065,094,087 | 38,794 |
| | Total cost (excl. lost productivity) | 615,244,768 | 10,508 | 489,818,889 | 8,366 |
| Eating disorders | Primary sector | 8,641,707 | 439 | 2,598,253 | 132 |
| | Hospital care | 146,421,706 | 7,434 | 113,859,541 | 5,781 |
| | Filled prescriptions | 5,778,497 | 293 | 2,767,124 | 140 |
| | Home care | 1,331,351 | 68 | 876,233 | 44 |
| | Nursing home / sheltered accomodation | 1,544,507 | 78 | 1,335,872 | 68 |
| | Lost productivity associated with illness | . | . | 128,547,028 | 7,148 |
| | Total cost (excl. lost productivity) | 163,717,768 | 8,313 | 121,437,022 | 6,166 |
| Epilepsy | Primary sector | 32,506,905 | 466 | 10,932,060 | 157 |
| | Hospital care | 341,166,561 | 4,889 | 194,302,905 | 2,784 |
| | Filled prescriptions | 61,579,885 | 882 | 43,400,452 | 622 |
| | Home care | 90,457,061 | 1,296 | 73,414,448 | 1,052 |
| | Nursing home / sheltered accomodation | 206,567,179 | 2,960 | 165,060,674 | 2,365 |
| | Lost productivity associated with illness | . | . | 957,869,365 | 22,423 |
| | Total cost (excl. lost productivity) | 732,277,592 | 10,493 | 487,110,538 | 6,980 |
| Headache | Primary sector | 58,947,995 | 456 | 18,072,368 | 140 |
| | Hospital care | 417,892,321 | 3,236 | 165,631,683 | 1,282 |
| | Filled prescriptions | 50,159,007 | 388 | 19,110,624 | 148 |
| | Home care | 29,415,537 | 228 | 9,132,867 | 71 |
| | Nursing home / sheltered accomodation | 53,480,951 | 414 | 12,046,264 | 93 |
| | Lost productivity associated with illness | . | . | 790,405,211 | 7,849 |
| | Total cost (excl. lost productivity) | 609,895,812 | 4,722 | 223,993,805 | 1,734 |
| Infections of the CNS | Primary sector | 11,588,045 | 385 | 2,468,849 | 82 |
| | Hospital care | 109,999,174 | 3,655 | 48,384,430 | 1,608 |
| | Filled prescriptions | 11,365,463 | 378 | 3,532,184 | 117 |
| | Home care | 15,292,530 | 508 | 8,430,911 | 280 |
| | Nursing home / sheltered accomodation | 33,441,847 | 1,111 | 17,133,976 | 569 |
| | Lost productivity associated with illness | . | . | 35,271,457 | 1,994 |
| | Total cost (excl. lost productivity) | 181,687,060 | 6,037 | 79,950,351 | 2,657 |
| Intellectual disability | Primary sector | 13,261,092 | 431 | 6,175,200 | 201 |
| | Hospital care | 163,662,708 | 5,321 | 121,404,278 | 3,947 |
| | Filled prescriptions | 23,887,092 | 777 | 19,295,306 | 627 |
| | Home care | 18,852,632 | 613 | 17,304,070 | 563 |
| | Nursing home / sheltered accomodation | 31,600,748 | 1,027 | 28,817,605 | 937 |
| | Lost productivity associated with illness | . | . | 830,207,840 | 37,060 |
| | Total cost (excl. lost productivity) | 251,264,272 | 8,170 | 192,996,459 | 6,275 |

***Sensitivity analysis 1 / Prevalent cohort 2021***

| *Brain disorder* | *Cost component* | *Actual costs (EUR)* | *Actual costs per person* | *Attributable costs (EUR)* | *Attributable costs per person* |
|---|---|---|---|---|---|
| Multiple sclerosis | Primary sector | 13,533,717 | 787 | 7,489,392 | 436 |
| | Hospital care | 124,524,169 | 7,245 | 86,356,566 | 5,024 |
| | Filled prescriptions | 9,993,905 | 581 | 4,934,470 | 287 |
| | Home care | 65,155,136 | 3,791 | 62,238,975 | 3,621 |
| | Nursing home / sheltered accomodation | 37,086,868 | 2,158 | 31,155,102 | 1,813 |
| | Lost productivity associated with illness | . | . | 272,092,544 | 20,861 |
| | Total cost (excl. lost productivity) | 250,293,795 | 14,562 | 192,174,505 | 11,181 |
| Neuromuscular disorders | Primary sector | 5,031,654 | 593 | 2,152,155 | 254 |
| | Hospital care | 50,903,497 | 6,003 | 30,426,104 | 3,588 |
| | Filled prescriptions | 5,436,854 | 641 | 2,801,366 | 330 |
| | Home care | 14,358,581 | 1,693 | 11,864,393 | 1,399 |
| | Nursing home / sheltered accomodation | 11,264,841 | 1,329 | 5,330,222 | 629 |
| | Lost productivity associated with illness | . | . | 88,739,681 | 17,590 |
| | Total cost (excl. lost productivity) | 86,995,427 | 10,260 | 52,574,241 | 6,200 |
| Other neurodegenerative disorders | Primary sector | 2,318,757 | 849 | 1,263,118 | 462 |
| | Hospital care | 22,993,496 | 8,419 | 15,314,169 | 5,607 |
| | Filled prescriptions | 2,316,301 | 848 | 1,287,127 | 471 |
| | Home care | 14,372,320 | 5,262 | 13,440,310 | 4,921 |
| | Nursing home / sheltered accomodation | 19,720,666 | 7,220 | 17,291,537 | 6,331 |
| | Lost productivity associated with illness | . | . | 46,444,116 | 36,484 |
| | Total cost (excl. lost productivity) | 61,721,541 | 22,598 | 48,596,261 | 17,793 |
| Parkinson's disease | Primary sector | 11,676,647 | 1,111 | 6,468,451 | 615 |
| | Hospital care | 76,776,110 | 7,302 | 33,864,178 | 3,221 |
| | Filled prescriptions | 34,207,043 | 3,253 | 28,475,428 | 2,708 |
| | Home care | 62,520,389 | 5,946 | 53,500,605 | 5,088 |
| | Nursing home / sheltered accomodation | 155,067,012 | 14,748 | 130,883,281 | 12,448 |
| | Lost productivity associated with illness | . | . | 49,769,138 | 32,508 |
| | Total cost (excl. lost productivity) | 340,247,200 | 32,359 | 253,191,943 | 24,080 |
| Personality disorders | Primary sector | 34,947,485 | 463 | 12,319,356 | 163 |
| | Hospital care | 506,465,838 | 6,704 | 378,072,309 | 5,005 |
| | Filled prescriptions | 36,991,862 | 490 | 22,491,472 | 298 |
| | Home care | 20,435,946 | 271 | 15,315,044 | 203 |
| | Nursing home / sheltered accomodation | 38,200,140 | 506 | 30,637,572 | 406 |
| | Lost productivity associated with illness | . | . | 2,389,716,719 | 34,148 |
| | Total cost (excl. lost productivity) | 637,041,269 | 8,433 | 458,835,754 | 6,074 |
| Polyneuropathy | Primary sector | 27,796,232 | 640 | 9,232,165 | 212 |
| | Hospital care | 336,539,484 | 7,745 | 192,526,256 | 4,431 |
| | Filled prescriptions | 45,063,592 | 1,037 | 25,545,645 | 588 |
| | Home care | 76,074,731 | 1,751 | 48,671,202 | 1,120 |

*Sensitivity analysis 1 / Prevalent cohort 2021*

| *Brain disorder* | *Cost component* | *Actual costs (EUR)* | *Actual costs per person* | *Attributable costs (EUR)* | *Attributable costs per person* |
|---|---|---|---|---|---|
| | Nursing home / sheltered accomodation | 126,302,376 | 2,907 | 53,490,542 | 1,231 |
| | Lost productivity associated with illness | . | . | 391,020,228 | 23,908 |
| | Total cost (excl. lost productivity) | 611,776,415 | 14,079 | 329,465,810 | 7,582 |
| Schizophrenia spectrum disorders | Primary sector | 27,818,015 | 399 | 7,757,907 | 111 |
| | Hospital care | 806,291,724 | 11,550 | 687,190,047 | 9,844 |
| | Filled prescriptions | 51,384,380 | 736 | 36,588,948 | 524 |
| | Home care | 46,322,900 | 664 | 36,935,829 | 529 |
| | Nursing home / sheltered accomodation | 140,161,981 | 2,008 | 121,682,604 | 1,743 |
| | Lost productivity associated with illness | . | . | 2,552,794,600 | 43,721 |
| | Total cost (excl. lost productivity) | 1,071,979,001 | 15,356 | 890,155,335 | 12,752 |
| Sleep disorders | Primary sector | 60,020,356 | 480 | 19,038,482 | 152 |
| | Hospital care | 544,321,190 | 4,350 | 236,440,800 | 1,889 |
| | Filled prescriptions | 88,411,359 | 706 | 49,021,682 | 392 |
| | Home care | 55,753,685 | 446 | 25,892,668 | 207 |
| | Nursing home / sheltered accomodation | 73,576,349 | 588 | 7,055,742 | 56 |
| | Lost productivity associated with illness | . | . | 1,018,071,679 | 13,640 |
| | Total cost (excl. lost productivity) | 822,082,939 | 6,569 | 337,449,374 | 2,697 |
| Stress related disorders | Primary sector | 108,404,313 | 426 | 34,736,261 | 137 |
| | Hospital care | 1,245,735,577 | 4,898 | 824,071,456 | 3,240 |
| | Filled prescriptions | 104,753,617 | 412 | 52,611,879 | 207 |
| | Home care | 72,280,953 | 284 | 45,542,248 | 179 |
| | Nursing home / sheltered accomodation | 166,517,118 | 655 | 118,067,867 | 464 |
| | Lost productivity associated with illness | . | . | 5,571,484,154 | 26,429 |
| | Total cost (excl. lost productivity) | 1,697,691,578 | 6,675 | 1,075,029,712 | 4,227 |
| Stroke | Primary sector | 77,577,444 | 614 | 20,311,441 | 161 |
| | Hospital care | 763,050,453 | 6,036 | 315,333,542 | 2,494 |
| | Filled prescriptions | 102,704,139 | 812 | 40,725,047 | 322 |
| | Home care | 308,795,566 | 2,443 | 215,929,137 | 1,708 |
| | Nursing home / sheltered accomodation | 759,625,321 | 6,009 | 500,300,815 | 3,958 |
| | Lost productivity associated with illness | . | . | 878,570,058 | 24,152 |
| | Total cost (excl. lost productivity) | 2,011,752,924 | 15,914 | 1,092,599,982 | 8,643 |
| Traumatic brain injury | Primary sector | 79,690,687 | 345 | 15,852,434 | 69 |
| | Hospital care | 707,650,901 | 3,065 | 291,149,340 | 1,261 |
| | Filled prescriptions | 74,379,951 | 322 | 23,860,014 | 103 |
| | Home care | 115,926,157 | 502 | 65,638,220 | 284 |
| | Nursing home / sheltered accomodation | 335,629,210 | 1,454 | 212,127,078 | 919 |
| | Lost productivity associated with illness | . | . | 1,340,901,388 | 8,676 |
| | Total cost (excl. lost productivity) | 1,313,276,906 | 5,688 | 608,627,086 | 2,636 |

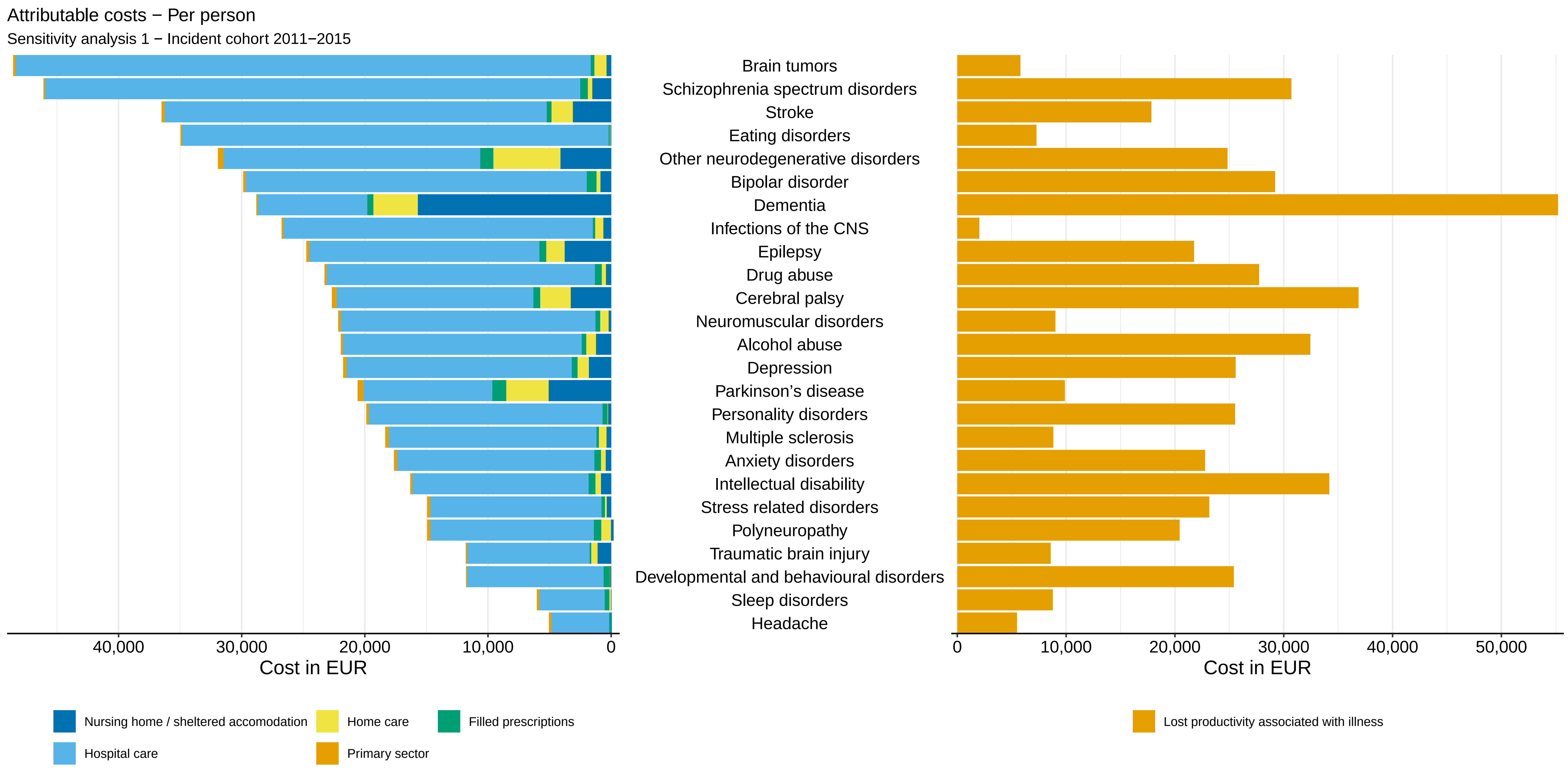
Attributable costs − Per person
Sensitivity analysis 1 − Incident cohort 2011−2015
Brain tumors
Schizophrenia spectrum disorders
Stroke
Eating disorders
Other neurodegenerative disorders
Bipolar disorder
Dementia
Infections of the CNS
Epilepsy
Drug abuse
Cerebral palsy
Neuromuscular disorders
Alcohol abuse
Depression
Parkinson’s disease
Personality disorders
Multiple sclerosis
Anxiety disorders
Intellectual disability
Stress related disorders
Polyneuropathy
Traumatic brain injury
Developmental and behavioural disorders
Sleep disorders
Headache
40,000
30,000
20,000
10,000
0
Cost in EUR
0
10,000
20,000
30,000
40,000
50,000
Cost in EUR
Nursing home / sheltered accomodation
Home care
Filled prescriptions
Hospital care
Primary sector
Lost productivity associated with illness

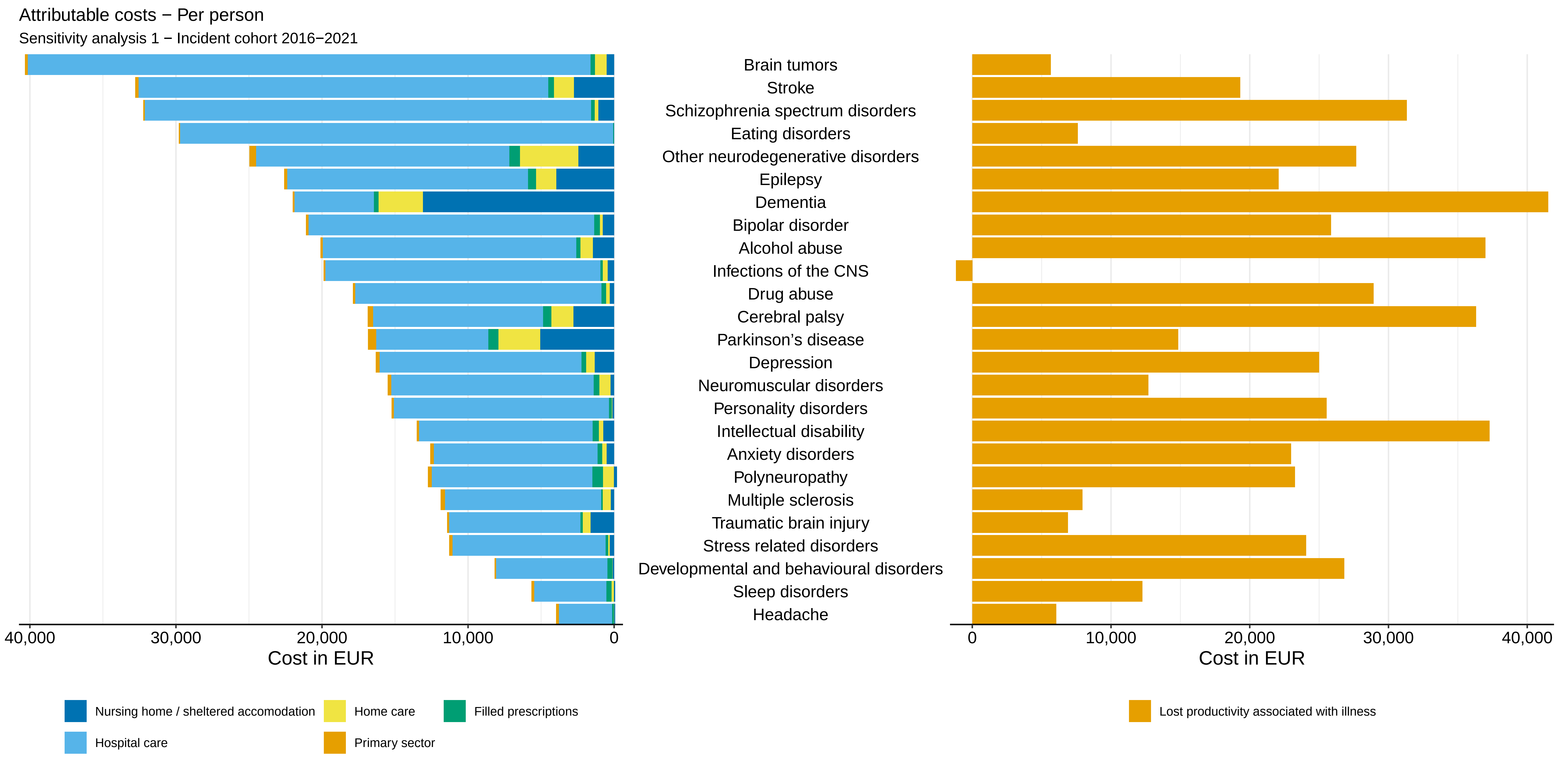

Attributable costs − Per person
Sensitivity analysis 1 − Incident cohort 2016−2021
Brain tumors
Stroke
Schizophrenia spectrum disorders
Eating disorders
Other neurodegenerative disorders
Epilepsy
Dementia
Bipolar disorder
Alcohol abuse
Infections of the CNS
Drug abuse
Cerebral palsy
Parkinson's disease
Depression
Neuromuscular disorders
Personality disorders
Intellectual disability
Anxiety disorders
Polyneuropathy
Multiple sclerosis
Traumatic brain injury
Stress related disorders
Developmental and behavioural disorders
Sleep disorders
Headache
40,000
30,000
20,000
10,000
0
Cost in EUR
0
10,000
20,000
30,000
40,000
Cost in EUR
Nursing home / sheltered accomodation
Home care
Filled prescriptions
Hospital care
Primary sector
Lost productivity associated with illness

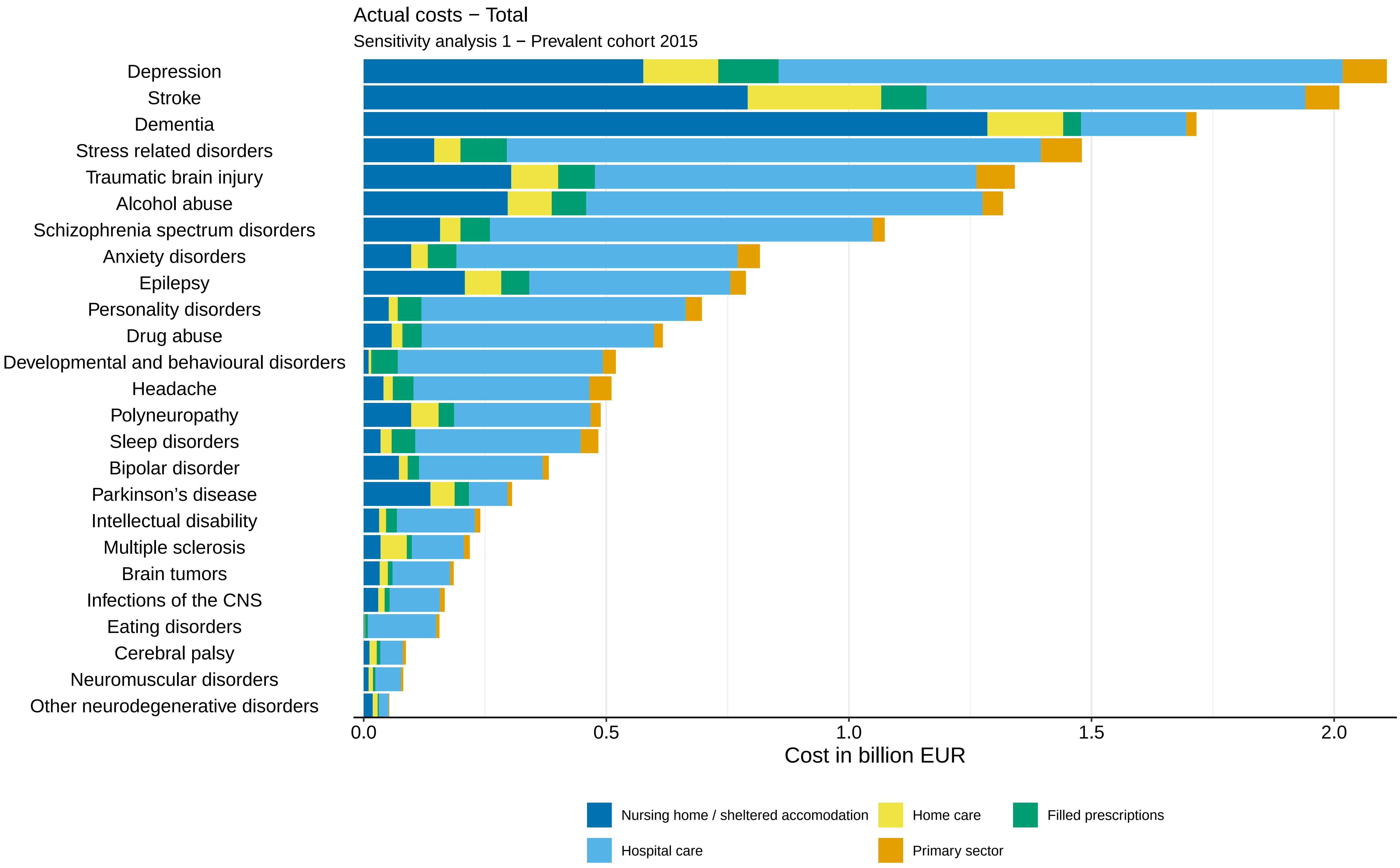

Actual costs − Total
Sensitivity analysis 1 − Prevalent cohort 2015
Depression
Stroke
Dementia
Stress related disorders
Traumatic brain injury
Alcohol abuse
Schizophrenia spectrum disorders
Anxiety disorders
Epilepsy
Personality disorders
Drug abuse
Developmental and behavioural disorders
Headache
Polyneuropathy
Sleep disorders
Bipolar disorder
Parkinson's disease
Intellectual disability
Multiple sclerosis
Brain tumors
Infections of the CNS
Eating disorders
Cerebral palsy
Neuromuscular disorders
Other neurodegenerative disorders
0.0
0.5
1.0
1.5
2.0
Cost in billion EUR
Nursing home / sheltered accomodation
Home care
Filled prescriptions
Hospital care
Primary sector

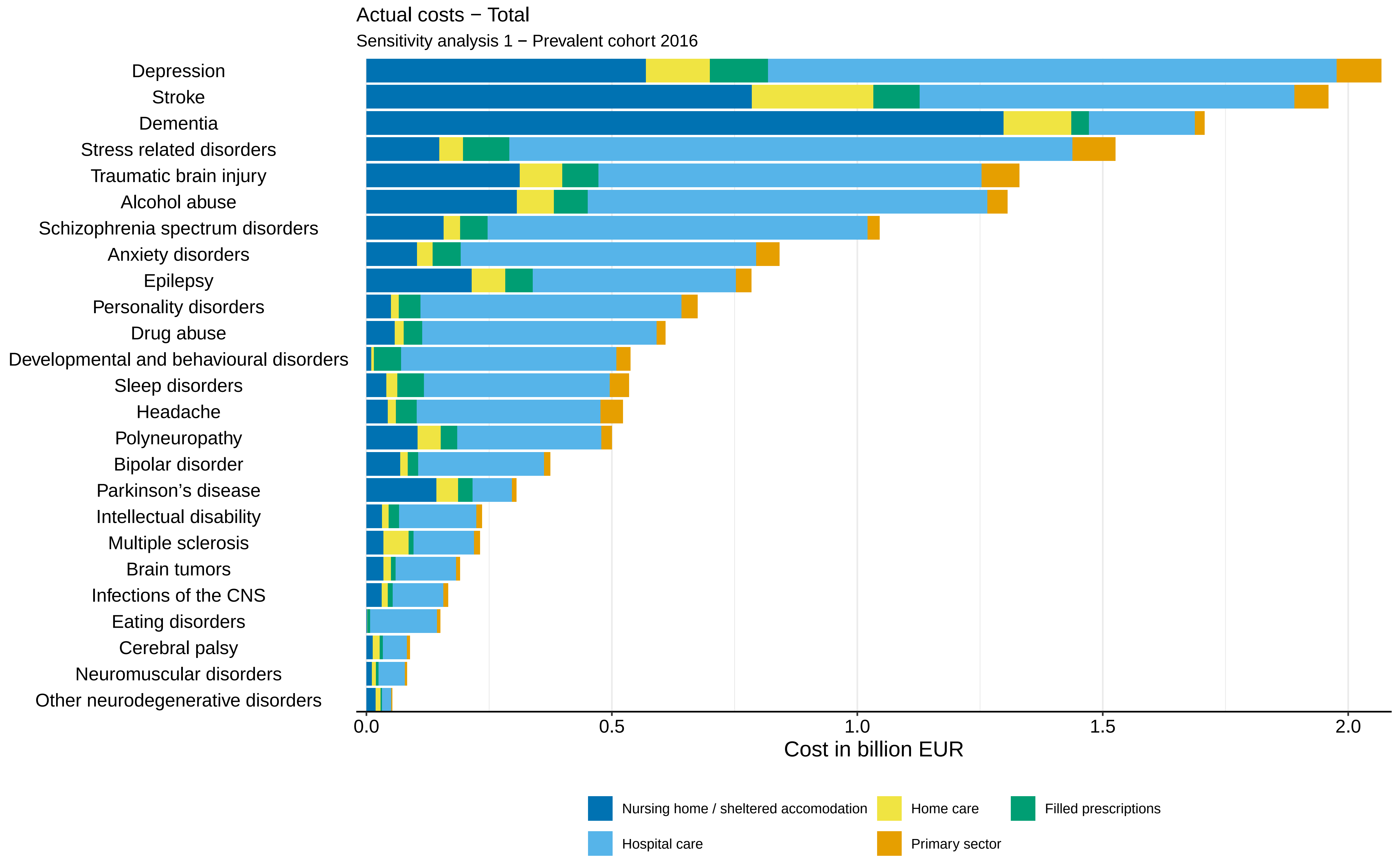

Actual costs − Total
Sensitivity analysis 1 − Prevalent cohort 2016
Depression
Stroke
Dementia
Stress related disorders
Traumatic brain injury
Alcohol abuse
Schizophrenia spectrum disorders
Anxiety disorders
Epilepsy
Personality disorders
Drug abuse
Developmental and behavioural disorders
Sleep disorders
Headache
Polyneuropathy
Bipolar disorder
Parkinson's disease
Intellectual disability
Multiple sclerosis
Brain tumors
Infections of the CNS
Eating disorders
Cerebral palsy
Neuromuscular disorders
Other neurodegenerative disorders
0.0
0.5
1.0
1.5
2.0
Cost in billion EUR
Nursing home / sheltered accomodation
Home care
Filled prescriptions
Hospital care
Primary sector

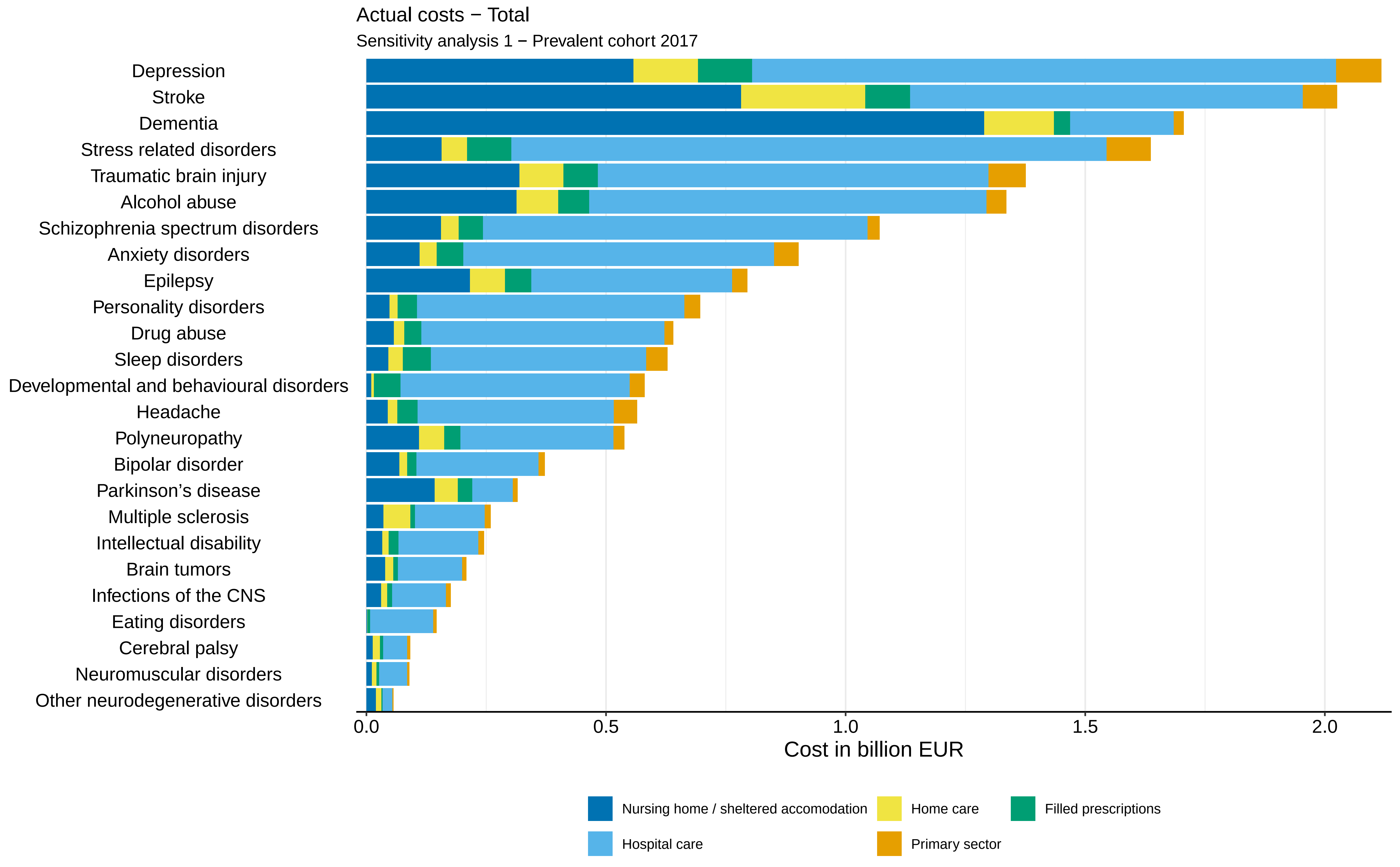

Actual costs − Total
Sensitivity analysis 1 − Prevalent cohort 2017
Depression
Stroke
Dementia
Stress related disorders
Traumatic brain injury
Alcohol abuse
Schizophrenia spectrum disorders
Anxiety disorders
Epilepsy
Personality disorders
Drug abuse
Sleep disorders
Developmental and behavioural disorders
Headache
Polyneuropathy
Bipolar disorder
Parkinson's disease
Multiple sclerosis
Intellectual disability
Brain tumors
Infections of the CNS
Eating disorders
Cerebral palsy
Neuromuscular disorders
Other neurodegenerative disorders
0.0
0.5
1.0
1.5
2.0
Cost in billion EUR
Nursing home / sheltered accomodation
Home care
Filled prescriptions
Hospital care
Primary sector

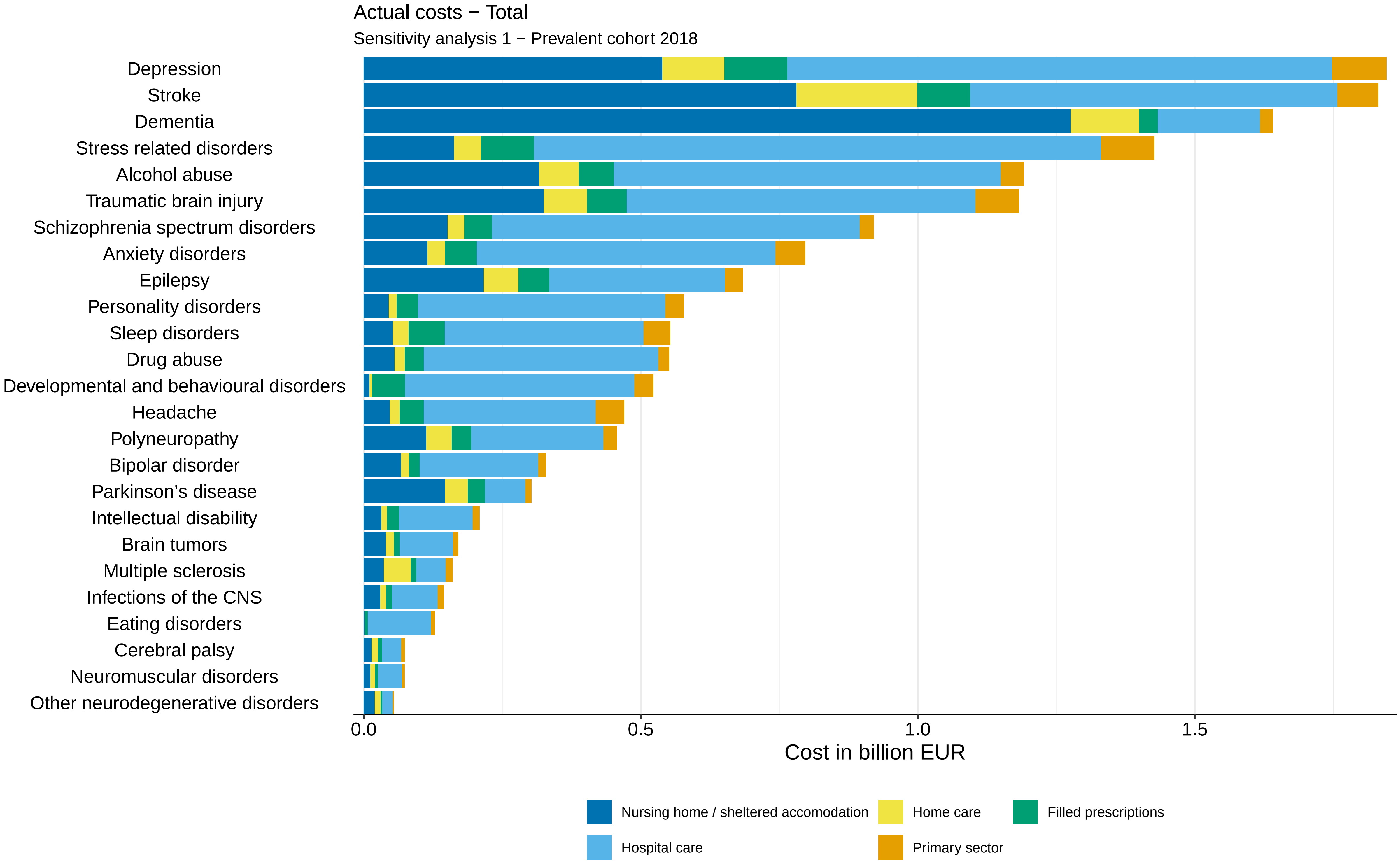

Actual costs − Total
Sensitivity analysis 1 − Prevalent cohort 2018
Depression
Stroke
Dementia
Stress related disorders
Alcohol abuse
Traumatic brain injury
Schizophrenia spectrum disorders
Anxiety disorders
Epilepsy
Personality disorders
Sleep disorders
Drug abuse
Developmental and behavioural disorders
Headache
Polyneuropathy
Bipolar disorder
Parkinson’s disease
Intellectual disability
Brain tumors
Multiple sclerosis
Infections of the CNS
Eating disorders
Cerebral palsy
Neuromuscular disorders
Other neurodegenerative disorders
0.0
0.5
1.0
1.5
Cost in billion EUR
Nursing home / sheltered accomodation
Home care
Filled prescriptions
Hospital care
Primary sector

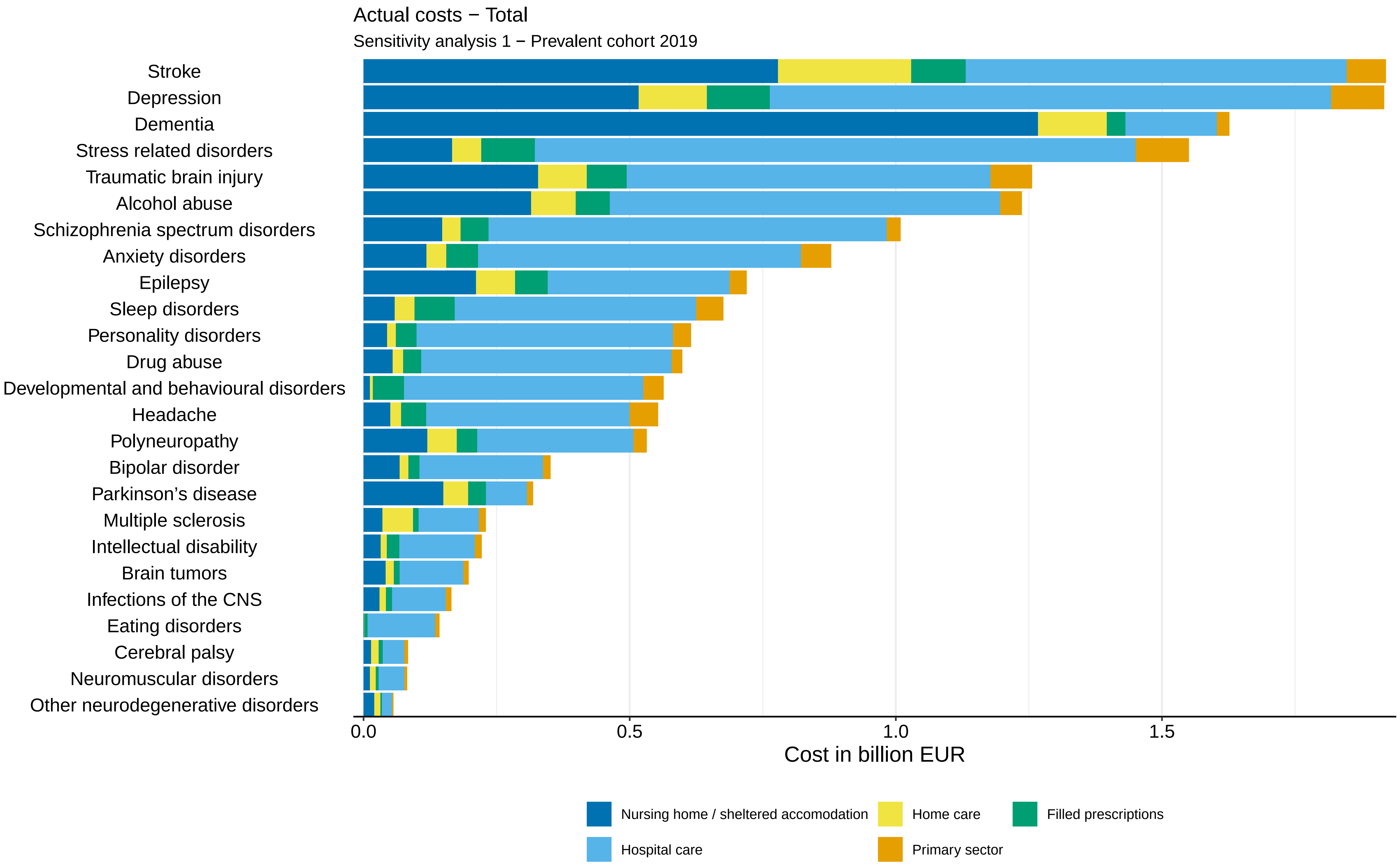

Actual costs − Total
Sensitivity analysis 1 − Prevalent cohort 2019
Stroke
Depression
Dementia
Stress related disorders
Traumatic brain injury
Alcohol abuse
Schizophrenia spectrum disorders
Anxiety disorders
Epilepsy
Sleep disorders
Personality disorders
Drug abuse
Developmental and behavioural disorders
Headache
Polyneuropathy
Bipolar disorder
Parkinson's disease
Multiple sclerosis
Intellectual disability
Brain tumors
Infections of the CNS
Eating disorders
Cerebral palsy
Neuromuscular disorders
Other neurodegenerative disorders
0.0
0.5
1.0
1.5
Cost in billion EUR
Nursing home / sheltered accomodation
Home care
Filled prescriptions
Hospital care
Primary sector

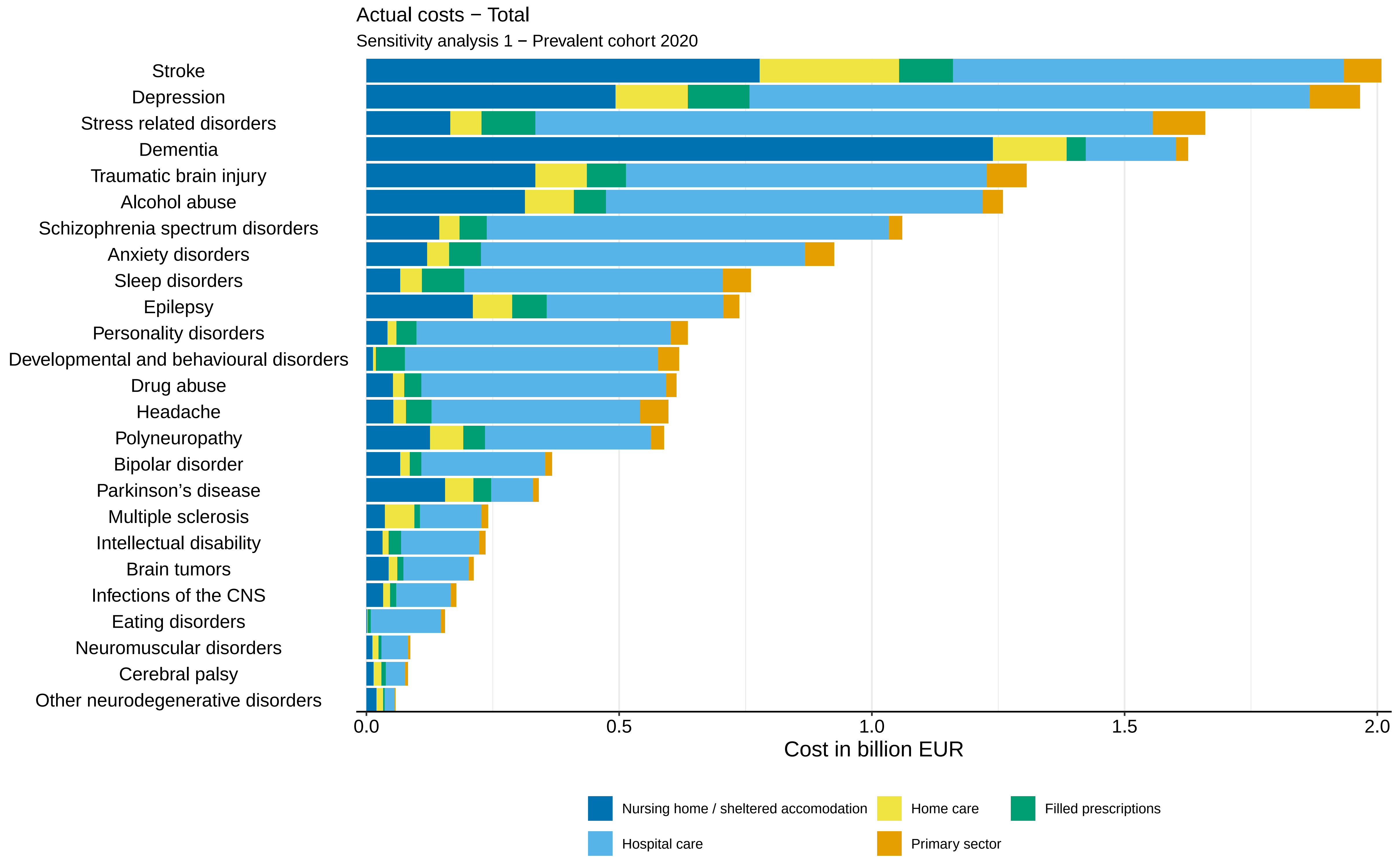

Actual costs − Total
Sensitivity analysis 1 − Prevalent cohort 2020
Stroke
Depression
Stress related disorders
Dementia
Traumatic brain injury
Alcohol abuse
Schizophrenia spectrum disorders
Anxiety disorders
Sleep disorders
Epilepsy
Personality disorders
Developmental and behavioural disorders
Drug abuse
Headache
Polyneuropathy
Bipolar disorder
Parkinson's disease
Multiple sclerosis
Intellectual disability
Brain tumors
Infections of the CNS
Eating disorders
Neuromuscular disorders
Cerebral palsy
Other neurodegenerative disorders
0.0
0.5
1.0
1.5
2.0
Cost in billion EUR
Nursing home / sheltered accomodation
Home care
Filled prescriptions
Hospital care
Primary sector

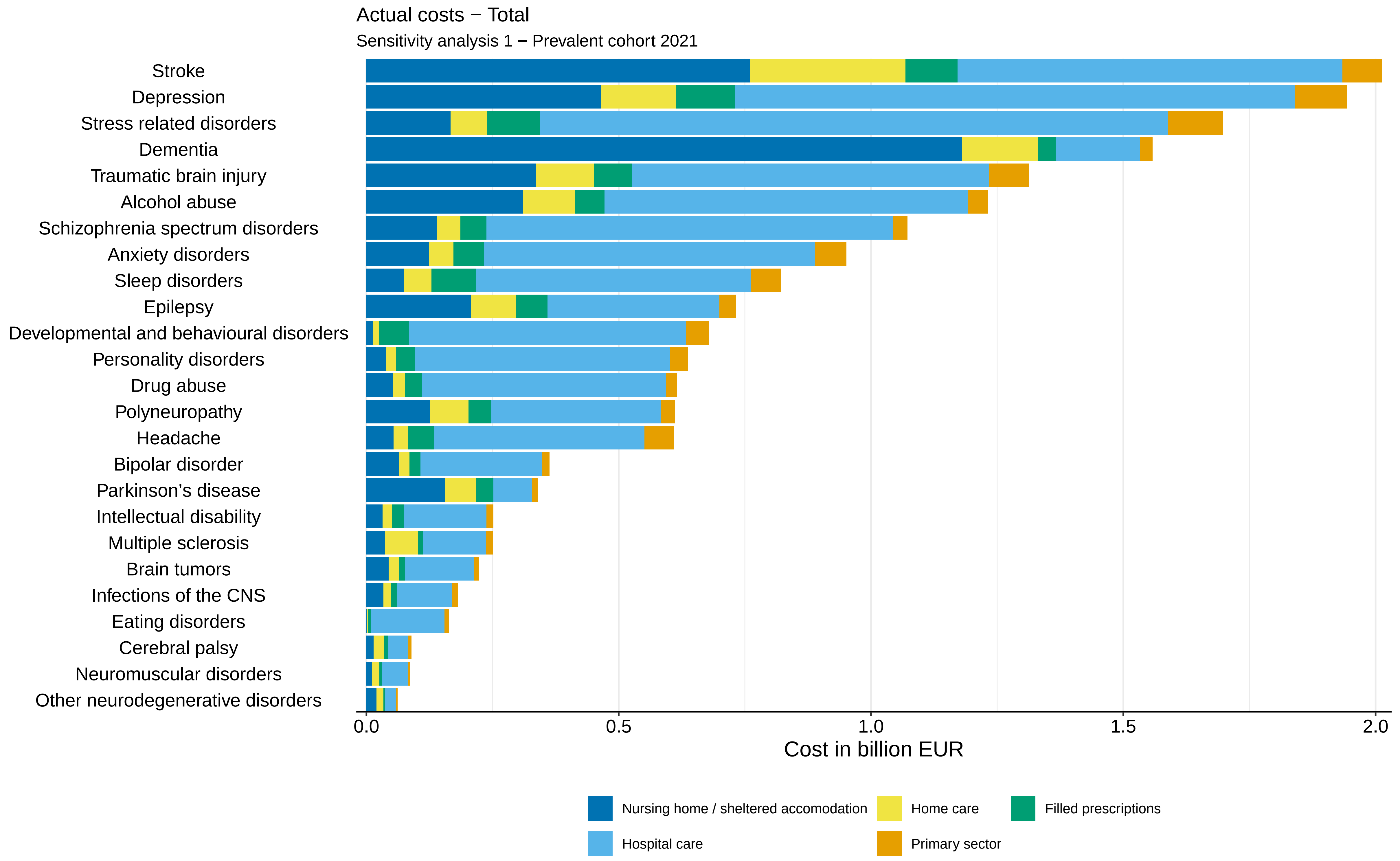

Actual costs − Total
Sensitivity analysis 1 − Prevalent cohort 2021
Stroke
Depression
Stress related disorders
Dementia
Traumatic brain injury
Alcohol abuse
Schizophrenia spectrum disorders
Anxiety disorders
Sleep disorders
Epilepsy
Developmental and behavioural disorders
Personality disorders
Drug abuse
Polyneuropathy
Headache
Bipolar disorder
Parkinson’s disease
Intellectual disability
Multiple sclerosis
Brain tumors
Infections of the CNS
Eating disorders
Cerebral palsy
Neuromuscular disorders
Other neurodegenerative disorders
0.0
0.5
1.0
1.5
2.0
Cost in billion EUR
Nursing home / sheltered accomodation
Home care
Filled prescriptions
Hospital care
Primary sector

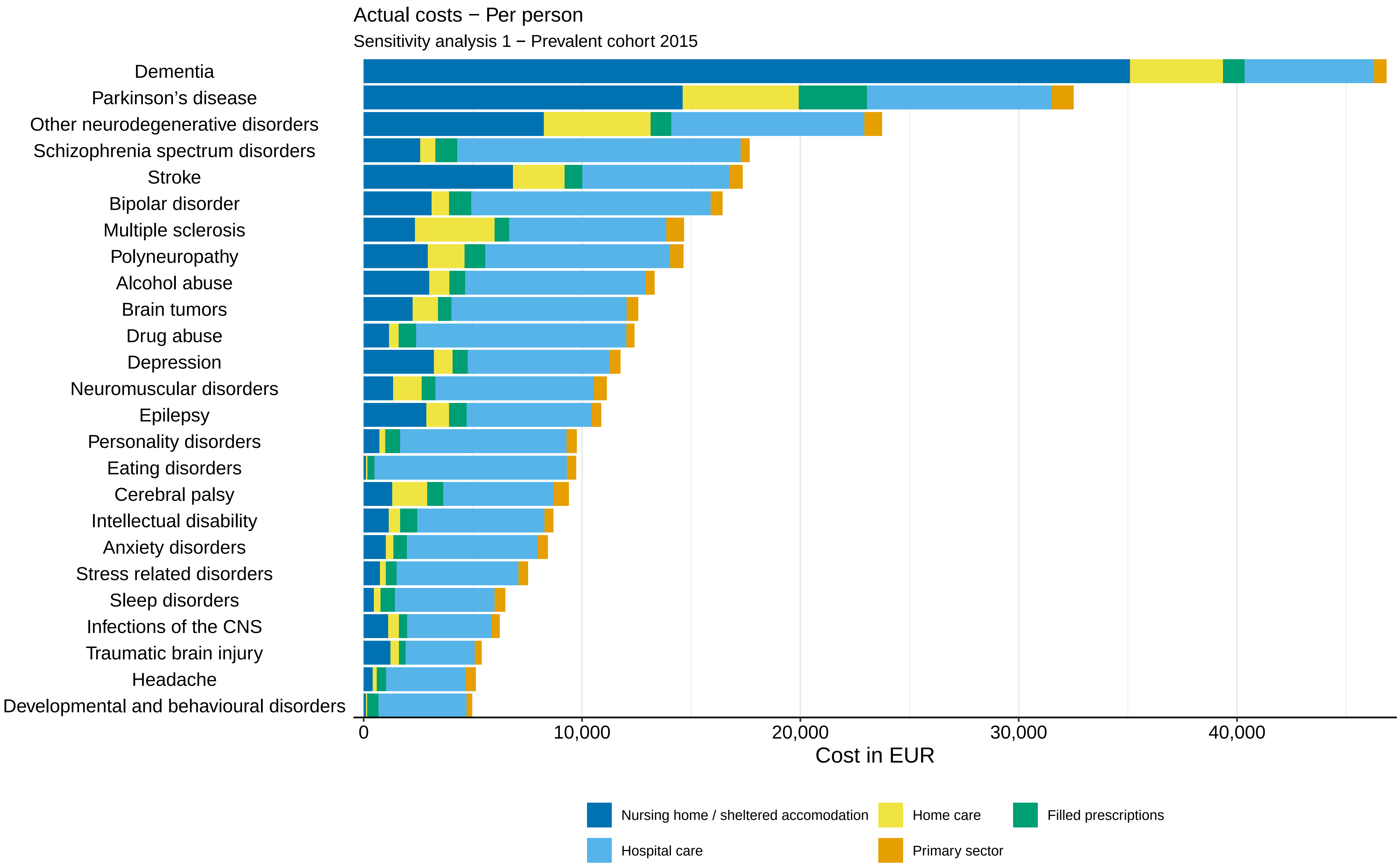

Actual costs − Per person
Sensitivity analysis 1 − Prevalent cohort 2015
Dementia
Parkinson’s disease
Other neurodegenerative disorders
Schizophrenia spectrum disorders
Stroke
Bipolar disorder
Multiple sclerosis
Polyneuropathy
Alcohol abuse
Brain tumors
Drug abuse
Depression
Neuromuscular disorders
Epilepsy
Personality disorders
Eating disorders
Cerebral palsy
Intellectual disability
Anxiety disorders
Stress related disorders
Sleep disorders
Infections of the CNS
Traumatic brain injury
Headache
Developmental and behavioural disorders
0
10,000
20,000
30,000
40,000
Cost in EUR
Nursing home / sheltered accomodation
Home care
Filled prescriptions
Hospital care
Primary sector

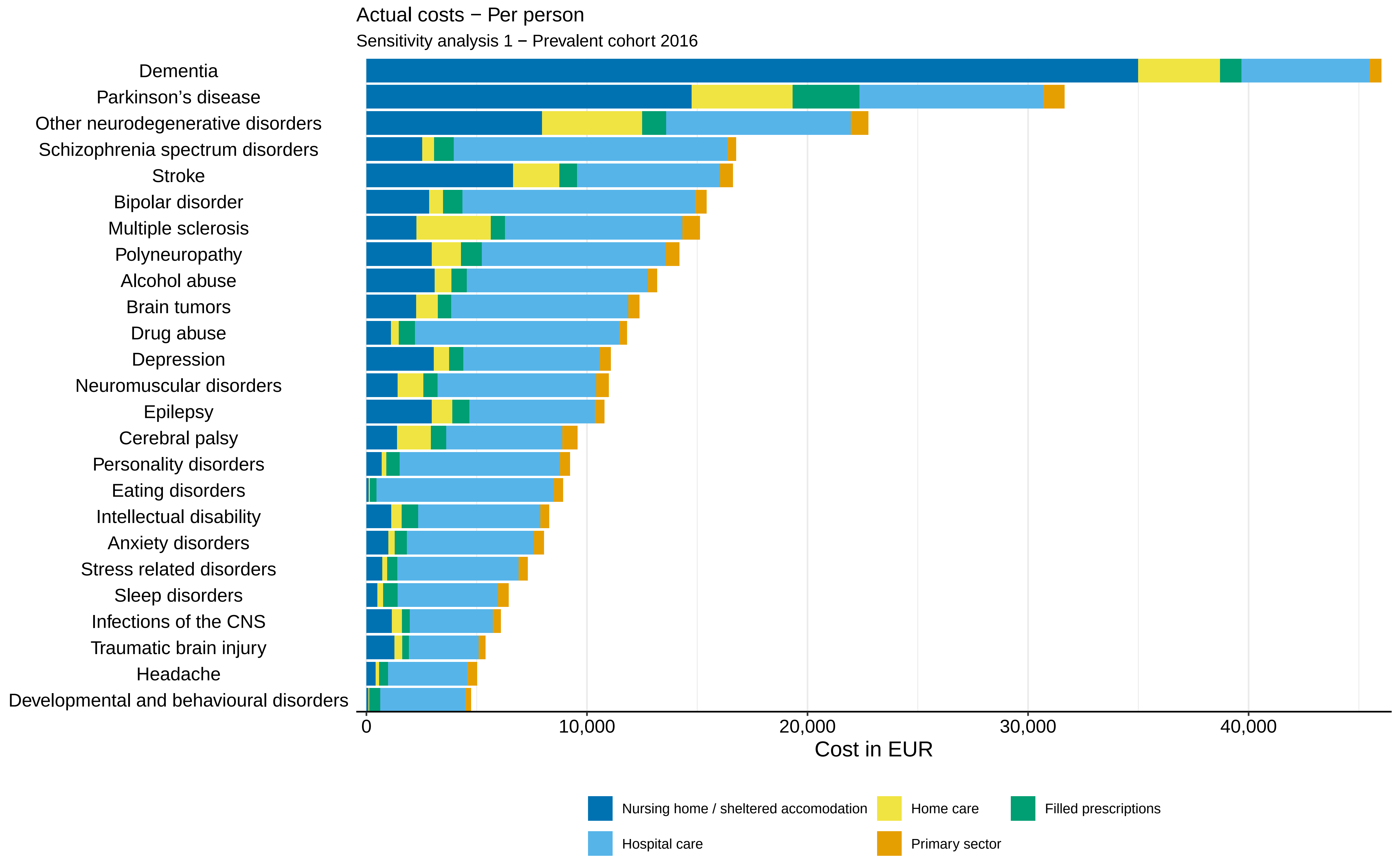

Actual costs − Per person
Sensitivity analysis 1 − Prevalent cohort 2016
Dementia
Parkinson’s disease
Other neurodegenerative disorders
Schizophrenia spectrum disorders
Stroke
Bipolar disorder
Multiple sclerosis
Polyneuropathy
Alcohol abuse
Brain tumors
Drug abuse
Depression
Neuromuscular disorders
Epilepsy
Cerebral palsy
Personality disorders
Eating disorders
Intellectual disability
Anxiety disorders
Stress related disorders
Sleep disorders
Infections of the CNS
Traumatic brain injury
Headache
Developmental and behavioural disorders
0
10,000
20,000
30,000
40,000
Cost in EUR
Nursing home / sheltered accomodation
Home care
Filled prescriptions
Hospital care
Primary sector

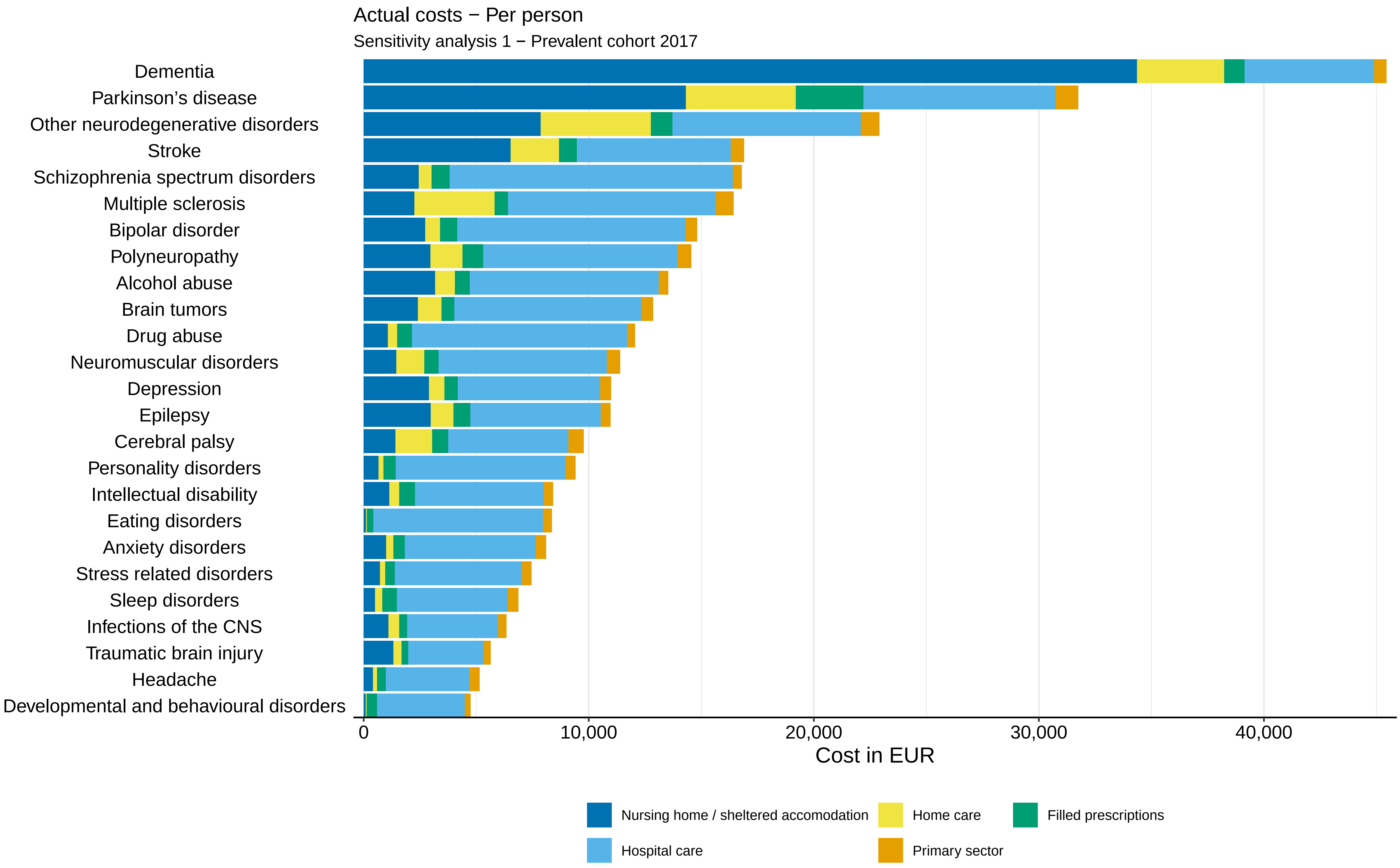
Actual costs − Per person
Sensitivity analysis 1 − Prevalent cohort 2017
Dementia
Parkinson's disease
Other neurodegenerative disorders
Stroke
Schizophrenia spectrum disorders
Multiple sclerosis
Bipolar disorder
Polyneuropathy
Alcohol abuse
Brain tumors
Drug abuse
Neuromuscular disorders
Depression
Epilepsy
Cerebral palsy
Personality disorders
Intellectual disability
Eating disorders
Anxiety disorders
Stress related disorders
Sleep disorders
Infections of the CNS
Traumatic brain injury
Headache
Developmental and behavioural disorders
0
10,000
20,000
30,000
40,000
Cost in EUR
Nursing home / sheltered accomodation
Home care
Filled prescriptions
Hospital care
Primary sector

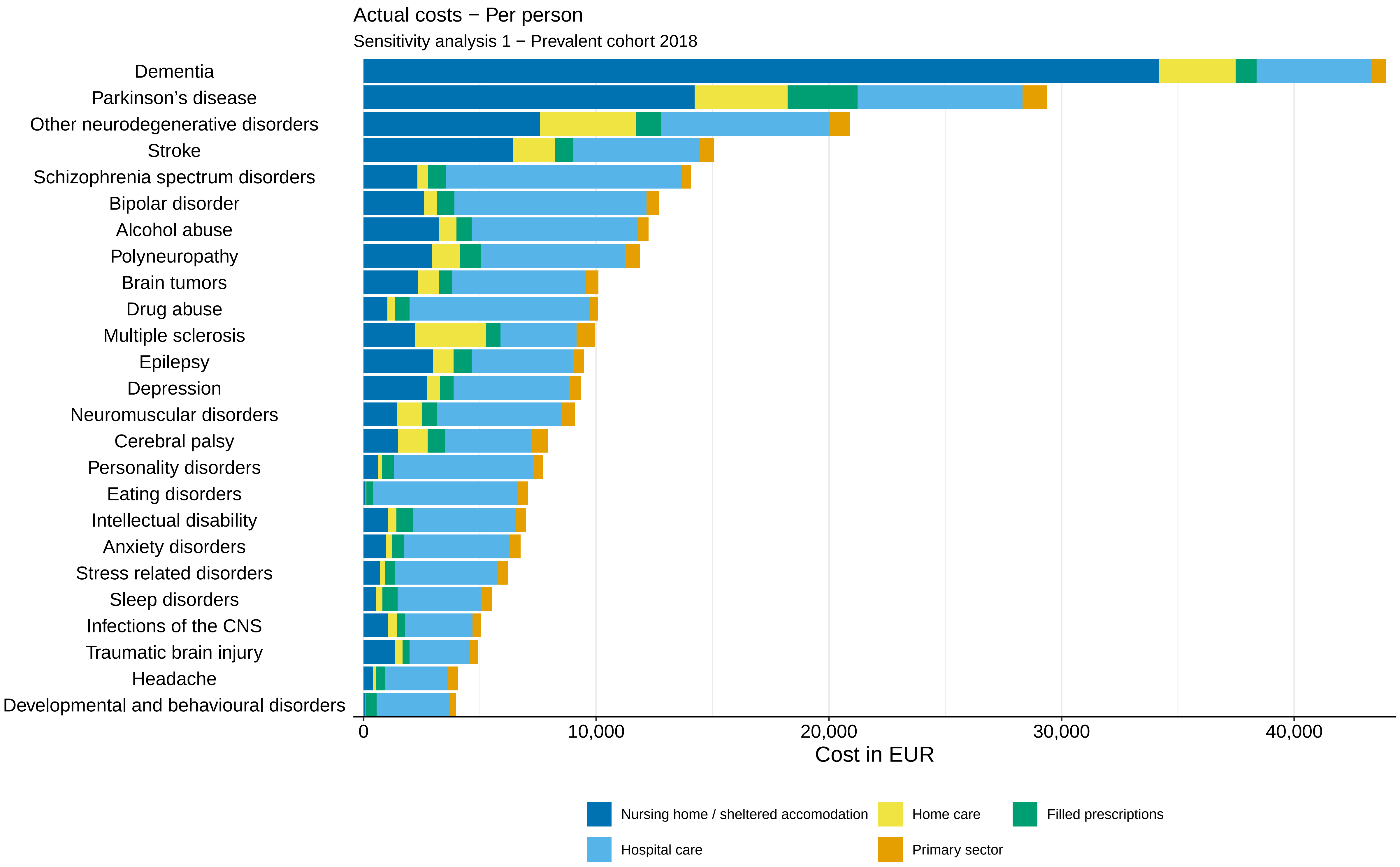

Actual costs − Per person
Sensitivity analysis 1 − Prevalent cohort 2018
Dementia
Parkinson's disease
Other neurodegenerative disorders
Stroke
Schizophrenia spectrum disorders
Bipolar disorder
Alcohol abuse
Polyneuropathy
Brain tumors
Drug abuse
Multiple sclerosis
Epilepsy
Depression
Neuromuscular disorders
Cerebral palsy
Personality disorders
Eating disorders
Intellectual disability
Anxiety disorders
Stress related disorders
Sleep disorders
Infections of the CNS
Traumatic brain injury
Headache
Developmental and behavioural disorders
0
10,000
20,000
30,000
40,000
Cost in EUR
Nursing home / sheltered accomodation
Home care
Filled prescriptions
Hospital care
Primary sector

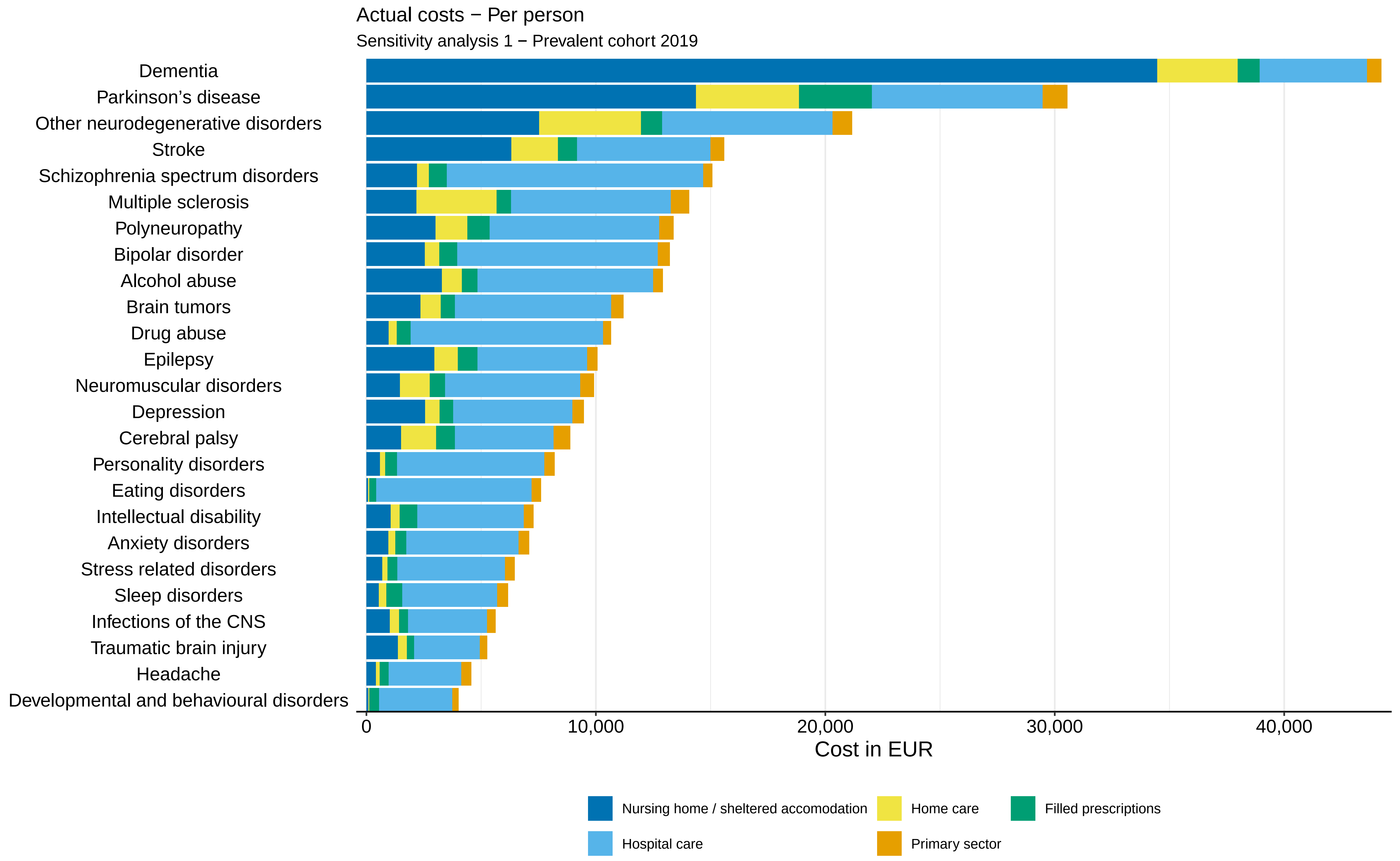
Actual costs − Per person
Sensitivity analysis 1 − Prevalent cohort 2019
Dementia
Parkinson’s disease
Other neurodegenerative disorders
Stroke
Schizophrenia spectrum disorders
Multiple sclerosis
Polyneuropathy
Bipolar disorder
Alcohol abuse
Brain tumors
Drug abuse
Epilepsy
Neuromuscular disorders
Depression
Cerebral palsy
Personality disorders
Eating disorders
Intellectual disability
Anxiety disorders
Stress related disorders
Sleep disorders
Infections of the CNS
Traumatic brain injury
Headache
Developmental and behavioural disorders
0
10,000
20,000
30,000
40,000
Cost in EUR
Nursing home / sheltered accomodation
Home care
Filled prescriptions
Hospital care
Primary sector

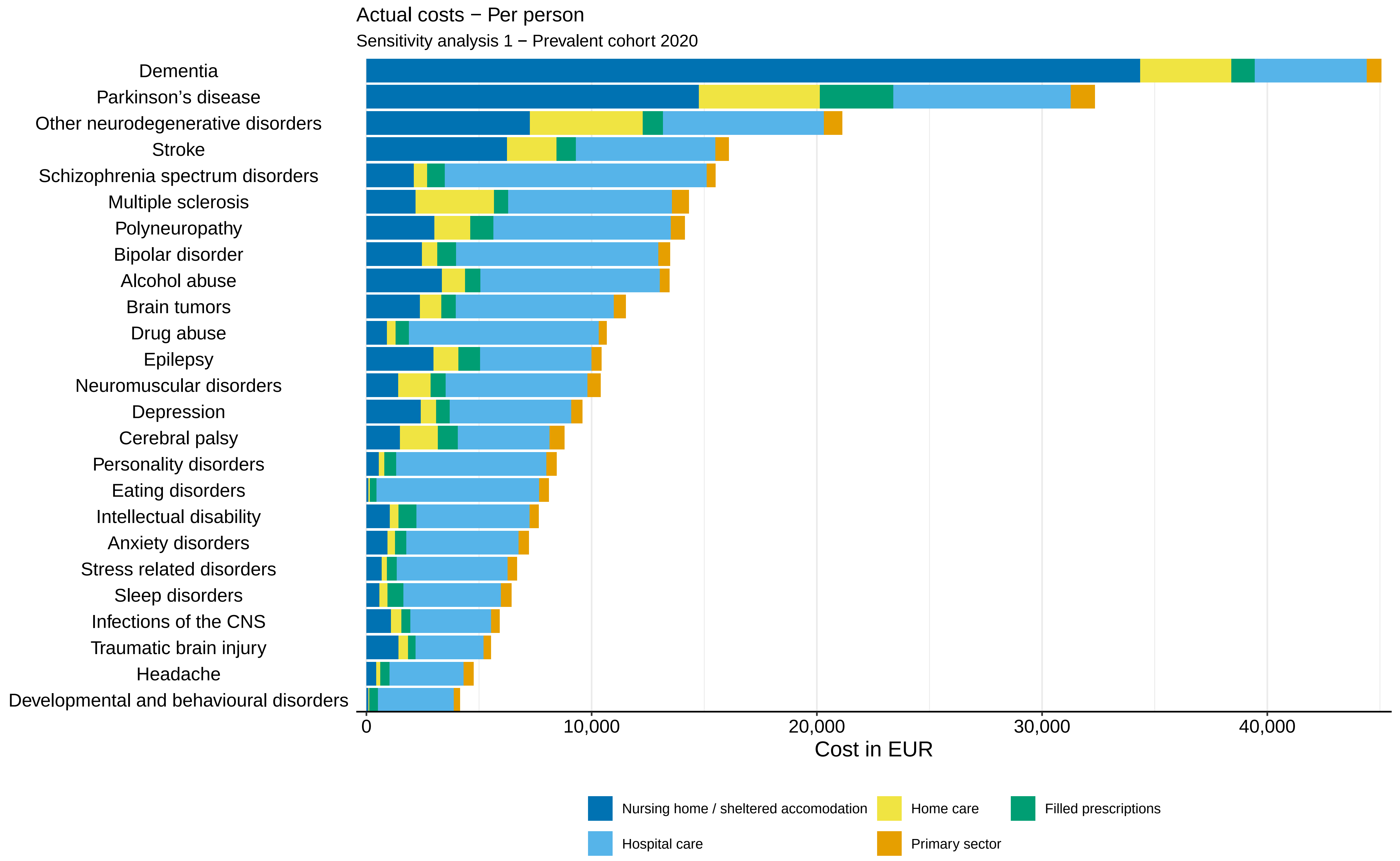

Actual costs − Per person
Sensitivity analysis 1 − Prevalent cohort 2020
Dementia
Parkinson’s disease
Other neurodegenerative disorders
Stroke
Schizophrenia spectrum disorders
Multiple sclerosis
Polyneuropathy
Bipolar disorder
Alcohol abuse
Brain tumors
Drug abuse
Epilepsy
Neuromuscular disorders
Depression
Cerebral palsy
Personality disorders
Eating disorders
Intellectual disability
Anxiety disorders
Stress related disorders
Sleep disorders
Infections of the CNS
Traumatic brain injury
Headache
Developmental and behavioural disorders
0
10,000
20,000
30,000
40,000
Cost in EUR
Nursing home / sheltered accomodation
Home care
Filled prescriptions
Hospital care
Primary sector

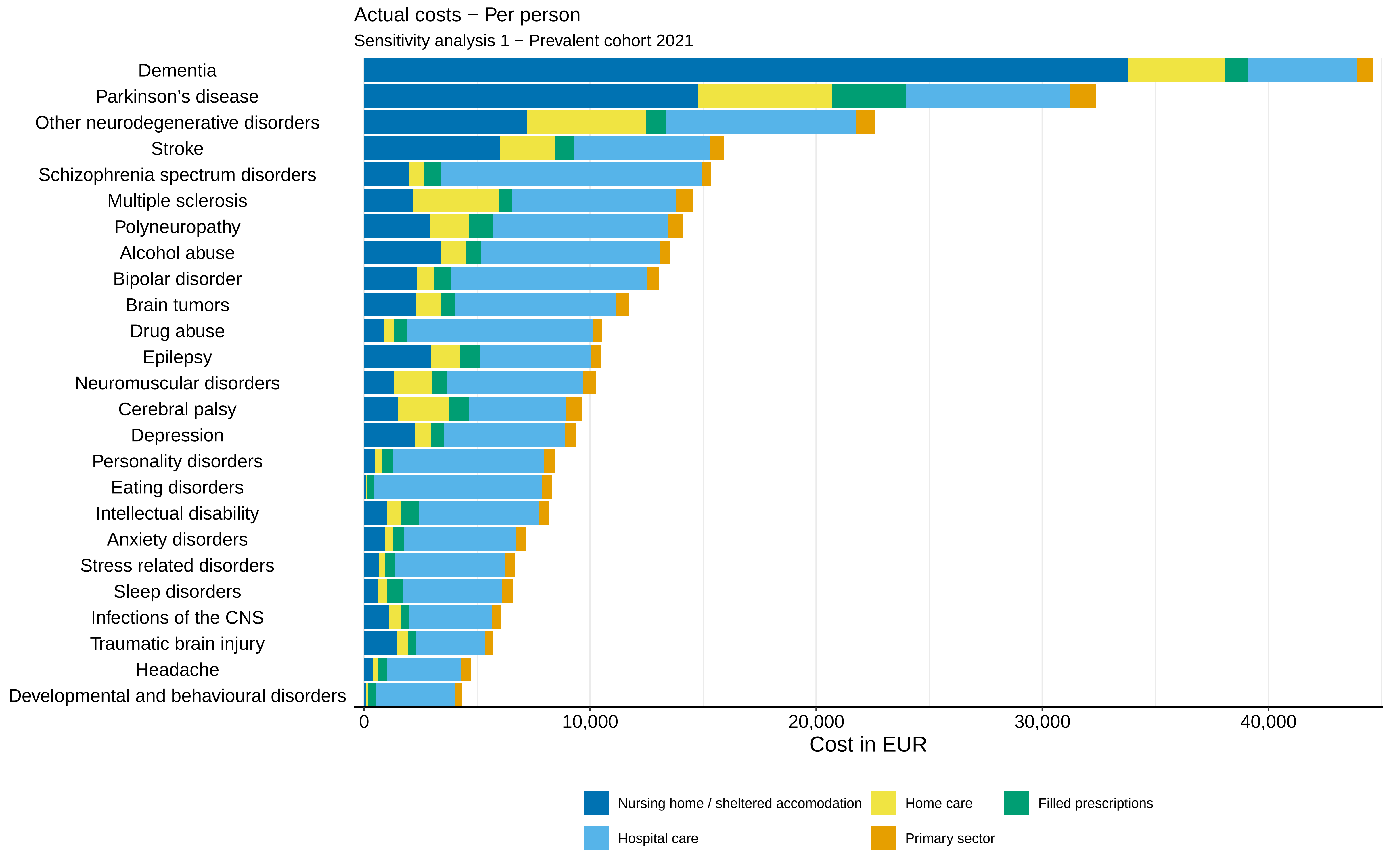

Actual costs − Per person
Sensitivity analysis 1 − Prevalent cohort 2021
Dementia
Parkinson’s disease
Other neurodegenerative disorders
Stroke
Schizophrenia spectrum disorders
Multiple sclerosis
Polyneuropathy
Alcohol abuse
Bipolar disorder
Brain tumors
Drug abuse
Epilepsy
Neuromuscular disorders
Cerebral palsy
Depression
Personality disorders
Eating disorders
Intellectual disability
Anxiety disorders
Stress related disorders
Sleep disorders
Infections of the CNS
Traumatic brain injury
Headache
Developmental and behavioural disorders
0
10,000
20,000
30,000
40,000
Cost in EUR
Nursing home / sheltered accomodation
Home care
Filled prescriptions
Hospital care
Primary sector

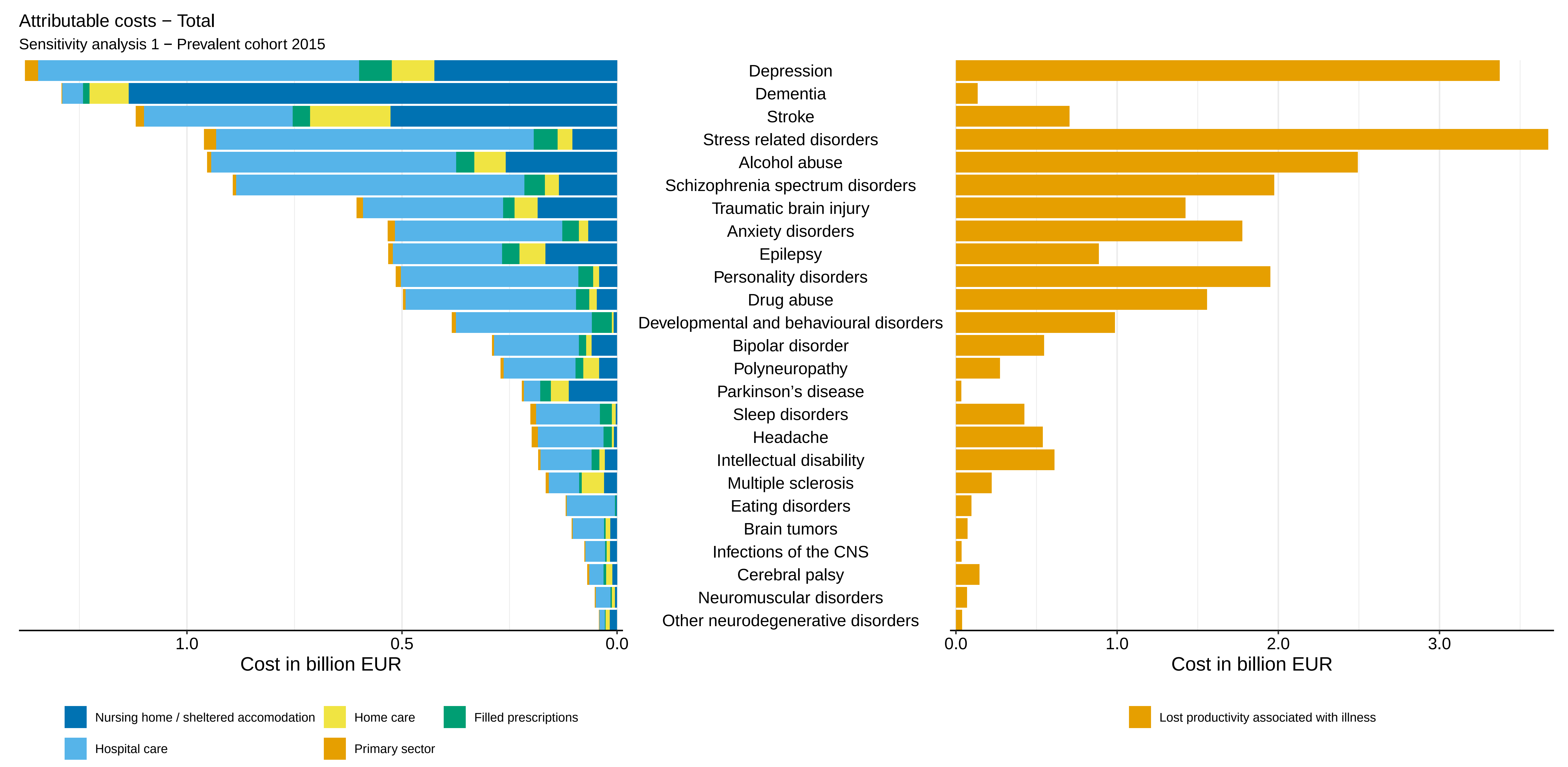
Attributable costs − Total
Sensitivity analysis 1 − Prevalent cohort 2015
Depression
Dementia
Stroke
Stress related disorders
Alcohol abuse
Schizophrenia spectrum disorders
Traumatic brain injury
Anxiety disorders
Epilepsy
Personality disorders
Drug abuse
Developmental and behavioural disorders
Bipolar disorder
Polyneuropathy
Parkinson’s disease
Sleep disorders
Headache
Intellectual disability
Multiple sclerosis
Eating disorders
Brain tumors
Infections of the CNS
Cerebral palsy
Neuromuscular disorders
Other neurodegenerative disorders
1.0
0.5
0.0
Cost in billion EUR
0.0
1.0
2.0
3.0
Cost in billion EUR
Nursing home / sheltered accomodation
Home care
Filled prescriptions
Hospital care
Primary sector
Lost productivity associated with illness

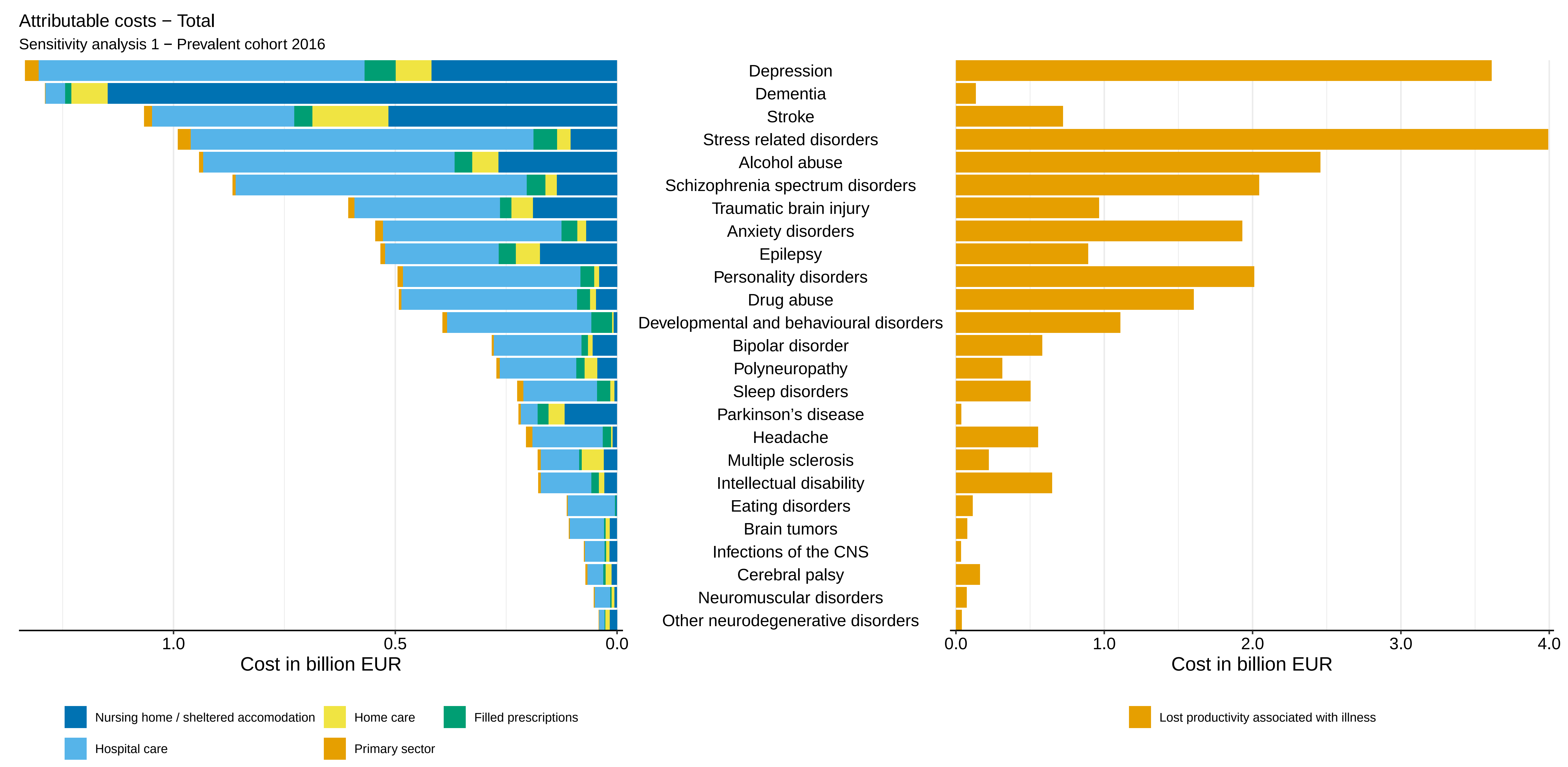

Attributable costs − Total
Sensitivity analysis 1 − Prevalent cohort 2016
Depression
Dementia
Stroke
Stress related disorders
Alcohol abuse
Schizophrenia spectrum disorders
Traumatic brain injury
Anxiety disorders
Epilepsy
Personality disorders
Drug abuse
Developmental and behavioural disorders
Bipolar disorder
Polyneuropathy
Sleep disorders
Parkinson’s disease
Headache
Multiple sclerosis
Intellectual disability
Eating disorders
Brain tumors
Infections of the CNS
Cerebral palsy
Neuromuscular disorders
Other neurodegenerative disorders
1.0
0.5
0.0
Cost in billion EUR
0.0
1.0
2.0
3.0
4.0
Cost in billion EUR
Nursing home / sheltered accomodation
Home care
Filled prescriptions
Hospital care
Primary sector
Lost productivity associated with illness

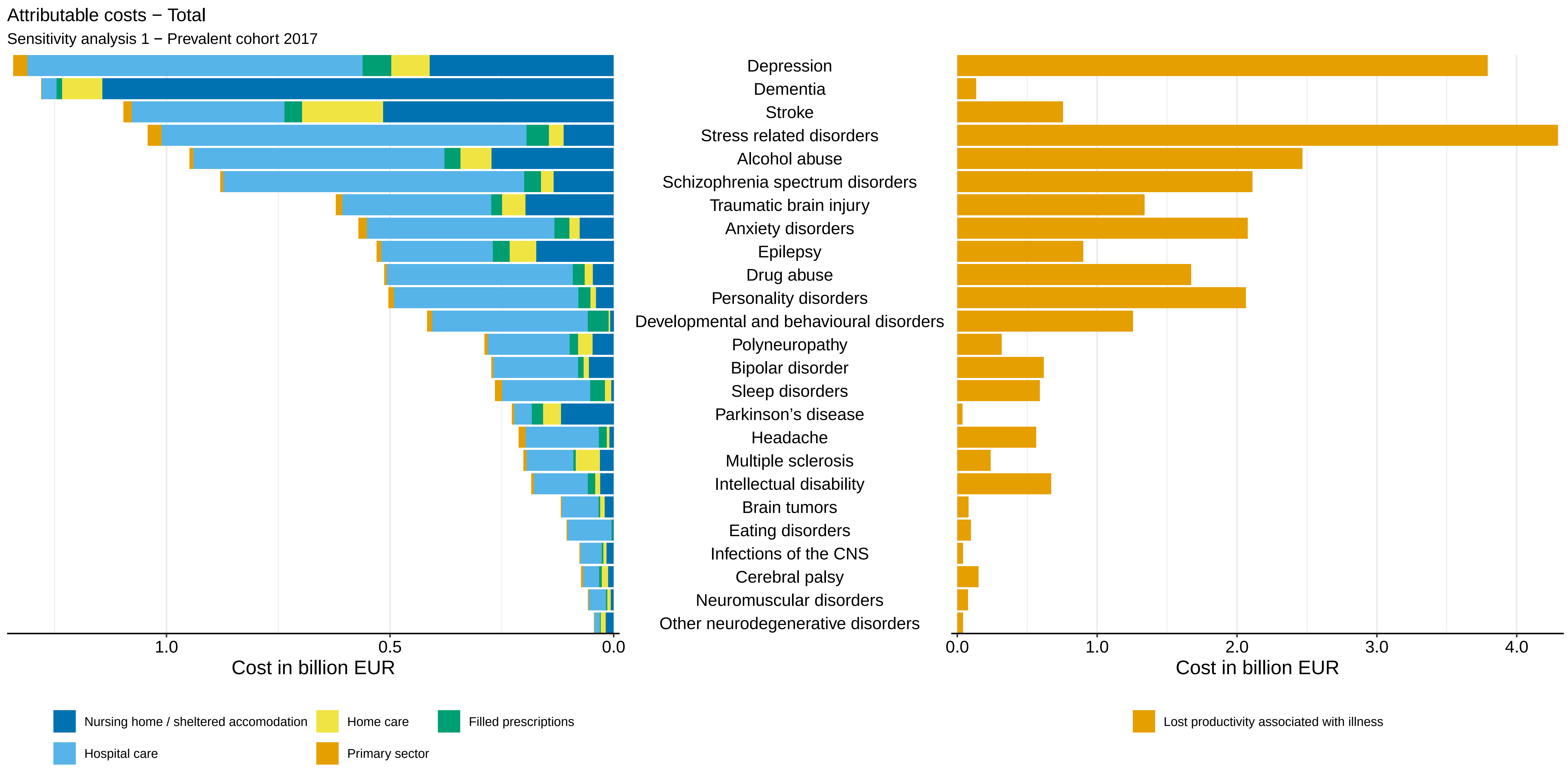

Attributable costs − Total
Sensitivity analysis 1 − Prevalent cohort 2017
Depression
Dementia
Stroke
Stress related disorders
Alcohol abuse
Schizophrenia spectrum disorders
Traumatic brain injury
Anxiety disorders
Epilepsy
Drug abuse
Personality disorders
Developmental and behavioural disorders
Polyneuropathy
Bipolar disorder
Sleep disorders
Parkinson’s disease
Headache
Multiple sclerosis
Intellectual disability
Brain tumors
Eating disorders
Infections of the CNS
Cerebral palsy
Neuromuscular disorders
Other neurodegenerative disorders
1.0
0.5
0.0
Cost in billion EUR
0.0
1.0
2.0
3.0
4.0
Cost in billion EUR
Nursing home / sheltered accomodation
Home care
Filled prescriptions
Hospital care
Primary sector
Lost productivity associated with illness

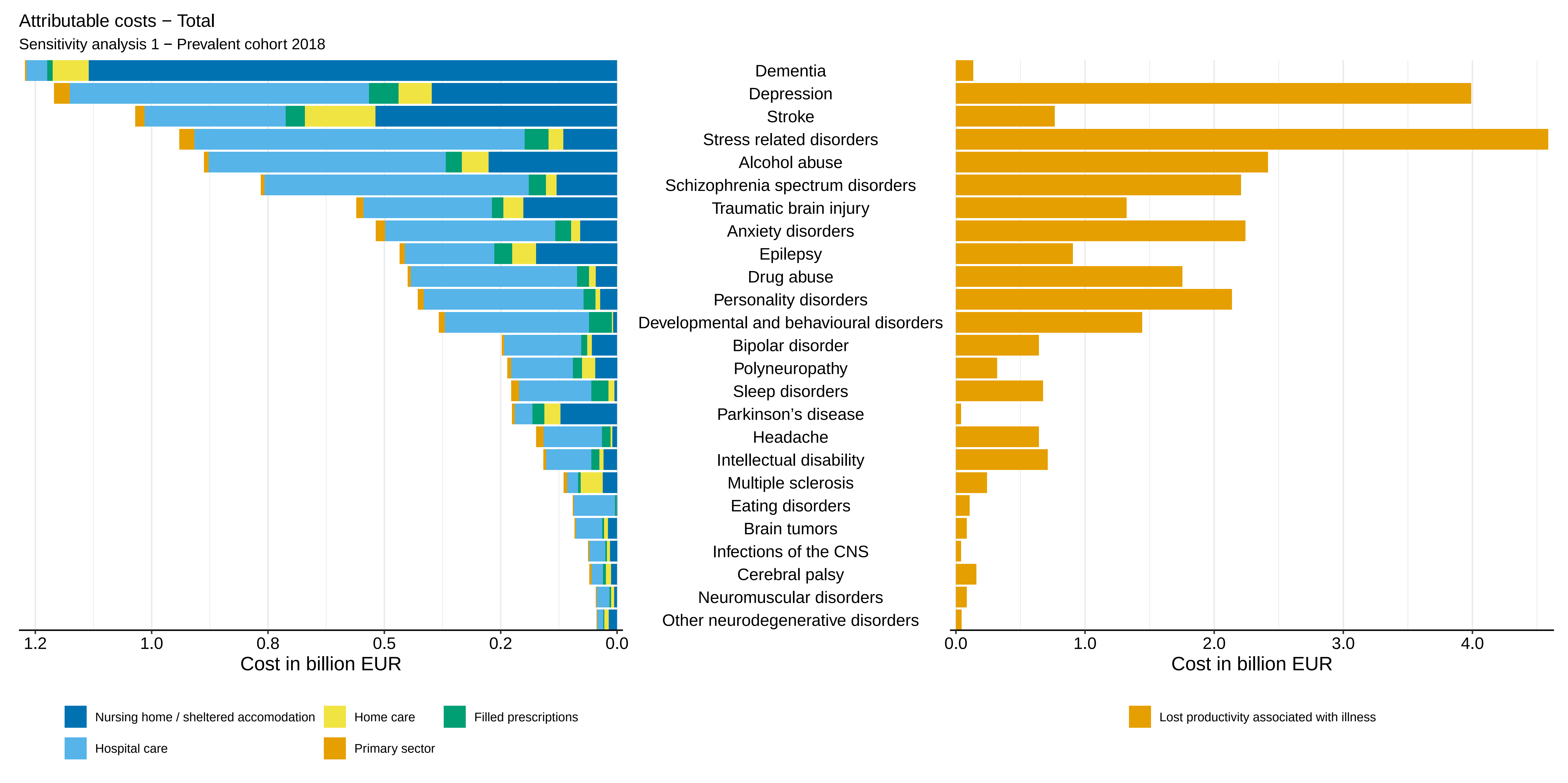
Attributable costs − Total
Sensitivity analysis 1 − Prevalent cohort 2018
Dementia
Depression
Stroke
Stress related disorders
Alcohol abuse
Schizophrenia spectrum disorders
Traumatic brain injury
Anxiety disorders
Epilepsy
Drug abuse
Personality disorders
Developmental and behavioural disorders
Bipolar disorder
Polyneuropathy
Sleep disorders
Parkinson’s disease
Headache
Intellectual disability
Multiple sclerosis
Eating disorders
Brain tumors
Infections of the CNS
Cerebral palsy
Neuromuscular disorders
Other neurodegenerative disorders
1.2
1.0
0.8
0.5
0.2
0.0
Cost in billion EUR
0.0
1.0
2.0
3.0
4.0
Cost in billion EUR
Nursing home / sheltered accomodation
Home care
Filled prescriptions
Hospital care
Primary sector
Lost productivity associated with illness

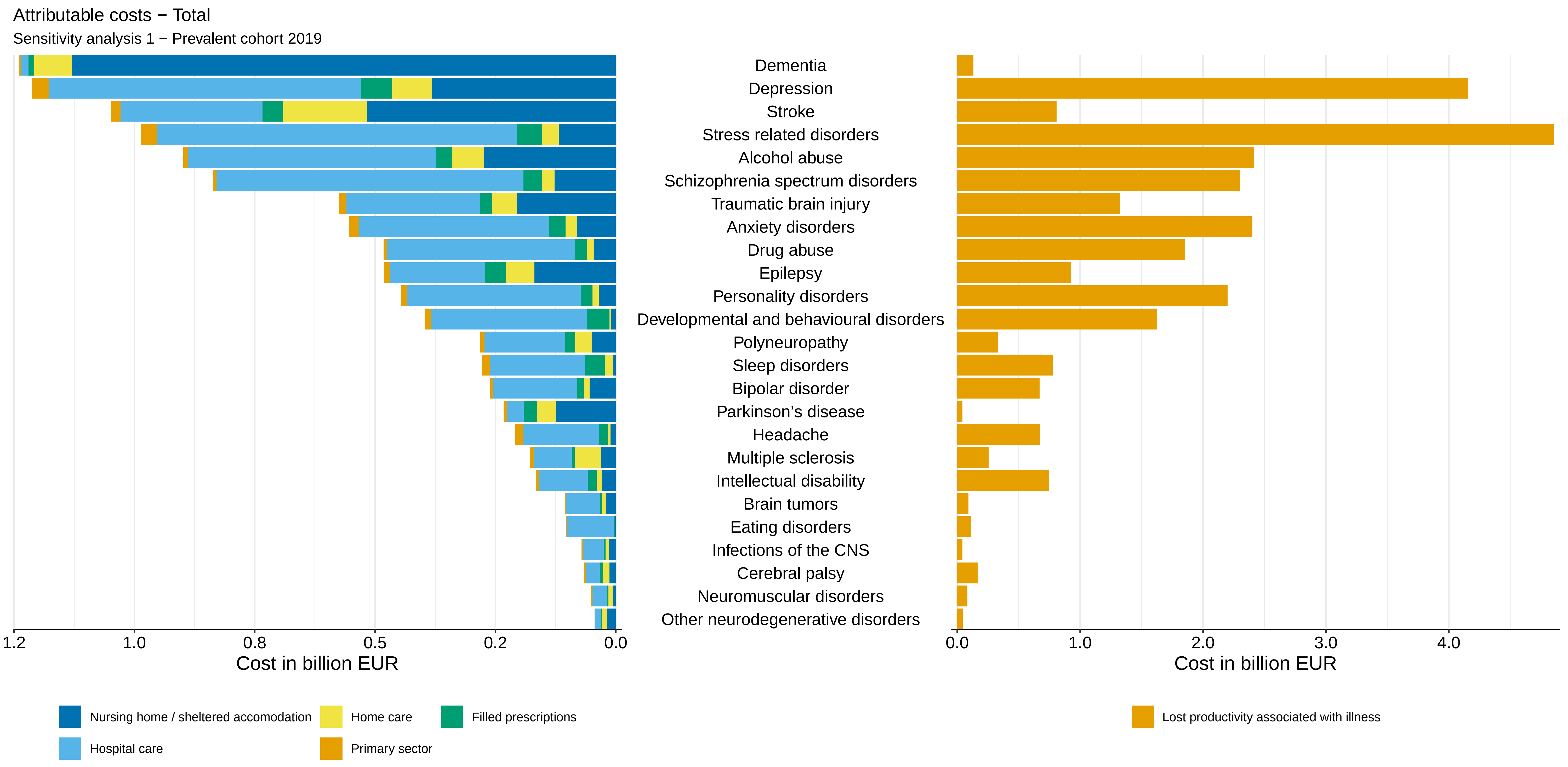

Attributable costs − Total
Sensitivity analysis 1 − Prevalent cohort 2019
Dementia
Depression
Stroke
Stress related disorders
Alcohol abuse
Schizophrenia spectrum disorders
Traumatic brain injury
Anxiety disorders
Drug abuse
Epilepsy
Personality disorders
Developmental and behavioural disorders
Polyneuropathy
Sleep disorders
Bipolar disorder
Parkinson’s disease
Headache
Multiple sclerosis
Intellectual disability
Brain tumors
Eating disorders
Infections of the CNS
Cerebral palsy
Neuromuscular disorders
Other neurodegenerative disorders
1.2
1.0
0.8
0.5
0.2
0.0
Cost in billion EUR
0.0
1.0
2.0
3.0
4.0
Cost in billion EUR
Nursing home / sheltered accomodation
Home care
Filled prescriptions
Hospital care
Primary sector
Lost productivity associated with illness

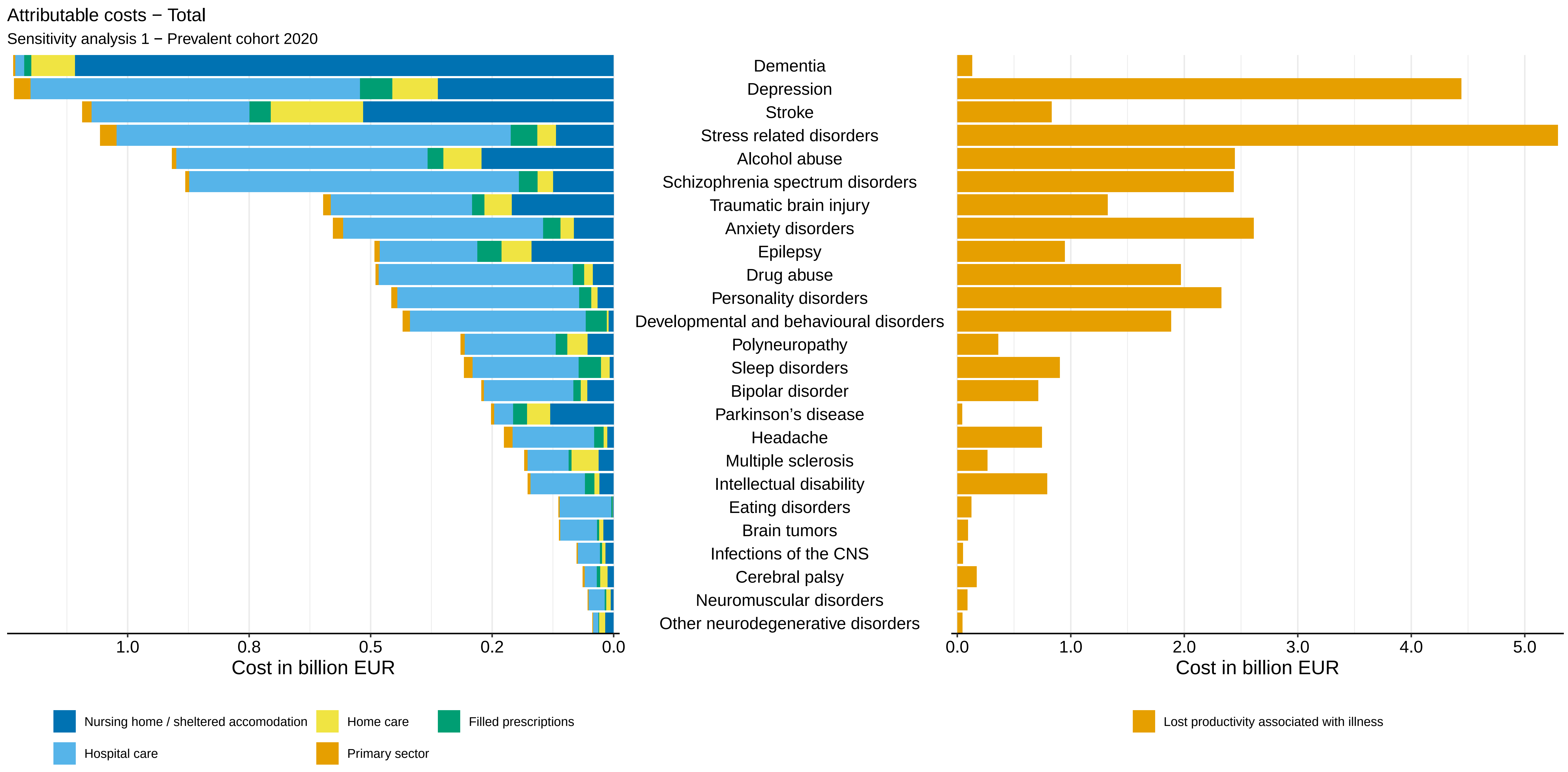

Attributable costs − Total
Sensitivity analysis 1 − Prevalent cohort 2020
Dementia
Depression
Stroke
Stress related disorders
Alcohol abuse
Schizophrenia spectrum disorders
Traumatic brain injury
Anxiety disorders
Epilepsy
Drug abuse
Personality disorders
Developmental and behavioural disorders
Polyneuropathy
Sleep disorders
Bipolar disorder
Parkinson's disease
Headache
Multiple sclerosis
Intellectual disability
Eating disorders
Brain tumors
Infections of the CNS
Cerebral palsy
Neuromuscular disorders
Other neurodegenerative disorders
1.0
0.8
0.5
0.2
0.0
Cost in billion EUR
0.0
1.0
2.0
3.0
4.0
5.0
Cost in billion EUR
Nursing home / sheltered accomodation
Home care
Filled prescriptions
Hospital care
Primary sector
Lost productivity associated with illness

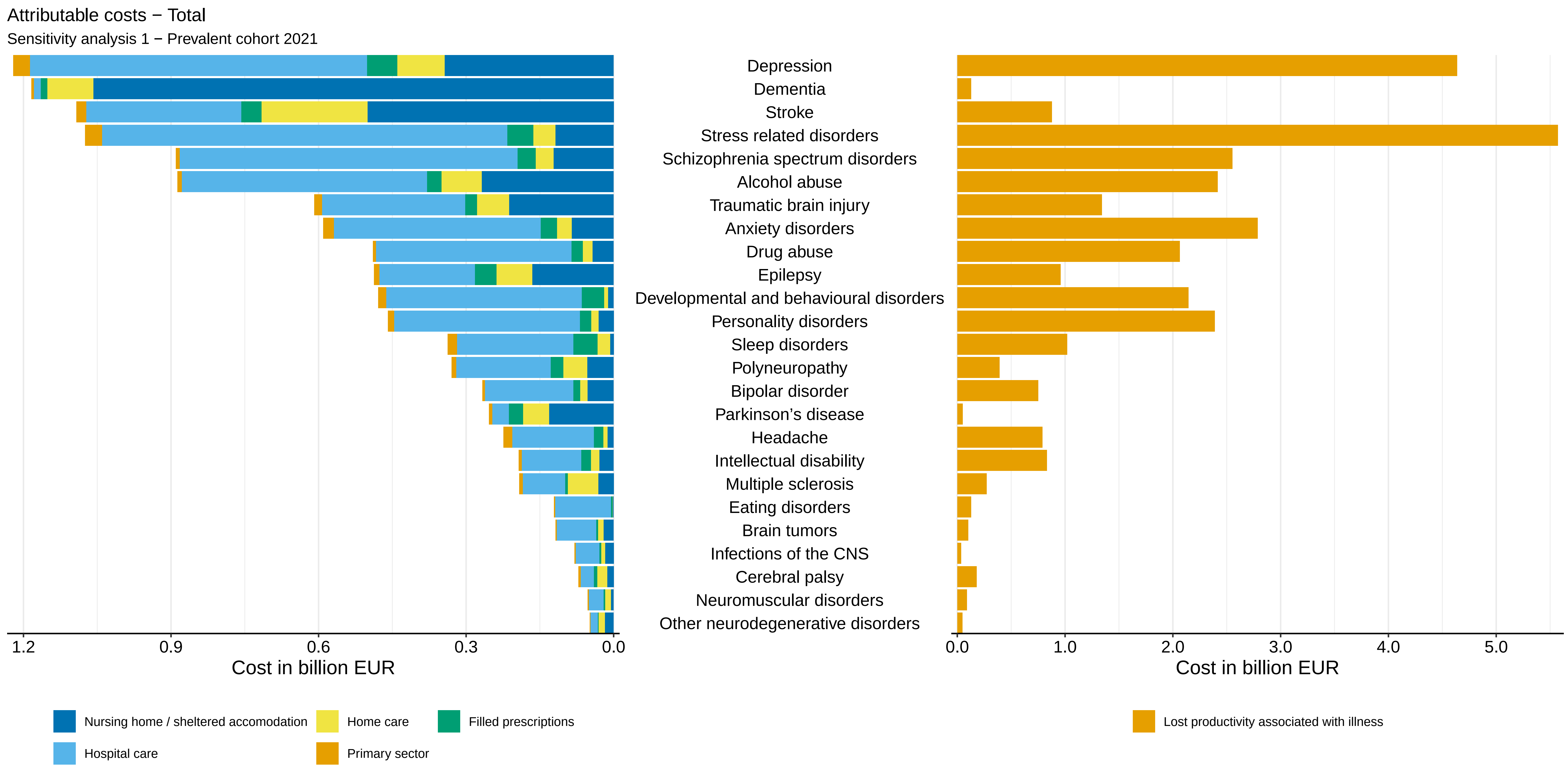
Attributable costs − Total
Sensitivity analysis 1 − Prevalent cohort 2021
Depression
Dementia
Stroke
Stress related disorders
Schizophrenia spectrum disorders
Alcohol abuse
Traumatic brain injury
Anxiety disorders
Drug abuse
Epilepsy
Developmental and behavioural disorders
Personality disorders
Sleep disorders
Polyneuropathy
Bipolar disorder
Parkinson’s disease
Headache
Intellectual disability
Multiple sclerosis
Eating disorders
Brain tumors
Infections of the CNS
Cerebral palsy
Neuromuscular disorders
Other neurodegenerative disorders
1.2
0.9
0.6
0.3
0.0
Cost in billion EUR
0.0
1.0
2.0
3.0
4.0
5.0
Cost in billion EUR
Nursing home / sheltered accomodation
Home care
Filled prescriptions
Hospital care
Primary sector
Lost productivity associated with illness

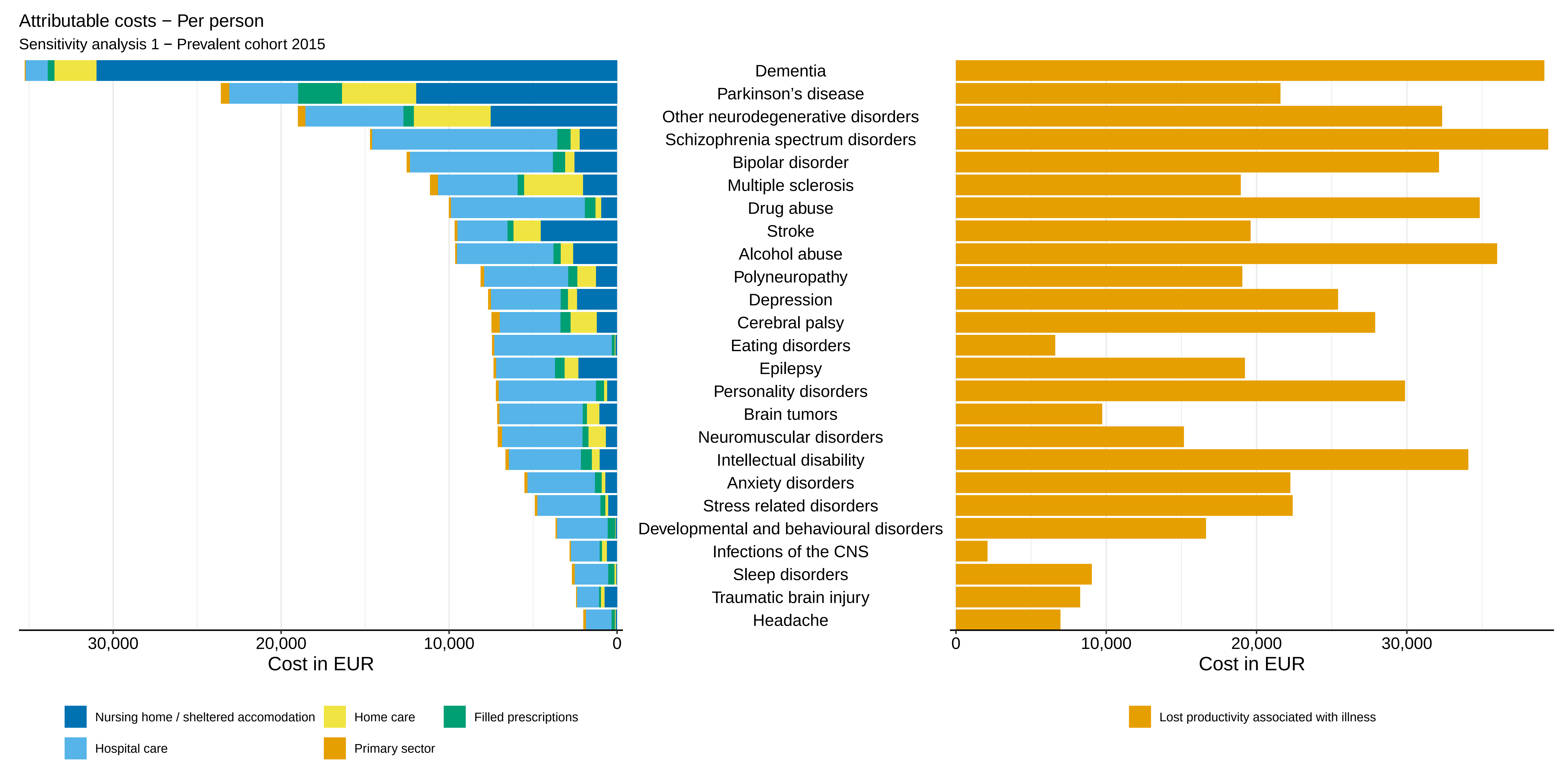

Attributable costs − Per person
Sensitivity analysis 1 − Prevalent cohort 2015
Dementia
Parkinson's disease
Other neurodegenerative disorders
Schizophrenia spectrum disorders
Bipolar disorder
Multiple sclerosis
Drug abuse
Stroke
Alcohol abuse
Polyneuropathy
Depression
Cerebral palsy
Eating disorders
Epilepsy
Personality disorders
Brain tumors
Neuromuscular disorders
Intellectual disability
Anxiety disorders
Stress related disorders
Developmental and behavioural disorders
Infections of the CNS
Sleep disorders
Traumatic brain injury
Headache
30,000
20,000
10,000
0
Cost in EUR
0
10,000
20,000
30,000
Cost in EUR
Nursing home / sheltered accomodation
Home care
Filled prescriptions
Hospital care
Primary sector
Lost productivity associated with illness

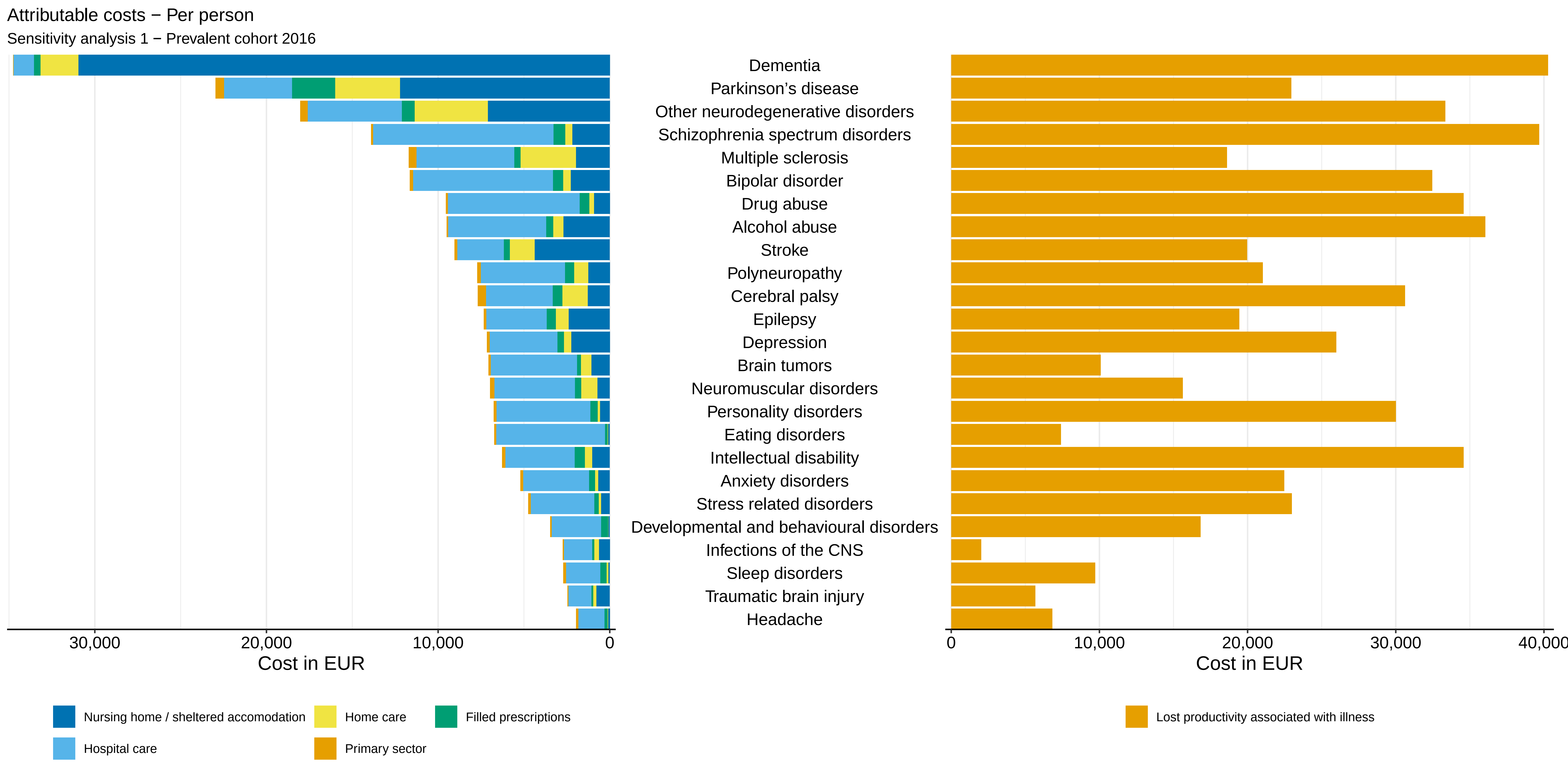
Attributable costs − Per person
Sensitivity analysis 1 − Prevalent cohort 2016
Dementia
Parkinson’s disease
Other neurodegenerative disorders
Schizophrenia spectrum disorders
Multiple sclerosis
Bipolar disorder
Drug abuse
Alcohol abuse
Stroke
Polyneuropathy
Cerebral palsy
Epilepsy
Depression
Brain tumors
Neuromuscular disorders
Personality disorders
Eating disorders
Intellectual disability
Anxiety disorders
Stress related disorders
Developmental and behavioural disorders
Infections of the CNS
Sleep disorders
Traumatic brain injury
Headache
30,000
20,000
10,000
0
Cost in EUR
0
10,000
20,000
30,000
40,000
Cost in EUR
Nursing home / sheltered accomodation
Home care
Filled prescriptions
Hospital care
Primary sector
Lost productivity associated with illness

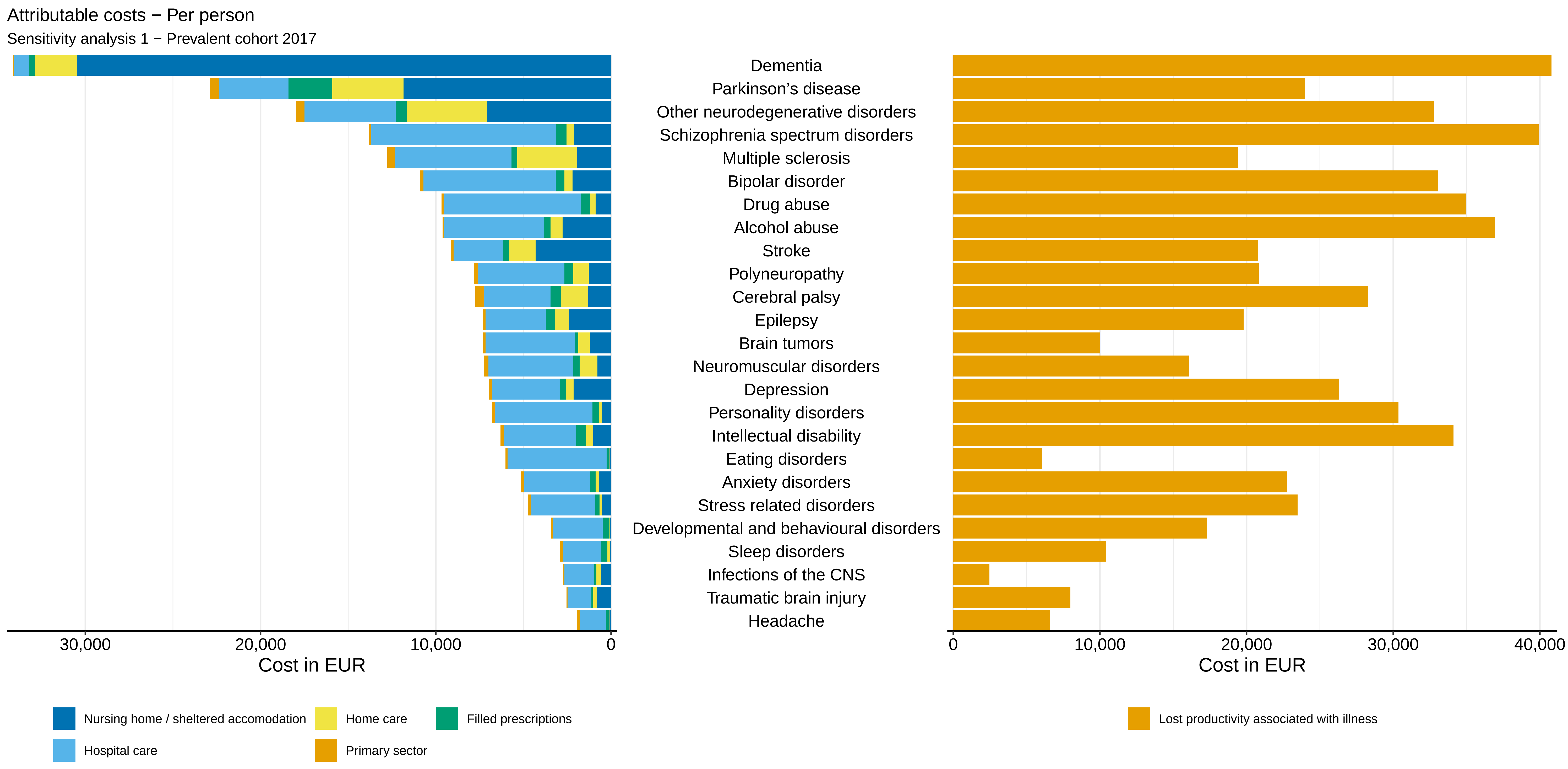

Attributable costs − Per person
Sensitivity analysis 1 − Prevalent cohort 2017
Dementia
Parkinson's disease
Other neurodegenerative disorders
Schizophrenia spectrum disorders
Multiple sclerosis
Bipolar disorder
Drug abuse
Alcohol abuse
Stroke
Polyneuropathy
Cerebral palsy
Epilepsy
Brain tumors
Neuromuscular disorders
Depression
Personality disorders
Intellectual disability
Eating disorders
Anxiety disorders
Stress related disorders
Developmental and behavioural disorders
Sleep disorders
Infections of the CNS
Traumatic brain injury
Headache
30,000
20,000
10,000
0
Cost in EUR
0
10,000
20,000
30,000
40,000
Cost in EUR
Nursing home / sheltered accomodation
Home care
Filled prescriptions
Hospital care
Primary sector
Lost productivity associated with illness

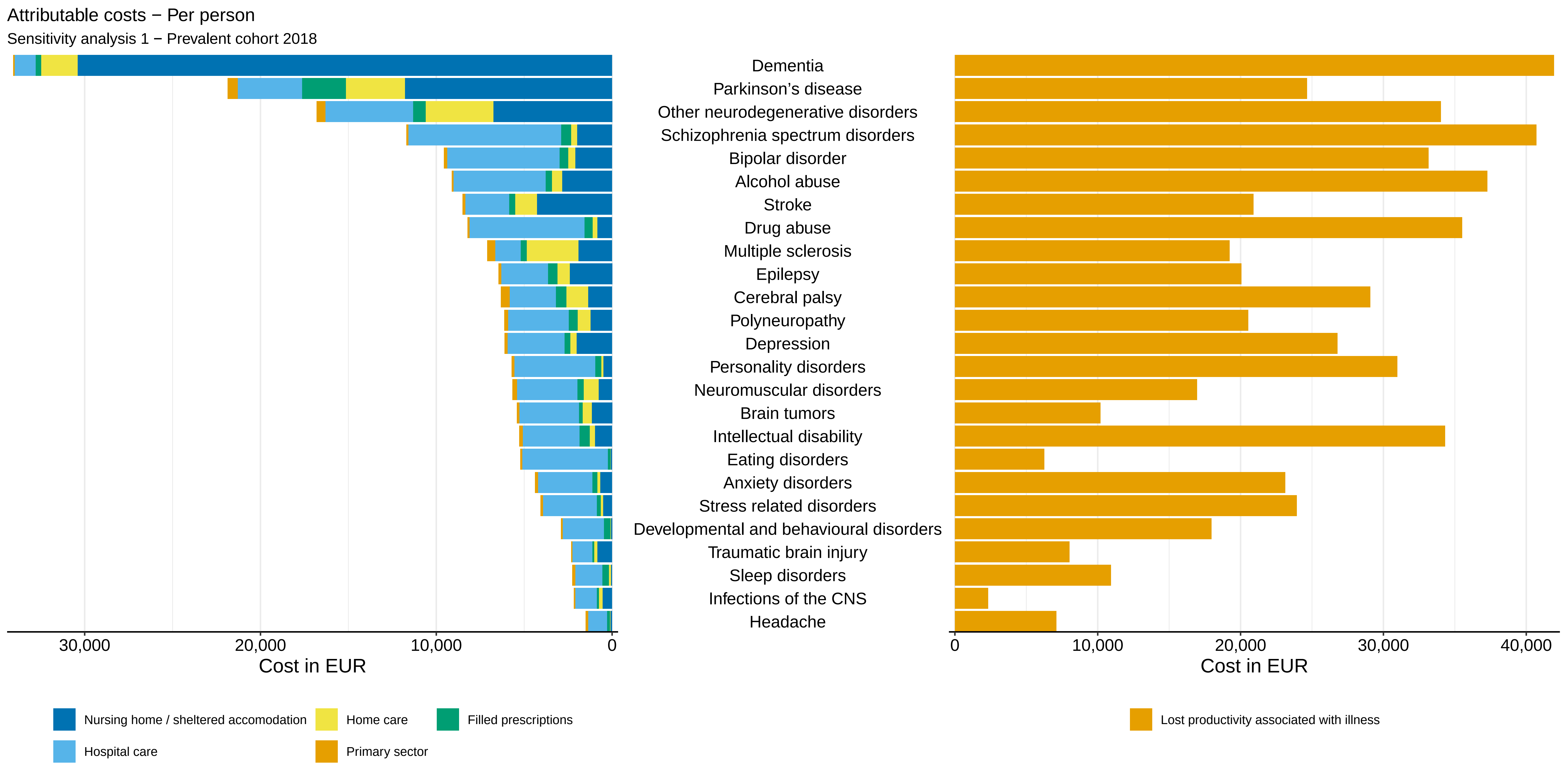
Attributable costs − Per person
Sensitivity analysis 1 − Prevalent cohort 2018
Dementia
Parkinson’s disease
Other neurodegenerative disorders
Schizophrenia spectrum disorders
Bipolar disorder
Alcohol abuse
Stroke
Drug abuse
Multiple sclerosis
Epilepsy
Cerebral palsy
Polyneuropathy
Depression
Personality disorders
Neuromuscular disorders
Brain tumors
Intellectual disability
Eating disorders
Anxiety disorders
Stress related disorders
Developmental and behavioural disorders
Traumatic brain injury
Sleep disorders
Infections of the CNS
Headache
30,000
20,000
10,000
0
Cost in EUR
0
10,000
20,000
30,000
40,000
Cost in EUR
Nursing home / sheltered accomodation
Home care
Filled prescriptions
Hospital care
Primary sector
Lost productivity associated with illness

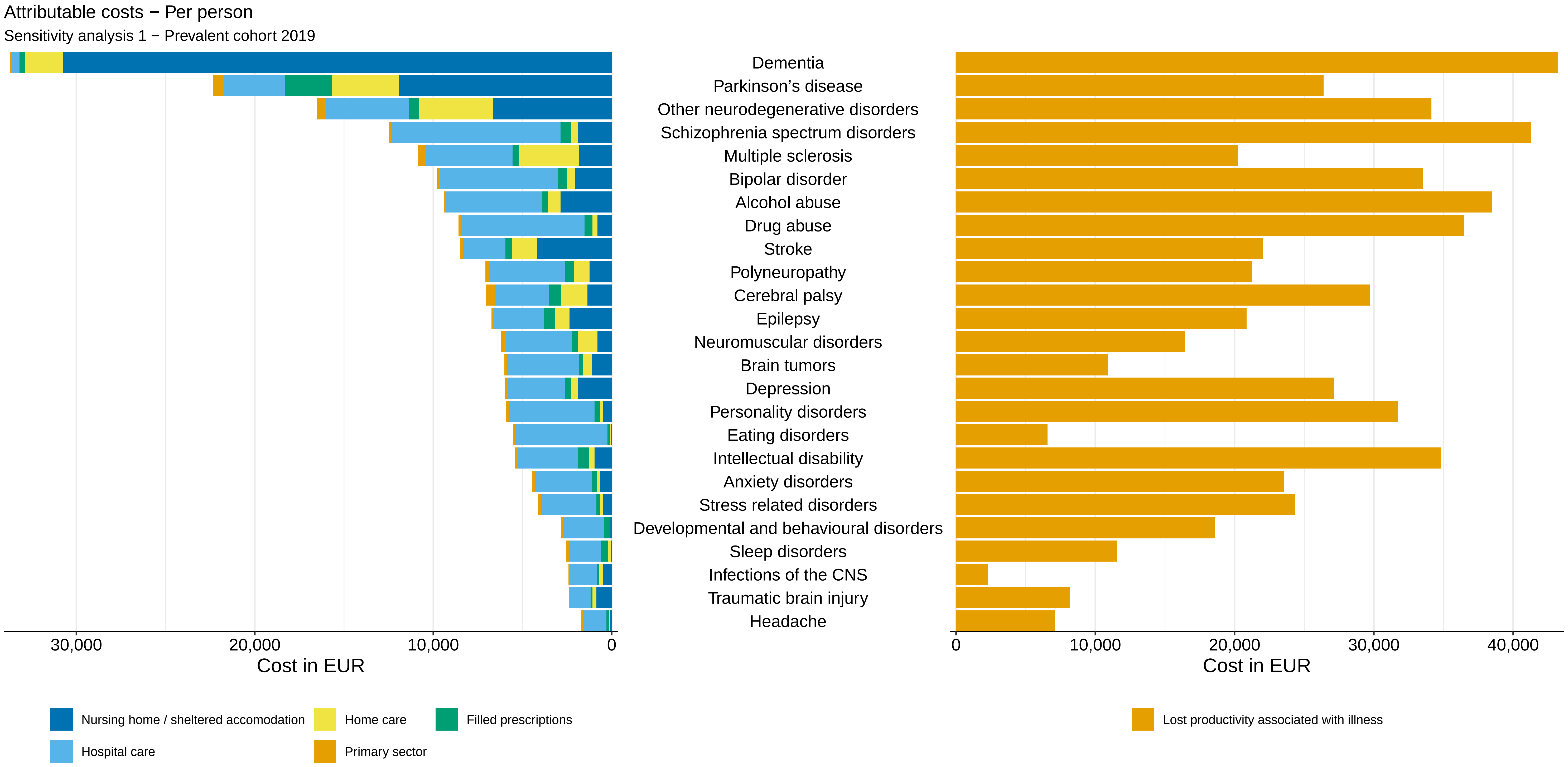

Attributable costs − Per person
Sensitivity analysis 1 − Prevalent cohort 2019
Dementia
Parkinson's disease
Other neurodegenerative disorders
Schizophrenia spectrum disorders
Multiple sclerosis
Bipolar disorder
Alcohol abuse
Drug abuse
Stroke
Polyneuropathy
Cerebral palsy
Epilepsy
Neuromuscular disorders
Brain tumors
Depression
Personality disorders
Eating disorders
Intellectual disability
Anxiety disorders
Stress related disorders
Developmental and behavioural disorders
Sleep disorders
Infections of the CNS
Traumatic brain injury
Headache
30,000
20,000
10,000
0
Cost in EUR
0
10,000
20,000
30,000
40,000
Cost in EUR
Nursing home / sheltered accomodation
Home care
Filled prescriptions
Hospital care
Primary sector
Lost productivity associated with illness

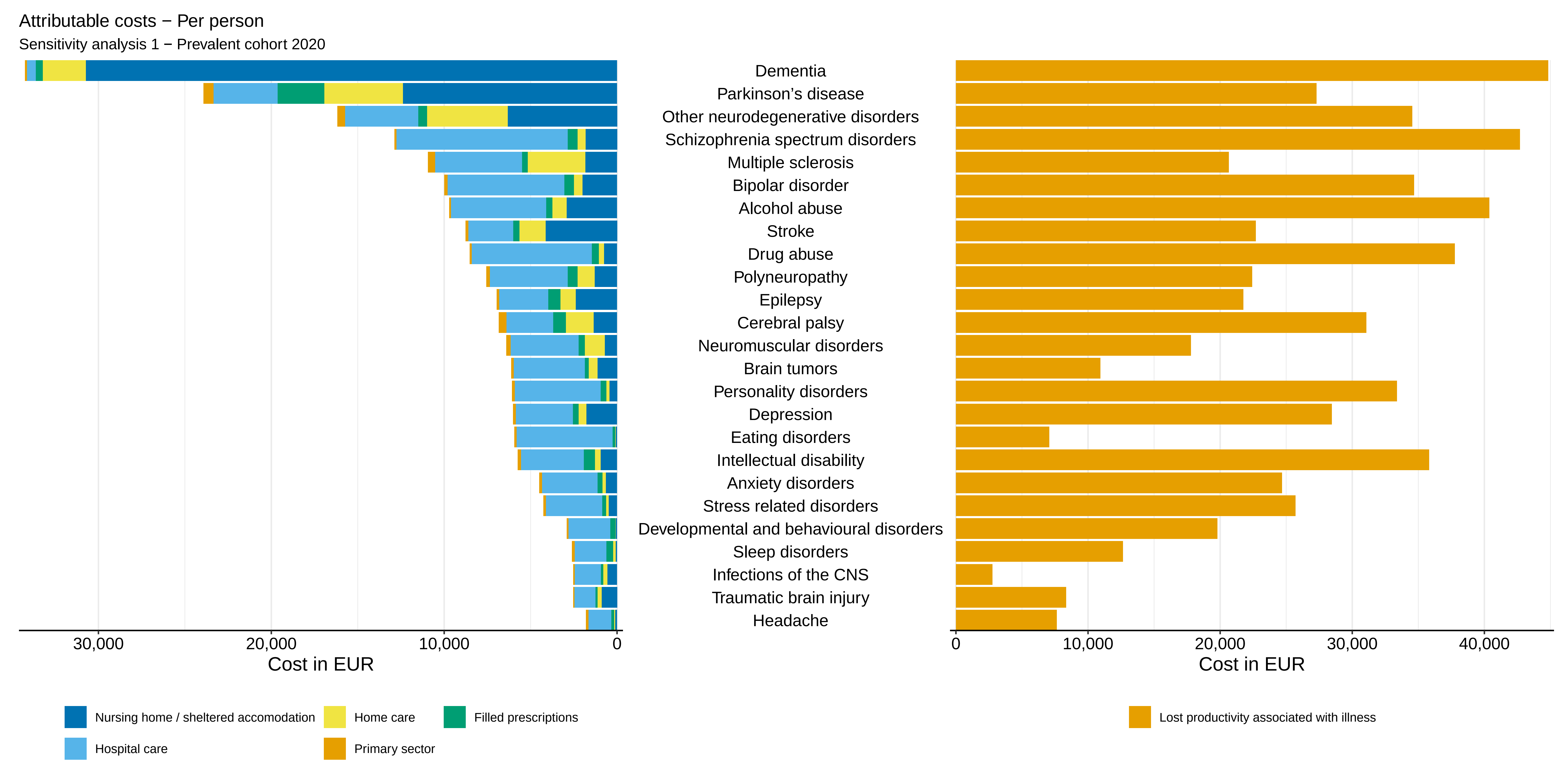

Attributable costs − Per person
Sensitivity analysis 1 − Prevalent cohort 2020
Dementia
Parkinson’s disease
Other neurodegenerative disorders
Schizophrenia spectrum disorders
Multiple sclerosis
Bipolar disorder
Alcohol abuse
Stroke
Drug abuse
Polyneuropathy
Epilepsy
Cerebral palsy
Neuromuscular disorders
Brain tumors
Personality disorders
Depression
Eating disorders
Intellectual disability
Anxiety disorders
Stress related disorders
Developmental and behavioural disorders
Sleep disorders
Infections of the CNS
Traumatic brain injury
Headache
30,000
20,000
10,000
0
Cost in EUR
0
10,000
20,000
30,000
40,000
Cost in EUR
Nursing home / sheltered accomodation
Home care
Filled prescriptions
Hospital care
Primary sector
Lost productivity associated with illness

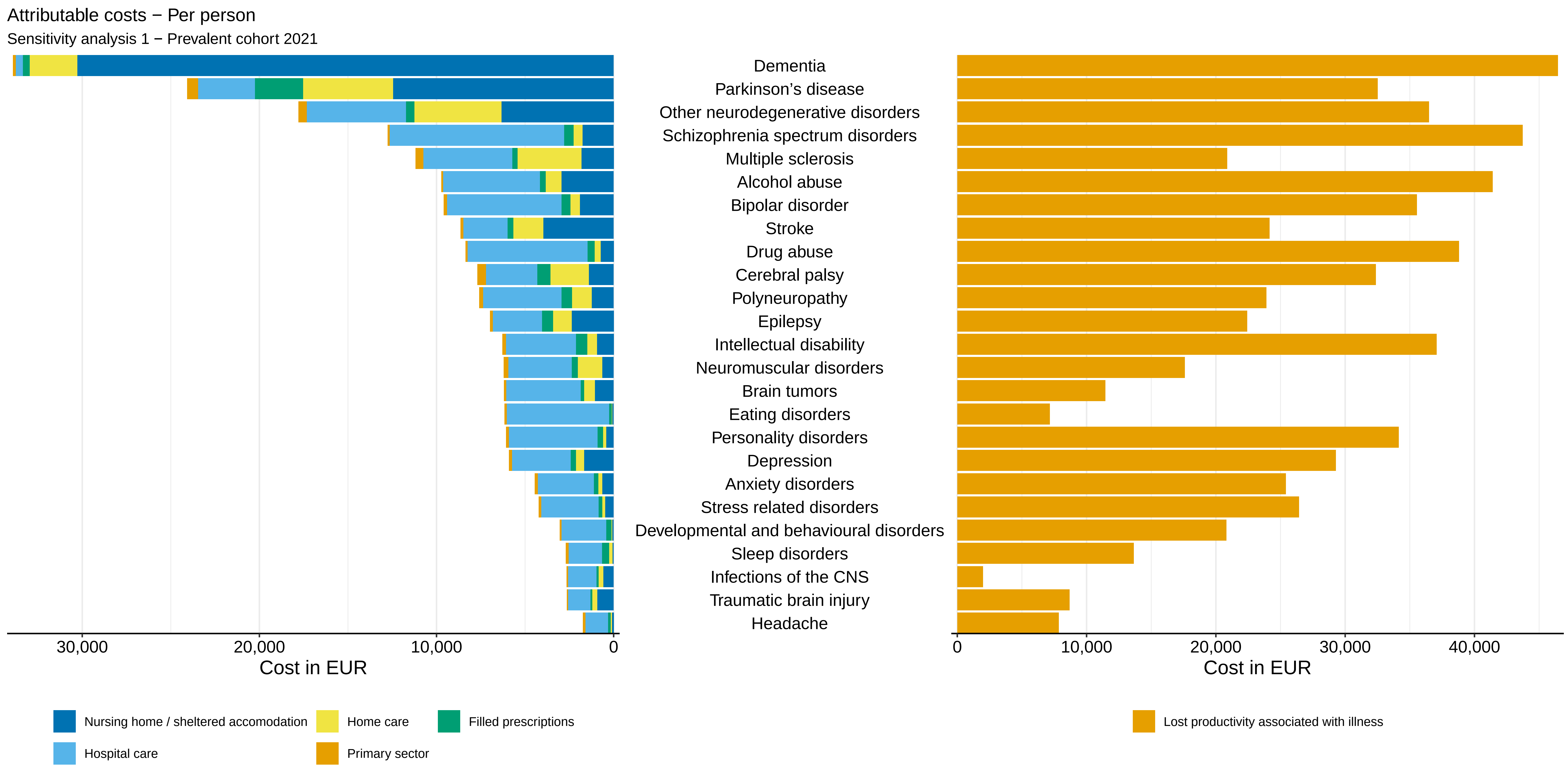
Attributable costs − Per person
Sensitivity analysis 1 − Prevalent cohort 2021
Dementia
Parkinson's disease
Other neurodegenerative disorders
Schizophrenia spectrum disorders
Multiple sclerosis
Alcohol abuse
Bipolar disorder
Stroke
Drug abuse
Cerebral palsy
Polyneuropathy
Epilepsy
Intellectual disability
Neuromuscular disorders
Brain tumors
Eating disorders
Personality disorders
Depression
Anxiety disorders
Stress related disorders
Developmental and behavioural disorders
Sleep disorders
Infections of the CNS
Traumatic brain injury
Headache
30,000
20,000
10,000
0
Cost in EUR
0
10,000
20,000
30,000
40,000
Cost in EUR
Nursing home / sheltered accomodation
Home care
Filled prescriptions
Hospital care
Primary sector
Lost productivity associated with illness

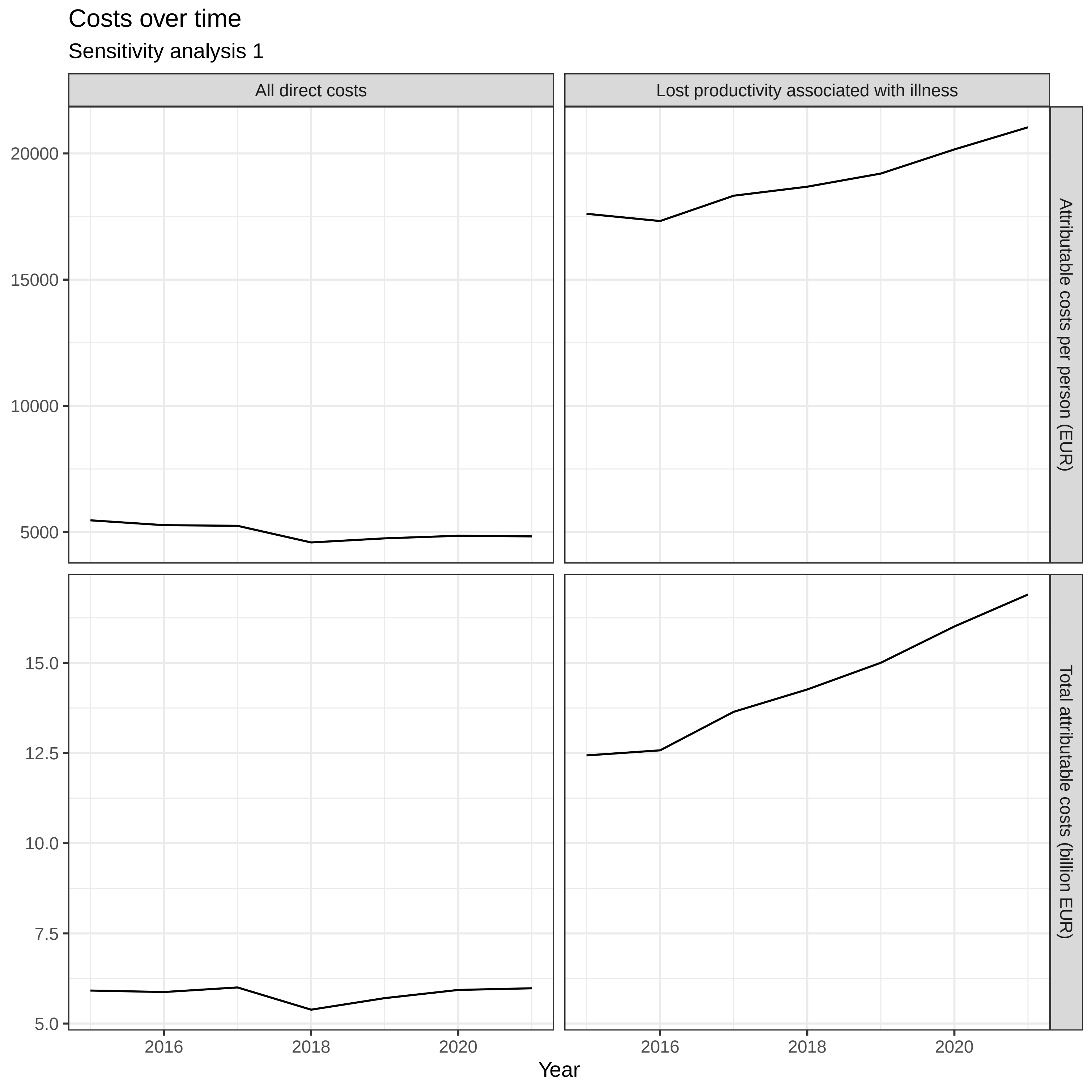
Costs over time
Sensitivity analysis 1
All direct costs
Lost productivity associated with illness
Attributable costs per person (EUR)
Total attributable costs (billion EUR)
20000
15000
10000
5000
15.0
12.5
10.0
7.5
5.0
2016
2018
2020
Year

***Sensitivity analysis 1 / Prevalent cohort 2015***

| | *CCI covariates included in model* | *No CCI covariates included in model* |
|---|---|---|
| Age 10-19 | -0.01 (-0.01 - -0.00) | -0.01 (-0.01 - -0.01) |
| Age 20-29 | 0.02 (0.02 - 0.02) | 0.01 (0.01 - 0.02) |
| Age 30-39 | 0.04 (0.03 - 0.04) | 0.04 (0.03 - 0.04) |
| Age 40-49 | 0.04 (0.03 - 0.04) | 0.05 (0.04 - 0.05) |
| Age 50-59 | 0.07 (0.07 - 0.08) | 0.12 (0.11 - 0.12) |
| Age 60-69 | 0.15 (0.15 - 0.15) | 0.26 (0.25 - 0.26) |
| Age 70-79 | 0.28 (0.27 - 0.28) | 0.47 (0.46 - 0.47) |
| Age 80-89 | 0.81 (0.80 - 0.82) | 1.08 (1.07 - 1.08) |
| Age 90+ | 2.21 (2.20 - 2.22) | 2.47 (2.46 - 2.48) |
| Myocardial infarction | 0.07 (0.06 - 0.08) | |
| Diabetes without end-organ damage | 0.28 (0.27 - 0.29) | |
| Hemiplegia | 1.16 (1.14 - 1.19) | |
| Moderate to severe renal disease | 0.78 (0.77 - 0.79) | |
| Diabetes with end-organ damage | 0.42 (0.41 - 0.43) | |
| Solid cancer tumour | 0.36 (0.35 - 0.36) | |
| Leukaemia | 1.09 (1.07 - 1.12) | |
| Lymphoma | 0.98 (0.96 - 1.00) | |
| Moderate to severe liver disease | 0.59 (0.56 - 0.62) | |
| Metastatic solid tumour | 1.06 (1.04 - 1.07) | |
| AIDS | 0.23 (0.20 - 0.27) | |
| Congestive heart failure | 0.51 (0.50 - 0.52) | |
| Peripheral vascular disease | 0.31 (0.30 - 0.32) | |
| Cerebrovascular disease | 0.40 (0.40 - 0.41) | |
| Chronic pulmonary disease | 0.29 (0.29 - 0.30) | |
| Connective tissue disease | 0.35 (0.35 - 0.36) | |
| Ulcer disease | 0.34 (0.33 - 0.35) | |
| Mild liver disease | 0.28 (0.26 - 0.29) | |
| Alcohol abuse | 0.30 (0.29 - 0.31) | 0.39 (0.39 - 0.40) |
| Anxiety disorders | 0.14 (0.13 - 0.15) | 0.16 (0.16 - 0.17) |
| Bipolar disorder | 0.42 (0.41 - 0.43) | 0.42 (0.41 - 0.44) |
| Brain tumors | 0.31 (0.30 - 0.32) | 0.37 (0.36 - 0.38) |
| Cerebral palsy | 0.25 (0.22 - 0.27) | 0.52 (0.50 - 0.54) |
| Dementia | 1.15 (1.15 - 1.16) | 1.16 (1.16 - 1.17) |
| Depression | 0.24 (0.23 - 0.24) | 0.27 (0.27 - 0.28) |
| Developmental and behavioural disorders | 0.05 (0.05 - 0.06) | 0.05 (0.04 - 0.05) |
| Drug abuse | 0.24 (0.23 - 0.25) | 0.25 (0.24 - 0.26) |
| Eating disorders | 0.41 (0.40 - 0.42) | 0.41 (0.40 - 0.43) |
| Epilepsy | 0.38 (0.37 - 0.38) | 0.43 (0.42 - 0.44) |
| Headache | 0.05 (0.05 - 0.06) | 0.07 (0.07 - 0.08) |
| Infections of the CNS | 0.15 (0.14 - 0.16) | 0.19 (0.18 - 0.20) |

***Sensitivity analysis 1 / Prevalent cohort 2015***

| | *CCI covariates included in model* | *No CCI covariates included in model* |
|---|---|---|
| Intellectual disability | 0.22 (0.21 - 0.24) | 0.24 (0.22 - 0.25) |
| Multiple sclerosis | 0.75 (0.74 - 0.77) | 0.77 (0.75 - 0.79) |
| Neuromuscular disorders | 0.36 (0.33 - 0.38) | 0.46 (0.44 - 0.48) |
| Other neurodegenerative disorders | 0.70 (0.67 - 0.73) | 0.74 (0.70 - 0.77) |
| Parkinson’s disease | 0.78 (0.76 - 0.79) | 0.78 (0.76 - 0.79) |
| Personality disorders | 0.13 (0.12 - 0.14) | 0.12 (0.11 - 0.13) |
| Polyneuropathy | 0.22 (0.22 - 0.23) | 0.42 (0.41 - 0.43) |
| Schizophrenia spectrum disorders | 0.78 (0.77 - 0.79) | 0.78 (0.77 - 0.79) |
| Sleep disorders | 0.09 (0.08 - 0.10) | 0.14 (0.13 - 0.15) |
| Stress related disorders | 0.07 (0.07 - 0.08) | 0.08 (0.07 - 0.08) |
| Stroke | 0.22 (0.21 - 0.23) | 0.45 (0.45 - 0.46) |
| Traumatic brain injury | 0.07 (0.07 - 0.08) | 0.08 (0.08 - 0.09) |
| Male | -0.05 (-0.05 - -0.05) | -0.04 (-0.04 - -0.04) |

***Sensitivity analysis 1 / Prevalent cohort 2016***

| | *CCI covariates included in model* | *No CCI covariates included in model* |
|---|---|---|
| Age 10-19 | -0.01 (-0.01 - -0.00) | -0.01 (-0.02 - -0.01) |
| Age 20-29 | 0.01 (0.01 - 0.02) | 0.01 (0.00 - 0.01) |
| Age 30-39 | 0.03 (0.03 - 0.04) | 0.03 (0.03 - 0.03) |
| Age 40-49 | 0.03 (0.03 - 0.03) | 0.04 (0.04 - 0.05) |
| Age 50-59 | 0.07 (0.06 - 0.07) | 0.11 (0.11 - 0.11) |
| Age 60-69 | 0.14 (0.14 - 0.14) | 0.24 (0.24 - 0.25) |
| Age 70-79 | 0.26 (0.26 - 0.27) | 0.45 (0.44 - 0.45) |
| Age 80-89 | 0.76 (0.75 - 0.77) | 1.02 (1.01 - 1.02) |
| Age 90+ | 2.13 (2.12 - 2.14) | 2.37 (2.36 - 2.38) |
| Myocardial infarction | 0.06 (0.05 - 0.07) | |
| Diabetes without end-organ damage | 0.28 (0.27 - 0.28) | |
| Hemiplegia | 1.10 (1.08 - 1.12) | |
| Moderate to severe renal disease | 0.71 (0.70 - 0.72) | |
| Diabetes with end-organ damage | 0.40 (0.39 - 0.41) | |
| Solid cancer tumour | 0.34 (0.33 - 0.34) | |
| Leukaemia | 0.94 (0.92 - 0.97) | |
| Lymphoma | 0.91 (0.89 - 0.92) | |
| Moderate to severe liver disease | 0.63 (0.60 - 0.66) | |
| Metastatic solid tumour | 0.98 (0.96 - 1.00) | |
| AIDS | 0.25 (0.22 - 0.28) | |
| Congestive heart failure | 0.51 (0.50 - 0.51) | |
| Peripheral vascular disease | 0.30 (0.29 - 0.31) | |
| Cerebrovascular disease | 0.35 (0.34 - 0.35) | |
| Chronic pulmonary disease | 0.28 (0.28 - 0.28) | |
| Connective tissue disease | 0.35 (0.34 - 0.35) | |
| Ulcer disease | 0.32 (0.30 - 0.33) | |
| Mild liver disease | 0.26 (0.25 - 0.27) | |
| Alcohol abuse | 0.31 (0.31 - 0.32) | 0.41 (0.40 - 0.42) |
| Anxiety disorders | 0.14 (0.13 - 0.14) | 0.16 (0.15 - 0.16) |
| Bipolar disorder | 0.40 (0.39 - 0.41) | 0.40 (0.39 - 0.41) |
| Brain tumors | 0.35 (0.34 - 0.36) | 0.41 (0.40 - 0.43) |
| Cerebral palsy | 0.26 (0.24 - 0.28) | 0.52 (0.50 - 0.54) |
| Dementia | 1.25 (1.25 - 1.26) | 1.27 (1.26 - 1.27) |
| Depression | 0.22 (0.22 - 0.23) | 0.26 (0.25 - 0.26) |
| Developmental and behavioural disorders | 0.06 (0.05 - 0.06) | 0.05 (0.05 - 0.06) |
| Drug abuse | 0.24 (0.24 - 0.25) | 0.25 (0.25 - 0.26) |
| Eating disorders | 0.40 (0.39 - 0.41) | 0.40 (0.39 - 0.42) |
| Epilepsy | 0.38 (0.37 - 0.39) | 0.43 (0.43 - 0.44) |
| Headache | 0.05 (0.05 - 0.06) | 0.08 (0.07 - 0.08) |
| Infections of the CNS | 0.16 (0.15 - 0.17) | 0.20 (0.19 - 0.21) |

***Sensitivity analysis 1 / Prevalent cohort 2016***

| | *CCI covariates included in model* | *No CCI covariates included in model* |
|---|---|---|
| Intellectual disability | 0.22 (0.21 - 0.24) | 0.24 (0.22 - 0.25) |
| Multiple sclerosis | 0.84 (0.82 - 0.85) | 0.86 (0.84 - 0.87) |
| Neuromuscular disorders | 0.36 (0.34 - 0.38) | 0.46 (0.44 - 0.48) |
| Other neurodegenerative disorders | 0.77 (0.74 - 0.80) | 0.81 (0.77 - 0.84) |
| Parkinson's disease | 0.83 (0.82 - 0.85) | 0.84 (0.82 - 0.85) |
| Personality disorders | 0.11 (0.10 - 0.12) | 0.10 (0.09 - 0.11) |
| Polyneuropathy | 0.23 (0.22 - 0.24) | 0.43 (0.42 - 0.44) |
| Schizophrenia spectrum disorders | 0.76 (0.75 - 0.77) | 0.76 (0.75 - 0.77) |
| Sleep disorders | 0.09 (0.09 - 0.10) | 0.15 (0.14 - 0.15) |
| Stress related disorders | 0.08 (0.08 - 0.09) | 0.09 (0.09 - 0.10) |
| Stroke | 0.25 (0.24 - 0.26) | 0.47 (0.47 - 0.48) |
| Traumatic brain injury | 0.08 (0.08 - 0.08) | 0.09 (0.09 - 0.10) |
| Male | -0.05 (-0.05 - -0.05) | -0.04 (-0.04 - -0.04) |

***Sensitivity analysis 1 / Prevalent cohort 2017***

| | *CCI covariates included in model* | *No CCI covariates included in model* |
|---|---|---|
| Age 10-19 | -0.01 (-0.01 - -0.00) | -0.01 (-0.01 - -0.00) |
| Age 20-29 | 0.02 (0.02 - 0.03) | 0.02 (0.01 - 0.02) |
| Age 30-39 | 0.04 (0.03 - 0.04) | 0.04 (0.03 - 0.04) |
| Age 40-49 | 0.04 (0.03 - 0.04) | 0.05 (0.05 - 0.06) |
| Age 50-59 | 0.08 (0.07 - 0.08) | 0.12 (0.12 - 0.12) |
| Age 60-69 | 0.16 (0.15 - 0.16) | 0.26 (0.26 - 0.27) |
| Age 70-79 | 0.28 (0.27 - 0.28) | 0.47 (0.47 - 0.47) |
| Age 80-89 | 0.74 (0.74 - 0.75) | 1.01 (1.00 - 1.01) |
| Age 90+ | 2.07 (2.06 - 2.08) | 2.32 (2.31 - 2.33) |
| Myocardial infarction | 0.06 (0.05 - 0.07) | |
| Diabetes without end-organ damage | 0.28 (0.28 - 0.29) | |
| Hemiplegia | 1.19 (1.17 - 1.22) | |
| Moderate to severe renal disease | 0.73 (0.72 - 0.74) | |
| Diabetes with end-organ damage | 0.42 (0.41 - 0.43) | |
| Solid cancer tumour | 0.36 (0.35 - 0.36) | |
| Leukaemia | 0.96 (0.94 - 0.99) | |
| Lymphoma | 0.97 (0.95 - 0.99) | |
| Moderate to severe liver disease | 0.65 (0.62 - 0.68) | |
| Metastatic solid tumour | 0.99 (0.97 - 1.00) | |
| AIDS | 0.23 (0.20 - 0.26) | |
| Congestive heart failure | 0.48 (0.47 - 0.49) | |
| Peripheral vascular disease | 0.30 (0.30 - 0.31) | |
| Cerebrovascular disease | 0.31 (0.30 - 0.31) | |
| Chronic pulmonary disease | 0.29 (0.28 - 0.29) | |
| Connective tissue disease | 0.34 (0.33 - 0.35) | |
| Ulcer disease | 0.31 (0.30 - 0.32) | |
| Mild liver disease | 0.26 (0.25 - 0.27) | |
| Alcohol abuse | 0.32 (0.31 - 0.33) | 0.42 (0.42 - 0.43) |
| Anxiety disorders | 0.14 (0.13 - 0.14) | 0.16 (0.15 - 0.17) |
| Bipolar disorder | 0.35 (0.34 - 0.37) | 0.36 (0.34 - 0.37) |
| Brain tumors | 0.38 (0.36 - 0.39) | 0.45 (0.44 - 0.46) |
| Cerebral palsy | 0.23 (0.21 - 0.26) | 0.51 (0.49 - 0.53) |
| Dementia | 1.37 (1.36 - 1.37) | 1.38 (1.37 - 1.38) |
| Depression | 0.22 (0.22 - 0.23) | 0.26 (0.25 - 0.26) |
| Developmental and behavioural disorders | 0.07 (0.06 - 0.07) | 0.06 (0.05 - 0.07) |
| Drug abuse | 0.25 (0.24 - 0.26) | 0.26 (0.25 - 0.27) |
| Eating disorders | 0.39 (0.38 - 0.41) | 0.39 (0.38 - 0.41) |
| Epilepsy | 0.38 (0.37 - 0.39) | 0.43 (0.43 - 0.44) |
| Headache | 0.05 (0.05 - 0.06) | 0.08 (0.07 - 0.08) |
| Infections of the CNS | 0.16 (0.15 - 0.17) | 0.20 (0.19 - 0.21) |

***Sensitivity analysis 1 / Prevalent cohort 2017***

| | *CCI covariates included in model* | *No CCI covariates included in model* |
|---|---|---|
| Intellectual disability | 0.21 (0.20 - 0.23) | 0.23 (0.22 - 0.24) |
| Multiple sclerosis | 0.96 (0.94 - 0.97) | 0.98 (0.96 - 0.99) |
| Neuromuscular disorders | 0.34 (0.32 - 0.36) | 0.46 (0.43 - 0.48) |
| Other neurodegenerative disorders | 0.81 (0.78 - 0.84) | 0.86 (0.83 - 0.89) |
| Parkinson’s disease | 0.94 (0.93 - 0.96) | 0.95 (0.94 - 0.97) |
| Personality disorders | 0.11 (0.10 - 0.12) | 0.10 (0.10 - 0.11) |
| Polyneuropathy | 0.27 (0.26 - 0.28) | 0.48 (0.48 - 0.49) |
| Schizophrenia spectrum disorders | 0.79 (0.78 - 0.80) | 0.79 (0.78 - 0.80) |
| Sleep disorders | 0.11 (0.10 - 0.12) | 0.17 (0.17 - 0.18) |
| Stress related disorders | 0.09 (0.08 - 0.09) | 0.10 (0.09 - 0.10) |
| Stroke | 0.31 (0.30 - 0.31) | 0.53 (0.52 - 0.53) |
| Traumatic brain injury | 0.09 (0.08 - 0.09) | 0.10 (0.10 - 0.10) |
| Male | -0.05 (-0.06 - -0.05) | -0.04 (-0.05 - -0.04) |

***Sensitivity analysis 1 / Prevalent cohort 2018***

| | *CCI covariates included in model* | *No CCI covariates included in model* |
|---|---|---|
| Age 10-19 | -0.01 (-0.01 - -0.00) | -0.01 (-0.01 - -0.00) |
| Age 20-29 | 0.01 (0.01 - 0.02) | 0.01 (0.00 - 0.01) |
| Age 30-39 | 0.02 (0.01 - 0.02) | 0.02 (0.01 - 0.02) |
| Age 40-49 | 0.01 (0.01 - 0.02) | 0.02 (0.02 - 0.03) |
| Age 50-59 | 0.05 (0.04 - 0.05) | 0.07 (0.07 - 0.08) |
| Age 60-69 | 0.12 (0.12 - 0.12) | 0.19 (0.19 - 0.20) |
| Age 70-79 | 0.23 (0.23 - 0.24) | 0.36 (0.36 - 0.37) |
| Age 80-89 | 0.69 (0.68 - 0.70) | 0.87 (0.87 - 0.88) |
| Age 90+ | 2.03 (2.02 - 2.04) | 2.20 (2.19 - 2.21) |
| Myocardial infarction | 0.04 (0.04 - 0.05) | |
| Diabetes without end-organ damage | 0.23 (0.23 - 0.24) | |
| Hemiplegia | 1.13 (1.11 - 1.16) | |
| Moderate to severe renal disease | 0.42 (0.41 - 0.43) | |
| Diabetes with end-organ damage | 0.32 (0.31 - 0.32) | |
| Solid cancer tumour | 0.18 (0.18 - 0.19) | |
| Leukaemia | 0.93 (0.90 - 0.95) | |
| Lymphoma | 0.52 (0.51 - 0.54) | |
| Moderate to severe liver disease | 0.61 (0.59 - 0.64) | |
| Metastatic solid tumour | 0.48 (0.46 - 0.49) | |
| AIDS | 0.06 (0.03 - 0.09) | |
| Congestive heart failure | 0.41 (0.41 - 0.42) | |
| Peripheral vascular disease | 0.25 (0.24 - 0.26) | |
| Cerebrovascular disease | 0.27 (0.26 - 0.27) | |
| Chronic pulmonary disease | 0.23 (0.23 - 0.24) | |
| Connective tissue disease | 0.12 (0.11 - 0.13) | |
| Ulcer disease | 0.31 (0.30 - 0.32) | |
| Mild liver disease | 0.20 (0.19 - 0.21) | |
| Alcohol abuse | 0.35 (0.35 - 0.36) | 0.44 (0.44 - 0.45) |
| Anxiety disorders | 0.10 (0.10 - 0.11) | 0.12 (0.11 - 0.12) |
| Bipolar disorder | 0.33 (0.31 - 0.34) | 0.33 (0.32 - 0.34) |
| Brain tumors | 0.34 (0.33 - 0.35) | 0.39 (0.37 - 0.40) |
| Cerebral palsy | 0.12 (0.10 - 0.14) | 0.38 (0.36 - 0.40) |
| Dementia | 1.52 (1.51 - 1.52) | 1.53 (1.52 - 1.54) |
| Depression | 0.20 (0.20 - 0.20) | 0.23 (0.22 - 0.23) |
| Developmental and behavioural disorders | 0.07 (0.06 - 0.07) | 0.06 (0.05 - 0.06) |
| Drug abuse | 0.21 (0.21 - 0.22) | 0.22 (0.21 - 0.23) |
| Eating disorders | 0.32 (0.31 - 0.33) | 0.32 (0.31 - 0.33) |
| Epilepsy | 0.33 (0.33 - 0.34) | 0.38 (0.37 - 0.38) |
| Headache | 0.03 (0.02 - 0.04) | 0.05 (0.04 - 0.05) |
| Infections of the CNS | 0.16 (0.15 - 0.17) | 0.18 (0.17 - 0.19) |

***Sensitivity analysis 1 / Prevalent cohort 2018***

| | *CCI covariates included in model* | *No CCI covariates included in model* |
|---|---|---|
| Intellectual disability | 0.17 (0.15 - 0.18) | 0.18 (0.17 - 0.19) |
| Multiple sclerosis | 0.51 (0.50 - 0.53) | 0.53 (0.52 - 0.55) |
| Neuromuscular disorders | 0.28 (0.26 - 0.30) | 0.36 (0.34 - 0.38) |
| Other neurodegenerative disorders | 0.84 (0.81 - 0.87) | 0.89 (0.86 - 0.92) |
| Parkinson’s disease | 1.00 (0.99 - 1.02) | 1.01 (0.99 - 1.02) |
| Personality disorders | 0.08 (0.08 - 0.09) | 0.08 (0.07 - 0.09) |
| Polyneuropathy | 0.20 (0.19 - 0.21) | 0.37 (0.36 - 0.38) |
| Schizophrenia spectrum disorders | 0.69 (0.69 - 0.70) | 0.69 (0.69 - 0.70) |
| Sleep disorders | 0.08 (0.08 - 0.09) | 0.14 (0.13 - 0.14) |
| Stress related disorders | 0.09 (0.08 - 0.09) | 0.09 (0.09 - 0.10) |
| Stroke | 0.33 (0.33 - 0.34) | 0.53 (0.52 - 0.53) |
| Traumatic brain injury | 0.08 (0.08 - 0.09) | 0.10 (0.09 - 0.10) |
| Male | -0.04 (-0.04 - -0.04) | -0.03 (-0.04 - -0.03) |

***Sensitivity analysis 1 / Prevalent cohort 2019***

| | *CCI covariates included in model* | *No CCI covariates included in model* |
|---|---|---|
| Age 10-19 | -0.01 (-0.01 - -0.01) | -0.01 (-0.01 - -0.01) |
| Age 20-29 | 0.02 (0.02 - 0.03) | 0.02 (0.02 - 0.02) |
| Age 30-39 | 0.03 (0.03 - 0.04) | 0.03 (0.03 - 0.04) |
| Age 40-49 | 0.02 (0.02 - 0.03) | 0.04 (0.03 - 0.04) |
| Age 50-59 | 0.05 (0.05 - 0.06) | 0.09 (0.09 - 0.10) |
| Age 60-69 | 0.13 (0.13 - 0.13) | 0.22 (0.22 - 0.23) |
| Age 70-79 | 0.25 (0.25 - 0.26) | 0.42 (0.41 - 0.42) |
| Age 80-89 | 0.68 (0.68 - 0.69) | 0.90 (0.90 - 0.91) |
| Age 90+ | 2.04 (2.03 - 2.05) | 2.25 (2.24 - 2.26) |
| Myocardial infarction | 0.04 (0.03 - 0.05) | |
| Diabetes without end-organ damage | 0.26 (0.26 - 0.27) | |
| Hemiplegia | 1.17 (1.15 - 1.20) | |
| Moderate to severe renal disease | 0.58 (0.57 - 0.59) | |
| Diabetes with end-organ damage | 0.39 (0.38 - 0.40) | |
| Solid cancer tumour | 0.29 (0.28 - 0.29) | |
| Leukaemia | 0.85 (0.83 - 0.87) | |
| Lymphoma | 0.88 (0.87 - 0.90) | |
| Moderate to severe liver disease | 0.61 (0.58 - 0.64) | |
| Metastatic solid tumour | 0.80 (0.79 - 0.82) | |
| AIDS | 0.14 (0.11 - 0.17) | |
| Congestive heart failure | 0.42 (0.41 - 0.43) | |
| Peripheral vascular disease | 0.27 (0.26 - 0.27) | |
| Cerebrovascular disease | 0.20 (0.19 - 0.21) | |
| Chronic pulmonary disease | 0.26 (0.26 - 0.27) | |
| Connective tissue disease | 0.23 (0.22 - 0.24) | |
| Ulcer disease | 0.28 (0.27 - 0.29) | |
| Mild liver disease | 0.21 (0.20 - 0.22) | |
| Alcohol abuse | 0.37 (0.36 - 0.38) | 0.47 (0.46 - 0.47) |
| Anxiety disorders | 0.09 (0.09 - 0.10) | 0.11 (0.10 - 0.11) |
| Bipolar disorder | 0.37 (0.36 - 0.38) | 0.37 (0.36 - 0.39) |
| Brain tumors | 0.39 (0.38 - 0.40) | 0.46 (0.45 - 0.47) |
| Cerebral palsy | 0.16 (0.14 - 0.18) | 0.42 (0.40 - 0.44) |
| Dementia | 1.67 (1.66 - 1.68) | 1.68 (1.67 - 1.69) |
| Depression | 0.18 (0.18 - 0.19) | 0.21 (0.21 - 0.22) |
| Developmental and behavioural disorders | 0.06 (0.05 - 0.06) | 0.05 (0.04 - 0.05) |
| Drug abuse | 0.24 (0.23 - 0.25) | 0.25 (0.24 - 0.26) |
| Eating disorders | 0.37 (0.35 - 0.38) | 0.37 (0.36 - 0.38) |
| Epilepsy | 0.36 (0.35 - 0.37) | 0.40 (0.40 - 0.41) |
| Headache | 0.04 (0.04 - 0.05) | 0.06 (0.06 - 0.07) |
| Infections of the CNS | 0.17 (0.16 - 0.18) | 0.20 (0.19 - 0.22) |

***Sensitivity analysis 1 / Prevalent cohort 2019***

| | *CCI covariates included in model* | *No CCI covariates included in model* |
|---|---|---|
| Intellectual disability | 0.16 (0.15 - 0.17) | 0.18 (0.16 - 0.19) |
| Multiple sclerosis | 0.86 (0.84 - 0.87) | 0.87 (0.86 - 0.89) |
| Neuromuscular disorders | 0.32 (0.30 - 0.34) | 0.42 (0.40 - 0.44) |
| Other neurodegenerative disorders | 0.89 (0.86 - 0.92) | 0.94 (0.91 - 0.98) |
| Parkinson’s disease | 1.14 (1.12 - 1.15) | 1.15 (1.13 - 1.16) |
| Personality disorders | 0.09 (0.08 - 0.10) | 0.09 (0.08 - 0.09) |
| Polyneuropathy | 0.26 (0.25 - 0.27) | 0.47 (0.46 - 0.48) |
| Schizophrenia spectrum disorders | 0.78 (0.77 - 0.78) | 0.78 (0.77 - 0.79) |
| Sleep disorders | 0.11 (0.10 - 0.11) | 0.17 (0.16 - 0.18) |
| Stress related disorders | 0.09 (0.08 - 0.09) | 0.10 (0.09 - 0.10) |
| Stroke | 0.41 (0.41 - 0.42) | 0.60 (0.59 - 0.60) |
| Traumatic brain injury | 0.09 (0.09 - 0.10) | 0.10 (0.10 - 0.11) |
| Male | -0.05 (-0.05 - -0.05) | -0.04 (-0.04 - -0.04) |

***Sensitivity analysis 1 / Prevalent cohort 2020***

| | *CCI covariates included in model* | *No CCI covariates included in model* |
|---|---|---|
| Age 10-19 | -0.00 (-0.01 - 0.00) | -0.00 (-0.01 - 0.00) |
| Age 20-29 | 0.03 (0.03 - 0.03) | 0.03 (0.02 - 0.03) |
| Age 30-39 | 0.04 (0.03 - 0.04) | 0.04 (0.03 - 0.04) |
| Age 40-49 | 0.02 (0.02 - 0.03) | 0.04 (0.04 - 0.04) |
| Age 50-59 | 0.06 (0.05 - 0.06) | 0.10 (0.10 - 0.11) |
| Age 60-69 | 0.14 (0.14 - 0.14) | 0.24 (0.23 - 0.24) |
| Age 70-79 | 0.27 (0.27 - 0.28) | 0.45 (0.44 - 0.45) |
| Age 80-89 | 0.69 (0.69 - 0.70) | 0.93 (0.92 - 0.93) |
| Age 90+ | 2.04 (2.03 - 2.05) | 2.26 (2.25 - 2.27) |
| Myocardial infarction | 0.05 (0.05 - 0.06) | |
| Diabetes without end-organ damage | 0.27 (0.26 - 0.27) | |
| Hemiplegia | 1.19 (1.17 - 1.21) | |
| Moderate to severe renal disease | 0.64 (0.63 - 0.64) | |
| Diabetes with end-organ damage | 0.38 (0.37 - 0.39) | |
| Solid cancer tumour | 0.32 (0.32 - 0.33) | |
| Leukaemia | 0.90 (0.88 - 0.92) | |
| Lymphoma | 0.97 (0.96 - 0.99) | |
| Moderate to severe liver disease | 0.64 (0.61 - 0.67) | |
| Metastatic solid tumour | 0.89 (0.88 - 0.91) | |
| AIDS | 0.12 (0.09 - 0.15) | |
| Congestive heart failure | 0.43 (0.42 - 0.44) | |
| Peripheral vascular disease | 0.28 (0.28 - 0.29) | |
| Cerebrovascular disease | 0.15 (0.14 - 0.16) | |
| Chronic pulmonary disease | 0.26 (0.25 - 0.26) | |
| Connective tissue disease | 0.24 (0.23 - 0.25) | |
| Ulcer disease | 0.32 (0.30 - 0.33) | |
| Mild liver disease | 0.22 (0.21 - 0.23) | |
| Alcohol abuse | 0.40 (0.39 - 0.40) | 0.50 (0.49 - 0.50) |
| Anxiety disorders | 0.10 (0.09 - 0.10) | 0.11 (0.10 - 0.11) |
| Bipolar disorder | 0.38 (0.37 - 0.40) | 0.39 (0.38 - 0.40) |
| Brain tumors | 0.43 (0.42 - 0.44) | 0.51 (0.50 - 0.52) |
| Cerebral palsy | 0.14 (0.12 - 0.16) | 0.40 (0.37 - 0.42) |
| Dementia | 1.92 (1.91 - 1.92) | 1.92 (1.91 - 1.93) |
| Depression | 0.19 (0.19 - 0.20) | 0.22 (0.21 - 0.22) |
| Developmental and behavioural disorders | 0.06 (0.06 - 0.07) | 0.06 (0.05 - 0.06) |
| Drug abuse | 0.23 (0.22 - 0.24) | 0.24 (0.23 - 0.25) |
| Eating disorders | 0.42 (0.40 - 0.43) | 0.42 (0.40 - 0.43) |
| Epilepsy | 0.39 (0.38 - 0.40) | 0.43 (0.42 - 0.44) |
| Headache | 0.05 (0.04 - 0.05) | 0.07 (0.06 - 0.07) |
| Infections of the CNS | 0.17 (0.15 - 0.18) | 0.20 (0.19 - 0.21) |

***Sensitivity analysis 1 / Prevalent cohort 2020***

| | *CCI covariates included in model* | *No CCI covariates included in model* |
|---|---|---|
| Intellectual disability | 0.18 (0.17 - 0.20) | 0.20 (0.18 - 0.21) |
| Multiple sclerosis | 0.90 (0.89 - 0.92) | 0.92 (0.91 - 0.94) |
| Neuromuscular disorders | 0.36 (0.34 - 0.38) | 0.46 (0.44 - 0.48) |
| Other neurodegenerative disorders | 0.96 (0.92 - 0.99) | 1.02 (0.98 - 1.05) |
| Parkinson's disease | 1.35 (1.33 - 1.37) | 1.36 (1.34 - 1.38) |
| Personality disorders | 0.10 (0.09 - 0.11) | 0.09 (0.09 - 0.10) |
| Polyneuropathy | 0.30 (0.29 - 0.31) | 0.53 (0.52 - 0.54) |
| Schizophrenia spectrum disorders | 0.85 (0.84 - 0.86) | 0.85 (0.84 - 0.86) |
| Sleep disorders | 0.11 (0.11 - 0.12) | 0.18 (0.17 - 0.19) |
| Stress related disorders | 0.09 (0.09 - 0.09) | 0.10 (0.10 - 0.11) |
| Stroke | 0.52 (0.51 - 0.52) | 0.69 (0.68 - 0.69) |
| Traumatic brain injury | 0.11 (0.11 - 0.11) | 0.12 (0.12 - 0.13) |
| Male | -0.05 (-0.05 - -0.05) | -0.04 (-0.04 - -0.04) |

***Sensitivity analysis 1 / Prevalent cohort 2021***

| | *CCI covariates included in model* | *No CCI covariates included in model* |
|---|---|---|
| Age 10-19 | -0.00 (-0.01 - 0.00) | -0.00 (-0.00 - 0.00) |
| Age 20-29 | 0.03 (0.03 - 0.03) | 0.03 (0.02 - 0.03) |
| Age 30-39 | 0.03 (0.03 - 0.04) | 0.04 (0.03 - 0.04) |
| Age 40-49 | 0.02 (0.02 - 0.03) | 0.04 (0.03 - 0.04) |
| Age 50-59 | 0.06 (0.05 - 0.06) | 0.10 (0.10 - 0.10) |
| Age 60-69 | 0.14 (0.13 - 0.14) | 0.23 (0.23 - 0.23) |
| Age 70-79 | 0.28 (0.27 - 0.28) | 0.44 (0.44 - 0.45) |
| Age 80-89 | 0.69 (0.69 - 0.70) | 0.92 (0.91 - 0.92) |
| Age 90+ | 2.03 (2.01 - 2.04) | 2.23 (2.22 - 2.25) |
| Myocardial infarction | 0.05 (0.04 - 0.06) | |
| Diabetes without end-organ damage | 0.28 (0.28 - 0.29) | |
| Hemiplegia | 1.26 (1.24 - 1.29) | |
| Moderate to severe renal disease | 0.63 (0.62 - 0.64) | |
| Diabetes with end-organ damage | 0.37 (0.36 - 0.38) | |
| Solid cancer tumour | 0.30 (0.29 - 0.30) | |
| Leukaemia | 0.80 (0.78 - 0.82) | |
| Lymphoma | 0.96 (0.95 - 0.98) | |
| Moderate to severe liver disease | 0.60 (0.57 - 0.63) | |
| Metastatic solid tumour | 0.90 (0.89 - 0.92) | |
| AIDS | 0.11 (0.08 - 0.14) | |
| Congestive heart failure | 0.44 (0.43 - 0.45) | |
| Peripheral vascular disease | 0.28 (0.27 - 0.29) | |
| Cerebrovascular disease | 0.08 (0.07 - 0.08) | |
| Chronic pulmonary disease | 0.26 (0.25 - 0.26) | |
| Connective tissue disease | 0.23 (0.22 - 0.24) | |
| Ulcer disease | 0.33 (0.32 - 0.34) | |
| Mild liver disease | 0.21 (0.20 - 0.22) | |
| Alcohol abuse | 0.41 (0.41 - 0.42) | 0.51 (0.51 - 0.52) |
| Anxiety disorders | 0.10 (0.09 - 0.10) | 0.11 (0.10 - 0.11) |
| Bipolar disorder | 0.37 (0.35 - 0.38) | 0.37 (0.36 - 0.39) |
| Brain tumors | 0.51 (0.49 - 0.52) | 0.59 (0.57 - 0.60) |
| Cerebral palsy | 0.20 (0.18 - 0.23) | 0.47 (0.45 - 0.49) |
| Dementia | 2.13 (2.12 - 2.14) | 2.13 (2.12 - 2.14) |
| Depression | 0.20 (0.19 - 0.20) | 0.22 (0.22 - 0.23) |
| Developmental and behavioural disorders | 0.09 (0.08 - 0.09) | 0.08 (0.08 - 0.09) |
| Drug abuse | 0.21 (0.20 - 0.22) | 0.21 (0.20 - 0.22) |
| Eating disorders | 0.48 (0.46 - 0.49) | 0.48 (0.46 - 0.49) |
| Epilepsy | 0.37 (0.36 - 0.38) | 0.41 (0.40 - 0.42) |
| Headache | 0.04 (0.03 - 0.05) | 0.06 (0.05 - 0.06) |
| Infections of the CNS | 0.18 (0.17 - 0.19) | 0.22 (0.21 - 0.23) |

***Sensitivity analysis 1 / Prevalent cohort 2021***

| | *CCI covariates included in model* | *No CCI covariates included in model* |
|---|---|---|
| Intellectual disability | 0.22 (0.21 - 0.23) | 0.23 (0.22 - 0.25) |
| Multiple sclerosis | 0.96 (0.95 - 0.98) | 0.98 (0.96 - 1.00) |
| Neuromuscular disorders | 0.36 (0.34 - 0.38) | 0.46 (0.44 - 0.49) |
| Other neurodegenerative disorders | 1.16 (1.12 - 1.19) | 1.22 (1.18 - 1.26) |
| Parkinson's disease | 1.52 (1.50 - 1.53) | 1.53 (1.51 - 1.54) |
| Personality disorders | 0.09 (0.08 - 0.10) | 0.09 (0.08 - 0.10) |
| Polyneuropathy | 0.32 (0.31 - 0.33) | 0.55 (0.54 - 0.56) |
| Schizophrenia spectrum disorders | 0.88 (0.87 - 0.89) | 0.89 (0.88 - 0.89) |
| Sleep disorders | 0.11 (0.11 - 0.12) | 0.18 (0.18 - 0.19) |
| Stress related disorders | 0.10 (0.09 - 0.10) | 0.11 (0.10 - 0.11) |
| Stroke | 0.61 (0.60 - 0.61) | 0.74 (0.74 - 0.75) |
| Traumatic brain injury | 0.13 (0.12 - 0.13) | 0.14 (0.13 - 0.14) |
| Male | -0.05 (-0.06 - -0.05) | -0.04 (-0.05 - -0.04) |

### Cost regression

Sensitivity analysis 1 / Prevalent cohort 2015 / No CCI variables in model

| Brain disorder | Estimate (95% CI) |
|---|---|
| Dementia | 1.16 (1.16 - 1.17) |
| Schizophrenia spectrum disorders | 0.78 (0.77 - 0.79) |
| Parkinson’s disease | 0.78 (0.76 - 0.79) |
| Multiple sclerosis | 0.77 (0.75 - 0.79) |
| Other neurodegenerative disorders | 0.74 (0.70 - 0.77) |
| Cerebral palsy | 0.52 (0.50 - 0.54) |
| Neuromuscular disorders | 0.46 (0.44 - 0.48) |
| Stroke | 0.45 (0.45 - 0.46) |
| Epilepsy | 0.43 (0.42 - 0.44) |
| Bipolar disorder | 0.42 (0.41 - 0.44) |
| Polyneuropathy | 0.42 (0.41 - 0.43) |
| Eating disorders | 0.41 (0.40 - 0.43) |
| Alcohol abuse | 0.39 (0.39 - 0.40) |
| Brain tumors | 0.37 (0.36 - 0.38) |
| Depression | 0.27 (0.27 - 0.28) |
| Drug abuse | 0.25 (0.24 - 0.26) |
| Intellectual disability | 0.24 (0.22 - 0.25) |
| Infections of the CNS | 0.19 (0.18 - 0.20) |
| Anxiety disorders | 0.16 (0.16 - 0.17) |
| Sleep disorders | 0.14 (0.13 - 0.15) |
| Personality disorders | 0.12 (0.11 - 0.13) |
| Traumatic brain injury | 0.08 (0.08 - 0.09) |
| Stress related disorders | 0.08 (0.07 - 0.08) |
| Headache | 0.07 (0.07 - 0.08) |
| Developmental and behavioural disorders | 0.05 (0.04 - 0.05) |

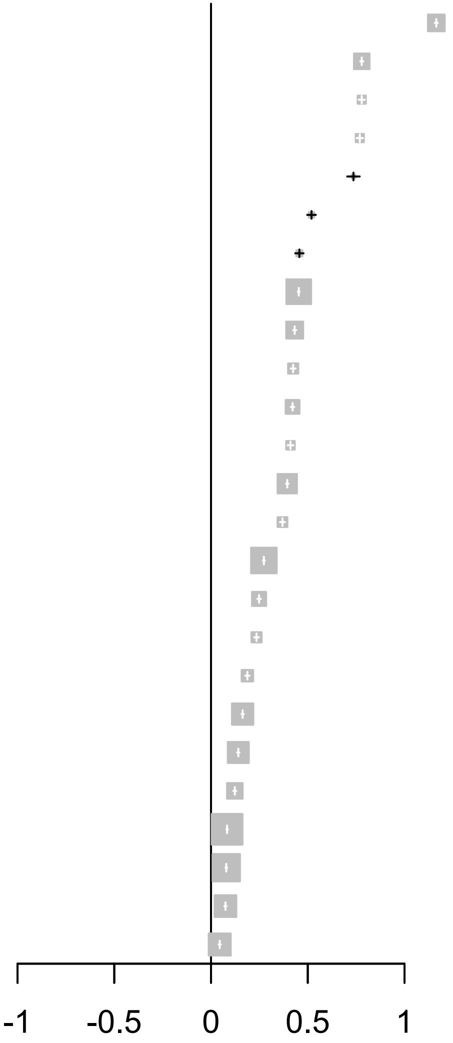

### Cost regression

#### Sensitivity analysis 1 / Prevalent cohort 2015 / CCI variables included in model

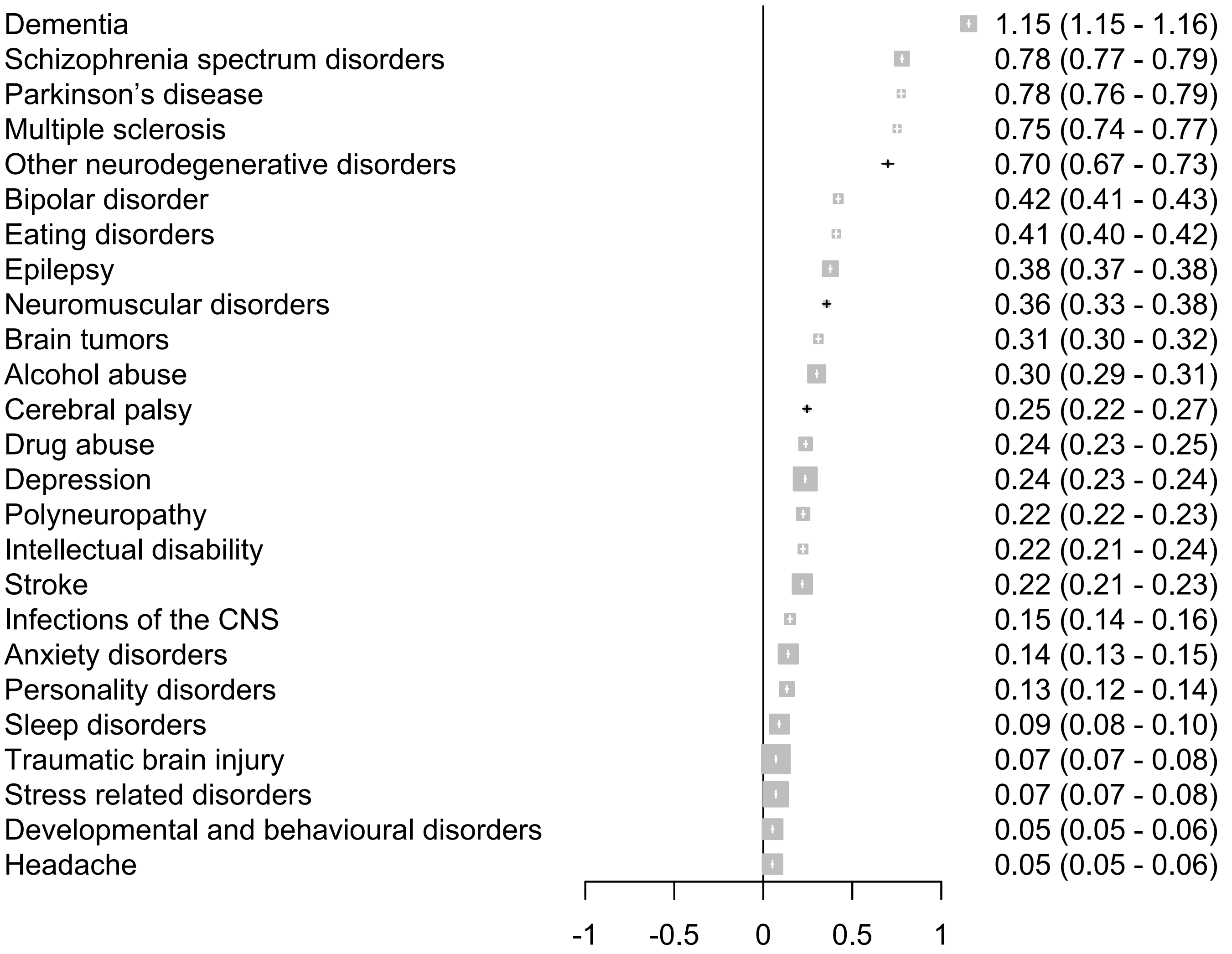

Cost regression

Sensitivity analysis 1 / Prevalent cohort 2016 / No CCI variables in model

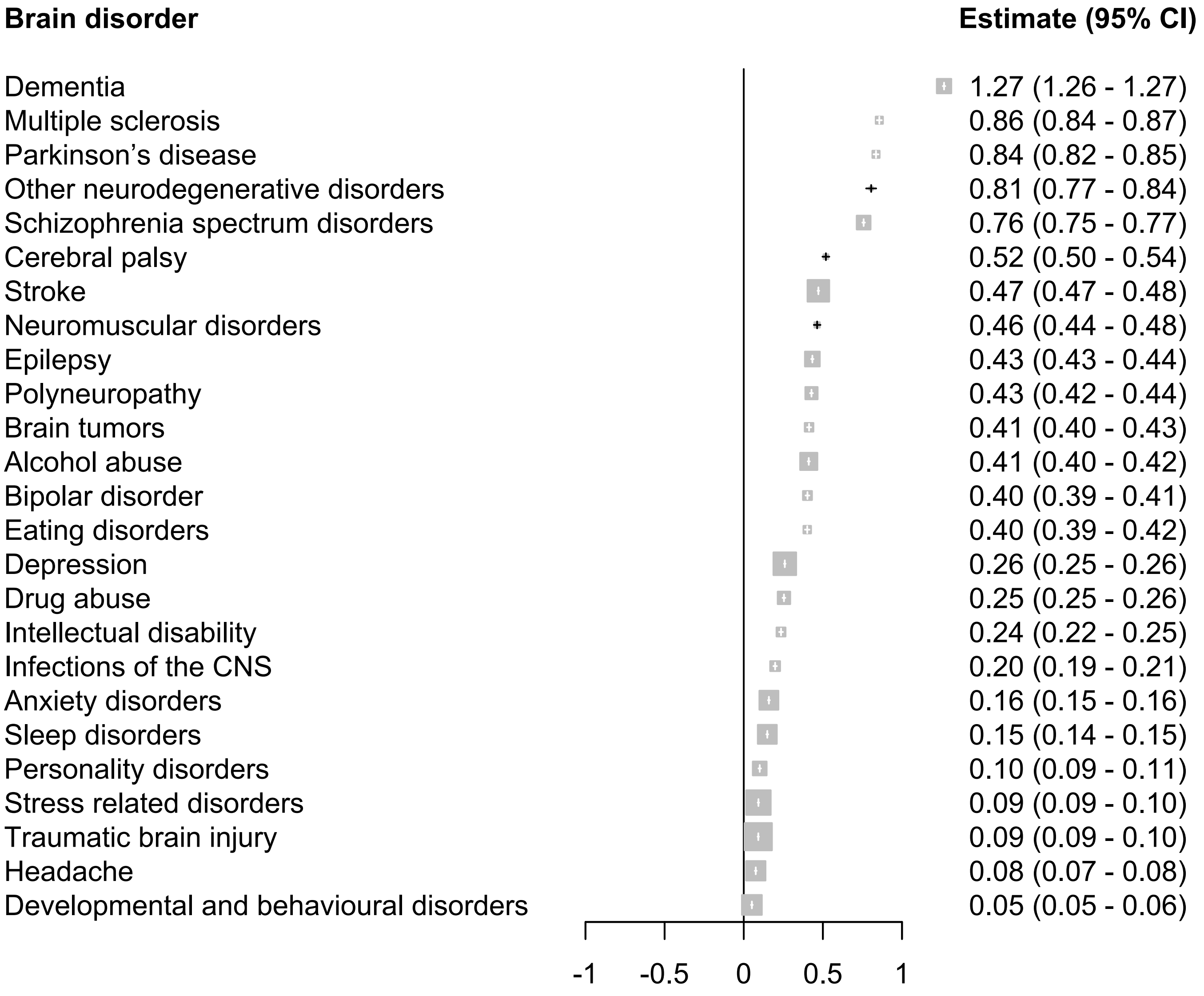

#### Cost regression

Sensitivity analysis 1 / Prevalent cohort 2016 / CCI variables included in model

| Brain disorder | Estimate (95% CI) |
|---|---|
| Dementia | 1.25 (1.25 - 1.26) |
| Multiple sclerosis | 0.84 (0.82 - 0.85) |
| Parkinson’s disease | 0.83 (0.82 - 0.85) |
| Other neurodegenerative disorders | 0.77 (0.74 - 0.80) |
| Schizophrenia spectrum disorders | 0.76 (0.75 - 0.77) |
| Eating disorders | 0.40 (0.39 - 0.41) |
| Bipolar disorder | 0.40 (0.39 - 0.41) |
| Epilepsy | 0.38 (0.37 - 0.39) |
| Neuromuscular disorders | 0.36 (0.34 - 0.38) |
| Brain tumors | 0.35 (0.34 - 0.36) |
| Alcohol abuse | 0.31 (0.31 - 0.32) |
| Cerebral palsy | 0.26 (0.24 - 0.28) |
| Stroke | 0.25 (0.24 - 0.26) |
| Drug abuse | 0.24 (0.24 - 0.25) |
| Polyneuropathy | 0.23 (0.22 - 0.24) |
| Depression | 0.22 (0.22 - 0.23) |
| Intellectual disability | 0.22 (0.21 - 0.24) |
| Infections of the CNS | 0.16 (0.15 - 0.17) |
| Anxiety disorders | 0.14 (0.13 - 0.14) |
| Personality disorders | 0.11 (0.10 - 0.12) |
| Sleep disorders | 0.09 (0.09 - 0.10) |
| Stress related disorders | 0.08 (0.08 - 0.09) |
| Traumatic brain injury | 0.08 (0.08 - 0.08) |
| Developmental and behavioural disorders | 0.06 (0.05 - 0.06) |
| Headache | 0.05 (0.05 - 0.06) |

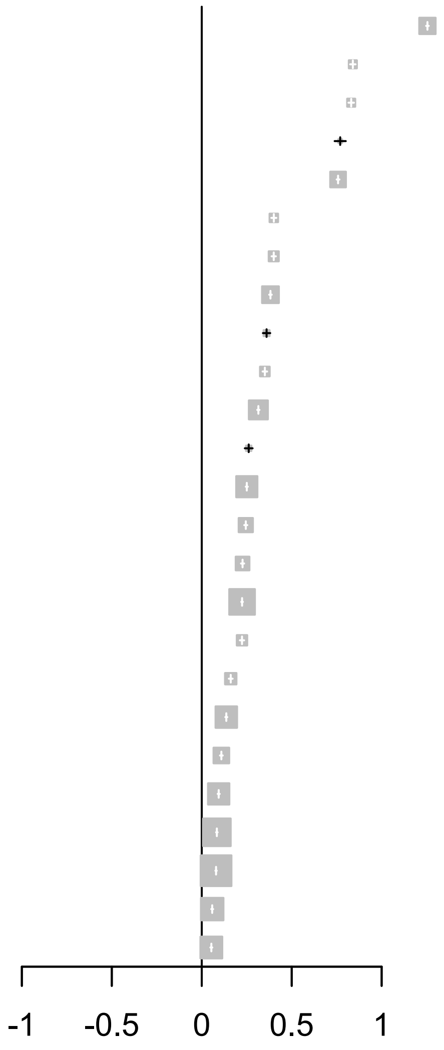

### Cost regression

Sensitivity analysis 1 / Prevalent cohort 2017 / No CCI variables in model

| Brain disorder | Estimate (95% CI) |
|---|---|
| Dementia | 1.38 (1.37 - 1.38) |
| Multiple sclerosis | 0.98 (0.96 - 0.99) |
| Parkinson’s disease | 0.95 (0.94 - 0.97) |
| Other neurodegenerative disorders | 0.86 (0.83 - 0.89) |
| Schizophrenia spectrum disorders | 0.79 (0.78 - 0.80) |
| Stroke | 0.53 (0.52 - 0.53) |
| Cerebral palsy | 0.51 (0.49 - 0.53) |
| Polyneuropathy | 0.48 (0.48 - 0.49) |
| Neuromuscular disorders | 0.46 (0.43 - 0.48) |
| Brain tumors | 0.45 (0.44 - 0.46) |
| Epilepsy | 0.43 (0.43 - 0.44) |
| Alcohol abuse | 0.42 (0.42 - 0.43) |
| Eating disorders | 0.39 (0.38 - 0.41) |
| Bipolar disorder | 0.36 (0.34 - 0.37) |
| Drug abuse | 0.26 (0.25 - 0.27) |
| Depression | 0.26 (0.25 - 0.26) |
| Intellectual disability | 0.23 (0.22 - 0.24) |
| Infections of the CNS | 0.20 (0.19 - 0.21) |
| Sleep disorders | 0.17 (0.17 - 0.18) |
| Anxiety disorders | 0.16 (0.15 - 0.17) |
| Personality disorders | 0.10 (0.10 - 0.11) |
| Traumatic brain injury | 0.10 (0.10 - 0.10) |
| Stress related disorders | 0.10 (0.09 - 0.10) |
| Headache | 0.08 (0.07 - 0.08) |
| Developmental and behavioural disorders | 0.06 (0.05 - 0.07) |

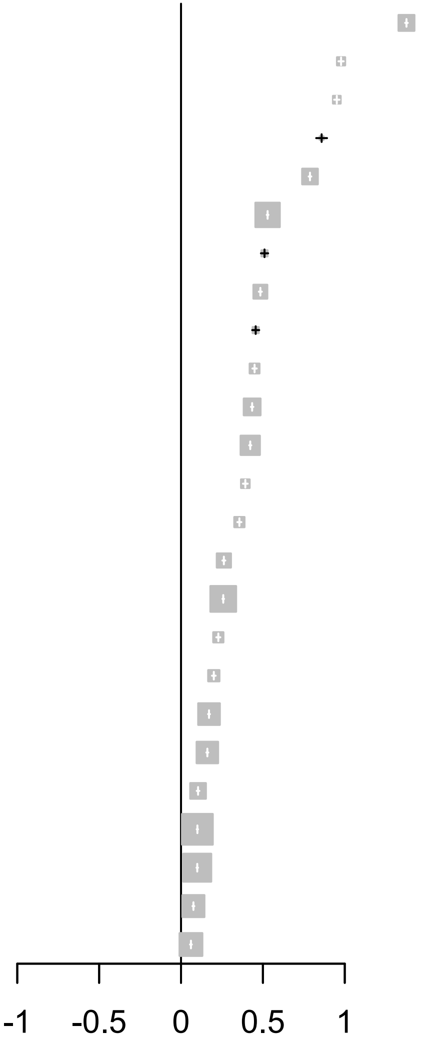

### Cost regression

Sensitivity analysis 1 / Prevalent cohort 2017 / CCI variables included in model

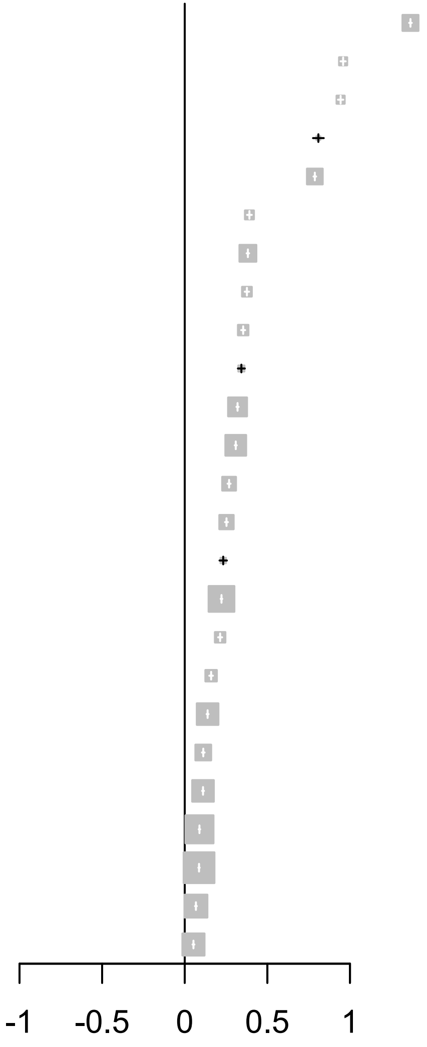


| **Brain disorder** | **Estimate (95% CI)** |
|---|---|
| Dementia | 1.37 (1.36 - 1.37) |
| Multiple sclerosis | 0.96 (0.94 - 0.97) |
| Parkinson’s disease | 0.94 (0.93 - 0.96) |
| Other neurodegenerative disorders | 0.81 (0.78 - 0.84) |
| Schizophrenia spectrum disorders | 0.79 (0.78 - 0.80) |
| Eating disorders | 0.39 (0.38 - 0.41) |
| Epilepsy | 0.38 (0.37 - 0.39) |
| Brain tumors | 0.38 (0.36 - 0.39) |
| Bipolar disorder | 0.35 (0.34 - 0.37) |
| Neuromuscular disorders | 0.34 (0.32 - 0.36) |
| Alcohol abuse | 0.32 (0.31 - 0.33) |
| Stroke | 0.31 (0.30 - 0.31) |
| Polyneuropathy | 0.27 (0.26 - 0.28) |
| Drug abuse | 0.25 (0.24 - 0.26) |
| Cerebral palsy | 0.23 (0.21 - 0.26) |
| Depression | 0.22 (0.22 - 0.23) |
| Intellectual disability | 0.21 (0.20 - 0.23) |
| Infections of the CNS | 0.16 (0.15 - 0.17) |
| Anxiety disorders | 0.14 (0.13 - 0.14) |
| Personality disorders | 0.11 (0.10 - 0.12) |
| Sleep disorders | 0.11 (0.10 - 0.12) |
| Stress related disorders | 0.09 (0.08 - 0.09) |
| Traumatic brain injury | 0.09 (0.08 - 0.09) |
| Developmental and behavioural disorders | 0.07 (0.06 - 0.07) |
| Headache | 0.05 (0.05 - 0.06) |

### Cost regression

#### Sensitivity analysis 1 / Prevalent cohort 2018 / No CCI variables in model

| **Brain disorder** | **Estimate (95% CI)** |
|---|---|
| Dementia | 1.53 (1.52 - 1.54) |
| Parkinson’s disease | 1.01 (0.99 - 1.02) |
| Other neurodegenerative disorders | 0.89 (0.86 - 0.92) |
| Schizophrenia spectrum disorders | 0.69 (0.69 - 0.70) |
| Multiple sclerosis | 0.53 (0.52 - 0.55) |
| Stroke | 0.53 (0.52 - 0.53) |
| Alcohol abuse | 0.44 (0.44 - 0.45) |
| Brain tumors | 0.39 (0.37 - 0.40) |
| Cerebral palsy | 0.38 (0.36 - 0.40) |
| Epilepsy | 0.38 (0.37 - 0.38) |
| Polyneuropathy | 0.37 (0.36 - 0.38) |
| Neuromuscular disorders | 0.36 (0.34 - 0.38) |
| Bipolar disorder | 0.33 (0.32 - 0.34) |
| Eating disorders | 0.32 (0.31 - 0.33) |
| Depression | 0.23 (0.22 - 0.23) |
| Drug abuse | 0.22 (0.21 - 0.23) |
| Infections of the CNS | 0.18 (0.17 - 0.19) |
| Intellectual disability | 0.18 (0.17 - 0.19) |
| Sleep disorders | 0.14 (0.13 - 0.14) |
| Anxiety disorders | 0.12 (0.11 - 0.12) |
| Traumatic brain injury | 0.10 (0.09 - 0.10) |
| Stress related disorders | 0.09 (0.09 - 0.10) |
| Personality disorders | 0.08 (0.07 - 0.09) |
| Developmental and behavioural disorders | 0.06 (0.05 - 0.06) |
| Headache | 0.05 (0.04 - 0.05) |

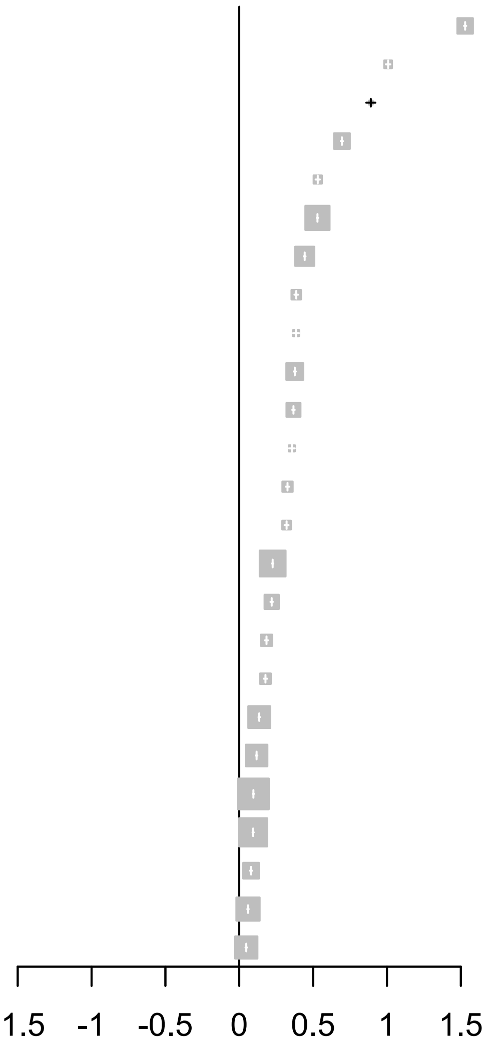

#### Cost regression

Sensitivity analysis 1 / Prevalent cohort 2018 / CCI variables included in model

| Brain disorder | Estimate (95% CI) |
|---|---|
| Dementia | 1.52 (1.51 - 1.52) |
| Parkinson’s disease | 1.00 (0.99 - 1.02) |
| Other neurodegenerative disorders | 0.84 (0.81 - 0.87) |
| Schizophrenia spectrum disorders | 0.69 (0.69 - 0.70) |
| Multiple sclerosis | 0.51 (0.50 - 0.53) |
| Alcohol abuse | 0.35 (0.35 - 0.36) |
| Brain tumors | 0.34 (0.33 - 0.35) |
| Epilepsy | 0.33 (0.33 - 0.34) |
| Stroke | 0.33 (0.33 - 0.34) |
| Bipolar disorder | 0.33 (0.31 - 0.34) |
| Eating disorders | 0.32 (0.31 - 0.33) |
| Neuromuscular disorders | 0.28 (0.26 - 0.30) |
| Drug abuse | 0.21 (0.21 - 0.22) |
| Polyneuropathy | 0.20 (0.19 - 0.21) |
| Depression | 0.20 (0.20 - 0.20) |
| Intellectual disability | 0.17 (0.15 - 0.18) |
| Infections of the CNS | 0.16 (0.15 - 0.17) |
| Cerebral palsy | 0.12 (0.10 - 0.14) |
| Anxiety disorders | 0.10 (0.10 - 0.11) |
| Stress related disorders | 0.09 (0.08 - 0.09) |
| Sleep disorders | 0.08 (0.08 - 0.09) |
| Personality disorders | 0.08 (0.08 - 0.09) |
| Traumatic brain injury | 0.08 (0.08 - 0.09) |
| Developmental and behavioural disorders | 0.07 (0.06 - 0.07) |
| Headache | 0.03 (0.02 - 0.04) |

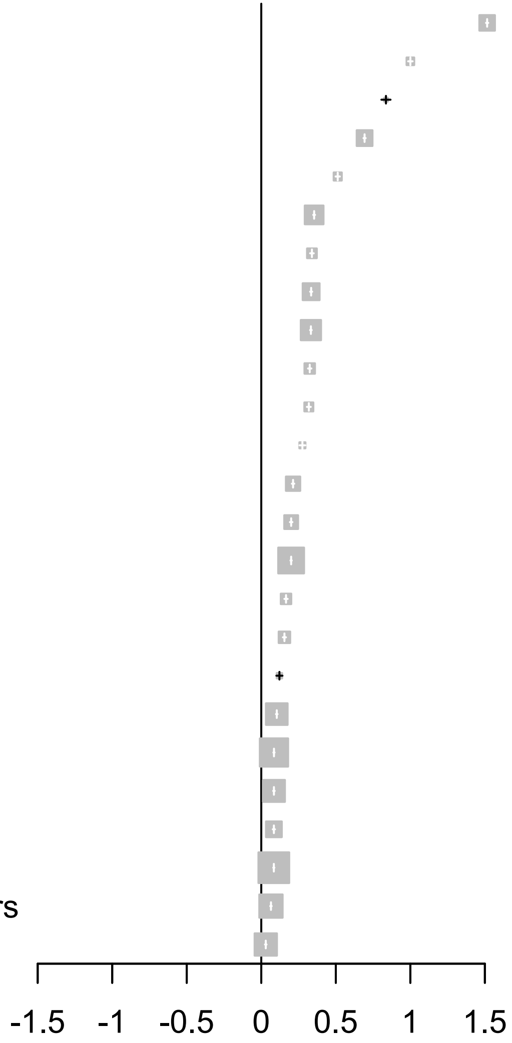

### Cost regression

Sensitivity analysis 1 / Prevalent cohort 2019 / No CCI variables in model

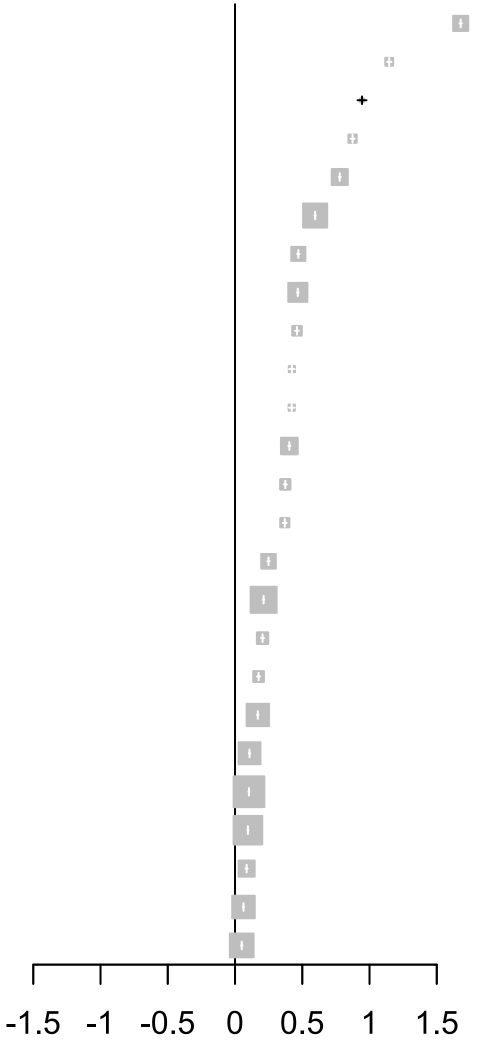


| Brain disorder | Estimate (95% CI) |
|---|---|
| Dementia | 1.68 (1.67 - 1.69) |
| Parkinson’s disease | 1.15 (1.13 - 1.16) |
| Other neurodegenerative disorders | 0.94 (0.91 - 0.98) |
| Multiple sclerosis | 0.87 (0.86 - 0.89) |
| Schizophrenia spectrum disorders | 0.78 (0.77 - 0.79) |
| Stroke | 0.60 (0.59 - 0.60) |
| Polyneuropathy | 0.47 (0.46 - 0.48) |
| Alcohol abuse | 0.47 (0.46 - 0.47) |
| Brain tumors | 0.46 (0.45 - 0.47) |
| Cerebral palsy | 0.42 (0.40 - 0.44) |
| Neuromuscular disorders | 0.42 (0.40 - 0.44) |
| Epilepsy | 0.40 (0.40 - 0.41) |
| Bipolar disorder | 0.37 (0.36 - 0.39) |
| Eating disorders | 0.37 (0.36 - 0.38) |
| Drug abuse | 0.25 (0.24 - 0.26) |
| Depression | 0.21 (0.21 - 0.22) |
| Infections of the CNS | 0.20 (0.19 - 0.22) |
| Intellectual disability | 0.18 (0.16 - 0.19) |
| Sleep disorders | 0.17 (0.16 - 0.18) |
| Anxiety disorders | 0.11 (0.10 - 0.11) |
| Traumatic brain injury | 0.10 (0.10 - 0.11) |
| Stress related disorders | 0.10 (0.09 - 0.10) |
| Personality disorders | 0.09 (0.08 - 0.09) |
| Headache | 0.06 (0.06 - 0.07) |
| Developmental and behavioural disorders | 0.05 (0.04 - 0.05) |

### Cost regression

Sensitivity analysis 1 / Prevalent cohort 2019 / CCI variables included in model

| **Brain disorder** | **Estimate (95% CI)** |
|---|---|
| Dementia | 1.67 (1.66 - 1.68) |
| Parkinson’s disease | 1.14 (1.12 - 1.15) |
| Other neurodegenerative disorders | 0.89 (0.86 - 0.92) |
| Multiple sclerosis | 0.86 (0.84 - 0.87) |
| Schizophrenia spectrum disorders | 0.78 (0.77 - 0.78) |
| Stroke | 0.41 (0.41 - 0.42) |
| Brain tumors | 0.39 (0.38 - 0.40) |
| Alcohol abuse | 0.37 (0.36 - 0.38) |
| Bipolar disorder | 0.37 (0.36 - 0.38) |
| Eating disorders | 0.37 (0.35 - 0.38) |
| Epilepsy | 0.36 (0.35 - 0.37) |
| Neuromuscular disorders | 0.32 (0.30 - 0.34) |
| Polyneuropathy | 0.26 (0.25 - 0.27) |
| Drug abuse | 0.24 (0.23 - 0.25) |
| Depression | 0.18 (0.18 - 0.19) |
| Infections of the CNS | 0.17 (0.16 - 0.18) |
| Intellectual disability | 0.16 (0.15 - 0.17) |
| Cerebral palsy | 0.16 (0.14 - 0.18) |
| Sleep disorders | 0.11 (0.10 - 0.11) |
| Anxiety disorders | 0.09 (0.09 - 0.10) |
| Traumatic brain injury | 0.09 (0.09 - 0.10) |
| Personality disorders | 0.09 (0.08 - 0.10) |
| Stress related disorders | 0.09 (0.08 - 0.09) |
| Developmental and behavioural disorders | 0.06 (0.05 - 0.06) |
| Headache | 0.04 (0.04 - 0.05) |

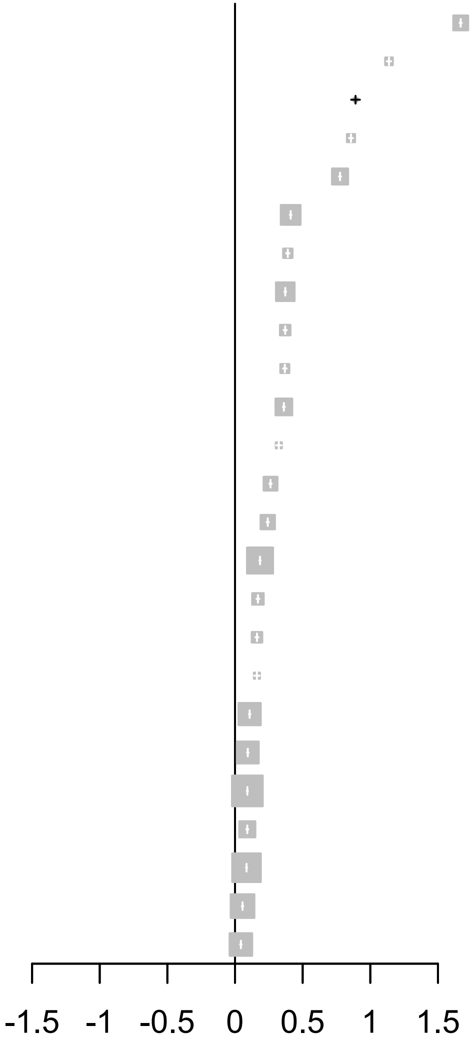

#### Cost regression

Sensitivity analysis 1 / Prevalent cohort 2020 / No CCI variables in model

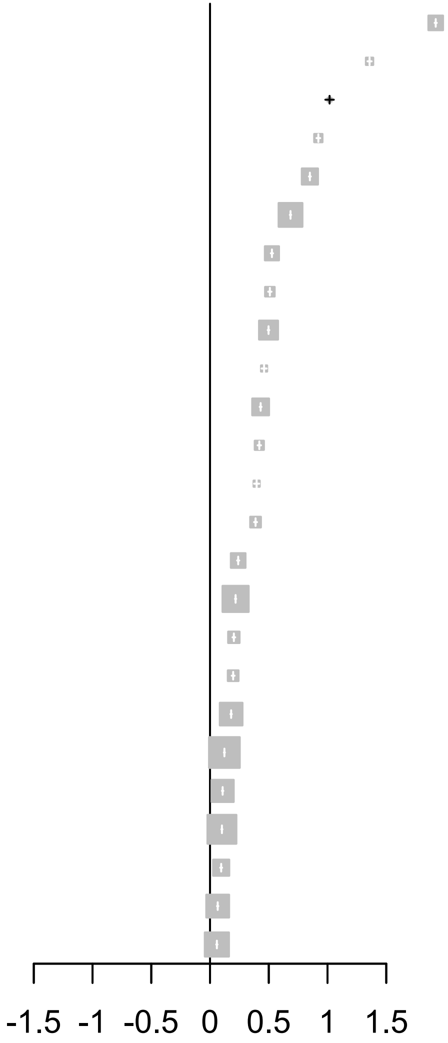


| **Brain disorder** | **Estimate (95% CI)** |
|---|---|
| Dementia | 1.92 (1.91 - 1.93) |
| Parkinson’s disease | 1.36 (1.34 - 1.38) |
| Other neurodegenerative disorders | 1.02 (0.98 - 1.05) |
| Multiple sclerosis | 0.92 (0.91 - 0.94) |
| Schizophrenia spectrum disorders | 0.85 (0.84 - 0.86) |
| Stroke | 0.69 (0.68 - 0.69) |
| Polyneuropathy | 0.53 (0.52 - 0.54) |
| Brain tumors | 0.51 (0.50 - 0.52) |
| Alcohol abuse | 0.50 (0.49 - 0.50) |
| Neuromuscular disorders | 0.46 (0.44 - 0.48) |
| Epilepsy | 0.43 (0.42 - 0.44) |
| Eating disorders | 0.42 (0.40 - 0.43) |
| Cerebral palsy | 0.40 (0.37 - 0.42) |
| Bipolar disorder | 0.39 (0.38 - 0.40) |
| Drug abuse | 0.24 (0.23 - 0.25) |
| Depression | 0.22 (0.21 - 0.22) |
| Infections of the CNS | 0.20 (0.19 - 0.21) |
| Intellectual disability | 0.20 (0.18 - 0.21) |
| Sleep disorders | 0.18 (0.17 - 0.19) |
| Traumatic brain injury | 0.12 (0.12 - 0.13) |
| Anxiety disorders | 0.11 (0.10 - 0.11) |
| Stress related disorders | 0.10 (0.10 - 0.11) |
| Personality disorders | 0.09 (0.09 - 0.10) |
| Headache | 0.07 (0.06 - 0.07) |
| Developmental and behavioural disorders | 0.06 (0.05 - 0.06) |

### Cost regression

Sensitivity analysis 1 / Prevalent cohort 2020 / CCI variables included in model

| **Brain disorder** | **Estimate (95% CI)** |
| --- | --- |
| Dementia | 1.92 (1.91 - 1.92) |
| Parkinson’s disease | 1.35 (1.33 - 1.37) |
| Other neurodegenerative disorders | 0.96 (0.92 - 0.99) |
| Multiple sclerosis | 0.90 (0.89 - 0.92) |
| Schizophrenia spectrum disorders | 0.85 (0.84 - 0.86) |
| Stroke | 0.52 (0.51 - 0.52) |
| Brain tumors | 0.43 (0.42 - 0.44) |
| Eating disorders | 0.42 (0.40 - 0.43) |
| Alcohol abuse | 0.40 (0.39 - 0.40) |
| Epilepsy | 0.39 (0.38 - 0.40) |
| Bipolar disorder | 0.38 (0.37 - 0.40) |
| Neuromuscular disorders | 0.36 (0.34 - 0.38) |
| Polyneuropathy | 0.30 (0.29 - 0.31) |
| Drug abuse | 0.23 (0.22 - 0.24) |
| Depression | 0.19 (0.19 - 0.20) |
| Intellectual disability | 0.18 (0.17 - 0.20) |
| Infections of the CNS | 0.17 (0.15 - 0.18) |
| Cerebral palsy | 0.14 (0.12 - 0.16) |
| Sleep disorders | 0.11 (0.11 - 0.12) |
| Traumatic brain injury | 0.11 (0.11 - 0.11) |
| Personality disorders | 0.10 (0.09 - 0.11) |
| Anxiety disorders | 0.10 (0.09 - 0.10) |
| Stress related disorders | 0.09 (0.09 - 0.09) |
| Developmental and behavioural disorders | 0.06 (0.06 - 0.07) |
| Headache | 0.05 (0.04 - 0.05) |

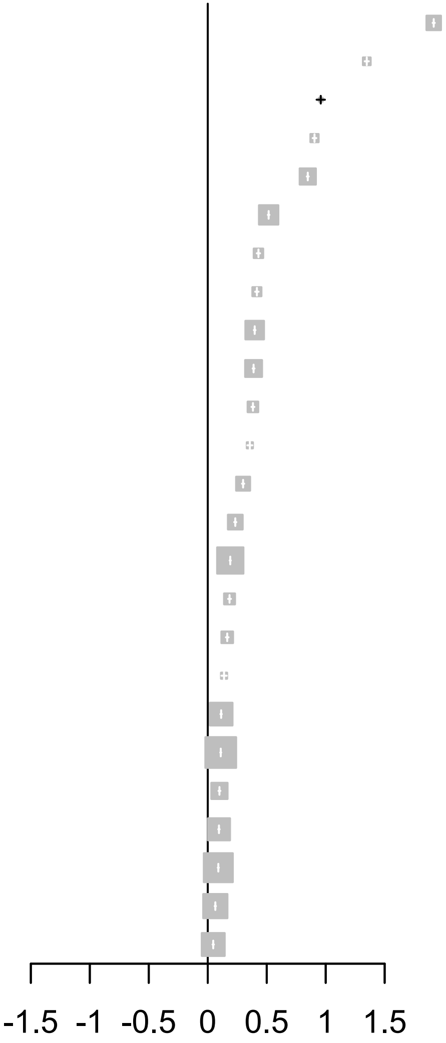

### Cost regression

Sensitivity analysis 1 / Prevalent cohort 2021 / No CCI variables in model

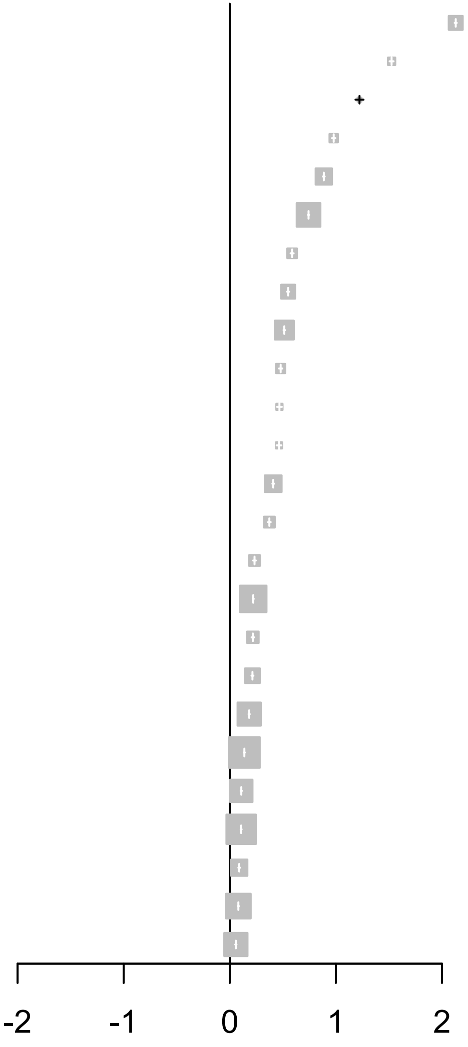


| **Brain disorder** | **Estimate (95% CI)** |
|---|---|
| Dementia | 2.13 (2.12 - 2.14) |
| Parkinson’s disease | 1.53 (1.51 - 1.54) |
| Other neurodegenerative disorders | 1.22 (1.18 - 1.26) |
| Multiple sclerosis | 0.98 (0.96 - 1.00) |
| Schizophrenia spectrum disorders | 0.89 (0.88 - 0.89) |
| Stroke | 0.74 (0.74 - 0.75) |
| Brain tumors | 0.59 (0.57 - 0.60) |
| Polyneuropathy | 0.55 (0.54 - 0.56) |
| Alcohol abuse | 0.51 (0.51 - 0.52) |
| Eating disorders | 0.48 (0.46 - 0.49) |
| Cerebral palsy | 0.47 (0.45 - 0.49) |
| Neuromuscular disorders | 0.46 (0.44 - 0.49) |
| Epilepsy | 0.41 (0.40 - 0.42) |
| Bipolar disorder | 0.37 (0.36 - 0.39) |
| Intellectual disability | 0.23 (0.22 - 0.25) |
| Depression | 0.22 (0.22 - 0.23) |
| Infections of the CNS | 0.22 (0.21 - 0.23) |
| Drug abuse | 0.21 (0.20 - 0.22) |
| Sleep disorders | 0.18 (0.18 - 0.19) |
| Traumatic brain injury | 0.14 (0.13 - 0.14) |
| Anxiety disorders | 0.11 (0.10 - 0.11) |
| Stress related disorders | 0.11 (0.10 - 0.11) |
| Personality disorders | 0.09 (0.08 - 0.10) |
| Developmental and behavioural disorders | 0.08 (0.08 - 0.09) |
| Headache | 0.06 (0.05 - 0.06) |

### Cost regression

Sensitivity analysis 1 / Prevalent cohort 2021 / CCI variables included in model

| **Brain disorder** | **Estimate (95% CI)** |
| --- | --- |
| Dementia | 2.13 (2.12 - 2.14) |
| Parkinson’s disease | 1.52 (1.50 - 1.53) |
| Other neurodegenerative disorders | 1.16 (1.12 - 1.19) |
| Multiple sclerosis | 0.96 (0.95 - 0.98) |
| Schizophrenia spectrum disorders | 0.88 (0.87 - 0.89) |
| Stroke | 0.61 (0.60 - 0.61) |
| Brain tumors | 0.51 (0.49 - 0.52) |
| Eating disorders | 0.48 (0.46 - 0.49) |
| Alcohol abuse | 0.41 (0.41 - 0.42) |
| Epilepsy | 0.37 (0.36 - 0.38) |
| Bipolar disorder | 0.37 (0.35 - 0.38) |
| Neuromuscular disorders | 0.36 (0.34 - 0.38) |
| Polyneuropathy | 0.32 (0.31 - 0.33) |
| Intellectual disability | 0.22 (0.21 - 0.23) |
| Drug abuse | 0.21 (0.20 - 0.22) |
| Cerebral palsy | 0.20 (0.18 - 0.23) |
| Depression | 0.20 (0.19 - 0.20) |
| Infections of the CNS | 0.18 (0.17 - 0.19) |
| Traumatic brain injury | 0.13 (0.12 - 0.13) |
| Sleep disorders | 0.11 (0.11 - 0.12) |
| Anxiety disorders | 0.10 (0.09 - 0.10) |
| Stress related disorders | 0.10 (0.09 - 0.10) |
| Personality disorders | 0.09 (0.08 - 0.10) |
| Developmental and behavioural disorders | 0.09 (0.08 - 0.09) |
| Headache | 0.04 (0.03 - 0.05) |

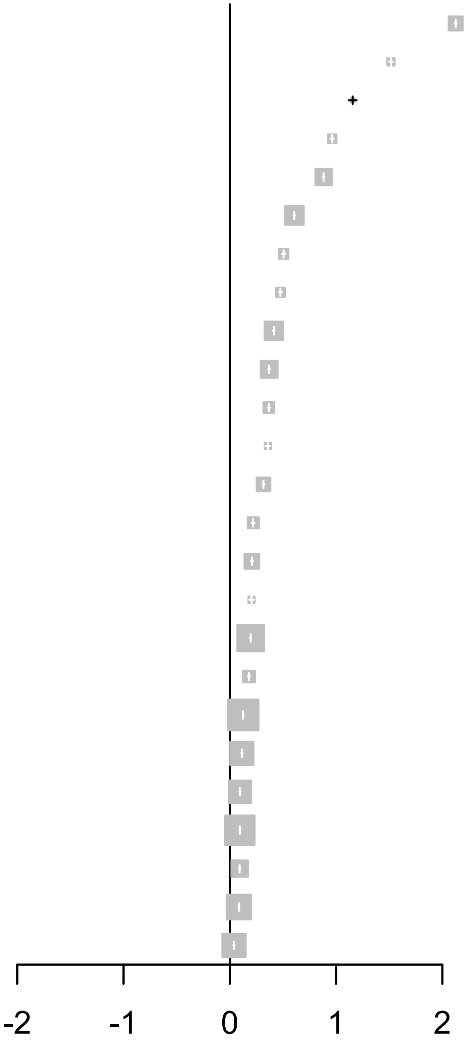
