## Appendix 11 for "Temporal trends in the occurrence, mortality, and cost of brain disorders in Denmark"

***Sensitivity analysis 3 / Incident cohort 2011-2015***

| *Disease group* | *Persons, N* | *Men, N (%)%* | *Age, median (Q1-Q3)* | *CCI: 0, N (%)* | *CCI: 1-2, N (%)* | *CCI: +3, N (%)* |
|---|---|---|---|---|---|---|
| **Any brain disorder** | 580,993 | 281,414 (48.4) | 40.7 (22.1;61.7) | 482,604 (83.1) | 76,216 (13.1) | 22,173 (3.8) |

***Sensitivity analysis 3 / Incident cohort 2016-2021***

| *Disease group* | *Persons, N* | *Men, N (%)%* | *Age, median (Q1-Q3)* | *CCI: 0, N (%)* | *CCI: 1-2, N (%)* | *CCI: +3, N (%)* |
|---|---|---|---|---|---|---|
| **Any brain disorder** | 636,299 | 313,181 (49.2) | 39.6 (21.4;63.3) | 527,781 (82.9) | 85,211 (13.4) | 23,307 (3.7) |

***Sensitivity analysis 3 / Prevalent cohort 2015***

| *Disease group* | *Persons, N* | *Men, N (%)%* | *Age, median (Q1-Q3)* | *CCI: 0, N (%)* | *CCI: 1-2, N (%)* | *CCI: +3, N (%)* |
|---|---|---|---|---|---|---|
| **Any brain disorder** | 2,361,015 | 1,038,065 (44.0) | 50.4 (34.8;65.5) | 1,759,716 (74.5) | 466,014 (19.7) | 135,285 (5.7) |

***Sensitivity analysis 3 / Prevalent cohort 2016***

| *Disease group* | *Persons, N* | *Men, N (%)%* | *Age, median (Q1-Q3)* | *CCI: 0, N (%)* | *CCI: 1-2, N (%)* | *CCI: +3, N (%)* |
|---|---|---|---|---|---|---|
| **Any brain disorder** | 2,402,356 | 1,057,715 (44.0) | 50.7 (34.7;65.7) | 1,782,902 (74.2) | 478,482 (19.9) | 140,972 (5.9) |

***Sensitivity analysis 3 / Prevalent cohort 2017***

| *Disease group* | *Persons, N* | *Men, N (%)%* | *Age, median (Q1-Q3)* | *CCI: 0, N (%)* | *CCI: 1-2, N (%)* | *CCI: +3, N (%)* |
|---|---|---|---|---|---|---|
| **Any brain disorder** | 2,440,500 | 1,076,519 (44.1) | 50.9 (34.6;65.9) | 1,802,986 (73.9) | 490,990 (20.1) | 146,524 (6.0) |

***Sensitivity analysis 3 / Prevalent cohort 2018***

| *Disease group* | *Persons, N* | *Men, N (%)%* | *Age, median (Q1-Q3)* | *CCI: 0, N (%)* | *CCI: 1-2, N (%)* | *CCI: +3, N (%)* |
|---|---|---|---|---|---|---|
| **Any brain disorder** | 2,476,561 | 1,094,261 (44.2) | 51.2 (34.5;66.1) | 1,822,472 (73.6) | 502,469 (20.3) | 151,620 (6.1) |

***Sensitivity analysis 3 / Prevalent cohort 2019***

| *Disease group* | *Persons, N* | *Men, N (%)%* | *Age, median (Q1-Q3)* | *CCI: 0, N (%)* | *CCI: 1-2, N (%)* | *CCI: +3, N (%)* |
|---|---|---|---|---|---|---|
| **Any brain disorder** | 2,507,423 | 1,109,739 (44.3) | 51.5 (34.5;66.3) | 1,844,853 (73.6) | 509,614 (20.3) | 152,956 (6.1) |

***Sensitivity analysis 3 / Prevalent cohort 2020***

| *Disease group* | *Persons, N* | *Men, N (%)%* | *Age, median (Q1-Q3)* | *CCI: 0, N (%)* | *CCI: 1-2, N (%)* | *CCI: +3, N (%)* |
|---|---|---|---|---|---|---|
| **Any brain disorder** | 2,533,726 | 1,122,539 (44.3) | 51.7 (34.4;66.5) | 1,864,425 (73.6) | 515,223 (20.3) | 154,078 (6.1) |

***Sensitivity analysis 3 / Prevalent cohort 2021***

| *Disease group* | *Persons, N* | *Men, N (%)%* | *Age, median (Q1-Q3)* | *CCI: 0, N (%)* | *CCI: 1-2, N (%)* | *CCI: +3, N (%)* |
|---|---|---|---|---|---|---|
| **Any brain disorder** | 2,557,051 | 1,132,842 (44.3) | 51.8 (34.4;66.7) | 1,882,135 (73.6) | 520,259 (20.3) | 154,657 (6.0) |

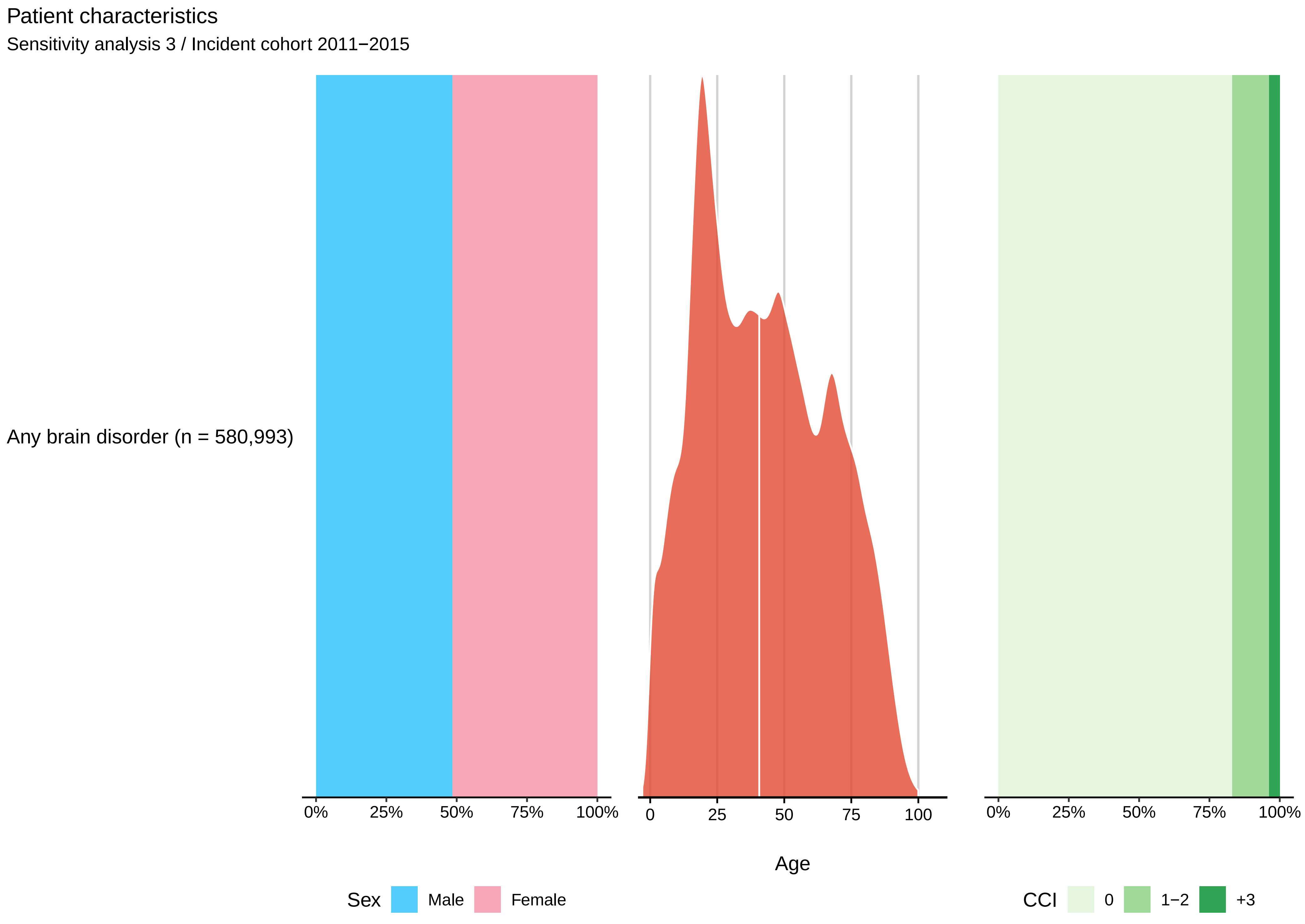
Patient characteristics
Sensitivity analysis 3 / Incident cohort 2011−2015
Any brain disorder (n = 580,993)
0%
25%
50%
75%
100%
0
25
50
75
100
Age
0%
25%
50%
75%
100%
Sex
Male
Female
CCI
0
1−2
+3

Patient characteristics

Sensitivity analysis 3 / Incident cohort 2016−2021

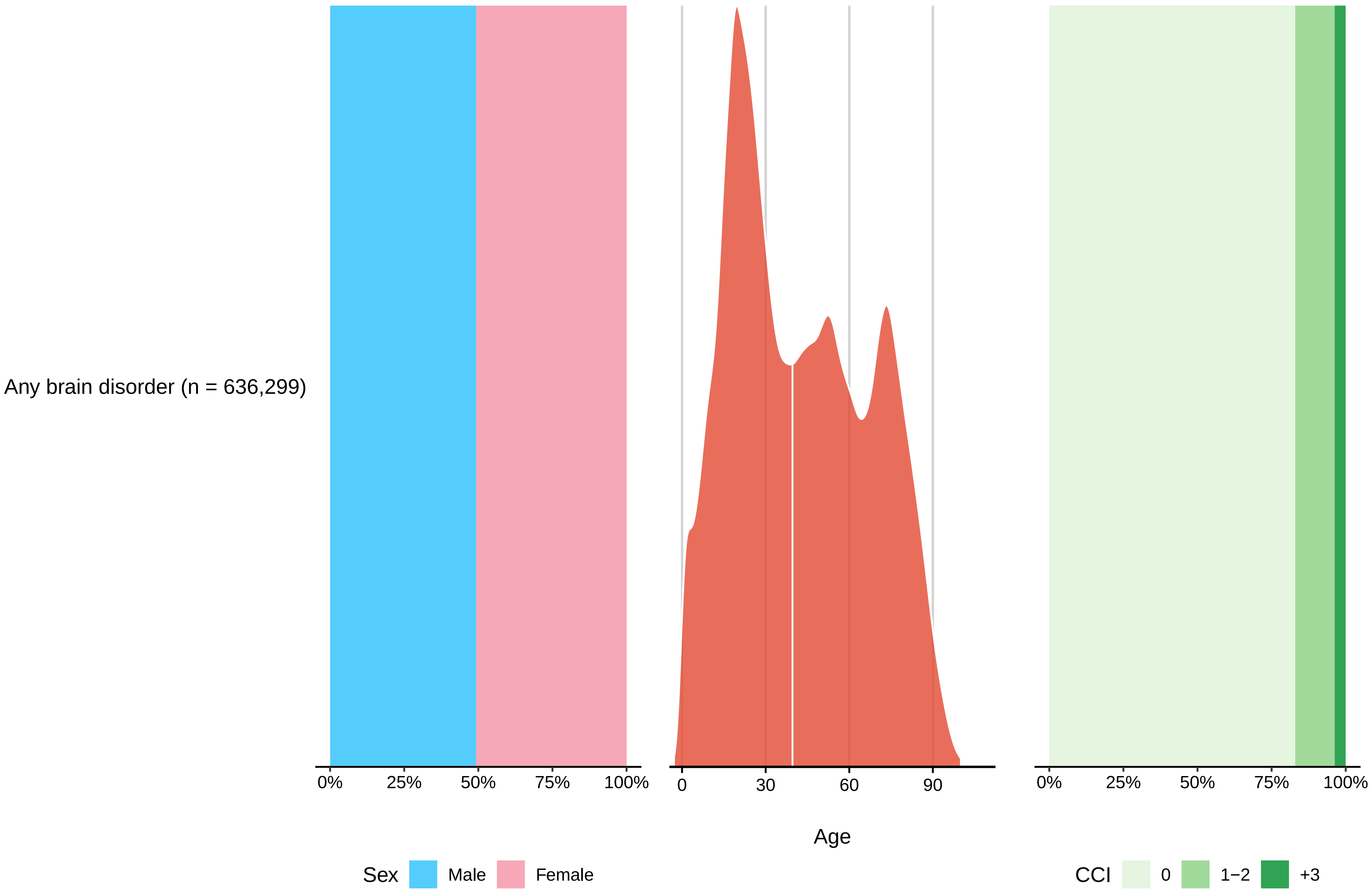

Patient characteristics

Sensitivity analysis 3 / Prevalent cohort 2015

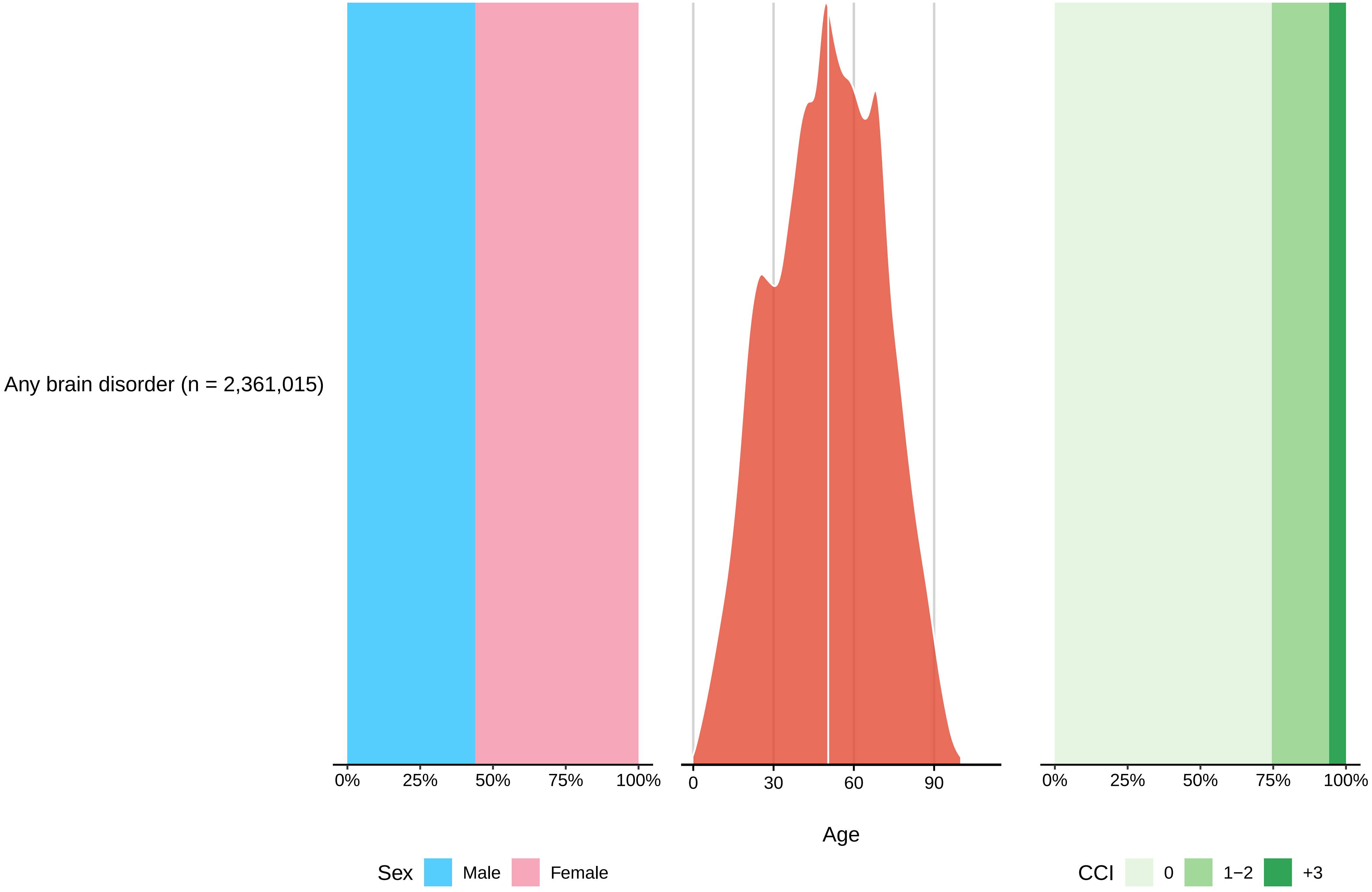

#### Patient characteristics

Sensitivity analysis 3 / Prevalent cohort 2016

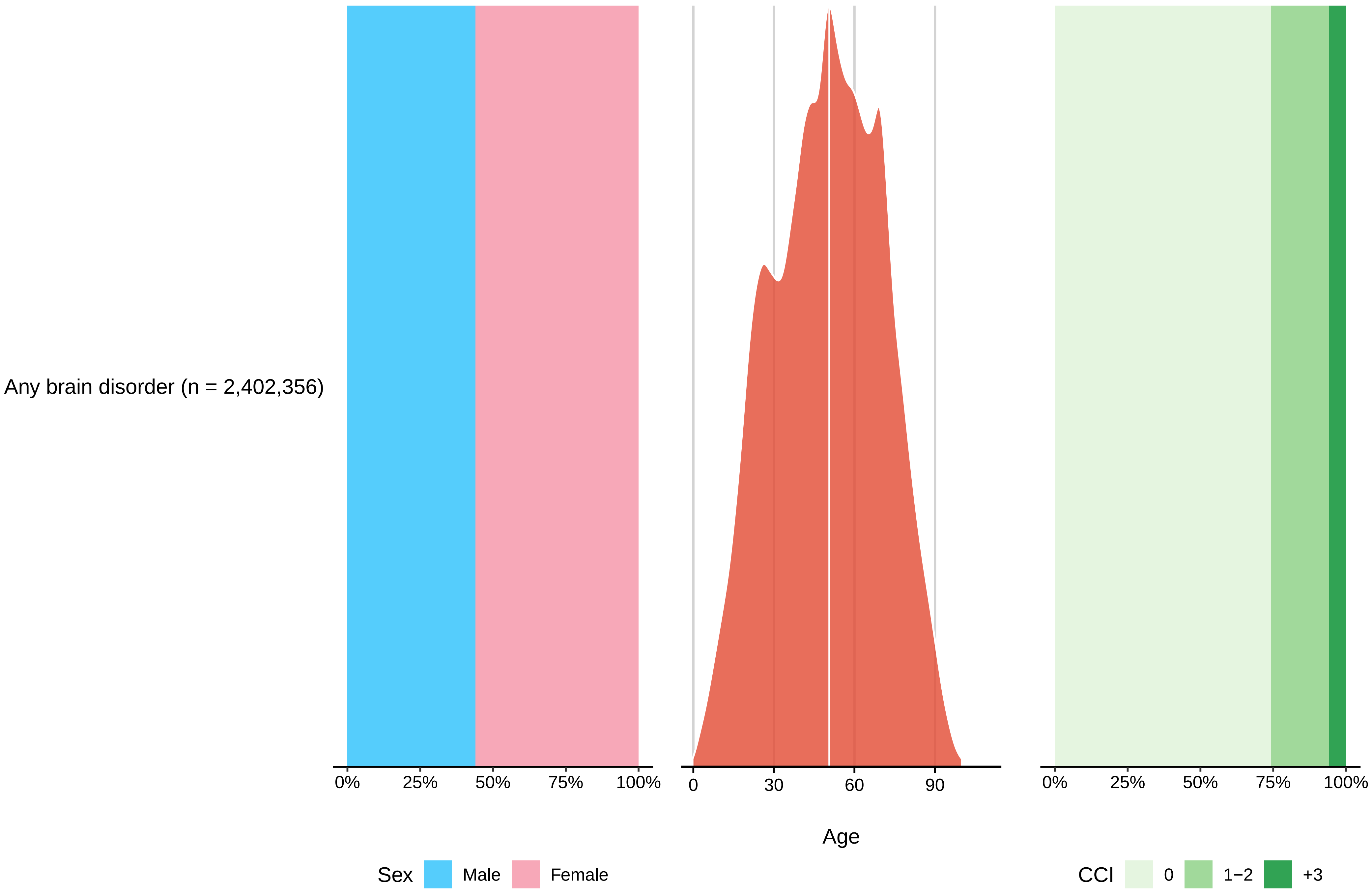

#### Patient characteristics

Sensitivity analysis 3 / Prevalent cohort 2017

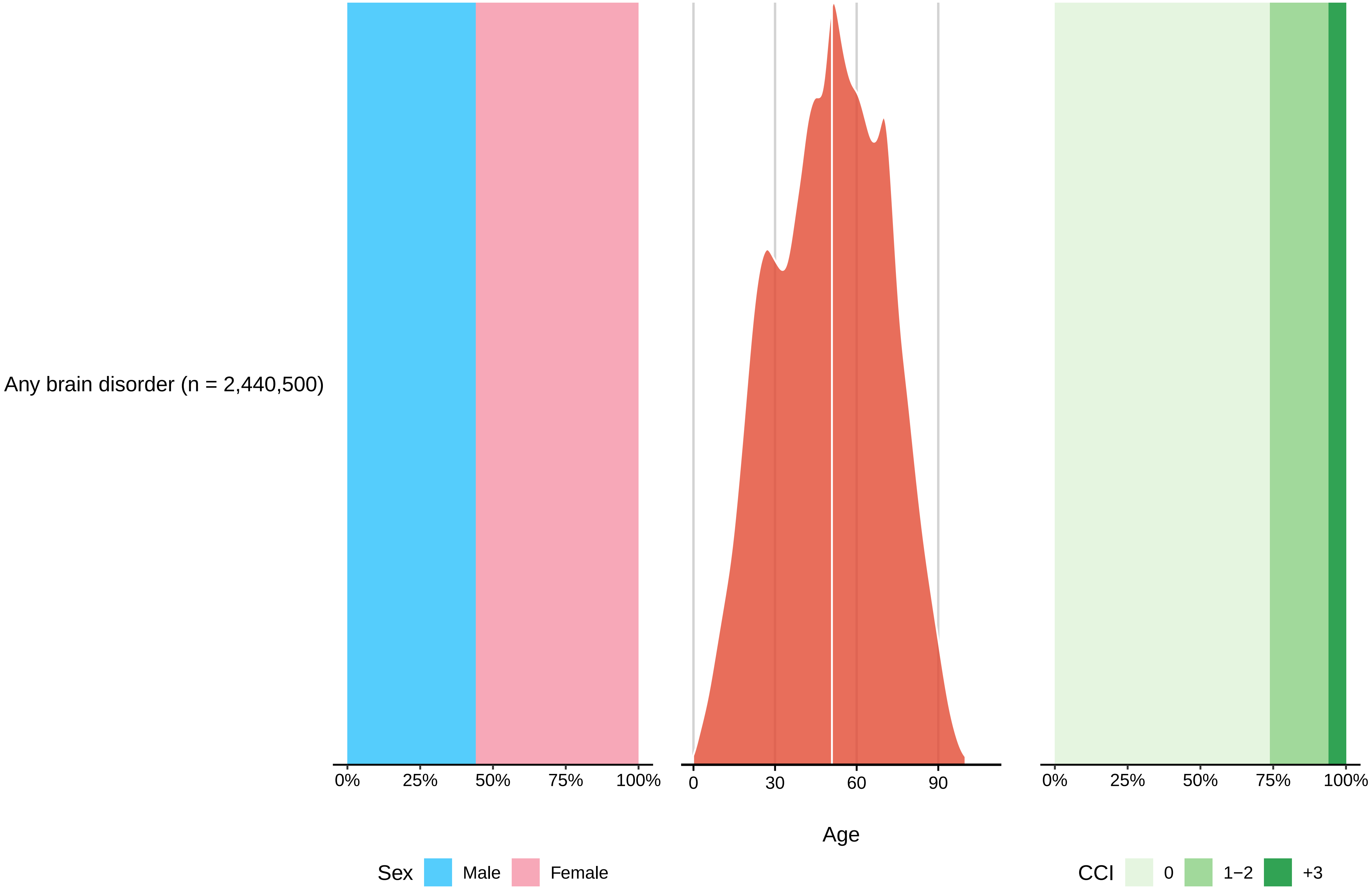

Patient characteristics

Sensitivity analysis 3 / Prevalent cohort 2018

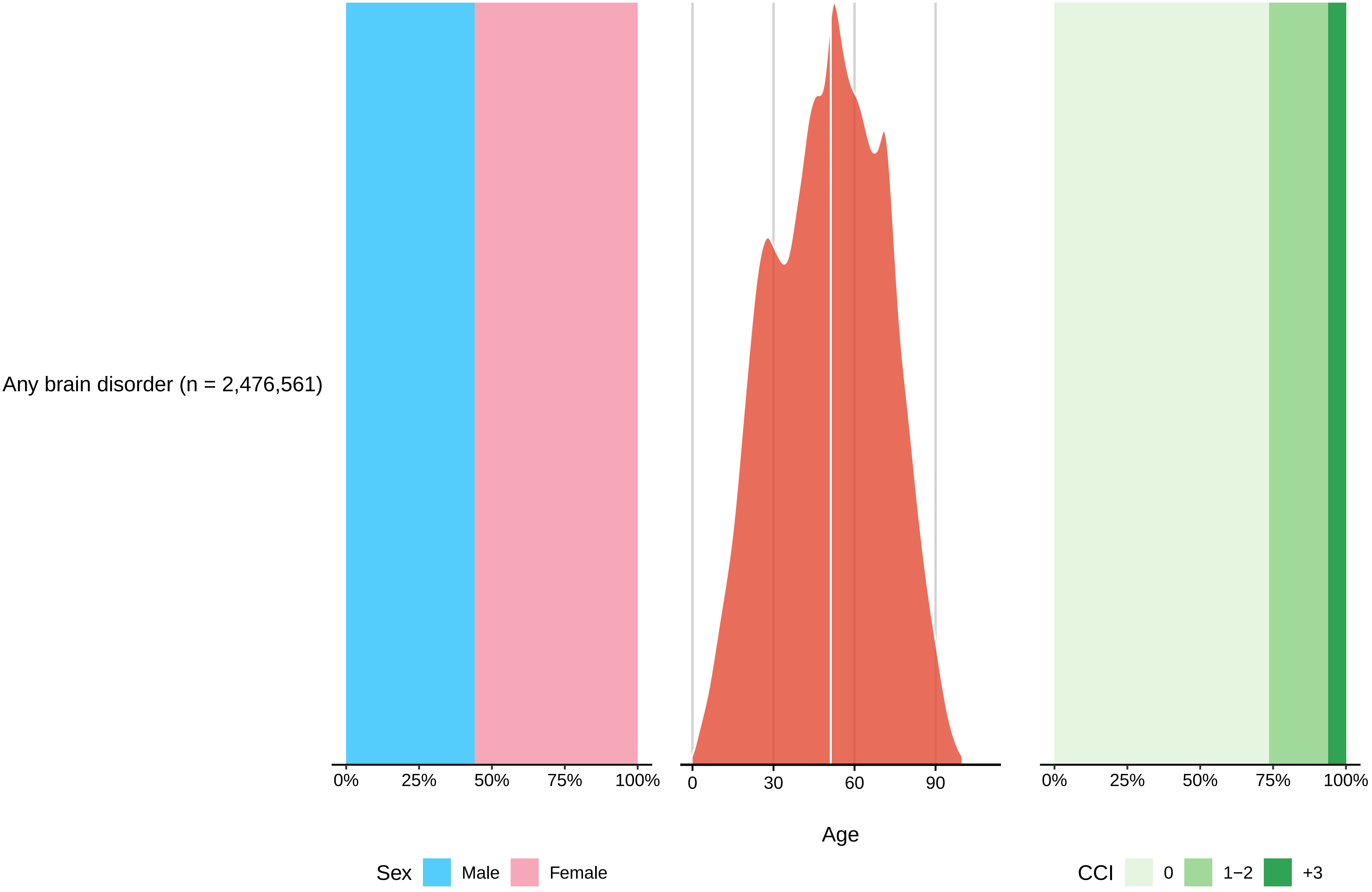

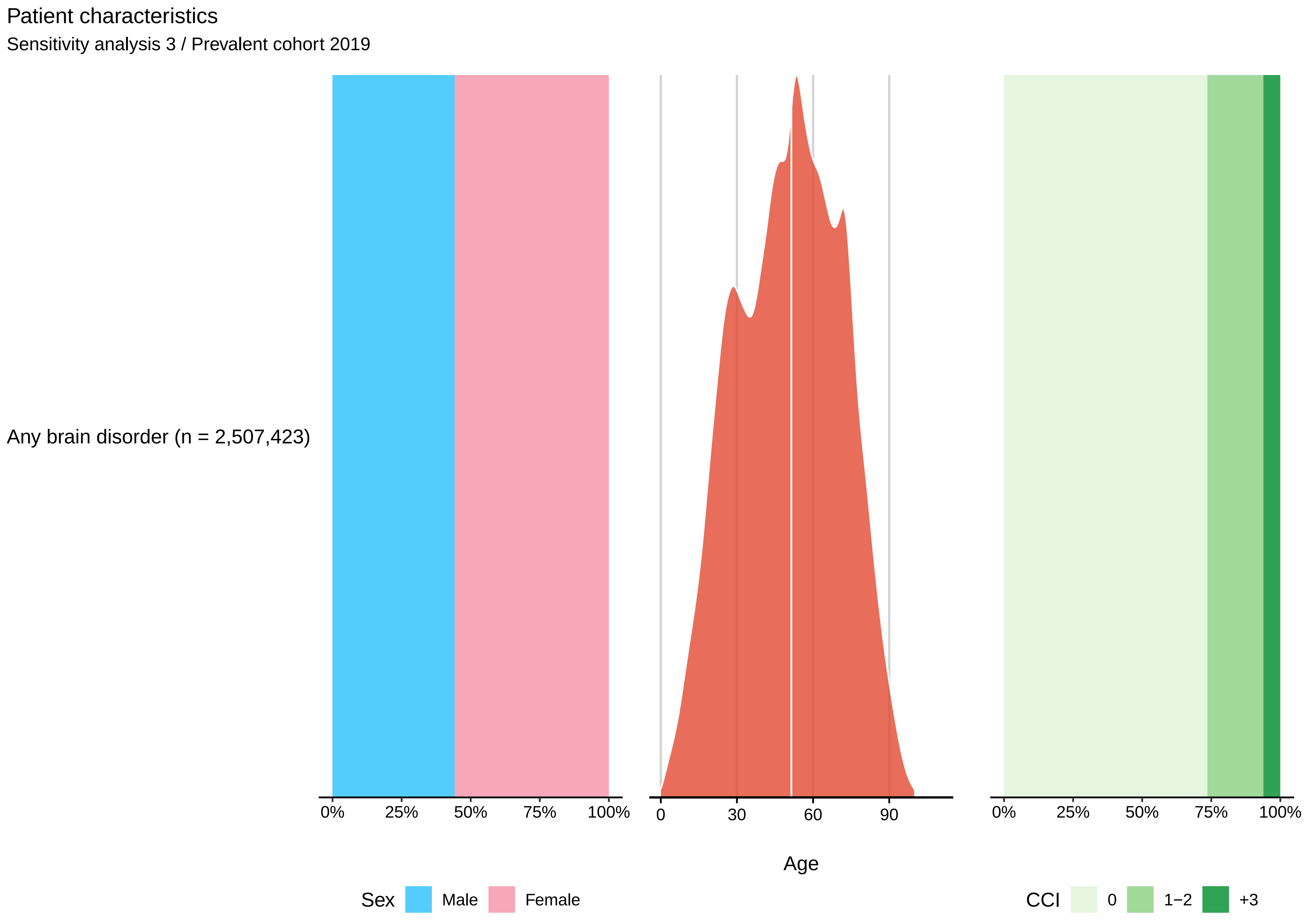
Patient characteristics
Sensitivity analysis 3 / Prevalent cohort 2019
Any brain disorder (n = 2,507,423)
0%
25%
50%
75%
100%
0
30
60
90
Age
0%
25%
50%
75%
100%
Sex
Male
Female
CCI
0
1−2
+3

#### Patient characteristics

Sensitivity analysis 3 / Prevalent cohort 2020

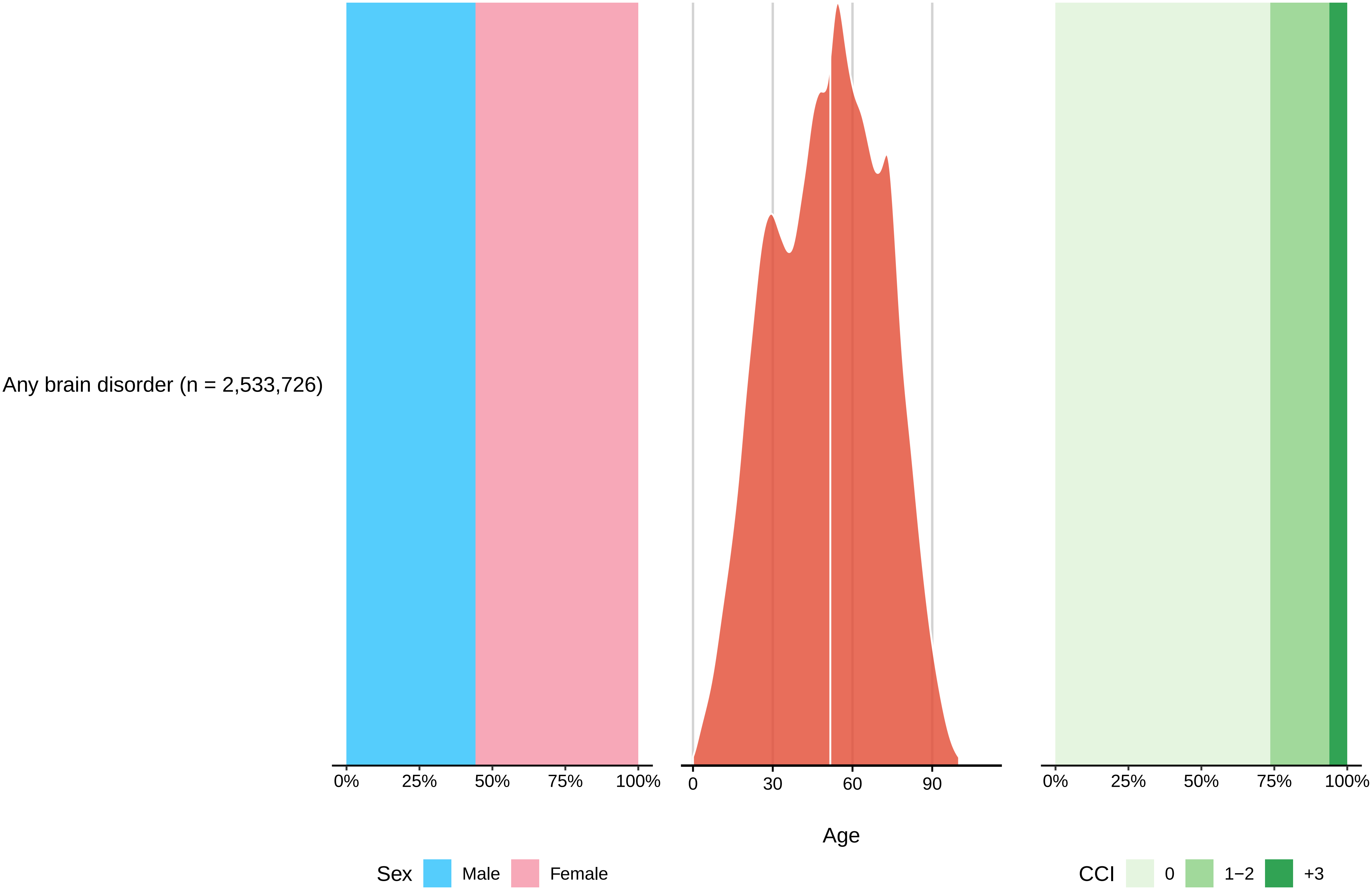

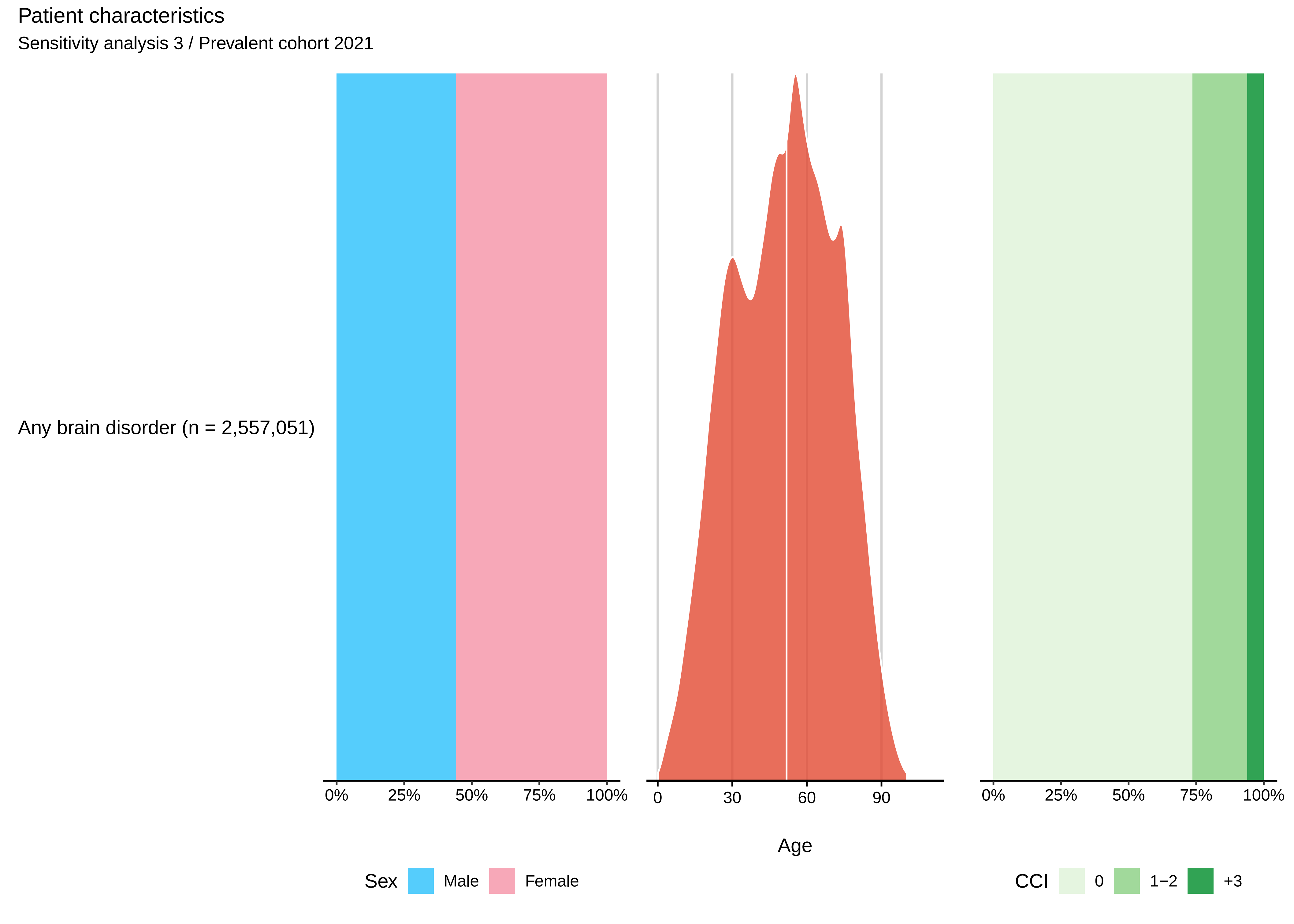

Patient characteristics
Sensitivity analysis 3 / Prevalent cohort 2021
Any brain disorder (n = 2,557,051)
0%
25%
50%
75%
100%
0
30
60
90
Age
0%
25%
50%
75%
100%
Sex
Male
Female
CCI
0
1−2
+3

***Sensitivity analysis 3 / Incident cohort 2011-2015***

| | *Incidence, N* | *Incidence rate per 100,000 person-years (95% CI)* |
|---|---|---|
| Any brain disorder | 580,993 | 2,050 (2,045-2,055) |

***Sensitivity analysis 3 / Incident cohort 2016-2021***

| | *Incidence, N* | *Incidence rate per 100,000 person-years (95% CI)* |
|---|---|---|
| Any brain disorder | 636,299 | 1,820 (1,816-1,825) |

***Sensitivity analysis 3 / Prevalent cohort 2015***

| | *Prevalence, N (%)* | *Prevalence per 100,000 persons (95% CI)* |
|---|---|---|
| Any brain disorder | 2,361,150 (41.4) | 41,383 (41,331-41,436) |

***Sensitivity analysis 3 / Prevalent cohort 2016***

| | *Prevalence, N (%)* | *Prevalence per 100,000 persons (95% CI)* |
|---|---|---|
| Any brain disorder | 2,402,477 (41.7) | 41,692 (41,639-41,744) |

***Sensitivity analysis 3 / Prevalent cohort 2017***

| | *Prevalence, N (%)* | *Prevalence per 100,000 persons (95% CI)* |
|---|---|---|
| Any brain disorder | 2,440,618 (42.2) | 42,209 (42,156-42,262) |

***Sensitivity analysis 3 / Prevalent cohort 2018***

| | *Prevalence, N (%)* | *Prevalence per 100,000 persons (95% CI)* |
|---|---|---|
| Any brain disorder | 2,476,670 (42.6) | 42,634 (42,581-42,687) |

***Sensitivity analysis 3 / Prevalent cohort 2019***

| | *Prevalence, N (%)* | *Prevalence per 100,000 persons (95% CI)* |
|---|---|---|
| Any brain disorder | 2,507,545 (43.0) | 43,017 (42,964-43,070) |

***Sensitivity analysis 3 / Prevalent cohort 2020***

| | *Prevalence, N (%)* | *Prevalence per 100,000 persons (95% CI)* |
|---|---|---|
| Any brain disorder | 2,533,805 (43.2) | 43,228 (43,174-43,281) |

***Sensitivity analysis 3 / Prevalent cohort 2021***

| | *Prevalence, N (%)* | *Prevalence per 100,000 persons (95% CI)* |
|---|---|---|
| Any brain disorder | 2,557,118 (43.7) | 43,662 (43,608-43,715) |

Prevalence

Sensitivity analysis 3

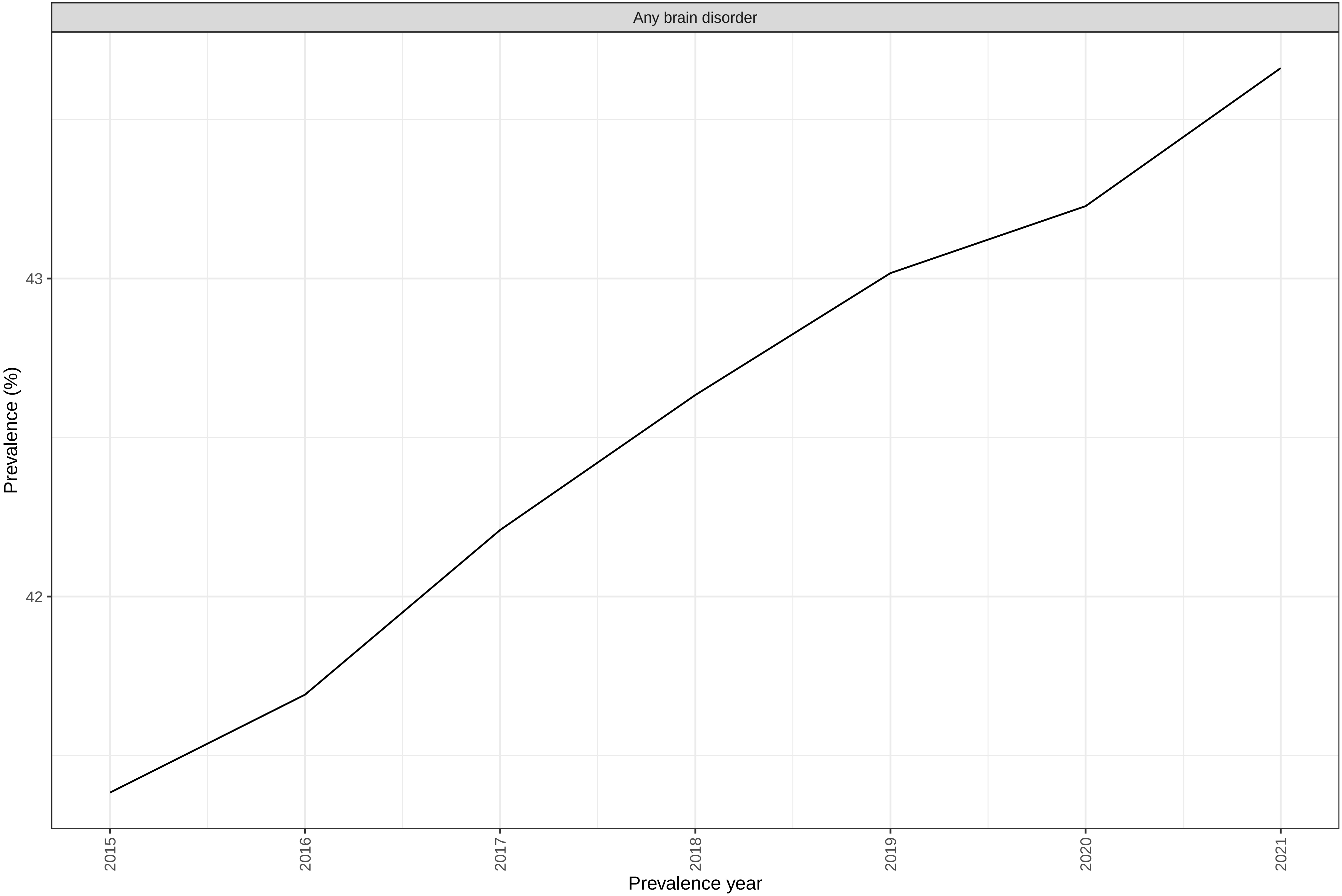

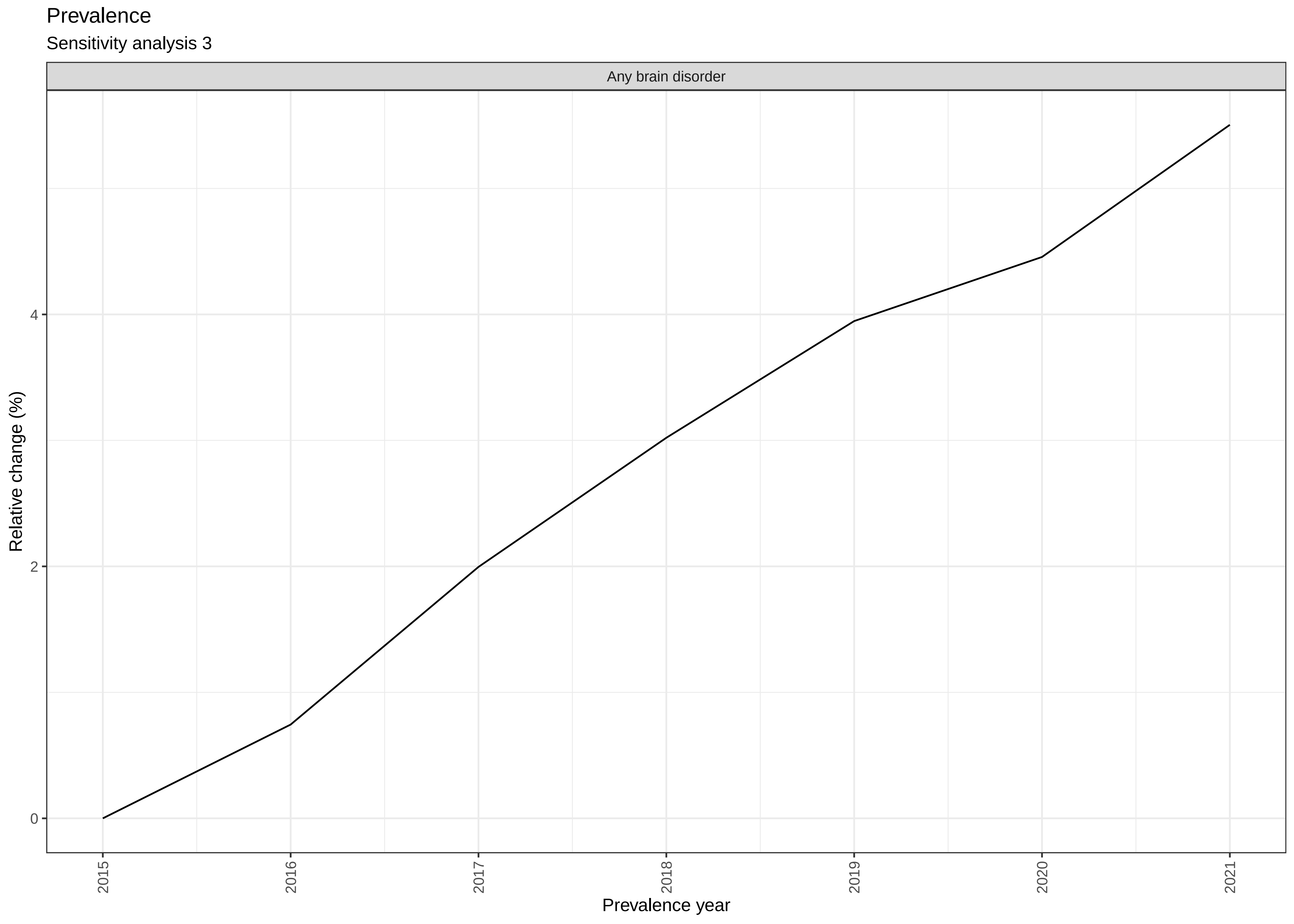

Prevalence
Sensitivity analysis 3
Any brain disorder
Relative change (%)
0
2
4
2015
2016
2017
2018
2019
2020
2021
Prevalence year

***Sensitivity analysis 3 / Incident cohort 2011-2015***

| *Brain disorder* | *Patient group* | | *Comparison group* | | |
|---|---|---|---|---|---|
| | *Persons, N* | *Deaths, N(%)* | *Persons, N* | *Deaths, N(%)* | *Adjusted HR (95% CI)* |
| Any brain disorder | 578,267 | 26,297 (4.5) | 5,767,566 | 46,347 (0.8) | 5.6 (5.5-5.6) |

***Sensitivity analysis 3 / Incident cohort 2016-2021***

| *Brain disorder* | *Patient group* | | *Comparison group* | | |
|---|---|---|---|---|---|
| | *Persons, N* | *Deaths, N(%)* | *Persons, N* | *Deaths, N(%)* | *Adjusted HR (95% CI)* |
| Any brain disorder | 632,009 | 27,586 (4.4) | 6,305,663 | 52,685 (0.8) | 5.4 (5.3-5.4) |

#### Mortality

Sensitivity analysis 3 / Incident cohort 2011-2015

| Brain disorder | Patient group -Deaths (%) | Comparison group- Deaths (%) | Hazard Ratio | Adjusted hazard ratio (95% CI) |
|---|---|---|---|---|
| Any brain disorder | 26,297 (4.5) | 46,347 (0.8) | 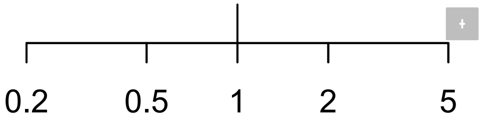 | 5.6 (5.5-5.6) |

### Mortality

Sensitivity analysis 3 / Incident cohort 2016-2021

| Brain disorder | Patient group -Deaths (%) | Comparison group- Deaths (%) | Hazard Ratio | Adjusted hazard ratio (95% CI) |
|---|---|---|---|---|
| Any brain disorder | 27,586 (4.4) | 52,685 (0.8) | 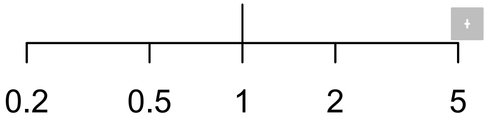 | 5.4 (5.3-5.4) |

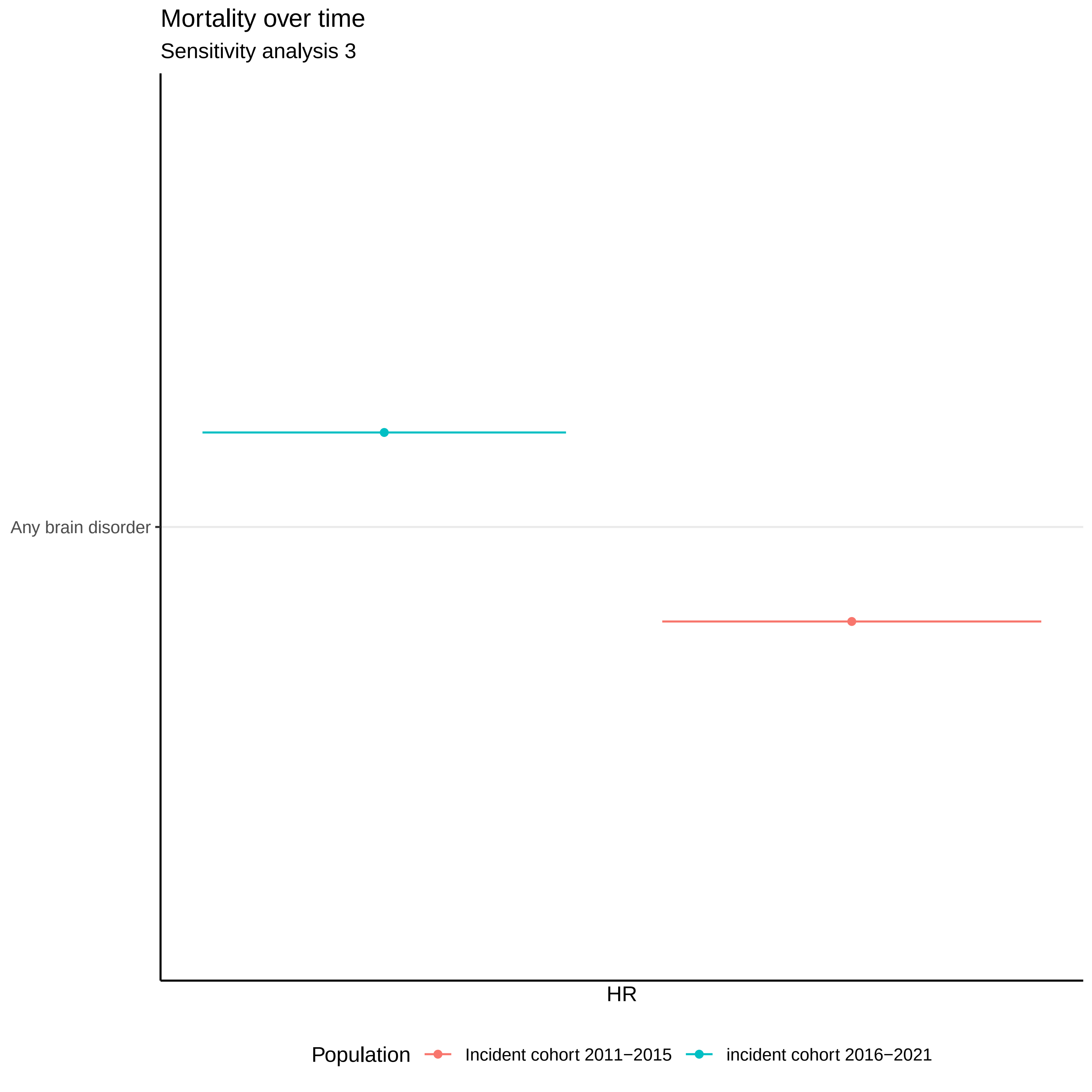
Mortality over time
Sensitivity analysis 3
Any brain disorder
HR
Population
Incident cohort 2011−2015
incident cohort 2016−2021

***Sensitivity analysis 3 / Incident cohort 2011-2015***

| *Brain disorder* | *Cost component* | *Actual costs (EUR)* | *Actual costs per person* | *Attributable costs (EUR)* | *Attributable costs per person* |
|---|---|---|---|---|---|
| Any brain disorder | Primary sector | 262,210,303 | 467 | 126,003,073 | 225 |
| | Hospital care | 4,666,044,988 | 8,316 | 3,839,240,569 | 6,843 |
| | Filled prescriptions | 166,580,032 | 297 | 86,851,764 | 155 |
| | Home care | 218,436,073 | 389 | 151,741,177 | 270 |
| | Nursing home / sheltered accomodation | 417,164,403 | 744 | 317,760,752 | 566 |
| | Lost productivity associated with illness | . | . | 2,672,278,648 | 7,854 |
| | Total cost (excl. lost productivity) | 5,730,435,800 | 10,213 | 4,521,597,335 | 8,059 |

***Sensitivity analysis 3 / Incident cohort 2016-2021***

| *Brain disorder* | *Cost component* | *Actual costs (EUR)* | *Actual costs per person* | *Attributable costs (EUR)* | *Attributable costs per person* |
|---|---|---|---|---|---|
| Any brain disorder | Primary sector | 292,643,489 | 476 | 147,677,488 | 240 |
| | Hospital care | 4,480,256,202 | 7,288 | 3,650,338,222 | 5,938 |
| | Filled prescriptions | 185,631,830 | 302 | 92,507,170 | 150 |
| | Home care | 202,630,194 | 330 | 142,386,503 | 232 |
| | Nursing home / sheltered accomodation | 461,567,357 | 751 | 354,455,075 | 577 |
| | Lost productivity associated with illness | . | . | 3,007,980,917 | 8,426 |
| | Total cost (excl. lost productivity) | 5,622,729,072 | 9,146 | 4,387,364,458 | 7,137 |

***Sensitivity analysis 3 / Prevalent cohort 2015***

| *Brain disorder* | *Cost component* | *Actual costs (EUR)* | *Actual costs per person* | *Attributable costs (EUR)* | *Attributable costs per person* |
|---|---|---|---|---|---|
| Any brain disorder | Primary sector | 946,466,695 | 405 | 331,011,998 | 142 |
| | Hospital care | 8,219,346,394 | 3,515 | 4,415,691,505 | 1,888 |
| | Filled prescriptions | 951,167,235 | 407 | 564,394,580 | 241 |
| | Home care | 1,003,926,433 | 429 | 714,540,808 | 306 |
| | Nursing home / sheltered accomodation | 2,858,340,074 | 1,222 | 2,394,551,245 | 1,024 |
| | Lost productivity associated with illness | . | . | 20,996,403,604 | 13,386 |
| | Total cost (excl. lost productivity) | 13,979,246,830 | 5,978 | 8,420,190,135 | 3,601 |

***Sensitivity analysis 3 / Prevalent cohort 2016***

| *Brain disorder* | *Cost component* | *Actual costs (EUR)* | *Actual costs per person* | *Attributable costs (EUR)* | *Attributable costs per person* |
|---|---|---|---|---|---|
| Any brain disorder | Primary sector | 937,622,509 | 394 | 328,431,304 | 138 |
| | Hospital care | 8,203,010,147 | 3,447 | 4,397,127,111 | 1,848 |
| | Filled prescriptions | 943,397,732 | 396 | 553,146,591 | 232 |
| | Home care | 882,400,184 | 371 | 632,186,154 | 266 |
| | Nursing home / sheltered accomodation | 2,879,277,358 | 1,210 | 2,420,942,511 | 1,017 |
| | Lost productivity associated with illness | . | . | 21,059,159,074 | 13,234 |
| | Total cost (excl. lost productivity) | 13,845,707,929 | 5,818 | 8,331,833,670 | 3,501 |

***Sensitivity analysis 3 / Prevalent cohort 2017***

| *Brain disorder* | *Cost component* | *Actual costs (EUR)* | *Actual costs per person* | *Attributable costs (EUR)* | *Attributable costs per person* |
|---|---|---|---|---|---|
| Any brain disorder | Primary sector | 964,198,454 | 399 | 337,997,069 | 140 |
| | Hospital care | 8,806,887,074 | 3,643 | 4,595,188,553 | 1,901 |
| | Filled prescriptions | 924,756,761 | 383 | 528,086,817 | 218 |
| | Home care | 923,146,831 | 382 | 674,376,981 | 279 |
| | Nursing home / sheltered accomodation | 2,881,032,546 | 1,192 | 2,418,998,901 | 1,001 |
| | Lost productivity associated with illness | . | . | 22,256,252,694 | 13,815 |
| | Total cost (excl. lost productivity) | 14,500,021,666 | 5,999 | 8,554,648,320 | 3,539 |

***Sensitivity analysis 3 / Prevalent cohort 2018***

| *Brain disorder* | *Cost component* | *Actual costs (EUR)* | *Actual costs per person* | *Attributable costs (EUR)* | *Attributable costs per person* |
|---|---|---|---|---|---|
| Any brain disorder | Primary sector | 977,676,498 | 399 | 356,016,207 | 145 |
| | Hospital care | 6,608,035,048 | 2,695 | 3,564,891,991 | 1,454 |
| | Filled prescriptions | 934,762,492 | 381 | 530,208,658 | 216 |
| | Home care | 781,661,901 | 319 | 568,176,890 | 232 |
| | Nursing home / sheltered accomodation | 2,869,918,942 | 1,170 | 2,413,869,603 | 984 |
| | Lost productivity associated with illness | . | . | 23,105,105,904 | 14,190 |
| | Total cost (excl. lost productivity) | 12,172,054,881 | 4,964 | 7,433,163,349 | 3,032 |

***Sensitivity analysis 3 / Prevalent cohort 2019***

| *Brain disorder* | *Cost component* | *Actual costs (EUR)* | *Actual costs per person* | *Attributable costs (EUR)* | *Attributable costs per person* |
|---|---|---|---|---|---|
| Any brain disorder | Primary sector | 987,931,568 | 398 | 358,065,139 | 144 |
| | Hospital care | 7,577,589,755 | 3,051 | 3,957,035,043 | 1,593 |
| | Filled prescriptions | 993,817,202 | 400 | 548,025,525 | 221 |
| | Home care | 901,346,766 | 363 | 659,320,518 | 265 |
| | Nursing home / sheltered accomodation | 2,884,732,810 | 1,162 | 2,427,524,881 | 977 |
| | Lost productivity associated with illness | . | . | 24,070,980,630 | 14,642 |
| | Total cost (excl. lost productivity) | 13,345,418,101 | 5,374 | 7,949,971,106 | 3,201 |

***Sensitivity analysis 3 / Prevalent cohort 2020***

| *Brain disorder* | *Cost component* | *Actual costs (EUR)* | *Actual costs per person* | *Attributable costs (EUR)* | *Attributable costs per person* |
|---|---|---|---|---|---|
| Any brain disorder | Primary sector | 1,005,601,461 | 401 | 362,539,440 | 144 |
| | Hospital care | 8,030,329,507 | 3,199 | 4,195,167,196 | 1,671 |
| | Filled prescriptions | 1,032,549,501 | 411 | 568,374,236 | 226 |
| | Home care | 1,013,199,924 | 404 | 734,344,379 | 293 |
| | Nursing home / sheltered accomodation | 2,902,488,657 | 1,156 | 2,456,664,965 | 979 |
| | Lost productivity associated with illness | . | . | 25,606,406,601 | 15,475 |
| | Total cost (excl. lost productivity) | 13,984,169,050 | 5,571 | 8,317,090,216 | 3,313 |

***Sensitivity analysis 3 / Prevalent cohort 2021***

| *Brain disorder* | *Cost component* | *Actual costs (EUR)* | *Actual costs per person* | *Attributable costs (EUR)* | *Attributable costs per person* |
|---|---|---|---|---|---|
| Any brain disorder | Primary sector | 1,042,449,950 | 412 | 376,531,582 | 149 |
| | Hospital care | 8,045,029,219 | 3,177 | 4,183,767,588 | 1,652 |
| | Filled prescriptions | 1,013,144,582 | 400 | 551,943,862 | 218 |
| | Home care | 1,143,324,621 | 451 | 838,792,375 | 331 |
| | Nursing home / sheltered accomodation | 2,883,224,432 | 1,139 | 2,441,861,589 | 964 |
| | Lost productivity associated with illness | . | . | 27,140,175,211 | 16,324 |
| | Total cost (excl. lost productivity) | 14,127,172,803 | 5,578 | 8,392,896,997 | 3,314 |

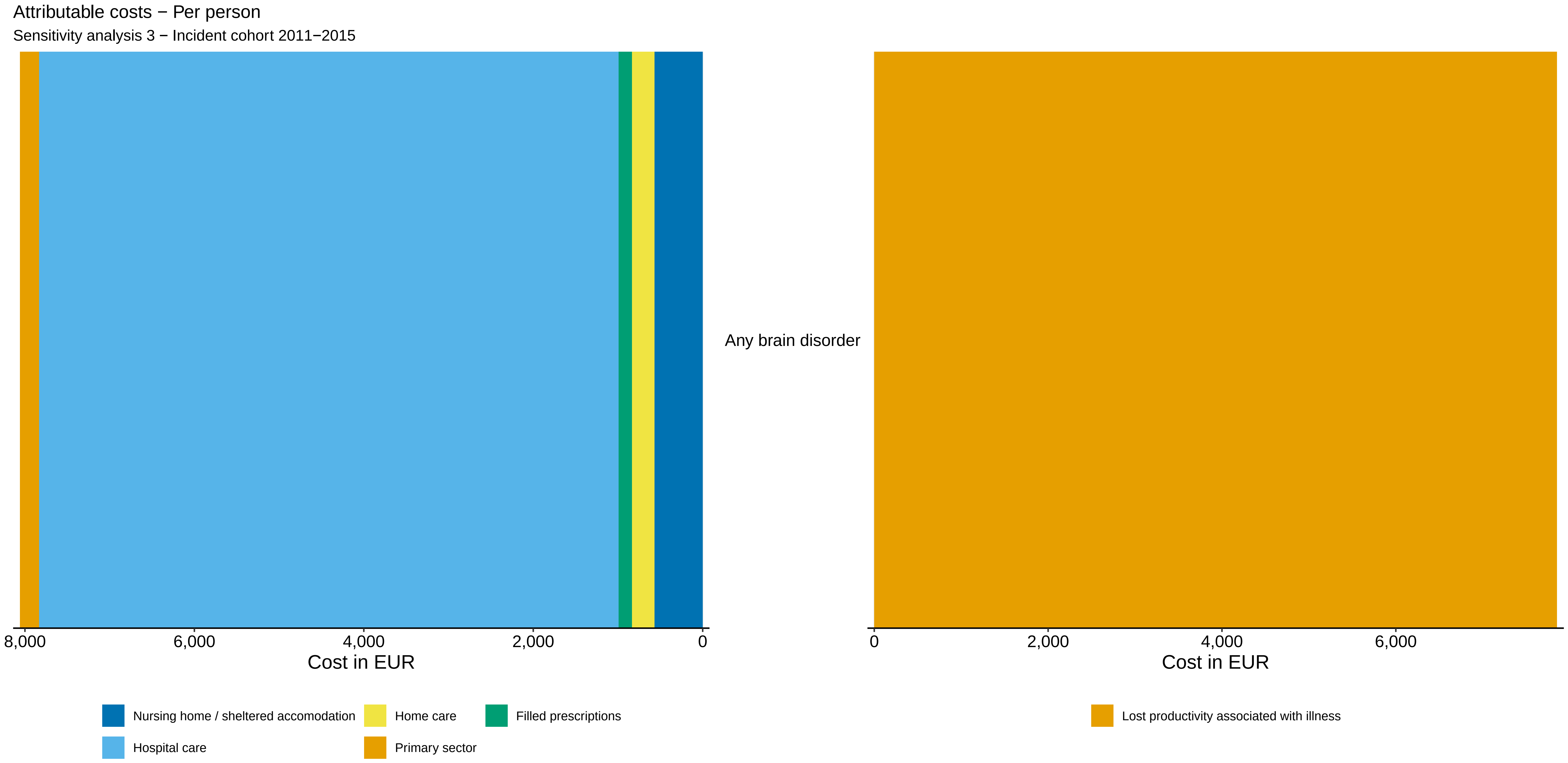

Attributable costs − Per person
Sensitivity analysis 3 − Incident cohort 2011−2015
Any brain disorder
8,000
6,000
4,000
2,000
0
Cost in EUR
0
2,000
4,000
6,000
Cost in EUR
Nursing home / sheltered accomodation
Home care
Filled prescriptions
Hospital care
Primary sector
Lost productivity associated with illness

Attributable costs − Per person

Sensitivity analysis 3 − Incident cohort 2016−2021

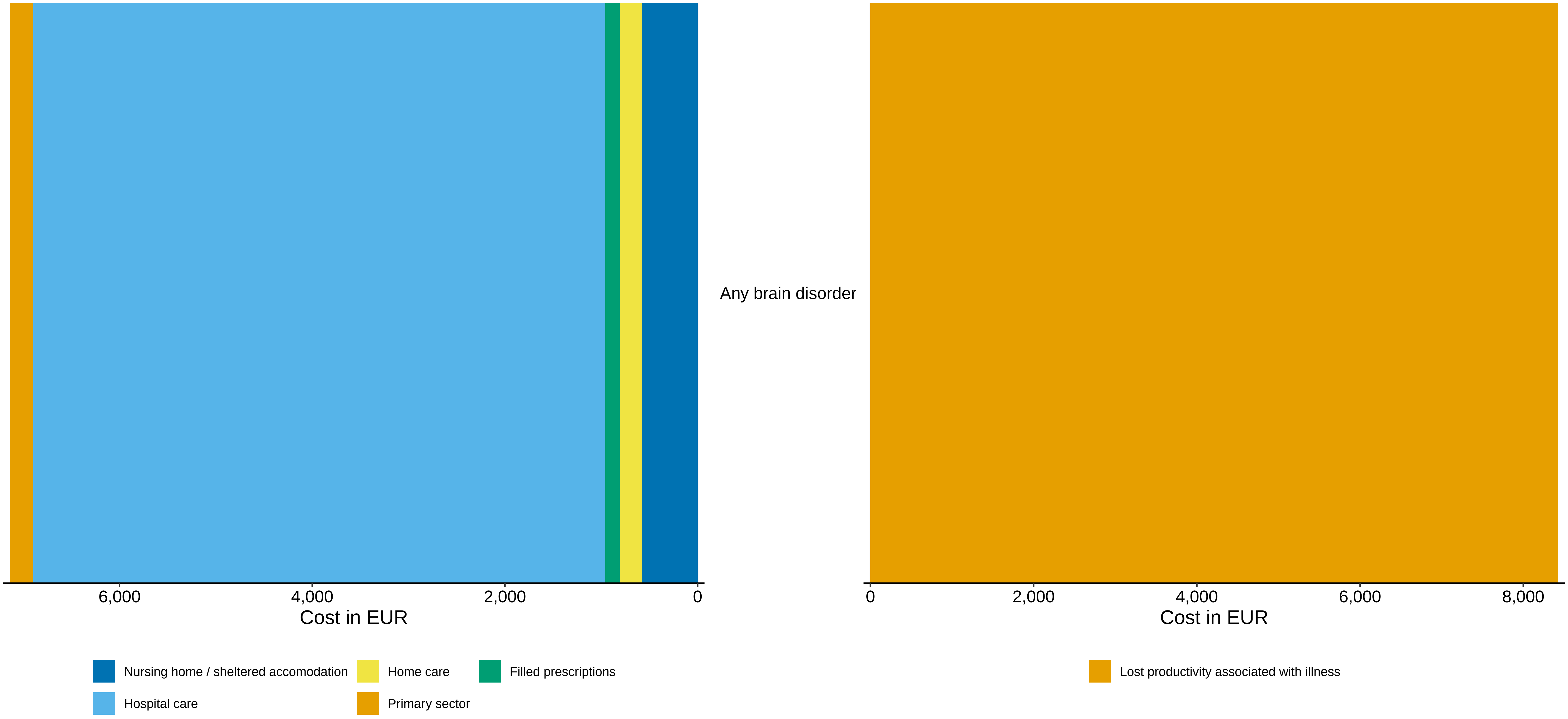

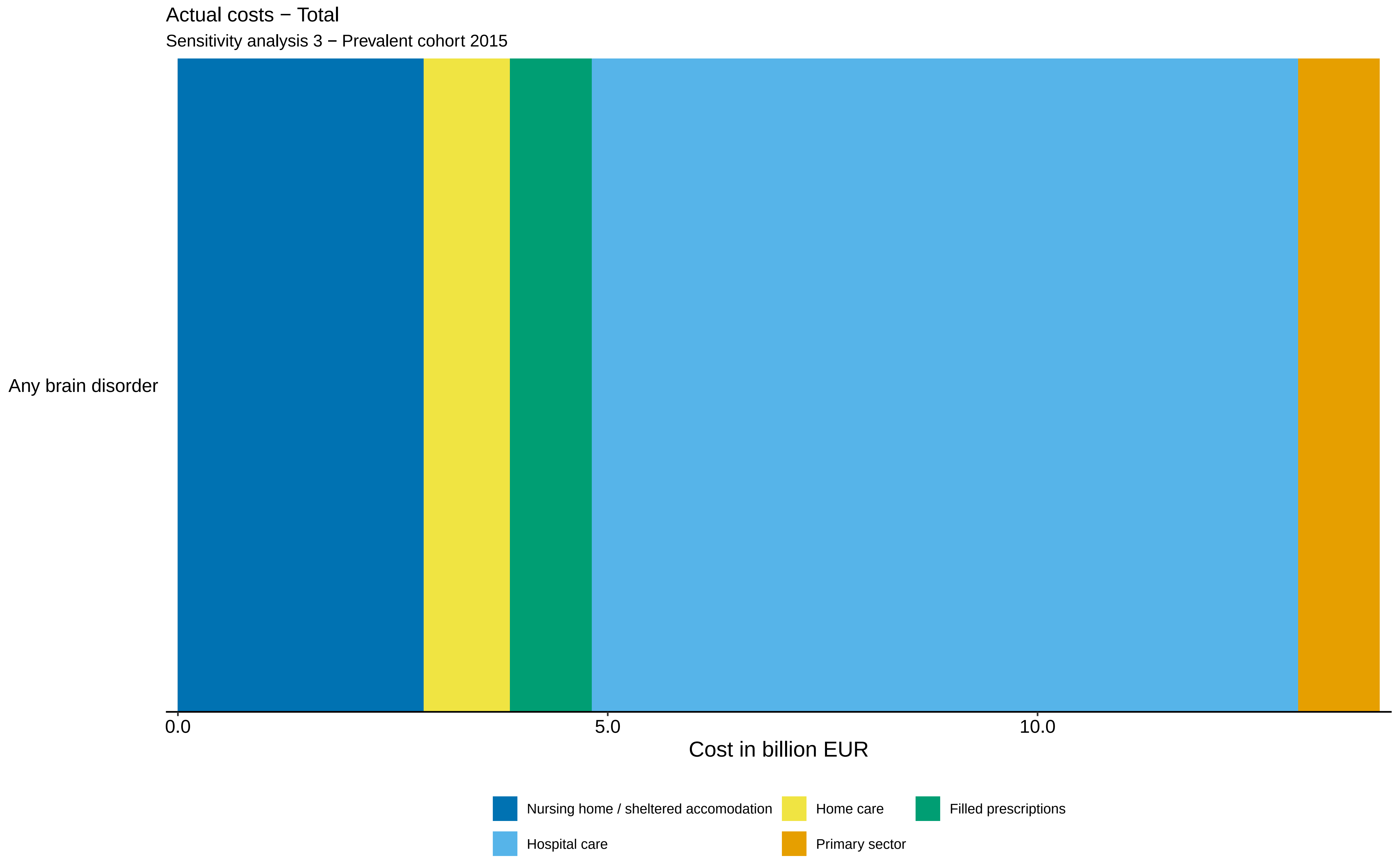

Actual costs − Total
Sensitivity analysis 3 − Prevalent cohort 2015
Any brain disorder
0.0
5.0
10.0
Cost in billion EUR
Nursing home / sheltered accomodation
Home care
Filled prescriptions
Hospital care
Primary sector

Actual costs − Total

Sensitivity analysis 3 − Prevalent cohort 2016

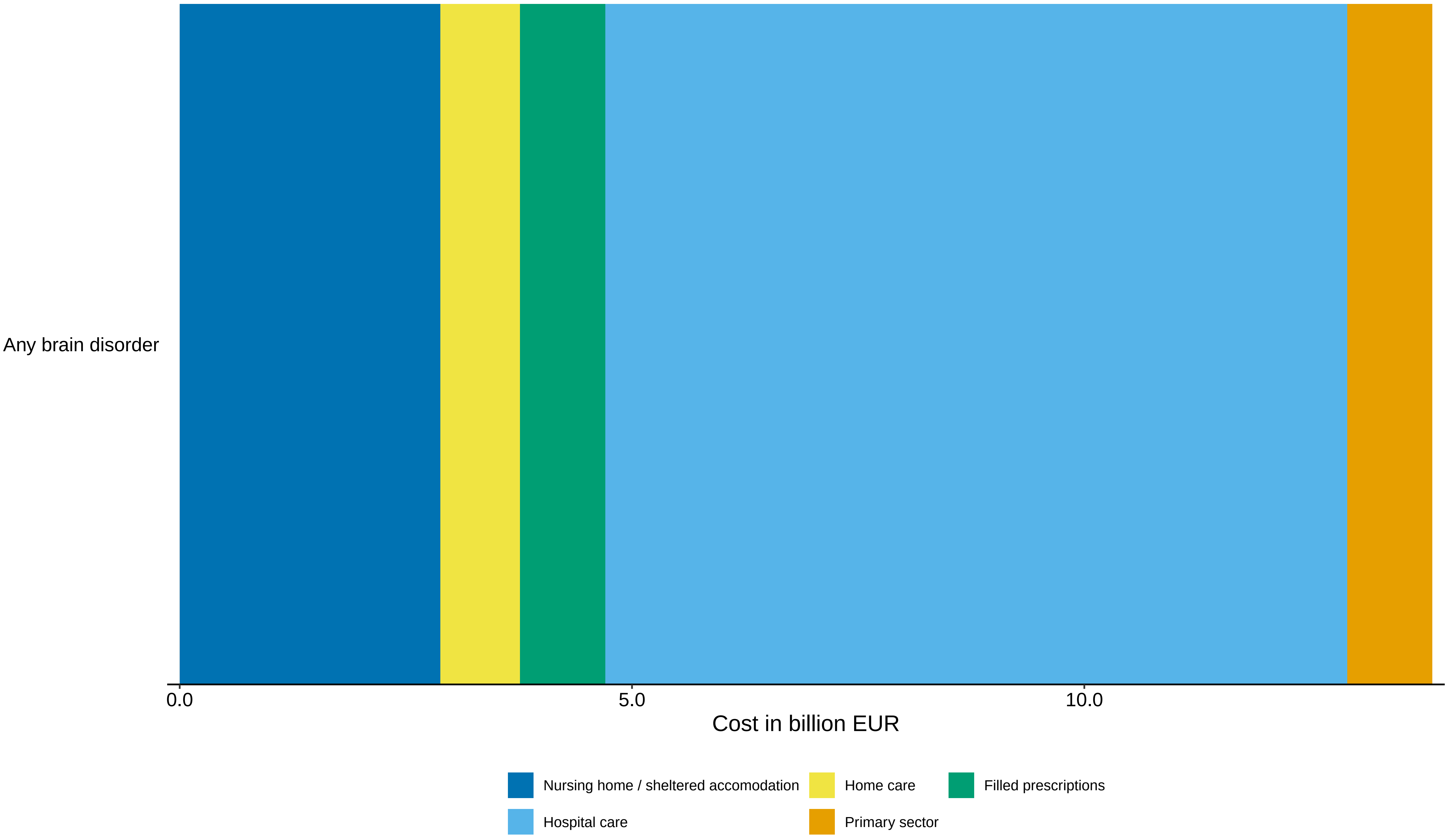

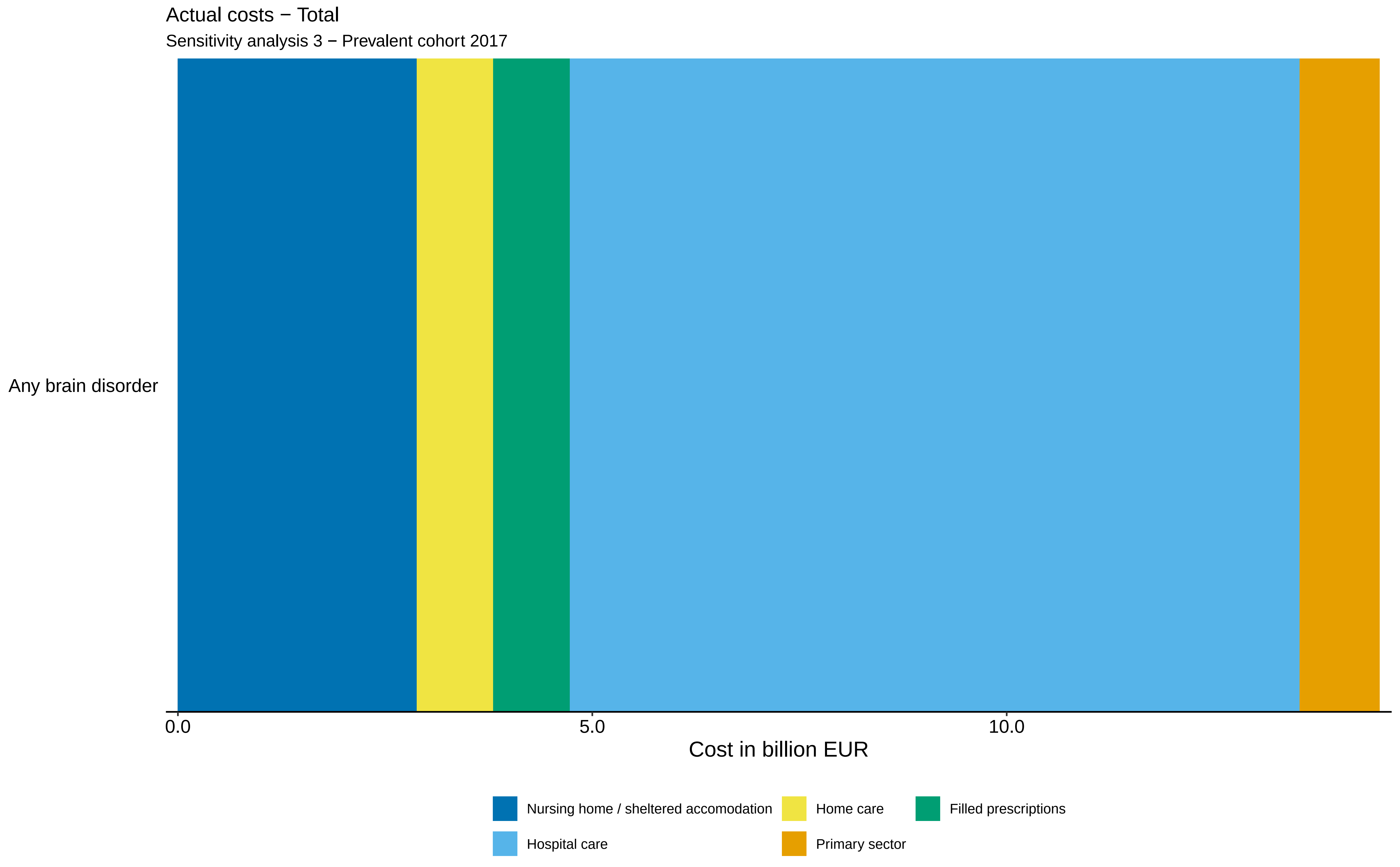
Actual costs − Total
Sensitivity analysis 3 − Prevalent cohort 2017
Any brain disorder
0.0
5.0
10.0
Cost in billion EUR
Nursing home / sheltered accomodation
Home care
Filled prescriptions
Hospital care
Primary sector

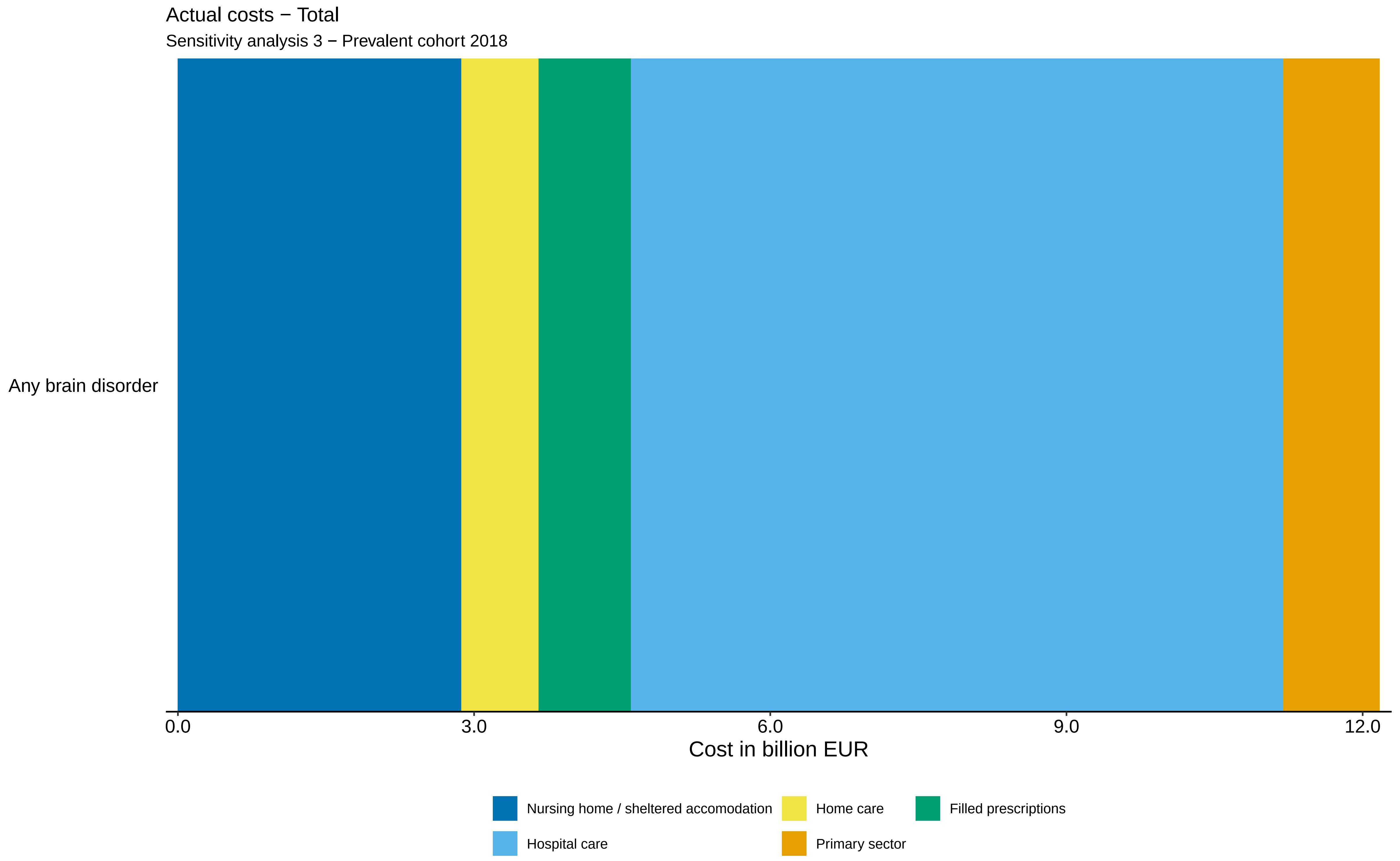
Actual costs − Total
Sensitivity analysis 3 − Prevalent cohort 2018
Any brain disorder
0.0
3.0
6.0
9.0
12.0
Cost in billion EUR
Nursing home / sheltered accomodation
Home care
Filled prescriptions
Hospital care
Primary sector

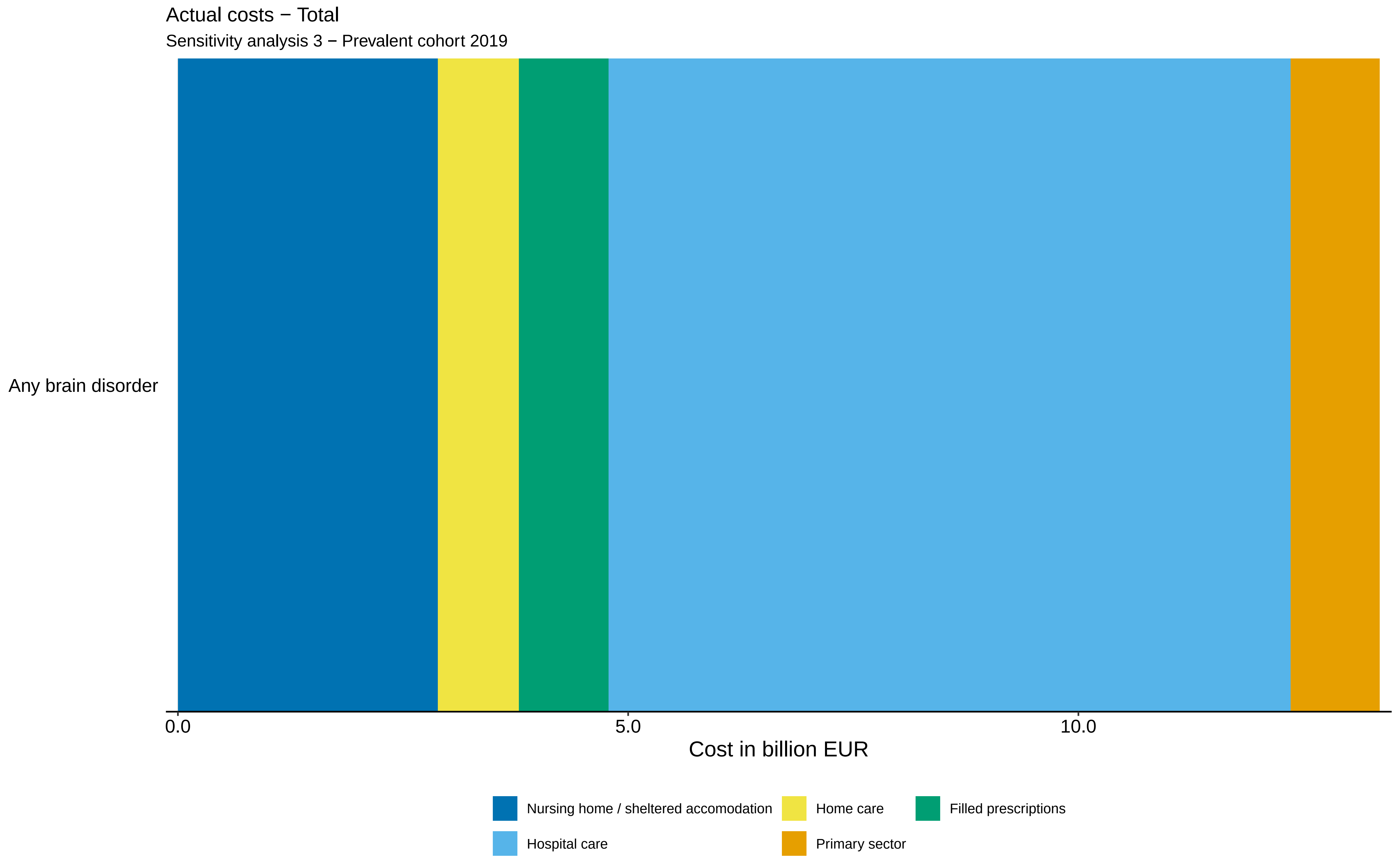
Actual costs − Total
Sensitivity analysis 3 − Prevalent cohort 2019
Any brain disorder
0.0
5.0
10.0
Cost in billion EUR
Nursing home / sheltered accomodation
Home care
Filled prescriptions
Hospital care
Primary sector

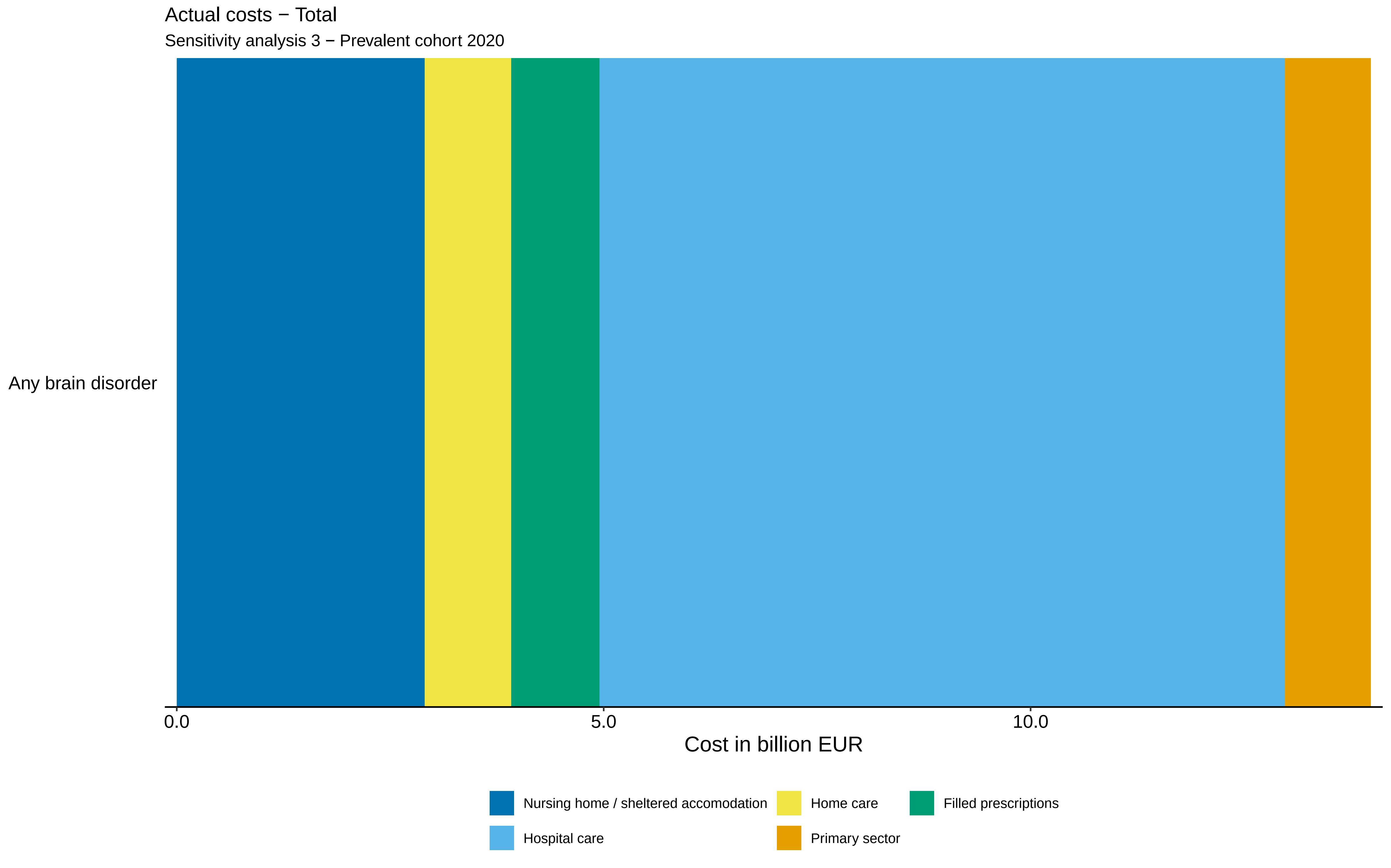

Actual costs − Total
Sensitivity analysis 3 − Prevalent cohort 2020
Any brain disorder
0.0
5.0
10.0
Cost in billion EUR
Nursing home / sheltered accomodation
Home care
Filled prescriptions
Hospital care
Primary sector

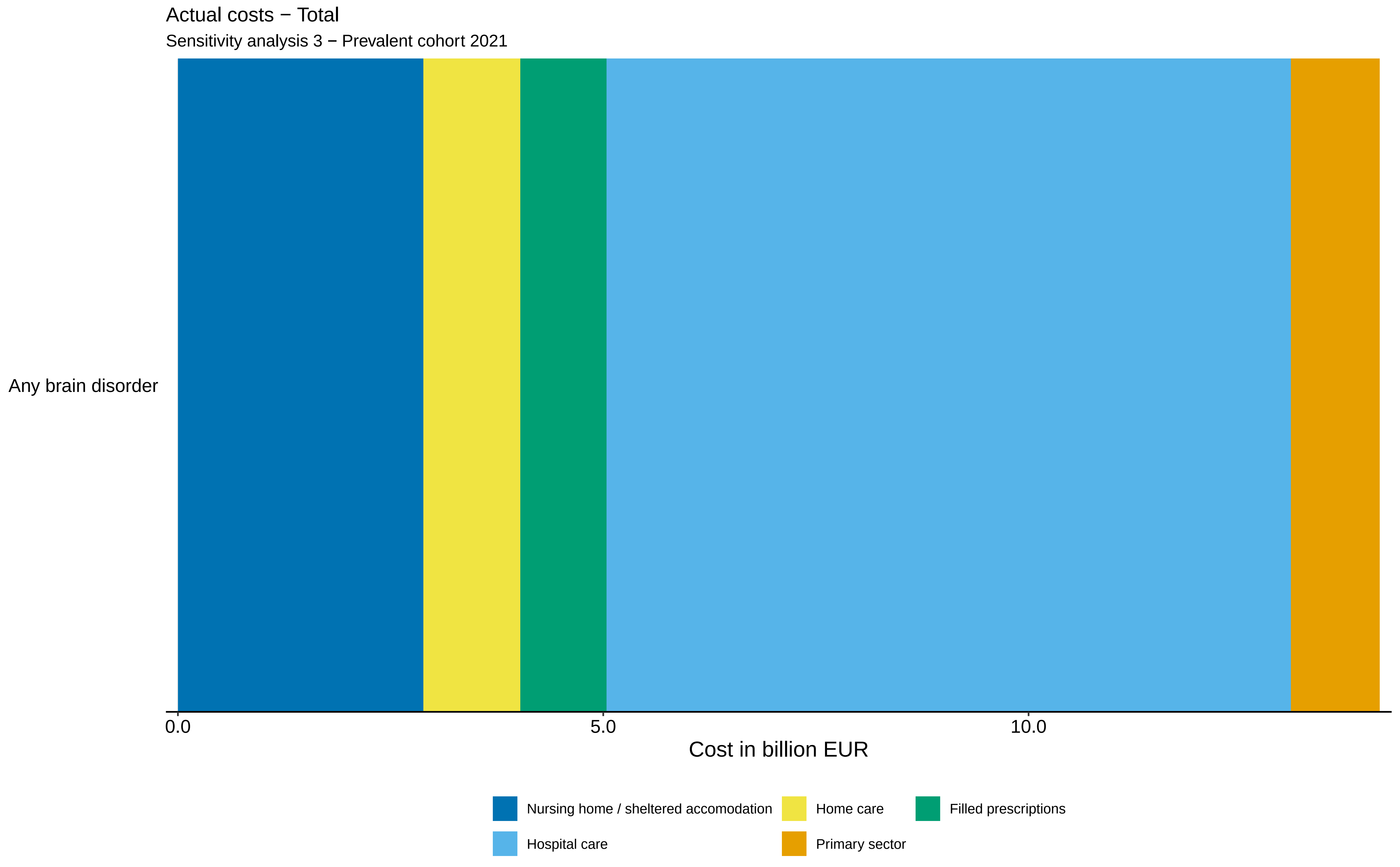
Actual costs − Total
Sensitivity analysis 3 − Prevalent cohort 2021
Any brain disorder
0.0
5.0
10.0
Cost in billion EUR
Nursing home / sheltered accomodation
Home care
Filled prescriptions
Hospital care
Primary sector

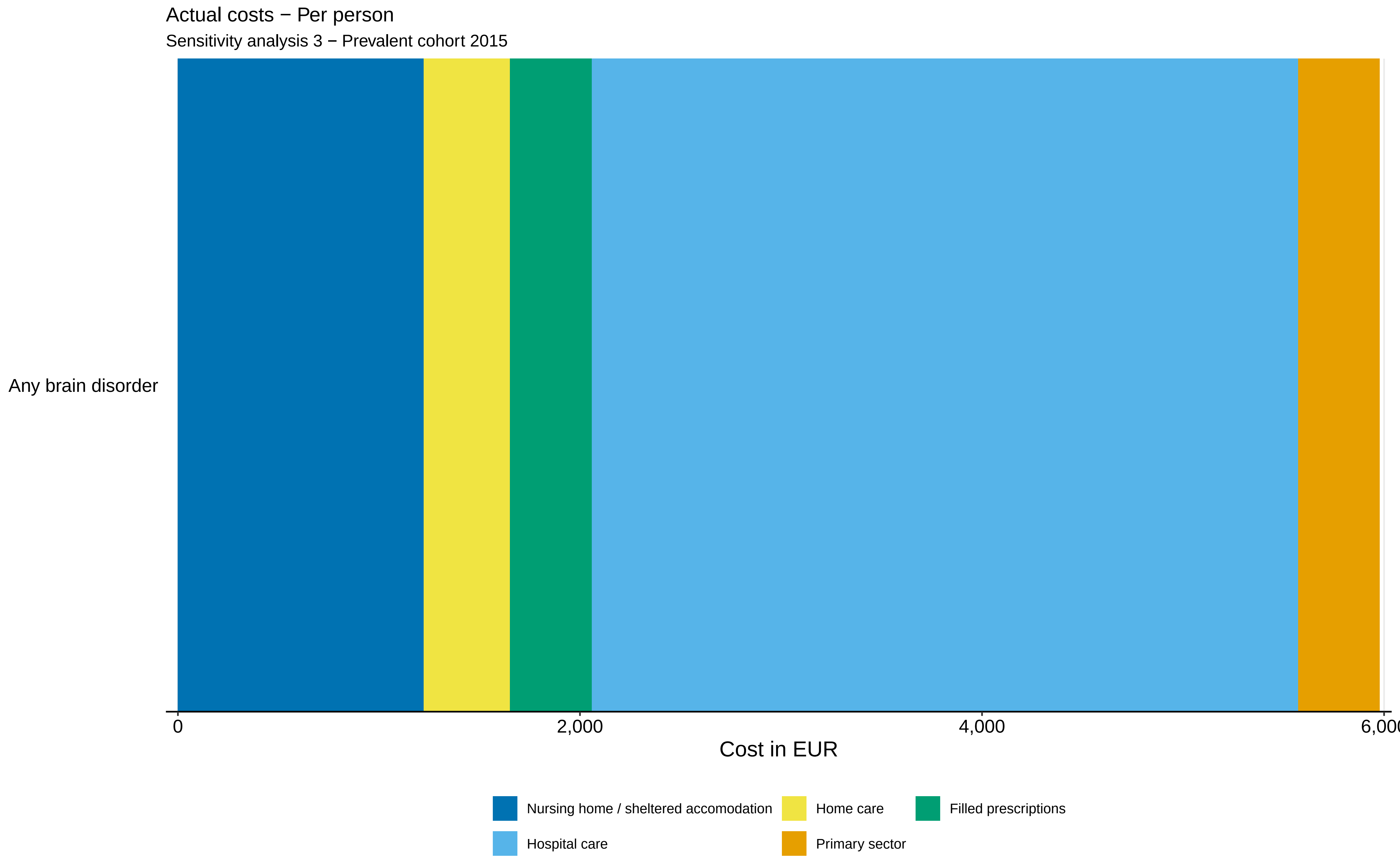

Actual costs − Per person
Sensitivity analysis 3 − Prevalent cohort 2015
Any brain disorder
0
2,000
4,000
6,000
Cost in EUR
Nursing home / sheltered accomodation
Home care
Filled prescriptions
Hospital care
Primary sector

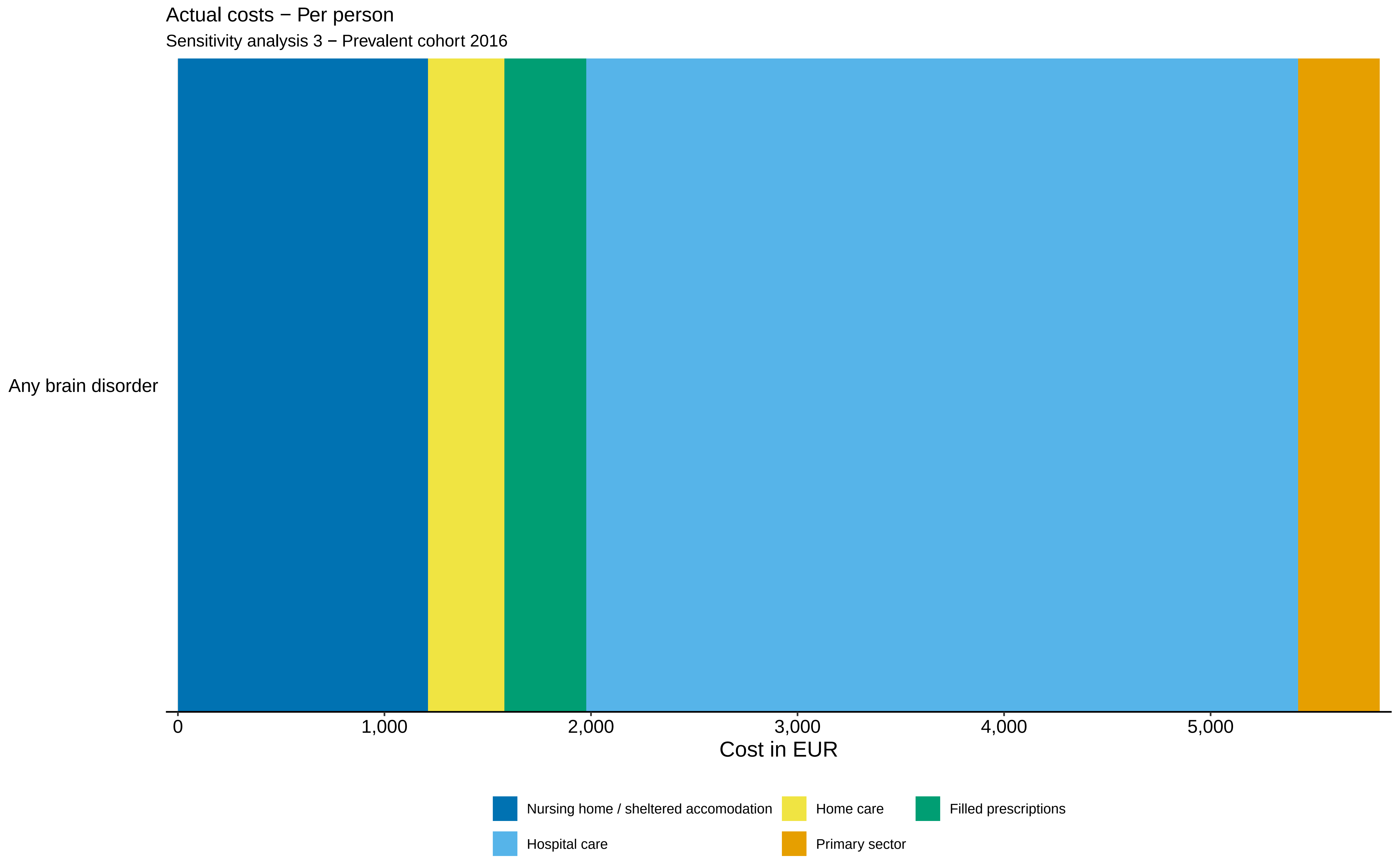
Actual costs − Per person
Sensitivity analysis 3 − Prevalent cohort 2016
Any brain disorder
0
1,000
2,000
3,000
4,000
5,000
Cost in EUR
Nursing home / sheltered accomodation
Home care
Filled prescriptions
Hospital care
Primary sector

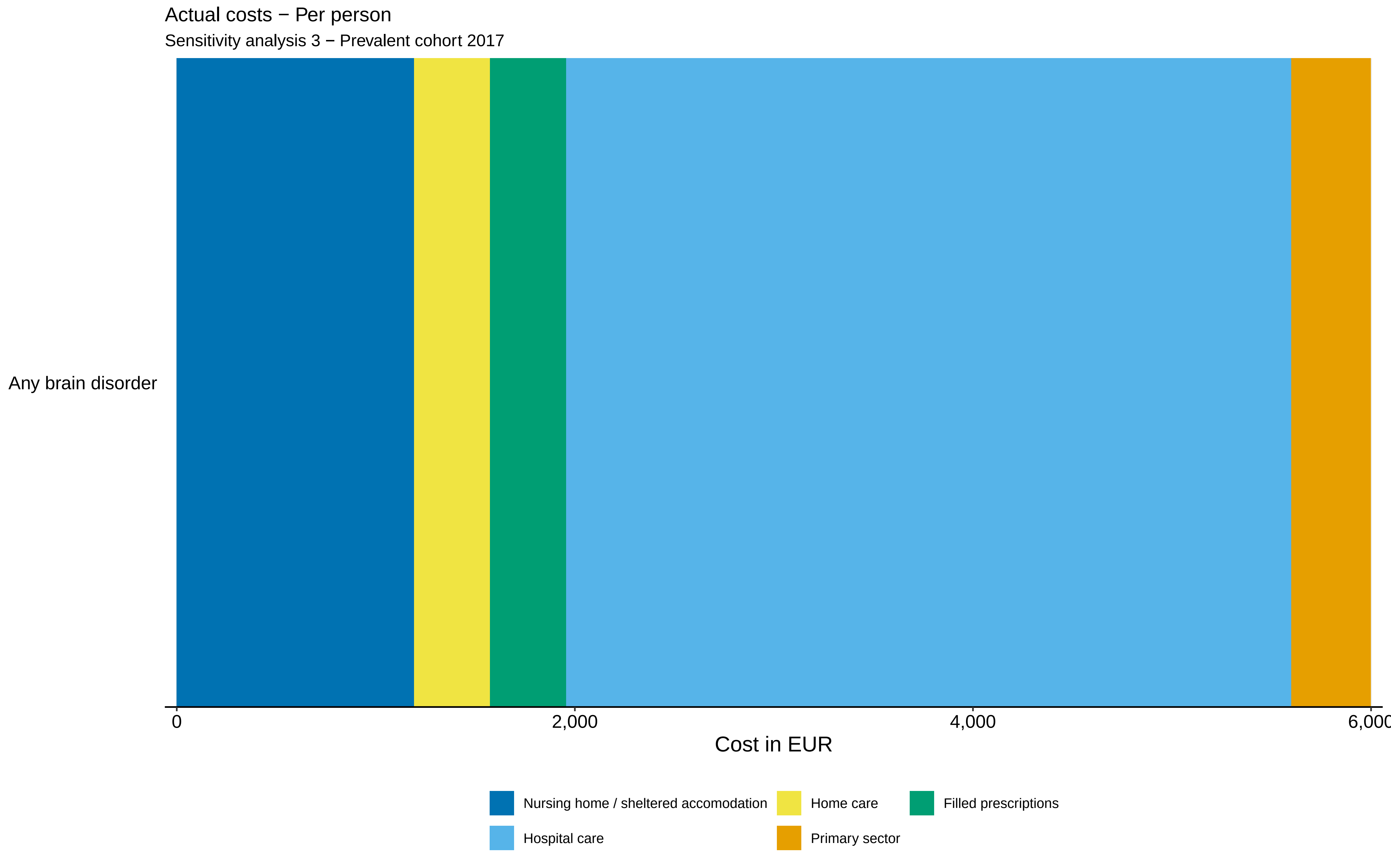

Actual costs − Per person
Sensitivity analysis 3 − Prevalent cohort 2017
Any brain disorder
0
2,000
4,000
6,000
Cost in EUR
Nursing home / sheltered accomodation
Home care
Filled prescriptions
Hospital care
Primary sector

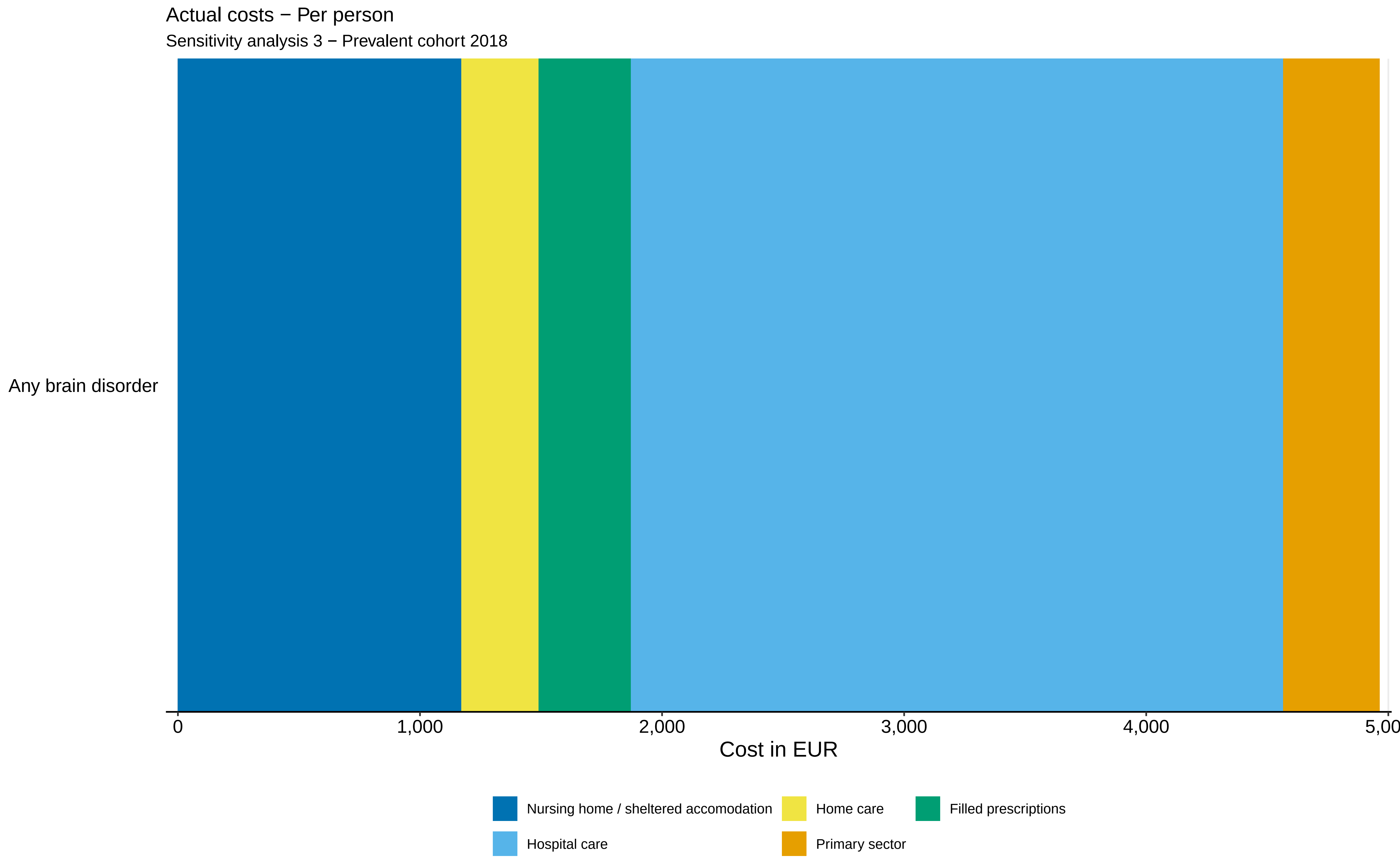
Actual costs − Per person
Sensitivity analysis 3 − Prevalent cohort 2018
Any brain disorder
0
1,000
2,000
3,000
4,000
5,00
Cost in EUR
Nursing home / sheltered accomodation
Home care
Filled prescriptions
Hospital care
Primary sector

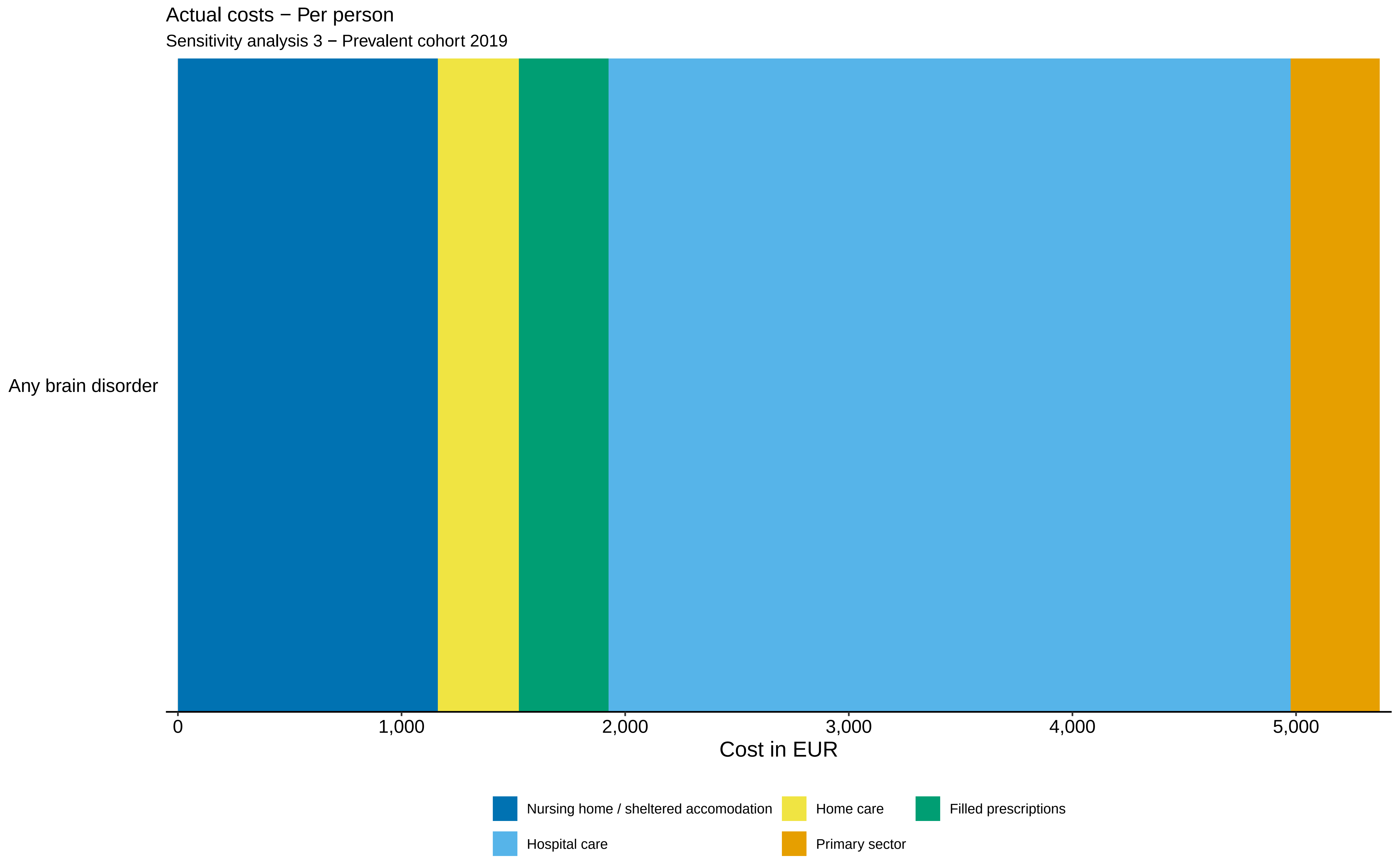

Actual costs − Per person
Sensitivity analysis 3 − Prevalent cohort 2019
Any brain disorder
0
1,000
2,000
3,000
4,000
5,000
Cost in EUR
Nursing home / sheltered accomodation
Home care
Filled prescriptions
Hospital care
Primary sector

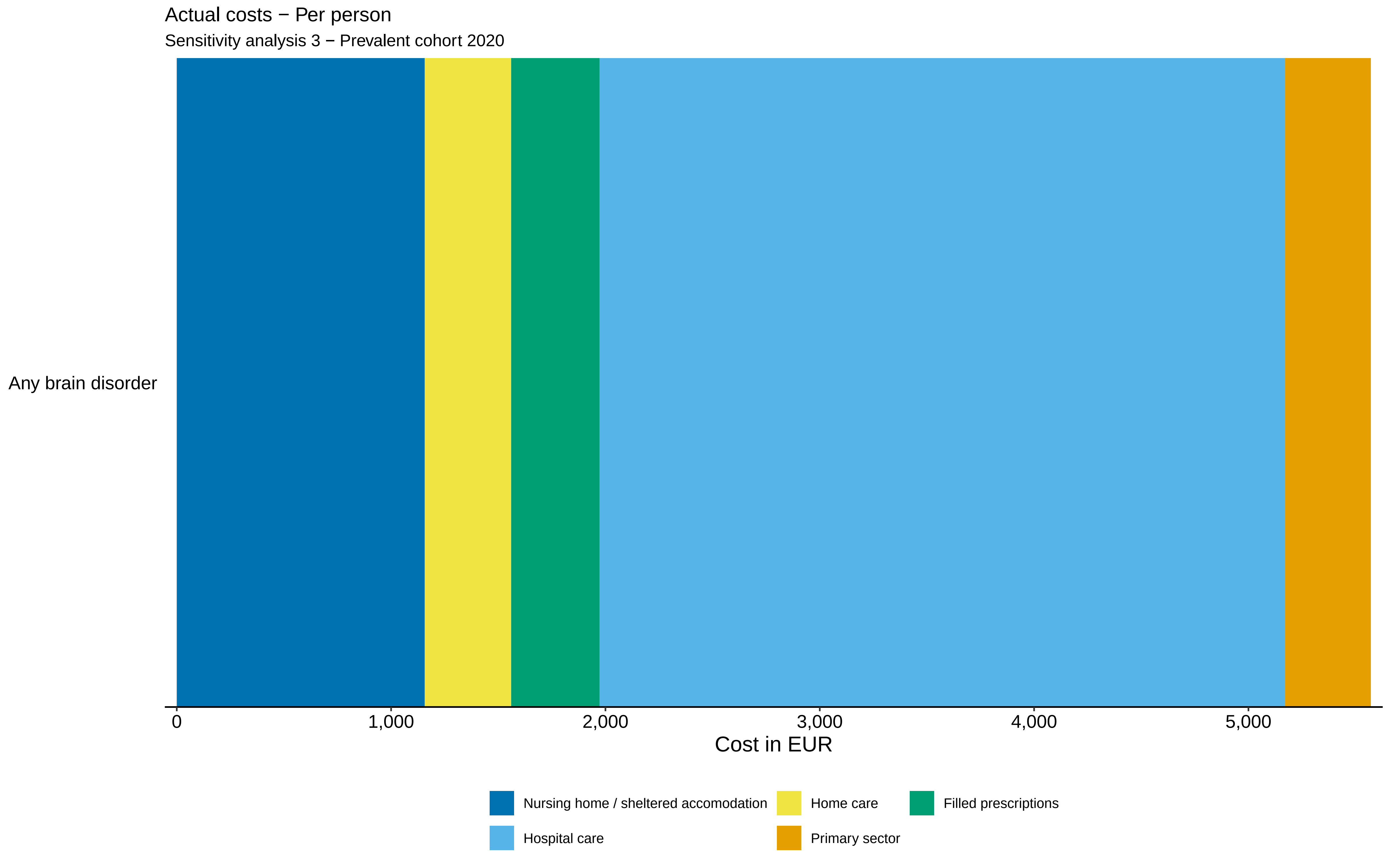
Actual costs − Per person
Sensitivity analysis 3 − Prevalent cohort 2020
Any brain disorder
0
1,000
2,000
3,000
4,000
5,000
Cost in EUR
Nursing home / sheltered accomodation
Home care
Filled prescriptions
Hospital care
Primary sector

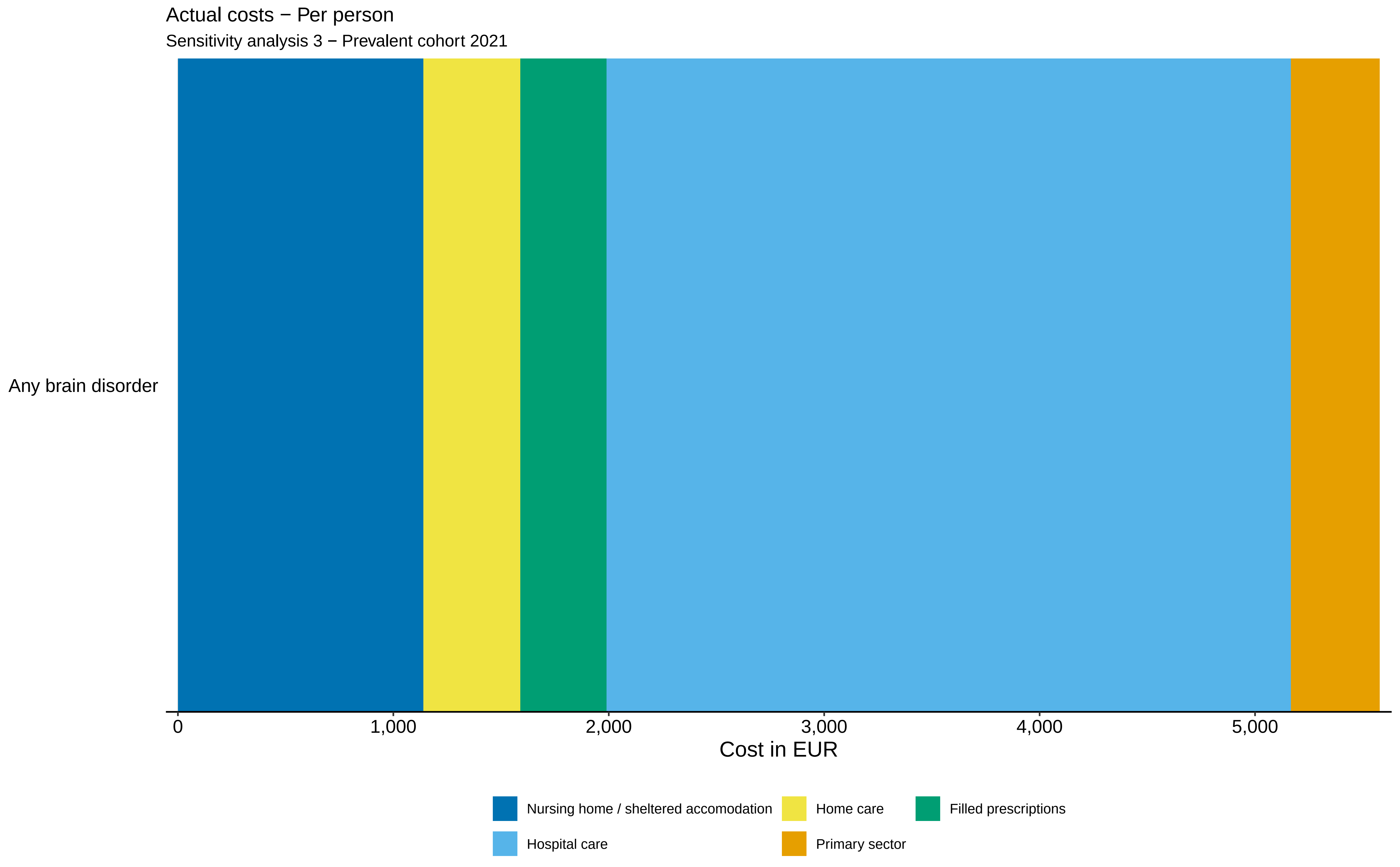

Actual costs − Per person
Sensitivity analysis 3 − Prevalent cohort 2021
Any brain disorder
0
1,000
2,000
3,000
4,000
5,000
Cost in EUR
Nursing home / sheltered accomodation
Home care
Filled prescriptions
Hospital care
Primary sector

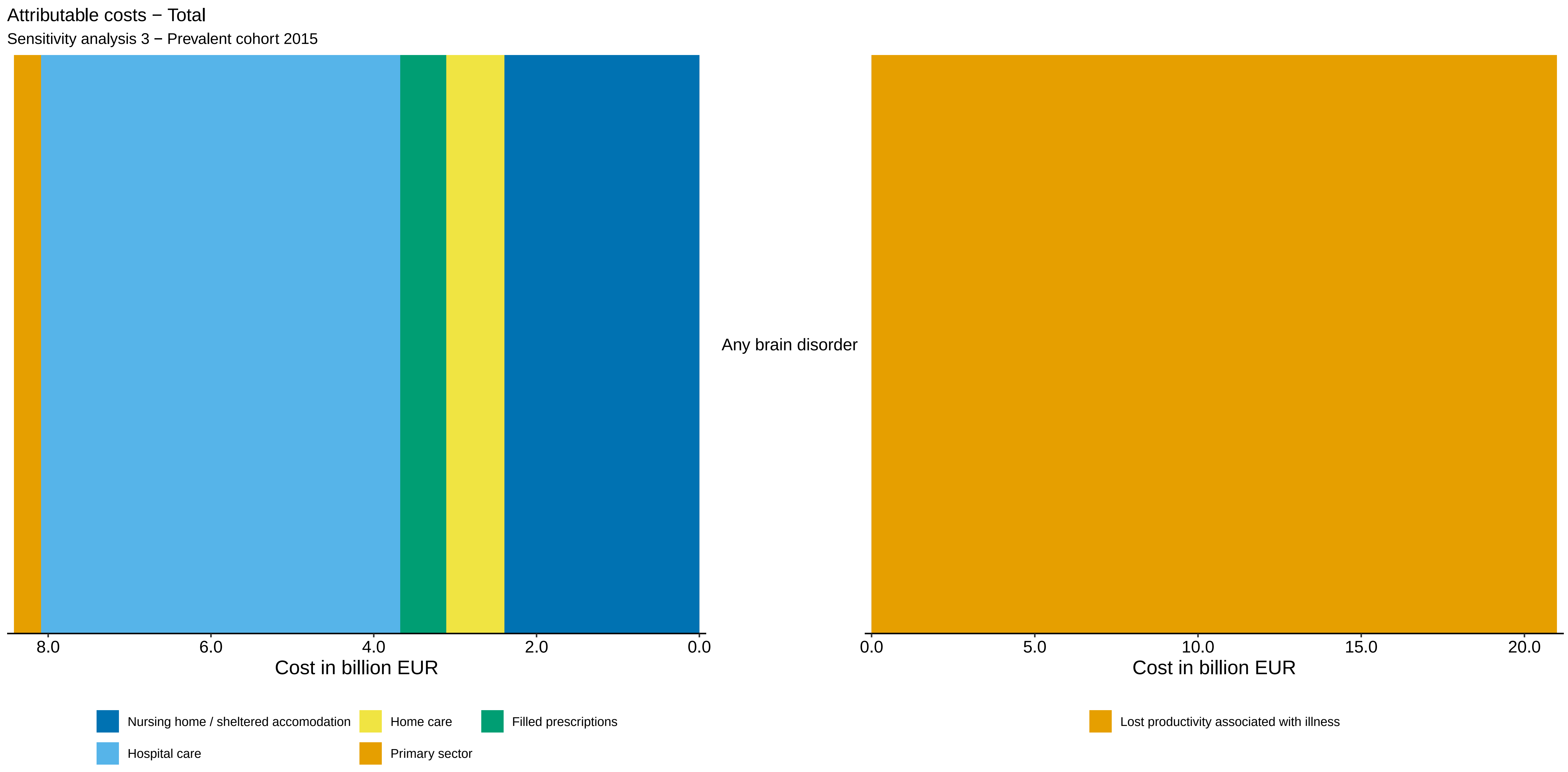

Attributable costs − Total
Sensitivity analysis 3 − Prevalent cohort 2015
Any brain disorder
8.0
6.0
4.0
2.0
0.0
Cost in billion EUR
0.0
5.0
10.0
15.0
20.0
Cost in billion EUR
Nursing home / sheltered accomodation
Home care
Filled prescriptions
Hospital care
Primary sector
Lost productivity associated with illness

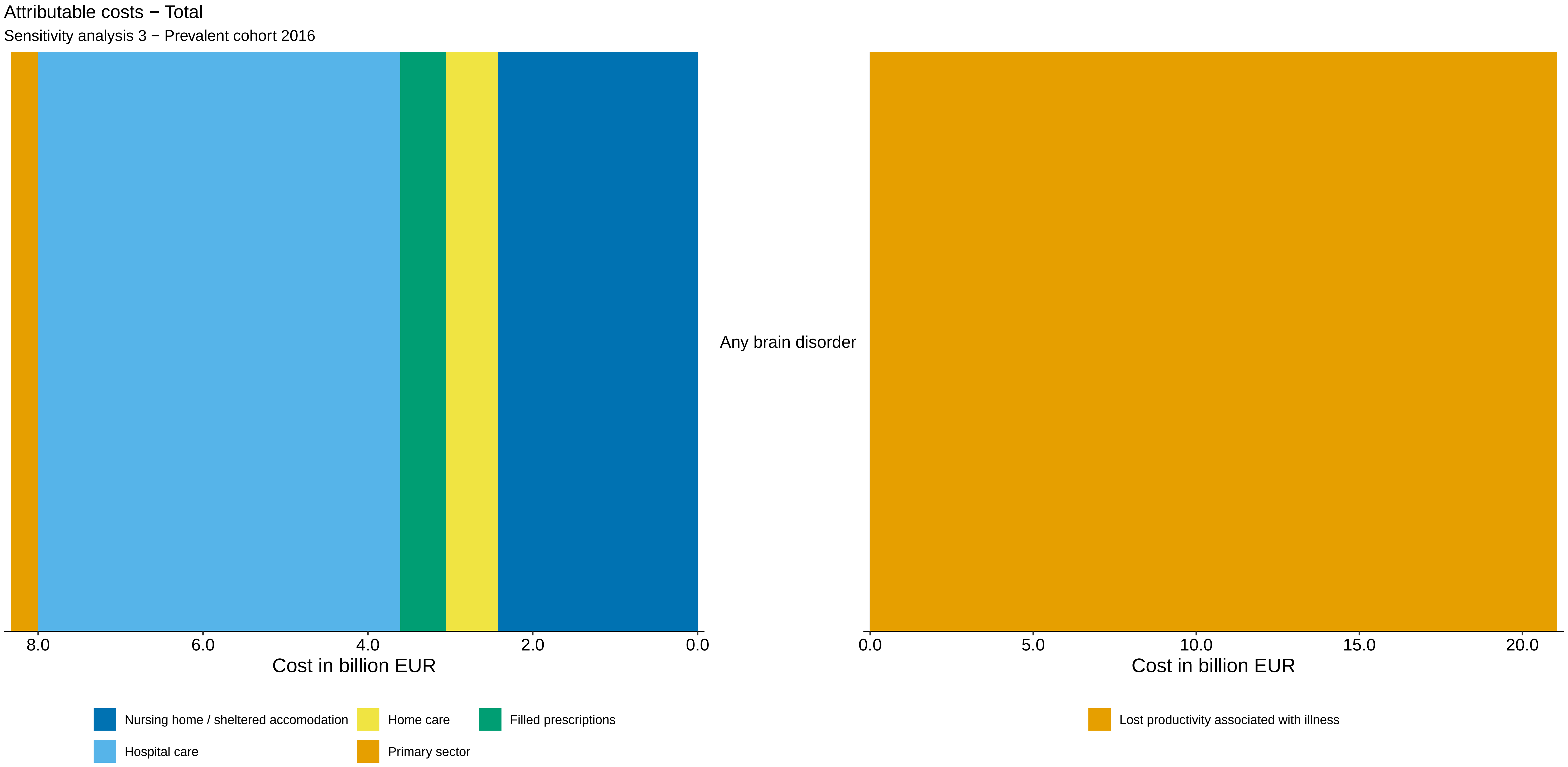

Attributable costs − Total
Sensitivity analysis 3 − Prevalent cohort 2016
Any brain disorder
8.0
6.0
4.0
2.0
0.0
Cost in billion EUR
0.0
5.0
10.0
15.0
20.0
Cost in billion EUR
Nursing home / sheltered accomodation
Home care
Filled prescriptions
Hospital care
Primary sector
Lost productivity associated with illness

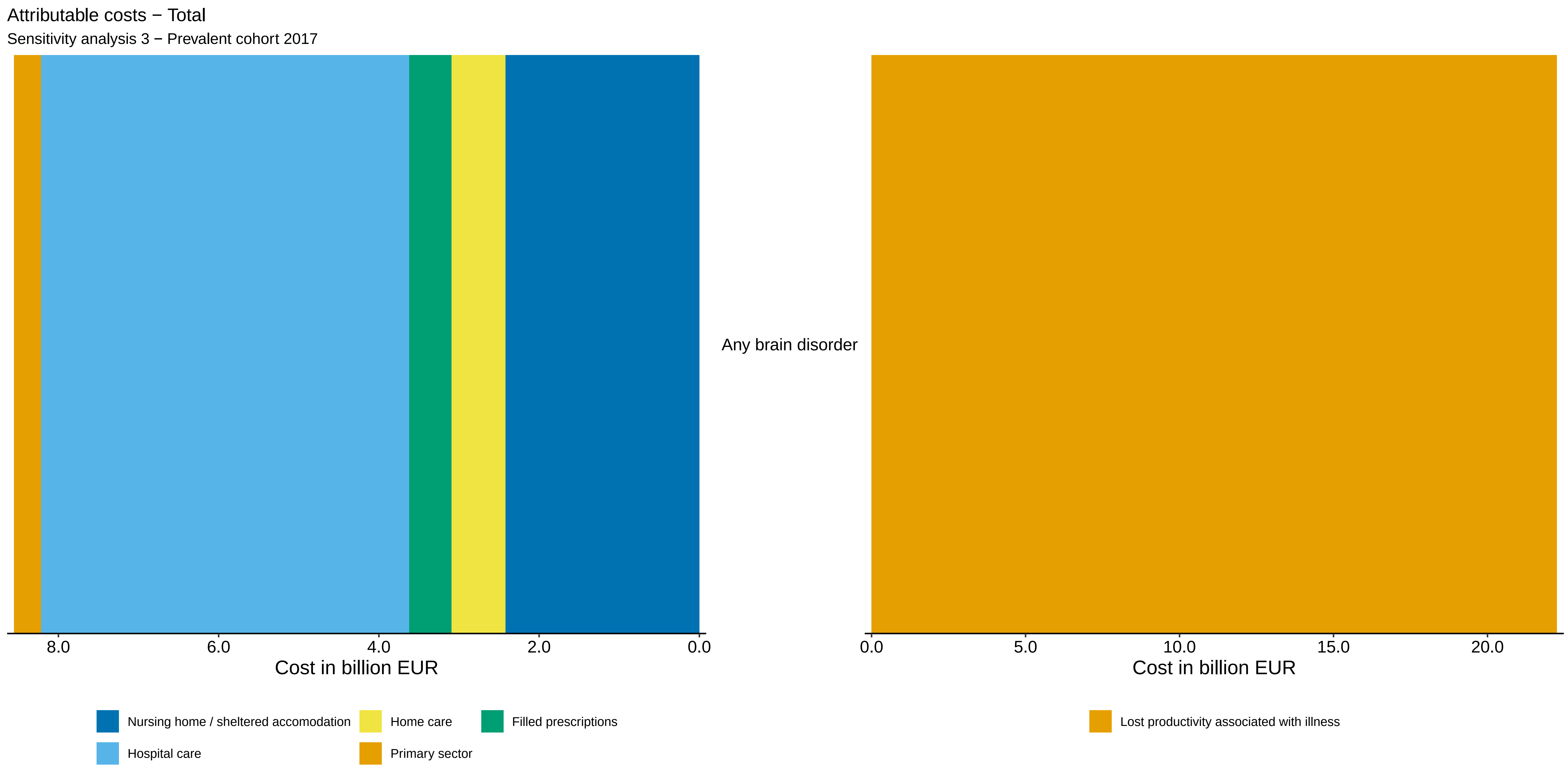
Attributable costs − Total
Sensitivity analysis 3 − Prevalent cohort 2017
Any brain disorder
8.0
6.0
4.0
2.0
0.0
Cost in billion EUR
0.0
5.0
10.0
15.0
20.0
Cost in billion EUR
Nursing home / sheltered accomodation
Home care
Filled prescriptions
Hospital care
Primary sector
Lost productivity associated with illness

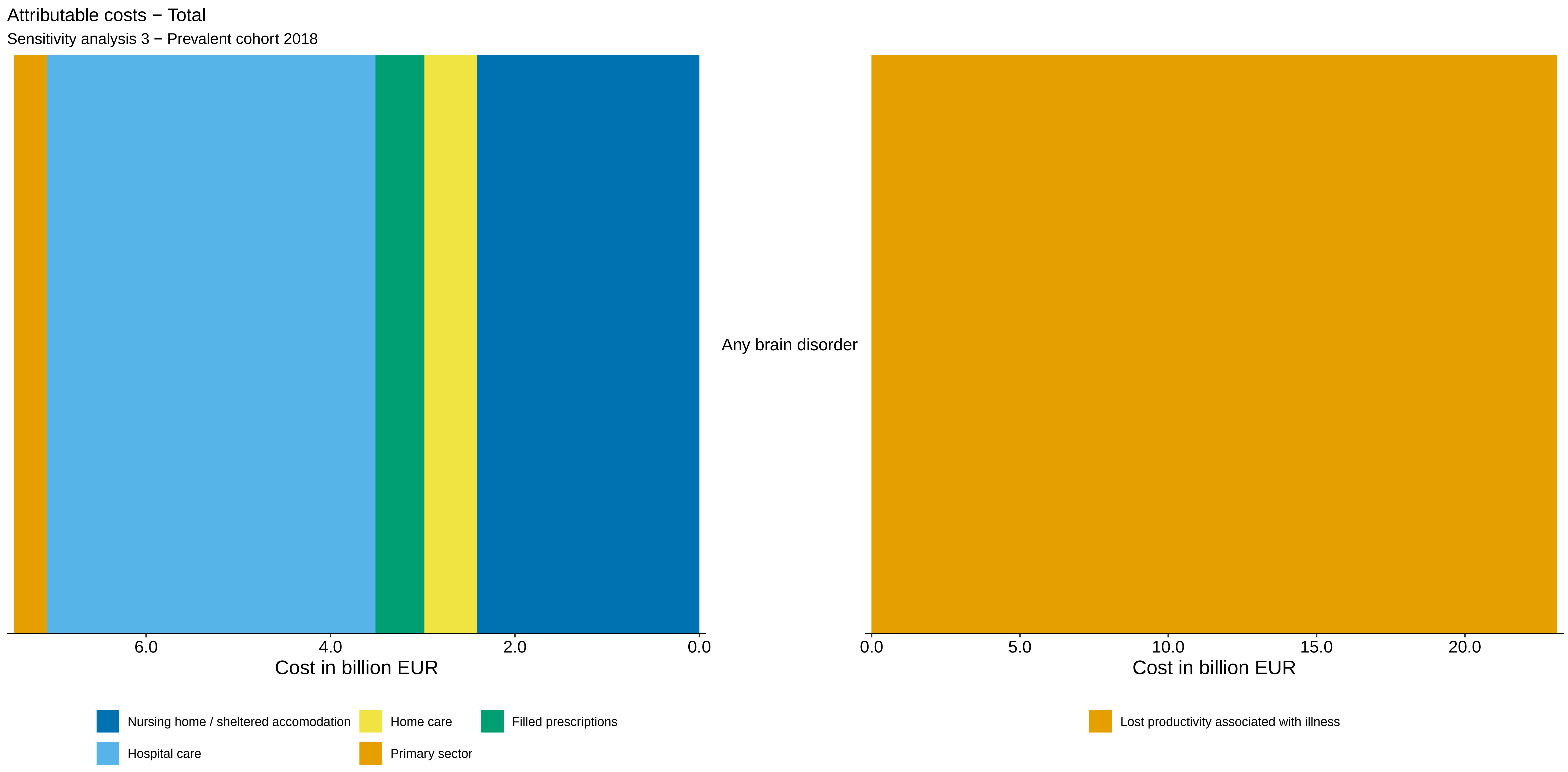
Attributable costs − Total
Sensitivity analysis 3 − Prevalent cohort 2018
Any brain disorder
6.0
4.0
2.0
0.0
Cost in billion EUR
0.0
5.0
10.0
15.0
20.0
Cost in billion EUR
Nursing home / sheltered accomodation
Home care
Filled prescriptions
Hospital care
Primary sector
Lost productivity associated with illness

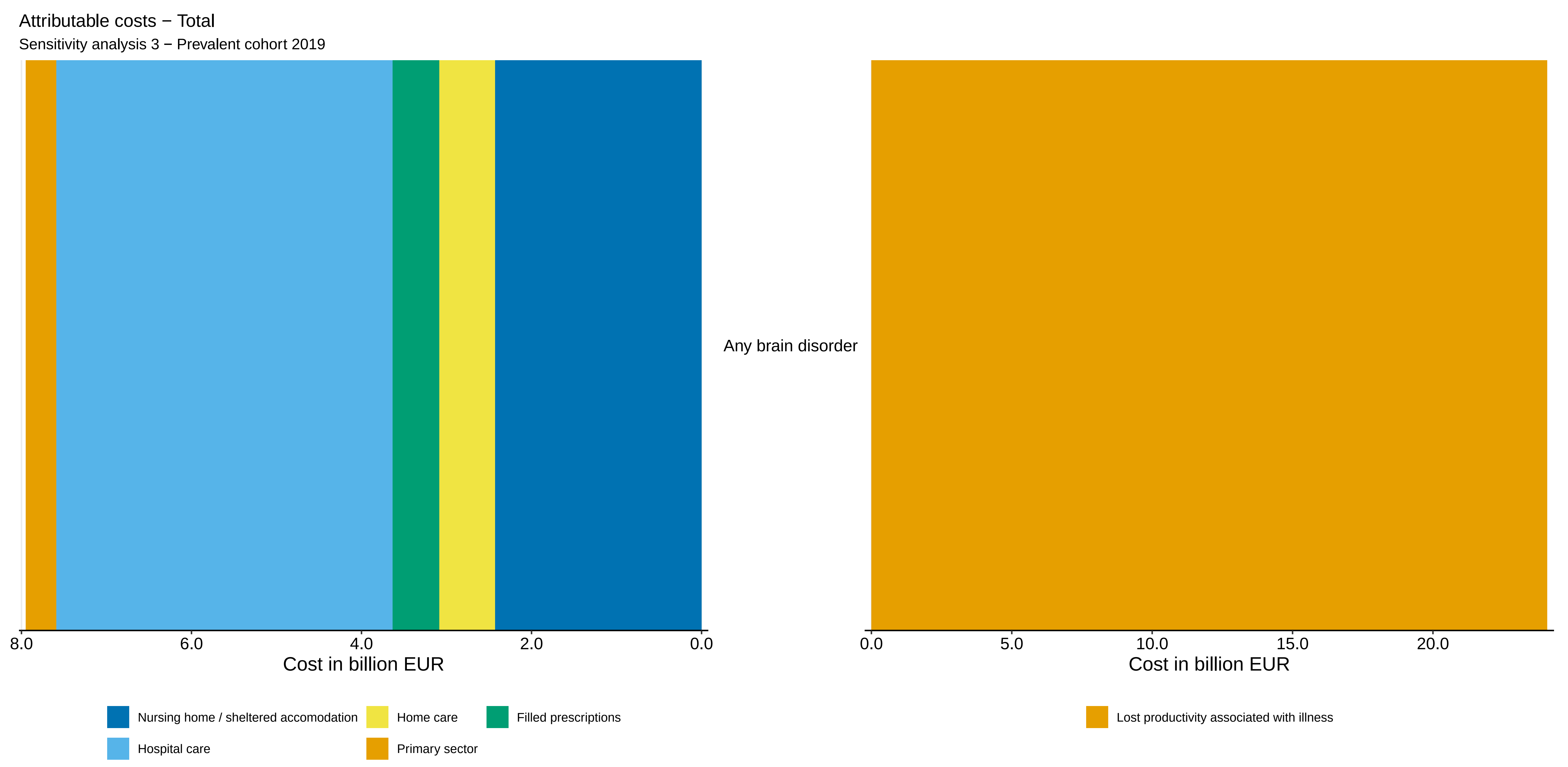

Attributable costs − Total
Sensitivity analysis 3 − Prevalent cohort 2019
Any brain disorder
8.0
6.0
4.0
2.0
0.0
Cost in billion EUR
0.0
5.0
10.0
15.0
20.0
Cost in billion EUR
Nursing home / sheltered accomodation
Home care
Filled prescriptions
Hospital care
Primary sector
Lost productivity associated with illness

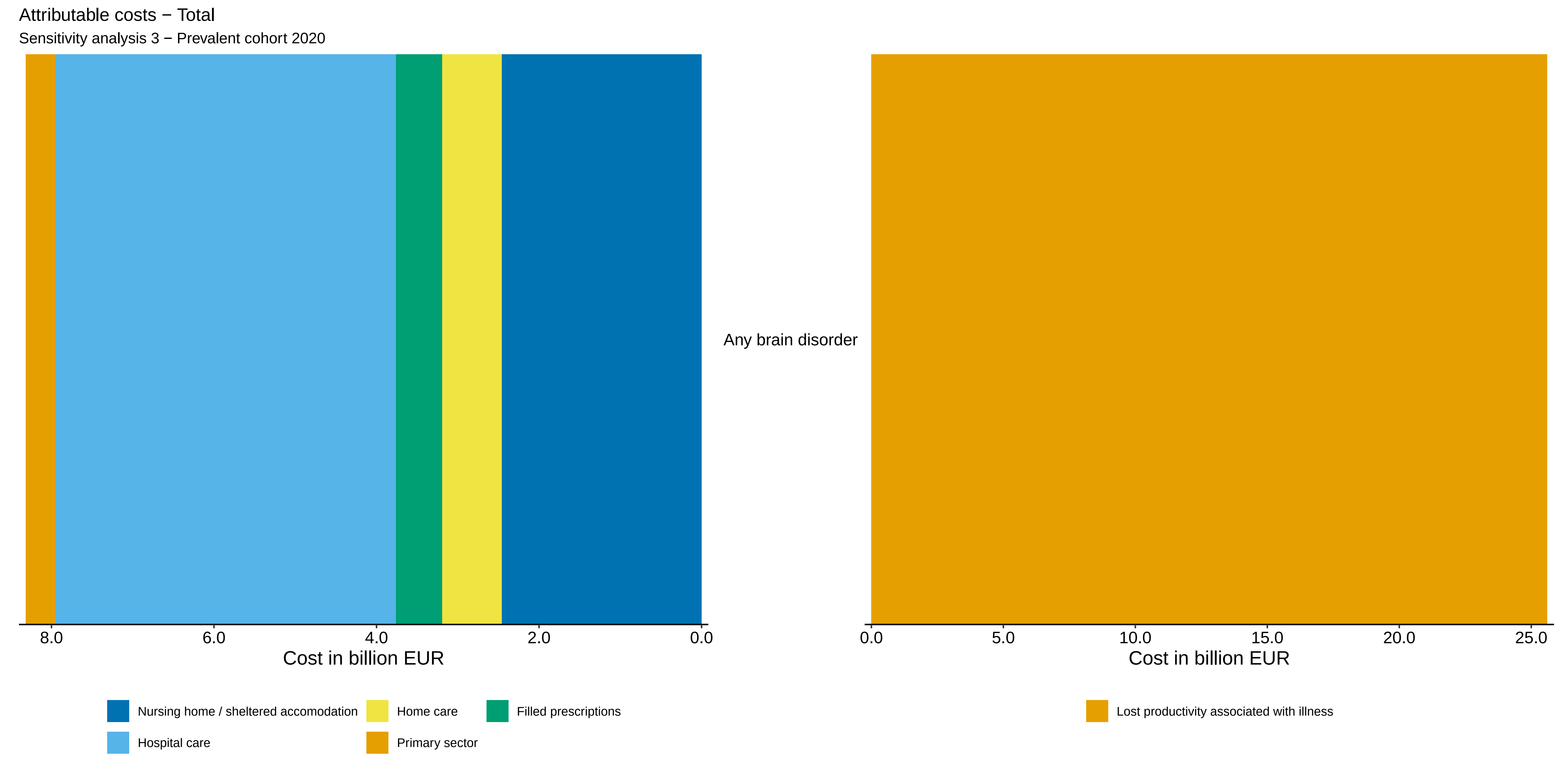

Attributable costs − Total
Sensitivity analysis 3 − Prevalent cohort 2020
Any brain disorder
8.0
6.0
4.0
2.0
0.0
Cost in billion EUR
0.0
5.0
10.0
15.0
20.0
25.0
Cost in billion EUR
Nursing home / sheltered accomodation
Home care
Filled prescriptions
Hospital care
Primary sector
Lost productivity associated with illness

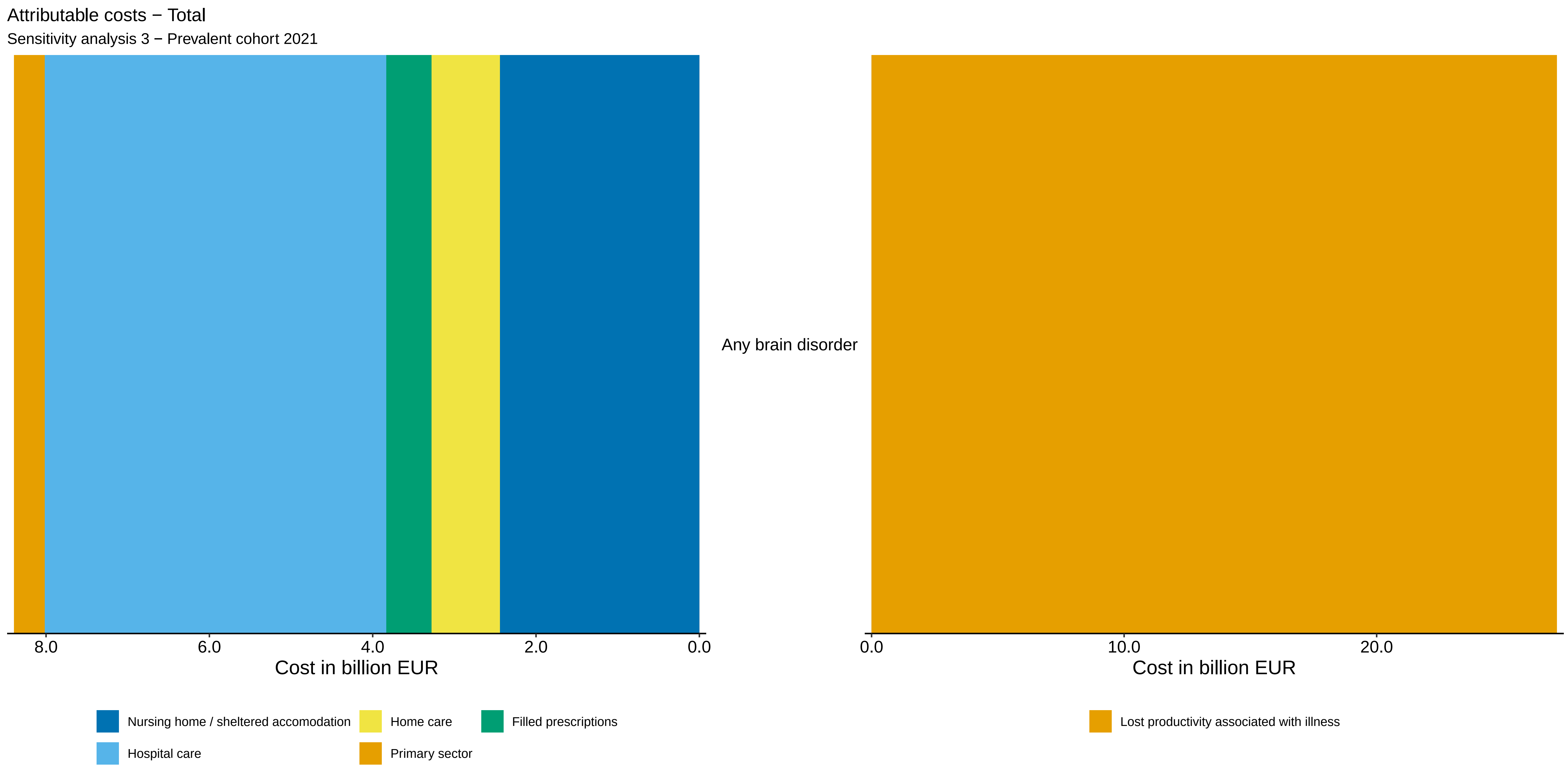
Attributable costs − Total
Sensitivity analysis 3 − Prevalent cohort 2021
Any brain disorder
8.0
6.0
4.0
2.0
0.0
Cost in billion EUR
0.0
10.0
20.0
Cost in billion EUR
Nursing home / sheltered accomodation
Home care
Filled prescriptions
Hospital care
Primary sector
Lost productivity associated with illness

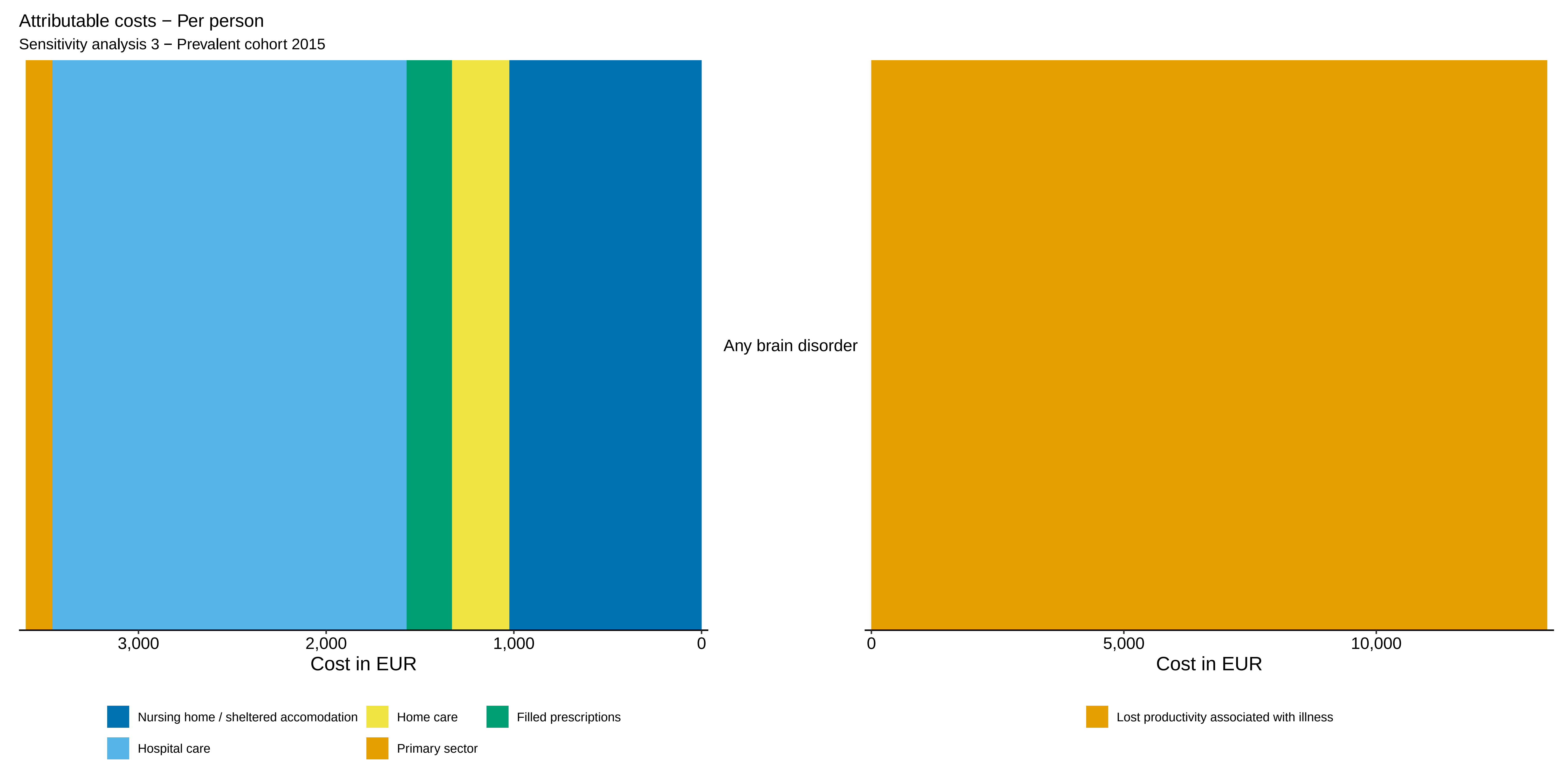

Attributable costs − Per person
Sensitivity analysis 3 − Prevalent cohort 2015
Any brain disorder
3,000
2,000
1,000
0
Cost in EUR
0
5,000
10,000
Cost in EUR
Nursing home / sheltered accomodation
Home care
Filled prescriptions
Hospital care
Primary sector
Lost productivity associated with illness

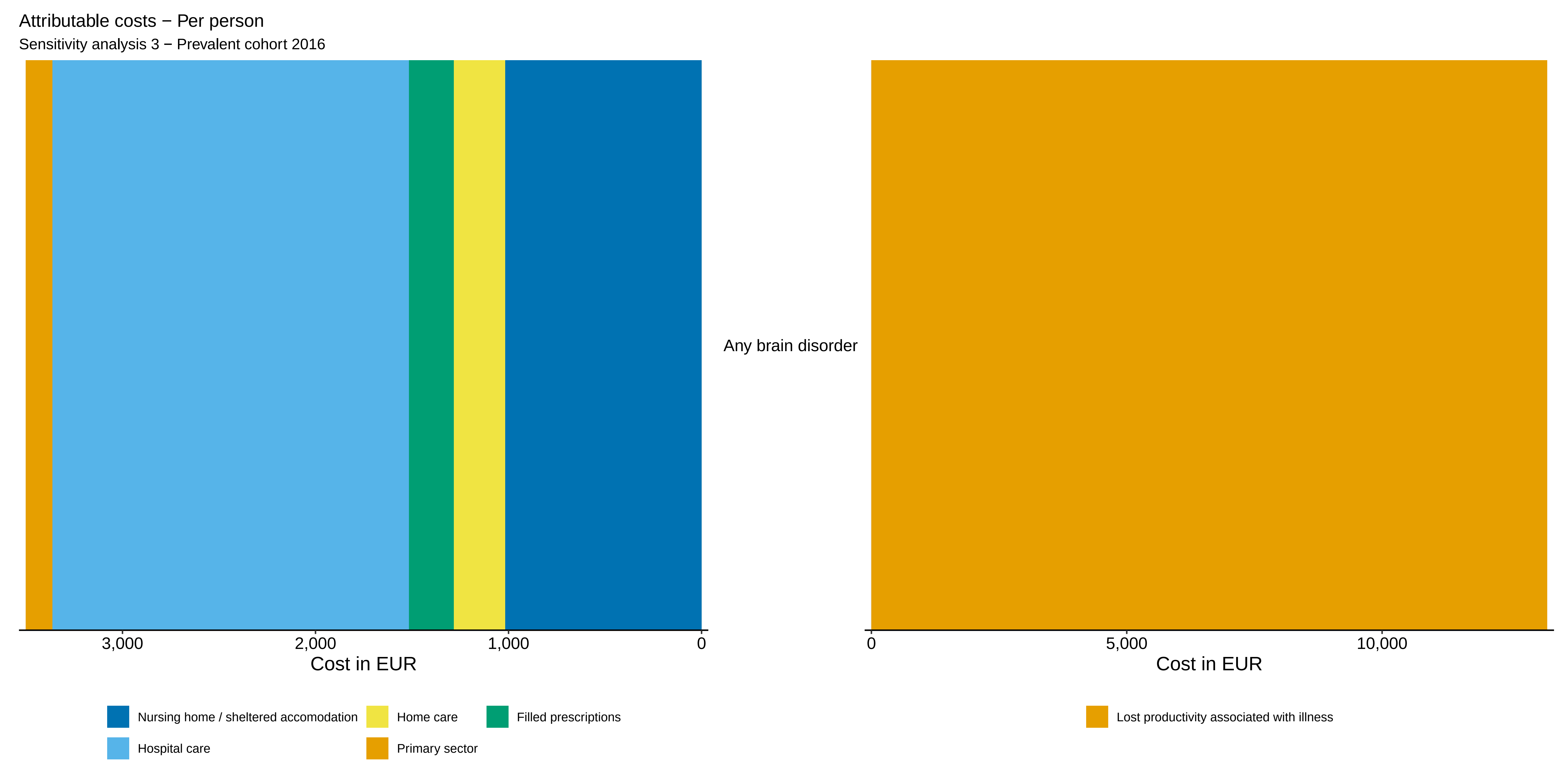

Attributable costs − Per person
Sensitivity analysis 3 − Prevalent cohort 2016
Any brain disorder
3,000
2,000
1,000
0
Cost in EUR
0
5,000
10,000
Cost in EUR
Nursing home / sheltered accomodation
Home care
Filled prescriptions
Hospital care
Primary sector
Lost productivity associated with illness

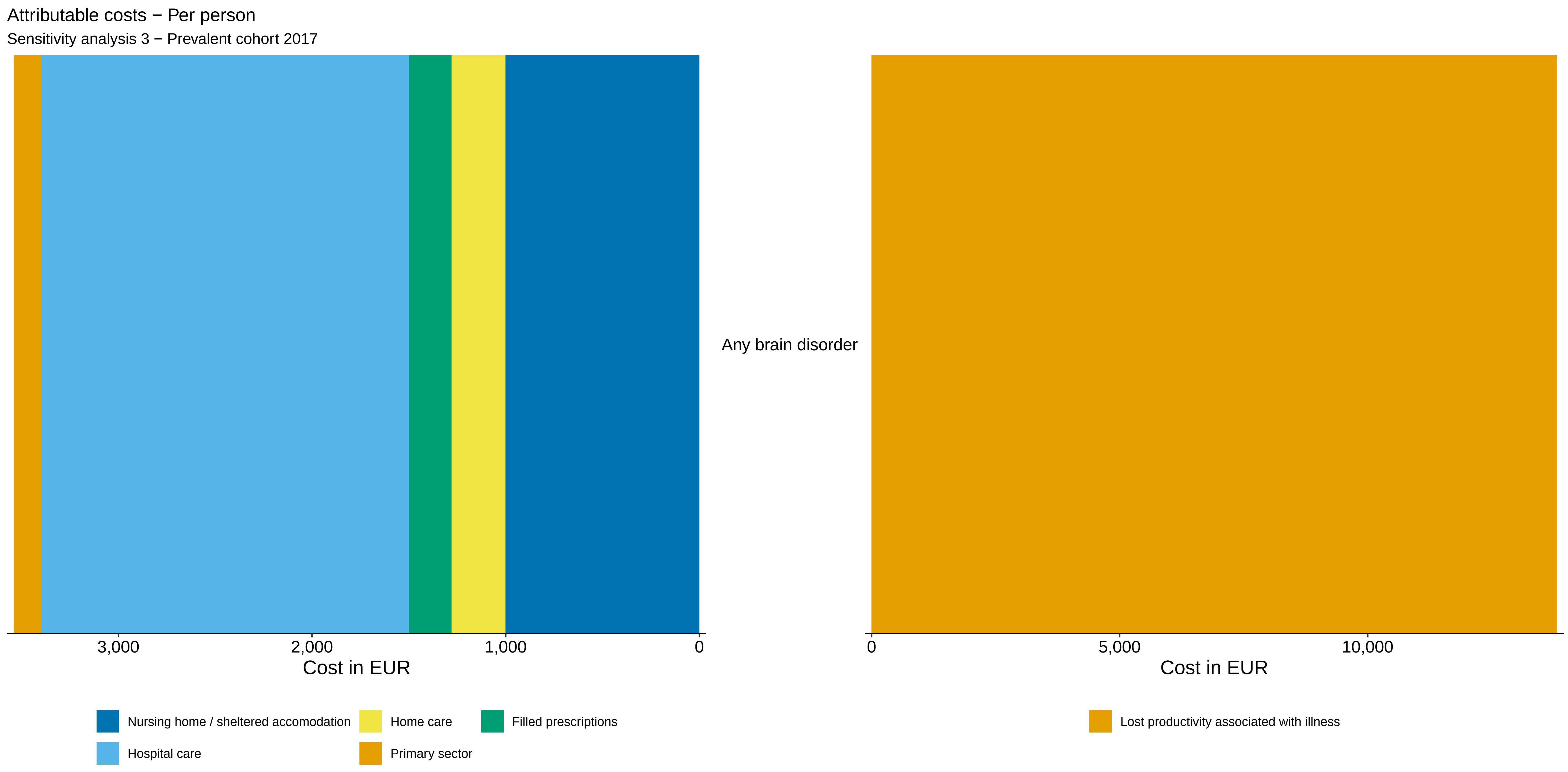

Attributable costs − Per person
Sensitivity analysis 3 − Prevalent cohort 2017
Any brain disorder
3,000
2,000
1,000
0
Cost in EUR
0
5,000
10,000
Cost in EUR
Nursing home / sheltered accomodation
Home care
Filled prescriptions
Hospital care
Primary sector
Lost productivity associated with illness

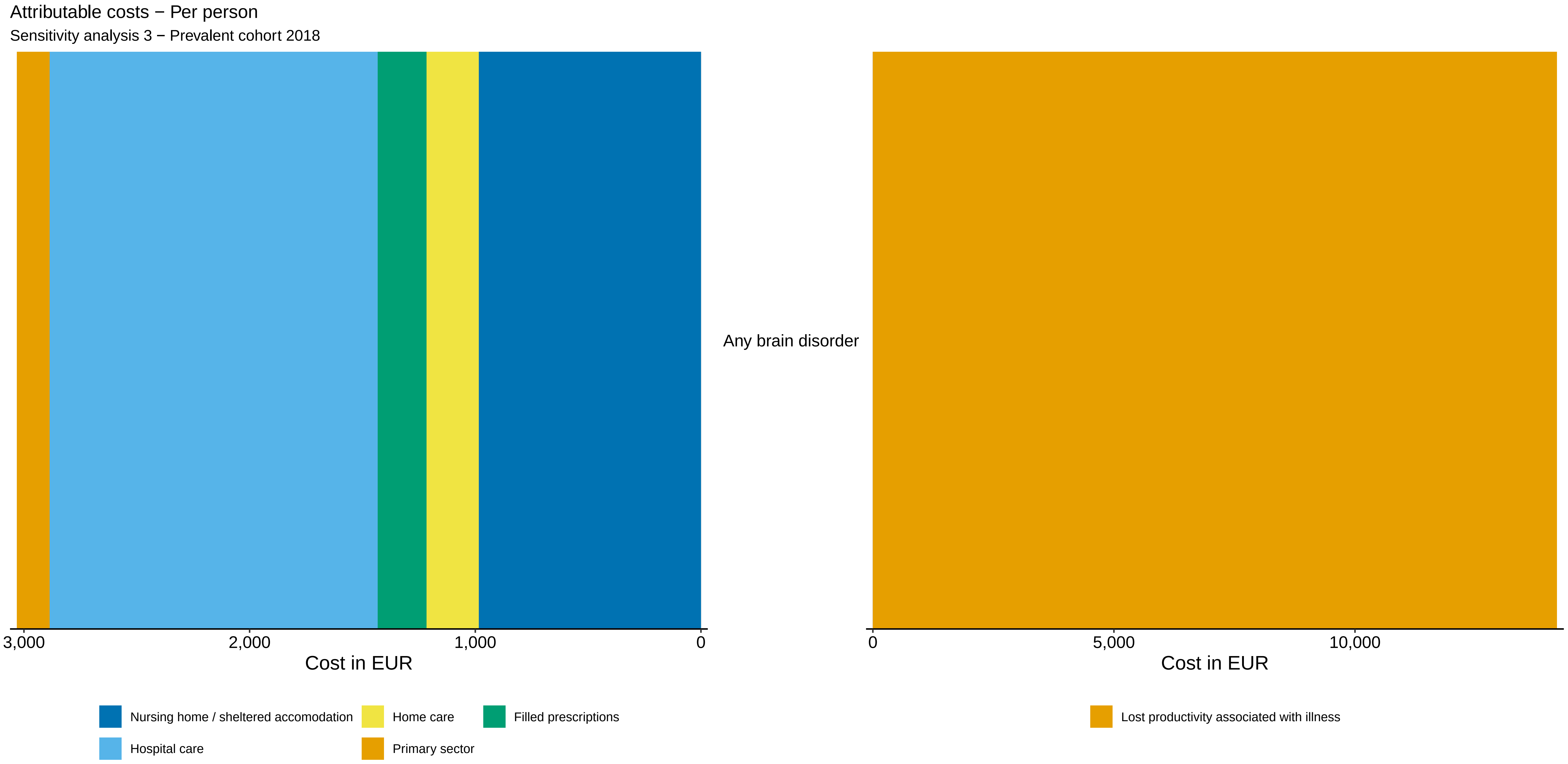
Attributable costs − Per person
Sensitivity analysis 3 − Prevalent cohort 2018
Any brain disorder
3,000
2,000
1,000
0
Cost in EUR
0
5,000
10,000
Cost in EUR
Nursing home / sheltered accomodation
Home care
Filled prescriptions
Hospital care
Primary sector
Lost productivity associated with illness

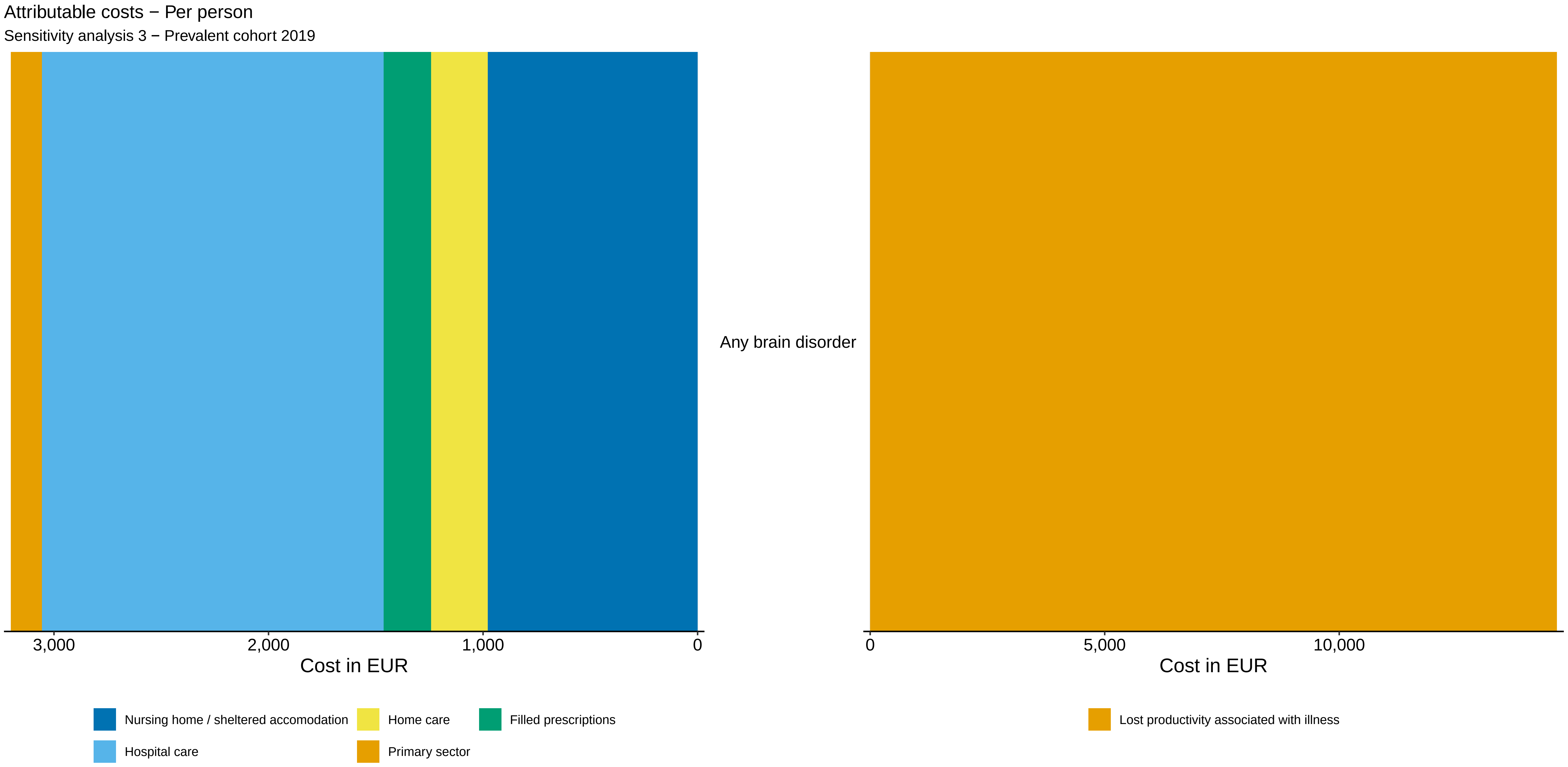

Attributable costs − Per person
Sensitivity analysis 3 − Prevalent cohort 2019
Any brain disorder
3,000
2,000
1,000
0
Cost in EUR
0
5,000
10,000
Cost in EUR
Nursing home / sheltered accomodation
Home care
Filled prescriptions
Hospital care
Primary sector
Lost productivity associated with illness

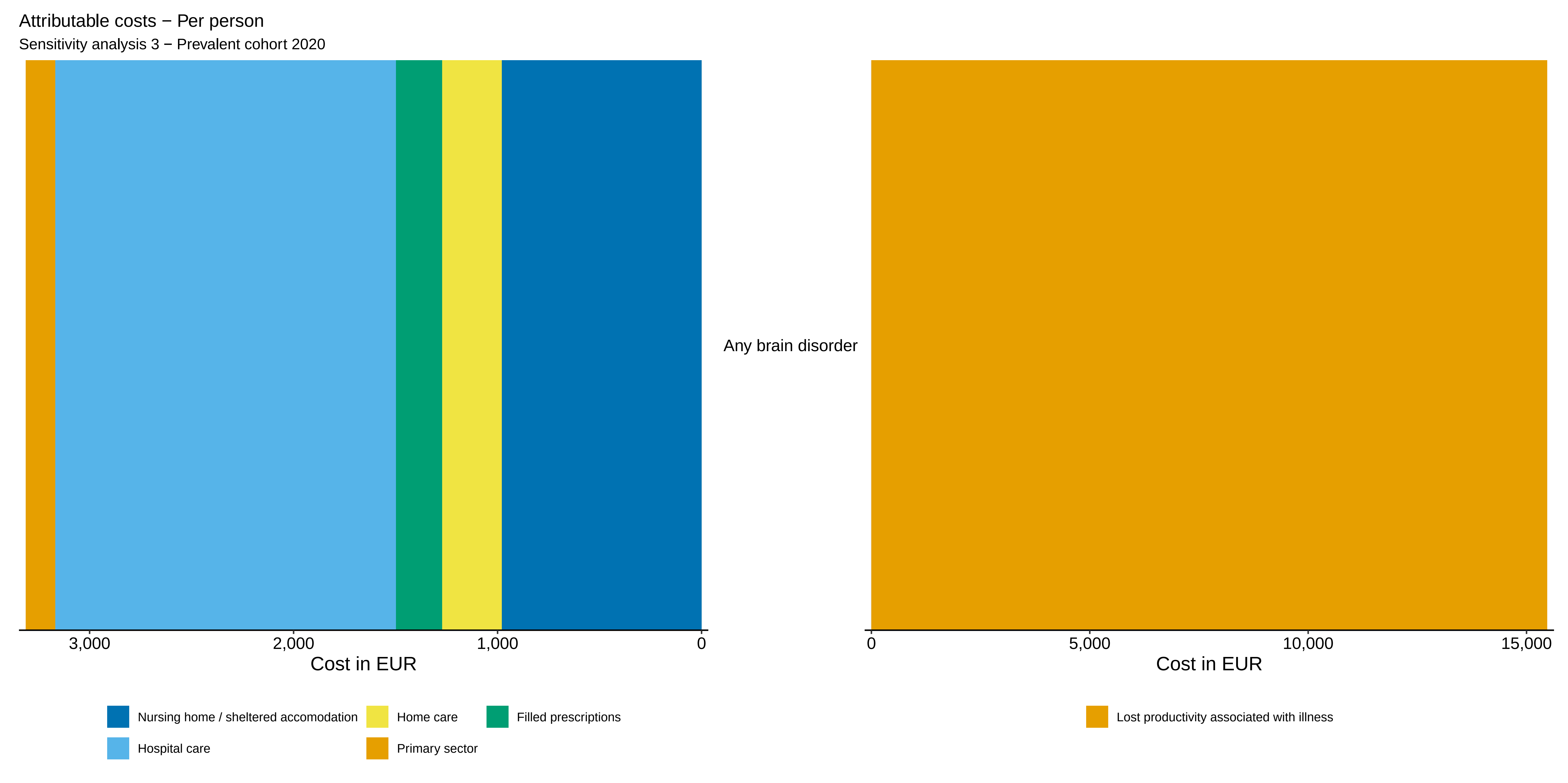

Attributable costs − Per person
Sensitivity analysis 3 − Prevalent cohort 2020
Any brain disorder
3,000
2,000
1,000
0
Cost in EUR
0
5,000
10,000
15,000
Cost in EUR
Nursing home / sheltered accomodation
Home care
Filled prescriptions
Hospital care
Primary sector
Lost productivity associated with illness

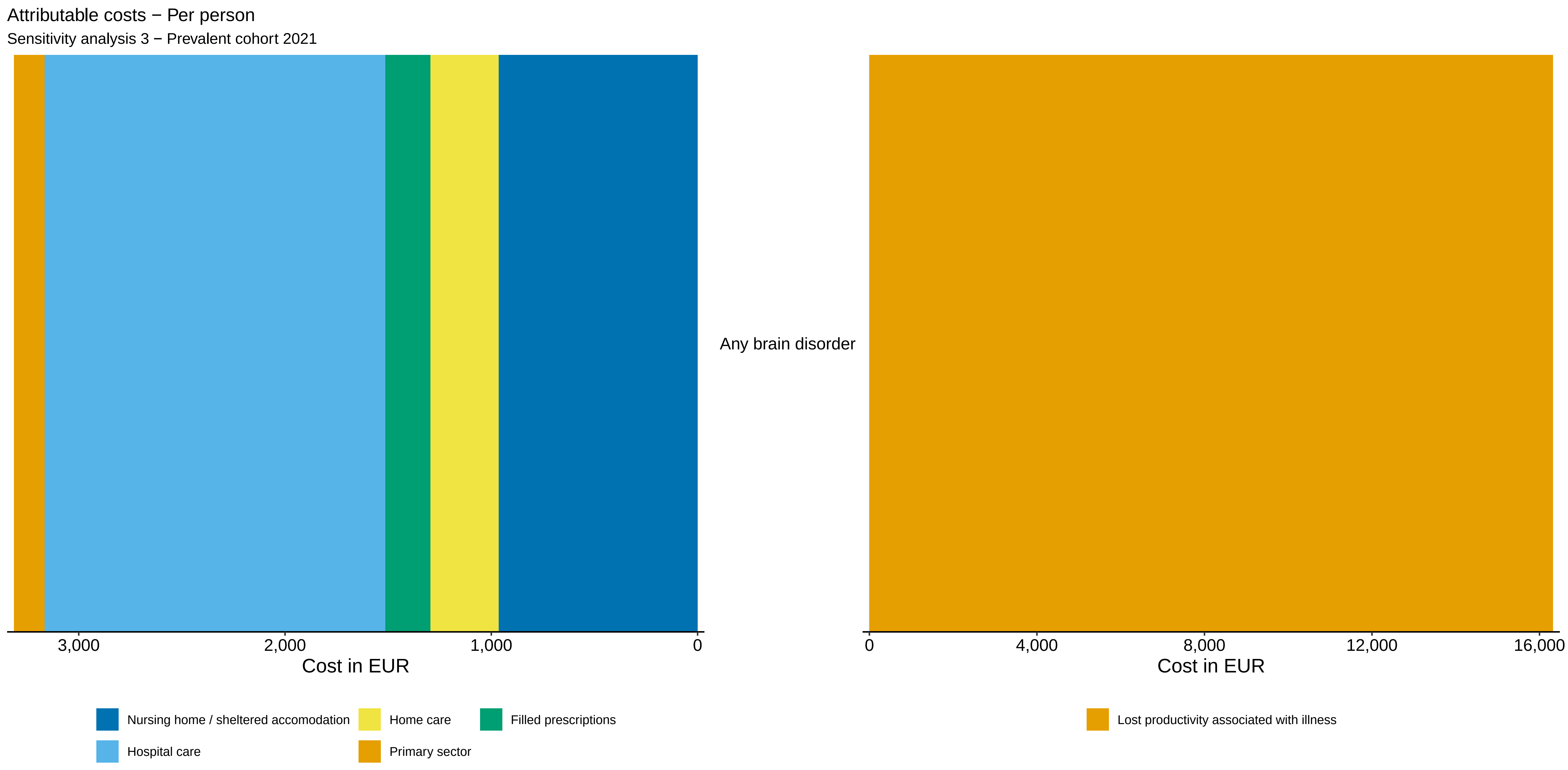

Attributable costs − Per person
Sensitivity analysis 3 − Prevalent cohort 2021
Any brain disorder
3,000
2,000
1,000
0
Cost in EUR
0
4,000
8,000
12,000
16,000
Cost in EUR
Nursing home / sheltered accomodation
Home care
Filled prescriptions
Hospital care
Primary sector
Lost productivity associated with illness

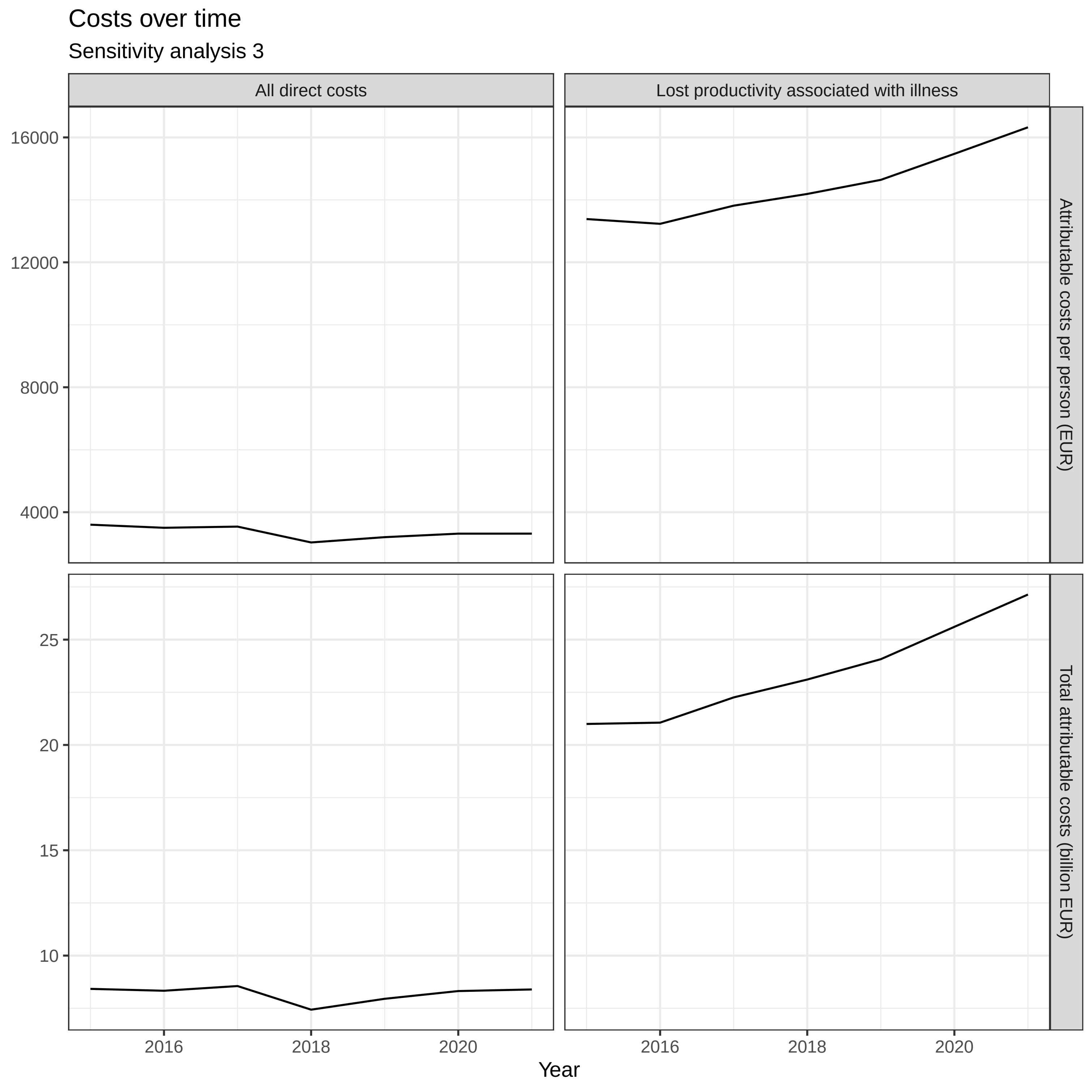
Costs over time
Sensitivity analysis 3
All direct costs
Lost productivity associated with illness
Attributable costs per person (EUR)
Total attributable costs (billion EUR)
16000
12000
8000
4000
25
20
15
10
2016
2018
2020
Year

***Sensitivity analysis 3 / Prevalent cohort 2015***

| | *CCI covariates included in model* | *No CCI covariates included in model* |
|---|---|---|
| Age 10-19 | -0.00 (-0.01 - 0.00) | -0.02 (-0.02 - -0.01) |
| Age 20-29 | 0.02 (0.02 - 0.03) | -0.00 (-0.01 - 0.00) |
| Age 30-39 | 0.03 (0.02 - 0.03) | 0.01 (0.01 - 0.02) |
| Age 40-49 | 0.02 (0.01 - 0.02) | 0.02 (0.02 - 0.02) |
| Age 50-59 | 0.05 (0.05 - 0.06) | 0.10 (0.10 - 0.11) |
| Age 60-69 | 0.14 (0.14 - 0.15) | 0.27 (0.27 - 0.27) |
| Age 70-79 | 0.33 (0.33 - 0.34) | 0.56 (0.56 - 0.57) |
| Age 80-89 | 0.97 (0.97 - 0.98) | 1.30 (1.30 - 1.31) |
| Age 90+ | 2.37 (2.36 - 2.38) | 2.69 (2.68 - 2.71) |
| Myocardial infarction | 0.06 (0.05 - 0.07) | |
| Diabetes without end-organ damage | 0.33 (0.32 - 0.34) | |
| Hemiplegia | 1.39 (1.37 - 1.42) | |
| Moderate to severe renal disease | 0.80 (0.79 - 0.81) | |
| Diabetes with end-organ damage | 0.46 (0.45 - 0.47) | |
| Solid cancer tumour | 0.35 (0.34 - 0.35) | |
| Leukaemia | 1.06 (1.03 - 1.09) | |
| Lymphoma | 0.96 (0.94 - 0.98) | |
| Moderate to severe liver disease | 0.74 (0.71 - 0.77) | |
| Metastatic solid tumour | 1.03 (1.01 - 1.05) | |
| AIDS | 0.29 (0.26 - 0.32) | |
| Congestive heart failure | 0.52 (0.51 - 0.53) | |
| Peripheral vascular disease | 0.31 (0.30 - 0.32) | |
| Cerebrovascular disease | 0.62 (0.62 - 0.63) | |
| Chronic pulmonary disease | 0.32 (0.32 - 0.33) | |
| Connective tissue disease | 0.35 (0.34 - 0.36) | |
| Ulcer disease | 0.43 (0.42 - 0.44) | |
| Mild liver disease | 0.49 (0.47 - 0.50) | |
| Any brain disorder | 0.21 (0.20 - 0.21) | 0.28 (0.27 - 0.28) |
| Male | -0.04 (-0.04 - -0.03) | -0.01 (-0.02 - -0.01) |

***Sensitivity analysis 3 / Prevalent cohort 2016***

| | *CCI covariates included in model* | *No CCI covariates included in model* |
|---|---|---|
| Age 10-19 | -0.01 (-0.01 - -0.00) | -0.02 (-0.02 - -0.02) |
| Age 20-29 | 0.02 (0.01 - 0.02) | -0.01 (-0.01 - -0.00) |
| Age 30-39 | 0.02 (0.02 - 0.03) | 0.00 (0.00 - 0.01) |
| Age 40-49 | 0.01 (0.00 - 0.01) | 0.01 (0.01 - 0.01) |
| Age 50-59 | 0.05 (0.04 - 0.05) | 0.09 (0.09 - 0.09) |
| Age 60-69 | 0.13 (0.13 - 0.14) | 0.25 (0.25 - 0.26) |
| Age 70-79 | 0.32 (0.31 - 0.32) | 0.54 (0.53 - 0.54) |
| Age 80-89 | 0.93 (0.92 - 0.93) | 1.25 (1.24 - 1.25) |
| Age 90+ | 2.29 (2.28 - 2.31) | 2.61 (2.59 - 2.62) |
| Myocardial infarction | 0.04 (0.03 - 0.05) | |
| Diabetes without end-organ damage | 0.33 (0.32 - 0.33) | |
| Hemiplegia | 1.34 (1.32 - 1.36) | |
| Moderate to severe renal disease | 0.73 (0.72 - 0.74) | |
| Diabetes with end-organ damage | 0.44 (0.43 - 0.45) | |
| Solid cancer tumour | 0.33 (0.33 - 0.34) | |
| Leukaemia | 0.91 (0.89 - 0.94) | |
| Lymphoma | 0.89 (0.88 - 0.91) | |
| Moderate to severe liver disease | 0.78 (0.75 - 0.81) | |
| Metastatic solid tumour | 0.95 (0.94 - 0.97) | |
| AIDS | 0.30 (0.27 - 0.34) | |
| Congestive heart failure | 0.51 (0.50 - 0.52) | |
| Peripheral vascular disease | 0.30 (0.29 - 0.31) | |
| Cerebrovascular disease | 0.59 (0.58 - 0.59) | |
| Chronic pulmonary disease | 0.31 (0.31 - 0.32) | |
| Connective tissue disease | 0.34 (0.33 - 0.35) | |
| Ulcer disease | 0.40 (0.39 - 0.41) | |
| Mild liver disease | 0.46 (0.45 - 0.48) | |
| Any brain disorder | 0.21 (0.21 - 0.21) | 0.28 (0.28 - 0.28) |
| Male | -0.03 (-0.03 - -0.03) | -0.01 (-0.01 - -0.01) |

***Sensitivity analysis 3 / Prevalent cohort 2017***

| | *CCI covariates included in model* | *No CCI covariates included in model* |
|---|---|---|
| Age 10-19 | -0.01 (-0.01 - -0.00) | -0.02 (-0.02 - -0.02) |
| Age 20-29 | 0.02 (0.01 - 0.02) | -0.01 (-0.01 - -0.00) |
| Age 30-39 | 0.02 (0.02 - 0.03) | 0.01 (0.00 - 0.01) |
| Age 40-49 | 0.01 (0.01 - 0.01) | 0.01 (0.01 - 0.02) |
| Age 50-59 | 0.05 (0.04 - 0.05) | 0.09 (0.09 - 0.10) |
| Age 60-69 | 0.14 (0.14 - 0.14) | 0.26 (0.26 - 0.27) |
| Age 70-79 | 0.32 (0.32 - 0.33) | 0.55 (0.55 - 0.56) |
| Age 80-89 | 0.91 (0.90 - 0.91) | 1.24 (1.23 - 1.24) |
| Age 90+ | 2.24 (2.23 - 2.25) | 2.56 (2.55 - 2.57) |
| Myocardial infarction | 0.05 (0.04 - 0.06) | |
| Diabetes without end-organ damage | 0.34 (0.33 - 0.34) | |
| Hemiplegia | 1.43 (1.40 - 1.45) | |
| Moderate to severe renal disease | 0.75 (0.74 - 0.76) | |
| Diabetes with end-organ damage | 0.47 (0.46 - 0.48) | |
| Solid cancer tumour | 0.35 (0.34 - 0.35) | |
| Leukaemia | 0.94 (0.91 - 0.96) | |
| Lymphoma | 0.95 (0.93 - 0.97) | |
| Moderate to severe liver disease | 0.80 (0.77 - 0.83) | |
| Metastatic solid tumour | 0.96 (0.95 - 0.98) | |
| AIDS | 0.28 (0.25 - 0.31) | |
| Congestive heart failure | 0.49 (0.48 - 0.50) | |
| Peripheral vascular disease | 0.31 (0.30 - 0.31) | |
| Cerebrovascular disease | 0.58 (0.57 - 0.59) | |
| Chronic pulmonary disease | 0.32 (0.32 - 0.33) | |
| Connective tissue disease | 0.33 (0.33 - 0.34) | |
| Ulcer disease | 0.40 (0.39 - 0.42) | |
| Mild liver disease | 0.46 (0.44 - 0.47) | |
| Any brain disorder | 0.22 (0.22 - 0.22) | 0.30 (0.29 - 0.30) |
| Male | -0.04 (-0.04 - -0.03) | -0.01 (-0.02 - -0.01) |

***Sensitivity analysis 3 / Prevalent cohort 2018***

| | *CCI covariates included in model* | *No CCI covariates included in model* |
|---|---|---|
| Age 10-19 | -0.01 (-0.01 - -0.00) | -0.02 (-0.02 - -0.02) |
| Age 20-29 | 0.01 (0.00 - 0.01) | -0.01 (-0.02 - -0.01) |
| Age 30-39 | 0.00 (0.00 - 0.01) | -0.01 (-0.02 - -0.01) |
| Age 40-49 | -0.01 (-0.02 - -0.01) | -0.01 (-0.02 - -0.01) |
| Age 50-59 | 0.02 (0.02 - 0.02) | 0.05 (0.05 - 0.05) |
| Age 60-69 | 0.11 (0.10 - 0.11) | 0.20 (0.19 - 0.20) |
| Age 70-79 | 0.28 (0.28 - 0.28) | 0.44 (0.44 - 0.45) |
| Age 80-89 | 0.87 (0.86 - 0.87) | 1.11 (1.10 - 1.11) |
| Age 90+ | 2.22 (2.21 - 2.23) | 2.47 (2.46 - 2.48) |
| Myocardial infarction | 0.03 (0.02 - 0.04) | |
| Diabetes without end-organ damage | 0.28 (0.28 - 0.29) | |
| Hemiplegia | 1.30 (1.28 - 1.33) | |
| Moderate to severe renal disease | 0.44 (0.43 - 0.45) | |
| Diabetes with end-organ damage | 0.35 (0.35 - 0.36) | |
| Solid cancer tumour | 0.17 (0.17 - 0.18) | |
| Leukaemia | 0.89 (0.87 - 0.92) | |
| Lymphoma | 0.51 (0.49 - 0.52) | |
| Moderate to severe liver disease | 0.77 (0.75 - 0.80) | |
| Metastatic solid tumour | 0.45 (0.44 - 0.47) | |
| AIDS | 0.10 (0.07 - 0.13) | |
| Congestive heart failure | 0.42 (0.41 - 0.43) | |
| Peripheral vascular disease | 0.25 (0.25 - 0.26) | |
| Cerebrovascular disease | 0.55 (0.55 - 0.56) | |
| Chronic pulmonary disease | 0.26 (0.26 - 0.27) | |
| Connective tissue disease | 0.11 (0.10 - 0.12) | |
| Ulcer disease | 0.40 (0.39 - 0.41) | |
| Mild liver disease | 0.38 (0.37 - 0.40) | |
| Any brain disorder | 0.19 (0.19 - 0.20) | 0.26 (0.25 - 0.26) |
| Male | -0.03 (-0.03 - -0.02) | -0.01 (-0.01 - -0.00) |

***Sensitivity analysis 3 / Prevalent cohort 2019***

| | *CCI covariates included in model* | *No CCI covariates included in model* |
|---|---|---|
| Age 10-19 | -0.02 (-0.02 - -0.01) | -0.03 (-0.03 - -0.02) |
| Age 20-29 | 0.02 (0.01 - 0.02) | -0.01 (-0.01 - -0.00) |
| Age 30-39 | 0.02 (0.01 - 0.02) | -0.00 (-0.00 - 0.00) |
| Age 40-49 | -0.01 (-0.01 - -0.00) | -0.01 (-0.01 - -0.00) |
| Age 50-59 | 0.03 (0.02 - 0.03) | 0.06 (0.06 - 0.07) |
| Age 60-69 | 0.11 (0.11 - 0.12) | 0.22 (0.21 - 0.22) |
| Age 70-79 | 0.29 (0.29 - 0.29) | 0.49 (0.48 - 0.49) |
| Age 80-89 | 0.85 (0.84 - 0.86) | 1.13 (1.13 - 1.14) |
| Age 90+ | 2.24 (2.23 - 2.25) | 2.51 (2.50 - 2.53) |
| Myocardial infarction | 0.03 (0.02 - 0.03) | |
| Diabetes without end-organ damage | 0.32 (0.31 - 0.32) | |
| Hemiplegia | 1.37 (1.35 - 1.40) | |
| Moderate to severe renal disease | 0.60 (0.60 - 0.61) | |
| Diabetes with end-organ damage | 0.44 (0.43 - 0.45) | |
| Solid cancer tumour | 0.28 (0.27 - 0.28) | |
| Leukaemia | 0.81 (0.79 - 0.84) | |
| Lymphoma | 0.87 (0.85 - 0.89) | |
| Moderate to severe liver disease | 0.78 (0.76 - 0.81) | |
| Metastatic solid tumour | 0.78 (0.76 - 0.80) | |
| AIDS | 0.18 (0.15 - 0.21) | |
| Congestive heart failure | 0.42 (0.41 - 0.43) | |
| Peripheral vascular disease | 0.27 (0.26 - 0.28) | |
| Cerebrovascular disease | 0.53 (0.52 - 0.53) | |
| Chronic pulmonary disease | 0.29 (0.29 - 0.30) | |
| Connective tissue disease | 0.22 (0.21 - 0.23) | |
| Ulcer disease | 0.37 (0.36 - 0.38) | |
| Mild liver disease | 0.39 (0.38 - 0.41) | |
| Any brain disorder | 0.21 (0.21 - 0.21) | 0.28 (0.28 - 0.28) |
| Male | -0.03 (-0.03 - -0.03) | -0.01 (-0.01 - -0.01) |

***Sensitivity analysis 3 / Prevalent cohort 2020***

| | *CCI covariates included in model* | *No CCI covariates included in model* |
|---|---|---|
| Age 10-19 | -0.01 (-0.01 - -0.01) | -0.02 (-0.03 - -0.02) |
| Age 20-29 | 0.02 (0.02 - 0.02) | -0.00 (-0.01 - 0.00) |
| Age 30-39 | 0.02 (0.01 - 0.02) | -0.00 (-0.01 - 0.00) |
| Age 40-49 | -0.01 (-0.01 - -0.00) | -0.01 (-0.01 - -0.01) |
| Age 50-59 | 0.02 (0.02 - 0.03) | 0.06 (0.06 - 0.07) |
| Age 60-69 | 0.12 (0.11 - 0.12) | 0.23 (0.22 - 0.23) |
| Age 70-79 | 0.30 (0.30 - 0.31) | 0.51 (0.51 - 0.52) |
| Age 80-89 | 0.86 (0.85 - 0.86) | 1.15 (1.15 - 1.16) |
| Age 90+ | 2.25 (2.24 - 2.26) | 2.54 (2.53 - 2.55) |
| Myocardial infarction | 0.04 (0.03 - 0.05) | |
| Diabetes without end-organ damage | 0.33 (0.32 - 0.33) | |
| Hemiplegia | 1.39 (1.37 - 1.42) | |
| Moderate to severe renal disease | 0.66 (0.65 - 0.67) | |
| Diabetes with end-organ damage | 0.43 (0.42 - 0.44) | |
| Solid cancer tumour | 0.31 (0.31 - 0.32) | |
| Leukaemia | 0.87 (0.84 - 0.89) | |
| Lymphoma | 0.96 (0.94 - 0.97) | |
| Moderate to severe liver disease | 0.82 (0.80 - 0.85) | |
| Metastatic solid tumour | 0.87 (0.85 - 0.88) | |
| AIDS | 0.16 (0.13 - 0.19) | |
| Congestive heart failure | 0.44 (0.43 - 0.45) | |
| Peripheral vascular disease | 0.29 (0.28 - 0.30) | |
| Cerebrovascular disease | 0.54 (0.53 - 0.55) | |
| Chronic pulmonary disease | 0.29 (0.28 - 0.29) | |
| Connective tissue disease | 0.23 (0.22 - 0.23) | |
| Ulcer disease | 0.40 (0.39 - 0.41) | |
| Mild liver disease | 0.40 (0.39 - 0.42) | |
| Any brain disorder | 0.23 (0.23 - 0.23) | 0.30 (0.30 - 0.30) |
| Male | -0.03 (-0.03 - -0.03) | -0.01 (-0.01 - -0.00) |

***Sensitivity analysis 3 / Prevalent cohort 2021***

| | *CCI covariates included in model* | *No CCI covariates included in model* |
|---|---|---|
| Age 10-19 | -0.01 (-0.01 - -0.01) | -0.02 (-0.02 - -0.02) |
| Age 20-29 | 0.02 (0.01 - 0.02) | -0.00 (-0.01 - -0.00) |
| Age 30-39 | 0.01 (0.01 - 0.02) | -0.01 (-0.01 - -0.00) |
| Age 40-49 | -0.02 (-0.02 - -0.01) | -0.02 (-0.02 - -0.01) |
| Age 50-59 | 0.02 (0.01 - 0.02) | 0.05 (0.05 - 0.06) |
| Age 60-69 | 0.10 (0.10 - 0.11) | 0.21 (0.21 - 0.22) |
| Age 70-79 | 0.30 (0.29 - 0.30) | 0.50 (0.50 - 0.50) |
| Age 80-89 | 0.84 (0.84 - 0.85) | 1.13 (1.12 - 1.13) |
| Age 90+ | 2.23 (2.22 - 2.25) | 2.52 (2.50 - 2.53) |
| Myocardial infarction | 0.03 (0.02 - 0.04) | |
| Diabetes without end-organ damage | 0.34 (0.34 - 0.35) | |
| Hemiplegia | 1.49 (1.46 - 1.51) | |
| Moderate to severe renal disease | 0.66 (0.65 - 0.67) | |
| Diabetes with end-organ damage | 0.41 (0.40 - 0.42) | |
| Solid cancer tumour | 0.29 (0.28 - 0.29) | |
| Leukaemia | 0.77 (0.74 - 0.79) | |
| Lymphoma | 0.95 (0.93 - 0.97) | |
| Moderate to severe liver disease | 0.79 (0.76 - 0.82) | |
| Metastatic solid tumour | 0.88 (0.86 - 0.89) | |
| AIDS | 0.15 (0.12 - 0.19) | |
| Congestive heart failure | 0.45 (0.44 - 0.46) | |
| Peripheral vascular disease | 0.28 (0.28 - 0.29) | |
| Cerebrovascular disease | 0.51 (0.51 - 0.52) | |
| Chronic pulmonary disease | 0.28 (0.28 - 0.29) | |
| Connective tissue disease | 0.22 (0.21 - 0.22) | |
| Ulcer disease | 0.42 (0.40 - 0.43) | |
| Mild liver disease | 0.39 (0.37 - 0.40) | |
| Any brain disorder | 0.24 (0.24 - 0.24) | 0.31 (0.31 - 0.32) |
| Male | -0.03 (-0.03 - -0.03) | -0.01 (-0.01 - -0.01) |

### Cost regression
Sensitivity analysis 3 / Prevalent cohort 2015 / No CCI variables in model

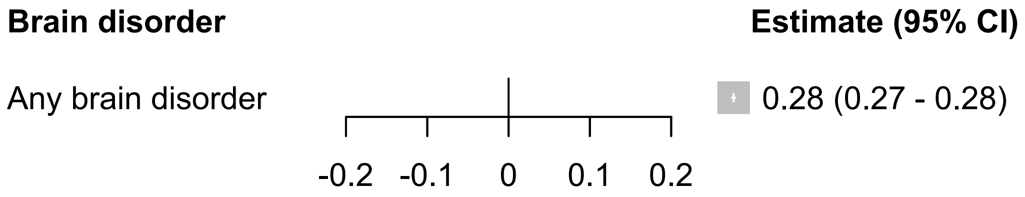

### Cost regression
Sensitivity analysis 3 / Prevalent cohort 2015 / CCI variables included in model

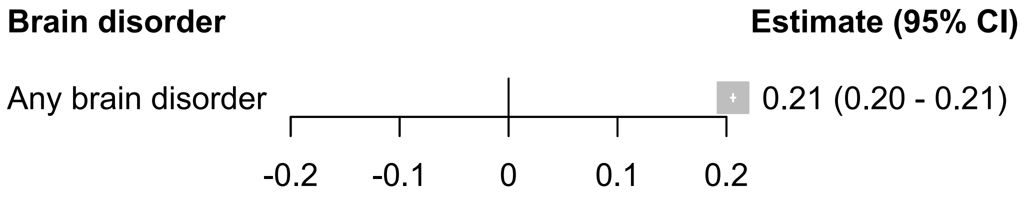

### Cost regression

Sensitivity analysis 3 / Prevalent cohort 2016 / No CCI variables in model

| Brain disorder | | Estimate (95% CI) |
|---|---|---|
| Any brain disorder | 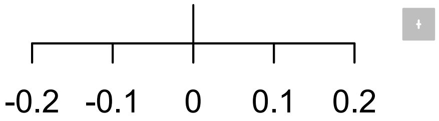 | 0.28 (0.28 - 0.28) |

### Cost regression

Sensitivity analysis 3 / Prevalent cohort 2016 / CCI variables included in model

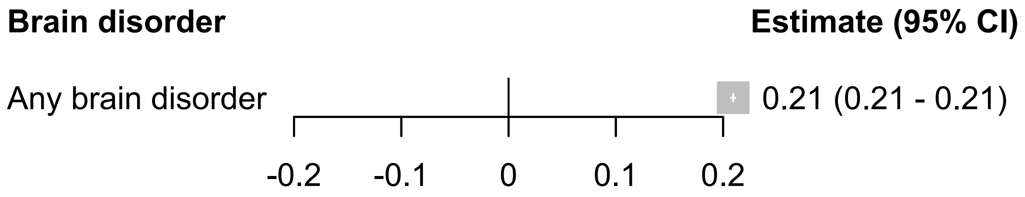

### Cost regression

#### Sensitivity analysis 3 / Prevalent cohort 2017 / No CCI variables in model

| Brain disorder | Estimate (95% CI) |
| --- | --- |
| Any brain disorder | 0.30 (0.29 - 0.30) |

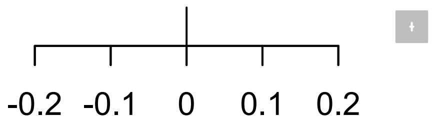

#### Cost regression

Sensitivity analysis 3 / Prevalent cohort 2017 / CCI variables included in model

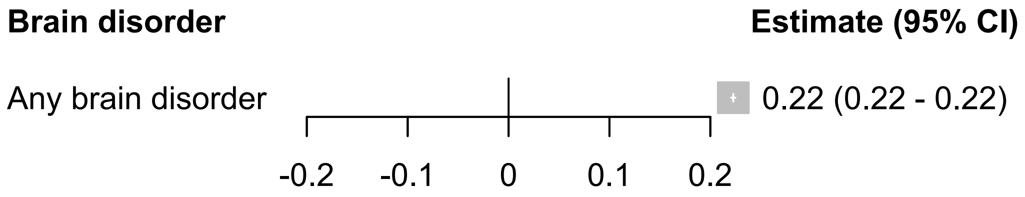

### Cost regression

Sensitivity analysis 3 / Prevalent cohort 2018 / No CCI variables in model

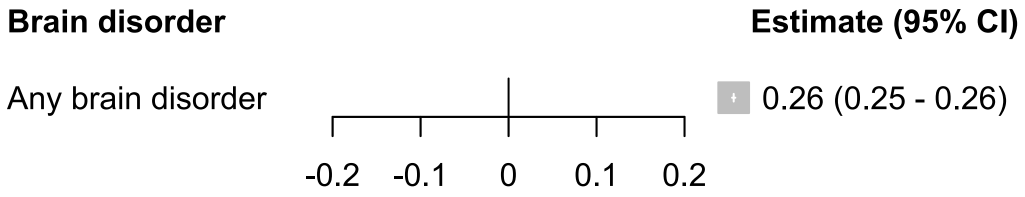

### Cost regression

Sensitivity analysis 3 / Prevalent cohort 2018 / CCI variables included in model

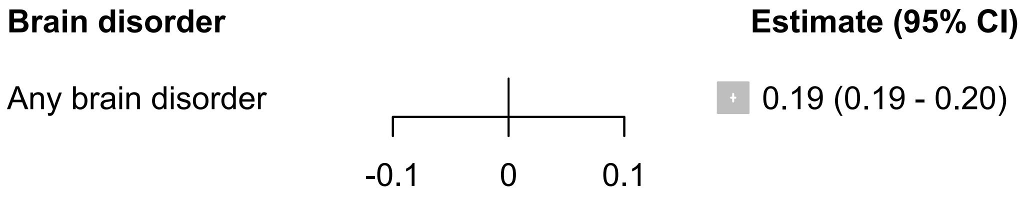

### Cost regression

#### Sensitivity analysis 3 / Prevalent cohort 2019 / No CCI variables in model

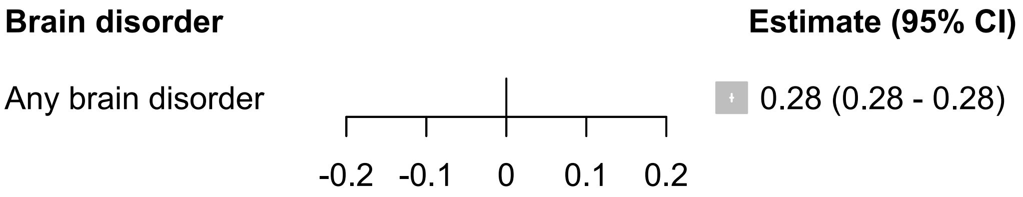

### Cost regression

Sensitivity analysis 3 / Prevalent cohort 2019 / CCI variables included in model

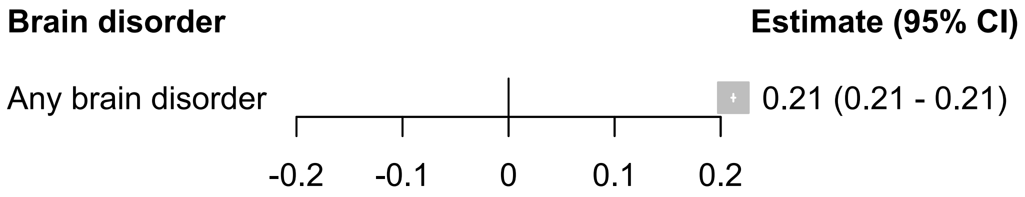

### Cost regression

Sensitivity analysis 3 / Prevalent cohort 2020 / No CCI variables in model

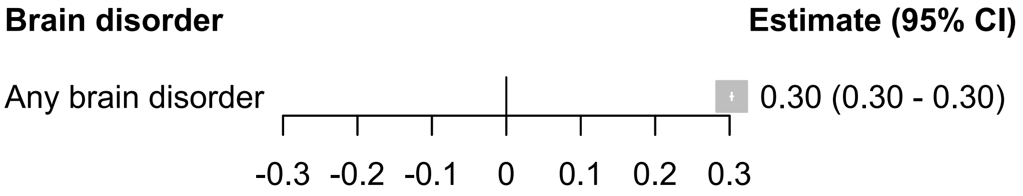

### Cost regression

Sensitivity analysis 3 / Prevalent cohort 2020 / CCI variables included in model

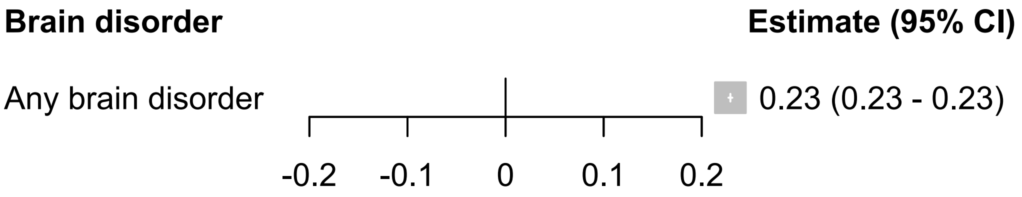

#### Cost regression
Sensitivity analysis 3 / Prevalent cohort 2021 / No CCI variables in model

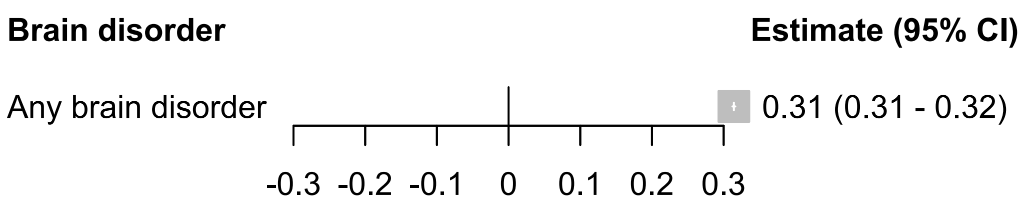

### Cost regression

Sensitivity analysis 3 / Prevalent cohort 2021 / CCI variables included in model

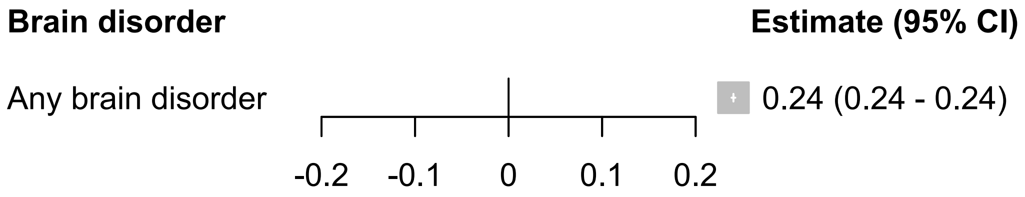
